# Disaggregating Asian Race Reveals COVID-19 Disparities among Asian Americans at New York City’s Public Hospital System

**DOI:** 10.1101/2020.11.23.20233155

**Authors:** Roopa Kalyanaraman Marcello, Johanna Dolle, Areeba Tariq, Sharanjit Kaur, Linda Wong, Joan Curcio, Rosy Thachil, Stella S. Yi, Nadia Islam

## Abstract

There is growing recognition of the burden of COVID-19 among Asian Americans, but data on outcomes among Asian ethnic subgroups remain extremely limited. We conducted a retrospective analysis of 85,328 patients tested for COVID-19 at New York City’s public hospital system between March 1 and May 31, 2020, to describe characteristics and COVID-19 outcomes of Asian ethnic subgroups compared to Asians overall and other racial/ethnic groups. South Asians had the highest rates of positivity and hospitalization among Asians, second only to Hispanics for positivity and Blacks for hospitalization. Chinese patients had the highest mortality rate of all groups and were nearly 1.5 times more likely to die than Whites. The high burden of COVID-19 among South Asian and Chinese Americans underscores the urgent needs for improved data collection and reporting as well as public health program and policy efforts to mitigate the disparate impact of COVID-19 among these communities.

---

New York City (NYC) was the first region in the United States (U.S.) to experience a significant burden of COVID-19, with more than 200,000 cases, 50,000 hospitalizations, and 17,000 deaths between March 1 and May 31, 2020.^1^ NYC Health + Hospitals (NYC H+H), the city’s public hospital system and the largest in the nation, was hit hardest in the city and the nation during this time, with many of its 11 hospitals having neared capacity and additional temporary facilities having opened to accommodate the thousands of patients with severe illness.

Evidence from NYC and the nation since that time, including from the Centers for Disease Control and Prevention (CDC), has shown that Blacks and Hispanics bear a substantially higher burden of COVID-19 than Whites, with Asian Americans reported as experiencing only a slightly higher burden than Whites.^2,3,4,5,6^ However, community leaders and experts in Asian American health in NYC – which is home to the largest overall Asian and South Asian populations in the nation – and across the country have voiced concerns about the lack of attention to the substantial burden of COVID-19 among Asian Americans, many of whom have similar clinical, social, and economic characteristics as other Americans of color.^7,8,9,10^ This is due in large part to two key factors: 1) an undercount of Asian Americans in health system records because of inadequate or inaccurate data collection and reporting of Asian Americans’ race and/or ethnicity (i.e., as “other,” “unknown,” or an incorrect race, e.g., American Indian/Native American race instead of Asian race and Indian ethnicity), and 2) aggregating all Asian ethnic groups into a single race category, thereby obscuring differences in characteristics and outcomes between these diverse groups.

A recent systematic review and meta-analysis of 50 studies from the U.S. and the United Kingdom (U.K.) found a higher risk of COVID-19 infection among Asians and Blacks are compared to Whites and a likely higher risk of intensive care unit admission and death only among Asians as compared to Whites.^11^ Several studies from the U.K., where there is a large Asian and especially South Asian population, have shown an increased burden of COVID-19 among certain Asian ethnic subgroups, mostly South Asian and Indo-Caribbean communities.^12^

Recent media reports in the U.S. have begun to illuminate the heretofore largely invisible burden of COVID-19 among Asian Americans, but data to this end are still lacking from health systems and health departments.^13,14,15^ A recent CDC analysis of excess deaths due to COVID-19 found a substantial burden among Asian Americans, with a 36.6% increase in deaths in 2020 as compared to the average from 2015-2019, far higher than the 11.9% increase among Whites and second only to the 53.6% increase among Hispanics.^16^ Still, the impact of COVID-19 among Asian Americans and distinct Asian ethnic subgroups has not yet been fully elucidated.

As such, this study has two key objectives. First, we sought to characterize COVID-19 clinical and demographic risk factors and outcomes among Asian Americans tested for COVID-19 at NYC’s public hospital system, which serves thousands of Asian Americans each year. Second, we sought to identify if differences exist in characteristics and outcomes between Asian ethnic subgroups and other racial groups.

## Study Data and Methods

### Data Sources

NYC H+H’s electronic health record (EHR) database was used to identify all patients who received a SARS-CoV-2 test at NYC H+H between March 1 and May 31, with follow up through August 15, 2020. Demographic data and select comorbidities were extracted for all patients, where available, as were COVID-19-related outcomes (test dates, test results, dates of hospitalization and discharge, and date of in-hospital death, where applicable). The primary variables of interest were race, ethnicity, age, and COVID-19-related outcomes (positive test, hospitalization, in-hospital mortality). NYC H+H classifies Hispanic ethnicity as a unique race category rather than an additional ethnicity.

Owing to known deficiencies in completeness of recorded race and ethnicity within the NYC H+H EHR, race, ethnicity, and language data in the EHR and validated Asian surname lists (**Supplement Table 1**) were used to classify individuals into one of three Asian ethnic groups (South Asian [Afghani, Bangladeshi, Indian, Nepalese, Pakistani, Sri Lankan], Chinese, and all other Asian).^17,18,19^ Because of the large overlap between Filipino and Hispanic surnames, we did not categorize any surnames as Filipino; however, Filipino patients whose race and/or ethnicity were recorded as Asian and/or Filipino in the EHR were categorized into the “other Asian” group.

**Table 1.** Characteristics and Comorbidities of Patients Tested for SARS-CoV-2 by Race and Ethnicity

|  |  |  |  |  |  |  | *d* |  |  |  |  |  |  |  |
| --- | --- | --- | --- | --- | --- | --- | --- | --- | --- | --- | --- | --- | --- | --- |
|  | **Total** | **White** | **Black** | **Hispanic** | **Asian** | *South Asian<sup>d</sup>* | *% of Asian* | *Chinese* | *% of Asian* | *Other Asian* | *% of Asian* | **Other** | **Unknown** | **P value** |
| No. (%) | 85328 (100.0) | 8037 (9.4) | 20827 (24.4) | 25119 (29.4) | 9971 (11.7) | 4804 (5.6) | 48.2% | 2214 (2.6) | 22.2% | 2953 (3.5) | 29.6% | 13505 (15.8) | 7869 (9.2) |  |
| **Sex** |  |  |  |  |  |  |  |  |  |  |  |  |  |  |
| Male | 40306 (47.2) | 4025 (50.1) | 8822 (42.4) | 12550 (50.0) | 4469 (44.8) | 2350 (48.9) |  | 1026 (46.3) |  | 1093 (37.0) |  | 6523 (48.3) | 3917 (49.8) | <0.01 |
| **Age** |  |  |  |  |  |  |  |  |  |  |  |  |  |  |
| Median [IQR] | 51 [38, 62] | 53 [37, 64] | 54 [40,64] | 51 [40,62] | 49 [35,61] | 49 [35,60] |  | 53 [38,64] |  | 47 [35,59] |  | 48 [34,60] | 49 [35,61] |  |
| 18-44 | 31381 (36.8) | 2903 (36.1) | 6510 (31.3) | 8662 (34.5) | 4124 (41.4) | 2054 (42.8) |  | 762 (34.4) |  | 1308 (44.3) |  | 5904 (43.7) | 3278 (41.7) | <0.01 |
| 45-64 | 36927 (43.3) | 3126 (38.9) | 9564 (45.9) | 11428 (45.5) | 4163 (41.8) | 1939 (40.4) |  | 936 (42.3) |  | 1288 (43.6) |  | 5462 (40.4) | 3184 (40.5) |  |
| 65-74 | 10647 (12.5) | 1067 (13.3) | 2964 (14.2) | 3188 (12.7) | 1079 (10.8) | 542 (11.3) |  | 293 (13.2) |  | 244 (8.3) |  | 1403 (10.4) | 946 (12.0) |  |
| 75+ | 6373 (7.5) | 941 (11.7) | 1789 (8.6) | 1841 (7.3) | 605 (6.1) | 269 (5.6) |  | 223 (10.1) |  | 113 (3.8) |  | 736 (5.4) | 461 (5.9) |  |
| **Payer** |  |  |  |  |  |  |  |  |  |  |  |  |  |  |
| Commercial | 22464 (26.3) | 3083 (38.4) | 6342 (30.5) | 3471 (13.8) | 3687 (37.0) | 1580 (32.9) |  | 766 (34.6) |  | 1341 (45.4) |  | 3965 (29.4) | 1916 (24.3) | <0.01 |
| Emergency Medicaid | 2409 (2.8) | 91 (1.1) | 254 (1.2) | 1559 (6.2) | 195 (2.0) | 104 (2.2) |  | 44 (2.0) |  | 47 (1.6) |  | 234 (1.7) | 76 (1.0) |  |
| Medicaid | 17809 (20.9) | 1200 (14.9) | 5154 (24.7) | 5180 (20.6) | 1801 (18.1) | 1128 (23.5) |  | 290 (13.1) |  | 383 (13.0) |  | 3379 (25.0) | 1095 (13.9) |  |
| Medicare | 12602 (14.8) | 1646 (20.5) | 3812 (18.3) | 3600 (14.3) | 1050 (10.5) | 500 (10.4) |  | 371 (16.8) |  | 179 (6.1) |  | 1733 (12.8) | 761 (9.7) |  |
| Self-Pay | 27393 (32.1) | 1790 (22.3) | 4659 (22.4) | 10449 (41.6) | 2986 (29.9) | 1363 (28.4) |  | 697 (31.5) |  | 926 (31.4) |  | 3684 (27.3) | 3825 (48.6) |  |
| Other | 2651 (3.1) | 227 (2.8) | 606 (2.9) | 860 (3.4) | 252 (2.5) | 129 (2.7) |  | 46 (2.1) |  | 77 (2.6) |  | 510 (3.8) | 196 (2.5) |  |
| **History with H+H** |  |  |  |  |  |  |  | 2214 |  |  |  |  |  |  |
| Acute Only | 8426 (9.9) | 849 (10.6) | 2801 (13.4) | 2336 (9.3) | 779 (7.8) | 468 (9.7) |  | 146 (6.6) |  | 165 (5.6) |  | 1418 (10.5) | 243 (3.1) | <0.01 |
| Outpatient | 36257 (42.5) | 3287 (40.9) | 10150 (48.7) | 11158 (44.4) | 5218 (52.3) | 2384 (49.6) |  | 948 (42.8) |  | 1886 (63.9) |  | 5382 (39.9) | 1062 (13.5) | <0.01 |
| New | 40645 | 3901 | 7876 | 11625 | 3974 | 1952 |  | 1120 |  | 902 |  | 6705 | 6564 |  |
|  | (47.6) | (48.5) | (37.8) | (46.3) | (39.9) | (40.6) |  | (50.6) |  | (30.5) |  | (49.6) | (83.4) |  |
| **Chronic conditions<sup>a</sup>** |  |  |  |  |  |  |  |  |  |  |  |  |  |  |
| Obesity<sup>b</sup> |  |  |  |  |  |  |  |  |  |  |  |  |  |  |
| BMI $\geq 30$ and $< 40$ | 12636<br>(14.8) | 967<br>(12.0) | 3898<br>(18.7) | 4545<br>(18.1) | 1085<br>(10.9) | 679<br>(14.1) | | 126<br>(5.7) | | 280 (9.5) | | 1735<br>(12.8) | 406<br>(5.2) | <0.01 |
| BMI $\geq 40$ | 2391 (2.8) | 202<br>(2.5) | 1020<br>(4.9) | 558 (2.2) | 127<br>(1.3) | 82 (1.7) | | 17 (0.8) | | 28 (0.9) | | 387 (2.9) | 97 (1.2) | |
| Chronic diseases<sup>c</sup> |  |  |  |  |  |  |  |  |  |  |  |  |  |  |
| Diabetes | 16499<br>(19.3) | 1138<br>(14.2) | 5058<br>(24.3) | 5657<br>(22.5) | 1900<br>(19.1) | 1108<br>(23.1) |  | 318<br>(14.4) |  | 474<br>(16.1) |  | 2238<br>(16.6) | 508<br>(6.5) | <0.01 |
| Hypertension | 21184<br>(24.8) | 1952<br>(24.3) | 7458<br>(35.8) | 5987<br>(23.8) | 2265<br>(22.7) | 1250<br>(26.0) |  | 452<br>(20.4) |  | 563<br>(19.1) |  | 2778<br>(20.6) | 744<br>(9.5) |  |
| Heart disease | 16535<br>(19.4) | 1786<br>(22.2) | 5407<br>(26.0) | 4600<br>(18.3) | 1862<br>(18.7) | 1055<br>(22.0) |  | 391<br>(17.7) |  | 416<br>(14.1) |  | 2316<br>(17.1) | 564<br>(7.2) |  |
| Cancer | 7082 (8.3) | 657<br>(8.2) | 2240<br>(10.8) | 2488<br>(9.9) | 706<br>(7.1) | 365 (7.6) |  | 167<br>(7.5) |  | 174 (5.9) |  | 822 (6.1) | 169(2.1) |  |
| Liver disease | 2622 (3.1) | 277<br>(3.4) | 671 (3.2) | 923 (3.7) | 276<br>(2.8) | 150 (3.1) |  | 66 (3.0) |  | 60 (2.0) |  | 404 (3.0) | 71 (0.9) |  |
| CKD | 4591 (5.4) | 413<br>(5.1) | 1952<br>(9.4) | 1101<br>(4.4) | 445<br>(4.5) | 255 (5.3) |  | 92 (4.2) |  | 98 (3.3) |  | 525 (3.9) | 155<br>(2.0) |  |

Abbreviations: IQR, inter quartile range; H+H, NYC Health + Hospitals; BMI, body mass index; CKD, chronic kidney disease.
<sup>a</sup>All percentages are based on the full population tested, not just individuals with available values.
<sup>b</sup>50.8% of patients tested did not have a BMI value available.
<sup>c</sup>Chronic disease status is based on the presence of a diagnosis in the patient's record.
<sup>d</sup>Overall percentage of Asian ethnic subgroups is among all Asians, not the full population tested.

The primary outcomes were a positive SARS-CoV-2 test, hospitalization for, and death from COVID-19. Secondary outcomes were patient demographics and comorbidities.

### Statistical Analysis

Patient demographics and comorbidities were summarized as descriptive statistics, with categorical data presented as frequency (percentage) and numeric data as mean (SD) or median (interquartile range [IQR]), as appropriate.

Pearson chi-square tests were used to examine differences between demographic and clinical characteristics by race and ethnicity. Multivariable logistic regression analyses were performed to assess whether racial/ethnic disparities in mortality persist after controlling for other demographics (i.e., sex, age) and comorbidities (e.g., diabetes, obesity) known to be associated with adverse outcomes from COVID-19 based on published literature. We ran two models: one aggregated all Asian ethnic subgroups into a single Asian race category and the other disaggregated Asian race into the three specified ethnic subgroups. We presented the association between risk factors and outcome of death as odds ratios and 95% confidence intervals. For all analyses, a p-value of <0.05 was considered statistically significant. All analyses were performed using R version 3.6 and SAS Enterprise Guide version 7.15.

This study was approved by the Biomedical Research Alliance of New York Institutional Review Board. Informed consent was not required because of the retrospective nature of this study.

### Limitations

This study had several limitations. First, it included only patients from a single health system; however, NYC H+H is the largest public hospital system in the nation, serving more than 1 million patients each year at more than 70 sites, including 11 hospitals, across the city’s five boroughs. NYC H+H serves a highly diverse patient population, including thousands of lower income Asian Americans, who are typically underrepresented in national datasets, so our findings can be generalized to other areas with similar Asian American populations that may be smaller or understudied.^20^ Second, we utilized only clinical and demographic data available from the EHR; social needs screening was rolled out across the system last year but collection and reporting remains limited, so we excluded these data. Third, nearly half of patients tested were new to NYC H+H, so a clinical history was not available for them. However, most new patients who were hospitalized had BMI and diagnoses entered in their EHR following their admission, so key clinical predictors of mortality were available for nearly all hospitalized patients. Fourth, we did not analyze outcomes for the group of patients categorized as Asian solely based on race entered in the EHR, as this group was nearly half the size of the group identified through surname-based ethnicity identification. Fifth, we did not classify as Filipino through surname matching any individuals who were not already identified as Filipino in the EHR; as such, this group is likely underrepresented in our analysis relative to other Asian ethnic subgroups, but, because Filipinos comprise a small proportion of all Asians in NYC, this likely does not bear any substantial impact upon our findings. Sixth, our reporting of test results and hospitalization by race/ethnicity was not adjusted for age. Seventh, we included only in-hospital mortality and did not include mortality for patients who died after discharge from the hospital. Finally, we did not adjust BMI categories for Asian ethnic groups to align with WHO recommendations, which categorize individuals of Asian race as overweight and obese at lower BMI values than the overall population. As such, the prevalence of overweight and obesity among individuals of Asian race is likely underestimated in our study.^10^

## Study Results

### Characteristics of Asian American Patients

Of the 85,328 adults tested for SARS-CoV-2, 9,971 (11.7%) were identified as Asian through a combination of EHR race and ethnicity data and surname matching. Of these, 4,804 (48.2%) were South Asian, 2,214 (22.2%) were Chinese, and 2,953 (29.6%) were of other Asian ethnic groups (**Table 1)**. Chinese patients were the oldest among Asians, with a median age of 53 years (IQR 38-64), and they were among the oldest of all racial and ethnic groups. Chinese patients were also most frequently new patients to NYC H+H (50.6%) among all Asian ethnic subgroups as well as all racial groups. South Asians were most likely among Asians to utilize Emergency Medicaid (2.2%) and Medicaid (23.6%), second overall only to Hispanics (6.2% and 24.7%, respectively). Among all racial groups, commercial insurance was utilized most frequently by White and Asian patients (38.4% and 37.0%, respectively), but this high rate among Asians was driven by individuals not of South Asian or Chinese descent (45.4%). Chronic diseases were prevalent among South Asians, who had a substantially higher rate of obesity (14.1% with BMI ≥30 and <40 and 1.7% with BMI ≥40), diabetes (23.1%), hypertension (26.0%), and heart disease (22.0%) than other Asian ethnic subgroups; these rates were comparable to and in some cases higher than those observed among Black and Hispanic patients.

### COVID-19 Outcomes of Asian American Patients

Among all racial groups, Asians had the second highest rate of testing positive (27.9%); South Asians had the highest rate among Asian ethnic groups (30.8%), second only to Hispanics (32.1%) overall (**Figure 1**). 51.6% of Asians who tested positive were hospitalized, a lower proportion than that of Blacks and Whites; however, rates were higher among Chinese (52.6%) and South Asian patients (54.7%), the latter of whom had the second highest rate among all groups.

**Figure 1.**
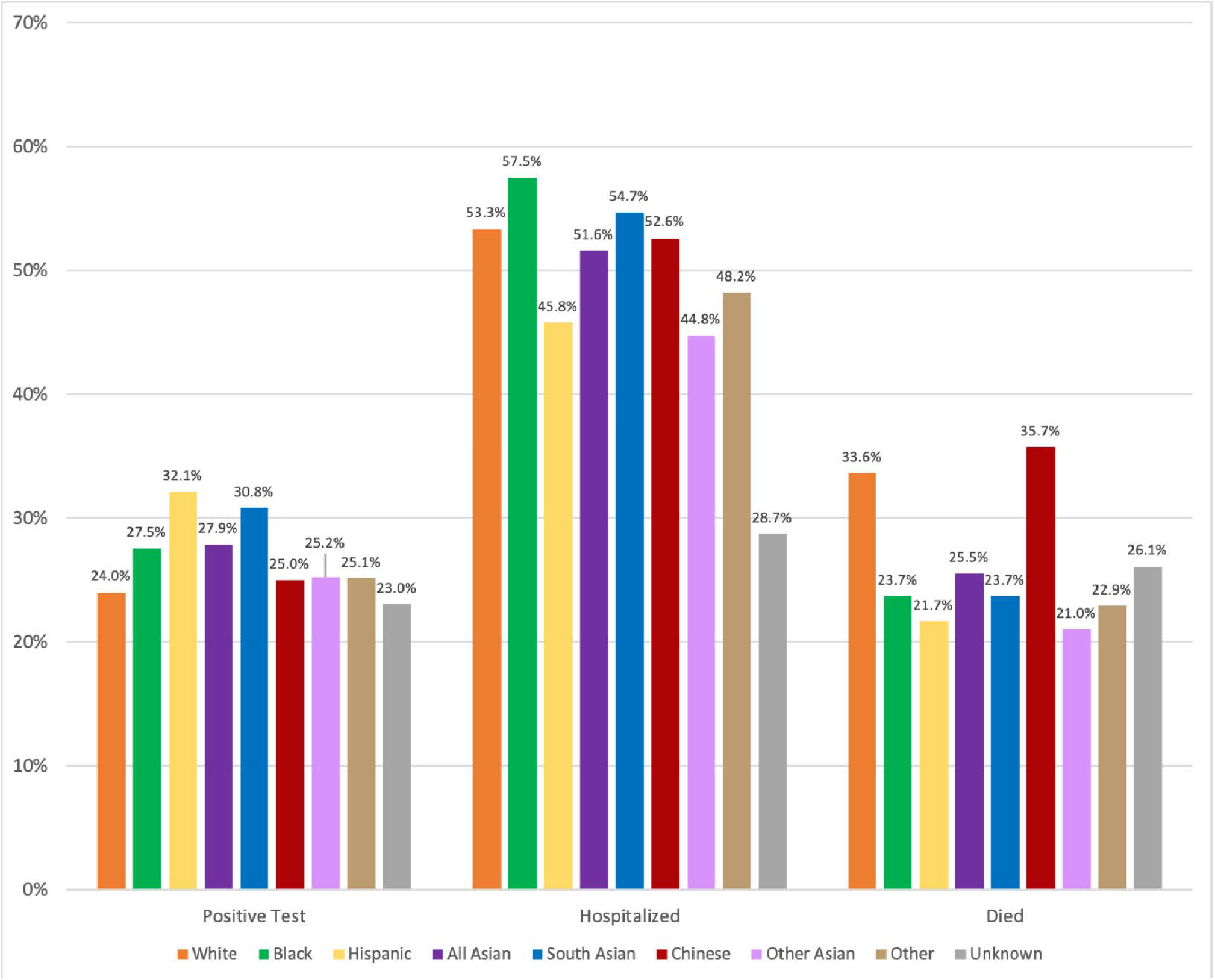
COVID-19 Outcomes by Race/Ethnicity. * *Rates are unadjusted*

The 25.5% mortality rate among Asians was second only to Whites (33.6%). Disaggregation into ethnic subgroups revealed that Chinese patients had a substantially higher mortality rate (35.7%) than South Asians (23.7%) and other Asians (21.0%), and this rate was the highest among all racial groups. This disparity in mortality among Chinese patients persisted even after adjusting for age, other demographics, and comorbidities (OR 1.44, 95% CI [1.035, 2.011], p=0.03) (**Table 2**). However, no disparity was observed between the aggregate Asian race group as compared to Whites (OR 1.15, 95% CI [0.93, 1.43], p=0.21). Use of Emergency Medicaid and being a new patient at NYC H+H were also associated with increased odds of death (OR 2.86, 95% CI [2.30, 3.54], p<0.001 and OR 2.03, 95% CI [1.79, 2.30], p<0.001, respectively).

**Table 2.** **Odds of Death among Patients Hospitalized with COVID-19 by Select Characteristics**

|  | **Adjusted OR (95% CI)**<br>**(n=9836)<sup>a</sup>** | **P-value** |
| --- | --- | --- |
| **Race/Ethnicity<sup>b</sup>** |  |  |
| White | Reference | --- |
| Black | 0.77 (0.64, 0.93) | 0.006 |
| Hispanic | 1.1 (0.91, 1.33) | 0.34 |
| Other | 0.99 (0.80, 1.23) | 0.95 |
| Unknown/Declined/Missing | 0.94 (0.69, 1.28) | 0.69 |
| *Asian<sup>c</sup>* | *1.15 (0.93, 1.43)* | *0.21* |
| South Asian | 1.06 (0.82, 1.37) | 0.68 |
| Chinese | 1.44 (1.04, 2.01) | 0.03 |
| Other Asian | 1.08 (0.76, 1.54) | 0.66 |
| **Payer<sup>d</sup>** |  |  |
| Commercial | Reference | --- |
| Emergency Medicaid | 2.86 (2.30, 3.54) | <0.001 |
| Medicaid | 1.03 (0.86, 1.24) | 0.76 |
| Medicare | 1.16 (0.97, 1.38) | 0.11 |
| Self-pay | 0.34 (0.26, 0.45) | <0.001 |
| Other | 0.86 (0.47, 1.58) | 0.62 |
| **History with H+H<sup>e</sup>** |  |  |
| Outpatient | Reference | --- |
| New | 2.03 (1.79, 2.30) | <0.001 |
| Acute Only | 1.36 (1.16, 1.60) | <0.001 |

Abbreviations: OR, odds ratio; CI, confidence interval; H+H, NYC Health + Hospitals.
<sup>a</sup>Only patients with a recorded BMI value were included.
<sup>b</sup>Reference group is White race.
<sup>c</sup>Results from Model 1 run with aggregated Asian race group; all other results are from Model 2 run with Asian race disaggregated into three ethnic groups.
<sup>d</sup>Reference group is commercial insurance.
<sup>e</sup>Reference group is patients with previous outpatient visits.

## Discussion

This study is the first to highlight disparities in COVID-19 outcomes among Asian American ethnic subgroups in the United States. We found a substantial burden of COVID-19 among South Asian and Chinese patients in NYC’s public hospital system, with rates similar to those observed among Blacks and Hispanics. Disaggregating Asian race into ethnic subgroups revealed a disproportionate burden of COVID-19 infection and hospitalization among South Asians and mortality among Chinese patients, with the latter having the highest likelihood of death among all racial/ethnic groups. Rates of positivity, hospitalization, and mortality were substantially lower among the overall Asian race group than among individual ethnic subgroups. To date, the disproportionate burden of COVID-19 among Asian ethnic subgroups has been masked in U.S.-based studies examining disparities by race/ethnicity by aggregating all Asian ethnic groups into a single Asian race category, which has subsequently hindered an appropriate public health response.

Asian Americans, especially those of South Asian and Chinese descent, have several key clinical risk factors in common with Blacks and Hispanics. Asians experience overweight and obesity at lower BMI values than individuals of other racial groups, resulting in a higher prevalence of these conditions than expected.^21^ Additionally, South Asians have high rates of diabetes and hypertension that are comparable to those observed among Black and Hispanic individuals, and they have a disproportionate burden of morbidity and mortality from cardiovascular disease. These factors are known to put individuals at elevated risk of COVID-19 infection, hospitalization, and death, and they are highly prevalent among many Asian Americans.^22,23^

Many Asian Americans experience social factors that are known to increase the risk of exposure for COVID-19, including living in multi-generational housing, jobs as frontline or essential workers, lack of paid sick leave, and limited access to linguistically and culturally appropriate healthcare.^24,25^ Furthermore, the social conditions that drive racial and ethnic disparities in diabetes and hypertension among Black and Hispanic Americans, including poverty, limited access to healthcare, limited English language proficiency, and other upstream determinants of health, are similar in the largely immigrant Asian patient population at NYC H+H.

The high rate of infection observed among South Asians may be due to factors that increase the likelihood of exposure (e.g., jobs in essential services) or impede the ability to isolate if infected (e.g., crowded housing, lack of paid sick leave) that are well-established as risk factors among other groups of color in the U.S. A recent analysis of community-level factors associated with racial and ethnic COVID-19 disparities found that “household size and food service occupation are strongly associated with the risk of COVID-19 infection” and that these factors “may be contributing to the higher number of COVID-19 cases in Black and Latino communities.”^26^ These factors are also common among New Yorkers of Asian descent, particularly among those who are recent immigrants, which may partially explain the high rate of infection we observed among South Asians. This same analysis also found that the proportion of foreign-born non-citizens was positively associated with the number of COVID-19 cases in a community, which may also explain higher rates of COVID-19 among Asian New Yorkers as well as among Hispanic New Yorkers, many of whom are also recent immigrants. Furthermore, Asian Americans are most likely of all racial groups to live in larger, multi-generational households, further driving their risk of COVID-19 exposure and infection.^27^

Data from the U.S. Bureau of Labor Statistics (BLS) indicating that Blacks and Hispanics are less able to work from home as compared to Whites and Asians have been cited to explain the social factors driving COVID-19, but these data do not take into account the socioeconomic variation within racial groups and further serve to mask the high burden of COVID-19 among Asian ethnic subgroups.^28^ NYC’s Asian diaspora is diverse both ethnically and socioeconomically, with many New Yorkers of South Asian descent working in low wage jobs for which working from home is not possible. U.S. BLS data show that only 9% of low-wage (<25th percentile) workers are able to work at home as compared to 62% of workers in the highest income quartile, which better explains higher infection rates among South Asians in NYC.

These social factors are even more common among recent immigrants, who may delay seeking care due to their immigration status. This is thought to have been exacerbated by the recent public charge rule, which puts legally present immigrants at risk of being denied permanent resident status if they utilize local, state, or federal government public benefits.^29^ An injunction was issued in July 2020 that prevented the U.S. Department of Homeland Security from enforcing the rule during the COVID-19 public health emergency, and the U.S. Customs and Immigration Services encouraged immigrants to seek care for COVID-19 symptoms, stating that they will “neither consider testing, treatment, nor preventative care (including vaccines, if a vaccine becomes available) related to COVID-19 as part of a public charge inadmissibility determination, nor as related to the public benefit condition applicable to certain nonimmigrants seeking an extension of stay or change of status, even if such treatment is provided or paid for by one or more public benefits, as defined in the rule (e.g. federally funded Medicaid).”^30^ However, anecdotes from community organizations in NYC that serve immigrants, especially those of Asian descent, and a recent study of immigrants in Texas found that immigrants avoided seeking medical care or enrolling in public benefits due to concerns about the public charge rule and the impact on their immigration status.^31,32,33^ This delay in or avoidance of seeking care may partially explain the high rates of hospitalization and mortality observed among South Asian and Chinese patients, respectively.

Our finding that Chinese Americans had the highest mortality rate of all racial/ethnic groups and were nearly 1.5 more times likely to die than Whites (OR 1.44, 95% CI [1.04, 2.01]) is concerning. This elevated burden was revealed only when the overall Asian race category was disaggregated into ethnic subgroups, as the overall Asian race group did not have a significantly higher likelihood of death than Whites (OR 1.15, 95% CI [0.93, 1.43], p=0.21). Since the emergence of COVID-19 in early 2020, Chinese and other Asian Americans have experienced increased xenophobia, discrimination, and harassment: one quarter of Asian New Yorkers surveyed reported witnessing or experiencing harassment, violence, or racism related to COVID-19, and more than half of Chinese American adults and their children across the U.S. reported being targeted by COVID-19 related racial discrimination either in person or online, which was associated with poorer mental health.^34,35,36^ Furthermore, this increase in harassment and racism may be leading to reluctance to and/or fear of leaving one’s home for care or testing, which may be exacerbating Chinese patients’ known reluctance to seek timely care, thereby leading to more severe illness that may be more difficult to treat successfully.^36,37^ Additionally, treatment patterns may have been influenced by early data on worse outcomes among Blacks and Hispanics and the “model minority” myth that Asians are typically healthier than other racial groups.^38^ Our findings underscore the urgency of additional research into the factors leading to higher mortality among Chinese Americans.

We found that patients utilizing Emergency Medicaid were nearly three times as likely to die as patients utilizing commercial insurance (OR 2.86, 95% CI [2.30, 3.54]). In our study, Emergency Medicaid use was most frequent among Hispanics (6.2%), followed by South Asians (2.2%) and both Asians overall and Chinese patients (2.0% for both), groups which contain large numbers of undocumented immigrants. The markedly higher likelihood of death observed among patients utilizing Emergency Medicaid likely reflects delayed care seeking among undocumented immigrants. In New York State, Emergency Medicaid for undocumented immigrants was expanded early in the pandemic to cover COVID-19 testing and treatment.^39^ However, it is likely that many undocumented immigrants delayed or even avoided care due to concerns over reporting of their immigration status.^40^

Similarly, we found that, as compared to patients with a history of outpatient primary or specialty care visits at NYC H+H, patients who were new to the NYC H+H system were twice as likely to die (OR 2.03, 95% CI [1.79, 2.30]) and patients who had only an inpatient or emergency department visit were 1.4 times more likely to die (OR 1.36, 95% CI [1.16, 1.60]). Observations from clinicians across the NYC H+H system indicate that this may be due in part to poorer health status among new patients and those who had previously only had an acute care visit, as they may not have been receiving regular care for chronic conditions or may have had poorly controlled and/or undiagnosed chronic conditions known to be a risk factor for COVID-19. These observations are aligned with research showing poorer health outcomes among patients who delay receiving care, particularly immigrants.^41,42,43^ These patients also may have presented with more severe COVID-19 illness owing to a lack of engagement with health care and concerns over presenting for care in light of federal rulings on public charge and immigration enforcement. We found that Chinese patients were most frequently new to NYC H+H (50.6%), which may reflect their known reluctance to and delay in seeking care, as described above.^36^

Asian Americans are facing unique challenges specific to COVID-19, but these have been largely overlooked to date by healthcare and public health agencies. As mentioned previously, Asian Americans, especially those of Chinese heritage, have experienced a significant increase in xenophobia and harassment, which affects not only their COVID-19 care seeking but also worsens mental health. Additionally, although our data do not report specifically on Filipino Americans, recent data show that Filipino nurses are dying at a disproportionately higher rate than other nurses: they comprise just 4% of the nursing workforce in the U.S. but nearly one third of COVID-19 deaths among this group.^44^ This is due in part to their overrepresentation in higher-risk roles in hospitals, including intensive care units, as well as to their frequent role as family caregivers.^45^ Finally, the COVID-19 pandemic has had an unprecedented economic impact on Asian Americans, many of whom are employees or owners of small businesses, with unemployment rates soaring due to business closures: more than one in 10 Asian Americans in the U.S. are currently unemployed.^46^ In New York City, the unemployment rate among Asians soared from 3.4% in February to a staggering 25.6% in May; although this has been slowly declining in recent months, this dramatic rise in unemployment is the largest among any racial/ethnic group.^47,48,49,50^

Our findings shed important light on the hidden burden of COVID-19 among Asian Americans. Community leaders and experts in Asian American health in NYC and across the country have voiced concerns about the substantial burden of COVID-19 among Asian Americans, particularly among specific ethnic subgroups, since the start of the pandemic in NYC, yet data to this end have been lacking.^7,8,15^ The lack of data on the burden of COVID-19 among Asian ethnic subgroups in the U.S. has impeded an appropriate public health response, including targeted communication campaigns on prevention and enhanced testing and tracing among the hardest hit communities.

These findings also provide critical insights that can help inform public health initiatives and policies to address the disproportionate burden of COVID-19 among communities of color, including Asian Americans. Specifically, policies and strategies that improve access to testing, isolation, and early care may help reduce disparities and mitigate the spread of COVID-19 among communities that have been hardest hit and will likely continue to be at high risk of exposure as a result of their employment and housing as well as at increased risk of hospitalization and death due to their immigration status.

## Conclusion

In New York City, substantial differences exist in COVID-19 outcomes between Asian ethnic subgroups, but these have been masked by reporting all Asian ethnic subgroups as a single Asian racial group. Specifically, South Asian and Chinese Americans experience adverse COVID-19 outcomes at high rates that are comparable to those observed – and known for several months – among Blacks and Hispanics. Our findings confirm and validate community observations and concerns of a disproportionate burden of COVID-19 among South Asian and Chinese Americans. Furthermore, our findings underscore the critical need for disaggregating Asian ethnic groups in data collection and reporting in order to appropriately allocate resources to the hardest hit communities, including testing and communication regarding seeking care as well as public health policies to mitigate risk factors and improve health equity.

## Data Availability

Data can be made available upon request.

