## Supplement Table 1 for "Disaggregating Asian Race Reveals COVID-19 Disparities among Asian Americans at New York City’s Public Hospital System"

**Supplemental Table 1. Surname List for Identifying Asian Race and Ethnic Groups**

| **Surname** | **Ethnicity** | **Subgroup** | **Source^1,2,3^** |
| --- | --- | --- | --- |
| A | Vietnamese | Other Asian | Wong et al. |
| Aala |  | South Asian | Lim et al. |
| Aasef |  | South Asian | Lim et al. |
| Aba | Filipino | Other Asian | Wong et al. |
| Ababa | Filipino | Other Asian | Wong et al. |
| Ababan | Filipino | Other Asian | Wong et al. |
| Ababon | Filipino | Other Asian | Wong et al. |
| Abac | Filipino | Other Asian | Wong et al. |
| Abaca | Filipino | Other Asian | Wong et al. |
| Abacan | Filipino | Other Asian | Wong et al. |
| Abad | Filipino | Other Asian | Wong et al. |
| Abada | Filipino | Other Asian | Wong et al. |
| Abadam | Filipino | Other Asian | Wong et al. |
| Abadeza | Filipino | Other Asian | Wong et al. |
| Abadi |  | South Asian | Lim et al. |
| Abadia | Filipino | Other Asian | Wong et al. |
| Abadiano | Filipino | Other Asian | Wong et al. |
| Abadilla | Filipino | Other Asian | Wong et al. |
| Abadines | Filipino | Other Asian | Wong et al. |
| Abadsantos | Filipino | Other Asian | Wong et al. |
| Abaga | Filipino | Other Asian | Wong et al. |
| Abagat | Filipino | Other Asian | Wong et al. |
| Abaigar | Filipino | Other Asian | Wong et al. |
| Abalahin | Filipino | Other Asian | Wong et al. |
| Aballe | Filipino | Other Asian | Wong et al. |
| Abalo | Filipino | Other Asian | Wong et al. |
| Abalos | Filipino | Other Asian | Wong et al. |
| Abamonga | Filipino | Other Asian | Wong et al. |
| Aban | Filipino | Other Asian | Wong et al. |
| Abanador | Filipino | Other Asian | Wong et al. |
| Abando | Filipino | Other Asian | Wong et al. |
| Abanes | Filipino | Other Asian | Wong et al. |
| Abang | Filipino | Other Asian | Wong et al. |
| Abangan | Filipino | Other Asian | Wong et al. |
| Abania | Filipino | Other Asian | Wong et al. |
| Abanilla | Filipino | Other Asian | Wong et al. |
| Abano | Filipino | Other Asian | Wong et al. |
| Abante | Filipino | Other Asian | Wong et al. |
| Abanto | Filipino | Other Asian | Wong et al. |
| Abao | Filipino | Other Asian | Wong et al. |
| Abaoag | Filipino | Other Asian | Wong et al. |
| Abapo | Filipino | Other Asian | Wong et al. |
| Abaquin | Filipino | Other Asian | Wong et al. |
| Abaquita | Filipino | Other Asian | Wong et al. |
| Abar | Filipino | Other Asian | Wong et al. |
| Abara | Filipino | Other Asian | Wong et al. |
| Abarabar | Filipino | Other Asian | Wong et al. |
| Abarca | Filipino | Other Asian | Wong et al. |
| Abarcar | Filipino | Other Asian | Wong et al. |
| Abarico | Filipino | Other Asian | Wong et al. |
| Abarientos | Filipino | Other Asian | Wong et al. |
| Abarintos | Filipino | Other Asian | Wong et al. |
| Abarquez | Filipino | Other Asian | Wong et al. |
| Abarra | Filipino | Other Asian | Wong et al. |
| Abarro | Filipino | Other Asian | Wong et al. |
| Abary | Filipino | Other Asian | Wong et al. |
| Abas | Filipino | Other Asian | Wong et al. |
| Abasolo | Filipino | Other Asian | Wong et al. |
| Abass |  | South Asian | Lim et al. |
| Abasta | Filipino | Other Asian | Wong et al. |
| Abastas | Filipino | Other Asian | Wong et al. |
| Abastillas | Filipino | Other Asian | Wong et al. |
| Abat | Filipino | Other Asian | Wong et al. |
| Abatayo | Filipino | Other Asian | Wong et al. |
| Abay | Filipino | Other Asian | Wong et al. |
| Abaya | Filipino | Other Asian | Wong et al. |
| Abayan | Filipino | Other Asian | Wong et al. |
| Abayon | Filipino | Other Asian | Wong et al. |
| Abbas |  | South Asian | Lim et al. |
| Abbi | Asian Indian | South Asian | Wong et al. |
| Abcede | Filipino | Other Asian | Wong et al. |
| Abdelrahman | Pakistani | South Asian | Kranick et al. |
| Abdin |  | South Asian | Lim et al. |
| Abdon | Filipino | Other Asian | Wong et al. |
| Abdou | Pakistani | South Asian | Kranick et al. |
| Abdul | Asian Indian | South Asian | Wong et al. |
| Abdulkhaleque |  | South Asian | Lim et al. |
| Abdulla | Asian Indian | South Asian | Wong et al. |
| Abdullah |  | South Asian | Lim et al. |
| Abe | Japanese | Other Asian | Wong et al. |
| Abear | Filipino | Other Asian | Wong et al. |
| Abecia | Filipino | Other Asian | Wong et al. |
| Abedes | Filipino | Other Asian | Wong et al. |
| Abedin | Bangladeshi | South Asian | Kranick et al. |
| Abejar | Filipino | Other Asian | Wong et al. |
| Abejero | Filipino | Other Asian | Wong et al. |
| Abejo | Filipino | Other Asian | Wong et al. |
| Abejuela | Filipino | Other Asian | Wong et al. |
| Abejuro | Filipino | Other Asian | Wong et al. |
| Abelardo | Filipino | Other Asian | Wong et al. |
| Abeleda | Filipino | Other Asian | Wong et al. |
| Abelgas | Filipino | Other Asian | Wong et al. |
| Abella | Filipino | Other Asian | Wong et al. |
| Abellada | Filipino | Other Asian | Wong et al. |
| Abellana | Filipino | Other Asian | Wong et al. |
| Abellanosa | Filipino | Other Asian | Wong et al. |
| Abellar | Filipino | Other Asian | Wong et al. |
| Abellera | Filipino | Other Asian | Wong et al. |
| Abello | Filipino | Other Asian | Wong et al. |
| Abellon | Filipino | Other Asian | Wong et al. |
| Abelon | Filipino | Other Asian | Wong et al. |
| Aben | Filipino | Other Asian | Wong et al. |
| Abendano | Filipino | Other Asian | Wong et al. |
| Abenes | Filipino | Other Asian | Wong et al. |
| Abenoja | Filipino | Other Asian | Wong et al. |
| Abenojar | Filipino | Other Asian | Wong et al. |
| Abergas | Filipino | Other Asian | Wong et al. |
| Aberin | Filipino | Other Asian | Wong et al. |
| Aberion | Filipino | Other Asian | Wong et al. |
| Abero | Filipino | Other Asian | Wong et al. |
| Abes | Filipino | Other Asian | Wong et al. |
| Abesamis | Filipino | Other Asian | Wong et al. |
| Abeto | Filipino | Other Asian | Wong et al. |
| Abia | Filipino | Other Asian | Wong et al. |
| Abiad | Filipino | Other Asian | Wong et al. |
| Abian | Filipino | Other Asian | Wong et al. |
| Abiang | Filipino | Other Asian | Wong et al. |
| Abidi | Asian Indian | South Asian | Wong et al. |
| Abidog | Filipino | Other Asian | Wong et al. |
| Abiera | Filipino | Other Asian | Wong et al. |
| Abigania | Filipino | Other Asian | Wong et al. |
| Abijay | Filipino | Other Asian | Wong et al. |
| Abiko | Japanese | Other Asian | Wong et al. |
| Abila | Filipino | Other Asian | Wong et al. |
| Abilar | Filipino | Other Asian | Wong et al. |
| Abilay | Filipino | Other Asian | Wong et al. |
| Abilla | Filipino | Other Asian | Wong et al. |
| Abille | Filipino | Other Asian | Wong et al. |
| Abilo | Filipino | Other Asian | Wong et al. |
| Abinales | Filipino | Other Asian | Wong et al. |
| Abines | Filipino | Other Asian | Wong et al. |
| Abing | Filipino | Other Asian | Wong et al. |
| Abinoja | Filipino | Other Asian | Wong et al. |
| Abinsay | Filipino | Other Asian | Wong et al. |
| Abiog | Filipino | Other Asian | Wong et al. |
| Abiru | Japanese | Other Asian | Wong et al. |
| Abis | Filipino | Other Asian | Wong et al. |
| Abiva | Filipino | Other Asian | Wong et al. |
| Ablan | Filipino | Other Asian | Wong et al. |
| Ablang | Filipino | Other Asian | Wong et al. |
| Ablao | Filipino | Other Asian | Wong et al. |
| Ablaza | Filipino | Other Asian | Wong et al. |
| Ablen | Filipino | Other Asian | Wong et al. |
| Ablian | Filipino | Other Asian | Wong et al. |
| Ablir | Filipino | Other Asian | Wong et al. |
| Ablog | Filipino | Other Asian | Wong et al. |
| Ablola | Filipino | Other Asian | Wong et al. |
| Ablong | Filipino | Other Asian | Wong et al. |
| Abo | Filipino | Other Asian | Wong et al. |
| Abobo | Filipino | Other Asian | Wong et al. |
| Abogadie | Filipino | Other Asian | Wong et al. |
| Abolencia | Filipino | Other Asian | Wong et al. |
| Abon | Filipino | Other Asian | Wong et al. |
| Abonales | Filipino | Other Asian | Wong et al. |
| Abong | Filipino | Other Asian | Wong et al. |
| Abordo | Filipino | Other Asian | Wong et al. |
| Aboy | Filipino | Other Asian | Wong et al. |
| Aboyme | Filipino | Other Asian | Wong et al. |
| Abracosa | Filipino | Other Asian | Wong et al. |
| Abraham | Bangladeshi | South Asian | Kranick et al. |
| Abrahan | Filipino | Other Asian | Wong et al. |
| Abrajano | Filipino | Other Asian | Wong et al. |
| Abrantes | Filipino | Other Asian | Wong et al. |
| Abrazado | Filipino | Other Asian | Wong et al. |
| Abrea | Filipino | Other Asian | Wong et al. |
| Abrecea | Filipino | Other Asian | Wong et al. |
| Abrena | Filipino | Other Asian | Wong et al. |
| Abrenica | Filipino | Other Asian | Wong et al. |
| Abrenilla | Filipino | Other Asian | Wong et al. |
| Abrenio | Filipino | Other Asian | Wong et al. |
| Abrera | Filipino | Other Asian | Wong et al. |
| Abreu | Filipino | Other Asian | Wong et al. |
| Abriam | Filipino | Other Asian | Wong et al. |
| Abrigo | Filipino | Other Asian | Wong et al. |
| Abril | Filipino | Other Asian | Wong et al. |
| Abrina | Filipino | Other Asian | Wong et al. |
| Abriol | Filipino | Other Asian | Wong et al. |
| Abro |  | South Asian | Lim et al. |
| Abrogar | Filipino | Other Asian | Wong et al. |
| Abrogena | Filipino | Other Asian | Wong et al. |
| Abrol | Asian Indian | South Asian | Wong et al. |
| Absalon | Filipino | Other Asian | Wong et al. |
| Absolor | Filipino | Other Asian | Wong et al. |
| Abua | Filipino | Other Asian | Wong et al. |
| Abuan | Filipino | Other Asian | Wong et al. |
| Abubo | Filipino | Other Asian | Wong et al. |
| Abucay | Filipino | Other Asian | Wong et al. |
| Abucejo | Filipino | Other Asian | Wong et al. |
| Abuda | Filipino | Other Asian | Wong et al. |
| Abude | Filipino | Other Asian | Wong et al. |
| Abueg | Filipino | Other Asian | Wong et al. |
| Abuel | Filipino | Other Asian | Wong et al. |
| Abueva | Filipino | Other Asian | Wong et al. |
| Abulencia | Filipino | Other Asian | Wong et al. |
| Abundo | Filipino | Other Asian | Wong et al. |
| Abungan | Filipino | Other Asian | Wong et al. |
| Abustan | Filipino | Other Asian | Wong et al. |
| Abut | Filipino | Other Asian | Wong et al. |
| Abutal | Filipino | Other Asian | Wong et al. |
| Abutaleb |  | South Asian | Lim et al. |
| Abutin | Filipino | Other Asian | Wong et al. |
| Abuton | Filipino | Other Asian | Wong et al. |
| Abuyan | Filipino | Other Asian | Wong et al. |
| Abuyen | Filipino | Other Asian | Wong et al. |
| Abuyo | Filipino | Other Asian | Wong et al. |
| Abuyuan | Filipino | Other Asian | Wong et al. |
| Acab | Filipino | Other Asian | Wong et al. |
| Acacio | Filipino | Other Asian | Wong et al. |
| Academia | Filipino | Other Asian | Wong et al. |
| Acain | Filipino | Other Asian | Wong et al. |
| Acal | Filipino | Other Asian | Wong et al. |
| Acampado | Filipino | Other Asian | Wong et al. |
| Acantilado | Filipino | Other Asian | Wong et al. |
| Acapulco | Filipino | Other Asian | Wong et al. |
| Acasio | Filipino | Other Asian | Wong et al. |
| Acay | Filipino | Other Asian | Wong et al. |
| Acayan | Filipino | Other Asian | Wong et al. |
| Acaylar | Filipino | Other Asian | Wong et al. |
| Acda | Filipino | Other Asian | Wong et al. |
| Acdal | Filipino | Other Asian | Wong et al. |
| Acdan | Filipino | Other Asian | Wong et al. |
| Acebedo | Filipino | Other Asian | Wong et al. |
| Acebo | Filipino | Other Asian | Wong et al. |
| Acedera | Filipino | Other Asian | Wong et al. |
| Acedillo | Filipino | Other Asian | Wong et al. |
| Acedo | Filipino | Other Asian | Wong et al. |
| Acejo | Filipino | Other Asian | Wong et al. |
| Acena | Filipino | Other Asian | Wong et al. |
| Acenas | Filipino | Other Asian | Wong et al. |
| Acera | Filipino | Other Asian | Wong et al. |
| Aceret | Filipino | Other Asian | Wong et al. |
| Acero | Filipino | Other Asian | Wong et al. |
| Aceron | Filipino | Other Asian | Wong et al. |
| Acevedo | Filipino | Other Asian | Wong et al. |
| Acha | Filipino | Other Asian | Wong et al. |
| Achacoso | Filipino | Other Asian | Wong et al. |
| Achang | Chinese | Chinese | Wong et al. |
| Achanzar | Filipino | Other Asian | Wong et al. |
| Achar | Asian Indian | South Asian | Wong et al. |
| Acharjee |  | South Asian | Lim et al. |
| Acharya | Asian Indian | South Asian | Wong et al. |
| Achas | Filipino | Other Asian | Wong et al. |
| Achurra | Filipino | Other Asian | Wong et al. |
| Achuthan | Asian Indian | South Asian | Wong et al. |
| Acidera | Filipino | Other Asian | Wong et al. |
| Acido | Filipino | Other Asian | Wong et al. |
| Acierto | Filipino | Other Asian | Wong et al. |
| Acio | Filipino | Other Asian | Wong et al. |
| Aclan | Filipino | Other Asian | Wong et al. |
| Aclaro | Filipino | Other Asian | Wong et al. |
| Acleema |  | South Asian | Lim et al. |
| Acob | Filipino | Other Asian | Wong et al. |
| Acoba | Filipino | Other Asian | Wong et al. |
| Acogido | Filipino | Other Asian | Wong et al. |
| Acojido | Filipino | Other Asian | Wong et al. |
| Acol | Filipino | Other Asian | Wong et al. |
| Acompanado | Filipino | Other Asian | Wong et al. |
| Acopiado | Filipino | Other Asian | Wong et al. |
| Acopio | Filipino | Other Asian | Wong et al. |
| Acorda | Filipino | Other Asian | Wong et al. |
| Acosta | Filipino | Other Asian | Kranick et al. |
| Acoymo | Filipino | Other Asian | Wong et al. |
| Acuario | Filipino | Other Asian | Wong et al. |
| Acuavera | Filipino | Other Asian | Wong et al. |
| Acuesta | Filipino | Other Asian | Wong et al. |
| Acuin | Filipino | Other Asian | Wong et al. |
| Acuna | Filipino | Other Asian | Wong et al. |
| Acupan | Filipino | Other Asian | Wong et al. |
| Acupido | Filipino | Other Asian | Wong et al. |
| Acuzar | Filipino | Other Asian | Wong et al. |
| Aczon | Filipino | Other Asian | Wong et al. |
| Ada | Filipino | Other Asian | Wong et al. |
| Adachi | Japanese | Other Asian | Wong et al. |
| Adad | Filipino | Other Asian | Wong et al. |
| Adajar | Filipino | Other Asian | Wong et al. |
| Adalid | Filipino | Other Asian | Wong et al. |
| Adalla | Filipino | Other Asian | Wong et al. |
| Adame | Filipino | Other Asian | Wong et al. |
| Adamjee | Asian Indian | South Asian | Wong et al. |
| Adamos | Filipino | Other Asian | Wong et al. |
| Adan | Filipino | Other Asian | Wong et al. |
| Adaniya | Japanese | Other Asian | Wong et al. |
| Adante | Filipino | Other Asian | Wong et al. |
| Adao | Filipino | Other Asian | Wong et al. |
| Adaoag | Filipino | Other Asian | Wong et al. |
| Adaon | Filipino | Other Asian | Wong et al. |
| Adap | Filipino | Other Asian | Wong et al. |
| Adapon | Filipino | Other Asian | Wong et al. |
| Adarkar | Asian Indian | South Asian | Wong et al. |
| Adarlo | Filipino | Other Asian | Wong et al. |
| Adarme | Filipino | Other Asian | Wong et al. |
| Adarna | Filipino | Other Asian | Wong et al. |
| Adarne | Filipino | Other Asian | Wong et al. |
| Adatia | Asian Indian | South Asian | Wong et al. |
| Adaya | Filipino | Other Asian | Wong et al. |
| Adaza | Filipino | Other Asian | Wong et al. |
| Adea | Filipino | Other Asian | Wong et al. |
| Adecer | Filipino | Other Asian | Wong et al. |
| Adefuin | Filipino | Other Asian | Wong et al. |
| Adelkarim | Asian Indian | South Asian | Kranick et al. |
| Adeva | Filipino | Other Asian | Wong et al. |
| Adevoso | Filipino | Other Asian | Wong et al. |
| Adhami | Asian Indian | South Asian | Wong et al. |
| Adhar |  | South Asian | Lim et al. |
| Adhikari | Asian Indian | South Asian | Wong et al. |
| Adhikary |  | South Asian | Lim et al. |
| Adhvaryu | Asian Indian | South Asian | Wong et al. |
| Adi | Filipino | Other Asian | Wong et al. |
| Adia | Filipino | Other Asian | Wong et al. |
| Adiarte | Filipino | Other Asian | Wong et al. |
| Adina | Filipino | Other Asian | Wong et al. |
| Adinig | Filipino | Other Asian | Wong et al. |
| Adion | Filipino | Other Asian | Wong et al. |
| Aditya | Asian Indian | South Asian | Wong et al. |
| Adlao | Filipino | Other Asian | Wong et al. |
| Adlaon | Filipino | Other Asian | Wong et al. |
| Adlawan | Filipino | Other Asian | Wong et al. |
| Admana | Filipino | Other Asian | Wong et al. |
| Ado | Filipino | Other Asian | Wong et al. |
| Adolfo | Filipino | Other Asian | Wong et al. |
| Adona | Filipino | Other Asian | Wong et al. |
| Adonay | Filipino | Other Asian | Wong et al. |
| Adonis | Filipino | Other Asian | Wong et al. |
| Adora | Filipino | Other Asian | Wong et al. |
| Adorable | Filipino | Other Asian | Wong et al. |
| Adorador | Filipino | Other Asian | Wong et al. |
| Adorna | Filipino | Other Asian | Wong et al. |
| Adrales | Filipino | Other Asian | Wong et al. |
| Adre | Filipino | Other Asian | Wong et al. |
| Adres | Filipino | Other Asian | Wong et al. |
| Adriano | Filipino | Other Asian | Wong et al. |
| Adrias | Filipino | Other Asian | Wong et al. |
| Adriatico | Filipino | Other Asian | Wong et al. |
| Adrid | Filipino | Other Asian | Wong et al. |
| Adsuara | Filipino | Other Asian | Wong et al. |
| Aduan | Filipino | Other Asian | Wong et al. |
| Aduca | Filipino | Other Asian | Wong et al. |
| Aduna | Filipino | Other Asian | Wong et al. |
| Adusumilli | Asian Indian | South Asian | Wong et al. |
| Advento | Filipino | Other Asian | Wong et al. |
| Adviento | Filipino | Other Asian | Wong et al. |
| Advincula | Filipino | Other Asian | Kranick et al. |
| Adya | Asian Indian | South Asian | Kranick et al. |
| Adzuara | Filipino | Other Asian | Wong et al. |
| Ae | Korean | Other Asian | Wong et al. |
| Aesquivel | Filipino | Other Asian | Wong et al. |
| Aeyasha |  | South Asian | Lim et al. |
| Afable | Filipino | Other Asian | Wong et al. |
| Afaga | Filipino | Other Asian | Wong et al. |
| Afalla | Filipino | Other Asian | Wong et al. |
| Afan | Filipino | Other Asian | Wong et al. |
| Afenir | Filipino | Other Asian | Wong et al. |
| Aficial | Filipino | Other Asian | Wong et al. |
| Africa | Filipino | Other Asian | Wong et al. |
| Africano | Filipino | Other Asian | Wong et al. |
| Afridi | Pakistani | South Asian | Kranick et al. |
| Afroilan | Filipino | Other Asian | Wong et al. |
| Afroz |  | South Asian | Lim et al. |
| Afroze | Bangladeshi | South Asian | Kranick et al. |
| Afsar |  | South Asian | Lim et al. |
| Aftabuddin |  | South Asian | Lim et al. |
| Afurong | Filipino | Other Asian | Wong et al. |
| Afzal | Pakistani | South Asian | Kranick et al. |
| Agabao | Filipino | Other Asian | Wong et al. |
| Agabon | Filipino | Other Asian | Wong et al. |
| Agacid | Filipino | Other Asian | Wong et al. |
| Agacite | Filipino | Other Asian | Wong et al. |
| Agad | Filipino | Other Asian | Wong et al. |
| Agag | Filipino | Other Asian | Wong et al. |
| Agagas | Filipino | Other Asian | Wong et al. |
| Agahan | Filipino | Other Asian | Wong et al. |
| Agaloos | Filipino | Other Asian | Wong et al. |
| Agamao | Filipino | Other Asian | Wong et al. |
| Agamata | Filipino | Other Asian | Wong et al. |
| Agana | Filipino | Other Asian | Wong et al. |
| Aganad | Filipino | Other Asian | Wong et al. |
| Aganon | Filipino | Other Asian | Wong et al. |
| Agapay | Filipino | Other Asian | Wong et al. |
| Agapito | Filipino | Other Asian | Wong et al. |
| Agar | Asian Indian | South Asian | Kranick et al. |
| Agaran | Filipino | Other Asian | Wong et al. |
| Agarano | Filipino | Other Asian | Wong et al. |
| Agarie | Japanese | Other Asian | Wong et al. |
| Agarpao | Filipino | Other Asian | Wong et al. |
| Agarwal | Asian Indian | South Asian | Kranick et al. |
| Agarwala | Asian Indian | South Asian | Wong et al. |
| Agas | Filipino | Other Asian | Wong et al. |
| Agatep | Filipino | Other Asian | Wong et al. |
| Agaton | Filipino | Other Asian | Wong et al. |
| Agawa | Japanese | Other Asian | Wong et al. |
| Agay | Filipino | Other Asian | Wong et al. |
| Agbada | Filipino | Other Asian | Wong et al. |
| Agbanlog | Filipino | Other Asian | Wong et al. |
| Agbay | Filipino | Other Asian | Wong et al. |
| Agbayani | Filipino | Other Asian | Wong et al. |
| Agbisit | Filipino | Other Asian | Wong et al. |
| Agbulos | Filipino | Other Asian | Wong et al. |
| Agbunag | Filipino | Other Asian | Wong et al. |
| Agbuya | Filipino | Other Asian | Wong et al. |
| Agcanas | Filipino | Other Asian | Wong et al. |
| Agcaoili | Filipino | Other Asian | Wong et al. |
| Agda | Filipino | Other Asian | Wong et al. |
| Agdamag | Filipino | Other Asian | Wong et al. |
| Agdeppa | Filipino | Other Asian | Wong et al. |
| Agdinaoay | Filipino | Other Asian | Wong et al. |
| Agdipa | Filipino | Other Asian | Wong et al. |
| Agduyeng | Filipino | Other Asian | Wong et al. |
| Agellon | Filipino | Other Asian | Wong et al. |
| Agena | Japanese | Other Asian | Wong et al. |
| Aggabao | Filipino | Other Asian | Wong et al. |
| Aggarwal | Asian Indian | South Asian | Wong et al. |
| Aggarwala | Asian Indian | South Asian | Wong et al. |
| Aggasid | Filipino | Other Asian | Wong et al. |
| Aghera | Asian Indian | South Asian | Wong et al. |
| Agi | Chinese | Chinese | Wong et al. |
| Agito | Filipino | Other Asian | Wong et al. |
| Agliam | Filipino | Other Asian | Wong et al. |
| Aglibot | Filipino | Other Asian | Wong et al. |
| Aglibut | Filipino | Other Asian | Wong et al. |
| Aglipay | Filipino | Other Asian | Wong et al. |
| Agloro | Filipino | Other Asian | Wong et al. |
| Agluba | Filipino | Other Asian | Wong et al. |
| Aglubat | Filipino | Other Asian | Wong et al. |
| Aglugub | Filipino | Other Asian | Wong et al. |
| Aglupos | Filipino | Other Asian | Wong et al. |
| Agmata | Filipino | Other Asian | Wong et al. |
| Agnes | Filipino | Other Asian | Wong et al. |
| Agngarayngay | Filipino | Other Asian | Wong et al. |
| Agni | Filipino | Other Asian | Wong et al. |
| Agnihotri | Asian Indian | South Asian | Wong et al. |
| Agnir | Filipino | Other Asian | Wong et al. |
| Agnis | Filipino | Other Asian | Wong et al. |
| Agno | Filipino | Other Asian | Wong et al. |
| Ago | Filipino | Other Asian | Wong et al. |
| Agodon | Filipino | Other Asian | Wong et al. |
| Agojo | Filipino | Other Asian | Wong et al. |
| Agoncillo | Filipino | Other Asian | Wong et al. |
| Agonias | Filipino | Other Asian | Wong et al. |
| Agonoy | Filipino | Other Asian | Wong et al. |
| Agoo | Filipino | Other Asian | Wong et al. |
| Agor | Filipino | Other Asian | Wong et al. |
| Agorilla | Filipino | Other Asian | Wong et al. |
| Agot | Filipino | Other Asian | Wong et al. |
| Agoto | Filipino | Other Asian | Wong et al. |
| Agoy | Filipino | Other Asian | Wong et al. |
| Agpalasin | Filipino | Other Asian | Wong et al. |
| Agpalo | Filipino | Other Asian | Wong et al. |
| Agpalza | Filipino | Other Asian | Wong et al. |
| Agpaoa | Filipino | Other Asian | Wong et al. |
| Agpawa | Filipino | Other Asian | Wong et al. |
| Agpoon | Filipino | Other Asian | Wong et al. |
| Agra | Filipino | Other Asian | Wong et al. |
| Agraan | Filipino | Other Asian | Wong et al. |
| Agravante | Filipino | Other Asian | Wong et al. |
| Agraviador | Filipino | Other Asian | Wong et al. |
| Agrawal | Asian Indian | South Asian | Wong et al. |
| Agrawala | Asian Indian | South Asian | Wong et al. |
| Agreda | Filipino | Other Asian | Wong et al. |
| Agregado | Filipino | Other Asian | Wong et al. |
| Agres | Filipino | Other Asian | Wong et al. |
| Agresor | Filipino | Other Asian | Wong et al. |
| Agriam | Filipino | Other Asian | Wong et al. |
| Agringo | Filipino | Other Asian | Wong et al. |
| Agron | Filipino | Other Asian | Wong et al. |
| Agsalda | Filipino | Other Asian | Wong et al. |
| Agsalog | Filipino | Other Asian | Wong et al. |
| Agsalud | Filipino | Other Asian | Wong et al. |
| Agsaulio | Filipino | Other Asian | Wong et al. |
| Agsunod | Filipino | Other Asian | Wong et al. |
| Agtang | Filipino | Other Asian | Wong et al. |
| Agtarap | Filipino | Other Asian | Wong et al. |
| Agtual | Filipino | Other Asian | Wong et al. |
| Agtuca | Filipino | Other Asian | Wong et al. |
| Aguada | Filipino | Other Asian | Wong et al. |
| Aguado | Filipino | Other Asian | Wong et al. |
| Aguas | Filipino | Other Asian | Wong et al. |
| Aguda | Filipino | Other Asian | Wong et al. |
| Agudelo | Filipino | Other Asian | Wong et al. |
| Agudo | Filipino | Other Asian | Wong et al. |
| Agudong | Filipino | Other Asian | Wong et al. |
| Aguhar | Filipino | Other Asian | Kranick et al. |
| Agui | Filipino | Other Asian | Wong et al. |
| Aguil | Filipino | Other Asian | Wong et al. |
| Aguila | Filipino | Other Asian | Wong et al. |
| Aguilan | Filipino | Other Asian | Wong et al. |
| Aguilar | Filipino | Other Asian | Wong et al. |
| Aguilera | Filipino | Other Asian | Wong et al. |
| Aguiling | Filipino | Other Asian | Kranick et al. |
| Aguillon | Filipino | Other Asian | Wong et al. |
| Aguilo | Filipino | Other Asian | Wong et al. |
| Aguilon | Filipino | Other Asian | Wong et al. |
| Aguilos | Filipino | Other Asian | Wong et al. |
| Aguilucho | Filipino | Other Asian | Wong et al. |
| Aguiluz | Filipino | Other Asian | Wong et al. |
| Aguimatang | Filipino | Other Asian | Wong et al. |
| Aguinaldo | Filipino | Other Asian | Kranick et al. |
| Aguino | Filipino | Other Asian | Wong et al. |
| Aguipo | Filipino | Other Asian | Wong et al. |
| Aguiran | Filipino | Other Asian | Wong et al. |
| Aguirre | Filipino | Other Asian | Wong et al. |
| Aguisanda | Filipino | Other Asian | Wong et al. |
| Aguja | Filipino | Other Asian | Wong et al. |
| Agujar | Filipino | Other Asian | Wong et al. |
| Agulan | Filipino | Other Asian | Wong et al. |
| Agulay | Filipino | Other Asian | Wong et al. |
| Agullana | Filipino | Other Asian | Wong et al. |
| Agulto | Filipino | Other Asian | Wong et al. |
| Agunoy | Filipino | Other Asian | Wong et al. |
| Agus | Filipino | Other Asian | Wong et al. |
| Agustin | Filipino | Other Asian | Wong et al. |
| Agustines | Filipino | Other Asian | Wong et al. |
| Agusto | Filipino | Other Asian | Wong et al. |
| Aguto | Filipino | Other Asian | Wong et al. |
| Ah | Chinese | Chinese | Wong et al. |
| Ahad | Asian Indian | South Asian | Wong et al. |
| Ahamad | Asian Indian | South Asian | Wong et al. |
| Ahamed |  | South Asian | Lim et al. |
| Ahammad | Asian Indian | South Asian | Wong et al. |
| Ahammed |  | South Asian | Lim et al. |
| Ahasan |  | South Asian | Lim et al. |
| Aherrera | Filipino | Other Asian | Wong et al. |
| Ahluwalia | Asian Indian | South Asian | Wong et al. |
| Ahmad | Pakistani; Asian Indian; Bangladeshi | South Asian | Kranick et al. |
| Ahmar |  | South Asian | Lim et al. |
| Ahmed | Pakistani; Asian Indian; Bangladeshi | South Asian | Kranick et al. |
| Ahmedullah | Asian Indian | South Asian | Wong et al. |
| Ahmmed |  | South Asian | Lim et al. |
| Ahn | Korean | Other Asian | Kranick et al. |
| Ahomed |  | South Asian | Lim et al. |
| Ahorro | Filipino | Other Asian | Wong et al. |
| Ahsab |  | South Asian | Lim et al. |
| Ahsan | Bangladeshi; Asian Indian | South Asian | Kranick et al. |
| Ahsanuddin | Asian Indian | South Asian | Wong et al. |
| Ahuja |  | South Asian | Lim et al. |
| Ahulwalia | Asian Indian | South Asian | Kranick et al. |
| Ai | Chinese | Chinese | Wong et al. |
| Aiba | Japanese | Other Asian | Wong et al. |
| Aich |  | South Asian | Lim et al. |
| Aida | Japanese | Other Asian | Wong et al. |
| Aihara | Japanese | Other Asian | Wong et al. |
| Aikawa | Japanese | Other Asian | Wong et al. |
| Aiko | Japanese | Other Asian | Wong et al. |
| Aisporna | Filipino | Other Asian | Wong et al. |
| Aiyar | Asian Indian | South Asian | Wong et al. |
| Aiyer | Asian Indian | South Asian | Wong et al. |
| Aizawa | Japanese | Other Asian | Wong et al. |
| Ajbarjamil |  | South Asian | Lim et al. |
| Ajero | Filipino | Other Asian | Wong et al. |
| Ajeto | Filipino | Other Asian | Wong et al. |
| Ajima | Japanese | Other Asian | Wong et al. |
| Ajmera | Asian Indian | South Asian | Wong et al. |
| Ajolo | Filipino | Other Asian | Wong et al. |
| Ajoste | Filipino | Other Asian | Wong et al. |
| Aka | Japanese | Other Asian | Wong et al. |
| Akabane | Japanese | Other Asian | Wong et al. |
| Akabori | Japanese | Other Asian | Wong et al. |
| Akada | Japanese | Other Asian | Wong et al. |
| Akagawa | Japanese | Other Asian | Wong et al. |
| Akagi | Japanese | Other Asian | Wong et al. |
| Akahane | Japanese | Other Asian | Wong et al. |
| Akahori | Japanese | Other Asian | Wong et al. |
| Akahoshi | Japanese | Other Asian | Wong et al. |
| Akai | Japanese | Other Asian | Wong et al. |
| Akaike | Japanese | Other Asian | Wong et al. |
| Akaishi | Japanese | Other Asian | Wong et al. |
| Akalrai |  | South Asian | Lim et al. |
| Akama | Japanese | Other Asian | Wong et al. |
| Akamatsu | Japanese | Other Asian | Wong et al. |
| Akamine | Japanese | Other Asian | Wong et al. |
| Akamura | Japanese | Other Asian | Wong et al. |
| Akao | Japanese | Other Asian | Wong et al. |
| Akasaka | Japanese | Other Asian | Wong et al. |
| Akasaki | Japanese | Other Asian | Wong et al. |
| Akashi | Japanese | Other Asian | Wong et al. |
| Akatsu | Japanese | Other Asian | Wong et al. |
| Akatsuka | Japanese | Other Asian | Wong et al. |
| Akazawa | Japanese | Other Asian | Wong et al. |
| Akbani | Asian Indian | South Asian | Wong et al. |
| Akbarali | Asian Indian | South Asian | Wong et al. |
| Akbari | Asian Indian | South Asian | Wong et al. |
| Akber | Asian Indian | South Asian | Wong et al. |
| Akbor |  | South Asian | Lim et al. |
| Akella | Asian Indian | South Asian | Wong et al. |
| Akemoto | Japanese | Other Asian | Wong et al. |
| Akhtar | Asian Indian; Pakistani | South Asian | Kranick et al. |
| Akhtary |  | South Asian | Lim et al. |
| Akhter | Asian Indian | South Asian | Wong et al. |
| Aki | Japanese | Other Asian | Wong et al. |
| Akiba | Japanese | Other Asian | Wong et al. |
| Akimoto | Japanese | Other Asian | Wong et al. |
| Akimura | Japanese | Other Asian | Wong et al. |
| Akinaga | Japanese | Other Asian | Wong et al. |
| Akino | Japanese | Other Asian | Wong et al. |
| Akira | Japanese | Other Asian | Wong et al. |
| Akita | Japanese | Other Asian | Wong et al. |
| Akiya | Japanese | Other Asian | Wong et al. |
| Akiyama | Japanese | Other Asian | Wong et al. |
| Akiyoshi | Japanese | Other Asian | Wong et al. |
| Akizawa | Japanese | Other Asian | Wong et al. |
| Akizuki | Japanese | Other Asian | Wong et al. |
| Akol | Filipino | Other Asian | Wong et al. |
| Akon |  | South Asian | Lim et al. |
| Akram | Pakistani | South Asian | Kranick et al. |
| Aktar |  | South Asian | Lim et al. |
| Akter | Bangladeshi | South Asian | Kranick et al. |
| Akthar |  | South Asian | Lim et al. |
| Akther |  | South Asian | Lim et al. |
| Akune | Japanese | Other Asian | Wong et al. |
| Akutagawa | Japanese | Other Asian | Wong et al. |
| Akutsu | Japanese | Other Asian | Wong et al. |
| Al |  | South Asian | Lim et al. |
| Ala | Filipino | Other Asian | Wong et al. |
| Alaan | Filipino | Other Asian | Wong et al. |
| Alaba | Filipino | Other Asian | Wong et al. |
| Alabab | Filipino | Other Asian | Wong et al. |
| Alabado | Filipino | Other Asian | Wong et al. |
| Alaban | Filipino | Other Asian | Wong et al. |
| Alabanza | Filipino | Other Asian | Wong et al. |
| Alabaso | Filipino | Other Asian | Wong et al. |
| Alabastro | Filipino | Other Asian | Wong et al. |
| Alabata | Filipino | Other Asian | Wong et al. |
| Alabe | Filipino | Other Asian | Wong et al. |
| Alacar | Filipino | Other Asian | Wong et al. |
| Aladin | Asian Indian | South Asian | Wong et al. |
| Alado | Filipino | Other Asian | Wong et al. |
| Alafriz | Filipino | Other Asian | Wong et al. |
| Alag | Filipino | Other Asian | Wong et al. |
| Alagao | Filipino | Other Asian | Wong et al. |
| Alagappan | Asian Indian | South Asian | Wong et al. |
| Alagar | Filipino | Other Asian | Wong et al. |
| Alagia | Asian Indian | South Asian | Wong et al. |
| Alagon | Filipino | Other Asian | Wong et al. |
| Alalay | Filipino | Other Asian | Wong et al. |
| Alam | Bangladeshi; Asian Indian | South Asian | Kranick et al. |
| Alamar | Filipino | Other Asian | Wong et al. |
| Alamares | Filipino | Other Asian | Wong et al. |
| Alambat | Filipino | Other Asian | Wong et al. |
| Alambra | Filipino | Other Asian | Wong et al. |
| Alamgir |  | South Asian | Lim et al. |
| Alamibinani |  | South Asian | Lim et al. |
| Alamillo | Filipino | Other Asian | Wong et al. |
| Alamo | Filipino | Other Asian | Wong et al. |
| Alamodin | Filipino | Other Asian | Wong et al. |
| Alandy | Filipino | Other Asian | Wong et al. |
| Alanes | Filipino | Other Asian | Wong et al. |
| Alanguilan | Filipino | Other Asian | Wong et al. |
| Alano | Filipino | Other Asian | Wong et al. |
| Alao | Filipino | Other Asian | Wong et al. |
| Alaras | Filipino | Other Asian | Wong et al. |
| Alarcio | Filipino | Other Asian | Wong et al. |
| Alarcon | Filipino | Other Asian | Wong et al. |
| Alarde | Filipino | Other Asian | Wong et al. |
| Alarva | Filipino | Other Asian | Wong et al. |
| Alas | Filipino | Other Asian | Wong et al. |
| Alauddin | Asian Indian | South Asian | Wong et al. |
| Alaura | Filipino | Other Asian | Wong et al. |
| Alava | Filipino | Other Asian | Wong et al. |
| Alavazo | Filipino | Other Asian | Wong et al. |
| Alavi | Asian Indian | South Asian | Wong et al. |
| Alay | Filipino | Other Asian | Wong et al. |
| Alayon | Filipino | Other Asian | Wong et al. |
| Alayvilla | Filipino | Other Asian | Wong et al. |
| Alba | Filipino | Other Asian | Wong et al. |
| Albaladejo | Filipino | Other Asian | Wong et al. |
| Albalos | Filipino | Other Asian | Wong et al. |
| Albana | Filipino | Other Asian | Wong et al. |
| Albania | Filipino | Other Asian | Wong et al. |
| Albaniel | Filipino | Other Asian | Wong et al. |
| Albano | Filipino | Other Asian | Kranick et al. |
| Albao | Filipino | Other Asian | Wong et al. |
| Albar | Filipino | Other Asian | Wong et al. |
| Albaran | Filipino | Other Asian | Wong et al. |
| Albarico | Filipino | Other Asian | Wong et al. |
| Albarillo | Filipino | Other Asian | Wong et al. |
| Albarracin | Filipino | Other Asian | Wong et al. |
| Albay | Filipino | Other Asian | Wong et al. |
| Albayalde | Filipino | Other Asian | Wong et al. |
| Albelda | Filipino | Other Asian | Wong et al. |
| Albert |  | South Asian | Lim et al. |
| Alberto | Filipino | Other Asian | Wong et al. |
| Albino | Filipino | Other Asian | Wong et al. |
| Albiso | Filipino | Other Asian | Wong et al. |
| Albito | Filipino | Other Asian | Wong et al. |
| Albornoz | Filipino | Other Asian | Wong et al. |
| Alburo | Filipino | Other Asian | Wong et al. |
| Alcaide | Filipino | Other Asian | Wong et al. |
| Alcairo | Filipino | Other Asian | Wong et al. |
| Alcala | Filipino | Other Asian | Wong et al. |
| Alcalde | Filipino | Other Asian | Wong et al. |
| Alcalen | Filipino | Other Asian | Wong et al. |
| Alcancia | Filipino | Other Asian | Wong et al. |
| Alcantara | Filipino | Other Asian | Wong et al. |
| Alcantra | Filipino | Other Asian | Wong et al. |
| Alcaparas | Filipino | Other Asian | Wong et al. |
| Alcaraz | Filipino | Other Asian | Wong et al. |
| Alcarion | Filipino | Other Asian | Wong et al. |
| Alcasid | Filipino | Other Asian | Wong et al. |
| Alcasio | Filipino | Other Asian | Wong et al. |
| Alcausin | Filipino | Other Asian | Wong et al. |
| Alcayaga | Filipino | Other Asian | Wong et al. |
| Alcayde | Filipino | Other Asian | Wong et al. |
| Alcazar | Filipino | Other Asian | Wong et al. |
| Alcazaren | Filipino | Other Asian | Wong et al. |
| Alcedo | Filipino | Other Asian | Wong et al. |
| Alcera | Filipino | Other Asian | Wong et al. |
| Alcid | Filipino | Other Asian | Wong et al. |
| Alcobendas | Filipino | Other Asian | Wong et al. |
| Alcober | Filipino | Other Asian | Wong et al. |
| Alcomendas | Filipino | Other Asian | Wong et al. |
| Alcon | Filipino | Other Asian | Wong et al. |
| Alconcel | Filipino | Other Asian | Wong et al. |
| Alcones | Filipino | Other Asian | Wong et al. |
| Alconis | Filipino | Other Asian | Wong et al. |
| Alcoran | Filipino | Other Asian | Wong et al. |
| Alcordo | Filipino | Other Asian | Wong et al. |
| Alcoriza | Filipino | Other Asian | Wong et al. |
| Alcos | Filipino | Other Asian | Wong et al. |
| Alcoseba | Filipino | Other Asian | Wong et al. |
| Alcova | Filipino | Other Asian | Wong et al. |
| Alcover | Filipino | Other Asian | Wong et al. |
| Alcoy | Filipino | Other Asian | Wong et al. |
| Alcuaz | Filipino | Other Asian | Wong et al. |
| Alcudia | Filipino | Other Asian | Wong et al. |
| Alda | Filipino | Other Asian | Wong et al. |
| Aldaba | Filipino | Other Asian | Wong et al. |
| Aldana | Filipino | Other Asian | Wong et al. |
| Aldas | Filipino | Other Asian | Wong et al. |
| Aldave | Filipino | Other Asian | Wong et al. |
| Alday | Filipino | Other Asian | Wong et al. |
| Alde | Filipino | Other Asian | Wong et al. |
| Aldea | Filipino | Other Asian | Wong et al. |
| Aldecoa | Filipino | Other Asian | Wong et al. |
| Aldeguer | Filipino | Other Asian | Wong et al. |
| Aldeza | Filipino | Other Asian | Wong et al. |
| Aldip | Filipino | Other Asian | Wong et al. |
| Aldover | Filipino | Other Asian | Wong et al. |
| Aldovino | Filipino | Other Asian | Wong et al. |
| Alducente | Filipino | Other Asian | Wong et al. |
| Ale | Asian Indian | South Asian | Wong et al. |
| Alea | Filipino | Other Asian | Wong et al. |
| Aledia | Filipino | Other Asian | Wong et al. |
| Aleem | Asian Indian | South Asian | Wong et al. |
| Alega | Filipino | Other Asian | Wong et al. |
| Alegado | Filipino | Other Asian | Wong et al. |
| Alegarbes | Filipino | Other Asian | Wong et al. |
| Alegrado | Filipino | Other Asian | Wong et al. |
| Alegre | Filipino | Other Asian | Wong et al. |
| Alegria | Filipino | Other Asian | Wong et al. |
| Alegro | Filipino | Other Asian | Wong et al. |
| Alejandre | Filipino | Other Asian | Wong et al. |
| Alejandria | Filipino | Other Asian | Wong et al. |
| Alejandrino | Filipino | Other Asian | Wong et al. |
| Alejandro | Filipino | Other Asian | Wong et al. |
| Alejar | Filipino | Other Asian | Wong et al. |
| Alejo | Filipino | Other Asian | Wong et al. |
| Alejos | Filipino | Other Asian | Wong et al. |
| Aleman | Filipino | Other Asian | Wong et al. |
| Alemania | Filipino | Other Asian | Wong et al. |
| Alerre | Filipino | Other Asian | Wong et al. |
| Alerta | Filipino | Other Asian | Wong et al. |
| Alesna | Filipino | Other Asian | Wong et al. |
| Aleta | Filipino | Other Asian | Wong et al. |
| Alfafara | Filipino | Other Asian | Wong et al. |
| Alfar | Filipino | Other Asian | Wong et al. |
| Alfaro | Filipino | Other Asian | Wong et al. |
| Alfeche | Filipino | Other Asian | Wong et al. |
| Alfelor | Filipino | Other Asian | Wong et al. |
| Alferez | Filipino | Other Asian | Wong et al. |
| Alferos | Filipino | Other Asian | Wong et al. |
| Alfiler | Filipino | Other Asian | Wong et al. |
| Alfon | Filipino | Other Asian | Wong et al. |
| Alfonso | Filipino | Other Asian | Wong et al. |
| Alfonzo | Filipino | Other Asian | Wong et al. |
| Alforque | Filipino | Other Asian | Wong et al. |
| Alforte | Filipino | Other Asian | Wong et al. |
| Algabre | Filipino | Other Asian | Wong et al. |
| Algas | Filipino | Other Asian | Wong et al. |
| Algenio | Filipino | Other Asian | Wong et al. |
| Algoso | Filipino | Other Asian | Wong et al. |
| Alhambra | Filipino | Other Asian | Wong et al. |
| Al-Hossain |  | South Asian | Lim et al. |
| Ali | Asian Indian; Bangladeshi | South Asian | Kranick et al. |
| Alia | Filipino | Other Asian | Wong et al. |
| Aliado | Filipino | Other Asian | Wong et al. |
| Alialy | Filipino | Other Asian | Wong et al. |
| Alianza | Filipino | Other Asian | Wong et al. |
| Alias | Filipino | Other Asian | Wong et al. |
| Aliazas | Filipino | Other Asian | Wong et al. |
| Alibangbang | Filipino | Other Asian | Wong et al. |
| Alibhai | Asian Indian | South Asian | Wong et al. |
| Alibin | Filipino | Other Asian | Wong et al. |
| Alibutod | Filipino | Other Asian | Wong et al. |
| Alicando | Filipino | Other Asian | Wong et al. |
| Alicante | Filipino | Other Asian | Wong et al. |
| Alicaya | Filipino | Other Asian | Wong et al. |
| Alicdan | Filipino | Other Asian | Wong et al. |
| Alido | Filipino | Other Asian | Wong et al. |
| Alidon | Filipino | Other Asian | Wong et al. |
| Alie | Asian Indian | South Asian | Wong et al. |
| Aliermo | Filipino | Other Asian | Wong et al. |
| Aliga | Filipino | Other Asian | Wong et al. |
| Aligada | Filipino | Other Asian | Wong et al. |
| Aligado | Filipino | Other Asian | Wong et al. |
| Aligaen | Filipino | Other Asian | Wong et al. |
| Aligato | Filipino | Other Asian | Wong et al. |
| Alignay | Filipino | Other Asian | Wong et al. |
| Alikhan | Asian Indian | South Asian | Wong et al. |
| Alikpala | Filipino | Other Asian | Wong et al. |
| Alilin | Filipino | Other Asian | Wong et al. |
| Alim | Bangladeshi | South Asian | Kranick et al. |
| Alimagno | Filipino | Other Asian | Wong et al. |
| Alimboyoguen | Filipino | Other Asian | Wong et al. |
| Alimohamed | Asian Indian | South Asian | Wong et al. |
| Alimohammed | Asian Indian | South Asian | Wong et al. |
| Alimorong | Filipino | Other Asian | Wong et al. |
| Alimuzzaman |  | South Asian | Lim et al. |
| Alina | Filipino | Other Asian | Wong et al. |
| Alindada | Filipino | Other Asian | Wong et al. |
| Alindayu | Filipino | Other Asian | Wong et al. |
| Alindogan | Filipino | Other Asian | Wong et al. |
| Alinea | Filipino | Other Asian | Wong et al. |
| Alingasa | Filipino | Other Asian | Wong et al. |
| Alino | Filipino | Other Asian | Wong et al. |
| Alinsod | Filipino | Other Asian | Wong et al. |
| Alip | Filipino | Other Asian | Wong et al. |
| Alipao | Filipino | Other Asian | Wong et al. |
| Alipio | Filipino | Other Asian | Wong et al. |
| Alishah | Asian Indian | South Asian | Wong et al. |
| Alisna | Filipino | Other Asian | Wong et al. |
| Alisuag | Filipino | Other Asian | Wong et al. |
| Alivia | Filipino | Other Asian | Wong et al. |
| Aliviado | Filipino | Other Asian | Wong et al. |
| Alivio | Filipino | Other Asian | Wong et al. |
| Aliwalas | Filipino | Other Asian | Wong et al. |
| Aliwarga | Uzbekistan | Other Asian | Kranick et al. |
| Alkuino | Filipino | Other Asian | Wong et al. |
| Alla | Asian Indian | South Asian | Wong et al. |
| Alladiin |  | South Asian | Lim et al. |
| Alladin | Asian Indian | South Asian | Wong et al. |
| Allado | Filipino | Other Asian | Wong et al. |
| Allah | Asian Indian | South Asian | Wong et al. |
| Allaham |  | South Asian | Lim et al. |
| Allam | Asian Indian | South Asian | Wong et al. |
| Allana | Asian Indian | South Asian | Wong et al. |
| Allanigue | Filipino | Other Asian | Wong et al. |
| Allarey | Filipino | Other Asian | Wong et al. |
| Allas | Filipino | Other Asian | Wong et al. |
| Allauddin | Asian Indian | South Asian | Wong et al. |
| Allego | Filipino | Other Asian | Wong et al. |
| Alli | Asian Indian | South Asian | Wong et al. |
| Allihosein |  | South Asian | Lim et al. |
| Allones | Filipino | Other Asian | Wong et al. |
| Ally | Asian Indian | South Asian | Wong et al. |
| Allyshaw |  | South Asian | Lim et al. |
| Almacen | Filipino | Other Asian | Wong et al. |
| Almachar | Filipino | Other Asian | Wong et al. |
| Almaden | Filipino | Other Asian | Wong et al. |
| Almadin | Filipino | Other Asian | Wong et al. |
| Almagro | Filipino | Other Asian | Wong et al. |
| Almalel | Filipino | Other Asian | Wong et al. |
| Almamun |  | South Asian | Lim et al. |
| Almanza | Filipino | Other Asian | Wong et al. |
| Almanzor | Filipino | Other Asian | Wong et al. |
| Almarez | Filipino | Other Asian | Wong et al. |
| Almaria | Filipino | Other Asian | Wong et al. |
| Almarines | Filipino | Other Asian | Wong et al. |
| Almario | Filipino | Other Asian | Wong et al. |
| Almase | Filipino | Other Asian | Wong et al. |
| Almaula | Asian Indian | South Asian | Wong et al. |
| Almazan | Filipino | Other Asian | Wong et al. |
| Almazar | Filipino | Other Asian | Wong et al. |
| Almeda | Filipino | Other Asian | Wong et al. |
| Almeida | Asian Indian | South Asian | Wong et al. |
| Almenario | Filipino | Other Asian | Wong et al. |
| Almendares | Filipino | Other Asian | Wong et al. |
| Almendra | Filipino | Other Asian | Wong et al. |
| Almendral | Filipino | Other Asian | Wong et al. |
| Almendrala | Filipino | Other Asian | Wong et al. |
| Almendras | Filipino | Other Asian | Wong et al. |
| Almeria | Filipino | Other Asian | Wong et al. |
| Almero | Filipino | Other Asian | Wong et al. |
| Almerol | Filipino | Other Asian | Wong et al. |
| Almeron | Filipino | Other Asian | Wong et al. |
| Alminar | Filipino | Other Asian | Wong et al. |
| Almiranez | Filipino | Other Asian | Wong et al. |
| Almirante | Filipino | Other Asian | Wong et al. |
| Almirez | Filipino | Other Asian | Wong et al. |
| Almirol | Filipino | Other Asian | Wong et al. |
| Almiron | Filipino | Other Asian | Wong et al. |
| Almodovar | Filipino | Other Asian | Wong et al. |
| Almoete | Filipino | Other Asian | Wong et al. |
| Almogela | Filipino | Other Asian | Wong et al. |
| Almoite | Filipino | Other Asian | Wong et al. |
| Almojera | Filipino | Other Asian | Wong et al. |
| Almojuela | Filipino | Other Asian | Wong et al. |
| Almoneda | Filipino | Other Asian | Wong et al. |
| Almonte | Filipino | Other Asian | Wong et al. |
| Almoradie | Filipino | Other Asian | Wong et al. |
| Almoro | Filipino | Other Asian | Wong et al. |
| Almuete | Filipino | Other Asian | Wong et al. |
| Alnas | Filipino | Other Asian | Wong et al. |
| Alo | Filipino | Other Asian | Wong et al. |
| Aloba | Filipino | Other Asian | Wong et al. |
| Alog | Filipino | Other Asian | Wong et al. |
| Alojado | Filipino | Other Asian | Wong et al. |
| Alojipan | Filipino | Other Asian | Wong et al. |
| Alombro | Filipino | Other Asian | Wong et al. |
| Alon | Filipino | Other Asian | Wong et al. |
| Alonso | Filipino | Other Asian | Wong et al. |
| Alonte | Filipino | Other Asian | Wong et al. |
| Alonzo | Filipino | Other Asian | Wong et al. |
| Aloot | Filipino | Other Asian | Wong et al. |
| Alop | Filipino | Other Asian | Wong et al. |
| Alorro | Filipino | Other Asian | Wong et al. |
| Alota | Filipino | Other Asian | Wong et al. |
| Alovera | Filipino | Other Asian | Wong et al. |
| Alpas | Filipino | Other Asian | Wong et al. |
| Alpay | Filipino | Other Asian | Wong et al. |
| Alphonso | Asian Indian | South Asian | Wong et al. |
| Alpuerto | Filipino | Other Asian | Wong et al. |
| Alquero | Filipino | Other Asian | Wong et al. |
| Alqueza | Filipino | Other Asian | Wong et al. |
| Alquilita | Filipino | Other Asian | Wong et al. |
| Alquiros | Filipino | Other Asian | Wong et al. |
| Alquiza | Filipino | Other Asian | Wong et al. |
| Alsaidi |  | South Asian | Lim et al. |
| Alsaybar | Filipino | Other Asian | Wong et al. |
| Alsisto | Filipino | Other Asian | Wong et al. |
| Alsol | Filipino | Other Asian | Wong et al. |
| Altaf |  | South Asian | Lim et al. |
| Altamero | Filipino | Other Asian | Wong et al. |
| Altamira | Filipino | Other Asian | Wong et al. |
| Altamirano | Filipino | Other Asian | Wong et al. |
| Altarejos | Filipino | Other Asian | Wong et al. |
| Altares | Filipino | Other Asian | Wong et al. |
| Altea | Filipino | Other Asian | Wong et al. |
| Alterado | Filipino | Other Asian | Wong et al. |
| Altiveros | Filipino | Other Asian | Wong et al. |
| Altre | Filipino | Other Asian | Wong et al. |
| Altuna | Filipino | Other Asian | Wong et al. |
| Altura | Filipino | Other Asian | Wong et al. |
| Aluag | Filipino | Other Asian | Wong et al. |
| Aludino | Filipino | Other Asian | Wong et al. |
| Alumia | Filipino | Other Asian | Wong et al. |
| Alumno | Filipino | Other Asian | Wong et al. |
| Alumpe | Filipino | Other Asian | Wong et al. |
| Alunan | Filipino | Other Asian | Wong et al. |
| Alupay | Filipino | Other Asian | Wong et al. |
| Alva | Filipino | Other Asian | Wong et al. |
| Alvarado | Filipino | Other Asian | Wong et al. |
| Alvaran | Filipino | Other Asian | Wong et al. |
| Alvares | Asian Indian | South Asian | Wong et al. |
| Alvarez | Filipino | Other Asian | Wong et al. |
| Alvarico | Filipino | Other Asian | Wong et al. |
| Alvaro | Filipino | Other Asian | Wong et al. |
| Alvear | Filipino | Other Asian | Wong et al. |
| Alvendia | Filipino | Other Asian | Wong et al. |
| Alvero | Filipino | Other Asian | Wong et al. |
| Alversado | Filipino | Other Asian | Wong et al. |
| Alvez | Filipino | Other Asian | Wong et al. |
| Alvi | Asian Indian | South Asian | Wong et al. |
| Alvia | Filipino | Other Asian | Wong et al. |
| Alviar | Filipino | Other Asian | Wong et al. |
| Alviedo | Filipino | Other Asian | Wong et al. |
| Alviento | Filipino | Other Asian | Wong et al. |
| Alviola | Filipino | Other Asian | Wong et al. |
| Alvior | Filipino | Other Asian | Wong et al. |
| Alvir | Filipino | Other Asian | Wong et al. |
| Alviz | Filipino | Other Asian | Wong et al. |
| Alwani | Asian Indian | South Asian | Wong et al. |
| Alwar | Asian Indian | South Asian | Wong et al. |
| Aly | Pakistani | South Asian | Kranick et al. |
| Alzate | Filipino | Other Asian | Wong et al. |
| Alzona | Filipino | Other Asian | Wong et al. |
| Ama | Filipino | Other Asian | Wong et al. |
| Amacio | Filipino | Other Asian | Wong et al. |
| Amada | Japanese | Other Asian | Wong et al. |
| Amadeo | Filipino | Other Asian | Wong et al. |
| Amado | Filipino | Other Asian | Kranick et al. |
| Amador | Filipino | Other Asian | Wong et al. |
| Amagai | Japanese | Other Asian | Wong et al. |
| Amagna | Filipino | Other Asian | Wong et al. |
| Amago | Filipino | Other Asian | Wong et al. |
| Amaguin | Filipino | Other Asian | Wong et al. |
| Amakawa | Japanese | Other Asian | Wong et al. |
| Amancio | Filipino | Other Asian | Wong et al. |
| Amando | Filipino | Other Asian | Wong et al. |
| Amano | Japanese | Other Asian | Wong et al. |
| Amansec | Filipino | Other Asian | Wong et al. |
| Amante | Filipino | Other Asian | Wong et al. |
| Amantillo | Filipino | Other Asian | Wong et al. |
| Amanullah | Asian Indian | South Asian | Wong et al. |
| Amar | Filipino | Other Asian | Wong et al. |
| Amaranto | Filipino | Other Asian | Wong et al. |
| Amarasinghe |  | South Asian | Lim et al. |
| Amarga | Filipino | Other Asian | Kranick et al. |
| Amargo | Filipino | Other Asian | Wong et al. |
| Amaria | Asian Indian | South Asian | Wong et al. |
| Amarillas | Filipino | Other Asian | Wong et al. |
| Amarillo | Filipino | Other Asian | Wong et al. |
| Amaro | Filipino | Other Asian | Wong et al. |
| Amat | Filipino | Other Asian | Wong et al. |
| Amatong | Filipino | Other Asian | Wong et al. |
| Amaya | Japanese | Other Asian | Wong et al. |
| Amazona | Filipino | Other Asian | Wong et al. |
| Ambal | Filipino | Other Asian | Wong et al. |
| Ambalal | Asian Indian | South Asian | Wong et al. |
| Ambalong | Filipino | Other Asian | Wong et al. |
| Ambas | Filipino | Other Asian | Wong et al. |
| Ambat | Filipino | Other Asian | Wong et al. |
| Ambata | Filipino | Other Asian | Wong et al. |
| Ambekar | Asian Indian | South Asian | Wong et al. |
| Ambia |  | South Asian | Lim et al. |
| Ambida | Filipino | Other Asian | Wong et al. |
| Ambion | Filipino | Other Asian | Wong et al. |
| Ambito | Filipino | Other Asian | Wong et al. |
| Amboy | Filipino | Other Asian | Wong et al. |
| Ambrocio | Filipino | Other Asian | Wong et al. |
| Ambulo | Filipino | Other Asian | Wong et al. |
| Ambwani | Asian Indian | South Asian | Wong et al. |
| Ame | Filipino | Other Asian | Wong et al. |
| Amechazurra | Filipino | Other Asian | Wong et al. |
| Ameeruddin | Asian Indian | South Asian | Wong et al. |
| Amemiya | Japanese | Other Asian | Wong et al. |
| Ametani | Japanese | Other Asian | Wong et al. |
| Ami | Filipino | Other Asian | Wong et al. |
| Amian | Filipino | Other Asian | Wong et al. |
| Amigable | Filipino | Other Asian | Wong et al. |
| Amigo | Filipino | Other Asian | Wong et al. |
| Amikura | Japanese | Other Asian | Wong et al. |
| Amil | Filipino | Other Asian | Wong et al. |
| Amimoto | Japanese | Other Asian | Wong et al. |
| Amin | Asian Indian | South Asian | Wong et al. |
| Amina |  | South Asian | Lim et al. |
| Amine |  | South Asian | Lim et al. |
| Amino | Japanese | Other Asian | Wong et al. |
| Amio | Filipino | Other Asian | Wong et al. |
| Amir |  | South Asian | Lim et al. |
| Amiri | Asian Indian | South Asian | Wong et al. |
| Amirzada | Afghanistan | South Asian | Kranick et al. |
| Amisola | Filipino | Other Asian | Wong et al. |
| Amistad | Filipino | Other Asian | Wong et al. |
| Amistoso | Filipino | Other Asian | Wong et al. |
| Amit | Filipino | Other Asian | Wong et al. |
| Amiya | Japanese | Other Asian | Wong et al. |
| Amjad | Asian Indian | South Asian | Wong et al. |
| Amlani | Asian Indian | South Asian | Wong et al. |
| Amma | Asian Indian | South Asian | Wong et al. |
| Ammad |  | South Asian | Lim et al. |
| Ammay | Filipino | Other Asian | Wong et al. |
| Amodia | Filipino | Other Asian | Wong et al. |
| Amodo | Filipino | Other Asian | Wong et al. |
| Amog | Filipino | Other Asian | Wong et al. |
| Amoguis | Filipino | Other Asian | Kranick et al. |
| Amor | Filipino | Other Asian | Wong et al. |
| Amora | Filipino | Other Asian | Wong et al. |
| Amoranto | Filipino | Other Asian | Wong et al. |
| Amores | Filipino | Other Asian | Wong et al. |
| Amorin | Filipino | Other Asian | Wong et al. |
| Amoroso | Filipino | Other Asian | Kranick et al. |
| Amorsolo | Filipino | Other Asian | Wong et al. |
| Amoyo | Filipino | Other Asian | Wong et al. |
| Amparado | Filipino | Other Asian | Wong et al. |
| Amparo | Filipino | Other Asian | Wong et al. |
| Amper | Filipino | Other Asian | Wong et al. |
| Ampil | Filipino | Other Asian | Wong et al. |
| Ampon | Filipino | Other Asian | Wong et al. |
| Amponin | Filipino | Other Asian | Wong et al. |
| Amposta | Filipino | Other Asian | Wong et al. |
| Amritlal | Asian Indian | South Asian | Wong et al. |
| Amuan | Filipino | Other Asian | Wong et al. |
| Amurao | Filipino | Other Asian | Wong et al. |
| An | Korean | Other Asian | Wong et al. |
| Ana | Filipino | Other Asian | Wong et al. |
| Anacan | Filipino | Other Asian | Wong et al. |
| Anacleto | Filipino | Other Asian | Wong et al. |
| Anacta | Filipino | Other Asian | Wong et al. |
| Anada | Japanese | Other Asian | Wong et al. |
| Anagaran | Filipino | Other Asian | Wong et al. |
| Anam | Indonesia; Bangladesh | Other Asian | Kranick et al. |
| Anama | Filipino | Other Asian | Wong et al. |
| Anan | Japanese | Other Asian | Wong et al. |
| Anana | Filipino | Other Asian | Wong et al. |
| Anand | Asian Indian | South Asian | Wong et al. |
| Ananda | Asian Indian | South Asian | Wong et al. |
| Anandam | Asian Indian | South Asian | Wong et al. |
| Anandan | Asian Indian | South Asian | Wong et al. |
| Ananthakrish | Asian Indian | South Asian | Wong et al. |
| Ananthanaray | Asian Indian | South Asian | Wong et al. |
| Anantharaman | Asian Indian | South Asian | Wong et al. |
| Anasco | Filipino | Other Asian | Wong et al. |
| Anastacio | Filipino | Other Asian | Wong et al. |
| Anatalio | Filipino | Other Asian | Wong et al. |
| Anaya | Filipino | Other Asian | Wong et al. |
| Ancajas | Filipino | Other Asian | Wong et al. |
| Ancheta | Filipino | Other Asian | Wong et al. |
| Anchita | Filipino | Other Asian | Wong et al. |
| Ancho | Filipino | Other Asian | Wong et al. |
| Anciano | Filipino | Other Asian | Wong et al. |
| Anciro | Filipino | Other Asian | Wong et al. |
| Ancog | Filipino | Other Asian | Wong et al. |
| Anda | Filipino | Other Asian | Wong et al. |
| Andaca | Filipino | Other Asian | Wong et al. |
| Andal | Filipino | Other Asian | Wong et al. |
| Andales | Filipino | Other Asian | Wong et al. |
| Andalis | Filipino | Other Asian | Wong et al. |
| Andallo | Filipino | Other Asian | Wong et al. |
| Andam | Filipino | Other Asian | Wong et al. |
| Anday | Filipino | Other Asian | Wong et al. |
| Andaya | Filipino | Other Asian | Wong et al. |
| Anden | Filipino | Other Asian | Wong et al. |
| Andia | Filipino | Other Asian | Wong et al. |
| Andico | Filipino | Other Asian | Wong et al. |
| Andiyappa |  | South Asian | Lim et al. |
| Ando | Japanese | Other Asian | Wong et al. |
| Andongo | Filipino | Other Asian | Wong et al. |
| Andoy | Filipino | Other Asian | Wong et al. |
| Andrada | Filipino | Other Asian | Wong et al. |
| Andrade | Filipino | Other Asian | Wong et al. |
| Andres | Filipino | Other Asian | Wong et al. |
| Andrion | Filipino | Other Asian | Wong et al. |
| Anfone | Filipino | Other Asian | Wong et al. |
| Angala | Filipino | Other Asian | Wong et al. |
| Angara | Filipino | Other Asian | Wong et al. |
| Angat | Filipino | Other Asian | Wong et al. |
| Angcaya | Filipino | Other Asian | Wong et al. |
| Angco | Filipino | Other Asian | Wong et al. |
| Angeles | Filipino | Other Asian | Wong et al. |
| Angelito | Filipino | Other Asian | Wong et al. |
| Angeo | Filipino | Other Asian | Wong et al. |
| Angkico | Filipino | Other Asian | Wong et al. |
| Anglo | Filipino | Other Asian | Wong et al. |
| Angoluan | Filipino | Other Asian | Wong et al. |
| Angsioco | Filipino | Other Asian | Wong et al. |
| Angtuaco | Filipino | Other Asian | Wong et al. |
| Angud | Filipino | Other Asian | Wong et al. |
| Angulo | Filipino | Other Asian | Wong et al. |
| Angustia | Filipino | Other Asian | Wong et al. |
| Anh | Vietnamese | Other Asian | Wong et al. |
| Ani | Filipino | Other Asian | Wong et al. |
| Aniag | Filipino | Other Asian | Wong et al. |
| Aniano | Filipino | Other Asian | Wong et al. |
| Aniban | Filipino | Other Asian | Wong et al. |
| Anicete | Filipino | Other Asian | Wong et al. |
| Aniceto | Filipino | Other Asian | Wong et al. |
| Aniciete | Filipino | Other Asian | Wong et al. |
| Aniel | Filipino | Other Asian | Wong et al. |
| Anies | Filipino | Other Asian | Wong et al. |
| Anilao | Filipino | Other Asian | Wong et al. |
| Aninag | Filipino | Other Asian | Wong et al. |
| Aninion | Filipino | Other Asian | Wong et al. |
| Anisuzzaman |  | South Asian | Lim et al. |
| Anit | Filipino | Other Asian | Wong et al. |
| Anjaria | Asian Indian | South Asian | Wong et al. |
| Anjum |  | South Asian | Lim et al. |
| Anklesaria | Asian Indian | South Asian | Wong et al. |
| Ann | Korean | Other Asian | Wong et al. |
| Annamalai | Asian Indian | South Asian | Wong et al. |
| Anoba | Filipino | Other Asian | Wong et al. |
| Anoche | Filipino | Other Asian | Wong et al. |
| Anolin | Filipino | Other Asian | Wong et al. |
| Anonas | Filipino | Other Asian | Wong et al. |
| Anongos | Filipino | Other Asian | Wong et al. |
| Anonuevo | Filipino | Other Asian | Wong et al. |
| Anor | Filipino | Other Asian | Wong et al. |
| Anorico | Filipino | Other Asian | Wong et al. |
| Anover | Filipino | Other Asian | Wong et al. |
| Anowar |  | South Asian | Lim et al. |
| Anraku | Japanese | Other Asian | Wong et al. |
| Ansagay | Filipino | Other Asian | Wong et al. |
| Ansari | Asian Indian; Pakistani | South Asian | Kranick et al. |
| Ansay | Filipino | Other Asian | Wong et al. |
| Antalan | Filipino | Other Asian | Wong et al. |
| Antani | Asian Indian | South Asian | Wong et al. |
| Antaran | Filipino | Other Asian | Wong et al. |
| Antazo | Filipino | Other Asian | Wong et al. |
| Ante | Filipino | Other Asian | Wong et al. |
| Anteola | Filipino | Other Asian | Wong et al. |
| Antero | Filipino | Other Asian | Wong et al. |
| Antia | Asian Indian | South Asian | Wong et al. |
| Antigua | Filipino | Other Asian | Wong et al. |
| Antillon | Filipino | Other Asian | Wong et al. |
| Antimano | Filipino | Other Asian | Wong et al. |
| Antioquia | Filipino | Other Asian | Wong et al. |
| Antipolo | Filipino | Other Asian | Wong et al. |
| Antiporda | Filipino | Other Asian | Wong et al. |
| Antiporta | Filipino | Other Asian | Wong et al. |
| Antipuesto | Filipino | Other Asian | Wong et al. |
| Antiquera | Filipino | Other Asian | Wong et al. |
| Antivo | Filipino | Other Asian | Wong et al. |
| Antolin | Filipino | Other Asian | Wong et al. |
| Antonio | Filipino | Other Asian | Wong et al. |
| Antony | Asian Indian | South Asian | Wong et al. |
| Anulacion | Filipino | Other Asian | Wong et al. |
| Anulao | Filipino | Other Asian | Wong et al. |
| Anunciacion | Filipino | Other Asian | Wong et al. |
| Anvernali |  | South Asian | Lim et al. |
| Anwar | Pakistani; Bangladeshi | South Asian | Kranick et al. |
| Anwara |  | South Asian | Lim et al. |
| Anwer |  | South Asian | Lim et al. |
| Anzai | Japanese | Other Asian | Wong et al. |
| Anzano | Filipino | Other Asian | Wong et al. |
| Anzures | Filipino | Other Asian | Wong et al. |
| Ao | Chinese | Chinese | Wong et al. |
| Aoanan | Filipino | Other Asian | Wong et al. |
| Aoay | Filipino | Other Asian | Wong et al. |
| Aoi | Japanese | Other Asian | Wong et al. |
| Aoki | Japanese | Other Asian | Wong et al. |
| Aono | Japanese | Other Asian | Wong et al. |
| Aonuma | Japanese | Other Asian | Wong et al. |
| Aoshima | Japanese | Other Asian | Wong et al. |
| Aoto | Japanese | Other Asian | Wong et al. |
| Aoyagi | Japanese | Other Asian | Wong et al. |
| Aoyama | Japanese | Other Asian | Wong et al. |
| Apacible | Filipino | Other Asian | Wong et al. |
| Apaga | Filipino | Other Asian | Wong et al. |
| Apalisok | Filipino | Other Asian | Wong et al. |
| Apalit | Filipino | Other Asian | Wong et al. |
| Apalla | Filipino | Other Asian | Wong et al. |
| Apan | Filipino | Other Asian | Wong et al. |
| Apanay | Filipino | Other Asian | Wong et al. |
| Apao | Filipino | Other Asian | Wong et al. |
| Aparici | Filipino | Other Asian | Wong et al. |
| Aparicio | Filipino | Other Asian | Wong et al. |
| Aparis | Filipino | Other Asian | Wong et al. |
| Aparri | Filipino | Other Asian | Wong et al. |
| Apelado | Filipino | Other Asian | Wong et al. |
| Apellanes | Filipino | Other Asian | Wong et al. |
| Apellido | Filipino | Other Asian | Wong et al. |
| Apelo | Filipino | Other Asian | Wong et al. |
| Apiado | Filipino | Other Asian | Wong et al. |
| Apigo | Filipino | Other Asian | Wong et al. |
| Apilado | Filipino | Other Asian | Wong et al. |
| Apillanes | Filipino | Other Asian | Wong et al. |
| Apis | Filipino | Other Asian | Wong et al. |
| Apo | Filipino | Other Asian | Wong et al. |
| Apolinar | Filipino | Other Asian | Wong et al. |
| Apolinario | Filipino | Other Asian | Wong et al. |
| Apolo | Filipino | Other Asian | Wong et al. |
| Apolona | Filipino | Other Asian | Wong et al. |
| Apolonio | Filipino | Other Asian | Wong et al. |
| Apongol | Filipino | Other Asian | Wong et al. |
| Apor | Filipino | Other Asian | Wong et al. |
| Apostol | Filipino | Other Asian | Wong et al. |
| Appadurai | Asian Indian | South Asian | Wong et al. |
| Apruebo | Filipino | Other Asian | Wong et al. |
| Apsay | Filipino | Other Asian | Wong et al. |
| Apuada | Filipino | Other Asian | Wong et al. |
| Apuan | Filipino | Other Asian | Wong et al. |
| Apura | Filipino | Other Asian | Wong et al. |
| Apuya | Filipino | Other Asian | Wong et al. |
| Apuyan | Filipino | Other Asian | Wong et al. |
| Aque | Filipino | Other Asian | Wong et al. |
| Aqui | Filipino | Other Asian | Wong et al. |
| Aquilar | Filipino | Other Asian | Wong et al. |
| Aquilizan | Filipino | Other Asian | Wong et al. |
| Aquinaldo | Filipino | Other Asian | Wong et al. |
| Aquinde | Filipino | Other Asian | Wong et al. |
| Aquino | Filipino | Other Asian | Kranick et al. |
| Aquitania | Filipino | Other Asian | Wong et al. |
| Ara |  | South Asian | Lim et al. |
| Arabe | Filipino | Other Asian | Wong et al. |
| Arabejo | Filipino | Other Asian | Wong et al. |
| Arabit | Filipino | Other Asian | Wong et al. |
| Arackal | Asian Indian | South Asian | Wong et al. |
| Arada | Filipino | Other Asian | Wong et al. |
| Aradanas | Filipino | Other Asian | Wong et al. |
| Arafat |  | South Asian | Lim et al. |
| Arafiles | Filipino | Other Asian | Wong et al. |
| Araga | Filipino | Other Asian | Wong et al. |
| Aragaki | Japanese | Other Asian | Kranick et al. |
| Arago | Filipino | Other Asian | Wong et al. |
| Aragon | Filipino | Other Asian | Wong et al. |
| Aragones | Filipino | Other Asian | Wong et al. |
| Aragoza | Filipino | Other Asian | Wong et al. |
| Arahaque |  | South Asian | Lim et al. |
| Arai | Japanese | Other Asian | Wong et al. |
| Arakaki | Japanese | Other Asian | Wong et al. |
| Arakawa | Japanese | Other Asian | Wong et al. |
| Araki | Japanese | Other Asian | Wong et al. |
| Arakkal | Asian Indian | South Asian | Wong et al. |
| Aral | Japanese | Other Asian | Wong et al. |
| Aralar | Filipino | Other Asian | Wong et al. |
| Aramaki | Japanese | Other Asian | Wong et al. |
| Arambulo | Filipino | Other Asian | Wong et al. |
| Arana | Filipino | Other Asian | Wong et al. |
| Aranas | Filipino | Other Asian | Wong et al. |
| Aranca | Filipino | Other Asian | Wong et al. |
| Arancillo | Filipino | Other Asian | Wong et al. |
| Aranda | Filipino | Other Asian | Wong et al. |
| Arandela | Filipino | Other Asian | Wong et al. |
| Arandez | Filipino | Other Asian | Wong et al. |
| Arandia | Filipino | Other Asian | Wong et al. |
| Arando | Filipino | Other Asian | Wong et al. |
| Araneta | Filipino | Other Asian | Wong et al. |
| Aranez | Filipino | Other Asian | Wong et al. |
| Arangorin | Filipino | Other Asian | Wong et al. |
| Aranha | Asian Indian | South Asian | Wong et al. |
| Aranico | Filipino | Other Asian | Wong et al. |
| Araniego | Filipino | Other Asian | Wong et al. |
| Aranilla | Filipino | Other Asian | Wong et al. |
| Arano | Filipino | Other Asian | Wong et al. |
| Arante | Filipino | Other Asian | Wong et al. |
| Aranton | Filipino | Other Asian | Wong et al. |
| Aranza | Filipino | Other Asian | Wong et al. |
| Aranzamendez | Filipino | Other Asian | Wong et al. |
| Aranzanso | Filipino | Other Asian | Wong et al. |
| Aranzaso | Filipino | Other Asian | Wong et al. |
| Arao | Filipino | Other Asian | Wong et al. |
| Araojo | Filipino | Other Asian | Wong et al. |
| Araos | Filipino | Other Asian | Wong et al. |
| Araquel | Filipino | Other Asian | Wong et al. |
| Araracap | Filipino | Other Asian | Wong et al. |
| Arasaki | Japanese | Other Asian | Wong et al. |
| Arase | Japanese | Other Asian | Wong et al. |
| Arashiro | Japanese | Other Asian | Wong et al. |
| Arastu | Asian Indian | South Asian | Wong et al. |
| Aratani | Japanese | Other Asian | Wong et al. |
| Aratea | Filipino | Other Asian | Wong et al. |
| Araujo | Filipino | Other Asian | Wong et al. |
| Araullo | Filipino | Other Asian | Wong et al. |
| Arayata | Filipino | Other Asian | Wong et al. |
| Araza | Filipino | Other Asian | Wong et al. |
| Arban | Filipino | Other Asian | Wong et al. |
| Arbas | Filipino | Other Asian | Wong et al. |
| Arbilo | Filipino | Other Asian | Wong et al. |
| Arbis | Filipino | Other Asian | Wong et al. |
| Arbitrario | Filipino | Other Asian | Wong et al. |
| Arbizo | Filipino | Other Asian | Wong et al. |
| Arbolado | Filipino | Other Asian | Wong et al. |
| Arboleda | Filipino | Other Asian | Wong et al. |
| Arbolente | Filipino | Other Asian | Wong et al. |
| Arbues | Filipino | Other Asian | Wong et al. |
| Arca | Filipino | Other Asian | Wong et al. |
| Arcadio | Filipino | Other Asian | Wong et al. |
| Arcaina | Filipino | Other Asian | Wong et al. |
| Arcala | Filipino | Other Asian | Wong et al. |
| Arcalas | Filipino | Other Asian | Wong et al. |
| Arcamo | Filipino | Other Asian | Wong et al. |
| Arcangel | Filipino | Other Asian | Wong et al. |
| Arcano | Filipino | Other Asian | Wong et al. |
| Arcaya | Filipino | Other Asian | Wong et al. |
| Arce | Filipino | Other Asian | Wong et al. |
| Arcebal | Filipino | Other Asian | Wong et al. |
| Arcebido | Filipino | Other Asian | Wong et al. |
| Arcega | Filipino | Other Asian | Wong et al. |
| Arcellana | Filipino | Other Asian | Wong et al. |
| Arcelo | Filipino | Other Asian | Wong et al. |
| Arcelona | Filipino | Other Asian | Wong et al. |
| Arcena | Filipino | Other Asian | Wong et al. |
| Arcenal | Filipino | Other Asian | Wong et al. |
| Arcenas | Filipino | Other Asian | Wong et al. |
| Arceno | Filipino | Other Asian | Wong et al. |
| Arceo | Filipino | Other Asian | Wong et al. |
| Arches | Filipino | Other Asian | Wong et al. |
| Arcia | Filipino | Other Asian | Wong et al. |
| Arciaga | Filipino | Other Asian | Wong et al. |
| Arcibal | Filipino | Other Asian | Wong et al. |
| Arciga | Filipino | Other Asian | Wong et al. |
| Arcilla | Filipino | Other Asian | Wong et al. |
| Arcillas | Filipino | Other Asian | Wong et al. |
| Arcinas | Filipino | Other Asian | Wong et al. |
| Arcitio | Filipino | Other Asian | Wong et al. |
| Arconado | Filipino | Other Asian | Wong et al. |
| Ardales | Filipino | Other Asian | Wong et al. |
| Arde | Filipino | Other Asian | Wong et al. |
| Ardiente | Filipino | Other Asian | Wong et al. |
| Arejola | Filipino | Other Asian | Wong et al. |
| Arellano | Filipino | Other Asian | Kranick et al. |
| Arenas | Filipino | Other Asian | Wong et al. |
| Areno | Filipino | Other Asian | Wong et al. |
| Arenzana | Filipino | Other Asian | Wong et al. |
| Areola | Filipino | Other Asian | Wong et al. |
| Ares | Filipino | Other Asian | Wong et al. |
| Arespacochag | Filipino | Other Asian | Wong et al. |
| Arevalo | Filipino | Other Asian | Kranick et al. |
| Argallon | Filipino | Other Asian | Wong et al. |
| Argame | Filipino | Other Asian | Wong et al. |
| Argana | Filipino | Other Asian | Wong et al. |
| Arganosa | Filipino | Other Asian | Wong et al. |
| Argao | Filipino | Other Asian | Wong et al. |
| Argarin | Filipino | Other Asian | Wong et al. |
| Argel | Filipino | Other Asian | Wong et al. |
| Argente | Filipino | Other Asian | Wong et al. |
| Argonza | Filipino | Other Asian | Wong et al. |
| Argosino | Filipino | Other Asian | Wong et al. |
| Arguelles | Filipino | Other Asian | Kranick et al. |
| Argueza | Filipino | Other Asian | Wong et al. |
| Arguilla | Filipino | Other Asian | Wong et al. |
| Arguilles | Filipino | Other Asian | Wong et al. |
| Arguson | Filipino | Other Asian | Wong et al. |
| Arias | Filipino | Other Asian | Wong et al. |
| Ariate | Filipino | Other Asian | Wong et al. |
| Aribon | Filipino | Other Asian | Wong et al. |
| Aricheta | Filipino | Other Asian | Wong et al. |
| Arie | Japanese | Other Asian | Wong et al. |
| Arif | Pakistani; Bangladeshi | South Asian | Kranick et al. |
| Arifadil |  | South Asian | Lim et al. |
| Ariga | Japanese | Other Asian | Wong et al. |
| Arii | Japanese | Other Asian | Wong et al. |
| Arikawa | Japanese | Other Asian | Wong et al. |
| Arillano | Filipino | Other Asian | Wong et al. |
| Arima | Japanese | Other Asian | Wong et al. |
| Arimori | Japanese | Other Asian | Wong et al. |
| Arimoto | Japanese | Other Asian | Wong et al. |
| Arimura | Japanese | Other Asian | Wong et al. |
| Arinduque | Filipino | Other Asian | Wong et al. |
| Arino | Filipino | Other Asian | Wong et al. |
| Arinto | Filipino | Other Asian | Wong et al. |
| Arioka | Japanese | Other Asian | Wong et al. |
| Ariola | Filipino | Other Asian | Wong et al. |
| Arios | Filipino | Other Asian | Wong et al. |
| Aris | Filipino | Other Asian | Wong et al. |
| Arisaka | Japanese | Other Asian | Wong et al. |
| Arisawa | Japanese | Other Asian | Wong et al. |
| Aristorenas | Filipino | Other Asian | Wong et al. |
| Arita | Japanese | Other Asian | Wong et al. |
| Ariyama | Japanese | Other Asian | Wong et al. |
| Ariyoshi | Japanese | Other Asian | Wong et al. |
| Ariz | Filipino | Other Asian | Wong et al. |
| Ariza | Filipino | Other Asian | Wong et al. |
| Arizabal | Filipino | Other Asian | Wong et al. |
| Arizala | Filipino | Other Asian | Wong et al. |
| Arizo | Filipino | Other Asian | Wong et al. |
| Arjona | Filipino | Other Asian | Wong et al. |
| Arju |  | South Asian | Lim et al. |
| Ark | Chinese | Chinese | Wong et al. |
| Arlantico | Filipino | Other Asian | Wong et al. |
| Armada | Filipino | Other Asian | Wong et al. |
| Armado | Filipino | Other Asian | Wong et al. |
| Armamento | Filipino | Other Asian | Wong et al. |
| Armandico | Filipino | Other Asian | Wong et al. |
| Armas | Filipino | Other Asian | Wong et al. |
| Armedilla | Filipino | Other Asian | Wong et al. |
| Armonio | Filipino | Other Asian | Wong et al. |
| Arnaiz | Filipino | Other Asian | Wong et al. |
| Arnaldo | Filipino | Other Asian | Wong et al. |
| Arnedo | Filipino | Other Asian | Wong et al. |
| Arneja | Asian Indian | South Asian | Wong et al. |
| Arnesto | Filipino | Other Asian | Wong et al. |
| Arnobit | Filipino | Other Asian | Wong et al. |
| Arnoco | Filipino | Other Asian | Wong et al. |
| Aroc | Filipino | Other Asian | Wong et al. |
| Arocena | Filipino | Other Asian | Wong et al. |
| Aromin | Filipino | Other Asian | Wong et al. |
| Arong | Filipino | Other Asian | Wong et al. |
| Arora | Asian Indian | South Asian | Wong et al. |
| Arpon | Filipino | Other Asian | Wong et al. |
| Arquero | Filipino | Other Asian | Wong et al. |
| Arquilada | Filipino | Other Asian | Wong et al. |
| Arquillano | Filipino | Other Asian | Wong et al. |
| Arquillo | Filipino | Other Asian | Wong et al. |
| Arquines | Filipino | Other Asian | Wong et al. |
| Arquiza | Filipino | Other Asian | Wong et al. |
| Arradaza | Filipino | Other Asian | Wong et al. |
| Arrastia | Filipino | Other Asian | Wong et al. |
| Arre | Filipino | Other Asian | Wong et al. |
| Arreola | Filipino | Other Asian | Wong et al. |
| Arreza | Filipino | Other Asian | Wong et al. |
| Arriesgado | Filipino | Other Asian | Wong et al. |
| Arrieta | Filipino | Other Asian | Wong et al. |
| Arriola | Filipino | Other Asian | Wong et al. |
| Arrocena | Filipino | Other Asian | Wong et al. |
| Arrogante | Filipino | Other Asian | Wong et al. |
| Arrojo | Filipino | Other Asian | Wong et al. |
| Arroyo | Filipino | Other Asian | Wong et al. |
| Arroz | Filipino | Other Asian | Wong et al. |
| Arrozal | Filipino | Other Asian | Wong et al. |
| Arsenal | Filipino | Other Asian | Wong et al. |
| Arshad | Pakistani | South Asian | Kranick et al. |
| Arsua | Filipino | Other Asian | Wong et al. |
| Artacho | Filipino | Other Asian | Wong et al. |
| Artadi | Filipino | Other Asian | Wong et al. |
| Artates | Filipino | Other Asian | Wong et al. |
| Arteche | Filipino | Other Asian | Wong et al. |
| Artiaga | Filipino | Other Asian | Wong et al. |
| Artienda | Filipino | Other Asian | Wong et al. |
| Artigas | Filipino | Other Asian | Wong et al. |
| Artugue | Filipino | Other Asian | Wong et al. |
| Artuz | Filipino | Other Asian | Wong et al. |
| Arucan | Filipino | Other Asian | Wong et al. |
| Aruga | Japanese | Other Asian | Wong et al. |
| Arugay | Filipino | Other Asian | Wong et al. |
| Arumainayaga | Asian Indian | South Asian | Wong et al. |
| Arumugam | Asian Indian | South Asian | Wong et al. |
| Arunachalam | Asian Indian | South Asian | Wong et al. |
| Arya | Asian Indian | South Asian | Kranick et al. |
| Arzadon | Filipino | Other Asian | Wong et al. |
| Arzaga | Filipino | Other Asian | Wong et al. |
| Asaba | Japanese | Other Asian | Wong et al. |
| Asad |  | South Asian | Lim et al. |
| Asada | Japanese | Other Asian | Wong et al. |
| Asadduzzaman |  | South Asian | Lim et al. |
| Asadon | Filipino | Other Asian | Wong et al. |
| Asadullah | Asian Indian | South Asian | Wong et al. |
| Asaga | Japanese | Other Asian | Wong et al. |
| Asahan | Filipino | Other Asian | Wong et al. |
| Asahara | Japanese | Other Asian | Wong et al. |
| Asahi | Japanese | Other Asian | Wong et al. |
| Asahina | Japanese | Other Asian | Wong et al. |
| Asai | Japanese | Other Asian | Wong et al. |
| Asaka | Japanese | Other Asian | Wong et al. |
| Asakawa | Japanese | Other Asian | Wong et al. |
| Asaki | Japanese | Other Asian | Wong et al. |
| Asako | Japanese | Other Asian | Wong et al. |
| Asakura | Japanese | Other Asian | Wong et al. |
| Asama | Japanese | Other Asian | Wong et al. |
| Asami | Japanese | Other Asian | Wong et al. |
| Asanion | Filipino | Other Asian | Wong et al. |
| Asano | Japanese | Other Asian | Wong et al. |
| Asanuma | Japanese | Other Asian | Wong et al. |
| Asanza | Filipino | Other Asian | Wong et al. |
| Asao | Japanese | Other Asian | Wong et al. |
| Asaoka | Japanese | Other Asian | Wong et al. |
| Asar | Asian Indian | South Asian | Wong et al. |
| Asari | Japanese | Other Asian | Wong et al. |
| Asato | Japanese | Other Asian | Wong et al. |
| Asayama | Japanese | Other Asian | Wong et al. |
| Asazuma | Japanese | Other Asian | Wong et al. |
| Ascalon | Filipino | Other Asian | Kranick et al. |
| Ascano | Filipino | Other Asian | Wong et al. |
| Ascue | Filipino | Other Asian | Wong et al. |
| Asejo | Filipino | Other Asian | Wong et al. |
| Asencion | Filipino | Other Asian | Wong et al. |
| Aseniero | Filipino | Other Asian | Wong et al. |
| Asensi | Filipino | Other Asian | Wong et al. |
| Asercion | Filipino | Other Asian | Wong et al. |
| Aseron | Filipino | Other Asian | Wong et al. |
| Asghar | Asian Indian | South Asian | Wong et al. |
| Asha |  | South Asian | Lim et al. |
| Ashabhai | Asian Indian | South Asian | Wong et al. |
| Ashar | Asian Indian | South Asian | Wong et al. |
| Ashfaq | Asian Indian | South Asian | Wong et al. |
| Ashida | Japanese | Other Asian | Wong et al. |
| Ashihara | Japanese | Other Asian | Wong et al. |
| Ashikaga | Japanese | Other Asian | Wong et al. |
| Ashikawa | Japanese | Other Asian | Wong et al. |
| Ashimine | Japanese | Other Asian | Wong et al. |
| Ashiq | Asian Indian | South Asian | Wong et al. |
| Ashizawa | Japanese | Other Asian | Wong et al. |
| Ashraf | Pakistani | South Asian | Kranick et al. |
| Ashraff | Asian Indian | South Asian | Wong et al. |
| Ashrafi |  | South Asian | Lim et al. |
| Ashrafunnahar |  | South Asian | Lim et al. |
| Ashram |  | South Asian | Lim et al. |
| Asi | Filipino | Other Asian | Wong et al. |
| Asia | Filipino | Other Asian | Wong et al. |
| Asiamah |  | South Asian | Lim et al. |
| Asias | Filipino | Other Asian | Wong et al. |
| Asiatico | Filipino | Other Asian | Wong et al. |
| Asiddao | Filipino | Other Asian | Wong et al. |
| Asido | Filipino | Other Asian | Wong et al. |
| Asif | Asian Indian | South Asian | Wong et al. |
| Asilo | Filipino | Other Asian | Wong et al. |
| Asim |  | South Asian | Lim et al. |
| Asinas | Filipino | Other Asian | Wong et al. |
| Asino | Filipino | Other Asian | Wong et al. |
| Asio | Filipino | Other Asian | Wong et al. |
| Asirifi |  | South Asian | Lim et al. |
| Asirvatham | Asian Indian | South Asian | Wong et al. |
| Asis | Filipino | Other Asian | Wong et al. |
| Asisten | Filipino | Other Asian | Wong et al. |
| Asistin | Filipino | Other Asian | Wong et al. |
| Asistio | Filipino | Other Asian | Wong et al. |
| Asistores | Filipino | Other Asian | Wong et al. |
| Askari | Asian Indian | South Asian | Wong et al. |
| Aslam | Pakistani | South Asian | Kranick et al. |
| Asmd |  | South Asian | Lim et al. |
| Aso | Japanese | Other Asian | Wong et al. |
| Asoncion | Filipino | Other Asian | Wong et al. |
| Aspa | Filipino | Other Asian | Wong et al. |
| Aspacio | Filipino | Other Asian | Wong et al. |
| Aspe | Filipino | Other Asian | Wong et al. |
| Asperas | Filipino | Other Asian | Wong et al. |
| Asperilla | Filipino | Other Asian | Wong et al. |
| Asperin | Filipino | Other Asian | Wong et al. |
| Aspili | Filipino | Other Asian | Wong et al. |
| Aspillera | Filipino | Other Asian | Wong et al. |
| Aspiras | Filipino | Other Asian | Wong et al. |
| Asprec | Filipino | Other Asian | Wong et al. |
| Asprer | Filipino | Other Asian | Wong et al. |
| Aspuria | Filipino | Other Asian | Wong et al. |
| Assar | Asian Indian | South Asian | Wong et al. |
| Asthana | Asian Indian | South Asian | Wong et al. |
| Astilla | Filipino | Other Asian | Wong et al. |
| Astillero | Filipino | Other Asian | Wong et al. |
| Astorga | Filipino | Other Asian | Wong et al. |
| Astrande | Filipino | Other Asian | Wong et al. |
| Astraquillo | Filipino | Other Asian | Wong et al. |
| Astrero | Filipino | Other Asian | Wong et al. |
| Astronomo | Filipino | Other Asian | Wong et al. |
| Astudillo | Filipino | Other Asian | Wong et al. |
| Asturias | Filipino | Other Asian | Wong et al. |
| Asu | Chinese | Chinese | Wong et al. |
| Asuelo | Filipino | Other Asian | Wong et al. |
| Asuncion | Filipino | Other Asian | Kranick et al. |
| Asuzano | Filipino | Other Asian | Wong et al. |
| Atabay | Filipino | Other Asian | Wong et al. |
| Atacador | Filipino | Other Asian | Wong et al. |
| Atad | Filipino | Other Asian | Wong et al. |
| Ataka | Japanese | Other Asian | Wong et al. |
| Atal | Asian Indian | South Asian | Wong et al. |
| Atanacio | Filipino | Other Asian | Wong et al. |
| Atangan | Filipino | Other Asian | Wong et al. |
| Atarashi | Japanese | Other Asian | Wong et al. |
| Atashi |  | South Asian | Lim et al. |
| Atay | Filipino | Other Asian | Wong et al. |
| Atayan | Filipino | Other Asian | Wong et al. |
| Atayde | Filipino | Other Asian | Wong et al. |
| Ate | Chinese | Chinese | Wong et al. |
| Atega | Filipino | Other Asian | Wong et al. |
| Atencio | Filipino | Other Asian | Wong et al. |
| Atencion | Filipino | Other Asian | Wong et al. |
| Atendido | Filipino | Other Asian | Wong et al. |
| Atengco | Filipino | Other Asian | Wong et al. |
| Atento | Filipino | Other Asian | Wong et al. |
| Aterrado | Filipino | Other Asian | Wong et al. |
| Athavale | Asian Indian | South Asian | Wong et al. |
| Athwal | Asian Indian | South Asian | Wong et al. |
| Atienza | Filipino | Other Asian | Wong et al. |
| Atiga | Filipino | Other Asian | Wong et al. |
| Atijera | Filipino | Other Asian | Wong et al. |
| Atik |  | South Asian | Lim et al. |
| Atilano | Filipino | Other Asian | Wong et al. |
| Atillo | Filipino | Other Asian | Wong et al. |
| Atis | Filipino | Other Asian | Wong et al. |
| Atluri | Asian Indian | South Asian | Wong et al. |
| Atmosfera | Filipino | Other Asian | Wong et al. |
| Atobe | Japanese | Other Asian | Wong et al. |
| Atos | Filipino | Other Asian | Wong et al. |
| Atrero | Filipino | Other Asian | Wong et al. |
| Atri | Asian Indian | South Asian | Wong et al. |
| Atsumi | Japanese | Other Asian | Wong et al. |
| Attar |  | South Asian | Lim et al. |
| Attique |  | South Asian | Lim et al. |
| Attri | Asian Indian | South Asian | Wong et al. |
| Atud | Filipino | Other Asian | Wong et al. |
| Atup | Filipino | Other Asian | Wong et al. |
| Atutubo | Filipino | Other Asian | Wong et al. |
| Atwal | Asian Indian | South Asian | Wong et al. |
| Au | Chinese | Chinese | Kranick et al. |
| Audije | Filipino | Other Asian | Wong et al. |
| Auditor | Filipino | Other Asian | Wong et al. |
| Auduong | Vietnamese | Other Asian | Wong et al. |
| Auh | Korean | Other Asian | Wong et al. |
| Aujero | Filipino | Other Asian | Wong et al. |
| Aujla | Asian Indian | South Asian | Wong et al. |
| Auk | Chinese | Chinese | Wong et al. |
| Aulakh | Asian Indian | South Asian | Wong et al. |
| Aumentado | Filipino | Other Asian | Wong et al. |
| Aung | Burmese | Other Asian | Kranick et al. |
| Aurangazeb |  | South Asian | Lim et al. |
| Aure | Filipino | Other Asian | Wong et al. |
| Aurea | Filipino | Other Asian | Wong et al. |
| Aurelio | Filipino | Other Asian | Wong et al. |
| Aurellado | Filipino | Other Asian | Wong et al. |
| Aurellano | Filipino | Other Asian | Wong et al. |
| Aureus | Filipino | Other Asian | Wong et al. |
| Aurigue | Filipino | Other Asian | Wong et al. |
| Auro | Filipino | Other Asian | Wong et al. |
| Ausa | Filipino | Other Asian | Wong et al. |
| Ausan | Filipino | Other Asian | Wong et al. |
| Austero | Filipino | Other Asian | Wong et al. |
| Austria | Filipino | Other Asian | Kranick et al. |
| Austriaco | Filipino | Other Asian | Wong et al. |
| Autar |  | South Asian | Lim et al. |
| Autor | Filipino | Other Asian | Wong et al. |
| Auyang | Chinese | Chinese | Wong et al. |
| Auyeung | Chinese | Chinese | Wong et al. |
| Au-Yeung | Chinese | Chinese | Kranick et al. |
| Auyong | Chinese | Chinese | Wong et al. |
| Auyoung | Chinese | Chinese | Wong et al. |
| Avadhani | Asian Indian | South Asian | Wong et al. |
| Avancena | Filipino | Other Asian | Wong et al. |
| Avanzado | Filipino | Other Asian | Wong et al. |
| Avasthi | Asian Indian | South Asian | Wong et al. |
| Avecilla | Filipino | Other Asian | Wong et al. |
| Avelino | Filipino | Other Asian | Wong et al. |
| Avellana | Filipino | Other Asian | Wong et al. |
| Avellano | Filipino | Other Asian | Wong et al. |
| Avellanosa | Filipino | Other Asian | Wong et al. |
| Avena | Filipino | Other Asian | Wong et al. |
| Avendano | Filipino | Other Asian | Wong et al. |
| Avenido | Filipino | Other Asian | Wong et al. |
| Avenir | Filipino | Other Asian | Wong et al. |
| Aveno | Filipino | Other Asian | Wong et al. |
| Averia | Filipino | Other Asian | Wong et al. |
| Averilla | Filipino | Other Asian | Wong et al. |
| Averion | Filipino | Other Asian | Wong et al. |
| Avestruz | Filipino | Other Asian | Wong et al. |
| Aviado | Filipino | Other Asian | Wong et al. |
| Avichal | Asian Indian | South Asian | Wong et al. |
| Avila | Filipino | Other Asian | Kranick et al. |
| Aviles | Filipino | Other Asian | Wong et al. |
| Avilla | Filipino | Other Asian | Wong et al. |
| Avillanoza | Filipino | Other Asian | Wong et al. |
| Aviso | Filipino | Other Asian | Wong et al. |
| Avula | Asian Indian | South Asian | Wong et al. |
| Awa | Filipino | Other Asian | Wong et al. |
| Awai | Japanese | Other Asian | Wong et al. |
| Awal |  | South Asian | Lim et al. |
| Awan |  | South Asian | Lim et al. |
| Awasthi | Asian Indian | South Asian | Wong et al. |
| Awata | Japanese | Other Asian | Wong et al. |
| Awaya | Japanese | Other Asian | Wong et al. |
| Awayan | Filipino | Other Asian | Wong et al. |
| Awit | Filipino | Other Asian | Wong et al. |
| Axibal | Filipino | Other Asian | Wong et al. |
| Ay | Chinese | Chinese | Wong et al. |
| Ayaay | Filipino | Other Asian | Wong et al. |
| Ayabe | Japanese | Other Asian | Wong et al. |
| Ayala | Filipino | Other Asian | Wong et al. |
| Ayalde | Filipino | Other Asian | Wong et al. |
| Ayalin | Filipino | Other Asian | Wong et al. |
| Ayap | Filipino | Other Asian | Wong et al. |
| Ayapana | Filipino | Other Asian | Wong et al. |
| Ayawarpet |  | South Asian | Lim et al. |
| Aycardo | Filipino | Other Asian | Wong et al. |
| Aye | Burmese | Other Asian | Kranick et al. |
| Ayengar | Asian Indian | South Asian | Wong et al. |
| Ayento | Filipino | Other Asian | Wong et al. |
| Ayeras | Filipino | Other Asian | Wong et al. |
| Ayes | Filipino | Other Asian | Wong et al. |
| Aying | Filipino | Other Asian | Wong et al. |
| Ayonayon | Filipino | Other Asian | Wong et al. |
| Ayop | Filipino | Other Asian | Wong et al. |
| Ayoung |  | Other Asian | Kranick et al. |
| Ayran | Filipino | Other Asian | Wong et al. |
| Ayroso | Filipino | Other Asian | Wong et al. |
| Ayson | Filipino | Other Asian | Wong et al. |
| Aytch |  | South Asian | Lim et al. |
| Aytona | Filipino | Other Asian | Wong et al. |
| Ayub |  | South Asian | Lim et al. |
| Ayube |  | South Asian | Lim et al. |
| Ayukawa | Japanese | Other Asian | Wong et al. |
| Ayuson | Filipino | Other Asian | Wong et al. |
| Ayuyao | Filipino | Other Asian | Wong et al. |
| Ayyangar | Asian Indian | South Asian | Wong et al. |
| Ayyar | Asian Indian | South Asian | Wong et al. |
| Azad | Bangladeshi | South Asian | Kranick et al. |
| Azada | Filipino | Other Asian | Wong et al. |
| Azam | Asian Indian | South Asian | Wong et al. |
| Azama | Japanese | Other Asian | Wong et al. |
| Azarcon | Filipino | Other Asian | Wong et al. |
| Azares | Filipino | Other Asian | Wong et al. |
| Azariah | Asian Indian | South Asian | Wong et al. |
| Azarias | Filipino | Other Asian | Wong et al. |
| Azarraga | Filipino | Other Asian | Wong et al. |
| Azaula | Filipino | Other Asian | Wong et al. |
| Azcarate | Filipino | Other Asian | Wong et al. |
| Azcarraga | Filipino | Other Asian | Wong et al. |
| Azcona | Filipino | Other Asian | Wong et al. |
| Azcueta | Filipino | Other Asian | Wong et al. |
| Azeem | Asian Indian | South Asian | Wong et al. |
| Azeez | Asian Indian | South Asian | Wong et al. |
| Azegami | Japanese | Other Asian | Wong et al. |
| Azhar | Asian Indian | South Asian | Wong et al. |
| Azim | Bangladeshi | South Asian | Kranick et al. |
| Azimulla |  | South Asian | Lim et al. |
| Aziz | Pakistani | South Asian | Kranick et al. |
| Azizi |  | South Asian | Lim et al. |
| Azizuddin | Asian Indian | South Asian | Wong et al. |
| Azmi | Asian Indian | South Asian | Wong et al. |
| Aznar | Filipino | Other Asian | Wong et al. |
| Azores | Filipino | Other Asian | Wong et al. |
| Azucena | Filipino | Other Asian | Wong et al. |
| Azucenas | Filipino | Other Asian | Wong et al. |
| Azuelo | Filipino | Other Asian | Wong et al. |
| Azul | Filipino | Other Asian | Wong et al. |
| Azuma | Japanese | Other Asian | Wong et al. |
| Azumi | Japanese | Other Asian | Wong et al. |
| Azurin | Filipino | Other Asian | Wong et al. |
| Ba | Vietnamese | Other Asian | Wong et al. |
| Baal | Asian Indian; Filipino | South Asian | Kranick et al. |
| Baay | Filipino | Other Asian | Wong et al. |
| Baba | Japanese | Other Asian | Wong et al. |
| Babao | Filipino | Other Asian | Wong et al. |
| Babaran | Filipino | Other Asian | Wong et al. |
| Babas | Filipino | Other Asian | Wong et al. |
| Babasa | Filipino | Other Asian | Wong et al. |
| Babbar | Asian Indian | South Asian | Wong et al. |
| Babia | Filipino | Other Asian | Wong et al. |
| Babida | Filipino | Other Asian | Wong et al. |
| Babiera | Filipino | Other Asian | Wong et al. |
| Babila | Filipino | Other Asian | Wong et al. |
| Babilonia | Filipino | Other Asian | Wong et al. |
| Babista | Filipino | Other Asian | Wong et al. |
| Babor |  | South Asian | Lim et al. |
| Babu | Asian Indian | South Asian | Kranick et al. |
| Babudial |  | South Asian | Lim et al. |
| Babul | Asian Indian | South Asian | Wong et al. |
| Baburam |  | South Asian | Lim et al. |
| Baby | Asian Indian | South Asian | Wong et al. |
| Bac | Vietnamese | Other Asian | Wong et al. |
| Bacabac | Filipino | Other Asian | Wong et al. |
| Bacala | Filipino | Other Asian | Wong et al. |
| Bacalan | Filipino | Other Asian | Wong et al. |
| Bacaling | Filipino | Other Asian | Wong et al. |
| Bacalla | Filipino | Other Asian | Wong et al. |
| Bacalso | Filipino | Other Asian | Wong et al. |
| Bacaltos | Filipino | Other Asian | Wong et al. |
| Bacalzo | Filipino | Other Asian | Wong et al. |
| Bacani | Filipino | Other Asian | Wong et al. |
| Bacanto | Filipino | Other Asian | Wong et al. |
| Bacareza | Filipino | Other Asian | Wong et al. |
| Bacarisas | Filipino | Other Asian | Wong et al. |
| Bacarro | Filipino | Other Asian | Wong et al. |
| Bacatan | Filipino | Other Asian | Wong et al. |
| Bacay | Filipino | Other Asian | Wong et al. |
| Bacayo | Filipino | Other Asian | Wong et al. |
| Baccam | Vietnamese | Other Asian | Wong et al. |
| Baccay | Filipino | Other Asian | Wong et al. |
| Bacchus |  | South Asian | Lim et al. |
| Bacdayan | Filipino | Other Asian | Wong et al. |
| Bacerra | Filipino | Other Asian | Wong et al. |
| Bach | Vietnamese | Other Asian | Wong et al. |
| Bachan | Asian Indian | South Asian | Kranick et al. |
| Bachiller | Filipino | Other Asian | Wong et al. |
| Bacho | Filipino | Other Asian | Wong et al. |
| Bachoco | Filipino | Other Asian | Kranick et al. |
| Bachoo |  | South Asian | Lim et al. |
| Back | Korean | Other Asian | Kranick et al. |
| Baclagan | Filipino | Other Asian | Wong et al. |
| Baclay | Filipino | Other Asian | Wong et al. |
| Baclayon | Filipino | Other Asian | Wong et al. |
| Baclig | Filipino | Other Asian | Wong et al. |
| Baclit | Filipino | Other Asian | Wong et al. |
| Bacnat | Filipino | Other Asian | Wong et al. |
| Bacod | Filipino | Other Asian | Wong et al. |
| Bacol | Filipino | Other Asian | Wong et al. |
| Bacolod | Filipino | Other Asian | Wong et al. |
| Bacolor | Filipino | Other Asian | Wong et al. |
| Bacomo | Filipino | Other Asian | Wong et al. |
| Bacong | Filipino | Other Asian | Wong et al. |
| Bacos | Filipino | Other Asian | Wong et al. |
| Bacosa | Filipino | Other Asian | Wong et al. |
| Bactad | Filipino | Other Asian | Wong et al. |
| Bactat | Filipino | Other Asian | Wong et al. |
| Bactol | Filipino | Other Asian | Wong et al. |
| Bacud | Filipino | Other Asian | Wong et al. |
| Baculi | Filipino | Other Asian | Wong et al. |
| Bacungan | Filipino | Other Asian | Wong et al. |
| Bada | Filipino | Other Asian | Wong et al. |
| Badajos | Filipino | Other Asian | Wong et al. |
| Badanguio | Filipino | Other Asian | Wong et al. |
| Badani | Asian Indian | South Asian | Wong et al. |
| Badar | Pakistani | South Asian | Kranick et al. |
| Badat | Asian Indian | South Asian | Wong et al. |
| Badayos | Filipino | Other Asian | Wong et al. |
| Badeo | Filipino | Other Asian | Wong et al. |
| Badhan | Asian Indian | South Asian | Wong et al. |
| Badhesha | Asian Indian | South Asian | Wong et al. |
| Badhwar | Asian Indian | South Asian | Wong et al. |
| Badilla | Filipino | Other Asian | Wong et al. |
| Badillo | Filipino | Other Asian | Wong et al. |
| Badiola | Filipino | Other Asian | Wong et al. |
| Bado | Filipino | Other Asian | Wong et al. |
| Badong | Filipino | Other Asian | Wong et al. |
| Badoy | Filipino | Other Asian | Wong et al. |
| Badrina | Filipino | Other Asian | Wong et al. |
| Badruddin | Asian Indian | South Asian | Wong et al. |
| Badrul |  | South Asian | Lim et al. |
| Badsha | Asian Indian | South Asian | Wong et al. |
| Badshah | Asian Indian | South Asian | Wong et al. |
| Badua | Filipino | Other Asian | Wong et al. |
| Baduria | Filipino | Other Asian | Wong et al. |
| Badwal | Asian Indian | South Asian | Wong et al. |
| Badyal | Asian Indian | South Asian | Wong et al. |
| Bae | Korean | Other Asian | Wong et al. |
| Baeg | Korean | Other Asian | Wong et al. |
| Baek | Korean | Other Asian | Kranick et al. |
| Bael | Filipino | Other Asian | Wong et al. |
| Baello | Filipino | Other Asian | Wong et al. |
| Baes | Filipino | Other Asian | Wong et al. |
| Baesa | Filipino | Other Asian | Wong et al. |
| Baet | Filipino | Other Asian | Wong et al. |
| Baetiong | Filipino | Other Asian | Wong et al. |
| Baga | Filipino | Other Asian | Wong et al. |
| Bagabaldo | Filipino | Other Asian | Wong et al. |
| Bagadiong | Filipino | Other Asian | Wong et al. |
| Bagain | Filipino | Other Asian | Wong et al. |
| Bagaipo | Filipino | Other Asian | Wong et al. |
| Bagalawis | Filipino | Other Asian | Wong et al. |
| Bagalay | Filipino | Other Asian | Wong et al. |
| Bagalayos | Filipino | Other Asian | Wong et al. |
| Bagam |  | South Asian | Lim et al. |
| Bagaman | Chinese | Chinese | Wong et al. |
| Bagamaspad | Filipino | Other Asian | Wong et al. |
| Bagang | Filipino | Other Asian | Wong et al. |
| Bagano | Filipino | Other Asian | Wong et al. |
| Bagao | Filipino | Other Asian | Wong et al. |
| Bagaoisan | Filipino | Other Asian | Wong et al. |
| Bagaporo | Filipino | Other Asian | Wong et al. |
| Bagares | Filipino | Other Asian | Wong et al. |
| Bagas | Filipino | Other Asian | Wong et al. |
| Bagasan | Filipino | Other Asian | Wong et al. |
| Bagasol | Filipino | Other Asian | Wong et al. |
| Bagat | Filipino | Other Asian | Wong et al. |
| Bagaum |  | South Asian | Lim et al. |
| Bagay | Filipino | Other Asian | Wong et al. |
| Bagayas | Filipino | Other Asian | Wong et al. |
| Bagaybagayan | Filipino | Other Asian | Wong et al. |
| Bagcal | Filipino | Other Asian | Wong et al. |
| Bagchi | Asian Indian | South Asian | Wong et al. |
| Bagga | Asian Indian | South Asian | Kranick et al. |
| Baggao | Filipino | Other Asian | Wong et al. |
| Baggay | Filipino | Other Asian | Wong et al. |
| Bagha | Asian Indian | South Asian | Wong et al. |
| Bagilthaya |  | South Asian | Lim et al. |
| Bagnas | Filipino | Other Asian | Wong et al. |
| Bagnol | Filipino | Other Asian | Wong et al. |
| Bago | Filipino | Other Asian | Wong et al. |
| Bagon | Filipino | Other Asian | Wong et al. |
| Bagood | Filipino | Other Asian | Wong et al. |
| Bagorio | Filipino | Other Asian | Wong et al. |
| Bagos | Filipino | Other Asian | Wong et al. |
| Bagoyo | Filipino | Other Asian | Wong et al. |
| Bagri | Asian Indian | South Asian | Wong et al. |
| Bagsic | Filipino | Other Asian | Wong et al. |
| Bagtas | Filipino | Other Asian | Wong et al. |
| Bague | Filipino | Other Asian | Wong et al. |
| Baguindoc | Filipino | Other Asian | Wong et al. |
| Baguinon | Filipino | Other Asian | Wong et al. |
| Baguio | Filipino | Other Asian | Wong et al. |
| Baguisi | Filipino | Other Asian | Wong et al. |
| Bagum |  | South Asian | Lim et al. |
| Bagunas | Filipino | Other Asian | Wong et al. |
| Baguyo | Filipino | Other Asian | Wong et al. |
| Baguyos | Filipino | Other Asian | Wong et al. |
| Bagwan | Filipino | Other Asian | Wong et al. |
| Bah | Chinese | Chinese | Wong et al. |
| Bahador |  | South Asian | Lim et al. |
| Bahadur | Asian Indian | South Asian | Wong et al. |
| Bahar |  | South Asian | Lim et al. |
| Bahgat |  | South Asian | Lim et al. |
| Bahia | Filipino | Other Asian | Wong et al. |
| Bahk | Korean | Other Asian | Wong et al. |
| Bahng | Korean | Other Asian | Wong et al. |
| Bahri | Asian Indian | South Asian | Wong et al. |
| Bai | Asian Indian; Chinese | South Asian | Kranick et al. |
| Baichu |  | South Asian | Lim et al. |
| Baidwan | Asian Indian | South Asian | Wong et al. |
| Baidya |  | South Asian | Lim et al. |
| Baig | Pakistani | South Asian | Kranick et al. |
| Baijal | Asian Indian | South Asian | Wong et al. |
| Baijnauth |  | South Asian | Lim et al. |
| Baik | Korean | Other Asian | Wong et al. |
| Baillo | Filipino | Other Asian | Wong et al. |
| Bailon | Filipino | Other Asian | Wong et al. |
| Bains | Asian Indian | South Asian | Wong et al. |
| Bairan | Filipino | Other Asian | Wong et al. |
| Bais | Filipino | Other Asian | Wong et al. |
| Baisa | Filipino | Other Asian | Wong et al. |
| Baisas | Filipino | Other Asian | Wong et al. |
| Baitan | Filipino | Other Asian | Wong et al. |
| Baiza | Filipino | Other Asian | Wong et al. |
| Baja | Filipino | Other Asian | Wong et al. |
| Bajada | Filipino | Other Asian | Wong et al. |
| Bajado | Filipino | Other Asian | Wong et al. |
| Bajaj | Asian Indian | South Asian | Wong et al. |
| Bajamunde | Filipino | Other Asian | Wong et al. |
| Bajao | Filipino | Other Asian | Wong et al. |
| Bajar | Filipino | Other Asian | Wong et al. |
| Bajarias | Filipino | Other Asian | Wong et al. |
| Baje | Filipino | Other Asian | Wong et al. |
| Bajet | Filipino | Other Asian | Wong et al. |
| Bajit | Filipino | Other Asian | Wong et al. |
| Bajo | Filipino | Other Asian | Wong et al. |
| Bajpai | Asian Indian | South Asian | Wong et al. |
| Bajracharya |  | South Asian | Lim et al. |
| Bajwa |  | South Asian | Lim et al. |
| Bak | Korean | Other Asian | Wong et al. |
| Bakar |  | South Asian | Lim et al. |
| Baker |  | South Asian | Lim et al. |
| Bakhash | Asian Indian | South Asian | Wong et al. |
| Bakhshi | Asian Indian | South Asian | Wong et al. |
| Baki |  | South Asian | Lim et al. |
| Baksh | Asian Indian | South Asian | Kranick et al. |
| Bakshi | Asian Indian | South Asian | Kranick et al. |
| Bal | Asian Indian | South Asian | Wong et al. |
| Bala |  | South Asian | Lim et al. |
| Balaan | Filipino | Other Asian | Wong et al. |
| Balaba | Filipino | Other Asian | Wong et al. |
| Balabis | Filipino | Other Asian | Wong et al. |
| Balacano | Filipino | Other Asian | Wong et al. |
| Balachandran | Asian Indian | South Asian | Wong et al. |
| Balacuit | Filipino | Other Asian | Wong et al. |
| Baladad | Filipino | Other Asian | Wong et al. |
| Balaga | Filipino | Other Asian | Wong et al. |
| Balagat | Filipino | Other Asian | Wong et al. |
| Balagot | Filipino | Other Asian | Wong et al. |
| Balagot-Tabile | Filipino | Other Asian | Kranick et al. |
| Balagso | Filipino | Other Asian | Wong et al. |
| Balagtas | Filipino | Other Asian | Kranick et al. |
| Balaguer | Filipino | Other Asian | Wong et al. |
| Balahadia | Filipino | Other Asian | Wong et al. |
| Balais | Filipino | Other Asian | Wong et al. |
| Balajadia | Filipino | Other Asian | Wong et al. |
| Balakrishna | Asian Indian | South Asian | Wong et al. |
| Balakrishnan | Asian Indian | South Asian | Wong et al. |
| Balala | Filipino | Other Asian | Wong et al. |
| Balalio | Filipino | Other Asian | Wong et al. |
| Balallo | Filipino | Other Asian | Wong et al. |
| Balan | Filipino | Other Asian | Wong et al. |
| Balana | Filipino | Other Asian | Wong et al. |
| Balanag | Filipino | Other Asian | Wong et al. |
| Balanay | Filipino | Other Asian | Wong et al. |
| Balance | Filipino | Other Asian | Wong et al. |
| Balancio | Filipino | Other Asian | Wong et al. |
| Balandra | Filipino | Other Asian | Wong et al. |
| Balane | Filipino | Other Asian | Wong et al. |
| Balang | Filipino | Other Asian | Wong et al. |
| Balanga | Filipino | Other Asian | Wong et al. |
| Balangatan | Filipino | Other Asian | Wong et al. |
| Balangon | Filipino | Other Asian | Wong et al. |
| Balangue | Filipino | Other Asian | Wong et al. |
| Balanon | Filipino | Other Asian | Wong et al. |
| Balansag | Filipino | Other Asian | Wong et al. |
| Balansay | Filipino | Other Asian | Wong et al. |
| Balantac | Filipino | Other Asian | Wong et al. |
| Balanza | Filipino | Other Asian | Wong et al. |
| Balao | Filipino | Other Asian | Wong et al. |
| Balaoing | Filipino | Other Asian | Wong et al. |
| Balaoro | Filipino | Other Asian | Wong et al. |
| Balaram | Asian Indian | South Asian | Wong et al. |
| Balaraman | Asian Indian | South Asian | Wong et al. |
| Balasabas | Filipino | Other Asian | Wong et al. |
| Balase | Filipino | Other Asian | Wong et al. |
| Balasta | Filipino | Other Asian | Wong et al. |
| Balasubraman | Asian Indian | South Asian | Wong et al. |
| Balasundaram | Asian Indian | South Asian | Wong et al. |
| Balat | Filipino | Other Asian | Wong et al. |
| Balatayo | Filipino | Other Asian | Wong et al. |
| Balatbat | Filipino | Other Asian | Wong et al. |
| Balatero | Filipino | Other Asian | Wong et al. |
| Balatico | Filipino | Other Asian | Wong et al. |
| Balauro | Filipino | Other Asian | Wong et al. |
| Balay | Filipino | Other Asian | Wong et al. |
| Balayan | Filipino | Other Asian | Wong et al. |
| Balba | Filipino | Other Asian | Wong et al. |
| Balbag | Filipino | Other Asian | Wong et al. |
| Balbago | Filipino | Other Asian | Wong et al. |
| Balbas | Filipino | Other Asian | Wong et al. |
| Balbastro | Filipino | Other Asian | Wong et al. |
| Balbido | Filipino | Other Asian | Wong et al. |
| Balbin | Filipino | Other Asian | Wong et al. |
| Balboa | Filipino | Other Asian | Wong et al. |
| Balbuena | Filipino | Other Asian | Wong et al. |
| Balce | Filipino | Other Asian | Wong et al. |
| Balcita | Filipino | Other Asian | Wong et al. |
| Balcobero | Filipino | Other Asian | Wong et al. |
| Balcorta | Filipino | Other Asian | Wong et al. |
| Balcos | Filipino | Other Asian | Wong et al. |
| Balcueva | Filipino | Other Asian | Wong et al. |
| Baldado | Filipino | Other Asian | Wong et al. |
| Baldago | Filipino | Other Asian | Wong et al. |
| Baldazo | Filipino | Other Asian | Wong et al. |
| Baldemor | Filipino | Other Asian | Wong et al. |
| Baldemoro | Filipino | Other Asian | Wong et al. |
| Baldeo |  | South Asian | Lim et al. |
| Balderama | Filipino | Other Asian | Wong et al. |
| Balderas | Filipino | Other Asian | Wong et al. |
| Balderrama | Filipino | Other Asian | Wong et al. |
| Baldevia | Filipino | Other Asian | Wong et al. |
| Baldeviso | Filipino | Other Asian | Wong et al. |
| Baldia | Filipino | Other Asian | Wong et al. |
| Baldivia | Filipino | Other Asian | Wong et al. |
| Baldivino | Filipino | Other Asian | Wong et al. |
| Baldogo | Filipino | Other Asian | Wong et al. |
| Baldomero | Filipino | Other Asian | Wong et al. |
| Baldonado | Filipino | Other Asian | Wong et al. |
| Baldoria | Filipino | Other Asian | Wong et al. |
| Baldos | Filipino | Other Asian | Wong et al. |
| Baldovi | Filipino | Other Asian | Wong et al. |
| Baldovino | Filipino | Other Asian | Wong et al. |
| Baldoz | Filipino | Other Asian | Wong et al. |
| Baldoza | Filipino | Other Asian | Wong et al. |
| Baldueza | Filipino | Other Asian | Wong et al. |
| Baldugo | Filipino | Other Asian | Wong et al. |
| Balecha | Filipino | Other Asian | Wong et al. |
| Balela | Filipino | Other Asian | Wong et al. |
| Baleros | Filipino | Other Asian | Wong et al. |
| Balete | Filipino | Other Asian | Wong et al. |
| Baleva | Filipino | Other Asian | Wong et al. |
| Balgobin |  | South Asian | Lim et al. |
| Balgos | Filipino | Other Asian | Wong et al. |
| Bali | Asian Indian | South Asian | Wong et al. |
| Balicanta | Filipino | Other Asian | Wong et al. |
| Balicao | Filipino | Other Asian | Wong et al. |
| Balicat | Filipino | Other Asian | Wong et al. |
| Balico | Filipino | Other Asian | Wong et al. |
| Balicoco | Filipino | Other Asian | Wong et al. |
| Balidio | Filipino | Other Asian | Wong et al. |
| Baliga | Asian Indian | South Asian | Wong et al. |
| Baligad | Filipino | Other Asian | Wong et al. |
| Balignasay | Filipino | Other Asian | Wong et al. |
| Baligod | Filipino | Other Asian | Wong et al. |
| Balila | Filipino | Other Asian | Wong et al. |
| Balili | Filipino | Other Asian | Wong et al. |
| Baliling | Filipino | Other Asian | Wong et al. |
| Balilo | Filipino | Other Asian | Wong et al. |
| Balina | Filipino | Other Asian | Wong et al. |
| Balinao | Filipino | Other Asian | Wong et al. |
| Balinas | Filipino | Other Asian | Wong et al. |
| Balinbin | Filipino | Other Asian | Wong et al. |
| Balingcos | Filipino | Other Asian | Wong et al. |
| Balingit | Filipino | Other Asian | Wong et al. |
| Balino | Filipino | Other Asian | Wong et al. |
| Balintona | Filipino | Other Asian | Wong et al. |
| Baliola | Filipino | Other Asian | Wong et al. |
| Balisacan | Filipino | Other Asian | Wong et al. |
| Balisalisa | Filipino | Other Asian | Wong et al. |
| Balisi | Filipino | Other Asian | Wong et al. |
| Balista | Filipino | Other Asian | Wong et al. |
| Balita | Filipino | Other Asian | Wong et al. |
| Balitaan | Filipino | Other Asian | Wong et al. |
| Balite | Filipino | Other Asian | Wong et al. |
| Baliton | Filipino | Other Asian | Wong et al. |
| Baliwag | Filipino | Other Asian | Wong et al. |
| Baliwala | Asian Indian | South Asian | Wong et al. |
| Baliza | Filipino | Other Asian | Wong et al. |
| Ballal | Asian Indian | South Asian | Wong et al. |
| Ballan | Filipino | Other Asian | Wong et al. |
| Ballares | Filipino | Other Asian | Wong et al. |
| Ballarta | Filipino | Other Asian | Wong et al. |
| Ballecer | Filipino | Other Asian | Wong et al. |
| Ballelos | Filipino | Other Asian | Wong et al. |
| Ballena | Filipino | Other Asian | Wong et al. |
| Ballera | Filipino | Other Asian | Wong et al. |
| Balleras | Filipino | Other Asian | Wong et al. |
| Balles | Filipino | Other Asian | Wong et al. |
| Ballesta | Filipino | Other Asian | Wong et al. |
| Ballesteros | Filipino | Other Asian | Wong et al. |
| Ballete | Filipino | Other Asian | Wong et al. |
| Balleza | Filipino | Other Asian | Wong et al. |
| Ballocanag | Filipino | Other Asian | Wong et al. |
| Ballon | Filipino | Other Asian | Wong et al. |
| Ballungay | Filipino | Other Asian | Wong et al. |
| Balmaceda | Filipino | Other Asian | Wong et al. |
| Balmania | Filipino | Other Asian | Wong et al. |
| Balmeo | Filipino | Other Asian | Wong et al. |
| Balmes | Filipino | Other Asian | Wong et al. |
| Balmilero | Filipino | Other Asian | Wong et al. |
| Balmocena | Filipino | Other Asian | Wong et al. |
| Balmonte | Filipino | Other Asian | Wong et al. |
| Balmores | Filipino | Other Asian | Wong et al. |
| Balmori | Filipino | Other Asian | Wong et al. |
| Balneg | Filipino | Other Asian | Wong et al. |
| Baloaloa | Filipino | Other Asian | Wong et al. |
| Baloca | Filipino | Other Asian | Wong et al. |
| Baloch |  | South Asian | Lim et al. |
| Balois | Filipino | Other Asian | Wong et al. |
| Balolong | Filipino | Other Asian | Wong et al. |
| Balonzo | Filipino | Other Asian | Wong et al. |
| Baloran | Filipino | Other Asian | Wong et al. |
| Balot | Filipino | Other Asian | Wong et al. |
| Baloy | Filipino | Other Asian | Wong et al. |
| Balquiedra | Filipino | Other Asian | Wong et al. |
| Balraj | Asian Indian | South Asian | Wong et al. |
| Balsara | Asian Indian | South Asian | Wong et al. |
| Baltao | Filipino | Other Asian | Wong et al. |
| Baltazar | Filipino | Other Asian | Kranick et al. |
| Baltero | Filipino | Other Asian | Wong et al. |
| Balu | Asian Indian | South Asian | Wong et al. |
| Balubar | Filipino | Other Asian | Wong et al. |
| Balugay | Filipino | Other Asian | Wong et al. |
| Balugo | Filipino | Other Asian | Wong et al. |
| Balunsat | Filipino | Other Asian | Wong et al. |
| Baluran | Filipino | Other Asian | Wong et al. |
| Balusu | Asian Indian | South Asian | Wong et al. |
| Baluyot | Filipino | Other Asian | Wong et al. |
| Baluyut | Filipino | Other Asian | Wong et al. |
| Balverde | Filipino | Other Asian | Wong et al. |
| Balwani | Asian Indian | South Asian | Wong et al. |
| Bam | Asian Indian | South Asian | Wong et al. |
| Bamba | Filipino | Other Asian | Wong et al. |
| Bambalan | Filipino | Other Asian | Wong et al. |
| Bambao | Filipino | Other Asian | Wong et al. |
| Bambico | Filipino | Other Asian | Wong et al. |
| Bambo | Filipino | Other Asian | Wong et al. |
| Bammi |  | South Asian | Lim et al. |
| Bana | Filipino | Other Asian | Wong et al. |
| Banaag | Filipino | Other Asian | Wong et al. |
| Banaban | Filipino | Other Asian | Wong et al. |
| Banados | Filipino | Other Asian | Wong et al. |
| Banag | Filipino | Other Asian | Wong et al. |
| Banaga | Filipino | Other Asian | Wong et al. |
| Banaji | Asian Indian | South Asian | Wong et al. |
| Banania | Filipino | Other Asian | Wong et al. |
| Banares | Filipino | Other Asian | Wong et al. |
| Banaria | Filipino | Other Asian | Wong et al. |
| Banatao | Filipino | Other Asian | Wong et al. |
| Banatwala | Asian Indian | South Asian | Wong et al. |
| Banawa | Filipino | Other Asian | Wong et al. |
| Banay | Filipino | Other Asian | Wong et al. |
| Banayad | Filipino | Other Asian | Wong et al. |
| Banayat | Filipino | Other Asian | Wong et al. |
| Banayo | Filipino | Other Asian | Wong et al. |
| Bancod | Filipino | Other Asian | Wong et al. |
| Banda | Filipino | Other Asian | Wong et al. |
| Bandala | Filipino | Other Asian | Wong et al. |
| Bandalan | Filipino | Other Asian | Wong et al. |
| Bandales | Filipino | Other Asian | Wong et al. |
| Bandali | Asian Indian | South Asian | Wong et al. |
| Bandayrel | Filipino | Other Asian | Wong et al. |
| Bande | Filipino | Other Asian | Wong et al. |
| Bandelaria | Filipino | Other Asian | Wong et al. |
| Bandiola | Filipino | Other Asian | Wong et al. |
| Bando | Japanese | Other Asian | Wong et al. |
| Bandoma | Filipino | Other Asian | Wong et al. |
| Bandong | Filipino | Other Asian | Wong et al. |
| Bandoquillo | Filipino | Other Asian | Wong et al. |
| Bandoy | Filipino | Other Asian | Wong et al. |
| Bandugula | Asian Indian | South Asian | Kranick et al. |
| Bandyopadhya | Asian Indian | South Asian | Wong et al. |
| Banerjea | Asian Indian | South Asian | Wong et al. |
| Banerjee | Asian Indian | South Asian | Kranick et al. |
| Banerji | Asian Indian | South Asian | Wong et al. |
| Banez | Filipino | Other Asian | Wong et al. |
| Bang | Korean | Other Asian | Kranick et al. |
| Bangal | Filipino | Other Asian | Wong et al. |
| Bangalan | Filipino | Other Asian | Wong et al. |
| Bangalore | Asian Indian | South Asian | Wong et al. |
| Bangaru | Asian Indian | South Asian | Kranick et al. |
| Bangasan | Filipino | Other Asian | Wong et al. |
| Bangayan | Filipino | Other Asian | Wong et al. |
| Bangco | Filipino | Other Asian | Wong et al. |
| Bangera | Asian Indian | South Asian | Wong et al. |
| Bangi | Filipino | Other Asian | Wong et al. |
| Bangia | Asian Indian | South Asian | Wong et al. |
| Bangilan | Filipino | Other Asian | Wong et al. |
| Bangit | Filipino | Other Asian | Wong et al. |
| Bangloy | Filipino | Other Asian | Wong et al. |
| Bangot | Filipino | Other Asian | Wong et al. |
| Banh | Vietnamese | Other Asian | Wong et al. |
| Baniaga | Filipino | Other Asian | Wong et al. |
| Banico | Filipino | Other Asian | Wong et al. |
| Baniel | Filipino | Other Asian | Wong et al. |
| Banik |  | South Asian | Lim et al. |
| Baniqued | Filipino | Other Asian | Wong et al. |
| Bannai | Japanese | Other Asian | Wong et al. |
| Banno | Japanese | Other Asian | Wong et al. |
| Bano | Asian Indian | South Asian | Kranick et al. |
| Banog | Filipino | Other Asian | Wong et al. |
| Banogon | Filipino | Other Asian | Wong et al. |
| Bans | Asian Indian | South Asian | Wong et al. |
| Bansal | Asian Indian | South Asian | Kranick et al. |
| Bansi |  | South Asian | Lim et al. |
| Bansil | Filipino | Other Asian | Wong et al. |
| Bantay | Filipino | Other Asian | Wong et al. |
| Bantegui | Filipino | Other Asian | Wong et al. |
| Bantigue | Filipino | Other Asian | Wong et al. |
| Bantilan | Filipino | Other Asian | Wong et al. |
| Bantiles | Filipino | Other Asian | Wong et al. |
| Banting | Filipino | Other Asian | Wong et al. |
| Bantique | Filipino | Other Asian | Wong et al. |
| Bantista | Filipino | Other Asian | Wong et al. |
| Bantog | Filipino | Other Asian | Wong et al. |
| Bantolino | Filipino | Other Asian | Wong et al. |
| Bantug | Filipino | Other Asian | Wong et al. |
| Bantugan | Filipino | Other Asian | Wong et al. |
| Banu |  | South Asian | Lim et al. |
| Banua | Filipino | Other Asian | Wong et al. |
| Banut | Filipino | Other Asian | Wong et al. |
| Banwait | Asian Indian | South Asian | Wong et al. |
| Banzon | Filipino | Other Asian | Wong et al. |
| Banzuela | Filipino | Other Asian | Wong et al. |
| Banzuelo | Filipino | Other Asian | Wong et al. |
| Bao | Chinese | Chinese | Wong et al. |
| Baoit | Filipino | Other Asian | Wong et al. |
| Baon | Filipino | Other Asian | Wong et al. |
| Bapat | Asian Indian | South Asian | Wong et al. |
| Bapna | Asian Indian | South Asian | Wong et al. |
| Baptista | Filipino | Other Asian | Wong et al. |
| Baqai | Asian Indian | South Asian | Wong et al. |
| Baqar | Asian Indian | South Asian | Wong et al. |
| Baquer | Asian Indian | South Asian | Wong et al. |
| Baqui | Asian Indian | South Asian | Wong et al. |
| Baquial | Filipino | Other Asian | Wong et al. |
| Baquir | Filipino | Other Asian | Wong et al. |
| Baquiran | Filipino | Other Asian | Wong et al. |
| Baraan | Filipino | Other Asian | Wong et al. |
| Barabad | Filipino | Other Asian | Wong et al. |
| Baraceros | Filipino | Other Asian | Wong et al. |
| Barachina | Filipino | Other Asian | Wong et al. |
| Baradi | Filipino | Other Asian | Wong et al. |
| Barai | Asian Indian | South Asian | Wong et al. |
| Baraiya | Asian Indian | South Asian | Wong et al. |
| Baranda | Filipino | Other Asian | Wong et al. |
| Barangan | Filipino | Other Asian | Wong et al. |
| Barangay | Filipino | Other Asian | Wong et al. |
| Baraoidan | Filipino | Other Asian | Wong et al. |
| Baraquiel | Filipino | Other Asian | Kranick et al. |
| Barasona | Filipino | Other Asian | Kranick et al. |
| Barata | Filipino | Other Asian | Wong et al. |
| Barayuga | Filipino | Other Asian | Wong et al. |
| Barba | Filipino | Other Asian | Wong et al. |
| Barbadillo | Filipino | Other Asian | Wong et al. |
| Barbasa | Filipino | Other Asian | Wong et al. |
| Barbero | Filipino | Other Asian | Wong et al. |
| Barbers | Filipino | Other Asian | Wong et al. |
| Barbieto | Filipino | Other Asian | Wong et al. |
| Barbo | Filipino | Other Asian | Kranick et al. |
| Barbon | Filipino | Other Asian | Wong et al. |
| Barbosa | Filipino | Other Asian | Wong et al. |
| Barboza | Filipino | Other Asian | Wong et al. |
| Barcarse | Filipino | Other Asian | Wong et al. |
| Barce | Filipino | Other Asian | Wong et al. |
| Barcega | Filipino | Other Asian | Wong et al. |
| Barcellano | Filipino | Other Asian | Wong et al. |
| Barcelo | Filipino | Other Asian | Wong et al. |
| Barcelon | Filipino | Other Asian | Wong et al. |
| Barcelona | Filipino | Other Asian | Wong et al. |
| Barcena | Filipino | Other Asian | Wong et al. |
| Barcenas | Filipino | Other Asian | Wong et al. |
| Barcenilla | Filipino | Other Asian | Wong et al. |
| Barcinas | Filipino | Other Asian | Wong et al. |
| Bardhan | Asian Indian; Bangladeshi | South Asian | Kranick et al. |
| Barela | Filipino | Other Asian | Wong et al. |
| Bareng | Filipino | Other Asian | Wong et al. |
| Bargas | Filipino | Other Asian | Wong et al. |
| Bari |  | South Asian | Lim et al. |
| Baria | Filipino | Other Asian | Wong et al. |
| Barias | Filipino | Other Asian | Wong et al. |
| Barican | Filipino | Other Asian | Wong et al. |
| Baricaua | Filipino | Other Asian | Wong et al. |
| Baricuatro | Filipino | Other Asian | Wong et al. |
| Barientos | Filipino | Other Asian | Wong et al. |
| Barik | Asian Indian | South Asian | Wong et al. |
| Barikdar |  | South Asian | Lim et al. |
| Barillo | Filipino | Other Asian | Wong et al. |
| Barin | Filipino | Other Asian | Wong et al. |
| Baring | Filipino | Other Asian | Wong et al. |
| Barinque | Filipino | Other Asian | Wong et al. |
| Bariso | Filipino | Other Asian | Wong et al. |
| Barit | Filipino | Other Asian | Wong et al. |
| Bariuan | Filipino | Other Asian | Wong et al. |
| Barizo | Filipino | Other Asian | Wong et al. |
| Barlaan | Filipino | Other Asian | Wong et al. |
| Barlan | Filipino | Other Asian | Wong et al. |
| Barles | Filipino | Other Asian | Wong et al. |
| Barleta | Filipino | Other Asian | Wong et al. |
| Barlis | Filipino | Other Asian | Wong et al. |
| Barlongo | Filipino | Other Asian | Wong et al. |
| Barnabas | Asian Indian | South Asian | Wong et al. |
| Barnacha | Filipino | Other Asian | Wong et al. |
| Barnachea | Filipino | Other Asian | Wong et al. |
| Baro | Filipino | Other Asian | Wong et al. |
| Baroi |  | South Asian | Lim et al. |
| Barolia | Asian Indian | South Asian | Wong et al. |
| Barona | Filipino | Other Asian | Wong et al. |
| Barongan | Filipino | Other Asian | Wong et al. |
| Baronia | Filipino | Other Asian | Wong et al. |
| Barot | Asian Indian | South Asian | Wong et al. |
| Barraca | Filipino | Other Asian | Wong et al. |
| Barradas | Chinese | Chinese | Wong et al. |
| Barral | Filipino | Other Asian | Wong et al. |
| Barrameda | Filipino | Other Asian | Wong et al. |
| Barranda | Filipino | Other Asian | Wong et al. |
| Barrantes | Filipino | Other Asian | Wong et al. |
| Barreda | Filipino | Other Asian | Wong et al. |
| Barredo | Filipino | Other Asian | Wong et al. |
| Barrenechea | Filipino | Other Asian | Wong et al. |
| Barrera | Filipino | Other Asian | Wong et al. |
| Barreras | Filipino | Other Asian | Wong et al. |
| Barreto | Filipino | Other Asian | Wong et al. |
| Barretto | Filipino | Other Asian | Kranick et al. |
| Barria | Filipino | Other Asian | Wong et al. |
| Barrido | Filipino | Other Asian | Wong et al. |
| Barrientos | Filipino | Other Asian | Wong et al. |
| Barriga | Filipino | Other Asian | Wong et al. |
| Barrion | Filipino | Other Asian | Wong et al. |
| Barrios | Filipino | Other Asian | Wong et al. |
| Barro | Filipino | Other Asian | Wong et al. |
| Barroga | Filipino | Other Asian | Wong et al. |
| Barros | Filipino | Other Asian | Wong et al. |
| Barroso | Filipino | Other Asian | Wong et al. |
| Barrozo | Filipino | Other Asian | Wong et al. |
| Barruga | Filipino | Other Asian | Wong et al. |
| Barsana | Filipino | Other Asian | Wong et al. |
| Barsatan | Filipino | Other Asian | Wong et al. |
| Barte | Filipino | Other Asian | Wong et al. |
| Bartido | Filipino | Other Asian | Wong et al. |
| Bartolata | Filipino | Other Asian | Wong et al. |
| Bartolo | Filipino | Other Asian | Wong et al. |
| Bartolome | Filipino | Other Asian | Wong et al. |
| Barua |  | South Asian | Lim et al. |
| Baruah | Asian Indian | South Asian | Wong et al. |
| Baruela | Filipino | Other Asian | Wong et al. |
| Barukh | Asian Indian | South Asian | Wong et al. |
| Barut | Filipino | Other Asian | Wong et al. |
| Baruwal |  | South Asian | Lim et al. |
| Barve | Asian Indian | South Asian | Wong et al. |
| Bary |  | South Asian | Lim et al. |
| Barzaga | Filipino | Other Asian | Wong et al. |
| Bas | Filipino | Other Asian | Wong et al. |
| Basa | Filipino | Other Asian | Wong et al. |
| Basada | Filipino | Other Asian | Wong et al. |
| Basak | Asian Indian; Bangladeshi | South Asian | Kranick et al. |
| Basallo | Filipino | Other Asian | Wong et al. |
| Basalo | Filipino | Other Asian | Wong et al. |
| Basan | Filipino | Other Asian | Wong et al. |
| Basanes | Filipino | Other Asian | Wong et al. |
| Basappa | Asian Indian | South Asian | Wong et al. |
| Basas | Filipino | Other Asian | Wong et al. |
| Basat | Filipino | Other Asian | Wong et al. |
| Basbas | Filipino | Other Asian | Wong et al. |
| Bascao | Filipino | Other Asian | Wong et al. |
| Bascara | Filipino | Other Asian | Wong et al. |
| Basco | Filipino | Other Asian | Wong et al. |
| Bascon | Filipino | Other Asian | Wong et al. |
| Basconcillo | Filipino | Other Asian | Wong et al. |
| Bascos | Filipino | Other Asian | Wong et al. |
| Basdeo |  | South Asian | Lim et al. |
| Basea | Filipino | Other Asian | Wong et al. |
| Bashar | Bangladeshi | South Asian | Kranick et al. |
| Basher |  | South Asian | Lim et al. |
| Bashir |  | South Asian | Lim et al. |
| Bashire |  | South Asian | Lim et al. |
| Bashiruddin | Asian Indian | South Asian | Wong et al. |
| Basi | Asian Indian | South Asian | Wong et al. |
| Basila | Filipino | Other Asian | Wong et al. |
| Basilan | Filipino | Other Asian | Wong et al. |
| Basilio | Filipino | Other Asian | Kranick et al. |
| Basith | Saudi Arabia; Asian Indian | South Asian | Kranick et al. |
| Basmayor | Filipino | Other Asian | Wong et al. |
| Basnet |  | South Asian | Lim et al. |
| Basobas | Filipino | Other Asian | Wong et al. |
| Basquez | Filipino | Other Asian | Wong et al. |
| Basra | Asian Indian | South Asian | Wong et al. |
| Basrai | Asian Indian | South Asian | Wong et al. |
| Basran | Asian Indian | South Asian | Wong et al. |
| Bassig | Filipino | Other Asian | Wong et al. |
| Bassit |  | South Asian | Lim et al. |
| Bastes | Filipino | Other Asian | Wong et al. |
| Basto | Filipino | Other Asian | Wong et al. |
| Basu | Asian Indian | South Asian | Wong et al. |
| Basubas | Filipino | Other Asian | Wong et al. |
| Basuel | Filipino | Other Asian | Wong et al. |
| Basug | Filipino | Other Asian | Wong et al. |
| Basuil | Filipino | Other Asian | Wong et al. |
| Bat | Vietnamese | Other Asian | Wong et al. |
| Batac | Filipino | Other Asian | Wong et al. |
| Batacan | Filipino | Other Asian | Wong et al. |
| Bataclan | Filipino | Other Asian | Wong et al. |
| Batad | Filipino | Other Asian | Wong et al. |
| Batalla | Filipino | Other Asian | Wong et al. |
| Bataller | Filipino | Other Asian | Kranick et al. |
| Batallones | Filipino | Other Asian | Wong et al. |
| Batalon | Filipino | Other Asian | Wong et al. |
| Batan | Filipino | Other Asian | Wong et al. |
| Batang | Filipino | Other Asian | Wong et al. |
| Batangan | Filipino | Other Asian | Wong et al. |
| Batara | Filipino | Other Asian | Wong et al. |
| Batarao | Filipino | Other Asian | Wong et al. |
| Batarina | Filipino | Other Asian | Wong et al. |
| Batario | Filipino | Other Asian | Wong et al. |
| Batas | Filipino | Other Asian | Wong et al. |
| Batayola | Filipino | Other Asian | Wong et al. |
| Batchoo |  | South Asian | Lim et al. |
| Baten |  | South Asian | Lim et al. |
| Batenga | Filipino | Other Asian | Wong et al. |
| Baterina | Filipino | Other Asian | Wong et al. |
| Baterna | Filipino | Other Asian | Wong et al. |
| Bathan | Filipino | Other Asian | Wong et al. |
| Batheja | Asian Indian | South Asian | Wong et al. |
| Bathla | Asian Indian | South Asian | Wong et al. |
| Bati | Filipino | Other Asian | Wong et al. |
| Batiller | Filipino | Other Asian | Wong et al. |
| Batilo | Filipino | Other Asian | Wong et al. |
| Batin | Filipino | Other Asian | Wong et al. |
| Batinga | Filipino | Other Asian | Wong et al. |
| Batingal | Filipino | Other Asian | Wong et al. |
| Batingan | Filipino | Other Asian | Wong et al. |
| Batino | Filipino | Other Asian | Wong et al. |
| Bation | Filipino | Other Asian | Wong et al. |
| Batiquin | Filipino | Other Asian | Wong et al. |
| Batista | Filipino | Other Asian | Wong et al. |
| Batliwala | Asian Indian | South Asian | Wong et al. |
| Bato | Filipino | Other Asian | Wong et al. |
| Batocabe | Filipino | Other Asian | Wong et al. |
| Batol | Filipino | Other Asian | Wong et al. |
| Batongbacal | Filipino | Other Asian | Wong et al. |
| Batoon | Filipino | Other Asian | Wong et al. |
| Battad | Filipino | Other Asian | Wong et al. |
| Batth | Asian Indian | South Asian | Wong et al. |
| Battung | Filipino | Other Asian | Wong et al. |
| Batucan | Filipino | Other Asian | Wong et al. |
| Batugal | Filipino | Other Asian | Wong et al. |
| Batugo | Filipino | Other Asian | Wong et al. |
| Batuhan | Filipino | Other Asian | Wong et al. |
| Batuigas | Filipino | Other Asian | Wong et al. |
| Batulan | Filipino | Other Asian | Wong et al. |
| Batungbacal | Filipino | Other Asian | Wong et al. |
| Batuyong | Filipino | Other Asian | Wong et al. |
| Bau | Chinese | Chinese | Wong et al. |
| Baula | Filipino | Other Asian | Wong et al. |
| Bausa | Filipino | Other Asian | Wong et al. |
| Bauson | Filipino | Other Asian | Wong et al. |
| Baustista | Filipino | Other Asian | Wong et al. |
| Bautista | Filipino | Other Asian | Wong et al. |
| Bautro | Filipino | Other Asian | Wong et al. |
| Bauzon | Filipino | Other Asian | Wong et al. |
| Bavishi | Asian Indian | South Asian | Wong et al. |
| Baw | Chinese | Chinese | Wong et al. |
| Bawa | Asian Indian | South Asian | Wong et al. |
| Bawalan | Filipino | Other Asian | Wong et al. |
| Bawany | Asian Indian | South Asian | Wong et al. |
| Baxa | Filipino | Other Asian | Wong et al. |
| Baxi | Asian Indian | South Asian | Kranick et al. |
| Baya | Filipino | Other Asian | Wong et al. |
| Bayaca | Filipino | Other Asian | Wong et al. |
| Bayacal | Filipino | Other Asian | Wong et al. |
| Bayag | Filipino | Other Asian | Wong et al. |
| Bayan | Filipino | Other Asian | Wong et al. |
| Bayang | Filipino | Other Asian | Wong et al. |
| Bayangos | Filipino | Other Asian | Wong et al. |
| Bayani | Filipino | Other Asian | Wong et al. |
| Bayas | Filipino | Other Asian | Wong et al. |
| Bayaua | Filipino | Other Asian | Wong et al. |
| Baybado | Filipino | Other Asian | Wong et al. |
| Baybay | Filipino | Other Asian | Wong et al. |
| Baybayan | Filipino | Other Asian | Wong et al. |
| Bayla | Filipino | Other Asian | Wong et al. |
| Bayle | Filipino | Other Asian | Wong et al. |
| Baylen | Filipino | Other Asian | Wong et al. |
| Baylon | Filipino | Other Asian | Wong et al. |
| Baylosis | Filipino | Other Asian | Wong et al. |
| Bayna | Filipino | Other Asian | Wong et al. |
| Bayog | Filipino | Other Asian | Wong et al. |
| Bayona | Filipino | Other Asian | Wong et al. |
| Bayoneta | Filipino | Other Asian | Wong et al. |
| Bayoneto | Filipino | Other Asian | Kranick et al. |
| Bayot | Filipino | Other Asian | Wong et al. |
| Bayquen | Filipino | Other Asian | Wong et al. |
| Bayron | Filipino | Other Asian | Wong et al. |
| Baysa | Filipino | Other Asian | Wong et al. |
| Baysic | Filipino | Other Asian | Wong et al. |
| Bayudan | Filipino | Other Asian | Wong et al. |
| Bayudang | Filipino | Other Asian | Wong et al. |
| Bayuga | Filipino | Other Asian | Wong et al. |
| Bayungan | Filipino | Other Asian | Wong et al. |
| Bayunus | Pakistani | South Asian | Kranick et al. |
| Bayya | Filipino | Other Asian | Wong et al. |
| Baz | Korean | Other Asian | Wong et al. |
| Bazaz | Asian Indian | South Asian | Wong et al. |
| Bazlu |  | South Asian | Lim et al. |
| Bea | Korean | Other Asian | Wong et al. |
| Beato | Filipino | Other Asian | Wong et al. |
| Bebing | Filipino | Other Asian | Wong et al. |
| Becerra | Chinese | Chinese | Wong et al. |
| Becharbhai | Asian Indian | South Asian | Wong et al. |
| Bechulal |  | South Asian | Lim et al. |
| Bedania | Filipino | Other Asian | Wong et al. |
| Bederio | Filipino | Other Asian | Wong et al. |
| Bedi | Asian Indian | South Asian | Wong et al. |
| Bedia | Filipino | Other Asian | Wong et al. |
| Bedico | Filipino | Other Asian | Wong et al. |
| Beer |  | South Asian | Lim et al. |
| Beg | Asian Indian | South Asian | Wong et al. |
| Begam |  | South Asian | Lim et al. |
| Begom |  | South Asian | Lim et al. |
| Begonia | Filipino | Other Asian | Wong et al. |
| Begum | Asian Indian; Bangladeshi | South Asian | Kranick et al. |
| Begume |  | South Asian | Lim et al. |
| Behal | Asian Indian | South Asian | Wong et al. |
| Beharry |  | South Asian | Lim et al. |
| Behera | Asian Indian | South Asian | Wong et al. |
| Bei | Chinese | Chinese | Wong et al. |
| Beig | Asian Indian | South Asian | Wong et al. |
| Bejar | Filipino | Other Asian | Wong et al. |
| Bejarin | Filipino | Other Asian | Wong et al. |
| Bejasa | Filipino | Other Asian | Wong et al. |
| Belal |  | South Asian | Lim et al. |
| Belamide | Filipino | Other Asian | Wong et al. |
| Belandres | Filipino | Other Asian | Wong et al. |
| Belangue | Filipino | Other Asian | Wong et al. |
| Belarde | Filipino | Other Asian | Wong et al. |
| Belardo | Filipino | Other Asian | Wong et al. |
| Belarmino | Filipino | Other Asian | Wong et al. |
| Belayet |  | South Asian | Lim et al. |
| Belda | Filipino | Other Asian | Wong et al. |
| Belderol | Filipino | Other Asian | Wong et al. |
| Belen | Filipino | Other Asian | Kranick et al. |
| Belena | Filipino | Other Asian | Wong et al. |
| Beleno | Filipino | Other Asian | Wong et al. |
| Belga | Filipino | Other Asian | Wong et al. |
| Belgado | Filipino | Other Asian | Wong et al. |
| Belino | Filipino | Other Asian | Wong et al. |
| Belisario |  | South Asian | Lim et al. |
| Beliso | Filipino | Other Asian | Wong et al. |
| Belizario | Filipino | Other Asian | Wong et al. |
| Belleza | Filipino | Other Asian | Wong et al. |
| Bello | Filipino | Other Asian | Wong et al. |
| Bellosillo | Filipino | Other Asian | Wong et al. |
| Belmes | Filipino | Other Asian | Wong et al. |
| Belmonte | Filipino | Other Asian | Wong et al. |
| Belnas | Filipino | Other Asian | Wong et al. |
| Belo | Filipino | Other Asian | Wong et al. |
| Belocura | Filipino | Other Asian | Wong et al. |
| Beloso | Filipino | Other Asian | Wong et al. |
| Belostrino | Filipino | Other Asian | Wong et al. |
| Beloy | Filipino | Other Asian | Wong et al. |
| Beltran | Filipino | Other Asian | Wong et al. |
| Beltrano | Filipino | Other Asian | Wong et al. |
| Beluso | Filipino | Other Asian | Wong et al. |
| Belvis | Filipino | Other Asian | Wong et al. |
| Belza | Filipino | Other Asian | Wong et al. |
| Belzunce | Filipino | Other Asian | Wong et al. |
| Bembo | Filipino | Other Asian | Wong et al. |
| Ben | Chinese | Chinese | Wong et al. |
| Benabese | Filipino | Other Asian | Wong et al. |
| Benavides | Filipino | Other Asian | Wong et al. |
| Benavidez | Filipino | Other Asian | Wong et al. |
| Bencio | Filipino | Other Asian | Wong et al. |
| Bencito | Filipino | Other Asian | Wong et al. |
| Bendana | Filipino | Other Asian | Wong et al. |
| Bendanillo | Filipino | Other Asian | Wong et al. |
| Bendijo | Filipino | Other Asian | Wong et al. |
| Bendo | Filipino | Other Asian | Wong et al. |
| Bendoy | Filipino | Other Asian | Wong et al. |
| Benedicto | Filipino | Other Asian | Kranick et al. |
| Benedictos | Filipino | Other Asian | Wong et al. |
| Benedito | Filipino | Other Asian | Wong et al. |
| Benemerito | Filipino | Other Asian | Wong et al. |
| Benesa | Filipino | Other Asian | Wong et al. |
| Benetua | Filipino | Other Asian | Wong et al. |
| Bengco | Filipino | Other Asian | Wong et al. |
| Bengzon | Filipino | Other Asian | Wong et al. |
| Benigno | Filipino | Other Asian | Wong et al. |
| Benipayo | Filipino | Other Asian | Wong et al. |
| Benitez | Filipino | Other Asian | Wong et al. |
| Benito | Filipino | Other Asian | Wong et al. |
| Benjamin |  | South Asian | Lim et al. |
| Benliro | Filipino | Other Asian | Wong et al. |
| Benoza | Filipino | Other Asian | Wong et al. |
| Bensan | Filipino | Other Asian | Wong et al. |
| Bentajado | Filipino | Other Asian | Wong et al. |
| Benzon | Filipino | Other Asian | Wong et al. |
| Beppu | Japanese | Other Asian | Wong et al. |
| Bequilla | Filipino | Other Asian | Wong et al. |
| Beralde | Filipino | Other Asian | Wong et al. |
| Berame | Filipino | Other Asian | Wong et al. |
| Berango | Filipino | Other Asian | Wong et al. |
| Berba | Filipino | Other Asian | Wong et al. |
| Berbano | Filipino | Other Asian | Wong et al. |
| Berberabe | Filipino | Other Asian | Wong et al. |
| Bercasio | Filipino | Other Asian | Wong et al. |
| Berces | Filipino | Other Asian | Wong et al. |
| Berdal | Filipino | Other Asian | Wong et al. |
| Berdiago | Filipino | Other Asian | Wong et al. |
| Berdin | Filipino | Other Asian | Wong et al. |
| Berdon | Filipino | Other Asian | Wong et al. |
| Berdos | Filipino | Other Asian | Wong et al. |
| Beredo | Filipino | Other Asian | Wong et al. |
| Berena | Filipino | Other Asian | Wong et al. |
| Berenguel | Filipino | Other Asian | Wong et al. |
| Berenguer | Filipino | Other Asian | Wong et al. |
| Bergado | Filipino | Other Asian | Wong et al. |
| Berganio | Filipino | Other Asian | Wong et al. |
| Bergantinos | Filipino | Other Asian | Wong et al. |
| Bergonia | Filipino | Other Asian | Wong et al. |
| Bergonio | Filipino | Other Asian | Wong et al. |
| Beri | Asian Indian | South Asian | Wong et al. |
| Berina | Filipino | Other Asian | Wong et al. |
| Beringuel | Filipino | Other Asian | Wong et al. |
| Beriones | Filipino | Other Asian | Wong et al. |
| Berioso | Filipino | Other Asian | Wong et al. |
| Berja | Filipino | Other Asian | Wong et al. |
| Berlanga | Filipino | Other Asian | Wong et al. |
| Bermas | Filipino | Other Asian | Wong et al. |
| Bermejo | Filipino | Other Asian | Wong et al. |
| Bermillo | Filipino | Other Asian | Wong et al. |
| Bermio | Filipino | Other Asian | Wong et al. |
| Bermoy | Filipino | Other Asian | Wong et al. |
| Bermudes | Filipino | Other Asian | Wong et al. |
| Bermudez | Filipino | Other Asian | Wong et al. |
| Bermudo | Filipino | Other Asian | Wong et al. |
| Bermundo | Filipino | Other Asian | Wong et al. |
| Bernabe | Filipino | Other Asian | Wong et al. |
| Bernadas | Filipino | Other Asian | Wong et al. |
| Bernados | Filipino | Other Asian | Wong et al. |
| Bernal | Filipino | Other Asian | Wong et al. |
| Bernaldes | Filipino | Other Asian | Wong et al. |
| Bernaldez | Filipino | Other Asian | Wong et al. |
| Bernaldo | Filipino | Other Asian | Wong et al. |
| Bernales | Filipino | Other Asian | Wong et al. |
| Bernandino | Filipino | Other Asian | Wong et al. |
| Bernardez | Filipino | Other Asian | Wong et al. |
| Bernardino | Filipino | Other Asian | Wong et al. |
| Bernardo | Filipino | Other Asian | Wong et al. |
| Bernarte | Filipino | Other Asian | Wong et al. |
| Bernil | Filipino | Other Asian | Wong et al. |
| Bernus | Filipino | Other Asian | Wong et al. |
| Berona | Filipino | Other Asian | Wong et al. |
| Beronilla | Filipino | Other Asian | Wong et al. |
| Berroya | Filipino | Other Asian | Wong et al. |
| Bersabal | Filipino | Other Asian | Wong et al. |
| Bersabe | Filipino | Other Asian | Wong et al. |
| Bersalona | Filipino | Other Asian | Wong et al. |
| Bersamin | Filipino | Other Asian | Wong et al. |
| Bersamina | Filipino | Other Asian | Wong et al. |
| Bersamira | Filipino | Other Asian | Wong et al. |
| Bersola | Filipino | Other Asian | Wong et al. |
| Bertubin | Filipino | Other Asian | Wong et al. |
| Bertulfo | Filipino | Other Asian | Wong et al. |
| Bertumen | Filipino | Other Asian | Wong et al. |
| Berzamina | Filipino | Other Asian | Wong et al. |
| Besa | Filipino | Other Asian | Wong et al. |
| Besana | Filipino | Other Asian | Wong et al. |
| Besas | Filipino | Other Asian | Wong et al. |
| Bessho | Japanese | Other Asian | Wong et al. |
| Betco | Filipino | Other Asian | Wong et al. |
| Betines | Chinese | Chinese | Wong et al. |
| Betita | Filipino | Other Asian | Wong et al. |
| Betonio | Filipino | Other Asian | Wong et al. |
| Beza | Filipino | Other Asian | Wong et al. |
| Bguyen | Vietnamese | Other Asian | Wong et al. |
| Bhadra | Asian Indian | South Asian | Wong et al. |
| Bhaduri | Asian Indian | South Asian | Wong et al. |
| Bhagabhai | Asian Indian | South Asian | Wong et al. |
| Bhagat | Asian Indian | South Asian | Wong et al. |
| Bhagavatula | Asian Indian | South Asian | Wong et al. |
| Bhagwan | Asian Indian | South Asian | Wong et al. |
| Bhagwanani | Asian Indian | South Asian | Wong et al. |
| Bhagwandas | Asian Indian | South Asian | Wong et al. |
| Bhagwanji | Asian Indian | South Asian | Wong et al. |
| Bhagwat | Asian Indian | South Asian | Wong et al. |
| Bhai | Asian Indian | South Asian | Wong et al. |
| Bhaidani | Asian Indian | South Asian | Wong et al. |
| Bhailalbhai | Asian Indian | South Asian | Wong et al. |
| Bhaipatel | Asian Indian | South Asian | Wong et al. |
| Bhaiyat | Asian Indian | South Asian | Wong et al. |
| Bhakta | Asian Indian | South Asian | Wong et al. |
| Bhala | Asian Indian | South Asian | Wong et al. |
| Bhalala | Asian Indian | South Asian | Wong et al. |
| Bhalani | Asian Indian | South Asian | Wong et al. |
| Bhalerao | Asian Indian | South Asian | Wong et al. |
| Bhalla | Asian Indian | South Asian | Wong et al. |
| Bhalodia | Asian Indian | South Asian | Wong et al. |
| Bhamani | Asian Indian | South Asian | Wong et al. |
| Bhambhani | Asian Indian | South Asian | Kranick et al. |
| Bhambri | Asian Indian | South Asian | Kranick et al. |
| Bhamra | Asian Indian | South Asian | Wong et al. |
| Bhan | Asian Indian | South Asian | Wong et al. |
| Bhana | Asian Indian | South Asian | Wong et al. |
| Bhandal | Asian Indian | South Asian | Wong et al. |
| Bhandari | Asian Indian | South Asian | Wong et al. |
| Bhandarkar | Asian Indian | South Asian | Wong et al. |
| Bhanderi | Asian Indian | South Asian | Wong et al. |
| Bhang | Korean | Other Asian | Wong et al. |
| Bhangal | Asian Indian | South Asian | Wong et al. |
| Bhangoo | Asian Indian | South Asian | Wong et al. |
| Bhangu | Asian Indian | South Asian | Wong et al. |
| Bhanji | Asian Indian | South Asian | Wong et al. |
| Bhanot | Asian Indian | South Asian | Wong et al. |
| Bhansali | Asian Indian | South Asian | Wong et al. |
| Bhar | Asian Indian | South Asian | Wong et al. |
| Bharadwaj | Asian Indian | South Asian | Wong et al. |
| Bharat |  | South Asian | Lim et al. |
| Bhardwaj | Asian Indian | South Asian | Kranick et al. |
| Bhargava | Asian Indian | South Asian | Wong et al. |
| Bharmal | Asian Indian | South Asian | Wong et al. |
| Bharrat |  | South Asian | Lim et al. |
| Bharucha | Asian Indian | South Asian | Wong et al. |
| Bhaskar | Asian Indian | South Asian | Wong et al. |
| Bhaskaran | Asian Indian | South Asian | Wong et al. |
| Bhasker | Asian Indian | South Asian | Wong et al. |
| Bhat | Asian Indian | South Asian | Wong et al. |
| Bhate | Asian Indian | South Asian | Wong et al. |
| Bhathal | Asian Indian | South Asian | Wong et al. |
| Bhati | Asian Indian | South Asian | Wong et al. |
| Bhatia | Asian Indian | South Asian | Wong et al. |
| Bhatji |  | South Asian | Lim et al. |
| Bhatnagar | Asian Indian | South Asian | Kranick et al. |
| Bhatt | Asian Indian | South Asian | Kranick et al. |
| Bhattacharje | Asian Indian | South Asian | Wong et al. |
| Bhattacharjee |  | South Asian | Lim et al. |
| Bhattacharji | Asian Indian | South Asian | Wong et al. |
| Bhattacharya | Asian Indian | South Asian | Wong et al. |
| Bhattacharyy | Asian Indian | South Asian | Wong et al. |
| Bhattacherje | Asian Indian | South Asian | Wong et al. |
| Bhattal | Asian Indian | South Asian | Wong et al. |
| Bhattarai |  | South Asian | Lim et al. |
| Bhatti | Pakistani | South Asian | Kranick et al. |
| Bhave | Asian Indian | South Asian | Wong et al. |
| Bhavsar | Asian Indian | South Asian | Wong et al. |
| Bhayana | Asian Indian | South Asian | Wong et al. |
| Bhayani | Asian Indian | South Asian | Wong et al. |
| Bhide | Asian Indian | South Asian | Wong et al. |
| Bhikhabhai | Asian Indian | South Asian | Wong et al. |
| Bhim | Asian Indian | South Asian | Wong et al. |
| Bhimani | Asian Indian | South Asian | Wong et al. |
| Bhimji | Asian Indian | South Asian | Wong et al. |
| Bhinder | Asian Indian | South Asian | Wong et al. |
| Bhise | Asian Indian | South Asian | Wong et al. |
| Bhogal | Asian Indian | South Asian | Wong et al. |
| Bhogilal | Asian Indian | South Asian | Wong et al. |
| Bhojani | Asian Indian | South Asian | Wong et al. |
| Bhola | Asian Indian | South Asian | Kranick et al. |
| Bhole | Asian Indian | South Asian | Wong et al. |
| Bhowmik | Asian Indian | South Asian | Wong et al. |
| Bhuian | Bangladeshi; Asian Indian | South Asian | Kranick et al. |
| Bhuiya |  | South Asian | Lim et al. |
| Bhuiyan | Bangladeshi; Asian Indian | South Asian | Kranick et al. |
| Bhukhanwala | Asian Indian | South Asian | Wong et al. |
| Bhula | Asian Indian | South Asian | Wong et al. |
| Bhulabhai | Asian Indian | South Asian | Wong et al. |
| Bhullar | Asian Indian | South Asian | Wong et al. |
| Bhusari | Asian Indian | South Asian | Wong et al. |
| Bhushan | Asian Indian | South Asian | Wong et al. |
| Bhut | Asian Indian | South Asian | Wong et al. |
| Bhuta | Asian Indian | South Asian | Wong et al. |
| Bhuva | Asian Indian | South Asian | Wong et al. |
| Bhuyan |  | South Asian | Lim et al. |
| Bhyan |  | South Asian | Lim et al. |
| Bi | Chinese | Chinese | Wong et al. |
| Bia |  | South Asian | Lim et al. |
| Biado | Filipino | Other Asian | Wong et al. |
| Biag | Filipino | Other Asian | Wong et al. |
| Biagtan | Filipino | Other Asian | Wong et al. |
| Biala | Filipino | Other Asian | Wong et al. |
| Bian | Chinese | Chinese | Wong et al. |
| Bianes | Filipino | Other Asian | Wong et al. |
| Biascan | Filipino | Other Asian | Wong et al. |
| Biason | Filipino | Other Asian | Wong et al. |
| Biazon | Filipino | Other Asian | Wong et al. |
| Bibat | Filipino | Other Asian | Wong et al. |
| Bibay | Filipino | Other Asian | Wong et al. |
| Bibera | Filipino | Other Asian | Wong et al. |
| Bibi | Asian Indian | South Asian | Kranick et al. |
| Bicera | Filipino | Other Asian | Wong et al. |
| Bich | Vietnamese | Other Asian | Wong et al. |
| Bicomong | Filipino | Other Asian | Wong et al. |
| Bides | Filipino | Other Asian | Wong et al. |
| Bie | Filipino | Other Asian | Wong et al. |
| Biete | Filipino | Other Asian | Wong et al. |
| Bieu | Vietnamese | Other Asian | Wong et al. |
| Big | Chinese | Chinese | Wong et al. |
| Bigalbal | Filipino | Other Asian | Wong et al. |
| Bigay | Filipino | Other Asian | Wong et al. |
| Bigcas | Filipino | Other Asian | Wong et al. |
| Biglangawa | Filipino | Other Asian | Wong et al. |
| Biglete | Filipino | Other Asian | Wong et al. |
| Bigornia | Filipino | Other Asian | Wong et al. |
| Bih | Chinese | Chinese | Wong et al. |
| Bihag | Filipino | Other Asian | Wong et al. |
| Bihis | Filipino | Other Asian | Wong et al. |
| Bijasa | Filipino | Other Asian | Wong et al. |
| Bilal | Pakistani | South Asian | Kranick et al. |
| Bilan | Filipino | Other Asian | Wong et al. |
| Bilbaeno | Filipino | Other Asian | Wong et al. |
| Bilbao | Filipino | Other Asian | Wong et al. |
| Bilen | Filipino | Other Asian | Wong et al. |
| Bilgera | Filipino | Other Asian | Wong et al. |
| Bilgrami | Asian Indian | South Asian | Wong et al. |
| Bilimoria | Asian Indian | South Asian | Wong et al. |
| Bilkis |  | South Asian | Lim et al. |
| Billah |  | South Asian | Lim et al. |
| Billedo | Filipino | Other Asian | Wong et al. |
| Billena | Filipino | Other Asian | Wong et al. |
| Billimoria | Asian Indian | South Asian | Wong et al. |
| Billones | Filipino | Other Asian | Wong et al. |
| Billoo | Pakistani | South Asian | Kranick et al. |
| Billote | Filipino | Other Asian | Wong et al. |
| Bilog | Filipino | Other Asian | Wong et al. |
| Binalinbing | Filipino | Other Asian | Wong et al. |
| Binarao | Filipino | Other Asian | Wong et al. |
| Binas | Filipino | Other Asian | Wong et al. |
| Binay | Filipino | Other Asian | Wong et al. |
| Bince | Filipino | Other Asian | Wong et al. |
| Bindra | Asian Indian | South Asian | Wong et al. |
| Bingcang | Filipino | Other Asian | Wong et al. |
| Binh | Vietnamese | Other Asian | Wong et al. |
| Binondo | Filipino | Other Asian | Wong et al. |
| Binongcal | Filipino | Other Asian | Wong et al. |
| Binoya | Filipino | Other Asian | Wong et al. |
| Binsol | Filipino | Other Asian | Wong et al. |
| Binuya | Filipino | Other Asian | Wong et al. |
| Bio | Filipino | Other Asian | Wong et al. |
| Bioc | Filipino | Other Asian | Wong et al. |
| Bionat | Filipino | Other Asian | Wong et al. |
| Biong | Filipino | Other Asian | Wong et al. |
| Birco | Filipino | Other Asian | Wong et al. |
| Biring | Asian Indian | South Asian | Wong et al. |
| Birla | Asian Indian | South Asian | Wong et al. |
| Birog | Filipino | Other Asian | Wong et al. |
| Birondo | Filipino | Other Asian | Wong et al. |
| Birung | Filipino | Other Asian | Wong et al. |
| Bisa | Filipino | Other Asian | Wong et al. |
| Bisarra | Filipino | Other Asian | Wong et al. |
| Bisco | Filipino | Other Asian | Wong et al. |
| Biscocho | Filipino | Other Asian | Wong et al. |
| Bisda | Filipino | Other Asian | Wong et al. |
| Bishnoi | Asian Indian | South Asian | Wong et al. |
| Bishowkarma |  | South Asian | Lim et al. |
| Bishwas |  | South Asian | Lim et al. |
| Bisla | Asian Indian | South Asian | Wong et al. |
| Bismonte | Filipino | Other Asian | Wong et al. |
| Bisnar | Filipino | Other Asian | Wong et al. |
| Bisnauth |  | South Asian | Lim et al. |
| Bisquera | Filipino | Other Asian | Wong et al. |
| Bisram |  | South Asian | Lim et al. |
| Bissoondai |  | South Asian | Lim et al. |
| Bissoondial |  | South Asian | Lim et al. |
| Biswas | Asian Indian; Bangladeshi | South Asian | Kranick et al. |
| Bitanga | Filipino | Other Asian | Wong et al. |
| Bitao | Filipino | Other Asian | Wong et al. |
| Biteng | Filipino | Other Asian | Wong et al. |
| Bithi |  | South Asian | Lim et al. |
| Bito | Filipino | Other Asian | Wong et al. |
| Bitong | Filipino | Other Asian | Wong et al. |
| Bitonio | Filipino | Other Asian | Wong et al. |
| Bituin | Filipino | Other Asian | Wong et al. |
| Biu | Chinese | Chinese | Wong et al. |
| Blacer | Filipino | Other Asian | Wong et al. |
| Blanca | Filipino | Other Asian | Wong et al. |
| Blancaflor | Filipino | Other Asian | Wong et al. |
| Blancas | Filipino | Other Asian | Wong et al. |
| Blancia | Filipino | Other Asian | Wong et al. |
| Blanco | Filipino | Other Asian | Wong et al. |
| Blando | Filipino | Other Asian | Wong et al. |
| Blanquera | Filipino | Other Asian | Wong et al. |
| Blanza | Filipino | Other Asian | Wong et al. |
| Blaquera | Filipino | Other Asian | Wong et al. |
| Blardony | Filipino | Other Asian | Wong et al. |
| Blas | Filipino | Other Asian | Wong et al. |
| Blasa | Filipino | Other Asian | Wong et al. |
| Blaza | Filipino | Other Asian | Wong et al. |
| Bleza | Filipino | Other Asian | Wong et al. |
| Blones | Filipino | Other Asian | Wong et al. |
| Blum |  | South Asian | Lim et al. |
| Bo | Chinese | Chinese | Wong et al. |
| Boac | Filipino | Other Asian | Wong et al. |
| Boado | Filipino | Other Asian | Wong et al. |
| Bobadilla | Filipino | Other Asian | Wong et al. |
| Bobba | Asian Indian | South Asian | Wong et al. |
| Bobila | Filipino | Other Asian | Wong et al. |
| Bobiles | Filipino | Other Asian | Wong et al. |
| Bobis | Filipino | Other Asian | Wong et al. |
| Bocala | Filipino | Other Asian | Wong et al. |
| Bocalan | Filipino | Other Asian | Wong et al. |
| Bocalbos | Filipino | Other Asian | Wong et al. |
| Boco | Filipino | Other Asian | Wong et al. |
| Bocobo | Filipino | Other Asian | Wong et al. |
| Bocoboc | Filipino | Other Asian | Wong et al. |
| Bodh | Asian Indian | South Asian | Kranick et al. |
| Boey | Chinese | Chinese | Wong et al. |
| Bofill | Filipino | Other Asian | Wong et al. |
| Boghani | Asian Indian | South Asian | Wong et al. |
| Bognot | Filipino | Other Asian | Wong et al. |
| Bohol | Filipino | Other Asian | Wong et al. |
| Boholst | Filipino | Other Asian | Wong et al. |
| Bohra | Asian Indian | South Asian | Wong et al. |
| Boi | Vietnamese | Other Asian | Wong et al. |
| Boiser | Filipino | Other Asian | Wong et al. |
| Bokar |  | South Asian | Lim et al. |
| Bokkor |  | South Asian | Lim et al. |
| Bokth |  | South Asian | Lim et al. |
| Bola | Asian Indian | South Asian | Wong et al. |
| Bolano | Filipino | Other Asian | Wong et al. |
| Bolanos | Filipino | Other Asian | Wong et al. |
| Bolante | Filipino | Other Asian | Wong et al. |
| Bolario | Filipino | Other Asian | Kranick et al. |
| Bolibol | Filipino | Other Asian | Wong et al. |
| Bolina | Filipino | Other Asian | Wong et al. |
| Bolinao | Filipino | Other Asian | Wong et al. |
| Bolinas | Filipino | Other Asian | Wong et al. |
| Bolisay | Filipino | Other Asian | Wong et al. |
| Bolivar |  | South Asian | Lim et al. |
| Bolloso | Filipino | Other Asian | Wong et al. |
| Bollozos | Filipino | Other Asian | Wong et al. |
| Bolo | Filipino | Other Asian | Wong et al. |
| Bologa | Filipino | Other Asian | Wong et al. |
| Bolong | Filipino | Other Asian | Wong et al. |
| Bolos | Filipino | Other Asian | Wong et al. |
| Bolosan | Filipino | Other Asian | Wong et al. |
| Bolotaolo | Filipino | Other Asian | Wong et al. |
| Bolusan | Filipino | Other Asian | Wong et al. |
| Bomagat | Filipino | Other Asian | Wong et al. |
| Bombarda | Filipino | Other Asian | Wong et al. |
| Bombase | Filipino | Other Asian | Wong et al. |
| Bombasi | Filipino | Other Asian | Wong et al. |
| Bombita | Filipino | Other Asian | Wong et al. |
| Bombon | Filipino | Other Asian | Wong et al. |
| Bonafe | Filipino | Other Asian | Wong et al. |
| Bonagua | Filipino | Other Asian | Wong et al. |
| Bonalos | Filipino | Other Asian | Wong et al. |
| Bonaobra | Filipino | Other Asian | Wong et al. |
| Boncato | Filipino | Other Asian | Wong et al. |
| Boncodin | Filipino | Other Asian | Wong et al. |
| Bondad | Filipino | Other Asian | Wong et al. |
| Bondame | Filipino | Other Asian | Wong et al. |
| Bondoc | Filipino | Other Asian | Wong et al. |
| Bonete | Filipino | Other Asian | Wong et al. |
| Bong | Korean | Other Asian | Wong et al. |
| Bonga | Filipino | Other Asian | Wong et al. |
| Bongat | Filipino | Other Asian | Wong et al. |
| Bongato | Filipino | Other Asian | Wong et al. |
| Bongbonga | Filipino | Other Asian | Wong et al. |
| Bongcaras | Filipino | Other Asian | Wong et al. |
| Bongcaron | Filipino | Other Asian | Wong et al. |
| Bongco | Filipino | Other Asian | Wong et al. |
| Bongo | Filipino | Other Asian | Wong et al. |
| Bongolan | Filipino | Other Asian | Wong et al. |
| Bongon | Filipino | Other Asian | Wong et al. |
| Bonifacio | Filipino | Other Asian | Wong et al. |
| Bonilla | Filipino | Other Asian | Wong et al. |
| Boniol | Filipino | Other Asian | Wong et al. |
| Bonita | Filipino | Other Asian | Wong et al. |
| Bonoan | Filipino | Other Asian | Wong et al. |
| Bonot | Filipino | Other Asian | Wong et al. |
| Bonotan | Filipino | Other Asian | Wong et al. |
| Bonsol | Filipino | Other Asian | Wong et al. |
| Bontia | Filipino | Other Asian | Wong et al. |
| Bontigao | Filipino | Other Asian | Wong et al. |
| Bontilao | Filipino | Other Asian | Wong et al. |
| Bontuyan | Filipino | Other Asian | Wong et al. |
| Bonus | Filipino | Other Asian | Kranick et al. |
| Bonzon | Filipino | Other Asian | Wong et al. |
| Booc | Filipino | Other Asian | Wong et al. |
| Boodoo |  | South Asian | Lim et al. |
| Bool | Filipino | Other Asian | Wong et al. |
| Boongaling | Filipino | Other Asian | Wong et al. |
| Bootwala | Asian Indian | South Asian | Wong et al. |
| Boparai | Asian Indian | South Asian | Wong et al. |
| Boquiren | Filipino | Other Asian | Wong et al. |
| Boquiron | Filipino | Other Asian | Wong et al. |
| Bor | Chinese | Chinese | Wong et al. |
| Borbe | Filipino | Other Asian | Wong et al. |
| Borbon | Filipino | Other Asian | Wong et al. |
| Borce | Filipino | Other Asian | Wong et al. |
| Borcena | Filipino | Other Asian | Wong et al. |
| Bordador | Filipino | Other Asian | Wong et al. |
| Bordeos | Filipino | Other Asian | Wong et al. |
| Bordon | Filipino | Other Asian | Wong et al. |
| Bordonada | Filipino | Other Asian | Wong et al. |
| Borela | Filipino | Other Asian | Wong et al. |
| Borgaonkar | Asian Indian | South Asian | Wong et al. |
| Borge | Filipino | Other Asian | Wong et al. |
| Borgonia | Filipino | Other Asian | Wong et al. |
| Borillo | Filipino | Other Asian | Wong et al. |
| Borinaga | Filipino | Other Asian | Wong et al. |
| Borja | Filipino | Other Asian | Wong et al. |
| Borjal | Filipino | Other Asian | Wong et al. |
| Borje | Filipino | Other Asian | Wong et al. |
| Borkar | Asian Indian | South Asian | Wong et al. |
| Borlagdan | Filipino | Other Asian | Wong et al. |
| Borlasa | Filipino | Other Asian | Wong et al. |
| Borlaza | Filipino | Other Asian | Wong et al. |
| Borlongan | Filipino | Other Asian | Wong et al. |
| Borneo | Filipino | Other Asian | Wong et al. |
| Borras | Filipino | Other Asian | Wong et al. |
| Borrero | Filipino | Other Asian | Wong et al. |
| Borres | Filipino | Other Asian | Wong et al. |
| Borro | Filipino | Other Asian | Wong et al. |
| Borromeo | Filipino | Other Asian | Wong et al. |
| Bose | Asian Indian | South Asian | Wong et al. |
| Bosque | Filipino | Other Asian | Wong et al. |
| Botacion | Filipino | Other Asian | Wong et al. |
| Botardo | Filipino | Other Asian | Wong et al. |
| Bote | Filipino | Other Asian | Wong et al. |
| Boter | Filipino | Other Asian | Wong et al. |
| Botin | Filipino | Other Asian | Wong et al. |
| Boton | Filipino | Other Asian | Wong et al. |
| Botones | Filipino | Other Asian | Wong et al. |
| Bow | Chinese | Chinese | Wong et al. |
| Boy | Chinese | Chinese | Wong et al. |
| Bracamonte | Filipino | Other Asian | Wong et al. |
| Braceros | Filipino | Other Asian | Wong et al. |
| Bragado | Filipino | Other Asian | Wong et al. |
| Bragais | Filipino | Other Asian | Wong et al. |
| Braganza | Filipino | Other Asian | Wong et al. |
| Bragas | Filipino | Other Asian | Wong et al. |
| Brah | Asian Indian | South Asian | Wong et al. |
| Brahmbhatt | Asian Indian | South Asian | Wong et al. |
| Braich | Asian Indian | South Asian | Wong et al. |
| Bramlall |  | South Asian | Lim et al. |
| Brana | Filipino | Other Asian | Wong et al. |
| Branzuela | Filipino | Other Asian | Wong et al. |
| Brar | Asian Indian | South Asian | Wong et al. |
| Brathwaite |  | South Asian | Lim et al. |
| Bravo | Filipino | Other Asian | Wong et al. |
| Braza | Filipino | Other Asian | Wong et al. |
| Brazal | Filipino | Other Asian | Wong et al. |
| Bretana | Filipino | Other Asian | Kranick et al. |
| Bridgemohan |  | South Asian | Lim et al. |
| Brigino | Filipino | Other Asian | Wong et al. |
| Brijall |  | South Asian | Lim et al. |
| Brijlall |  | South Asian | Lim et al. |
| Brijraj |  | South Asian | Lim et al. |
| Brillante | Filipino | Other Asian | Wong et al. |
| Brillantes | Filipino | Other Asian | Wong et al. |
| Brillo | Filipino | Other Asian | Wong et al. |
| Brinas | Filipino | Other Asian | Wong et al. |
| Bringas | Filipino | Other Asian | Wong et al. |
| Brion | Filipino | Other Asian | Wong et al. |
| Briones | Filipino | Other Asian | Wong et al. |
| Brioso | Filipino | Other Asian | Wong et al. |
| Britanico | Filipino | Other Asian | Wong et al. |
| Brito | Asian Indian | South Asian | Wong et al. |
| Britos | Filipino | Other Asian | Wong et al. |
| Briz | Filipino | Other Asian | Wong et al. |
| Brizuela | Filipino | Other Asian | Wong et al. |
| Broas | Filipino | Other Asian | Wong et al. |
| Brodeth | Filipino | Other Asian | Wong et al. |
| Brodit | Filipino | Other Asian | Wong et al. |
| Broncano | Filipino | Other Asian | Wong et al. |
| Brosas | Filipino | Other Asian | Wong et al. |
| Brotonel | Filipino | Other Asian | Wong et al. |
| Brual | Filipino | Other Asian | Wong et al. |
| Bruan | Filipino | Other Asian | Wong et al. |
| Brucal | Filipino | Other Asian | Wong et al. |
| Brucelas | Filipino | Other Asian | Wong et al. |
| Bu | Chinese | Chinese | Wong et al. |
| Buada | Filipino | Other Asian | Wong et al. |
| Buado | Filipino | Other Asian | Wong et al. |
| Bual | Filipino | Other Asian | Wong et al. |
| Buan | Filipino | Other Asian | Wong et al. |
| Buaya | Filipino | Other Asian | Wong et al. |
| Buban | Filipino | Other Asian | Wong et al. |
| Bucad | Filipino | Other Asian | Wong et al. |
| Bucago | Filipino | Other Asian | Wong et al. |
| Bucaneg | Filipino | Other Asian | Wong et al. |
| Bucao | Filipino | Other Asian | Wong et al. |
| Bucasas | Filipino | Other Asian | Wong et al. |
| Buccat | Filipino | Other Asian | Wong et al. |
| Buclatin | Filipino | Other Asian | Wong et al. |
| Bucoy | Filipino | Other Asian | Wong et al. |
| Bucsit | Filipino | Other Asian | Wong et al. |
| Bucu | Filipino | Other Asian | Wong et al. |
| Bucud | Filipino | Other Asian | Wong et al. |
| Budhathoki |  | South Asian | Lim et al. |
| Budhraja | Asian Indian | South Asian | Wong et al. |
| Budhu | Asian Indian | South Asian | Kranick et al. |
| Budhwani | Asian Indian | South Asian | Wong et al. |
| Budomo | Filipino | Other Asian | Wong et al. |
| Buduan | Filipino | Other Asian | Wong et al. |
| Buela | Filipino | Other Asian | Wong et al. |
| Buen | Filipino | Other Asian | Wong et al. |
| Buena | Filipino | Other Asian | Wong et al. |
| Buenafe | Filipino | Other Asian | Wong et al. |
| Buenaflor | Filipino | Other Asian | Wong et al. |
| Buenaluz | Filipino | Other Asian | Wong et al. |
| Buenaobra | Filipino | Other Asian | Wong et al. |
| Buenaseda | Filipino | Other Asian | Wong et al. |
| Buenaventura | Filipino | Other Asian | Wong et al. |
| Buenavides | Filipino | Other Asian | Wong et al. |
| Buenavista | Filipino | Other Asian | Wong et al. |
| Buencamino | Filipino | Other Asian | Wong et al. |
| Buenconsejo | Filipino | Other Asian | Wong et al. |
| Buendia | Filipino | Other Asian | Wong et al. |
| Bueno | Filipino | Other Asian | Wong et al. |
| Buensalido | Filipino | Other Asian | Wong et al. |
| Buensuceso | Filipino | Other Asian | Wong et al. |
| Buentipo | Filipino | Other Asian | Wong et al. |
| Buenvenida | Filipino | Other Asian | Wong et al. |
| Buenviaje | Filipino | Other Asian | Wong et al. |
| Buera | Filipino | Other Asian | Wong et al. |
| Buerano | Filipino | Other Asian | Wong et al. |
| Buesa | Filipino | Other Asian | Wong et al. |
| Bueza | Filipino | Other Asian | Wong et al. |
| Bugante | Filipino | Other Asian | Wong et al. |
| Bugaoan | Filipino | Other Asian | Wong et al. |
| Bugaoisan | Filipino | Other Asian | Wong et al. |
| Bugarin | Filipino | Other Asian | Wong et al. |
| Bugawan | Filipino | Other Asian | Wong et al. |
| Bugay | Filipino | Other Asian | Wong et al. |
| Bugayong | Filipino | Other Asian | Wong et al. |
| Bugtong | Filipino | Other Asian | Wong et al. |
| Buguis | Filipino | Other Asian | Wong et al. |
| Buhain | Filipino | Other Asian | Wong et al. |
| Buhat | Filipino | Other Asian | Wong et al. |
| Buhay | Filipino | Other Asian | Wong et al. |
| Buhisan | Filipino | Other Asian | Wong et al. |
| Bui | Vietnamese | Other Asian | Kranick et al. |
| Buison | Filipino | Other Asian | Wong et al. |
| Buizon | Filipino | Other Asian | Wong et al. |
| Bukhary |  | South Asian | Lim et al. |
| Bukish | Asian Indian | South Asian | Wong et al. |
| Buksh | Asian Indian | South Asian | Wong et al. |
| Bulacan | Filipino | Other Asian | Wong et al. |
| Bulaclac | Filipino | Other Asian | Wong et al. |
| Bulala | Filipino | Other Asian | Wong et al. |
| Bulalacao | Filipino | Other Asian | Wong et al. |
| Bulan | Filipino | Other Asian | Wong et al. |
| Bulanadi | Filipino | Other Asian | Wong et al. |
| Bulanhagui | Filipino | Other Asian | Wong et al. |
| Bulante | Filipino | Other Asian | Wong et al. |
| Bulaon | Filipino | Other Asian | Wong et al. |
| Bulaong | Filipino | Other Asian | Wong et al. |
| Bulatao | Filipino | Other Asian | Wong et al. |
| Bulawan | Filipino | Other Asian | Wong et al. |
| Bulawin | Filipino | Other Asian | Wong et al. |
| Bulbul |  | South Asian | Lim et al. |
| Bulda | Filipino | Other Asian | Wong et al. |
| Bullecer | Filipino | Other Asian | Wong et al. |
| Bulos | Filipino | Other Asian | Wong et al. |
| Bulosan | Filipino | Other Asian | Wong et al. |
| Bulquerin | Filipino | Other Asian | Wong et al. |
| Bulsara | Asian Indian | South Asian | Wong et al. |
| Bulseco | Filipino | Other Asian | Wong et al. |
| Buluran | Filipino | Other Asian | Wong et al. |
| Bulusan | Filipino | Other Asian | Wong et al. |
| Bulusu | Asian Indian | South Asian | Wong et al. |
| Bum | Korean | Other Asian | Wong et al. |
| Bumacod | Filipino | Other Asian | Wong et al. |
| Bumagat | Filipino | Other Asian | Wong et al. |
| Bumanglag | Filipino | Other Asian | Wong et al. |
| Bumanlag | Filipino | Other Asian | Wong et al. |
| Bumatay | Filipino | Other Asian | Wong et al. |
| Bun | Chinese | Chinese | Wong et al. |
| Buna | Filipino | Other Asian | Wong et al. |
| Bunac | Filipino | Other Asian | Wong et al. |
| Bunag | Filipino | Other Asian | Wong et al. |
| Bunagan | Filipino | Other Asian | Wong et al. |
| Bunales | Filipino | Other Asian | Wong et al. |
| Bunao | Filipino | Other Asian | Wong et al. |
| Bunda | Filipino | Other Asian | Wong et al. |
| Bundalian | Filipino | Other Asian | Wong et al. |
| Bundang | Filipino | Other Asian | Wong et al. |
| Bundoc | Filipino | Other Asian | Wong et al. |
| Bungay | Filipino | Other Asian | Wong et al. |
| Bungcayao | Filipino | Other Asian | Wong et al. |
| Buni | Filipino | Other Asian | Wong et al. |
| Buniao | Filipino | Other Asian | Wong et al. |
| Bunnag | Thai | Other Asian | Kranick et al. |
| Buno | Filipino | Other Asian | Wong et al. |
| Bunoan | Filipino | Other Asian | Wong et al. |
| Bunuan | Filipino | Other Asian | Wong et al. |
| Bunya | Japanese | Other Asian | Wong et al. |
| Bunyi | Filipino | Other Asian | Wong et al. |
| Buot | Filipino | Other Asian | Wong et al. |
| Buquid | Filipino | Other Asian | Wong et al. |
| Buquing | Filipino | Other Asian | Wong et al. |
| Buraga | Filipino | Other Asian | Wong et al. |
| Burayag | Filipino | Other Asian | Wong et al. |
| Burbank |  | South Asian | Lim et al. |
| Burbano | Filipino | Other Asian | Wong et al. |
| Burce | Filipino | Other Asian | Wong et al. |
| Burdeos | Filipino | Other Asian | Wong et al. |
| Burgonio | Filipino | Other Asian | Wong et al. |
| Burgos | Filipino | Other Asian | Kranick et al. |
| Burguillos | Filipino | Other Asian | Wong et al. |
| Burlaza | Filipino | Other Asian | Wong et al. |
| Burog | Filipino | Other Asian | Wong et al. |
| Busacay | Filipino | Other Asian | Wong et al. |
| Busante | Filipino | Other Asian | Wong et al. |
| Bushra |  | South Asian | Lim et al. |
| Busine | Filipino | Other Asian | Wong et al. |
| Busjit |  | South Asian | Lim et al. |
| Buslon | Filipino | Other Asian | Wong et al. |
| Bustalino | Filipino | Other Asian | Wong et al. |
| Bustamante | Filipino | Other Asian | Wong et al. |
| Bustarde | Filipino | Other Asian | Wong et al. |
| Bustillo | Filipino | Other Asian | Wong et al. |
| Bustillos | Filipino | Other Asian | Wong et al. |
| Busto | Filipino | Other Asian | Wong et al. |
| Bustos | Filipino | Other Asian | Wong et al. |
| Bustria | Filipino | Other Asian | Wong et al. |
| Busuego | Filipino | Other Asian | Wong et al. |
| But | Chinese | Chinese | Wong et al. |
| Butac | Filipino | Other Asian | Wong et al. |
| Butalid | Filipino | Other Asian | Wong et al. |
| Butardo | Filipino | Other Asian | Wong et al. |
| Butay | Filipino | Other Asian | Wong et al. |
| Buted | Filipino | Other Asian | Wong et al. |
| Butial | Filipino | Other Asian | Wong et al. |
| Butin | Filipino | Other Asian | Wong et al. |
| Butiong | Filipino | Other Asian | Wong et al. |
| Butiu | Filipino | Other Asian | Wong et al. |
| Butt |  | South Asian | Lim et al. |
| Buttan |  | South Asian | Lim et al. |
| Buttar | Asian Indian | South Asian | Wong et al. |
| Buu | Vietnamese | Other Asian | Wong et al. |
| Buy | Chinese | Chinese | Wong et al. |
| Buyao | Filipino | Other Asian | Wong et al. |
| Buyco | Filipino | Other Asian | Wong et al. |
| Buyson | Filipino | Other Asian | Wong et al. |
| Buzon | Filipino | Other Asian | Wong et al. |
| Byeon | Korean | Other Asian | Wong et al. |
| Byon | Korean | Other Asian | Wong et al. |
| Byong | Korean | Other Asian | Wong et al. |
| Byun | Korean | Other Asian | Kranick et al. |
| Byung | Korean | Other Asian | Wong et al. |
| Ca | Vietnamese | Other Asian | Wong et al. |
| Caabay | Filipino | Other Asian | Wong et al. |
| Caalim | Filipino | Other Asian | Wong et al. |
| Caamic | Filipino | Other Asian | Wong et al. |
| Caampued | Filipino | Other Asian | Wong et al. |
| Caasi | Filipino | Other Asian | Wong et al. |
| Caayon | Filipino | Other Asian | Wong et al. |
| Caba | Filipino | Other Asian | Wong et al. |
| Cababa | Filipino | Other Asian | Wong et al. |
| Cababat | Filipino | Other Asian | Wong et al. |
| Cabacang | Filipino | Other Asian | Wong et al. |
| Cabacar | Filipino | Other Asian | Wong et al. |
| Cabaccang | Filipino | Other Asian | Wong et al. |
| Cabacoy | Filipino | Other Asian | Wong et al. |
| Cabacungan | Filipino | Other Asian | Wong et al. |
| Cabada | Filipino | Other Asian | Wong et al. |
| Cabaddu | Filipino | Other Asian | Wong et al. |
| Cabading | Filipino | Other Asian | Wong et al. |
| Cabael | Filipino | Other Asian | Wong et al. |
| Cabaero | Filipino | Other Asian | Wong et al. |
| Cabagbag | Filipino | Other Asian | Wong et al. |
| Cabagnot | Filipino | Other Asian | Wong et al. |
| Cabagua | Filipino | Other Asian | Wong et al. |
| Cabahit | Filipino | Other Asian | Wong et al. |
| Cabahug | Filipino | Other Asian | Kranick et al. |
| Cabais | Filipino | Other Asian | Wong et al. |
| Cabal | Filipino | Other Asian | Wong et al. |
| Cabalar | Filipino | Other Asian | Wong et al. |
| Cabalatungan | Filipino | Other Asian | Kranick et al. |
| Cabalbag | Filipino | Other Asian | Wong et al. |
| Cabalce | Filipino | Other Asian | Wong et al. |
| Cabales | Filipino | Other Asian | Wong et al. |
| Cabalfin | Filipino | Other Asian | Wong et al. |
| Caballa | Filipino | Other Asian | Wong et al. |
| Caballero | Filipino | Other Asian | Wong et al. |
| Caballes | Filipino | Other Asian | Wong et al. |
| Caballo | Filipino | Other Asian | Wong et al. |
| Cabalo | Filipino | Other Asian | Wong et al. |
| Cabalona | Filipino | Other Asian | Wong et al. |
| Cabalquinto | Filipino | Other Asian | Wong et al. |
| Cabalse | Filipino | Other Asian | Wong et al. |
| Cabalteja | Filipino | Other Asian | Wong et al. |
| Cabaltera | Filipino | Other Asian | Wong et al. |
| Cabaltica | Filipino | Other Asian | Wong et al. |
| Cabalu | Filipino | Other Asian | Wong et al. |
| Cabaluna | Filipino | Other Asian | Wong et al. |
| Cabamongan | Filipino | Other Asian | Wong et al. |
| Caban | Filipino | Other Asian | Wong et al. |
| Cabana | Filipino | Other Asian | Wong et al. |
| Cabanada | Filipino | Other Asian | Wong et al. |
| Cabanag | Filipino | Other Asian | Wong et al. |
| Cabanas | Filipino | Other Asian | Wong et al. |
| Cabanatan | Filipino | Other Asian | Wong et al. |
| Cabanayan | Filipino | Other Asian | Wong et al. |
| Cabanban | Filipino | Other Asian | Wong et al. |
| Cabanero | Filipino | Other Asian | Wong et al. |
| Cabanes | Filipino | Other Asian | Wong et al. |
| Cabanez | Filipino | Other Asian | Wong et al. |
| Cabang | Filipino | Other Asian | Wong et al. |
| Cabangbang | Filipino | Other Asian | Wong et al. |
| Cabangis | Filipino | Other Asian | Wong et al. |
| Cabangon | Filipino | Other Asian | Wong et al. |
| Cabanilla | Filipino | Other Asian | Wong et al. |
| Cabanit | Filipino | Other Asian | Wong et al. |
| Cabanlig | Filipino | Other Asian | Wong et al. |
| Cabanlit | Filipino | Other Asian | Wong et al. |
| Cabanos | Filipino | Other Asian | Wong et al. |
| Cabansag | Filipino | Other Asian | Wong et al. |
| Cabantac | Filipino | Other Asian | Wong et al. |
| Cabanting | Filipino | Other Asian | Wong et al. |
| Cabantog | Filipino | Other Asian | Wong et al. |
| Cabaong | Filipino | Other Asian | Wong et al. |
| Cabardo | Filipino | Other Asian | Wong et al. |
| Cabarle | Filipino | Other Asian | Wong et al. |
| Cabarles | Filipino | Other Asian | Wong et al. |
| Cabarlo | Filipino | Other Asian | Wong et al. |
| Cabarloc | Filipino | Other Asian | Wong et al. |
| Cabaron | Filipino | Other Asian | Wong et al. |
| Cabarrubias | Filipino | Other Asian | Wong et al. |
| Cabarrus | Filipino | Other Asian | Wong et al. |
| Cabasa | Filipino | Other Asian | Wong et al. |
| Cabasag | Filipino | Other Asian | Wong et al. |
| Cabasal | Filipino | Other Asian | Wong et al. |
| Cabase | Filipino | Other Asian | Kranick et al. |
| Cabasug | Filipino | Other Asian | Wong et al. |
| Cabatan | Filipino | Other Asian | Wong et al. |
| Cabatbat | Filipino | Other Asian | Wong et al. |
| Cabatic | Filipino | Other Asian | Wong et al. |
| Cabatingan | Filipino | Other Asian | Wong et al. |
| Cabatit | Filipino | Other Asian | Wong et al. |
| Cabato | Filipino | Other Asian | Wong et al. |
| Cabatu | Filipino | Other Asian | Wong et al. |
| Cabatuan | Filipino | Other Asian | Kranick et al. |
| Cabatuando | Filipino | Other Asian | Wong et al. |
| Cabauatan | Filipino | Other Asian | Wong et al. |
| Cabaya | Filipino | Other Asian | Wong et al. |
| Cabayan | Filipino | Other Asian | Wong et al. |
| Cabayao | Filipino | Other Asian | Wong et al. |
| Cabbab | Filipino | Other Asian | Wong et al. |
| Cabbat | Filipino | Other Asian | Wong et al. |
| Cabebe | Filipino | Other Asian | Wong et al. |
| Cabel | Filipino | Other Asian | Wong et al. |
| Cabelin | Filipino | Other Asian | Wong et al. |
| Cabeliza | Filipino | Other Asian | Wong et al. |
| Cabello | Filipino | Other Asian | Wong et al. |
| Cabellon | Filipino | Other Asian | Wong et al. |
| Cabero | Filipino | Other Asian | Wong et al. |
| Caberte | Filipino | Other Asian | Wong et al. |
| Caberto | Filipino | Other Asian | Wong et al. |
| Cabezas | Filipino | Other Asian | Wong et al. |
| Cabiao | Filipino | Other Asian | Wong et al. |
| Cabias | Filipino | Other Asian | Wong et al. |
| Cabico | Filipino | Other Asian | Wong et al. |
| Cabida | Filipino | Other Asian | Wong et al. |
| Cabides | Filipino | Other Asian | Wong et al. |
| Cabie | Filipino | Other Asian | Wong et al. |
| Cabigao | Filipino | Other Asian | Wong et al. |
| Cabigas | Filipino | Other Asian | Wong et al. |
| Cabigon | Filipino | Other Asian | Wong et al. |
| Cabigting | Filipino | Other Asian | Wong et al. |
| Cabilao | Filipino | Other Asian | Wong et al. |
| Cabilatazan | Filipino | Other Asian | Wong et al. |
| Cabildo | Filipino | Other Asian | Wong et al. |
| Cabiles | Filipino | Other Asian | Wong et al. |
| Cabili | Filipino | Other Asian | Wong et al. |
| Cabilin | Filipino | Other Asian | Wong et al. |
| Cabiling | Filipino | Other Asian | Wong et al. |
| Cabillo | Filipino | Other Asian | Wong et al. |
| Cabiltes | Filipino | Other Asian | Wong et al. |
| Cabinian | Filipino | Other Asian | Wong et al. |
| Cabinta | Filipino | Other Asian | Wong et al. |
| Cabio | Filipino | Other Asian | Wong et al. |
| Cabitac | Filipino | Other Asian | Wong et al. |
| Cablay | Filipino | Other Asian | Wong et al. |
| Cablayan | Filipino | Other Asian | Kranick et al. |
| Cabling | Filipino | Other Asian | Wong et al. |
| Cabo | Filipino | Other Asian | Wong et al. |
| Cabochan | Filipino | Other Asian | Wong et al. |
| Cabonce | Filipino | Other Asian | Wong et al. |
| Cabontocan | Filipino | Other Asian | Wong et al. |
| Cabotage | Filipino | Other Asian | Wong et al. |
| Cabotaje | Filipino | Other Asian | Wong et al. |
| Cabote | Filipino | Other Asian | Wong et al. |
| Cabradilla | Filipino | Other Asian | Wong et al. |
| Cabral | Filipino | Other Asian | Wong et al. |
| Cabrales | Filipino | Other Asian | Wong et al. |
| Cabras | Filipino | Other Asian | Wong et al. |
| Cabreira | Filipino | Other Asian | Wong et al. |
| Cabrera | Filipino | Other Asian | Wong et al. |
| Cabreros | Filipino | Other Asian | Wong et al. |
| Cabreza | Filipino | Other Asian | Wong et al. |
| Cabrido | Filipino | Other Asian | Wong et al. |
| Cabrillas | Filipino | Other Asian | Wong et al. |
| Cabrillos | Filipino | Other Asian | Wong et al. |
| Cabrito | Filipino | Other Asian | Wong et al. |
| Cabuag | Filipino | Other Asian | Wong et al. |
| Cabuang | Filipino | Other Asian | Wong et al. |
| Cabuco | Filipino | Other Asian | Wong et al. |
| Cabudol | Filipino | Other Asian | Wong et al. |
| Cabudoy | Filipino | Other Asian | Wong et al. |
| Cabuena | Filipino | Other Asian | Wong et al. |
| Cabuenos | Filipino | Other Asian | Wong et al. |
| Cabug | Filipino | Other Asian | Wong et al. |
| Cabugao | Filipino | Other Asian | Wong et al. |
| Cabugos | Filipino | Other Asian | Wong et al. |
| Cabugwason | Filipino | Other Asian | Wong et al. |
| Cabuhat | Filipino | Other Asian | Wong et al. |
| Cabulisan | Filipino | Other Asian | Wong et al. |
| Cabulong | Filipino | Other Asian | Wong et al. |
| Cabuloy | Filipino | Other Asian | Wong et al. |
| Cabungcal | Filipino | Other Asian | Wong et al. |
| Cabunoc | Filipino | Other Asian | Wong et al. |
| Cabural | Filipino | Other Asian | Wong et al. |
| Caburao | Filipino | Other Asian | Wong et al. |
| Caburian | Filipino | Other Asian | Wong et al. |
| Caburnay | Filipino | Other Asian | Wong et al. |
| Cabus | Filipino | Other Asian | Wong et al. |
| Cabusao | Filipino | Other Asian | Wong et al. |
| Cabusas | Filipino | Other Asian | Wong et al. |
| Cabuslay | Filipino | Other Asian | Wong et al. |
| Cabusora | Filipino | Other Asian | Wong et al. |
| Cabutaje | Filipino | Other Asian | Wong et al. |
| Cabutotan | Filipino | Other Asian | Wong et al. |
| Cabuyadao | Filipino | Other Asian | Wong et al. |
| Cabuyao | Filipino | Other Asian | Wong et al. |
| Cac | Filipino | Other Asian | Wong et al. |
| Cacabelos | Filipino | Other Asian | Wong et al. |
| Cacacho | Filipino | Other Asian | Wong et al. |
| Cacafranca | Filipino | Other Asian | Wong et al. |
| Cacal | Filipino | Other Asian | Wong et al. |
| Cacalda | Filipino | Other Asian | Wong et al. |
| Cacananta | Filipino | Other Asian | Wong et al. |
| Cacanindin | Filipino | Other Asian | Kranick et al. |
| Cacao | Filipino | Other Asian | Wong et al. |
| Cacas | Filipino | Other Asian | Wong et al. |
| Cacatian | Filipino | Other Asian | Wong et al. |
| Cacayan | Filipino | Other Asian | Wong et al. |
| Cacayorin | Filipino | Other Asian | Wong et al. |
| Cacayuran | Filipino | Other Asian | Wong et al. |
| Caccam | Filipino | Other Asian | Wong et al. |
| Cacdac | Filipino | Other Asian | Wong et al. |
| Caceres | Filipino | Other Asian | Wong et al. |
| Caces | Filipino | Other Asian | Wong et al. |
| Cacha | Filipino | Other Asian | Wong et al. |
| Cachaper | Filipino | Other Asian | Wong et al. |
| Cachapero | Filipino | Other Asian | Wong et al. |
| Cachero | Filipino | Other Asian | Wong et al. |
| Cachin | Filipino | Other Asian | Wong et al. |
| Cacho | Filipino | Other Asian | Wong et al. |
| Cachola | Filipino | Other Asian | Wong et al. |
| Cachuela | Filipino | Other Asian | Wong et al. |
| Cacnio | Filipino | Other Asian | Wong et al. |
| Cacpal | Filipino | Other Asian | Wong et al. |
| Caculitan | Filipino | Other Asian | Wong et al. |
| Cadabona | Filipino | Other Asian | Wong et al. |
| Cadacio | Filipino | Other Asian | Wong et al. |
| Cadag | Filipino | Other Asian | Wong et al. |
| Cadalig | Filipino | Other Asian | Wong et al. |
| Cadalin | Filipino | Other Asian | Wong et al. |
| Cadalzo | Filipino | Other Asian | Wong et al. |
| Cadang | Filipino | Other Asian | Wong et al. |
| Cadano | Filipino | Other Asian | Wong et al. |
| Cadaoas | Filipino | Other Asian | Wong et al. |
| Cadapan | Filipino | Other Asian | Wong et al. |
| Cadatal | Filipino | Other Asian | Wong et al. |
| Cadavona | Filipino | Other Asian | Wong et al. |
| Cadawas | Filipino | Other Asian | Wong et al. |
| Caday | Filipino | Other Asian | Wong et al. |
| Cadelina | Filipino | Other Asian | Wong et al. |
| Cadelinia | Filipino | Other Asian | Wong et al. |
| Cadena | Filipino | Other Asian | Wong et al. |
| Cadiang | Filipino | Other Asian | Wong et al. |
| Cadiente | Filipino | Other Asian | Wong et al. |
| Cadigal | Filipino | Other Asian | Wong et al. |
| Cadion | Filipino | Other Asian | Wong et al. |
| Cadiz | Filipino | Other Asian | Wong et al. |
| Cadorna | Filipino | Other Asian | Wong et al. |
| Cadua | Filipino | Other Asian | Wong et al. |
| Cadungog | Filipino | Other Asian | Wong et al. |
| Caeg | Filipino | Other Asian | Wong et al. |
| Cafe | Filipino | Other Asian | Wong et al. |
| Cafirma | Filipino | Other Asian | Wong et al. |
| Caga | Filipino | Other Asian | Wong et al. |
| Cagaanan | Filipino | Other Asian | Wong et al. |
| Cagalawan | Filipino | Other Asian | Wong et al. |
| Cagampan | Filipino | Other Asian | Wong et al. |
| Cagampang | Filipino | Other Asian | Wong et al. |
| Caganap | Filipino | Other Asian | Wong et al. |
| Cagandahan | Filipino | Other Asian | Wong et al. |
| Cagaoan | Filipino | Other Asian | Wong et al. |
| Cagape | Filipino | Other Asian | Wong et al. |
| Cagara | Filipino | Other Asian | Wong et al. |
| Cagas | Filipino | Other Asian | Wong et al. |
| Cagat | Filipino | Other Asian | Wong et al. |
| Cagayat | Filipino | Other Asian | Kranick et al. |
| Caguia | Filipino | Other Asian | Wong et al. |
| Caguiat | Filipino | Other Asian | Wong et al. |
| Caguicla | Filipino | Other Asian | Wong et al. |
| Caguimbaga | Filipino | Other Asian | Wong et al. |
| Caguin | Filipino | Other Asian | Wong et al. |
| Caguioa | Filipino | Other Asian | Wong et al. |
| Cagulada | Filipino | Other Asian | Kranick et al. |
| Cagungun | Filipino | Other Asian | Wong et al. |
| Cahanap | Filipino | Other Asian | Wong et al. |
| Cahigas | Filipino | Other Asian | Wong et al. |
| Cahilig | Filipino | Other Asian | Wong et al. |
| Cahiwat | Filipino | Other Asian | Wong et al. |
| Cai | Chinese | Chinese | Kranick et al. |
| Cailao | Filipino | Other Asian | Wong et al. |
| Cailing | Filipino | Other Asian | Wong et al. |
| Cailles | Filipino | Other Asian | Wong et al. |
| Caimol | Filipino | Other Asian | Wong et al. |
| Caina | Filipino | Other Asian | Wong et al. |
| Caindec | Filipino | Other Asian | Wong et al. |
| Caingat | Filipino | Other Asian | Wong et al. |
| Cainglet | Filipino | Other Asian | Wong et al. |
| Cainglit | Filipino | Other Asian | Wong et al. |
| Cainoy | Filipino | Other Asian | Wong et al. |
| Caintic | Filipino | Other Asian | Wong et al. |
| Cairme | Filipino | Other Asian | Wong et al. |
| Caisido | Filipino | Other Asian | Wong et al. |
| Caisip | Filipino | Other Asian | Wong et al. |
| Cajayon | Filipino | Other Asian | Wong et al. |
| Cajes | Filipino | Other Asian | Wong et al. |
| Cajigal | Filipino | Other Asian | Wong et al. |
| Cajigas | Filipino | Other Asian | Wong et al. |
| Cajiles | Filipino | Other Asian | Wong et al. |
| Cajili | Filipino | Other Asian | Wong et al. |
| Cajilig | Filipino | Other Asian | Wong et al. |
| Cajimat | Filipino | Other Asian | Wong et al. |
| Cajiuat | Filipino | Other Asian | Wong et al. |
| Cajucom | Filipino | Other Asian | Wong et al. |
| Cajudoy | Filipino | Other Asian | Wong et al. |
| Cajulao | Filipino | Other Asian | Wong et al. |
| Cajulis | Filipino | Other Asian | Wong et al. |
| Calabia | Filipino | Other Asian | Wong et al. |
| Calabio | Filipino | Other Asian | Wong et al. |
| Calabucal | Filipino | Other Asian | Wong et al. |
| Calacal | Filipino | Other Asian | Wong et al. |
| Calacsan | Filipino | Other Asian | Wong et al. |
| Calad | Filipino | Other Asian | Wong et al. |
| Caladiao | Filipino | Other Asian | Wong et al. |
| Calaguas | Filipino | Other Asian | Wong et al. |
| Calagui | Filipino | Other Asian | Wong et al. |
| Calalang | Filipino | Other Asian | Wong et al. |
| Calalay | Filipino | Other Asian | Wong et al. |
| Calalo | Filipino | Other Asian | Wong et al. |
| Calamaan | Filipino | Other Asian | Wong et al. |
| Calamayan | Filipino | Other Asian | Wong et al. |
| Calamba | Filipino | Other Asian | Wong et al. |
| Calamiong | Filipino | Other Asian | Wong et al. |
| Calandria | Filipino | Other Asian | Wong et al. |
| Calangan | Filipino | Other Asian | Wong et al. |
| Calangi | Filipino | Other Asian | Wong et al. |
| Calano | Filipino | Other Asian | Wong et al. |
| Calanog | Filipino | Other Asian | Wong et al. |
| Calantas | Filipino | Other Asian | Wong et al. |
| Calantoc | Filipino | Other Asian | Wong et al. |
| Calantog | Filipino | Other Asian | Wong et al. |
| Calaoagan | Filipino | Other Asian | Wong et al. |
| Calapan | Filipino | Other Asian | Wong et al. |
| Calapini | Filipino | Other Asian | Wong et al. |
| Calapit | Filipino | Other Asian | Wong et al. |
| Calar | Filipino | Other Asian | Wong et al. |
| Calara | Filipino | Other Asian | Wong et al. |
| Calaramo | Filipino | Other Asian | Wong et al. |
| Calaranan | Filipino | Other Asian | Wong et al. |
| Calaro | Filipino | Other Asian | Wong et al. |
| Calata | Filipino | Other Asian | Wong et al. |
| Calaunan | Filipino | Other Asian | Wong et al. |
| Calaustro | Filipino | Other Asian | Wong et al. |
| Calayag | Filipino | Other Asian | Wong et al. |
| Calayan | Filipino | Other Asian | Wong et al. |
| Calaycay | Filipino | Other Asian | Wong et al. |
| Calbay | Filipino | Other Asian | Wong et al. |
| Calceta | Filipino | Other Asian | Wong et al. |
| Caldea | Filipino | Other Asian | Wong et al. |
| Caldejon | Filipino | Other Asian | Wong et al. |
| Caldeo | Filipino | Other Asian | Wong et al. |
| Calderon | Filipino | Other Asian | Wong et al. |
| Caldito | Filipino | Other Asian | Wong et al. |
| Caldo | Filipino | Other Asian | Wong et al. |
| Caldona | Filipino | Other Asian | Wong et al. |
| Caldoza | Filipino | Other Asian | Wong et al. |
| Calero | Filipino | Other Asian | Wong et al. |
| Calibo | Filipino | Other Asian | Wong et al. |
| Caliboso | Filipino | Other Asian | Wong et al. |
| Calibuso | Filipino | Other Asian | Wong et al. |
| Calica | Filipino | Other Asian | Wong et al. |
| Calicdan | Filipino | Other Asian | Wong et al. |
| Calija | Filipino | Other Asian | Wong et al. |
| Calilan | Filipino | Other Asian | Wong et al. |
| Calilao | Filipino | Other Asian | Wong et al. |
| Calilap | Filipino | Other Asian | Wong et al. |
| Calilung | Filipino | Other Asian | Wong et al. |
| Calimag | Filipino | Other Asian | Wong et al. |
| Calimbas | Filipino | Other Asian | Wong et al. |
| Calimlim | Filipino | Other Asian | Wong et al. |
| Calimpong | Filipino | Other Asian | Wong et al. |
| Calimquim | Filipino | Other Asian | Wong et al. |
| Calina | Filipino | Other Asian | Wong et al. |
| Calinao | Filipino | Other Asian | Wong et al. |
| Calindas | Filipino | Other Asian | Wong et al. |
| Caling | Filipino | Other Asian | Wong et al. |
| Calinga | Filipino | Other Asian | Wong et al. |
| Calingasan | Filipino | Other Asian | Wong et al. |
| Calingo | Filipino | Other Asian | Wong et al. |
| Calinisan | Filipino | Other Asian | Wong et al. |
| Calinog | Filipino | Other Asian | Wong et al. |
| Caliolio | Filipino | Other Asian | Wong et al. |
| Calip | Filipino | Other Asian | Wong et al. |
| Calisterio | Filipino | Other Asian | Wong et al. |
| Caliva | Filipino | Other Asian | Wong et al. |
| Calivoso | Filipino | Other Asian | Wong et al. |
| Caliwag | Filipino | Other Asian | Wong et al. |
| Caliwara | Filipino | Other Asian | Wong et al. |
| Calixterio | Filipino | Other Asian | Wong et al. |
| Calixto | Filipino | Other Asian | Wong et al. |
| Calixtro | Filipino | Other Asian | Wong et al. |
| Caliyo | Filipino | Other Asian | Wong et al. |
| Calizar | Filipino | Other Asian | Wong et al. |
| Calizo | Filipino | Other Asian | Wong et al. |
| Callado | Filipino | Other Asian | Wong et al. |
| Callangan | Filipino | Other Asian | Wong et al. |
| Callanta | Filipino | Other Asian | Wong et al. |
| Callao | Filipino | Other Asian | Wong et al. |
| Calle | Filipino | Other Asian | Wong et al. |
| Calleja | Filipino | Other Asian | Wong et al. |
| Callejas | Filipino | Other Asian | Wong et al. |
| Callejo | Filipino | Other Asian | Wong et al. |
| Callera | Filipino | Other Asian | Wong et al. |
| Callo | Filipino | Other Asian | Wong et al. |
| Callorina | Filipino | Other Asian | Wong et al. |
| Callos | Filipino | Other Asian | Wong et al. |
| Callueng | Filipino | Other Asian | Wong et al. |
| Calma | Filipino | Other Asian | Wong et al. |
| Calnea | Filipino | Other Asian | Wong et al. |
| Calo | Filipino | Other Asian | Wong et al. |
| Calon | Filipino | Other Asian | Wong et al. |
| Calonge | Filipino | Other Asian | Wong et al. |
| Calonsag | Filipino | Other Asian | Wong et al. |
| Calope | Filipino | Other Asian | Wong et al. |
| Caloza | Filipino | Other Asian | Wong et al. |
| Calpito | Filipino | Other Asian | Wong et al. |
| Calpo | Filipino | Other Asian | Wong et al. |
| Calso | Filipino | Other Asian | Wong et al. |
| Caluag | Filipino | Other Asian | Wong et al. |
| Calub | Filipino | Other Asian | Wong et al. |
| Calubag | Filipino | Other Asian | Wong et al. |
| Calubaquib | Filipino | Other Asian | Wong et al. |
| Calucag | Filipino | Other Asian | Wong et al. |
| Calucin | Filipino | Other Asian | Wong et al. |
| Calugay | Filipino | Other Asian | Wong et al. |
| Caluma | Filipino | Other Asian | Wong et al. |
| Calumpit | Filipino | Other Asian | Wong et al. |
| Calungsod | Filipino | Other Asian | Wong et al. |
| Calunod | Filipino | Other Asian | Wong et al. |
| Caluya | Filipino | Other Asian | Kranick et al. |
| Caluza | Filipino | Other Asian | Wong et al. |
| Calvan | Filipino | Other Asian | Wong et al. |
| Calvario | Filipino | Other Asian | Wong et al. |
| Calvelo | Filipino | Other Asian | Wong et al. |
| Calvero | Filipino | Other Asian | Wong et al. |
| Calvo | Filipino | Other Asian | Wong et al. |
| Calzada | Filipino | Other Asian | Wong et al. |
| Calzado | Filipino | Other Asian | Wong et al. |
| Calzo | Filipino | Other Asian | Wong et al. |
| Cam | Vietnamese | Other Asian | Wong et al. |
| Camacho | Filipino | Other Asian | Wong et al. |
| Camaclang | Filipino | Other Asian | Wong et al. |
| Camagay | Filipino | Other Asian | Kranick et al. |
| Camahort | Filipino | Other Asian | Wong et al. |
| Camaisa | Filipino | Other Asian | Wong et al. |
| Camama | Filipino | Other Asian | Wong et al. |
| Camanag | Filipino | Other Asian | Wong et al. |
| Camanga | Filipino | Other Asian | Wong et al. |
| Camangeg | Filipino | Other Asian | Wong et al. |
| Camangian | Filipino | Other Asian | Wong et al. |
| Camantigue | Filipino | Other Asian | Wong et al. |
| Camaquin | Filipino | Other Asian | Wong et al. |
| Camara |  | South Asian | Lim et al. |
| Camarao | Filipino | Other Asian | Wong et al. |
| Camarce | Filipino | Other Asian | Wong et al. |
| Camarillo | Filipino | Other Asian | Wong et al. |
| Camat | Filipino | Other Asian | Wong et al. |
| Camaya | Filipino | Other Asian | Wong et al. |
| Camba | Filipino | Other Asian | Wong et al. |
| Cambaliza | Filipino | Other Asian | Wong et al. |
| Cambare | Filipino | Other Asian | Wong et al. |
| Cambay | Filipino | Other Asian | Wong et al. |
| Cambe | Filipino | Other Asian | Wong et al. |
| Cambel | Filipino | Other Asian | Wong et al. |
| Cambia | Filipino | Other Asian | Wong et al. |
| Cambronero | Filipino | Other Asian | Wong et al. |
| Camcam | Filipino | Other Asian | Wong et al. |
| Camello | Filipino | Other Asian | Wong et al. |
| Camerino | Filipino | Other Asian | Wong et al. |
| Camero | Filipino | Other Asian | Wong et al. |
| Camia | Filipino | Other Asian | Wong et al. |
| Camilo | Filipino | Other Asian | Wong et al. |
| Camilon | Filipino | Other Asian | Wong et al. |
| Camiloza | Filipino | Other Asian | Wong et al. |
| Camina | Filipino | Other Asian | Wong et al. |
| Camit | Filipino | Other Asian | Wong et al. |
| Camomot | Filipino | Other Asian | Wong et al. |
| Camonayan | Filipino | Other Asian | Wong et al. |
| Camoras | Filipino | Other Asian | Wong et al. |
| Campanano | Filipino | Other Asian | Wong et al. |
| Campania | Filipino | Other Asian | Wong et al. |
| Campanilla | Filipino | Other Asian | Wong et al. |
| Campilan | Filipino | Other Asian | Wong et al. |
| Campollo | Filipino | Other Asian | Wong et al. |
| Campomanes | Filipino | Other Asian | Wong et al. |
| Campos | Filipino | Other Asian | Wong et al. |
| Camposagrado | Filipino | Other Asian | Wong et al. |
| Camposano | Filipino | Other Asian | Wong et al. |
| Campoy | Filipino | Other Asian | Wong et al. |
| Camu | Filipino | Other Asian | Wong et al. |
| Camua | Filipino | Other Asian | Wong et al. |
| Can | Vietnamese | Other Asian | Wong et al. |
| Cana | Filipino | Other Asian | Wong et al. |
| Canales | Filipino | Other Asian | Wong et al. |
| Canalita | Filipino | Other Asian | Wong et al. |
| Canapi | Filipino | Other Asian | Wong et al. |
| Canare | Filipino | Other Asian | Wong et al. |
| Canares | Filipino | Other Asian | Kranick et al. |
| Canaria | Filipino | Other Asian | Wong et al. |
| Canas | Filipino | Other Asian | Wong et al. |
| Canasa | Filipino | Other Asian | Wong et al. |
| Canave | Filipino | Other Asian | Wong et al. |
| Canaveral | Filipino | Other Asian | Wong et al. |
| Canaya | Filipino | Other Asian | Wong et al. |
| Canaynay | Filipino | Other Asian | Wong et al. |
| Cancel | Filipino | Other Asian | Wong et al. |
| Canciller | Filipino | Other Asian | Wong et al. |
| Cancino | Filipino | Other Asian | Wong et al. |
| Cancio | Filipino | Other Asian | Wong et al. |
| Canda | Filipino | Other Asian | Wong et al. |
| Candari | Filipino | Other Asian | Wong et al. |
| Candaza | Filipino | Other Asian | Wong et al. |
| Candelaria | Filipino | Other Asian | Wong et al. |
| Candelario | Filipino | Other Asian | Wong et al. |
| Candia | Filipino | Other Asian | Wong et al. |
| Cando | Filipino | Other Asian | Wong et al. |
| Caneda | Filipino | Other Asian | Wong et al. |
| Canedo | Filipino | Other Asian | Wong et al. |
| Caneja | Filipino | Other Asian | Wong et al. |
| Canero | Filipino | Other Asian | Wong et al. |
| Caneso | Filipino | Other Asian | Wong et al. |
| Canete | Filipino | Other Asian | Wong et al. |
| Canezal | Filipino | Other Asian | Kranick et al. |
| Cangco | Filipino | Other Asian | Wong et al. |
| Canh | Vietnamese | Other Asian | Wong et al. |
| Canicosa | Filipino | Other Asian | Wong et al. |
| Canicula | Filipino | Other Asian | Wong et al. |
| Canido | Filipino | Other Asian | Wong et al. |
| Canieso | Filipino | Other Asian | Wong et al. |
| Canilang | Filipino | Other Asian | Wong et al. |
| Canilao | Filipino | Other Asian | Wong et al. |
| Canillas | Filipino | Other Asian | Kranick et al. |
| Canillo | Filipino | Other Asian | Wong et al. |
| Canio | Filipino | Other Asian | Wong et al. |
| Canites | Filipino | Other Asian | Wong et al. |
| Canizares | Filipino | Other Asian | Wong et al. |
| Canja | Filipino | Other Asian | Wong et al. |
| Canlapan | Filipino | Other Asian | Wong et al. |
| Canlas | Filipino | Other Asian | Wong et al. |
| Cano | Filipino | Other Asian | Wong et al. |
| Canonigo | Filipino | Other Asian | Wong et al. |
| Canonizado | Filipino | Other Asian | Wong et al. |
| Canono | Filipino | Other Asian | Wong et al. |
| Canos | Filipino | Other Asian | Wong et al. |
| Canosa | Filipino | Other Asian | Wong et al. |
| Canotal | Filipino | Other Asian | Wong et al. |
| Canovas | Filipino | Other Asian | Wong et al. |
| Canoy | Filipino | Other Asian | Wong et al. |
| Canoza | Filipino | Other Asian | Wong et al. |
| Canseco | Filipino | Other Asian | Wong et al. |
| Cansino | Filipino | Other Asian | Wong et al. |
| Canson | Filipino | Other Asian | Wong et al. |
| Canta | Filipino | Other Asian | Wong et al. |
| Cantada | Filipino | Other Asian | Wong et al. |
| Cantal | Filipino | Other Asian | Wong et al. |
| Cantano | Filipino | Other Asian | Wong et al. |
| Cantero | Filipino | Other Asian | Wong et al. |
| Cantiga | Filipino | Other Asian | Wong et al. |
| Cantil | Filipino | Other Asian | Wong et al. |
| Cantila | Filipino | Other Asian | Wong et al. |
| Cantillas | Filipino | Other Asian | Wong et al. |
| Cantiller | Filipino | Other Asian | Wong et al. |
| Cantillo | Filipino | Other Asian | Wong et al. |
| Canto | Filipino | Other Asian | Wong et al. |
| Cantong | Filipino | Other Asian | Wong et al. |
| Cantoria | Filipino | Other Asian | Wong et al. |
| Cantorna | Filipino | Other Asian | Wong et al. |
| Cantos | Filipino | Other Asian | Wong et al. |
| Cantu | Chinese | Chinese | Wong et al. |
| Cantuba | Filipino | Other Asian | Wong et al. |
| Canuto | Filipino | Other Asian | Wong et al. |
| Cao | Chinese | Chinese | Kranick et al. |
| Caoagas | Filipino | Other Asian | Wong et al. |
| Caoagdan | Filipino | Other Asian | Wong et al. |
| Caoile | Filipino | Other Asian | Wong et al. |
| Caoili | Filipino | Other Asian | Wong et al. |
| Capa | Filipino | Other Asian | Wong et al. |
| Capacillo | Filipino | Other Asian | Wong et al. |
| Capacite | Filipino | Other Asian | Wong et al. |
| Capada | Filipino | Other Asian | Wong et al. |
| Capadocia | Filipino | Other Asian | Wong et al. |
| Capalad | Filipino | Other Asian | Wong et al. |
| Capalaran | Filipino | Other Asian | Wong et al. |
| Capalla | Filipino | Other Asian | Wong et al. |
| Capapas | Filipino | Other Asian | Wong et al. |
| Caparas | Filipino | Other Asian | Wong et al. |
| Caparaz | Filipino | Other Asian | Wong et al. |
| Caparino | Filipino | Other Asian | Wong et al. |
| Caparra | Filipino | Other Asian | Wong et al. |
| Caparros | Filipino | Other Asian | Wong et al. |
| Capati | Filipino | Other Asian | Wong et al. |
| Capayas | Filipino | Other Asian | Wong et al. |
| Capellan | Filipino | Other Asian | Wong et al. |
| Capiendo | Filipino | Other Asian | Wong et al. |
| Capil | Filipino | Other Asian | Wong et al. |
| Capili | Filipino | Other Asian | Wong et al. |
| Capilitan | Filipino | Other Asian | Wong et al. |
| Capina | Filipino | Other Asian | Wong et al. |
| Capinia | Filipino | Other Asian | Wong et al. |
| Capino | Filipino | Other Asian | Wong et al. |
| Capinpin | Filipino | Other Asian | Wong et al. |
| Capio | Filipino | Other Asian | Wong et al. |
| Capiral | Filipino | Other Asian | Wong et al. |
| Capistrano | Filipino | Other Asian | Wong et al. |
| Capitan | Filipino | Other Asian | Wong et al. |
| Capito | Filipino | Other Asian | Wong et al. |
| Capitulo | Filipino | Other Asian | Wong et al. |
| Capobres | Filipino | Other Asian | Wong et al. |
| Caponpon | Filipino | Other Asian | Wong et al. |
| Caposagrado | Filipino | Other Asian | Kranick et al. |
| Captain | Asian Indian | South Asian | Wong et al. |
| Capua | Filipino | Other Asian | Wong et al. |
| Capucao | Filipino | Other Asian | Wong et al. |
| Capuchino | Filipino | Other Asian | Wong et al. |
| Capul | Filipino | Other Asian | Wong et al. |
| Capule | Filipino | Other Asian | Wong et al. |
| Capuli | Filipino | Other Asian | Wong et al. |
| Capulong | Filipino | Other Asian | Wong et al. |
| Capunitan | Filipino | Other Asian | Wong et al. |
| Capuno | Filipino | Other Asian | Wong et al. |
| Capuyan | Filipino | Other Asian | Wong et al. |
| Capuz | Filipino | Other Asian | Wong et al. |
| Caraan | Filipino | Other Asian | Wong et al. |
| Caraang | Filipino | Other Asian | Wong et al. |
| Caraballo | Filipino | Other Asian | Wong et al. |
| Carabeo | Filipino | Other Asian | Wong et al. |
| Carabio | Filipino | Other Asian | Wong et al. |
| Carag | Filipino | Other Asian | Wong et al. |
| Caragan | Filipino | Other Asian | Wong et al. |
| Caragay | Filipino | Other Asian | Wong et al. |
| Caraig | Filipino | Other Asian | Wong et al. |
| Caramat | Filipino | Other Asian | Wong et al. |
| Carambas | Filipino | Other Asian | Wong et al. |
| Caranay | Filipino | Other Asian | Wong et al. |
| Carancho | Filipino | Other Asian | Wong et al. |
| Carandang | Filipino | Other Asian | Wong et al. |
| Carantes | Filipino | Other Asian | Wong et al. |
| Caranto | Filipino | Other Asian | Wong et al. |
| Caranza | Filipino | Other Asian | Wong et al. |
| Caraos | Filipino | Other Asian | Wong et al. |
| Carasig | Filipino | Other Asian | Wong et al. |
| Caravana | Filipino | Other Asian | Wong et al. |
| Carbajal | Filipino | Other Asian | Wong et al. |
| Carballo | Filipino | Other Asian | Wong et al. |
| Carbillon | Filipino | Other Asian | Wong et al. |
| Carbonel | Filipino | Other Asian | Wong et al. |
| Carbonell | Filipino | Other Asian | Wong et al. |
| Carbungco | Filipino | Other Asian | Wong et al. |
| Cardano | Filipino | Other Asian | Wong et al. |
| Cardenas | Filipino | Other Asian | Wong et al. |
| Cardeno | Filipino | Other Asian | Wong et al. |
| Cardinas | Filipino | Other Asian | Wong et al. |
| Cardines | Filipino | Other Asian | Wong et al. |
| Cardinez | Filipino | Other Asian | Wong et al. |
| Cardona | Filipino | Other Asian | Wong et al. |
| Cardoza | Filipino | Other Asian | Wong et al. |
| Carganilla | Filipino | Other Asian | Wong et al. |
| Carganillo | Filipino | Other Asian | Wong et al. |
| Cariaga | Filipino | Other Asian | Wong et al. |
| Carian | Filipino | Other Asian | Wong et al. |
| Cariaso | Filipino | Other Asian | Wong et al. |
| Cariazo | Filipino | Other Asian | Wong et al. |
| Caricungan | Filipino | Other Asian | Wong et al. |
| Caridad | Filipino | Other Asian | Wong et al. |
| Carido | Filipino | Other Asian | Wong et al. |
| Carig | Filipino | Other Asian | Wong et al. |
| Carigma | Filipino | Other Asian | Wong et al. |
| Carigo | Filipino | Other Asian | Wong et al. |
| Carillo | Filipino | Other Asian | Wong et al. |
| Carin | Filipino | Other Asian | Wong et al. |
| Caringal | Filipino | Other Asian | Wong et al. |
| Carinio | Filipino | Other Asian | Wong et al. |
| Carino | Filipino | Other Asian | Wong et al. |
| Carion | Chinese | Chinese | Wong et al. |
| Carizal | Filipino | Other Asian | Wong et al. |
| Carlay | Filipino | Other Asian | Wong et al. |
| Carlos | Filipino | Other Asian | Wong et al. |
| Carlota | Filipino | Other Asian | Wong et al. |
| Carmelo | Filipino | Other Asian | Wong et al. |
| Carmona | Filipino | Other Asian | Wong et al. |
| Carnate | Filipino | Other Asian | Wong et al. |
| Carnero | Filipino | Other Asian | Wong et al. |
| Caro | Filipino | Other Asian | Wong et al. |
| Carolino | Filipino | Other Asian | Wong et al. |
| Caronan | Filipino | Other Asian | Wong et al. |
| Caronongan | Filipino | Other Asian | Wong et al. |
| Carpena | Filipino | Other Asian | Wong et al. |
| Carpentero | Filipino | Other Asian | Wong et al. |
| Carpio | Filipino | Other Asian | Wong et al. |
| Carpizo | Filipino | Other Asian | Wong et al. |
| Carpo | Filipino | Other Asian | Wong et al. |
| Carracedo | Filipino | Other Asian | Wong et al. |
| Carrancho | Filipino | Other Asian | Wong et al. |
| Carranza | Filipino | Other Asian | Wong et al. |
| Carrasco | Filipino | Other Asian | Wong et al. |
| Carreon | Filipino | Other Asian | Wong et al. |
| Carrera | Filipino | Other Asian | Wong et al. |
| Carriaga | Filipino | Other Asian | Wong et al. |
| Carrido | Filipino | Other Asian | Wong et al. |
| Carriedo | Filipino | Other Asian | Wong et al. |
| Carrillo | Filipino | Other Asian | Wong et al. |
| Carrion | Filipino | Other Asian | Wong et al. |
| Carsola | Filipino | Other Asian | Wong et al. |
| Cartagena | Filipino | Other Asian | Wong et al. |
| Cartago | Filipino | Other Asian | Wong et al. |
| Carteciano | Filipino | Other Asian | Wong et al. |
| Cartel | Filipino | Other Asian | Wong et al. |
| Carubio | Filipino | Other Asian | Wong et al. |
| Carungay | Filipino | Other Asian | Wong et al. |
| Carungcong | Filipino | Other Asian | Wong et al. |
| Carvajal | Filipino | Other Asian | Wong et al. |
| Carvalho | Asian Indian | South Asian | Wong et al. |
| Carzon | Filipino | Other Asian | Wong et al. |
| Cas | Filipino | Other Asian | Wong et al. |
| Casaba | Filipino | Other Asian | Wong et al. |
| Casabal | Filipino | Other Asian | Wong et al. |
| Casabar | Filipino | Other Asian | Wong et al. |
| Casabay | Filipino | Other Asian | Wong et al. |
| Casabuena | Filipino | Other Asian | Wong et al. |
| Casaclang | Filipino | Other Asian | Wong et al. |
| Casado | Filipino | Other Asian | Wong et al. |
| Casaje | Filipino | Other Asian | Wong et al. |
| Casal | Filipino | Other Asian | Wong et al. |
| Casalla | Filipino | Other Asian | Wong et al. |
| Casallo | Filipino | Other Asian | Wong et al. |
| Casals | Filipino | Other Asian | Wong et al. |
| Casamina | Filipino | Other Asian | Wong et al. |
| Casana | Filipino | Other Asian | Wong et al. |
| Casanas | Filipino | Other Asian | Wong et al. |
| Casanova | Filipino | Other Asian | Wong et al. |
| Casapao | Filipino | Other Asian | Wong et al. |
| Casareno | Filipino | Other Asian | Wong et al. |
| Casas | Filipino | Other Asian | Wong et al. |
| Casasola | Filipino | Other Asian | Wong et al. |
| Casasos | Filipino | Other Asian | Wong et al. |
| Casayuran | Filipino | Other Asian | Wong et al. |
| Cascayan | Filipino | Other Asian | Wong et al. |
| Casco | Filipino | Other Asian | Wong et al. |
| Cascolan | Filipino | Other Asian | Wong et al. |
| Casel | Filipino | Other Asian | Wong et al. |
| Casela | Filipino | Other Asian | Wong et al. |
| Casem | Filipino | Other Asian | Wong et al. |
| Casenas | Filipino | Other Asian | Wong et al. |
| Caseres | Filipino | Other Asian | Wong et al. |
| Casero | Filipino | Other Asian | Wong et al. |
| Cases | Filipino | Other Asian | Wong et al. |
| Casia | Filipino | Other Asian | Wong et al. |
| Casiano | Filipino | Other Asian | Wong et al. |
| Casibu | Filipino | Other Asian | Wong et al. |
| Casica | Filipino | Other Asian | Wong et al. |
| Casidsid | Filipino | Other Asian | Wong et al. |
| Casil | Filipino | Other Asian | Wong et al. |
| Casila | Filipino | Other Asian | Wong et al. |
| Casilan | Filipino | Other Asian | Wong et al. |
| Casilang | Filipino | Other Asian | Wong et al. |
| Casilao | Filipino | Other Asian | Wong et al. |
| Casilla | Filipino | Other Asian | Wong et al. |
| Casillan | Filipino | Other Asian | Wong et al. |
| Casillano | Filipino | Other Asian | Wong et al. |
| Casim | Filipino | Other Asian | Wong et al. |
| Casimiro | Filipino | Other Asian | Wong et al. |
| Casing | Filipino | Other Asian | Wong et al. |
| Casingal | Filipino | Other Asian | Wong et al. |
| Casino | Filipino | Other Asian | Wong et al. |
| Casio | Filipino | Other Asian | Wong et al. |
| Casipe | Filipino | Other Asian | Wong et al. |
| Casipit | Filipino | Other Asian | Wong et al. |
| Casiple | Filipino | Other Asian | Wong et al. |
| Casis | Filipino | Other Asian | Wong et al. |
| Casison | Filipino | Other Asian | Wong et al. |
| Caspe | Filipino | Other Asian | Wong et al. |
| Caspellan | Filipino | Other Asian | Wong et al. |
| Caspillan | Filipino | Other Asian | Wong et al. |
| Caspillo | Filipino | Other Asian | Wong et al. |
| Casquejo | Filipino | Other Asian | Wong et al. |
| Cassum | Asian Indian | South Asian | Wong et al. |
| Castanares | Filipino | Other Asian | Wong et al. |
| Castaneda | Filipino | Other Asian | Wong et al. |
| Castaneto | Filipino | Other Asian | Wong et al. |
| Castano | Filipino | Other Asian | Wong et al. |
| Castanos | Filipino | Other Asian | Wong et al. |
| Castelino | Asian Indian | South Asian | Wong et al. |
| Castelo | Filipino | Other Asian | Wong et al. |
| Castil | Filipino | Other Asian | Wong et al. |
| Castilan | Filipino | Other Asian | Wong et al. |
| Castillano | Filipino | Other Asian | Wong et al. |
| Castillejo | Filipino | Other Asian | Wong et al. |
| Castillejos | Filipino | Other Asian | Wong et al. |
| Castillio | Filipino | Other Asian | Wong et al. |
| Castillo | Filipino | Other Asian | Wong et al. |
| Castillon | Filipino | Other Asian | Wong et al. |
| Castillote | Filipino | Other Asian | Wong et al. |
| Castisimo | Filipino | Other Asian | Wong et al. |
| Castrence | Filipino | Other Asian | Wong et al. |
| Castrillo | Filipino | Other Asian | Wong et al. |
| Castro | Filipino | Other Asian | Wong et al. |
| Castromayor | Filipino | Other Asian | Wong et al. |
| Castronero | Filipino | Other Asian | Wong et al. |
| Castronuevo | Filipino | Other Asian | Wong et al. |
| Castroverde | Filipino | Other Asian | Wong et al. |
| Castueras | Filipino | Other Asian | Wong et al. |
| Castulo | Filipino | Other Asian | Wong et al. |
| Casuga | Filipino | Other Asian | Wong et al. |
| Casugay | Filipino | Other Asian | Wong et al. |
| Casulla | Filipino | Other Asian | Wong et al. |
| Casumpang | Filipino | Other Asian | Wong et al. |
| Casupanan | Filipino | Other Asian | Wong et al. |
| Casupang | Filipino | Other Asian | Wong et al. |
| Cat | Vietnamese | Other Asian | Wong et al. |
| Catabas | Filipino | Other Asian | Wong et al. |
| Catabay | Filipino | Other Asian | Wong et al. |
| Catacutan | Filipino | Other Asian | Wong et al. |
| Catada | Filipino | Other Asian | Wong et al. |
| Cataga | Filipino | Other Asian | Wong et al. |
| Catahan | Filipino | Other Asian | Wong et al. |
| Catajan | Filipino | Other Asian | Wong et al. |
| Catalan | Filipino | Other Asian | Wong et al. |
| Catalasan | Filipino | Other Asian | Wong et al. |
| Catalla | Filipino | Other Asian | Wong et al. |
| Catalon | Filipino | Other Asian | Wong et al. |
| Cataluna | Filipino | Other Asian | Wong et al. |
| Catambay | Filipino | Other Asian | Wong et al. |
| Catamisan | Filipino | Other Asian | Wong et al. |
| Catan | Filipino | Other Asian | Wong et al. |
| Catane | Filipino | Other Asian | Wong et al. |
| Catanes | Filipino | Other Asian | Wong et al. |
| Catangay | Filipino | Other Asian | Wong et al. |
| Catanghal | Filipino | Other Asian | Wong et al. |
| Catangui | Filipino | Other Asian | Wong et al. |
| Catanyag | Filipino | Other Asian | Wong et al. |
| Catap | Filipino | Other Asian | Wong et al. |
| Catapang | Filipino | Other Asian | Wong et al. |
| Catapusan | Filipino | Other Asian | Wong et al. |
| Catarroja | Filipino | Other Asian | Wong et al. |
| Catayong | Filipino | Other Asian | Wong et al. |
| Catbagan | Filipino | Other Asian | Wong et al. |
| Catedral | Filipino | Other Asian | Wong et al. |
| Catibayan | Filipino | Other Asian | Wong et al. |
| Catibog | Filipino | Other Asian | Wong et al. |
| Catig | Filipino | Other Asian | Wong et al. |
| Catiggay | Filipino | Other Asian | Wong et al. |
| Catiis | Filipino | Other Asian | Wong et al. |
| Catimbang | Filipino | Other Asian | Wong et al. |
| Catindig | Filipino | Other Asian | Wong et al. |
| Catingub | Filipino | Other Asian | Wong et al. |
| Catipay | Filipino | Other Asian | Wong et al. |
| Catipon | Filipino | Other Asian | Wong et al. |
| Catli | Filipino | Other Asian | Wong et al. |
| Catolico | Filipino | Other Asian | Wong et al. |
| Catolos | Filipino | Other Asian | Wong et al. |
| Catral | Filipino | Other Asian | Wong et al. |
| Catu | Filipino | Other Asian | Wong et al. |
| Catubay | Filipino | Other Asian | Wong et al. |
| Catubig | Filipino | Other Asian | Wong et al. |
| Catubo | Filipino | Other Asian | Wong et al. |
| Catudan | Filipino | Other Asian | Wong et al. |
| Catuira | Filipino | Other Asian | Wong et al. |
| Catunao | Filipino | Other Asian | Wong et al. |
| Catungal | Filipino | Other Asian | Wong et al. |
| Catura | Filipino | Other Asian | Wong et al. |
| Caturay | Filipino | Other Asian | Wong et al. |
| Cau | Vietnamese | Other Asian | Wong et al. |
| Caudilla | Filipino | Other Asian | Wong et al. |
| Cauilan | Filipino | Other Asian | Wong et al. |
| Caunan | Filipino | Other Asian | Wong et al. |
| Causapin | Filipino | Other Asian | Wong et al. |
| Causing | Filipino | Other Asian | Wong et al. |
| Cautivar | Filipino | Other Asian | Wong et al. |
| Cauton | Filipino | Other Asian | Wong et al. |
| Cavan | Filipino | Other Asian | Wong et al. |
| Cavero | Filipino | Other Asian | Wong et al. |
| Caviles | Filipino | Other Asian | Wong et al. |
| Cavinta | Filipino | Other Asian | Wong et al. |
| Cavosora | Filipino | Other Asian | Wong et al. |
| Cawagas | Filipino | Other Asian | Wong et al. |
| Cawaling | Filipino | Other Asian | Wong et al. |
| Cawile | Filipino | Other Asian | Wong et al. |
| Cawili | Filipino | Other Asian | Wong et al. |
| Cay | Vietnamese | Other Asian | Wong et al. |
| Cayaba | Filipino | Other Asian | Wong et al. |
| Cayaban | Filipino | Other Asian | Wong et al. |
| Cayabo | Filipino | Other Asian | Wong et al. |
| Cayabyab | Filipino | Other Asian | Wong et al. |
| Cayamanda | Filipino | Other Asian | Wong et al. |
| Cayanan | Filipino | Other Asian | Wong et al. |
| Cayanong | Filipino | Other Asian | Wong et al. |
| Cayas | Filipino | Other Asian | Wong et al. |
| Cayetano | Filipino | Other Asian | Wong et al. |
| Caylao | Filipino | Other Asian | Wong et al. |
| Cayme | Filipino | Other Asian | Wong et al. |
| Caymol | Filipino | Other Asian | Wong et al. |
| Caynap | Filipino | Other Asian | Wong et al. |
| Cayosa | Filipino | Other Asian | Wong et al. |
| Cazar | Filipino | Other Asian | Wong et al. |
| Cea | Filipino | Other Asian | Wong et al. |
| Ceballos | Filipino | Other Asian | Wong et al. |
| Cebedo | Filipino | Other Asian | Wong et al. |
| Cebu | Filipino | Other Asian | Wong et al. |
| Cecilio | Filipino | Other Asian | Kranick et al. |
| Cedeno | Filipino | Other Asian | Wong et al. |
| Cedilla | Filipino | Other Asian | Wong et al. |
| Cedo | Filipino | Other Asian | Wong et al. |
| Cedro | Filipino | Other Asian | Wong et al. |
| Cedron | Filipino | Other Asian | Wong et al. |
| Cefre | Filipino | Other Asian | Wong et al. |
| Celebrado | Filipino | Other Asian | Wong et al. |
| Celemin | Filipino | Other Asian | Wong et al. |
| Celeridad | Filipino | Other Asian | Wong et al. |
| Celeste | Filipino | Other Asian | Wong et al. |
| Celestial | Filipino | Other Asian | Wong et al. |
| Celestino | Filipino | Other Asian | Wong et al. |
| Celi | Filipino | Other Asian | Wong et al. |
| Celino | Filipino | Other Asian | Wong et al. |
| Celio | Filipino | Other Asian | Wong et al. |
| Celis | Filipino | Other Asian | Wong et al. |
| Celiz | Filipino | Other Asian | Wong et al. |
| Cellona | Filipino | Other Asian | Wong et al. |
| Celo | Filipino | Other Asian | Wong et al. |
| Celso | Filipino | Other Asian | Wong et al. |
| Celzo | Filipino | Other Asian | Wong et al. |
| Cen | Chinese | Chinese | Kranick et al. |
| Cena | Filipino | Other Asian | Wong et al. |
| Cendana | Filipino | Other Asian | Wong et al. |
| Cendreda | Filipino | Other Asian | Wong et al. |
| Ceng | Chinese | Chinese | Wong et al. |
| Cenidoza | Filipino | Other Asian | Wong et al. |
| Cenina | Filipino | Other Asian | Wong et al. |
| Cenit | Filipino | Other Asian | Wong et al. |
| Ceniza | Filipino | Other Asian | Kranick et al. |
| Cenizal | Filipino | Other Asian | Wong et al. |
| Cenon | Filipino | Other Asian | Wong et al. |
| Centenera | Filipino | Other Asian | Wong et al. |
| Centeno | Filipino | Other Asian | Wong et al. |
| Centillas | Filipino | Other Asian | Wong et al. |
| Centino | Filipino | Other Asian | Wong et al. |
| Cenzon | Filipino | Other Asian | Wong et al. |
| Ceon | Filipino | Other Asian | Wong et al. |
| Cepe | Filipino | Other Asian | Wong et al. |
| Cepeda | Filipino | Other Asian | Wong et al. |
| Cera | Filipino | Other Asian | Wong et al. |
| Ceralde | Filipino | Other Asian | Wong et al. |
| Ceraos | Filipino | Other Asian | Wong et al. |
| Cerbas | Filipino | Other Asian | Wong et al. |
| Cerbolles | Filipino | Other Asian | Wong et al. |
| Cercado | Filipino | Other Asian | Wong et al. |
| Cerda | Filipino | Other Asian | Wong et al. |
| Cerdena | Filipino | Other Asian | Wong et al. |
| Cerdenola | Filipino | Other Asian | Wong et al. |
| Cerdon | Filipino | Other Asian | Wong et al. |
| Cerenado | Filipino | Other Asian | Wong et al. |
| Cereno | Filipino | Other Asian | Wong et al. |
| Cerezo | Filipino | Other Asian | Wong et al. |
| Ceria | Filipino | Other Asian | Wong et al. |
| Cerilles | Filipino | Other Asian | Wong et al. |
| Cerna | Filipino | Other Asian | Wong et al. |
| Certeza | Filipino | Other Asian | Wong et al. |
| Cervana | Filipino | Other Asian | Wong et al. |
| Cervania | Filipino | Other Asian | Wong et al. |
| Cervantes | Filipino | Other Asian | Wong et al. |
| Cervera | Filipino | Other Asian | Wong et al. |
| Cesar | Filipino | Other Asian | Wong et al. |
| Cespedes | Filipino | Other Asian | Wong et al. |
| Cezar | Filipino | Other Asian | Wong et al. |
| Cha | Korean | Other Asian | Kranick et al. |
| Chac | Vietnamese | Other Asian | Wong et al. |
| Chacko | Asian Indian | South Asian | Kranick et al. |
| Chacon | Filipino | Other Asian | Wong et al. |
| Chadda | Asian Indian | South Asian | Wong et al. |
| Chaddha | Asian Indian | South Asian | Wong et al. |
| Chae | Korean | Other Asian | Wong et al. |
| Chaeko | Asian Indian | South Asian | Wong et al. |
| Chagani | Asian Indian | South Asian | Wong et al. |
| Chahal | Asian Indian | South Asian | Wong et al. |
| Chahil | Asian Indian | South Asian | Wong et al. |
| Chai | Korean | Other Asian | Kranick et al. |
| Chaing | Chinese | Chinese | Wong et al. |
| Chak | Chinese | Chinese | Wong et al. |
| Chakko | Asian Indian | South Asian | Wong et al. |
| Chaklader |  | South Asian | Lim et al. |
| Chakrabarti | Asian Indian | South Asian | Kranick et al. |
| Chakrabarty | Asian Indian; Bangladeshi | South Asian | Kranick et al. |
| Chakraborti | Asian Indian | South Asian | Wong et al. |
| Chakraborty | Asian Indian | South Asian | Wong et al. |
| Chakravarthi | Asian Indian | South Asian | Wong et al. |
| Chakravarthy | Asian Indian | South Asian | Wong et al. |
| Chakravarti | Asian Indian | South Asian | Wong et al. |
| Chakravarty | Asian Indian | South Asian | Wong et al. |
| Chakraverty | Asian Indian | South Asian | Wong et al. |
| Chakravorty | Asian Indian | South Asian | Wong et al. |
| Chakroborty |  | South Asian | Lim et al. |
| Chalasani | Asian Indian | South Asian | Wong et al. |
| Challa | Asian Indian | South Asian | Wong et al. |
| Challapalli | Asian Indian | South Asian | Wong et al. |
| Champaneri | Asian Indian | South Asian | Wong et al. |
| Chan | Chinese | Chinese | Kranick et al. |
| Chana | Asian Indian | South Asian | Wong et al. |
| Chanchal |  | South Asian | Lim et al. |
| Chanco | Filipino | Other Asian | Wong et al. |
| Chand | Asian Indian | South Asian | Wong et al. |
| Chandan | Asian Indian | South Asian | Wong et al. |
| Chandani | Asian Indian; Pakistani | South Asian | Kranick et al. |
| Chandapilla | Asian Indian | South Asian | Wong et al. |
| Chandar | Asian Indian | South Asian | Wong et al. |
| Chandarana | Asian Indian | South Asian | Wong et al. |
| Chandavarkar | Asian Indian | South Asian | Wong et al. |
| Chande | Asian Indian | South Asian | Wong et al. |
| Chander |  | South Asian | Lim et al. |
| Chanderban |  | South Asian | Lim et al. |
| Chanderpaul |  | South Asian | Lim et al. |
| Chandhoke | Asian Indian | South Asian | Wong et al. |
| Chandi | Asian Indian | South Asian | Wong et al. |
| Chandler |  | South Asian | Lim et al. |
| Chandna | Asian Indian | South Asian | Wong et al. |
| Chandra | Asian Indian; Bangladeshi; Indonesian | South Asian | Kranick et al. |
| Chandran | Asian Indian | South Asian | Wong et al. |
| Chandrasekar | Asian Indian | South Asian | Wong et al. |
| Chandrasekha | Asian Indian | South Asian | Wong et al. |
| Chandrashekh | Asian Indian | South Asian | Wong et al. |
| Chandulal | Asian Indian | South Asian | Wong et al. |
| Chandy | Asian Indian | South Asian | Wong et al. |
| Chane | Chinese | Chinese | Wong et al. |
| Chang | Chinese; Taiwanese; Korean | Chinese | Kranick et al. |
| Changchien | Chinese | Chinese | Wong et al. |
| Changco | Filipino | Other Asian | Wong et al. |
| Changcoco | Filipino | Other Asian | Wong et al. |
| Change | Chinese | Chinese | Wong et al. |
| Changela | Asian Indian | South Asian | Wong et al. |
| Channa | Asian Indian | South Asian | Wong et al. |
| Channabasapp | Asian Indian | South Asian | Wong et al. |
| Chantengco | Filipino | Other Asian | Kranick et al. |
| Chao | Chinese | Chinese | Kranick et al. |
| Chapagain |  | South Asian | Lim et al. |
| Chapatwala | Asian Indian | South Asian | Wong et al. |
| Char | Chinese | Chinese | Wong et al. |
| Charan | Asian Indian | South Asian | Wong et al. |
| Charania | Asian Indian | South Asian | Wong et al. |
| Charavorty |  | South Asian | Lim et al. |
| Chari | Asian Indian | South Asian | Wong et al. |
| Chark | Chinese | Chinese | Wong et al. |
| Charng | Chinese | Chinese | Wong et al. |
| Charvet | Filipino | Other Asian | Wong et al. |
| Charyulu | Asian Indian | South Asian | Wong et al. |
| Chasmawala | Asian Indian | South Asian | Wong et al. |
| Chatani | Japanese | Other Asian | Wong et al. |
| Chatarpal |  | South Asian | Lim et al. |
| Chatha | Asian Indian | South Asian | Wong et al. |
| Chatoor | Asian Indian | South Asian | Wong et al. |
| Chattergoon |  | South Asian | Lim et al. |
| Chatterjea | Asian Indian | South Asian | Wong et al. |
| Chatterjee | Asian Indian | South Asian | Kranick et al. |
| Chatterji | Asian Indian | South Asian | Wong et al. |
| Chattha | Asian Indian | South Asian | Wong et al. |
| Chattopadhya | Asian Indian | South Asian | Wong et al. |
| Chaturbhai | Asian Indian | South Asian | Wong et al. |
| Chaturvedi | Asian Indian | South Asian | Wong et al. |
| Chau | Chinese | Chinese | Kranick et al. |
| Chaubal | Asian Indian | South Asian | Wong et al. |
| Chaubey | Asian Indian | South Asian | Wong et al. |
| Chaud | Asian Indian | South Asian | Wong et al. |
| Chaudhari | Asian Indian | South Asian | Wong et al. |
| Chaudhary | Asian Indian | South Asian | Kranick et al. |
| Chaudhry | Pakistani | South Asian | Kranick et al. |
| Chaudhuri | Asian Indian; Bangladeshi | South Asian | Kranick et al. |
| Chaudhury | Bangladeshi; Asian Indian | South Asian | Kranick et al. |
| Chauhan | Asian Indian | South Asian | Wong et al. |
| Chaun | Chinese | Chinese | Wong et al. |
| Chaung | Chinese | Chinese | Kranick et al. |
| Chavan | Asian Indian | South Asian | Wong et al. |
| Chavarria | Filipino | Other Asian | Wong et al. |
| Chavda | Asian Indian | South Asian | Wong et al. |
| Chaves | Filipino | Other Asian | Wong et al. |
| Chavez | Filipino | Other Asian | Wong et al. |
| Chavit | Filipino | Other Asian | Wong et al. |
| Chaw | Chinese | Chinese | Wong et al. |
| Chawala | Asian Indian | South Asian | Wong et al. |
| Chawda | Asian Indian | South Asian | Wong et al. |
| Chawla | Asian Indian | South Asian | Kranick et al. |
| Che | Chinese | Chinese | Wong et al. |
| Chea | Cambodian | Other Asian | Kranick et al. |
| Cheang | Cambodian | Other Asian | Kranick et al. |
| Cheaz | Chinese | Chinese | Wong et al. |
| Chebat | Filipino | Other Asian | Wong et al. |
| Chediram |  | South Asian | Lim et al. |
| Chee | Chinese | Chinese | Wong et al. |
| Cheema | Pakistani; Asian Indian | South Asian | Kranick et al. |
| Cheen | Chinese | Chinese | Wong et al. |
| Cheer | Chinese | Chinese | Wong et al. |
| Cheh | Chinese | Chinese | Wong et al. |
| Chehal | Asian Indian | South Asian | Wong et al. |
| Chei | Chinese | Chinese | Wong et al. |
| Cheigh | Korean | Other Asian | Wong et al. |
| Chein | Chinese | Chinese | Wong et al. |
| Cheing | Chinese | Chinese | Wong et al. |
| Chek | Chinese | Chinese | Wong et al. |
| Chellam | Asian Indian | South Asian | Wong et al. |
| Chelliah | Asian Indian | South Asian | Wong et al. |
| Chen | Chinese | Chinese | Kranick et al. |
| Chenchang | Chinese | Chinese | Wong et al. |
| Cheng | Chinese | Chinese | Kranick et al. |
| Chenh | Vietnamese | Other Asian | Wong et al. |
| Chenng | Chinese | Chinese | Wong et al. |
| Cheo | Chinese | Chinese | Wong et al. |
| Cheok | Chinese | Chinese | Wong et al. |
| Cheon | Korean | Other Asian | Wong et al. |
| Cheong | Chinese; Malaysian; Korean | Chinese | Kranick et al. |
| Cher | Chinese | Chinese | Wong et al. |
| Cheria | Asian Indian | South Asian | Wong et al. |
| Cherian | Asian Indian | South Asian | Wong et al. |
| Cheriyan | Asian Indian | South Asian | Wong et al. |
| Chern | Chinese | Chinese | Wong et al. |
| Cherng | Chinese | Chinese | Wong et al. |
| Cherukara | Asian Indian | South Asian | Wong et al. |
| Cherukuri | Asian Indian | South Asian | Wong et al. |
| Cheruvu | Asian Indian | South Asian | Wong et al. |
| Chervu | Asian Indian | South Asian | Wong et al. |
| Chettiar | Asian Indian | South Asian | Wong et al. |
| Chetty | Asian Indian | South Asian | Wong et al. |
| Cheu | Chinese | Chinese | Wong et al. |
| Cheuck | Chinese | Chinese | Wong et al. |
| Cheug | Chinese | Chinese | Wong et al. |
| Cheuk | Chinese | Chinese | Wong et al. |
| Cheun | Chinese | Chinese | Wong et al. |
| Cheung | Chinese | Chinese | Kranick et al. |
| Chew | Chinese; Malaysian | Chinese | Kranick et al. |
| Chez | Chinese | Chinese | Wong et al. |
| Chhabildas | Asian Indian | South Asian | Wong et al. |
| Chhaganbhai | Asian Indian | South Asian | Wong et al. |
| Chhaganlal | Asian Indian | South Asian | Wong et al. |
| Chhatrapati | Asian Indian | South Asian | Wong et al. |
| Chhatwal | Asian Indian | South Asian | Wong et al. |
| Chhaya | Asian Indian | South Asian | Wong et al. |
| Chheda | Asian Indian | South Asian | Wong et al. |
| Chhetri |  | South Asian | Lim et al. |
| Chhibbar |  | South Asian | Lim et al. |
| Chhina | Asian Indian | South Asian | Wong et al. |
| Chhokar | Asian Indian | South Asian | Wong et al. |
| Chhotabhai | Asian Indian | South Asian | Wong et al. |
| Chhotalal | Asian Indian | South Asian | Wong et al. |
| Chi | Chinese; Vietnamese | Chinese | Kranick et al. |
| Chia | Chinese | Chinese | Wong et al. |
| Chian | Chinese | Chinese | Wong et al. |
| Chiang | Chinese | Chinese | Kranick et al. |
| Chiao | Chinese | Chinese | Wong et al. |
| Chiau | Chinese | Chinese | Wong et al. |
| Chib | Asian Indian | South Asian | Wong et al. |
| Chiba | Japanese | Other Asian | Wong et al. |
| Chibana | Japanese | Other Asian | Wong et al. |
| Chicano | Filipino | Other Asian | Wong et al. |
| Chich | Chinese | Chinese | Wong et al. |
| Chichibu | Japanese | Other Asian | Wong et al. |
| Chichioco | Filipino | Other Asian | Wong et al. |
| Chico | Filipino | Other Asian | Wong et al. |
| Chicote | Filipino | Other Asian | Wong et al. |
| Chida | Japanese | Other Asian | Wong et al. |
| Chidambaram | Asian Indian | South Asian | Wong et al. |
| Chie | Chinese | Chinese | Wong et al. |
| Chieh | Chinese | Chinese | Wong et al. |
| Chiem | Vietnamese | Other Asian | Wong et al. |
| Chien | Chinese | Chinese | Kranick et al. |
| Chieng | Chinese; Taiwanese; Vietnam | Chinese | Kranick et al. |
| Chieu | Vietnamese | Other Asian | Wong et al. |
| Chiew | Chinese | Chinese | Wong et al. |
| Chih | Chinese | Chinese | Wong et al. |
| Chihara | Japanese | Other Asian | Wong et al. |
| Chii | Chinese | Chinese | Wong et al. |
| Chik | Chinese | Chinese | Wong et al. |
| Chiku | Japanese | Other Asian | Wong et al. |
| Chil | Korean | Other Asian | Wong et al. |
| Chili | Chinese | Chinese | Wong et al. |
| Chim | Cambodian | Other Asian | Kranick et al. |
| Chima | Asian Indian | South Asian | Wong et al. |
| Chimandel |  | South Asian | Lim et al. |
| Chimanlal | Asian Indian | South Asian | Wong et al. |
| Chime |  | South Asian | Lim et al. |
| Chimoto | Japanese | Other Asian | Wong et al. |
| Chimura | Japanese | Other Asian | Wong et al. |
| Chin | Chinese; Malaysian | Chinese | Kranick et al. |
| China |  | South Asian | Lim et al. |
| Chinai | Asian Indian | South Asian | Wong et al. |
| Chine | Chinese | Chinese | Wong et al. |
| Chinen | Japanese | Other Asian | Wong et al. |
| Ching | Chinese | Chinese | Kranick et al. |
| Chinh | Vietnamese | Other Asian | Wong et al. |
| Chinn | Chinese | Chinese | Wong et al. |
| Chinna | Asian Indian | South Asian | Wong et al. |
| Chinnappa | Asian Indian | South Asian | Wong et al. |
| Chinnaswamy | Asian Indian | South Asian | Wong et al. |
| Chino | Japanese | Other Asian | Wong et al. |
| Chinoy | Asian Indian | South Asian | Wong et al. |
| Chio | Chinese | Chinese | Wong et al. |
| Chiou | Chinese | Chinese | Wong et al. |
| Chipongian | Filipino | Other Asian | Wong et al. |
| Chirayil | Asian Indian | South Asian | Wong et al. |
| Chishti | Pakistani | South Asian | Kranick et al. |
| Chisty |  | South Asian | Lim et al. |
| Chitale | Asian Indian | South Asian | Wong et al. |
| Chitalia | Asian Indian | South Asian | Wong et al. |
| Chitambar | Asian Indian | South Asian | Wong et al. |
| Chiting | Chinese | Chinese | Wong et al. |
| Chitnis | Asian Indian | South Asian | Wong et al. |
| Chitose | Japanese | Other Asian | Wong et al. |
| Chitre | Asian Indian | South Asian | Wong et al. |
| Chitturi | Asian Indian | South Asian | Wong et al. |
| Chityala | Asian Indian | South Asian | Kranick et al. |
| Chiu | Chinese | Chinese | Kranick et al. |
| Chiung | Chinese | Chinese | Wong et al. |
| Chiw | Chinese | Chinese | Wong et al. |
| Cho | Korean; Burmese; Chinese | Other Asian | Kranick et al. |
| Chock | Chinese | Chinese | Wong et al. |
| Chockalingam | Asian Indian | South Asian | Wong et al. |
| Chodha | Asian Indian | South Asian | Wong et al. |
| Chodon |  | South Asian | Lim et al. |
| Choe | Korean | Other Asian | Kranick et al. |
| Choeng | Chinese | Chinese | Wong et al. |
| Chog | Korean | Other Asian | Wong et al. |
| Choh | Korean | Other Asian | Wong et al. |
| Chohan | Pakistani | South Asian | Kranick et al. |
| Choi | Korean; Chinese | Other Asian | Kranick et al. |
| Choie | Korean | Other Asian | Wong et al. |
| Choing | Chinese | Chinese | Wong et al. |
| Chois | Chinese | Chinese | Wong et al. |
| Chok | Chinese | Chinese | Wong et al. |
| Choksey | Asian Indian | South Asian | Wong et al. |
| Chokshi | Asian Indian | South Asian | Wong et al. |
| Choksi | Asian Indian | South Asian | Wong et al. |
| Chol | Korean | Other Asian | Wong et al. |
| Chom | Chinese | Chinese | Wong et al. |
| Chon | Korean | Other Asian | Kranick et al. |
| Chonan | Japanese | Other Asian | Wong et al. |
| Chong | Chinese; Malaysian | Chinese | Kranick et al. |
| Choo | Chinese; Malaysian; Korean | Chinese | Kranick et al. |
| Chool | Korean | Other Asian | Wong et al. |
| Choon |  | South Asian | Lim et al. |
| Chopra | Asian Indian | South Asian | Kranick et al. |
| Chor | Chinese | Chinese | Wong et al. |
| Chot | Korean | Other Asian | Wong et al. |
| Chotoo |  | South Asian | Lim et al. |
| Chou | Chinese | Chinese | Kranick et al. |
| Choudary | Asian Indian | South Asian | Wong et al. |
| Choudhari | Asian Indian | South Asian | Wong et al. |
| Choudhary | Asian Indian | South Asian | Kranick et al. |
| Choudhry | Asian Indian | South Asian | Kranick et al. |
| Choudhury | Bangladeshi; Asian Indian | South Asian | Kranick et al. |
| Chough | Korean | Other Asian | Wong et al. |
| Chouhan | Asian Indian | South Asian | Wong et al. |
| Choung | Korean | Other Asian | Wong et al. |
| Chow | Chinese | Chinese | Kranick et al. |
| Chowdary | Asian Indian | South Asian | Wong et al. |
| Chowdhary | Asian Indian | South Asian | Wong et al. |
| Chowdhaury |  | South Asian | Lim et al. |
| Chowdhoury |  | South Asian | Lim et al. |
| Chowdhry | Asian Indian | South Asian | Wong et al. |
| Chowdhuri | Asian Indian | South Asian | Wong et al. |
| Chowdhury | Bangladeshi; Asian Indian | South Asian | Kranick et al. |
| Chowhury |  | South Asian | Lim et al. |
| Choy | Chinese | Chinese | Kranick et al. |
| Choz | Korean | Other Asian | Wong et al. |
| Chu | Chinese; Vietnamese | Chinese | Kranick et al. |
| Chua | Filipino; Chinese | Other Asian | Kranick et al. |
| Chuakay | Filipino | Other Asian | Wong et al. |
| Chuan | Chinese | Chinese | Wong et al. |
| Chuang | Chinese | Chinese | Wong et al. |
| Chubachi | Japanese | Other Asian | Wong et al. |
| Chuc | Vietnamese | Other Asian | Kranick et al. |
| Chuck | Chinese | Chinese | Wong et al. |
| Chudasama | Asian Indian | South Asian | Wong et al. |
| Chudgar | Asian Indian | South Asian | Wong et al. |
| Chue | Chinese | Chinese | Wong et al. |
| Chueh | Chinese | Chinese | Wong et al. |
| Chuek | Chinese | Chinese | Wong et al. |
| Chuen | Chinese | Chinese | Wong et al. |
| Chueng | Chinese | Chinese | Wong et al. |
| Chuey | Chinese | Chinese | Wong et al. |
| Chughtai |  | South Asian | Lim et al. |
| Chuh | Chinese | Chinese | Wong et al. |
| Chui | Chinese | Chinese | Kranick et al. |
| Chuidian | Filipino | Other Asian | Wong et al. |
| Chuk | Chinese | Chinese | Wong et al. |
| Chull | Korean | Other Asian | Wong et al. |
| Chumacera | Filipino | Other Asian | Wong et al. |
| Chuman | Japanese | Other Asian | Wong et al. |
| Chummar | Asian Indian | South Asian | Wong et al. |
| Chun | Chinese; Korean | Chinese | Kranick et al. |
| Chunara | Asian Indian | South Asian | Wong et al. |
| Chunawala | Asian Indian | South Asian | Wong et al. |
| Chune | Chinese | Chinese | Wong et al. |
| Chung | Chinese; Korean | Chinese | Kranick et al. |
| Chunilal | Asian Indian | South Asian | Wong et al. |
| Chunnulal |  | South Asian | Lim et al. |
| Chuo | Chinese | Chinese | Wong et al. |
| Chuong | Vietnamese | Other Asian | Wong et al. |
| Churaman |  | South Asian | Lim et al. |
| Chut | Chinese | Chinese | Wong et al. |
| Chuu | Chinese | Chinese | Wong et al. |
| Chuy | Chinese | Chinese | Wong et al. |
| Chuyen | Vietnamese | Other Asian | Wong et al. |
| Chwang | Chinese | Chinese | Wong et al. |
| Chwe | Korean | Other Asian | Wong et al. |
| Chy |  | South Asian | Lim et al. |
| Chyan | Chinese | Chinese | Wong et al. |
| Chye | Chinese | Chinese | Wong et al. |
| Chyi | Chinese | Chinese | Wong et al. |
| Chyn | Chinese | Chinese | Wong et al. |
| Chyou | Chinese | Chinese | Wong et al. |
| Chyu | Chinese | Chinese | Wong et al. |
| Chyun | Korean | Other Asian | Wong et al. |
| Chyung | Korean | Other Asian | Wong et al. |
| Chzung | Chinese | Chinese | Wong et al. |
| Ci | Chinese | Chinese | Wong et al. |
| Cid | Filipino | Other Asian | Wong et al. |
| Cidro | Filipino | Other Asian | Wong et al. |
| Cielo | Filipino | Other Asian | Wong et al. |
| Cifra | Filipino | Other Asian | Wong et al. |
| Cimafranca | Filipino | Other Asian | Wong et al. |
| Cimatu | Filipino | Other Asian | Wong et al. |
| Cinco | Filipino | Other Asian | Wong et al. |
| Cinense | Filipino | Other Asian | Wong et al. |
| Ciocon | Filipino | Other Asian | Wong et al. |
| Cipres | Filipino | Other Asian | Wong et al. |
| Cipriaso | Filipino | Other Asian | Wong et al. |
| Cirera | Filipino | Other Asian | Wong et al. |
| Ciriaco | Filipino | Other Asian | Wong et al. |
| Cirilo | Filipino | Other Asian | Wong et al. |
| Cirineo | Filipino | Other Asian | Wong et al. |
| Cisnero | Filipino | Other Asian | Wong et al. |
| Cisneros | Filipino | Other Asian | Wong et al. |
| Ciubal | Filipino | Other Asian | Wong et al. |
| Ciudadano | Filipino | Other Asian | Wong et al. |
| Clamor | Filipino | Other Asian | Wong et al. |
| Clamucha | Filipino | Other Asian | Wong et al. |
| Clanor | Filipino | Other Asian | Wong et al. |
| Claravall | Filipino | Other Asian | Wong et al. |
| Claridad | Filipino | Other Asian | Wong et al. |
| Clarin | Filipino | Other Asian | Wong et al. |
| Clarino | Filipino | Other Asian | Wong et al. |
| Clarion | Filipino | Other Asian | Wong et al. |
| Clarito | Filipino | Other Asian | Wong et al. |
| Claro | Filipino | Other Asian | Wong et al. |
| Claros | Filipino | Other Asian | Wong et al. |
| Claudio | Filipino | Other Asian | Wong et al. |
| Claustro | Filipino | Other Asian | Wong et al. |
| Clava | Filipino | Other Asian | Wong et al. |
| Clavano | Filipino | Other Asian | Wong et al. |
| Clavel | Filipino | Other Asian | Wong et al. |
| Claveria | Filipino | Other Asian | Wong et al. |
| Clemena | Filipino | Other Asian | Wong et al. |
| Clemente | Filipino | Other Asian | Kranick et al. |
| Cleofas | Filipino | Other Asian | Wong et al. |
| Cleofe | Filipino | Other Asian | Wong et al. |
| Cleope | Filipino | Other Asian | Wong et al. |
| Clerigo | Filipino | Other Asian | Wong et al. |
| Cleto | Filipino | Other Asian | Wong et al. |
| Climaco | Filipino | Other Asian | Wong et al. |
| Climacosa | Filipino | Other Asian | Wong et al. |
| Cloma | Filipino | Other Asian | Wong et al. |
| Clomera | Filipino | Other Asian | Wong et al. |
| Clores | Filipino | Other Asian | Wong et al. |
| Closa | Filipino | Other Asian | Wong et al. |
| Clutario | Filipino | Other Asian | Wong et al. |
| Cobangbang | Filipino | Other Asian | Wong et al. |
| Cobarrubia | Filipino | Other Asian | Wong et al. |
| Cobarrubias | Filipino | Other Asian | Wong et al. |
| Coc | Vietnamese | Other Asian | Wong et al. |
| Coca | Filipino | Other Asian | Wong et al. |
| Cocadiz | Filipino | Other Asian | Wong et al. |
| Cocal | Filipino | Other Asian | Wong et al. |
| Coching | Filipino | Other Asian | Wong et al. |
| Cocjin | Filipino | Other Asian | Wong et al. |
| Codilla | Filipino | Other Asian | Wong et al. |
| Codina | Filipino | Other Asian | Wong et al. |
| Codog | Filipino | Other Asian | Wong et al. |
| Coi | Korean | Other Asian | Wong et al. |
| Cojuangco | Filipino | Other Asian | Wong et al. |
| Cok | Chinese | Chinese | Wong et al. |
| Colaco | Asian Indian | South Asian | Wong et al. |
| Colcol | Filipino | Other Asian | Wong et al. |
| Colet | Filipino | Other Asian | Wong et al. |
| Coleto | Filipino | Other Asian | Wong et al. |
| Coligado | Filipino | Other Asian | Wong et al. |
| Colina | Filipino | Other Asian | Wong et al. |
| Colinares | Filipino | Other Asian | Wong et al. |
| Colinco | Filipino | Other Asian | Wong et al. |
| Colis | Filipino | Other Asian | Wong et al. |
| Collado | Filipino | Other Asian | Wong et al. |
| Collantes | Filipino | Other Asian | Wong et al. |
| Collera | Filipino | Other Asian | Wong et al. |
| Collo | Filipino | Other Asian | Wong et al. |
| Colmenar | Filipino | Other Asian | Wong et al. |
| Colmenares | Filipino | Other Asian | Wong et al. |
| Colobong | Filipino | Other Asian | Wong et al. |
| Colocado | Filipino | Other Asian | Wong et al. |
| Coloma | Filipino | Other Asian | Wong et al. |
| Colon | Filipino | Other Asian | Wong et al. |
| Colorado | Filipino | Other Asian | Wong et al. |
| Colorina | Filipino | Other Asian | Wong et al. |
| Coloso | Filipino | Other Asian | Wong et al. |
| Coloyan | Filipino | Other Asian | Wong et al. |
| Columbres | Filipino | Other Asian | Wong et al. |
| Columna | Filipino | Other Asian | Wong et al. |
| Coma | Filipino | Other Asian | Wong et al. |
| Comandante | Filipino | Other Asian | Wong et al. |
| Comar | Asian Indian | South Asian | Wong et al. |
| Combate | Filipino | Other Asian | Wong et al. |
| Combis | Filipino | Other Asian | Wong et al. |
| Comendador | Filipino | Other Asian | Wong et al. |
| Cometa | Filipino | Other Asian | Wong et al. |
| Comia | Filipino | Other Asian | Wong et al. |
| Comilang | Filipino | Other Asian | Wong et al. |
| Comiso | Filipino | Other Asian | Wong et al. |
| Comoda | Filipino | Other Asian | Wong et al. |
| Competente | Filipino | Other Asian | Wong et al. |
| Compuesto | Filipino | Other Asian | Wong et al. |
| Comsti | Filipino | Other Asian | Wong et al. |
| Con | Vietnamese | Other Asian | Wong et al. |
| Conanan | Filipino | Other Asian | Wong et al. |
| Concengco | Filipino | Other Asian | Wong et al. |
| Concepcion | Filipino | Other Asian | Wong et al. |
| Conception | Filipino | Other Asian | Wong et al. |
| Concha | Filipino | Other Asian | Wong et al. |
| Concon | Filipino | Other Asian | Wong et al. |
| Conde | Filipino | Other Asian | Wong et al. |
| Condes | Filipino | Other Asian | Wong et al. |
| Condevillama | Filipino | Other Asian | Wong et al. |
| Condez | Filipino | Other Asian | Wong et al. |
| Conejero | Filipino | Other Asian | Wong et al. |
| Confesor | Filipino | Other Asian | Wong et al. |
| Conlu | Filipino | Other Asian | Wong et al. |
| Conol | Filipino | Other Asian | Wong et al. |
| Conopio | Filipino | Other Asian | Wong et al. |
| Consengco | Filipino | Other Asian | Wong et al. |
| Consignado | Filipino | Other Asian | Wong et al. |
| Consing | Filipino | Other Asian | Wong et al. |
| Consolacion | Filipino | Other Asian | Wong et al. |
| Constante | Filipino | Other Asian | Wong et al. |
| Constantino | Filipino | Other Asian | Wong et al. |
| Consul | Filipino | Other Asian | Wong et al. |
| Consunji | Filipino | Other Asian | Wong et al. |
| Contapay | Filipino | Other Asian | Wong et al. |
| Contawe | Filipino | Other Asian | Wong et al. |
| Contemprato | Filipino | Other Asian | Wong et al. |
| Contractor | Asian Indian | South Asian | Wong et al. |
| Contreras | Filipino | Other Asian | Wong et al. |
| Conui | Filipino | Other Asian | Wong et al. |
| Convento | Filipino | Other Asian | Wong et al. |
| Conwi | Filipino | Other Asian | Wong et al. |
| Coo | Chinese | Chinese | Wong et al. |
| Cooc | Vietnamese | Other Asian | Wong et al. |
| Copon | Filipino | Other Asian | Wong et al. |
| Coquia | Filipino | Other Asian | Wong et al. |
| Coquilla | Filipino | Other Asian | Wong et al. |
| Coral | Filipino | Other Asian | Wong et al. |
| Coralde | Filipino | Other Asian | Wong et al. |
| Corales | Filipino | Other Asian | Wong et al. |
| Corbilla | Filipino | Other Asian | Wong et al. |
| Corcega | Filipino | Other Asian | Wong et al. |
| Corcino | Filipino | Other Asian | Wong et al. |
| Corcoro | Filipino | Other Asian | Wong et al. |
| Corcuera | Filipino | Other Asian | Wong et al. |
| Cordero | Filipino | Other Asian | Wong et al. |
| Cordoba | Filipino | Other Asian | Wong et al. |
| Cordova | Filipino | Other Asian | Wong et al. |
| Cordovez | Filipino | Other Asian | Wong et al. |
| Cordoviz | Filipino | Other Asian | Wong et al. |
| Coria | Filipino | Other Asian | Wong et al. |
| Corla | Filipino | Other Asian | Wong et al. |
| Cornejo | Filipino | Other Asian | Wong et al. |
| Cornel | Filipino | Other Asian | Wong et al. |
| Cornelio | Filipino | Other Asian | Kranick et al. |
| Cornista | Filipino | Other Asian | Wong et al. |
| Corominas | Filipino | Other Asian | Wong et al. |
| Corona | Filipino | Other Asian | Wong et al. |
| Coronado | Filipino | Other Asian | Wong et al. |
| Coronel | Filipino | Other Asian | Wong et al. |
| Corotan | Filipino | Other Asian | Wong et al. |
| Corpin | Filipino | Other Asian | Wong et al. |
| Corporal | Filipino | Other Asian | Wong et al. |
| Corpoz | Filipino | Other Asian | Wong et al. |
| Corpus | Filipino | Other Asian | Wong et al. |
| Corpuz | Filipino | Other Asian | Kranick et al. |
| Corral | Filipino | Other Asian | Wong et al. |
| Corrales | Filipino | Other Asian | Wong et al. |
| Corre | Filipino | Other Asian | Wong et al. |
| Correa | Filipino | Other Asian | Wong et al. |
| Correos | Filipino | Other Asian | Wong et al. |
| Corres | Filipino | Other Asian | Wong et al. |
| Corro | Filipino | Other Asian | Wong et al. |
| Corsino | Filipino | Other Asian | Wong et al. |
| Cortado | Filipino | Other Asian | Wong et al. |
| Cortel | Filipino | Other Asian | Wong et al. |
| Cortes | Filipino | Other Asian | Wong et al. |
| Cortez | Filipino | Other Asian | Wong et al. |
| Corteza | Filipino | Other Asian | Wong et al. |
| Cortezano | Filipino | Other Asian | Wong et al. |
| Cortina | Filipino | Other Asian | Wong et al. |
| Corvera | Filipino | Other Asian | Wong et al. |
| Cosare | Filipino | Other Asian | Wong et al. |
| Cosca | Filipino | Other Asian | Wong et al. |
| Coscolluela | Filipino | Other Asian | Wong et al. |
| Cosep | Filipino | Other Asian | Wong et al. |
| Cosico | Filipino | Other Asian | Wong et al. |
| Cosio | Filipino | Other Asian | Wong et al. |
| Cosme | Filipino | Other Asian | Wong et al. |
| Cosmiano | Filipino | Other Asian | Wong et al. |
| Coson | Filipino | Other Asian | Wong et al. |
| Costales | Filipino | Other Asian | Wong et al. |
| Costelo | Filipino | Other Asian | Wong et al. |
| Costes | Filipino | Other Asian | Wong et al. |
| Costillo | Filipino | Other Asian | Wong et al. |
| Costiniano | Filipino | Other Asian | Wong et al. |
| Costosa | Filipino | Other Asian | Wong et al. |
| Costuna | Filipino | Other Asian | Wong et al. |
| Cotaco | Filipino | Other Asian | Wong et al. |
| Cotoner | Filipino | Other Asian | Wong et al. |
| Cou | Chinese | Chinese | Wong et al. |
| Covita | Filipino | Other Asian | Wong et al. |
| Cow | Chinese | Chinese | Wong et al. |
| Coyoca | Filipino | Other Asian | Wong et al. |
| Cravotto | Filipino | Other Asian | Wong et al. |
| Credo | Filipino | Other Asian | Wong et al. |
| Creencia | Filipino | Other Asian | Wong et al. |
| Crescini | Filipino | Other Asian | Wong et al. |
| Cresencia | Filipino | Other Asian | Wong et al. |
| Cresencio | Filipino | Other Asian | Wong et al. |
| Crespo | Filipino | Other Asian | Wong et al. |
| Creus | Filipino | Other Asian | Wong et al. |
| Crisanto | Filipino | Other Asian | Wong et al. |
| Crisol | Filipino | Other Asian | Wong et al. |
| Crisolo | Filipino | Other Asian | Wong et al. |
| Crisologo | Filipino | Other Asian | Wong et al. |
| Crisostomo | Filipino | Other Asian | Wong et al. |
| Cristal | Filipino | Other Asian | Wong et al. |
| Cristales | Filipino | Other Asian | Wong et al. |
| Criste | Filipino | Other Asian | Wong et al. |
| Cristi | Filipino | Other Asian | Wong et al. |
| Cristobal | Filipino | Other Asian | Wong et al. |
| Crizaldo | Filipino | Other Asian | Wong et al. |
| Crucena | Filipino | Other Asian | Wong et al. |
| Cruces | Filipino | Other Asian | Wong et al. |
| Crucillo | Filipino | Other Asian | Wong et al. |
| Cruspero | Filipino | Other Asian | Wong et al. |
| Cruz | Filipino | Other Asian | Wong et al. |
| Cruzada | Filipino | Other Asian | Wong et al. |
| Cruzado | Filipino | Other Asian | Wong et al. |
| Cruzat | Filipino | Other Asian | Wong et al. |
| Cruzdela | Filipino | Other Asian | Wong et al. |
| Cruzet | Filipino | Other Asian | Wong et al. |
| Cu | Vietnamese | Other Asian | Wong et al. |
| Cua | Filipino | Other Asian | Wong et al. |
| Cuachon | Filipino | Other Asian | Wong et al. |
| Cuaderno | Filipino | Other Asian | Wong et al. |
| Cuadra | Filipino | Other Asian | Wong et al. |
| Cuadro | Filipino | Other Asian | Wong et al. |
| Cuago | Filipino | Other Asian | Wong et al. |
| Cuajunco | Filipino | Other Asian | Wong et al. |
| Cuan | Chinese | Chinese | Wong et al. |
| Cuano | Filipino | Other Asian | Wong et al. |
| Cuaresma | Filipino | Other Asian | Wong et al. |
| Cuario | Filipino | Other Asian | Wong et al. |
| Cuartelon | Filipino | Other Asian | Wong et al. |
| Cuartero | Filipino | Other Asian | Wong et al. |
| Cuarteros | Filipino | Other Asian | Wong et al. |
| Cuarto | Filipino | Other Asian | Wong et al. |
| Cuasay | Filipino | Other Asian | Wong et al. |
| Cuasito | Filipino | Other Asian | Wong et al. |
| Cuaycong | Filipino | Other Asian | Wong et al. |
| Cubacub | Filipino | Other Asian | Wong et al. |
| Cubangbang | Filipino | Other Asian | Wong et al. |
| Cube | Filipino | Other Asian | Wong et al. |
| Cubelo | Filipino | Other Asian | Wong et al. |
| Cubero | Filipino | Other Asian | Wong et al. |
| Cubilla | Filipino | Other Asian | Wong et al. |
| Cubillas | Filipino | Other Asian | Wong et al. |
| Cubillo | Filipino | Other Asian | Wong et al. |
| Cubio | Filipino | Other Asian | Wong et al. |
| Cubol | Filipino | Other Asian | Wong et al. |
| Cubos | Filipino | Other Asian | Wong et al. |
| Cuc | Vietnamese | Other Asian | Wong et al. |
| Cuchapin | Filipino | Other Asian | Wong et al. |
| Cucio | Filipino | Other Asian | Wong et al. |
| Cucueco | Filipino | Other Asian | Wong et al. |
| Cudal | Filipino | Other Asian | Wong et al. |
| Cudia | Filipino | Other Asian | Wong et al. |
| Cudiamat | Filipino | Other Asian | Wong et al. |
| Cudilla | Filipino | Other Asian | Wong et al. |
| Cuello | Filipino | Other Asian | Wong et al. |
| Cuenca | Filipino | Other Asian | Wong et al. |
| Cuenco | Filipino | Other Asian | Wong et al. |
| Cuenta | Filipino | Other Asian | Wong et al. |
| Cuento | Filipino | Other Asian | Wong et al. |
| Cuenza | Filipino | Other Asian | Wong et al. |
| Cuerpo | Filipino | Other Asian | Wong et al. |
| Cuesta | Filipino | Other Asian | Wong et al. |
| Cueto | Filipino | Other Asian | Wong et al. |
| Cueva | Filipino | Other Asian | Wong et al. |
| Cuevas | Filipino | Other Asian | Kranick et al. |
| Cuevo | Filipino | Other Asian | Wong et al. |
| Cui | Chinese | Chinese | Kranick et al. |
| Cuico | Filipino | Other Asian | Wong et al. |
| Cuisia | Filipino | Other Asian | Wong et al. |
| Cuison | Filipino | Other Asian | Wong et al. |
| Cuizon | Filipino | Other Asian | Wong et al. |
| Culala | Filipino | Other Asian | Wong et al. |
| Culanag | Filipino | Other Asian | Wong et al. |
| Culannay | Filipino | Other Asian | Wong et al. |
| Culata | Filipino | Other Asian | Wong et al. |
| Culla | Filipino | Other Asian | Wong et al. |
| Cumagun | Filipino | Other Asian | Wong et al. |
| Cumlat | Filipino | Other Asian | Wong et al. |
| Cumpio | Filipino | Other Asian | Wong et al. |
| Cun | Vietnamese | Other Asian | Wong et al. |
| Cuna | Filipino | Other Asian | Wong et al. |
| Cunada | Filipino | Other Asian | Wong et al. |
| Cunanan | Filipino | Other Asian | Wong et al. |
| Cundangan | Filipino | Other Asian | Wong et al. |
| Cuneta | Filipino | Other Asian | Wong et al. |
| Cung | Vietnamese | Other Asian | Wong et al. |
| Cuntapay | Filipino | Other Asian | Wong et al. |
| Cuong | Vietnamese | Other Asian | Wong et al. |
| Cupat | Filipino | Other Asian | Wong et al. |
| Cupay | Filipino | Other Asian | Wong et al. |
| Cupino | Filipino | Other Asian | Wong et al. |
| Cura | Filipino | Other Asian | Wong et al. |
| Curamen | Filipino | Other Asian | Wong et al. |
| Curameng | Filipino | Other Asian | Wong et al. |
| Curammeng | Filipino | Other Asian | Wong et al. |
| Cureg | Filipino | Other Asian | Wong et al. |
| Curilan | Filipino | Other Asian | Wong et al. |
| Curimao | Filipino | Other Asian | Wong et al. |
| Curioso | Filipino | Other Asian | Wong et al. |
| Currim | Asian Indian | South Asian | Wong et al. |
| Cusi | Filipino | Other Asian | Wong et al. |
| Custodia | Filipino | Other Asian | Wong et al. |
| Custodio | Filipino | Other Asian | Wong et al. |
| Cutaran | Filipino | Other Asian | Wong et al. |
| Cutillar | Filipino | Other Asian | Wong et al. |
| Cuu | Vietnamese | Other Asian | Wong et al. |
| Cuunjieng | Filipino | Other Asian | Wong et al. |
| Cuvin | Filipino | Other Asian | Wong et al. |
| Cuya | Filipino | Other Asian | Wong et al. |
| Cuyo | Filipino | Other Asian | Wong et al. |
| Cuyos | Filipino | Other Asian | Wong et al. |
| Cuyugan | Filipino | Other Asian | Wong et al. |
| Cynn | Korean | Other Asian | Wong et al. |
| Daan | Filipino | Other Asian | Wong et al. |
| Daanton | Filipino | Other Asian | Wong et al. |
| Dabalos | Filipino | Other Asian | Wong et al. |
| Dabalus | Filipino | Other Asian | Wong et al. |
| Dabao | Filipino | Other Asian | Wong et al. |
| Dabas | Asian Indian | South Asian | Wong et al. |
| Dabholkar | Asian Indian | South Asian | Kranick et al. |
| Dabiu |  | South Asian | Lim et al. |
| Dablo | Filipino | Other Asian | Wong et al. |
| Dabu | Filipino | Other Asian | Wong et al. |
| Dabuet | Filipino | Other Asian | Wong et al. |
| Dac | Vietnamese | Other Asian | Wong et al. |
| Dacalos | Filipino | Other Asian | Wong et al. |
| Dacanay | Filipino | Other Asian | Kranick et al. |
| Dacasin | Filipino | Other Asian | Wong et al. |
| Dacay | Filipino | Other Asian | Wong et al. |
| Dacayanan | Filipino | Other Asian | Wong et al. |
| Dacayo | Filipino | Other Asian | Wong et al. |
| Dacillo | Filipino | Other Asian | Wong et al. |
| Dacio | Filipino | Other Asian | Wong et al. |
| Daclan | Filipino | Other Asian | Wong et al. |
| Daco | Filipino | Other Asian | Wong et al. |
| Dacoco | Filipino | Other Asian | Wong et al. |
| Dacones | Filipino | Other Asian | Wong et al. |
| Dacpano | Filipino | Other Asian | Wong et al. |
| Dacquel | Filipino | Other Asian | Wong et al. |
| Dacruz | Chinese | Chinese | Wong et al. |
| Dacula | Filipino | Other Asian | Wong et al. |
| Dacuma | Filipino | Other Asian | Wong et al. |
| Dacumos | Filipino | Other Asian | Wong et al. |
| Dacuycuy | Filipino | Other Asian | Wong et al. |
| Dada | Asian Indian | South Asian | Wong et al. |
| Dadi | Asian Indian | South Asian | Wong et al. |
| Dadia | Filipino | Other Asian | Wong et al. |
| Dadis | Filipino | Other Asian | Wong et al. |
| Dadivas | Filipino | Other Asian | Wong et al. |
| Dadiz | Filipino | Other Asian | Wong et al. |
| Dador | Filipino | Other Asian | Wong et al. |
| Dadufalza | Filipino | Other Asian | Wong et al. |
| Dadula | Filipino | Other Asian | Wong et al. |
| Dadulla | Filipino | Other Asian | Wong et al. |
| Dael | Filipino | Other Asian | Wong et al. |
| Daen | Filipino | Other Asian | Wong et al. |
| Daet | Filipino | Other Asian | Wong et al. |
| Daez | Filipino | Other Asian | Wong et al. |
| Daffon | Filipino | Other Asian | Wong et al. |
| Daftary | Asian Indian | South Asian | Wong et al. |
| Daga | Filipino | Other Asian | Wong et al. |
| Dagal | Filipino | Other Asian | Wong et al. |
| Dagamac | Filipino | Other Asian | Wong et al. |
| Dagami | Filipino | Other Asian | Wong et al. |
| Dagampat | Filipino | Other Asian | Wong et al. |
| Dagan | Filipino | Other Asian | Wong et al. |
| Dagang | Filipino | Other Asian | Wong et al. |
| Dagar | Filipino | Other Asian | Wong et al. |
| Dagatan | Filipino | Other Asian | Wong et al. |
| Dagdag | Filipino | Other Asian | Wong et al. |
| Dagdagan | Filipino | Other Asian | Wong et al. |
| Dagdayan | Filipino | Other Asian | Wong et al. |
| Dagli | Asian Indian | South Asian | Wong et al. |
| Dagondon | Filipino | Other Asian | Wong et al. |
| Dagsaan | Filipino | Other Asian | Wong et al. |
| Dagucon | Filipino | Other Asian | Wong et al. |
| Daguio | Filipino | Other Asian | Wong et al. |
| Dagulo | Filipino | Other Asian | Wong et al. |
| Daguman | Filipino | Other Asian | Wong et al. |
| Daguno | Filipino | Other Asian | Wong et al. |
| Dagupion | Filipino | Other Asian | Wong et al. |
| Daguro | Filipino | Other Asian | Wong et al. |
| Dahilig | Filipino | Other Asian | Wong et al. |
| Dahilog | Filipino | Other Asian | Wong et al. |
| Dahiya | Asian Indian | South Asian | Wong et al. |
| Dahodwala | Asian Indian | South Asian | Wong et al. |
| Dahya | Asian Indian | South Asian | Wong et al. |
| Dahyabhai | Asian Indian | South Asian | Wong et al. |
| Dai | Chinese | Chinese | Wong et al. |
| Daigo | Japanese | Other Asian | Wong et al. |
| Daikoku | Japanese | Other Asian | Wong et al. |
| Daileg | Filipino | Other Asian | Wong et al. |
| Dailo | Filipino | Other Asian | Wong et al. |
| Daimally |  | South Asian | Lim et al. |
| Dairo |  | South Asian | Lim et al. |
| Dait | Filipino | Other Asian | Wong et al. |
| Daite | Filipino | Other Asian | Wong et al. |
| Dajao | Filipino | Other Asian | Wong et al. |
| Dajay | Filipino | Other Asian | Wong et al. |
| Dajose | Filipino | Other Asian | Wong et al. |
| Dal | Korean | Other Asian | Wong et al. |
| Dalagan | Filipino | Other Asian | Wong et al. |
| Dalal | Asian Indian | South Asian | Wong et al. |
| Dalan | Filipino | Other Asian | Wong et al. |
| Dalangin | Filipino | Other Asian | Wong et al. |
| Dalao | Filipino | Other Asian | Wong et al. |
| Dalay | Filipino | Other Asian | Wong et al. |
| Dalena | Filipino | Other Asian | Wong et al. |
| Dalere | Filipino | Other Asian | Wong et al. |
| Dalida | Filipino | Other Asian | Wong et al. |
| Daligcon | Filipino | Other Asian | Wong et al. |
| Daligdig | Filipino | Other Asian | Wong et al. |
| Dalija | Filipino | Other Asian | Wong et al. |
| Dalim |  | South Asian | Lim et al. |
| Dalina | Filipino | Other Asian | Wong et al. |
| Dalindin | Filipino | Other Asian | Wong et al. |
| Dalipe | Filipino | Other Asian | Wong et al. |
| Dalire | Filipino | Other Asian | Wong et al. |
| Dalisay | Filipino | Other Asian | Wong et al. |
| Dalit | Filipino | Other Asian | Wong et al. |
| Daliva | Filipino | Other Asian | Wong et al. |
| Dallo | Filipino | Other Asian | Wong et al. |
| Dalmacio | Filipino | Other Asian | Wong et al. |
| Dalope | Filipino | Other Asian | Wong et al. |
| Dalsania | Asian Indian | South Asian | Wong et al. |
| Dalumpines | Filipino | Other Asian | Wong et al. |
| Dalupan | Filipino | Other Asian | Wong et al. |
| Daluraya | Filipino | Other Asian | Wong et al. |
| Daluro | Filipino | Other Asian | Wong et al. |
| Dalusong | Filipino | Other Asian | Wong et al. |
| Dalusung | Filipino | Other Asian | Wong et al. |
| Daluz | Filipino | Other Asian | Wong et al. |
| Dalvi | Asian Indian | South Asian | Wong et al. |
| Dalwadi | Asian Indian | South Asian | Wong et al. |
| Dam | Vietnamese | Other Asian | Wong et al. |
| Damalerio | Filipino | Other Asian | Wong et al. |
| Damani | Asian Indian | South Asian | Wong et al. |
| Damania | Asian Indian | South Asian | Wong et al. |
| Damasco | Filipino | Other Asian | Wong et al. |
| Damasen | Filipino | Other Asian | Wong et al. |
| Damaso | Filipino | Other Asian | Wong et al. |
| Damatan | Filipino | Other Asian | Wong et al. |
| Damazo | Filipino | Other Asian | Wong et al. |
| Damian | Filipino | Other Asian | Wong et al. |
| Damji | Asian Indian | South Asian | Wong et al. |
| Damle | Asian Indian | South Asian | Wong et al. |
| Damo | Filipino | Other Asian | Wong et al. |
| Damodaran | Asian Indian | South Asian | Wong et al. |
| Dampil | Filipino | Other Asian | Wong et al. |
| Dan | Chinese | Chinese | Wong et al. |
| Danak | Asian Indian | South Asian | Wong et al. |
| Danan | Filipino | Other Asian | Wong et al. |
| Danao | Filipino | Other Asian | Wong et al. |
| Dancel | Filipino | Other Asian | Wong et al. |
| Dancil | Filipino | Other Asian | Wong et al. |
| Dandan | Filipino | Other Asian | Wong et al. |
| Dandekar | Asian Indian | South Asian | Wong et al. |
| Dandin | Filipino | Other Asian | Wong et al. |
| Dandoy | Filipino | Other Asian | Wong et al. |
| Danetaras | Filipino | Other Asian | Wong et al. |
| Dang | Vietnamese | Other Asian | Wong et al. |
| Danga | Filipino | Other Asian | Wong et al. |
| Dangan | Filipino | Other Asian | Wong et al. |
| Danganan | Filipino | Other Asian | Wong et al. |
| Dangaran | Filipino | Other Asian | Wong et al. |
| Dangca | Filipino | Other Asian | Wong et al. |
| Dango | Filipino | Other Asian | Wong et al. |
| Dangoy | Filipino | Other Asian | Wong et al. |
| Danguilan | Filipino | Other Asian | Wong et al. |
| Danh | Vietnamese | Other Asian | Wong et al. |
| Danila | Filipino | Other Asian | Wong et al. |
| Danlag | Filipino | Other Asian | Wong et al. |
| Dannog | Filipino | Other Asian | Wong et al. |
| Dannug | Filipino | Other Asian | Wong et al. |
| Dano | Filipino | Other Asian | Wong et al. |
| Danque | Filipino | Other Asian | Wong et al. |
| Dansalan | Filipino | Other Asian | Wong et al. |
| Dantes | Filipino | Other Asian | Wong et al. |
| Dantic | Filipino | Other Asian | Wong et al. |
| Dantis | Filipino | Other Asian | Wong et al. |
| Dantoc | Filipino | Other Asian | Wong et al. |
| Dao | Vietnamese | Other Asian | Wong et al. |
| Daoang | Filipino | Other Asian | Wong et al. |
| Daomilas | Filipino | Other Asian | Wong et al. |
| Daos | Filipino | Other Asian | Wong et al. |
| Dap | Chinese | Chinese | Wong et al. |
| Dapar | Filipino | Other Asian | Wong et al. |
| Dapat | Filipino | Other Asian | Wong et al. |
| Daproza | Filipino | Other Asian | Wong et al. |
| Daquep | Filipino | Other Asian | Wong et al. |
| Daquigan | Filipino | Other Asian | Wong et al. |
| Daquil | Filipino | Other Asian | Wong et al. |
| Daquioag | Filipino | Other Asian | Wong et al. |
| Daquis | Filipino | Other Asian | Wong et al. |
| Daquiz | Filipino | Other Asian | Wong et al. |
| Daral | Filipino | Other Asian | Wong et al. |
| Daranciang | Filipino | Other Asian | Wong et al. |
| Darang | Filipino | Other Asian | Wong et al. |
| Darantinao | Filipino | Other Asian | Wong et al. |
| Darbari | Asian Indian | South Asian | Wong et al. |
| Daria | Filipino | Other Asian | Wong et al. |
| Dario | Filipino | Other Asian | Wong et al. |
| Darji | Asian Indian | South Asian | Wong et al. |
| Darlucio | Filipino | Other Asian | Wong et al. |
| Daroowalla | Asian Indian | South Asian | Wong et al. |
| Darosa | Chinese | Chinese | Wong et al. |
| Daroy | Filipino | Other Asian | Wong et al. |
| Daroya | Filipino | Other Asian | Wong et al. |
| Daroza | Chinese | Chinese | Wong et al. |
| Darunday | Filipino | Other Asian | Wong et al. |
| Daruvala | Asian Indian | South Asian | Wong et al. |
| Daruvalla | Asian Indian | South Asian | Wong et al. |
| Daruwala | Asian Indian | South Asian | Wong et al. |
| Daruwalla | Asian Indian | South Asian | Wong et al. |
| Das | Asian Indian; Bangladeshi | South Asian | Kranick et al. |
| Dasal | Filipino | Other Asian | Wong et al. |
| Dasalla | Filipino | Other Asian | Wong et al. |
| Dasappa | Asian Indian | South Asian | Wong et al. |
| Dasari | Asian Indian | South Asian | Wong et al. |
| Dasgupta |  | South Asian | Lim et al. |
| Dasig | Filipino | Other Asian | Wong et al. |
| Dasmarinas | Filipino | Other Asian | Wong et al. |
| Dass | Asian Indian | South Asian | Kranick et al. |
| Dastur | Asian Indian | South Asian | Wong et al. |
| Dat | Vietnamese; Asian Indian | Other Asian | Kranick et al. |
| Datangel | Filipino | Other Asian | Wong et al. |
| Datar | Asian Indian | South Asian | Wong et al. |
| Date | Japanese | Other Asian | Wong et al. |
| Datiles | Filipino | Other Asian | Wong et al. |
| Dato | Filipino | Other Asian | Wong et al. |
| Datoc | Filipino | Other Asian | Wong et al. |
| Datoon | Filipino | Other Asian | Wong et al. |
| Dator | Filipino | Other Asian | Wong et al. |
| Datt | Asian Indian | South Asian | Kranick et al. |
| Datta | Asian Indian; Bangladeshi | South Asian | Kranick et al. |
| Dattani | Asian Indian | South Asian | Wong et al. |
| Datu | Filipino | Other Asian | Wong et al. |
| Datugan | Filipino | Other Asian | Wong et al. |
| Datuin | Filipino | Other Asian | Wong et al. |
| Dau | Vietnamese | Other Asian | Wong et al. |
| Daud | Pakistani; Malaysian; Indonesian | South Asian | Kranick et al. |
| Daudi | Asian Indian | South Asian | Wong et al. |
| Daug | Filipino | Other Asian | Wong et al. |
| Dauglash | Filipino | Other Asian | Wong et al. |
| Dauz | Filipino | Other Asian | Wong et al. |
| Davadilla | Filipino | Other Asian | Wong et al. |
| Davalos | Filipino | Other Asian | Wong et al. |
| Davar | Asian Indian | South Asian | Wong et al. |
| Dave | Asian Indian | South Asian | Wong et al. |
| Daver | Asian Indian | South Asian | Wong et al. |
| Davila | Filipino | Other Asian | Wong et al. |
| Davo | Filipino | Other Asian | Wong et al. |
| Davocol | Filipino | Other Asian | Wong et al. |
| Dawa | Filipino | Other Asian | Wong et al. |
| Dawan |  | South Asian | Lim et al. |
| Dawang | Filipino | Other Asian | Wong et al. |
| Dawar | Asian Indian | South Asian | Wong et al. |
| Dawis | Filipino | Other Asian | Wong et al. |
| Dawood | Asian Indian | South Asian | Wong et al. |
| Day |  | South Asian | Lim et al. |
| Daya | Asian Indian | South Asian | Wong et al. |
| Dayabhai | Asian Indian | South Asian | Wong et al. |
| Dayag | Filipino | Other Asian | Wong et al. |
| Dayal | Asian Indian | South Asian | Wong et al. |
| Dayalji | Asian Indian | South Asian | Wong et al. |
| Dayan | Filipino | Other Asian | Wong et al. |
| Dayanan | Filipino | Other Asian | Wong et al. |
| Dayandante | Filipino | Other Asian | Wong et al. |
| Dayanghirang | Filipino | Other Asian | Wong et al. |
| Dayani | Asian Indian | South Asian | Wong et al. |
| Dayao | Filipino | Other Asian | Wong et al. |
| Dayap | Filipino | Other Asian | Wong et al. |
| Dayaram | Asian Indian | South Asian | Wong et al. |
| Dayco | Filipino | Other Asian | Wong et al. |
| Dayday | Filipino | Other Asian | Wong et al. |
| Daylo | Filipino | Other Asian | Wong et al. |
| Dayo | Filipino | Other Asian | Wong et al. |
| Dayoan | Filipino | Other Asian | Wong et al. |
| Dayok | Filipino | Other Asian | Wong et al. |
| Dayot | Filipino | Other Asian | Wong et al. |
| Dayrit | Filipino | Other Asian | Wong et al. |
| Daysog | Filipino | Other Asian | Wong et al. |
| Daza | Filipino | Other Asian | Wong et al. |
| Dazo | Filipino | Other Asian | Wong et al. |
| Dazon | Filipino | Other Asian | Wong et al. |
| Dcosta | Asian Indian | South Asian | Wong et al. |
| Dcruz | Asian Indian | South Asian | Wong et al. |
| Dcunha | Asian Indian | South Asian | Wong et al. |
| de Castro | Filipino | Other Asian | Kranick et al. |
| de Dios | Filipino | Other Asian | Kranick et al. |
| Dea | Chinese | Chinese | Wong et al. |
| Deala | Filipino | Other Asian | Wong et al. |
| Deang | Filipino | Other Asian | Wong et al. |
| Deano | Filipino | Other Asian | Wong et al. |
| Deaquino | Filipino | Other Asian | Wong et al. |
| Dearce | Filipino | Other Asian | Wong et al. |
| Deasis | Filipino | Other Asian | Wong et al. |
| Deato | Filipino | Other Asian | Wong et al. |
| Deauna | Filipino | Other Asian | Wong et al. |
| Deausen | Filipino | Other Asian | Wong et al. |
| Deb | Asian Indian | South Asian | Wong et al. |
| Debelen | Filipino | Other Asian | Wong et al. |
| Debi |  | South Asian | Lim et al. |
| Debiprashad |  | South Asian | Lim et al. |
| Debnath |  | South Asian | Lim et al. |
| Deboo | Asian Indian | South Asian | Wong et al. |
| Deborja | Filipino | Other Asian | Wong et al. |
| Decano | Filipino | Other Asian | Wong et al. |
| Decasa | Filipino | Other Asian | Wong et al. |
| Decastro | Filipino | Other Asian | Wong et al. |
| Deceased | Filipino | Other Asian | Wong et al. |
| Decelis | Filipino | Other Asian | Wong et al. |
| Decena | Filipino | Other Asian | Wong et al. |
| Decepida | Filipino | Other Asian | Wong et al. |
| Dechavez | Filipino | Other Asian | Wong et al. |
| Declaro | Filipino | Other Asian | Wong et al. |
| Decouto | Japanese | Other Asian | Wong et al. |
| Dedal | Filipino | Other Asian | Wong et al. |
| Dedicatoria | Filipino | Other Asian | Wong et al. |
| Dedios | Filipino | Other Asian | Wong et al. |
| Deen | Asian Indian | South Asian | Kranick et al. |
| Deepa |  | South Asian | Lim et al. |
| Deer | Chinese | Chinese | Wong et al. |
| Defensor | Filipino | Other Asian | Wong et al. |
| Defiesta | Filipino | Other Asian | Wong et al. |
| Defrancia | Filipino | Other Asian | Wong et al. |
| Defu | Chinese | Chinese | Wong et al. |
| Defuntorum | Filipino | Other Asian | Wong et al. |
| Degala | Filipino | Other Asian | Wong et al. |
| Degalicia | Filipino | Other Asian | Wong et al. |
| Degamo | Filipino | Other Asian | Wong et al. |
| Degarcia | Filipino | Other Asian | Wong et al. |
| Degayo | Filipino | Other Asian | Wong et al. |
| Degollado | Filipino | Other Asian | Wong et al. |
| Degoma | Filipino | Other Asian | Wong et al. |
| Degorostiza | Filipino | Other Asian | Wong et al. |
| Degracia | Filipino | Other Asian | Wong et al. |
| Degrano | Filipino | Other Asian | Wong et al. |
| Degroot |  | South Asian | Lim et al. |
| Deguchi | Japanese | Other Asian | Wong et al. |
| Deguia | Filipino | Other Asian | Wong et al. |
| Deguino | Filipino | Other Asian | Wong et al. |
| Degula | Filipino | Other Asian | Wong et al. |
| Degusman | Filipino | Other Asian | Wong et al. |
| Deguzman | Filipino | Other Asian | Kranick et al. |
| Dehesa | Filipino | Other Asian | Wong et al. |
| Dehitta | Filipino | Other Asian | Wong et al. |
| Dehlavi | Asian Indian | South Asian | Wong et al. |
| Deiparine | Filipino | Other Asian | Wong et al. |
| Dejano | Filipino | Other Asian | Wong et al. |
| Dejesa | Filipino | Other Asian | Wong et al. |
| Dejesus | Filipino | Other Asian | Wong et al. |
| Dejos | Filipino | Other Asian | Wong et al. |
| Dejoya | Filipino | Other Asian | Wong et al. |
| Dejuan | Filipino | Other Asian | Wong et al. |
| Dela Rosa | Filipino | Other Asian | Kranick et al. |
| Delacalzad | Filipino | Other Asian | Wong et al. |
| Delacalzada | Filipino | Other Asian | Wong et al. |
| Delacerna | Filipino | Other Asian | Wong et al. |
| Delacruz | Filipino | Other Asian | Wong et al. |
| Delacuadra | Filipino | Other Asian | Wong et al. |
| Delacuesta | Filipino | Other Asian | Wong et al. |
| Delacueva | Filipino | Other Asian | Wong et al. |
| Delacurz | Filipino | Other Asian | Wong et al. |
| Delafuente | Filipino | Other Asian | Wong et al. |
| Delagana | Filipino | Other Asian | Wong et al. |
| Delaisla | Filipino | Other Asian | Wong et al. |
| Delalcazar | Filipino | Other Asian | Wong et al. |
| Delamerced | Filipino | Other Asian | Wong et al. |
| Delansig | Filipino | Other Asian | Wong et al. |
| Delantar | Filipino | Other Asian | Wong et al. |
| Delapasion | Filipino | Other Asian | Wong et al. |
| Delapaz | Filipino | Other Asian | Wong et al. |
| Delapena | Filipino | Other Asian | Wong et al. |
| Delapina | Filipino | Other Asian | Wong et al. |
| Delara | Filipino | Other Asian | Wong et al. |
| Delarama | Filipino | Other Asian | Wong et al. |
| Delarea | Filipino | Other Asian | Wong et al. |
| Delaresma | Filipino | Other Asian | Wong et al. |
| Delarmente | Filipino | Other Asian | Wong et al. |
| Delaroca | Filipino | Other Asian | Wong et al. |
| Delarosa | Filipino | Other Asian | Wong et al. |
| Delasalas | Filipino | Other Asian | Wong et al. |
| Delaserna | Filipino | Other Asian | Wong et al. |
| Delatorre | Filipino | Other Asian | Wong et al. |
| Delavega | Filipino | Other Asian | Wong et al. |
| Delavictori | Filipino | Other Asian | Wong et al. |
| Delavictoria | Filipino | Other Asian | Wong et al. |
| Delavin | Filipino | Other Asian | Wong et al. |
| Delayon | Filipino | Other Asian | Wong et al. |
| Delbarrio | Filipino | Other Asian | Wong et al. |
| Delcarmen | Filipino | Other Asian | Wong et al. |
| Delcastillo | Filipino | Other Asian | Wong et al. |
| Delda | Filipino | Other Asian | Wong et al. |
| Delemos | Filipino | Other Asian | Wong et al. |
| Deleon | Filipino | Other Asian | Kranick et al. |
| Deleoz | Filipino | Other Asian | Wong et al. |
| Deles | Filipino | Other Asian | Wong et al. |
| Delfierro | Filipino | Other Asian | Wong et al. |
| Delfin | Filipino | Other Asian | Kranick et al. |
| Delfinado | Filipino | Other Asian | Wong et al. |
| Delgado | Filipino | Other Asian | Wong et al. |
| Delgallego | Filipino | Other Asian | Wong et al. |
| Deligero | Filipino | Other Asian | Wong et al. |
| Delim | Filipino | Other Asian | Wong et al. |
| Delima | Filipino | Other Asian | Wong et al. |
| Delizo | Filipino | Other Asian | Wong et al. |
| Della | Filipino | Other Asian | Wong et al. |
| Delloro | Filipino | Other Asian | Wong et al. |
| Delmendo | Filipino | Other Asian | Wong et al. |
| Delmoral | Filipino | Other Asian | Wong et al. |
| Delmundo | Filipino | Other Asian | Wong et al. |
| Deloria | Filipino | Other Asian | Wong et al. |
| Delosangel | Filipino | Other Asian | Wong et al. |
| Delosangele | Filipino | Other Asian | Wong et al. |
| Delosangeles | Filipino | Other Asian | Wong et al. |
| Delosantos | Filipino | Other Asian | Wong et al. |
| Deloso | Filipino | Other Asian | Wong et al. |
| Delosreyes | Filipino | Other Asian | Wong et al. |
| Delossanto | Filipino | Other Asian | Wong et al. |
| Delossantos | Filipino | Other Asian | Wong et al. |
| Delostrino | Filipino | Other Asian | Wong et al. |
| Delostrinos | Filipino | Other Asian | Wong et al. |
| Delovino | Filipino | Other Asian | Wong et al. |
| Delpilar | Filipino | Other Asian | Wong et al. |
| Delprado | Filipino | Other Asian | Wong et al. |
| Delpuerto | Filipino | Other Asian | Wong et al. |
| Delrio | Filipino | Other Asian | Wong et al. |
| Delrosario | Filipino | Other Asian | Wong et al. |
| Delumen | Filipino | Other Asian | Wong et al. |
| Delumpa | Filipino | Other Asian | Wong et al. |
| Deluna | Filipino | Other Asian | Wong et al. |
| Delute | Filipino | Other Asian | Wong et al. |
| Deluzuriaga | Filipino | Other Asian | Wong et al. |
| Delvalle | Filipino | Other Asian | Wong et al. |
| Delvillar | Filipino | Other Asian | Wong et al. |
| Demaala | Filipino | Other Asian | Wong et al. |
| Demafeliz | Filipino | Other Asian | Wong et al. |
| Demaisip | Filipino | Other Asian | Wong et al. |
| Demandante | Filipino | Other Asian | Wong et al. |
| Demanuel | Filipino | Other Asian | Wong et al. |
| Demee | Chinese | Chinese | Wong et al. |
| Demello | Asian Indian | South Asian | Wong et al. |
| Demerin | Filipino | Other Asian | Wong et al. |
| Demesa | Filipino | Other Asian | Wong et al. |
| Demetillo | Filipino | Other Asian | Wong et al. |
| Demetria | Filipino | Other Asian | Wong et al. |
| Demira | Filipino | Other Asian | Wong et al. |
| Demonteverde | Filipino | Other Asian | Wong et al. |
| Demoreta | Filipino | Other Asian | Wong et al. |
| Den | Chinese | Chinese | Wong et al. |
| Deng | Chinese | Chinese | Kranick et al. |
| Denh | Vietnamese | Other Asian | Wong et al. |
| Deniega | Filipino | Other Asian | Wong et al. |
| Denieva | Filipino | Other Asian | Wong et al. |
| Denina | Filipino | Other Asian | Wong et al. |
| Denosta | Filipino | Other Asian | Wong et al. |
| Deo | Asian Indian | South Asian | Wong et al. |
| Deocales | Filipino | Other Asian | Wong et al. |
| Deocampo | Filipino | Other Asian | Wong et al. |
| Deocares | Filipino | Other Asian | Wong et al. |
| Deocariza | Filipino | Other Asian | Wong et al. |
| Deocera | Filipino | Other Asian | Wong et al. |
| Deodat |  | South Asian | Lim et al. |
| Deodhar | Asian Indian | South Asian | Wong et al. |
| Deogracias | Filipino | Other Asian | Wong et al. |
| Deoki |  | South Asian | Lim et al. |
| Deokiesingh |  | South Asian | Lim et al. |
| Deol | Asian Indian | South Asian | Wong et al. |
| Deona | Filipino | Other Asian | Wong et al. |
| Deonarain |  | South Asian | Lim et al. |
| Deonarine |  | South Asian | Lim et al. |
| Deowdharrie |  | South Asian | Lim et al. |
| Dep | Chinese | Chinese | Wong et al. |
| Depadua | Filipino | Other Asian | Wong et al. |
| Depakakibo | Filipino | Other Asian | Wong et al. |
| Depano | Filipino | Other Asian | Wong et al. |
| Depante | Filipino | Other Asian | Wong et al. |
| Depaz | Filipino | Other Asian | Wong et al. |
| Deperalta | Filipino | Other Asian | Wong et al. |
| Deperio | Filipino | Other Asian | Wong et al. |
| Depra | Filipino | Other Asian | Wong et al. |
| Deputron |  | South Asian | Lim et al. |
| Dequilla | Filipino | Other Asian | Wong et al. |
| Dequina | Filipino | Other Asian | Wong et al. |
| Dequiroz | Filipino | Other Asian | Wong et al. |
| Dequit | Filipino | Other Asian | Wong et al. |
| Dequito | Filipino | Other Asian | Wong et al. |
| Der | Chinese | Chinese | Wong et al. |
| Derama | Filipino | Other Asian | Wong et al. |
| Deramos | Filipino | Other Asian | Wong et al. |
| Dere | Chinese | Chinese | Wong et al. |
| Derecho | Filipino | Other Asian | Wong et al. |
| Deregla | Filipino | Other Asian | Wong et al. |
| Derequito | Filipino | Other Asian | Wong et al. |
| Deriada | Filipino | Other Asian | Wong et al. |
| Derige | Filipino | Other Asian | Wong et al. |
| Derilo | Filipino | Other Asian | Wong et al. |
| Deriquito | Filipino | Other Asian | Wong et al. |
| Derivera | Filipino | Other Asian | Wong et al. |
| Derla | Filipino | Other Asian | Wong et al. |
| Derosas | Filipino | Other Asian | Wong et al. |
| Deroxas | Filipino | Other Asian | Wong et al. |
| Desa | Asian Indian | South Asian | Wong et al. |
| Desagun | Filipino | Other Asian | Wong et al. |
| Desai | Asian Indian | South Asian | Kranick et al. |
| Desal | Asian Indian | South Asian | Wong et al. |
| Desales | Filipino | Other Asian | Wong et al. |
| Desamero | Filipino | Other Asian | Wong et al. |
| Desamito | Filipino | Other Asian | Wong et al. |
| Desantos | Filipino | Other Asian | Wong et al. |
| Descallar | Filipino | Other Asian | Wong et al. |
| Descalzo | Filipino | Other Asian | Wong et al. |
| Descargar | Filipino | Other Asian | Wong et al. |
| Descartin | Filipino | Other Asian | Wong et al. |
| Desembrana | Filipino | Other Asian | Wong et al. |
| Desenna | Chinese | Chinese | Wong et al. |
| Deseo | Filipino | Other Asian | Wong et al. |
| Deshmukh | Asian Indian | South Asian | Wong et al. |
| Deshpande | Asian Indian | South Asian | Wong et al. |
| Desierto | Filipino | Other Asian | Wong et al. |
| Desikachar | Asian Indian | South Asian | Wong et al. |
| Deslate | Filipino | Other Asian | Wong et al. |
| Desouza | Asian Indian | South Asian | Wong et al. |
| Despe | Filipino | Other Asian | Wong et al. |
| Despi | Filipino | Other Asian | Wong et al. |
| Despues | Filipino | Other Asian | Wong et al. |
| Desquitado | Filipino | Other Asian | Wong et al. |
| Destacamento | Filipino | Other Asian | Wong et al. |
| Destajo | Filipino | Other Asian | Wong et al. |
| Destura | Filipino | Other Asian | Wong et al. |
| Desuyo | Filipino | Other Asian | Wong et al. |
| Desvarro | Filipino | Other Asian | Wong et al. |
| Detablan | Filipino | Other Asian | Wong et al. |
| Detera | Filipino | Other Asian | Wong et al. |
| Detorres | Filipino | Other Asian | Wong et al. |
| Deuk | Korean | Other Asian | Wong et al. |
| Deungria | Filipino | Other Asian | Wong et al. |
| Deuz | Filipino | Other Asian | Wong et al. |
| Dev |  | South Asian | Lim et al. |
| Devadas | Asian Indian | South Asian | Wong et al. |
| Devadoss | Asian Indian | South Asian | Wong et al. |
| Devaprasad | Asian Indian | South Asian | Wong et al. |
| Devaraj | Asian Indian | South Asian | Wong et al. |
| Devarajan | Asian Indian | South Asian | Wong et al. |
| Devasagayam | Asian Indian | South Asian | Wong et al. |
| Devasahayam | Asian Indian | South Asian | Wong et al. |
| Devasia | Asian Indian | South Asian | Wong et al. |
| Devassy | Asian Indian | South Asian | Wong et al. |
| Deveas | Filipino | Other Asian | Wong et al. |
| Devega | Filipino | Other Asian | Wong et al. |
| Devela | Filipino | Other Asian | Wong et al. |
| Devenecia | Filipino | Other Asian | Wong et al. |
| Devera | Filipino | Other Asian | Wong et al. |
| Deveras | Filipino | Other Asian | Wong et al. |
| Deveyra | Filipino | Other Asian | Wong et al. |
| Deveza | Filipino | Other Asian | Wong et al. |
| Devi | Asian Indian | South Asian | Wong et al. |
| Devika |  | South Asian | Lim et al. |
| Devilla | Filipino | Other Asian | Wong et al. |
| Devitre | Asian Indian | South Asian | Wong et al. |
| Devji | Asian Indian | South Asian | Wong et al. |
| Devnath |  | South Asian | Lim et al. |
| Devraj | Asian Indian | South Asian | Wong et al. |
| Dewa | Japanese | Other Asian | Wong et al. |
| Dewan | Asian Indian | South Asian | Wong et al. |
| Dey | Asian Indian; Bangladeshi | South Asian | Kranick et al. |
| Deypalubos | Filipino | Other Asian | Wong et al. |
| Deyro | Filipino | Other Asian | Wong et al. |
| Deyto | Filipino | Other Asian | Wong et al. |
| Deza | Filipino | Other Asian | Wong et al. |
| Dhadda | Asian Indian | South Asian | Wong et al. |
| Dhagat | Asian Indian | South Asian | Wong et al. |
| Dhali |  | South Asian | Lim et al. |
| Dhaliwal | Asian Indian | South Asian | Wong et al. |
| Dhall | Asian Indian | South Asian | Wong et al. |
| Dham | Vietnamese | Other Asian | Wong et al. |
| Dhamani | Asian Indian | South Asian | Wong et al. |
| Dhami | Asian Indian | South Asian | Wong et al. |
| Dhamija | Asian Indian | South Asian | Wong et al. |
| Dhan | Vietnamese | Other Asian | Wong et al. |
| Dhanani | Asian Indian | South Asian | Wong et al. |
| Dhanapala | Sri Lankan | South Asian | Kranick et al. |
| Dhanaraj | Asian Indian | South Asian | Wong et al. |
| Dhand | Asian Indian | South Asian | Wong et al. |
| Dhanda | Asian Indian | South Asian | Wong et al. |
| Dhanjal | Asian Indian | South Asian | Wong et al. |
| Dhanji | Asian Indian | South Asian | Wong et al. |
| Dhanoa | Asian Indian | South Asian | Wong et al. |
| Dhanoya | Asian Indian | South Asian | Wong et al. |
| Dhanraj |  | South Asian | Lim et al. |
| Dhar | Asian Indian | South Asian | Wong et al. |
| Dharamsey | Asian Indian | South Asian | Wong et al. |
| Dharia | Asian Indian | South Asian | Wong et al. |
| Dhariwal | Asian Indian | South Asian | Wong et al. |
| Dharmarajan | Asian Indian | South Asian | Wong et al. |
| Dhatt | Asian Indian | South Asian | Wong et al. |
| Dhawan | Asian Indian | South Asian | Wong et al. |
| Dheer | Asian Indian | South Asian | Wong et al. |
| Dhesi | Asian Indian | South Asian | Wong et al. |
| Dhillon | Asian Indian | South Asian | Wong et al. |
| Dhiman | Asian Indian | South Asian | Wong et al. |
| Dhindsa | Asian Indian | South Asian | Wong et al. |
| Dhir | Asian Indian | South Asian | Wong et al. |
| Dhirsingh |  | South Asian | Lim et al. |
| Dhody | Asian Indian | South Asian | Wong et al. |
| Dholakia | Asian Indian | South Asian | Wong et al. |
| Dhond | Asian Indian | South Asian | Wong et al. |
| Dhondy | Asian Indian | South Asian | Wong et al. |
| Dhoot | Asian Indian | South Asian | Wong et al. |
| Dhruv | Asian Indian | South Asian | Wong et al. |
| Dhruva | Asian Indian | South Asian | Wong et al. |
| Dhruve | Asian Indian | South Asian | Wong et al. |
| Dhungel |  | South Asian | Lim et al. |
| Dhurandhar | Asian Indian | South Asian | Wong et al. |
| Dhursan |  | South Asian | Lim et al. |
| Dhuru | Asian Indian | South Asian | Wong et al. |
| Di | Chinese | Chinese | Wong et al. |
| Dia | Filipino | Other Asian | Wong et al. |
| Diacuna | Filipino | Other Asian | Wong et al. |
| Diagan | Filipino | Other Asian | Wong et al. |
| Diago | Filipino | Other Asian | Wong et al. |
| Diala | Filipino | Other Asian | Wong et al. |
| Diamante | Filipino | Other Asian | Wong et al. |
| Diamsay | Filipino | Other Asian | Wong et al. |
| Diangson | Filipino | Other Asian | Wong et al. |
| Diano | Filipino | Other Asian | Wong et al. |
| Diao | Chinese | Chinese | Wong et al. |
| Diapo | Filipino | Other Asian | Wong et al. |
| Dias | Asian Indian | South Asian | Wong et al. |
| Diaz | Filipino | Other Asian | Wong et al. |
| Dicang | Filipino | Other Asian | Wong et al. |
| Diccion | Filipino | Other Asian | Wong et al. |
| Dicen | Filipino | Other Asian | Wong et al. |
| Dich | Vietnamese | Other Asian | Wong et al. |
| Dichosa | Filipino | Other Asian | Wong et al. |
| Dichoso | Filipino | Other Asian | Wong et al. |
| Dicion | Filipino | Other Asian | Wong et al. |
| Diec | Chinese | Chinese | Wong et al. |
| Diego | Filipino | Other Asian | Wong et al. |
| Diem | Vietnamese | Other Asian | Wong et al. |
| Dien | Vietnamese | Other Asian | Wong et al. |
| Dieng | Vietnamese | Other Asian | Wong et al. |
| Diep | Vietnamese | Other Asian | Wong et al. |
| Diesta | Filipino | Other Asian | Wong et al. |
| Diesto | Filipino | Other Asian | Wong et al. |
| Diestro | Filipino | Other Asian | Wong et al. |
| Diet | Vietnamese | Other Asian | Wong et al. |
| Dieu | Vietnamese | Other Asian | Wong et al. |
| Diez | Filipino | Other Asian | Wong et al. |
| Diezmo | Filipino | Other Asian | Wong et al. |
| Difuntorum | Filipino | Other Asian | Wong et al. |
| Dig | Filipino | Other Asian | Wong et al. |
| Diga | Filipino | Other Asian | Wong et al. |
| Digal | Filipino | Other Asian | Wong et al. |
| Digap | Filipino | Other Asian | Wong et al. |
| Digdigan | Filipino | Other Asian | Wong et al. |
| Dighe | Asian Indian | South Asian | Wong et al. |
| Digma | Filipino | Other Asian | Wong et al. |
| Dignadice | Filipino | Other Asian | Wong et al. |
| Diguangco | Filipino | Other Asian | Wong et al. |
| Dijamco | Filipino | Other Asian | Wong et al. |
| Dikit | Filipino | Other Asian | Wong et al. |
| Dikitanan | Filipino | Other Asian | Wong et al. |
| Dikshit | Asian Indian | South Asian | Wong et al. |
| Dila | Filipino | Other Asian | Wong et al. |
| Dilag | Filipino | Other Asian | Wong et al. |
| Dilan | Filipino | Other Asian | Wong et al. |
| Dilao | Filipino | Other Asian | Wong et al. |
| Dilara |  | South Asian | Lim et al. |
| Dilawari | Asian Indian | South Asian | Wong et al. |
| Dilay | Filipino | Other Asian | Wong et al. |
| Dilidili | Filipino | Other Asian | Wong et al. |
| Dilig | Filipino | Other Asian | Wong et al. |
| Diligencia | Filipino | Other Asian | Wong et al. |
| Dilla | Filipino | Other Asian | Wong et al. |
| Dillera | Filipino | Other Asian | Wong et al. |
| Diloy | Filipino | Other Asian | Wong et al. |
| Dimaandal | Filipino | Other Asian | Wong et al. |
| Dimaano | Filipino | Other Asian | Kranick et al. |
| Dimaapi | Filipino | Other Asian | Wong et al. |
| Dimacale | Filipino | Other Asian | Wong et al. |
| Dimacali | Filipino | Other Asian | Wong et al. |
| Dimaculangan | Filipino | Other Asian | Kranick et al. |
| Dimagiba | Filipino | Other Asian | Wong et al. |
| Dimagmaliw | Filipino | Other Asian | Wong et al. |
| Dimaguila | Filipino | Other Asian | Wong et al. |
| Dimailig | Filipino | Other Asian | Wong et al. |
| Dimaisip | Filipino | Other Asian | Wong et al. |
| Dimal | Filipino | Other Asian | Wong et al. |
| Dimalaluan | Filipino | Other Asian | Wong et al. |
| Dimalanta | Filipino | Other Asian | Kranick et al. |
| Dimanlig | Filipino | Other Asian | Wong et al. |
| Dimapasoc | Filipino | Other Asian | Wong et al. |
| Dimapilis | Filipino | Other Asian | Wong et al. |
| Dimaranan | Filipino | Other Asian | Wong et al. |
| Dimarucot | Filipino | Other Asian | Wong et al. |
| Dimarucut | Filipino | Other Asian | Wong et al. |
| Dimasuay | Filipino | Other Asian | Wong et al. |
| Dimatulac | Filipino | Other Asian | Wong et al. |
| Dimaunahan | Filipino | Other Asian | Wong et al. |
| Dimaya | Filipino | Other Asian | Wong et al. |
| Dimayuga | Filipino | Other Asian | Wong et al. |
| Dimazana | Filipino | Other Asian | Wong et al. |
| Dime | Filipino | Other Asian | Wong et al. |
| Dimen | Filipino | Other Asian | Wong et al. |
| Dimla | Filipino | Other Asian | Wong et al. |
| Dimson | Filipino | Other Asian | Wong et al. |
| Dimzon | Filipino | Other Asian | Wong et al. |
| Din |  | South Asian | Lim et al. |
| Dindial |  | South Asian | Lim et al. |
| Dinero | Filipino | Other Asian | Wong et al. |
| Dineros | Filipino | Other Asian | Wong et al. |
| Ding | Chinese | Chinese | Wong et al. |
| Dingal | Filipino | Other Asian | Wong et al. |
| Dingco | Filipino | Other Asian | Wong et al. |
| Dingcong | Filipino | Other Asian | Wong et al. |
| Dinglas | Filipino | Other Asian | Wong et al. |
| Dinglasa | Filipino | Other Asian | Wong et al. |
| Dinglasan | Filipino | Other Asian | Wong et al. |
| Dinh | Vietnamese | Other Asian | Wong et al. |
| Diniega | Filipino | Other Asian | Wong et al. |
| Dinio | Filipino | Other Asian | Wong et al. |
| Dino | Filipino | Other Asian | Wong et al. |
| Dinong | Filipino | Other Asian | Wong et al. |
| Dinoso | Filipino | Other Asian | Wong et al. |
| Dinsay | Filipino | Other Asian | Wong et al. |
| Dinshaw | Asian Indian | South Asian | Wong et al. |
| Dio | Filipino | Other Asian | Wong et al. |
| Diocares | Filipino | Other Asian | Wong et al. |
| Diocson | Filipino | Other Asian | Wong et al. |
| Diokno | Filipino | Other Asian | Wong et al. |
| Diola | Filipino | Other Asian | Wong et al. |
| Diolazo | Filipino | Other Asian | Wong et al. |
| Diolola | Filipino | Other Asian | Wong et al. |
| Diomampo | Filipino | Other Asian | Wong et al. |
| Dionaldo | Filipino | Other Asian | Wong et al. |
| Dioneda | Filipino | Other Asian | Wong et al. |
| Dionela | Filipino | Other Asian | Wong et al. |
| Diones | Filipino | Other Asian | Wong et al. |
| Dionglay | Filipino | Other Asian | Wong et al. |
| Diongzon | Filipino | Other Asian | Wong et al. |
| Dionicio | Filipino | Other Asian | Wong et al. |
| Dionido | Filipino | Other Asian | Wong et al. |
| Dionio | Filipino | Other Asian | Wong et al. |
| Dionisio | Filipino | Other Asian | Wong et al. |
| Dionson | Filipino | Other Asian | Wong et al. |
| Dioquino | Filipino | Other Asian | Wong et al. |
| Diosana | Filipino | Other Asian | Wong et al. |
| Dioso | Filipino | Other Asian | Wong et al. |
| Diosomito | Filipino | Other Asian | Wong et al. |
| Dip | Chinese | Chinese | Wong et al. |
| Dipa |  | South Asian | Lim et al. |
| Dipaling | Filipino | Other Asian | Wong et al. |
| Dipasupil | Filipino | Other Asian | Wong et al. |
| Diputado | Filipino | Other Asian | Wong et al. |
| Dira | Filipino | Other Asian | Wong et al. |
| Directo | Filipino | Other Asian | Wong et al. |
| Dirige | Filipino | Other Asian | Wong et al. |
| Discipulo | Filipino | Other Asian | Wong et al. |
| Dismaya | Filipino | Other Asian | Wong et al. |
| Disonglo | Filipino | Other Asian | Wong et al. |
| Disouza | Asian Indian | South Asian | Wong et al. |
| Dispo | Filipino | Other Asian | Wong et al. |
| Distajo | Filipino | Other Asian | Wong et al. |
| Distor | Filipino | Other Asian | Wong et al. |
| Ditalo | Filipino | Other Asian | Wong et al. |
| Ditching | Filipino | Other Asian | Wong et al. |
| Ditona | Filipino | Other Asian | Wong et al. |
| Diu | Chinese | Chinese | Wong et al. |
| Diuco | Filipino | Other Asian | Wong et al. |
| Divakaran | Asian Indian | South Asian | Wong et al. |
| Divatia | Asian Indian | South Asian | Wong et al. |
| Divecha | Asian Indian | South Asian | Wong et al. |
| Divekar | Asian Indian | South Asian | Wong et al. |
| Divina | Filipino | Other Asian | Wong et al. |
| Divinagracia | Filipino | Other Asian | Wong et al. |
| Divino | Filipino | Other Asian | Wong et al. |
| Diwa | Filipino | Other Asian | Wong et al. |
| Diwadkar | Asian Indian | South Asian | Wong et al. |
| Diwan | Asian Indian | South Asian | Wong et al. |
| Dixit | Asian Indian | South Asian | Wong et al. |
| Diy | Filipino | Other Asian | Wong et al. |
| Diza | Filipino | Other Asian | Wong et al. |
| Dizon | Filipino | Other Asian | Kranick et al. |
| Djang | Chinese | Chinese | Wong et al. |
| Djung | Chinese | Chinese | Wong et al. |
| Dlima | Asian Indian | South Asian | Wong et al. |
| Dmello | Asian Indian | South Asian | Wong et al. |
| Do | Vietnamese | Other Asian | Wong et al. |
| Doan | Vietnamese | Other Asian | Wong et al. |
| Dobashi | Japanese | Other Asian | Wong et al. |
| Doblon | Filipino | Other Asian | Wong et al. |
| Docena | Filipino | Other Asian | Wong et al. |
| Dock | Chinese | Chinese | Wong et al. |
| Docto | Filipino | Other Asian | Wong et al. |
| Doctolero | Filipino | Other Asian | Wong et al. |
| Doctor | Filipino | Other Asian | Wong et al. |
| Doctora | Filipino | Other Asian | Wong et al. |
| Doculan | Filipino | Other Asian | Wong et al. |
| Docuyanan | Filipino | Other Asian | Wong et al. |
| Dodia | Asian Indian | South Asian | Wong et al. |
| Dofredo | Filipino | Other Asian | Wong et al. |
| Dogar | Asian Indian | South Asian | Wong et al. |
| Dogoldogol | Filipino | Other Asian | Wong et al. |
| Dogra | Asian Indian | South Asian | Wong et al. |
| Doh | Korean | Other Asian | Wong et al. |
| Dohi | Japanese | Other Asian | Wong et al. |
| Doi | Japanese | Other Asian | Wong et al. |
| Doki | Japanese | Other Asian | Wong et al. |
| Dokko | Korean | Other Asian | Wong et al. |
| Dolar | Filipino | Other Asian | Wong et al. |
| Dolatre | Filipino | Other Asian | Wong et al. |
| Dolendo | Filipino | Other Asian | Wong et al. |
| Doliente | Filipino | Other Asian | Wong et al. |
| Doligosa | Filipino | Other Asian | Wong et al. |
| Dolino | Filipino | Other Asian | Wong et al. |
| Dolit | Filipino | Other Asian | Wong et al. |
| Dollaga | Filipino | Other Asian | Wong et al. |
| Dollente | Filipino | Other Asian | Wong et al. |
| Dollete | Filipino | Other Asian | Wong et al. |
| Dolleton | Filipino | Other Asian | Wong et al. |
| Dollosa | Filipino | Other Asian | Wong et al. |
| Dolma |  | South Asian | Lim et al. |
| Dolojan | Filipino | Other Asian | Wong et al. |
| Dolor | Filipino | Other Asian | Wong et al. |
| Dolores | Filipino | Other Asian | Wong et al. |
| Dolorfino | Filipino | Other Asian | Wong et al. |
| Dolormente | Filipino | Other Asian | Wong et al. |
| Doloroso | Filipino | Other Asian | Wong et al. |
| Doma | Filipino | Other Asian | Wong et al. |
| Domacena | Filipino | Other Asian | Wong et al. |
| Domadia | Asian Indian | South Asian | Wong et al. |
| Domagas | Filipino | Other Asian | Wong et al. |
| Domalanta | Filipino | Other Asian | Wong et al. |
| Domalaon | Filipino | Other Asian | Wong et al. |
| Domantay | Filipino | Other Asian | Wong et al. |
| Domaoal | Filipino | Other Asian | Wong et al. |
| Domaoan | Filipino | Other Asian | Wong et al. |
| Domasig | Filipino | Other Asian | Wong et al. |
| Domdom | Filipino | Other Asian | Wong et al. |
| Domen | Japanese | Other Asian | Wong et al. |
| Domenden | Filipino | Other Asian | Wong et al. |
| Domil | Filipino | Other Asian | Wong et al. |
| Dominado | Filipino | Other Asian | Wong et al. |
| Domincel | Filipino | Other Asian | Wong et al. |
| Domine | Filipino | Other Asian | Wong et al. |
| Domingcil | Filipino | Other Asian | Wong et al. |
| Domingo | Filpino | Other Asian | Kranick et al. |
| Domingsil | Filipino | Other Asian | Wong et al. |
| Domingues | Filipino | Other Asian | Wong et al. |
| Dominguez | Filipino | Other Asian | Wong et al. |
| Dominisac | Filipino | Other Asian | Wong et al. |
| Dominno | Filipino | Other Asian | Wong et al. |
| Dominquez | Filipino | Other Asian | Wong et al. |
| Domogma | Filipino | Other Asian | Wong et al. |
| Domondon | Filipino | Other Asian | Wong et al. |
| Domoto | Japanese | Other Asian | Wong et al. |
| Dompor | Filipino | Other Asian | Wong et al. |
| Domulot | Filipino | Other Asian | Wong et al. |
| Don | Pakistani; Bangladeshi; Sri Lankan | South Asian | Kranick et al. |
| Dona | Filipino | Other Asian | Wong et al. |
| Donado | Filipino | Other Asian | Wong et al. |
| Donasco | Filipino | Other Asian | Wong et al. |
| Donayre | Filipino | Other Asian | Wong et al. |
| Donde | Asian Indian | South Asian | Wong et al. |
| Dones | Filipino | Other Asian | Wong et al. |
| Donesa | Filipino | Other Asian | Wong et al. |
| Doneza | Filipino | Other Asian | Wong et al. |
| Dong | Chinese | Chinese | Kranick et al. |
| Dongallo | Filipino | Other Asian | Wong et al. |
| Dongon | Filipino | Other Asian | Wong et al. |
| Dongre | Asian Indian | South Asian | Wong et al. |
| Donguines | Filipino | Other Asian | Wong et al. |
| Donia | Filipino | Other Asian | Wong et al. |
| Donor | Filipino | Other Asian | Wong et al. |
| Donoso | Chinese | Chinese | Wong et al. |
| Doo | Chinese | Chinese | Wong et al. |
| Doobay |  | South Asian | Lim et al. |
| Doodnauth |  | South Asian | Lim et al. |
| Dooma | Filipino | Other Asian | Wong et al. |
| Doon | Chinese | Chinese | Wong et al. |
| Doong | Chinese | Chinese | Wong et al. |
| Dor | Chinese | Chinese | Wong et al. |
| Dorado | Filipino | Other Asian | Wong et al. |
| Dorairaj | Asian Indian | South Asian | Wong et al. |
| Doraiswami | Asian Indian | South Asian | Wong et al. |
| Doraiswamy | Asian Indian | South Asian | Wong et al. |
| Doral | Filipino | Other Asian | Wong et al. |
| Doras | Filipino | Other Asian | Wong et al. |
| Doria |  | South Asian | Lim et al. |
| Dorilag | Filipino | Other Asian | Wong et al. |
| Dorjee |  | South Asian | Lim et al. |
| Dormitorio | Filipino | Other Asian | Wong et al. |
| Doroja | Filipino | Other Asian | Wong et al. |
| Doromal | Filipino | Other Asian | Wong et al. |
| Doronila | Filipino | Other Asian | Wong et al. |
| Doronio | Filipino | Other Asian | Wong et al. |
| Doropan | Filipino | Other Asian | Wong et al. |
| Doroteo | Filipino | Other Asian | Wong et al. |
| Dorotheo | Filipino | Other Asian | Wong et al. |
| Dosado | Filipino | Other Asian | Wong et al. |
| Dosani | Asian Indian | South Asian | Wong et al. |
| Dosanjh | Asian Indian | South Asian | Wong et al. |
| Dosdos | Filipino | Other Asian | Wong et al. |
| Doshi | Asian Indian | South Asian | Kranick et al. |
| Dosono | Filipino | Other Asian | Wong et al. |
| Dosremedios | Chinese | Chinese | Wong et al. |
| Dossani | Asian Indian | South Asian | Wong et al. |
| Dossantos | Chinese | Chinese | Wong et al. |
| Dot | Chinese | Chinese | Wong et al. |
| Dote | Japanese | Other Asian | Wong et al. |
| Dotimas | Filipino | Other Asian | Wong et al. |
| Doton | Filipino | Other Asian | Wong et al. |
| Dou | Chinese | Chinese | Wong et al. |
| Doy | Chinese | Chinese | Wong et al. |
| Dozono | Japanese | Other Asian | Wong et al. |
| Draculan | Filipino | Other Asian | Wong et al. |
| Draksharam | Asian Indian | South Asian | Wong et al. |
| Drapiza | Filipino | Other Asian | Wong et al. |
| Draviam | Asian Indian | South Asian | Wong et al. |
| Dravid | Asian Indian | South Asian | Wong et al. |
| Drequito | Filipino | Other Asian | Wong et al. |
| Dres | Filipino | Other Asian | Wong et al. |
| Drilon | Filipino | Other Asian | Wong et al. |
| Driz | Filipino | Other Asian | Wong et al. |
| Drozario | Asian Indian | South Asian | Wong et al. |
| Dsa | Asian Indian | South Asian | Wong et al. |
| Dsilva | Asian Indian | South Asian | Wong et al. |
| Dsouza | Asian Indian | South Asian | Wong et al. |
| Dsylva | Asian Indian | South Asian | Wong et al. |
| Du | Chinese | Chinese | Kranick et al. |
| Dualan | Filipino | Other Asian | Wong et al. |
| Duan | Chinese | Chinese | Wong et al. |
| Duarosan | Filipino | Other Asian | Wong et al. |
| Duarte | Filipino | Other Asian | Wong et al. |
| Duatin | Filipino | Other Asian | Wong et al. |
| Duazo | Filipino | Other Asian | Wong et al. |
| Dubal | Asian Indian | South Asian | Wong et al. |
| Dubash | Asian Indian | South Asian | Wong et al. |
| Dube | Asian Indian | South Asian | Kranick et al. |
| Dubey | Asian Indian | South Asian | Kranick et al. |
| Duc | Vietnamese | Other Asian | Wong et al. |
| Ducay | Filipino | Other Asian | Wong et al. |
| Duclayan | Filipino | Other Asian | Wong et al. |
| Duco | Filipino | Other Asian | Wong et al. |
| Ducosin | Filipino | Other Asian | Wong et al. |
| Duculan | Filipino | Other Asian | Wong et al. |
| Ducusin | Filipino | Other Asian | Wong et al. |
| Ducut | Filipino | Other Asian | Wong et al. |
| Dudang | Filipino | Other Asian | Wong et al. |
| Dudhia | Asian Indian | South Asian | Wong et al. |
| Dudhnath |  | South Asian | Lim et al. |
| Duen | Chinese | Chinese | Wong et al. |
| Duenas | Filipino | Other Asian | Wong et al. |
| Dueno | Filipino | Other Asian | Wong et al. |
| Duero | Filipino | Other Asian | Wong et al. |
| Dugay | Filipino | Other Asian | Wong et al. |
| Dugenia | Filipino | Other Asian | Wong et al. |
| Duggal | Asian Indian | South Asian | Wong et al. |
| Duggirala | Asian Indian | South Asian | Wong et al. |
| Dugue | Filipino | Other Asian | Wong et al. |
| Duhaylongsod | Filipino | Other Asian | Wong et al. |
| Dui | Vietnamese | Other Asian | Wong et al. |
| Duk | Korean | Other Asian | Wong et al. |
| Duka | Filipino | Other Asian | Wong et al. |
| Dulai | Asian Indian | South Asian | Wong et al. |
| Dulalia | Filipino | Other Asian | Wong et al. |
| Dulatre | Filipino | Other Asian | Wong et al. |
| Dulay | Filipino | Other Asian | Wong et al. |
| Dulce | Filipino | Other Asian | Wong et al. |
| Duldulao | Filipino | Other Asian | Wong et al. |
| Dulnuan | Filipino | Other Asian | Wong et al. |
| Dulog | Filipino | Other Asian | Wong et al. |
| Dulos | Filipino | Other Asian | Wong et al. |
| Dumadag | Filipino | Other Asian | Wong et al. |
| Dumadaug | Filipino | Other Asian | Wong et al. |
| Dumag | Filipino | Other Asian | Wong et al. |
| Dumagan | Filipino | Other Asian | Wong et al. |
| Dumago | Filipino | Other Asian | Wong et al. |
| Dumaguin | Filipino | Other Asian | Wong et al. |
| Dumaguing | Filipino | Other Asian | Wong et al. |
| Dumalag | Filipino | Other Asian | Wong et al. |
| Dumalanta | Filipino | Other Asian | Wong et al. |
| Dumale | Filipino | Other Asian | Wong et al. |
| Dumancas | Filipino | Other Asian | Wong et al. |
| Dumandan | Filipino | Other Asian | Wong et al. |
| Dumangas | Filipino | Other Asian | Wong et al. |
| Dumanon | Filipino | Other Asian | Wong et al. |
| Dumaop | Filipino | Other Asian | Wong et al. |
| Dumapias | Filipino | Other Asian | Wong et al. |
| Dumapit | Filipino | Other Asian | Wong et al. |
| Dumaplin | Filipino | Other Asian | Wong et al. |
| Dumaran | Filipino | Other Asian | Wong et al. |
| Dumaual | Filipino | Other Asian | Wong et al. |
| Dumayag | Filipino | Other Asian | Wong et al. |
| Dumayas | Filipino | Other Asian | Wong et al. |
| Dumbrigue | Filipino | Other Asian | Wong et al. |
| Dumbrique | Filipino | Other Asian | Wong et al. |
| Dumdum | Filipino | Other Asian | Wong et al. |
| Dumlao | Filipino | Other Asian | Wong et al. |
| Dumo | Filipino | Other Asian | Wong et al. |
| Dumol | Filipino | Other Asian | Wong et al. |
| Dumot | Filipino | Other Asian | Wong et al. |
| Dumpit | Filipino | Other Asian | Wong et al. |
| Dumrique | Filipino | Other Asian | Wong et al. |
| Dumuk | Filipino | Other Asian | Wong et al. |
| Dun | Chinese | Chinese | Wong et al. |
| Dung | Chinese | Chinese | Wong et al. |
| Dungao | Filipino | Other Asian | Wong et al. |
| Dungca | Filipino | Other Asian | Wong et al. |
| Dungo | Filipino | Other Asian | Wong et al. |
| Dungog | Filipino | Other Asian | Wong et al. |
| Dunuan | Filipino | Other Asian | Wong et al. |
| Duoc | Vietnamese | Other Asian | Wong et al. |
| Duong | Vietnamese; Cambodian | Other Asian | Kranick et al. |
| Dupay |  | South Asian | Lim et al. |
| Dupaya | Filipino | Other Asian | Wong et al. |
| Duque | Filipino | Other Asian | Wong et al. |
| Duquilla | Filipino | Other Asian | Wong et al. |
| Durairaj | Asian Indian | South Asian | Wong et al. |
| Dural | Filipino | Other Asian | Wong et al. |
| Duran | Filipino | Other Asian | Wong et al. |
| Durana | Filipino | Other Asian | Wong et al. |
| Durano | Filipino | Other Asian | Wong et al. |
| Durbeej |  | South Asian | Lim et al. |
| Dureg | Filipino | Other Asian | Wong et al. |
| Duremdes | Filipino | Other Asian | Wong et al. |
| Dureza | Filipino | Other Asian | Wong et al. |
| Duria | Filipino | Other Asian | Wong et al. |
| Durias | Filipino | Other Asian | Wong et al. |
| Duro | Filipino | Other Asian | Wong et al. |
| Duropan | Filipino | Other Asian | Wong et al. |
| Durvasula | Asian Indian | South Asian | Wong et al. |
| Durve | Asian Indian | South Asian | Wong et al. |
| Dusaban | Filipino | Other Asian | Wong et al. |
| Duschane | Filipino | Other Asian | Wong et al. |
| Dusich | Filipino | Other Asian | Wong et al. |
| Duterte | Filipino | Other Asian | Wong et al. |
| Dutt | Asian Indian | South Asian | Wong et al. |
| Dutta | Asian Indian; Bangladeshi | South Asian | Kranick et al. |
| Duttaroy | Asian Indian | South Asian | Wong et al. |
| Duvvuri | Asian Indian | South Asian | Wong et al. |
| Duy | Vietnamese | Other Asian | Wong et al. |
| Duya | Filipino | Other Asian | Wong et al. |
| Duyag | Filipino | Other Asian | Wong et al. |
| Duyan | Filipino | Other Asian | Wong et al. |
| Duyanen | Filipino | Other Asian | Wong et al. |
| Duyao | Filipino | Other Asian | Wong et al. |
| Duyen | Vietnamese | Other Asian | Wong et al. |
| Duyet | Vietnamese | Other Asian | Wong et al. |
| Duyongco | Filipino | Other Asian | Wong et al. |
| Duzon | Filipino | Other Asian | Wong et al. |
| Dwarkadas | Asian Indian | South Asian | Wong et al. |
| Dwivedi | Asian Indian | South Asian | Wong et al. |
| Dy | Filipino | Other Asian | Wong et al. |
| Dychangco | Filipino | Other Asian | Wong et al. |
| Dychitan | Filipino | Other Asian | Wong et al. |
| Dycoco | Filipino | Other Asian | Wong et al. |
| Dyliacco | Filipino | Other Asian | Wong et al. |
| Dyogi | Filipino | Other Asian | Wong et al. |
| Dyreyes | Filipino | Other Asian | Wong et al. |
| Dysangco | Filipino | Other Asian | Wong et al. |
| Eachempati | Asian Indian | South Asian | Wong et al. |
| Eala | Filipino | Other Asian | Wong et al. |
| Eamiguel | Filipino | Other Asian | Wong et al. |
| Eamilao | Filipino | Other Asian | Wong et al. |
| Eapen | Asian Indian | South Asian | Wong et al. |
| Eappen | Asian Indian | South Asian | Wong et al. |
| Eaqub |  | South Asian | Lim et al. |
| Easaw | Asian Indian | South Asian | Wong et al. |
| Easmin |  | South Asian | Lim et al. |
| Easo | Asian Indian | South Asian | Wong et al. |
| Easow | Asian Indian | South Asian | Wong et al. |
| Eay | Filipino | Other Asian | Wong et al. |
| Ebalo | Filipino | Other Asian | Wong et al. |
| Ebalobo | Filipino | Other Asian | Wong et al. |
| Eban | Vietnamese | Other Asian | Wong et al. |
| Ebara | Japanese | Other Asian | Wong et al. |
| Ebarle | Filipino | Other Asian | Wong et al. |
| Ebarvia | Filipino | Other Asian | Wong et al. |
| Ebashi | Japanese | Other Asian | Wong et al. |
| Ebata | Japanese | Other Asian | Wong et al. |
| Ebba | Filipino | Other Asian | Wong et al. |
| Ebbat | Filipino | Other Asian | Wong et al. |
| Ebenezer | Asian Indian | South Asian | Wong et al. |
| Ebesu | Japanese | Other Asian | Wong et al. |
| Ebido | Filipino | Other Asian | Wong et al. |
| Ebihara | Japanese | Other Asian | Wong et al. |
| Ebilane | Filipino | Other Asian | Wong et al. |
| Ebina | Japanese | Other Asian | Wong et al. |
| Ebio | Filipino | Other Asian | Wong et al. |
| Ebisawa | Japanese | Other Asian | Wong et al. |
| Ebitner | Filipino | Other Asian | Wong et al. |
| Ebizuka | Japanese | Other Asian | Wong et al. |
| Ebojo | Filipino | Other Asian | Wong et al. |
| Ebon | Filipino | Other Asian | Wong et al. |
| Ebora | Filipino | Other Asian | Wong et al. |
| Ebrahim | Asian Indian | South Asian | Wong et al. |
| Ebreo | Filipino | Other Asian | Wong et al. |
| Ebro | Filipino | Other Asian | Wong et al. |
| Ebue | Filipino | Other Asian | Wong et al. |
| Ebuen | Filipino | Other Asian | Wong et al. |
| Ebueng | Filipino | Other Asian | Wong et al. |
| Ebuenga | Filipino | Other Asian | Wong et al. |
| Ecal | Filipino | Other Asian | Wong et al. |
| Ecaldre | Filipino | Other Asian | Wong et al. |
| Ecarma | Filipino | Other Asian | Wong et al. |
| Echague | Filipino | Other Asian | Wong et al. |
| Echalar | Filipino | Other Asian | Wong et al. |
| Echanes | Filipino | Other Asian | Wong et al. |
| Echanis | Filipino | Other Asian | Wong et al. |
| Echano | Filipino | Other Asian | Wong et al. |
| Echaure | Filipino | Other Asian | Wong et al. |
| Echave | Filipino | Other Asian | Wong et al. |
| Echavez | Filipino | Other Asian | Wong et al. |
| Echevarria | Filipino | Other Asian | Wong et al. |
| Echeverria | Chinese | Chinese | Wong et al. |
| Echigo | Japanese | Other Asian | Wong et al. |
| Echinique | Filipino | Other Asian | Wong et al. |
| Echipare | Filipino | Other Asian | Wong et al. |
| Echivarre | Filipino | Other Asian | Wong et al. |
| Echiverri | Filipino | Other Asian | Wong et al. |
| Echizen | Japanese | Other Asian | Wong et al. |
| Echon | Filipino | Other Asian | Wong et al. |
| Ecija | Filipino | Other Asian | Wong et al. |
| Ecijan | Filipino | Other Asian | Wong et al. |
| Eclar | Filipino | Other Asian | Wong et al. |
| Eclarin | Filipino | Other Asian | Wong et al. |
| Eclarinal | Filipino | Other Asian | Wong et al. |
| Eclarino | Filipino | Other Asian | Wong et al. |
| Eclavea | Filipino | Other Asian | Wong et al. |
| Ecle | Filipino | Other Asian | Wong et al. |
| Ecleo | Filipino | Other Asian | Wong et al. |
| Eclevia | Filipino | Other Asian | Wong et al. |
| Eclipse | Filipino | Other Asian | Wong et al. |
| Eco | Filipino | Other Asian | Wong et al. |
| Eda | Japanese | Other Asian | Wong et al. |
| Edades | Filipino | Other Asian | Wong et al. |
| Edagawa | Japanese | Other Asian | Wong et al. |
| Edamatsu | Japanese | Other Asian | Wong et al. |
| Edano | Filipino | Other Asian | Wong et al. |
| Edar | Filipino | Other Asian | Wong et al. |
| Edejer | Filipino | Other Asian | Wong et al. |
| Edillo | Filipino | Other Asian | Wong et al. |
| Edillon | Filipino | Other Asian | Wong et al. |
| Edillor | Filipino | Other Asian | Wong et al. |
| Edjan | Filipino | Other Asian | Wong et al. |
| Ednacot | Filipino | Other Asian | Wong et al. |
| Ednalino | Filipino | Other Asian | Wong et al. |
| Ednave | Filipino | Other Asian | Wong et al. |
| Ednilao | Filipino | Other Asian | Wong et al. |
| Edora | Filipino | Other Asian | Wong et al. |
| Edoria | Filipino | Other Asian | Wong et al. |
| Edpao | Filipino | Other Asian | Wong et al. |
| Edquiban | Filipino | Other Asian | Wong et al. |
| Edquid | Filipino | Other Asian | Wong et al. |
| Edquilang | Filipino | Other Asian | Wong et al. |
| Edra | Filipino | Other Asian | Wong et al. |
| Edrada | Filipino | Other Asian | Wong et al. |
| Edralin | Filipino | Other Asian | Wong et al. |
| Edrosa | Filipino | Other Asian | Wong et al. |
| Edrosolan | Filipino | Other Asian | Wong et al. |
| Edrosolo | Filipino | Other Asian | Wong et al. |
| Edrozo | Filipino | Other Asian | Wong et al. |
| Edu | Filipino | Other Asian | Wong et al. |
| Eduardo | Filipino | Other Asian | Wong et al. |
| Eduarte | Filipino | Other Asian | Wong et al. |
| Eduave | Filipino | Other Asian | Wong et al. |
| Eduria | Filipino | Other Asian | Wong et al. |
| Edusada | Filipino | Other Asian | Wong et al. |
| Edwin | Asian Indian | South Asian | Wong et al. |
| Ee | Chinese | Chinese | Wong et al. |
| Effendi | Asian Indian | South Asian | Wong et al. |
| Efondo | Filipino | Other Asian | Wong et al. |
| Egami | Japanese | Other Asian | Wong et al. |
| Egamino | Filipino | Other Asian | Wong et al. |
| Egana | Filipino | Other Asian | Wong et al. |
| Egashira | Japanese | Other Asian | Wong et al. |
| Egawa | Japanese | Other Asian | Wong et al. |
| Egbalic | Filipino | Other Asian | Wong et al. |
| Egenias | Filipino | Other Asian | Wong et al. |
| Egipto | Filipino | Other Asian | Wong et al. |
| Egnisaban | Filipino | Other Asian | Wong et al. |
| Egos | Filipino | Other Asian | Wong et al. |
| Eguchi | Japanese | Other Asian | Wong et al. |
| Eguia | Filipino | Other Asian | Wong et al. |
| Egusa | Japanese | Other Asian | Wong et al. |
| Ehara | Japanese | Other Asian | Wong et al. |
| Ehtasham |  | South Asian | Lim et al. |
| Eisma | Filipino | Other Asian | Wong et al. |
| Ejanda | Filipino | Other Asian | Wong et al. |
| Ejercito | Filipino | Other Asian | Wong et al. |
| Ejima | Japanese | Other Asian | Wong et al. |
| Ejiri | Japanese | Other Asian | Wong et al. |
| Ekambaram | Asian Indian | South Asian | Wong et al. |
| Elacion | Filipino | Other Asian | Wong et al. |
| Elad | Filipino | Other Asian | Wong et al. |
| Elago | Filipino | Other Asian | Wong et al. |
| Elahi | Pakistani; Bangladeshi | South Asian | Kranick et al. |
| Elamparo | Filipino | Other Asian | Wong et al. |
| Elane | Filipino | Other Asian | Wong et al. |
| Elarco | Filipino | Other Asian | Wong et al. |
| Elarmo | Filipino | Other Asian | Wong et al. |
| Elasigue | Filipino | Other Asian | Wong et al. |
| Elauria | Filipino | Other Asian | Wong et al. |
| Elavia | Asian Indian | South Asian | Wong et al. |
| Elayda | Filipino | Other Asian | Wong et al. |
| Elaydo | Filipino | Other Asian | Wong et al. |
| Elazegui | Filipino | Other Asian | Wong et al. |
| Elbo | Filipino | Other Asian | Wong et al. |
| Elcano | Filipino | Other Asian | Wong et al. |
| Ele | Filipino | Other Asian | Wong et al. |
| Eleazar | Filipino | Other Asian | Wong et al. |
| Eleccion | Filipino | Other Asian | Wong et al. |
| Elefante | Filipino | Other Asian | Wong et al. |
| Elegado | Filipino | Other Asian | Wong et al. |
| Elegino | Filipino | Other Asian | Wong et al. |
| Elemento | Filipino | Other Asian | Wong et al. |
| Eleno | Filipino | Other Asian | Wong et al. |
| Elento | Filipino | Other Asian | Wong et al. |
| Elenzano | Filipino | Other Asian | Wong et al. |
| Elepano | Filipino | Other Asian | Wong et al. |
| Elevado | Filipino | Other Asian | Wong et al. |
| Elevazo | Filipino | Other Asian | Wong et al. |
| Elgar | Filipino | Other Asian | Wong et al. |
| Elgarico | Filipino | Other Asian | Wong et al. |
| Elgincolin | Filipino | Other Asian | Wong et al. |
| Elguira | Filipino | Other Asian | Wong et al. |
| Eliazo | Filipino | Other Asian | Wong et al. |
| Elican | Filipino | Other Asian | Wong et al. |
| Elicano | Filipino | Other Asian | Wong et al. |
| Elido | Filipino | Other Asian | Wong et al. |
| Eligio | Filipino | Other Asian | Wong et al. |
| Elisan | Filipino | Other Asian | Wong et al. |
| Elizaga | Filipino | Other Asian | Wong et al. |
| Elizalde | Filipino | Other Asian | Wong et al. |
| Elizarde | Filipino | Other Asian | Wong et al. |
| Ella | Filipino | Other Asian | Wong et al. |
| Ellazar | Filipino | Other Asian | Wong et al. |
| Elle | Filipino | Other Asian | Wong et al. |
| Ellema | Filipino | Other Asian | Wong et al. |
| Ellescas | Filipino | Other Asian | Wong et al. |
| Ellias |  | South Asian | Lim et al. |
| Ello | Filipino | Other Asian | Wong et al. |
| Ellorin | Filipino | Other Asian | Wong et al. |
| Elma | Filipino | Other Asian | Wong et al. |
| Elomina | Filipino | Other Asian | Wong et al. |
| Elopre | Filipino | Other Asian | Wong et al. |
| Elorde | Filipino | Other Asian | Wong et al. |
| Eloriaga | Filipino | Other Asian | Wong et al. |
| Elpedes | Filipino | Other Asian | Wong et al. |
| Elumba | Filipino | Other Asian | Wong et al. |
| Elumir | Filipino | Other Asian | Wong et al. |
| Elvena | Filipino | Other Asian | Wong et al. |
| Elvina | Filipino | Other Asian | Wong et al. |
| Ema | Japanese | Other Asian | Wong et al. |
| Emano | Filipino | Other Asian | Wong et al. |
| Emata | Filipino | Other Asian | Wong et al. |
| Embalsado | Filipino | Other Asian | Wong et al. |
| Embernate | Filipino | Other Asian | Wong et al. |
| Embrador | Filipino | Other Asian | Wong et al. |
| Embudo | Filipino | Other Asian | Wong et al. |
| Embuscado | Filipino | Other Asian | Wong et al. |
| Emi | Japanese | Other Asian | Wong et al. |
| Emiliano | Filipino | Other Asian | Wong et al. |
| Emnace | Filipino | Other Asian | Wong et al. |
| Emocling | Filipino | Other Asian | Wong et al. |
| Emori | Japanese | Other Asian | Wong et al. |
| Emoto | Japanese | Other Asian | Wong et al. |
| Empaynado | Filipino | Other Asian | Wong et al. |
| Empeno | Filipino | Other Asian | Wong et al. |
| Emperado | Filipino | Other Asian | Wong et al. |
| Emperador | Filipino | Other Asian | Wong et al. |
| Empeynado | Filipino | Other Asian | Wong et al. |
| Empleo | Filipino | Other Asian | Wong et al. |
| Emralino | Filipino | Other Asian | Wong et al. |
| Emran |  | South Asian | Lim et al. |
| Emura | Japanese | Other Asian | Wong et al. |
| En | Chinese | Chinese | Wong et al. |
| Enad | Filipino | Other Asian | Wong et al. |
| Enage | Filipino | Other Asian | Wong et al. |
| Enaje | Filipino | Other Asian | Wong et al. |
| Enami | Japanese | Other Asian | Wong et al. |
| Enand | Asian Indian | South Asian | Wong et al. |
| Enano | Filipino | Other Asian | Wong et al. |
| Enate | Filipino | Other Asian | Wong et al. |
| Encabo | Filipino | Other Asian | Wong et al. |
| Encanto | Filipino | Other Asian | Wong et al. |
| Encarnacao | Chinese | Chinese | Wong et al. |
| Encarnacion | Filipino | Other Asian | Wong et al. |
| Encarnado | Filipino | Other Asian | Wong et al. |
| Encina | Filipino | Other Asian | Wong et al. |
| Encinares | Filipino | Other Asian | Wong et al. |
| Encinas | Filipino | Other Asian | Wong et al. |
| Encio | Filipino | Other Asian | Wong et al. |
| Enciso | Filipino | Other Asian | Wong et al. |
| Encomienda | Filipino | Other Asian | Wong et al. |
| Endaya | Filipino | Other Asian | Wong et al. |
| Endo | Japanese | Other Asian | Wong et al. |
| Endoh | Japanese | Other Asian | Wong et al. |
| Endoso | Filipino | Other Asian | Wong et al. |
| Endow | Japanese | Other Asian | Wong et al. |
| Endozo | Filipino | Other Asian | Wong et al. |
| Endriga | Filipino | Other Asian | Wong et al. |
| Endrina | Filipino | Other Asian | Wong et al. |
| Endrinal | Filipino | Other Asian | Wong et al. |
| Enerio | Filipino | Other Asian | Wong et al. |
| Enerlan | Filipino | Other Asian | Wong et al. |
| Enero | Filipino | Other Asian | Wong et al. |
| Enerva | Filipino | Other Asian | Wong et al. |
| Eng | Chinese; Cambodian; Malaysian | Chinese | Kranick et al. |
| Engalla | Filipino | Other Asian | Wong et al. |
| Engineer | Asian Indian | South Asian | Wong et al. |
| Engracia | Filipino | Other Asian | Wong et al. |
| Enjoji | Japanese | Other Asian | Wong et al. |
| Enojo | Filipino | Other Asian | Wong et al. |
| Enoki | Japanese | Other Asian | Wong et al. |
| Enokida | Japanese | Other Asian | Wong et al. |
| Enokido | Japanese | Other Asian | Wong et al. |
| Enomoto | Japanese | Other Asian | Wong et al. |
| Enrico | Filipino | Other Asian | Wong et al. |
| Enriguez | Filipino | Other Asian | Wong et al. |
| Enrile | Filipino | Other Asian | Wong et al. |
| Enrique | Filipino | Other Asian | Wong et al. |
| Enriques | Filipino | Other Asian | Wong et al. |
| Enriquez | Filipino | Other Asian | Wong et al. |
| Enteria | Filipino | Other Asian | Wong et al. |
| Enverga | Filipino | Other Asian | Wong et al. |
| Eo | Korean | Other Asian | Wong et al. |
| Eom | Korean | Other Asian | Kranick et al. |
| Eong | Chinese | Chinese | Wong et al. |
| Eoyang | Chinese | Chinese | Wong et al. |
| Epino | Filipino | Other Asian | Wong et al. |
| Epistola | Filipino | Other Asian | Wong et al. |
| Equila | Filipino | Other Asian | Wong et al. |
| Erana | Filipino | Other Asian | Wong et al. |
| Eraso | Filipino | Other Asian | Wong et al. |
| Erasquin | Filipino | Other Asian | Wong et al. |
| Erazo | Chinese | Chinese | Wong et al. |
| Erece | Filipino | Other Asian | Wong et al. |
| Erese | Filipino | Other Asian | Wong et al. |
| Ereso | Filipino | Other Asian | Wong et al. |
| Erestain | Filipino | Other Asian | Wong et al. |
| Erfe | Filipino | Other Asian | Wong et al. |
| Ergina | Filipino | Other Asian | Wong et al. |
| Ergino | Filipino | Other Asian | Wong et al. |
| Erguiza | Filipino | Other Asian | Wong et al. |
| Erh | Chinese | Chinese | Wong et al. |
| Erice | Filipino | Other Asian | Wong et al. |
| Erive | Filipino | Other Asian | Wong et al. |
| Ermino | Filipino | Other Asian | Wong et al. |
| Ermita | Filipino | Other Asian | Wong et al. |
| Ermitanio | Filipino | Other Asian | Wong et al. |
| Ermitano | Filipino | Other Asian | Wong et al. |
| Ermosilla | Filipino | Other Asian | Wong et al. |
| Eroles | Filipino | Other Asian | Wong et al. |
| Erulkar | Asian Indian | South Asian | Wong et al. |
| Erum | Filipino | Other Asian | Wong et al. |
| Esaki | Japanese | Other Asian | Wong et al. |
| Escala | Filipino | Other Asian | Wong et al. |
| Escalada | Filipino | Other Asian | Wong et al. |
| Escalante | Filipino | Other Asian | Wong et al. |
| Escalera | Filipino | Other Asian | Wong et al. |
| Escalona | Filipino | Other Asian | Wong et al. |
| Escamilla | Filipino | Other Asian | Wong et al. |
| Escandor | Filipino | Other Asian | Wong et al. |
| Escanilla | Filipino | Other Asian | Wong et al. |
| Escanlar | Filipino | Other Asian | Wong et al. |
| Escano | Filipino | Other Asian | Wong et al. |
| Escarcha | Filipino | Other Asian | Wong et al. |
| Escarez | Filipino | Other Asian | Wong et al. |
| Escarilla | Filipino | Other Asian | Wong et al. |
| Escario | Filipino | Other Asian | Wong et al. |
| Escarrilla | Filipino | Other Asian | Wong et al. |
| Escartin | Filipino | Other Asian | Wong et al. |
| Escasa | Filipino | Other Asian | Wong et al. |
| Escay | Filipino | Other Asian | Wong et al. |
| Esclamado | Filipino | Other Asian | Wong et al. |
| Escobal | Filipino | Other Asian | Wong et al. |
| Escobar | Filipino | Other Asian | Wong et al. |
| Escobedo | Filipino | Other Asian | Wong et al. |
| Escober | Filipino | Other Asian | Wong et al. |
| Escobia | Filipino | Other Asian | Wong et al. |
| Escobido | Filipino | Other Asian | Wong et al. |
| Escolano | Filipino | Other Asian | Wong et al. |
| Escolar | Filipino | Other Asian | Wong et al. |
| Esconde | Filipino | Other Asian | Wong et al. |
| Escondo | Filipino | Other Asian | Wong et al. |
| Escorpiso | Filipino | Other Asian | Wong et al. |
| Escorpizo | Filipino | Other Asian | Wong et al. |
| Escosio | Filipino | Other Asian | Wong et al. |
| Escota | Filipino | Other Asian | Wong et al. |
| Escoto | Filipino | Other Asian | Wong et al. |
| Escribano | Filipino | Other Asian | Wong et al. |
| Escritor | Filipino | Other Asian | Wong et al. |
| Escuadra | Filipino | Other Asian | Wong et al. |
| Escuadro | Filipino | Other Asian | Wong et al. |
| Escudero | Filipino | Other Asian | Wong et al. |
| Escueta | Filipino | Other Asian | Wong et al. |
| Escullar | Filipino | Other Asian | Wong et al. |
| Escurel | Filipino | Other Asian | Wong et al. |
| Escuro | Filipino | Other Asian | Wong et al. |
| Escusa | Filipino | Other Asian | Wong et al. |
| Escutin | Filipino | Other Asian | Wong et al. |
| Esguerra | Filipino | Other Asian | Wong et al. |
| Eshak |  | South Asian | Lim et al. |
| Eslabon | Filipino | Other Asian | Wong et al. |
| Eslao | Filipino | Other Asian | Wong et al. |
| Eslava | Filipino | Other Asian | Wong et al. |
| Eslit | Filipino | Other Asian | Wong et al. |
| Esmade | Filipino | Other Asian | Wong et al. |
| Esmail | Asian Indian | South Asian | Wong et al. |
| Esmane | Filipino | Other Asian | Wong et al. |
| Esmele | Filipino | Other Asian | Wong et al. |
| Esmeralda | Filipino | Other Asian | Wong et al. |
| Esmero | Filipino | Other Asian | Wong et al. |
| Esmilla | Filipino | Other Asian | Wong et al. |
| Esmino | Filipino | Other Asian | Wong et al. |
| Espada | Filipino | Other Asian | Wong et al. |
| Espaldon | Filipino | Other Asian | Wong et al. |
| Espana | Filipino | Other Asian | Wong et al. |
| Espano | Filipino | Other Asian | Wong et al. |
| Espanol | Filipino | Other Asian | Wong et al. |
| Espanola | Filipino | Other Asian | Wong et al. |
| Espanto | Filipino | Other Asian | Wong et al. |
| Esparas | Filipino | Other Asian | Wong et al. |
| Espares | Filipino | Other Asian | Wong et al. |
| Esparrago | Filipino | Other Asian | Wong et al. |
| Espartero | Filipino | Other Asian | Wong et al. |
| Espayos | Filipino | Other Asian | Wong et al. |
| Espedido | Filipino | Other Asian | Wong et al. |
| Espejo | Filipino | Other Asian | Wong et al. |
| Espeleta | Filipino | Other Asian | Wong et al. |
| Espena | Filipino | Other Asian | Wong et al. |
| Espenilla | Filipino | Other Asian | Wong et al. |
| Espera | Filipino | Other Asian | Wong et al. |
| Esperancilla | Filipino | Other Asian | Wong et al. |
| Esperante | Filipino | Other Asian | Wong et al. |
| Esperanza | Filipino | Other Asian | Wong et al. |
| Esperanzate | Filipino | Other Asian | Wong et al. |
| Esperas | Filipino | Other Asian | Wong et al. |
| Espero | Filipino | Other Asian | Wong et al. |
| Esperon | Filipino | Other Asian | Wong et al. |
| Espidol | Filipino | Other Asian | Wong et al. |
| Espina | Filipino | Other Asian | Wong et al. |
| Espinas | Filipino | Other Asian | Wong et al. |
| Espineli | Filipino | Other Asian | Wong et al. |
| Espino | Filipino | Other Asian | Wong et al. |
| Espinosa | Filipino | Other Asian | Wong et al. |
| Espinoza | Filipino | Other Asian | Wong et al. |
| Espinueva | Filipino | Other Asian | Wong et al. |
| Espique | Filipino | Other Asian | Wong et al. |
| Espirito | Filipino | Other Asian | Wong et al. |
| Espiritu | Filipino | Other Asian | Wong et al. |
| Espirity | Filipino | Other Asian | Wong et al. |
| Esplana | Filipino | Other Asian | Wong et al. |
| Esplanada | Filipino | Other Asian | Wong et al. |
| Espolong | Filipino | Other Asian | Wong et al. |
| Esporlas | Filipino | Other Asian | Wong et al. |
| Espos | Filipino | Other Asian | Wong et al. |
| Esposo | Filipino | Other Asian | Wong et al. |
| Esquejo | Filipino | Other Asian | Wong et al. |
| Esquerra | Filipino | Other Asian | Wong et al. |
| Esquibel | Filipino | Other Asian | Wong et al. |
| Esquieres | Filipino | Other Asian | Wong et al. |
| Esquillo | Filipino | Other Asian | Wong et al. |
| Esquivel | Filipino | Other Asian | Wong et al. |
| Esta | Filipino | Other Asian | Wong et al. |
| Estabaya | Filipino | Other Asian | Wong et al. |
| Estabillo | Filipino | Other Asian | Wong et al. |
| Estacio | Filipino | Other Asian | Wong et al. |
| Estacion | Filipino | Other Asian | Wong et al. |
| Estal | Filipino | Other Asian | Wong et al. |
| Estalilla | Filipino | Other Asian | Wong et al. |
| Estandarte | Filipino | Other Asian | Wong et al. |
| Estanislao | Filipino | Other Asian | Wong et al. |
| Estante | Filipino | Other Asian | Wong et al. |
| Estantino | Filipino | Other Asian | Wong et al. |
| Estares | Filipino | Other Asian | Wong et al. |
| Estarija | Filipino | Other Asian | Wong et al. |
| Estaris | Filipino | Other Asian | Wong et al. |
| Estavillo | Filipino | Other Asian | Wong et al. |
| Estayo | Filipino | Other Asian | Wong et al. |
| Esteban | Filipino | Other Asian | Wong et al. |
| Estela | Filipino | Other Asian | Wong et al. |
| Estella | Filipino | Other Asian | Wong et al. |
| Estepa | Filipino | Other Asian | Wong et al. |
| Estera | Filipino | Other Asian | Wong et al. |
| Esternon | Filipino | Other Asian | Wong et al. |
| Esteron | Filipino | Other Asian | Wong et al. |
| Esteva | Filipino | Other Asian | Wong et al. |
| Esteves | Filipino | Other Asian | Wong et al. |
| Estevez | Filipino | Other Asian | Wong et al. |
| Estiban | Filipino | Other Asian | Wong et al. |
| Estigoy | Filipino | Other Asian | Wong et al. |
| Estil | Filipino | Other Asian | Wong et al. |
| Estillore | Filipino | Other Asian | Wong et al. |
| Estilo | Filipino | Other Asian | Wong et al. |
| Estimo | Filipino | Other Asian | Wong et al. |
| Estioco | Filipino | Other Asian | Wong et al. |
| Estioko | Filipino | Other Asian | Wong et al. |
| Estipona | Filipino | Other Asian | Wong et al. |
| Estipular | Filipino | Other Asian | Wong et al. |
| Estira | Filipino | Other Asian | Wong et al. |
| Estiva | Filipino | Other Asian | Wong et al. |
| Estocapio | Filipino | Other Asian | Wong et al. |
| Estoesta | Filipino | Other Asian | Wong et al. |
| Estolano | Filipino | Other Asian | Wong et al. |
| Estolas | Filipino | Other Asian | Wong et al. |
| Estonactoc | Filipino | Other Asian | Wong et al. |
| Estonilo | Filipino | Other Asian | Wong et al. |
| Estopare | Filipino | Other Asian | Wong et al. |
| Estoque | Filipino | Other Asian | Wong et al. |
| Estores | Filipino | Other Asian | Wong et al. |
| Estorque | Filipino | Other Asian | Wong et al. |
| Estoy | Filipino | Other Asian | Wong et al. |
| Estrada | Filipino | Other Asian | Wong et al. |
| Estrala | Filipino | Other Asian | Wong et al. |
| Estranero | Filipino | Other Asian | Wong et al. |
| Estrella | Filipino | Other Asian | Wong et al. |
| Estrellado | Filipino | Other Asian | Wong et al. |
| Estrellas | Filipino | Other Asian | Wong et al. |
| Estrellon | Filipino | Other Asian | Wong et al. |
| Estrera | Filipino | Other Asian | Wong et al. |
| Estuar | Filipino | Other Asian | Wong et al. |
| Estuart | Filipino | Other Asian | Wong et al. |
| Estudillo | Filipino | Other Asian | Wong et al. |
| Estuita | Filipino | Other Asian | Wong et al. |
| Eto | Japanese | Other Asian | Wong et al. |
| Etrata | Filipino | Other Asian | Wong et al. |
| Etwarrow |  | South Asian | Lim et al. |
| Eugenio | Filipino | Other Asian | Wong et al. |
| Eulalia | Filipino | Other Asian | Wong et al. |
| Eulalio | Filipino | Other Asian | Wong et al. |
| Eum | Korean | Other Asian | Kranick et al. |
| Eun | Korean | Other Asian | Wong et al. |
| Europa | Filipino | Other Asian | Wong et al. |
| Eusantos | Filipino | Other Asian | Wong et al. |
| Eusebio | Filipino | Other Asian | Wong et al. |
| Eustaquio | Filipino | Other Asian | Wong et al. |
| Evaldez | Filipino | Other Asian | Wong et al. |
| Evalle | Filipino | Other Asian | Wong et al. |
| Evangelio | Filipino | Other Asian | Wong et al. |
| Evangelista | Filpino | Other Asian | Kranick et al. |
| Evans |  | South Asian | Lim et al. |
| Evaristo | Filipino | Other Asian | Wong et al. |
| Evasco | Filipino | Other Asian | Wong et al. |
| Evia | Filipino | Other Asian | Wong et al. |
| Evidente | Filipino | Other Asian | Wong et al. |
| Eviota | Filipino | Other Asian | Wong et al. |
| Evora | Filipino | Other Asian | Wong et al. |
| Exala | Filipino | Other Asian | Wong et al. |
| Exconde | Filipino | Other Asian | Wong et al. |
| Exito | Filipino | Other Asian | Wong et al. |
| Ezaki | Japanese | Other Asian | Wong et al. |
| Ezawa | Japanese | Other Asian | Wong et al. |
| Ezoe | Japanese | Other Asian | Wong et al. |
| Ezpeleta | Filipino | Other Asian | Wong et al. |
| Fa | Chinese | Chinese | Wong et al. |
| Fababier | Filipino | Other Asian | Wong et al. |
| Fabay | Filipino | Other Asian | Wong et al. |
| Fabela | Filipino | Other Asian | Wong et al. |
| Fabella | Filipino | Other Asian | Wong et al. |
| Fabello | Filipino | Other Asian | Wong et al. |
| Fabi | Filipino | Other Asian | Wong et al. |
| Fabia | Filipino | Other Asian | Wong et al. |
| Fabico | Filipino | Other Asian | Wong et al. |
| Fabie | Filipino | Other Asian | Wong et al. |
| Fabila | Filipino | Other Asian | Wong et al. |
| Fabillar | Filipino | Other Asian | Wong et al. |
| Fabillaran | Filipino | Other Asian | Wong et al. |
| Fabio | Filipino | Other Asian | Wong et al. |
| Fabionar | Filipino | Other Asian | Wong et al. |
| Fabon | Filipino | Other Asian | Wong et al. |
| Fabra | Filipino | Other Asian | Wong et al. |
| Fabregas | Filipino | Other Asian | Wong et al. |
| Fabricante | Filipino | Other Asian | Wong et al. |
| Fabrigas | Filipino | Other Asian | Wong et al. |
| Fabro | Filipino | Other Asian | Wong et al. |
| Fabros | Filipino | Other Asian | Wong et al. |
| Fabula | Filipino | Other Asian | Wong et al. |
| Fabunan | Filipino | Other Asian | Wong et al. |
| Facelo | Filipino | Other Asian | Wong et al. |
| Factor | Filipino | Other Asian | Wong et al. |
| Factora | Filipino | Other Asian | Wong et al. |
| Factura | Filipino | Other Asian | Wong et al. |
| Facun | Filipino | Other Asian | Wong et al. |
| Facundo | Filipino | Other Asian | Wong et al. |
| Facunla | Filipino | Other Asian | Wong et al. |
| Fadera | Filipino | Other Asian | Wong et al. |
| Faderan | Filipino | Other Asian | Wong et al. |
| Faderon | Filipino | Other Asian | Wong et al. |
| Fadia | Asian Indian | South Asian | Wong et al. |
| Fadri | Filipino | Other Asian | Wong et al. |
| Fadrigo | Filipino | Other Asian | Wong et al. |
| Fadul | Filipino | Other Asian | Wong et al. |
| Faelnar | Filipino | Other Asian | Wong et al. |
| Fagaragan | Filipino | Other Asian | Wong et al. |
| Fagarang | Filipino | Other Asian | Wong et al. |
| Fagel | Filipino | Other Asian | Wong et al. |
| Fah | Chinese | Chinese | Wong et al. |
| Fahmin |  | South Asian | Lim et al. |
| Fai | Chinese | Chinese | Wong et al. |
| Faigal | Filipino | Other Asian | Wong et al. |
| Failano | Filipino | Other Asian | Wong et al. |
| Failma | Filipino | Other Asian | Wong et al. |
| Faisal |  | South Asian | Lim et al. |
| Faiz |  | South Asian | Lim et al. |
| Fajardo | Filipino | Other Asian | Wong et al. |
| Fajarillo | Filipino | Other Asian | Wong et al. |
| Fajatin | Filipino | Other Asian | Wong et al. |
| Fajayan | Filipino | Other Asian | Wong et al. |
| Fajota | Filipino | Other Asian | Wong et al. |
| Fajotina | Filipino | Other Asian | Wong et al. |
| Fakih | Asian Indian | South Asian | Wong et al. |
| Fakir |  | South Asian | Lim et al. |
| Fakirbhai | Asian Indian | South Asian | Wong et al. |
| Falaminiano | Filipino | Other Asian | Wong et al. |
| Falcis | Filipino | Other Asian | Wong et al. |
| Falcon | Filipino | Other Asian | Wong et al. |
| Falek | Filipino | Other Asian | Wong et al. |
| Falgui | Filipino | Other Asian | Wong et al. |
| Fallaria | Filipino | Other Asian | Wong et al. |
| Fallarme | Filipino | Other Asian | Wong et al. |
| Fallejo | Filipino | Other Asian | Wong et al. |
| Falloran | Filipino | Other Asian | Wong et al. |
| Fallorina | Filipino | Other Asian | Wong et al. |
| Falsario | Filipino | Other Asian | Wong et al. |
| Falsis | Filipino | Other Asian | Wong et al. |
| Faltado | Filipino | Other Asian | Wong et al. |
| Fam | Chinese | Chinese | Wong et al. |
| Famador | Filipino | Other Asian | Wong et al. |
| Famanilay | Filipino | Other Asian | Wong et al. |
| Familar | Filipino | Other Asian | Wong et al. |
| Familara | Filipino | Other Asian | Wong et al. |
| Famisan | Filipino | Other Asian | Wong et al. |
| Famorca | Filipino | Other Asian | Wong et al. |
| Famularcano | Filipino | Other Asian | Wong et al. |
| Fan | Chinese | Chinese | Kranick et al. |
| Fanchiang | Chinese | Chinese | Wong et al. |
| Fandialan | Filipino | Other Asian | Wong et al. |
| Fandino | Filipino | Other Asian | Wong et al. |
| Faner | Filipino | Other Asian | Wong et al. |
| Fang | Chinese | Chinese | Kranick et al. |
| Fangon | Filipino | Other Asian | Wong et al. |
| Fangonil | Filipino | Other Asian | Wong et al. |
| Fangonilo | Filipino | Other Asian | Wong et al. |
| Fanibanda | Asian Indian | South Asian | Wong et al. |
| Fanlo | Filipino | Other Asian | Wong et al. |
| Fantone | Filipino | Other Asian | Wong et al. |
| Fanugao | Filipino | Other Asian | Wong et al. |
| Far | Chinese | Chinese | Wong et al. |
| Farajee |  | South Asian | Lim et al. |
| Farala | Filipino | Other Asian | Wong et al. |
| Faralan | Filipino | Other Asian | Wong et al. |
| Faraon | Filipino | Other Asian | Wong et al. |
| Faraque |  | South Asian | Lim et al. |
| Fardoosi |  | South Asian | Lim et al. |
| Fareed | Asian Indian | South Asian | Wong et al. |
| Farid | Asian Indian | South Asian | Wong et al. |
| Faridi | Asian Indian | South Asian | Wong et al. |
| Farin | Filipino | Other Asian | Wong et al. |
| Farinas | Filipino | Other Asian | Wong et al. |
| Farishta | Asian Indian | South Asian | Wong et al. |
| Farjana |  | South Asian | Lim et al. |
| Farnacio | Filipino | Other Asian | Wong et al. |
| Farnal | Filipino | Other Asian | Wong et al. |
| Farne | Filipino | Other Asian | Wong et al. |
| Farol | Filipino | Other Asian | Wong et al. |
| Farolan | Filipino | Other Asian | Wong et al. |
| Farooki | Asian Indian | South Asian | Wong et al. |
| Farooq | Asian Indian | South Asian | Wong et al. |
| Farooqi | Asian Indian | South Asian | Wong et al. |
| Farooque |  | South Asian | Lim et al. |
| Farooqui | Asian Indian | South Asian | Wong et al. |
| Faroque |  | South Asian | Lim et al. |
| Faroquee |  | South Asian | Lim et al. |
| Farrales | Filipino | Other Asian | Wong et al. |
| Faruk |  | South Asian | Lim et al. |
| Farukhi | Asian Indian | South Asian | Wong et al. |
| Faruki |  | South Asian | Lim et al. |
| Faruqi | Asian Indian | South Asian | Wong et al. |
| Faruque |  | South Asian | Lim et al. |
| Faruqui | Asian Indian | South Asian | Wong et al. |
| Farzana |  | South Asian | Lim et al. |
| Fasihuddin | Asian Indian | South Asian | Wong et al. |
| Fat | Chinese | Chinese | Wong et al. |
| Fatama |  | South Asian | Lim et al. |
| Fatema |  | South Asian | Lim et al. |
| Fathadayetulla |  | South Asian | Lim et al. |
| Fatima | Pakistani; Asian Indian | South Asian | Kranick et al. |
| Fatt | Chinese | Chinese | Wong et al. |
| Faun | Chinese | Chinese | Wong et al. |
| Faung | Chinese | Chinese | Wong et al. |
| Fauni | Filipino | Other Asian | Wong et al. |
| Faustino | Filipino | Other Asian | Wong et al. |
| Fausto | Filipino | Other Asian | Wong et al. |
| Favila | Filipino | Other Asian | Wong et al. |
| Favis | Filipino | Other Asian | Wong et al. |
| Fawad |  | South Asian | Lim et al. |
| Fayloga | Filipino | Other Asian | Wong et al. |
| Faylona | Filipino | Other Asian | Wong et al. |
| Faypon | Filipino | Other Asian | Wong et al. |
| Fayyaz |  | South Asian | Lim et al. |
| Fazal | Asian Indian | South Asian | Wong et al. |
| Fazalbhoy | Asian Indian | South Asian | Wong et al. |
| Fazil | Asian Indian | South Asian | Wong et al. |
| Fazli | Asian Indian | South Asian | Wong et al. |
| Fe | Chinese | Chinese | Wong et al. |
| Febre | Filipino | Other Asian | Wong et al. |
| Fedalizo | Filipino | Other Asian | Wong et al. |
| Federe | Filipino | Other Asian | Wong et al. |
| Federis | Filipino | Other Asian | Wong et al. |
| Federizo | Filipino | Other Asian | Wong et al. |
| Fei | Chinese | Chinese | Wong et al. |
| Feir | Filipino | Other Asian | Wong et al. |
| Felarca | Filipino | Other Asian | Wong et al. |
| Feleo | Filipino | Other Asian | Wong et al. |
| Felicano | Filipino | Other Asian | Wong et al. |
| Felices | Filipino | Other Asian | Wong et al. |
| Felicia | Filipino | Other Asian | Wong et al. |
| Feliciano | Filipino | Other Asian | Wong et al. |
| Felicidario | Filipino | Other Asian | Wong et al. |
| Felicilda | Filipino | Other Asian | Wong et al. |
| Felicisimo | Filipino | Other Asian | Wong et al. |
| Felicitas | Filipino | Other Asian | Wong et al. |
| Felipe | Filipino | Other Asian | Wong et al. |
| Felisilda | Filipino | Other Asian | Wong et al. |
| Felix | Filipino | Other Asian | Wong et al. |
| Felizardo | Filipino | Other Asian | Wong et al. |
| Felizmena | Filipino | Other Asian | Wong et al. |
| Fen | Chinese | Chinese | Wong et al. |
| Feng | Chinese | Chinese | Kranick et al. |
| Feniquito | Filipino | Other Asian | Wong et al. |
| Fenol | Filipino | Other Asian | Wong et al. |
| Feraer | Filipino | Other Asian | Wong et al. |
| Feraren | Filipino | Other Asian | Wong et al. |
| Feraro | Filipino | Other Asian | Wong et al. |
| Ferdausi |  | South Asian | Lim et al. |
| Ferdous |  | South Asian | Lim et al. |
| Ferdoushi |  | South Asian | Lim et al. |
| Ferdousi |  | South Asian | Lim et al. |
| Ferdus |  | South Asian | Lim et al. |
| Feria | Filipino | Other Asian | Wong et al. |
| Ferido | Filipino | Other Asian | Wong et al. |
| Feril | Filipino | Other Asian | Wong et al. |
| Ferma | Filipino | Other Asian | Wong et al. |
| Fermil | Filipino | Other Asian | Wong et al. |
| Fermin | Filipino | Other Asian | Wong et al. |
| Fermo | Filipino | Other Asian | Wong et al. |
| Fernadez | Filipino | Other Asian | Wong et al. |
| Fernandes | Asian Indian | South Asian | Wong et al. |
| Fernandez | Filipino | Other Asian | Wong et al. |
| Fernandis | Asian Indian | South Asian | Wong et al. |
| Fernando | Filipino | Other Asian | Wong et al. |
| Ferng | Chinese | Chinese | Wong et al. |
| Ferolin | Filipino | Other Asian | Wong et al. |
| Ferolino | Filipino | Other Asian | Wong et al. |
| Feroze |  | South Asian | Lim et al. |
| Ferrao | Asian Indian | South Asian | Wong et al. |
| Ferraren | Filipino | Other Asian | Wong et al. |
| Ferras | Chinese | Chinese | Wong et al. |
| Ferrer | Filipino | Other Asian | Wong et al. |
| Ferreras | Filipino | Other Asian | Wong et al. |
| Ferreria | Filipino | Other Asian | Wong et al. |
| Ferreros | Filipino | Other Asian | Wong et al. |
| Ferriols | Filipino | Other Asian | Wong et al. |
| Festejo | Filipino | Other Asian | Wong et al. |
| Festin | Filipino | Other Asian | Wong et al. |
| Fetalino | Filipino | Other Asian | Wong et al. |
| Fetalvero | Filipino | Other Asian | Wong et al. |
| Fi | Chinese | Chinese | Wong et al. |
| Fidel | Filipino | Other Asian | Wong et al. |
| Fidelino | Filipino | Other Asian | Wong et al. |
| Fider | Filipino | Other Asian | Wong et al. |
| Fiel | Filipino | Other Asian | Wong et al. |
| Fiesta | Filipino | Other Asian | Wong et al. |
| Figueras | Filipino | Other Asian | Wong et al. |
| Figueroa | Filipino | Other Asian | Wong et al. |
| Figuerres | Filipino | Other Asian | Wong et al. |
| Figuracion | Filipino | Other Asian | Wong et al. |
| Filamor | Filipino | Other Asian | Wong et al. |
| Filart | Filipino | Other Asian | Wong et al. |
| Filio | Filipino | Other Asian | Wong et al. |
| Filipinas | Filipino | Other Asian | Wong et al. |
| Fillon | Filipino | Other Asian | Wong et al. |
| Filomeno | Filipino | Other Asian | Wong et al. |
| Filoteo | Filipino | Other Asian | Wong et al. |
| Fin | Chinese | Chinese | Wong et al. |
| Fines | Filipino | Other Asian | Wong et al. |
| Fineza | Filipino | Other Asian | Wong et al. |
| Fing | Chinese | Chinese | Wong et al. |
| Finones | Filipino | Other Asian | Wong et al. |
| Finuliar | Filipino | Other Asian | Wong et al. |
| Firdous | Pakistani | South Asian | Kranick et al. |
| Firme | Filipino | Other Asian | Wong et al. |
| Firmeza | Filipino | Other Asian | Wong et al. |
| Flandez | Filipino | Other Asian | Wong et al. |
| Flauta | Filipino | Other Asian | Wong et al. |
| Flaviano | Filipino | Other Asian | Wong et al. |
| Flavier | Filipino | Other Asian | Wong et al. |
| Floirendo | Filipino | Other Asian | Wong et al. |
| Flojo | Filipino | Other Asian | Wong et al. |
| Flor | Filipino | Other Asian | Wong et al. |
| Floralde | Filipino | Other Asian | Wong et al. |
| Floranda | Filipino | Other Asian | Wong et al. |
| Florano | Filipino | Other Asian | Wong et al. |
| Flordelis | Filipino | Other Asian | Wong et al. |
| Flordeliza | Filipino | Other Asian | Wong et al. |
| Florenciano | Filipino | Other Asian | Wong et al. |
| Florencio | Filipino | Other Asian | Wong et al. |
| Florendo | Filipino | Other Asian | Wong et al. |
| Florentin | Filipino | Other Asian | Wong et al. |
| Florentino | Filipino | Other Asian | Wong et al. |
| Flores | Filipino | Other Asian | Wong et al. |
| Floresca | Filipino | Other Asian | Wong et al. |
| Florez | Filipino | Other Asian | Wong et al. |
| Floreza | Filipino | Other Asian | Wong et al. |
| Florida | Filipino | Other Asian | Wong et al. |
| Florido | Filipino | Other Asian | Wong et al. |
| Florita | Filipino | Other Asian | Wong et al. |
| Flormata | Filipino | Other Asian | Wong et al. |
| Fnu |  | South Asian | Lim et al. |
| Fo | Chinese | Chinese | Wong et al. |
| Fock | Chinese | Chinese | Wong et al. |
| Fogata | Filipino | Other Asian | Wong et al. |
| Fojas | Filipino | Other Asian | Wong et al. |
| Fok | Chinese | Chinese | Wong et al. |
| Fokhor |  | South Asian | Lim et al. |
| Foliente | Filipino | Other Asian | Wong et al. |
| Follosco | Filipino | Other Asian | Wong et al. |
| Folloso | Filipino | Other Asian | Wong et al. |
| Fon | Chinese | Chinese | Wong et al. |
| Fonacier | Filipino | Other Asian | Wong et al. |
| Fond | Chinese | Chinese | Wong et al. |
| Fone | Chinese | Chinese | Wong et al. |
| Fong | Chinese | Chinese | Kranick et al. |
| Font | Chinese | Chinese | Wong et al. |
| Fontamillas | Filipino | Other Asian | Wong et al. |
| Fontanares | Filipino | Other Asian | Wong et al. |
| Fontanilla | Filipino | Other Asian | Wong et al. |
| Fontanos | Filipino | Other Asian | Wong et al. |
| Fontecha | Filipino | Other Asian | Wong et al. |
| Fontejon | Filipino | Other Asian | Wong et al. |
| Fontela | Filipino | Other Asian | Wong et al. |
| Fontelera | Filipino | Other Asian | Wong et al. |
| Fontilea | Filipino | Other Asian | Wong et al. |
| Fontillas | Filipino | Other Asian | Wong et al. |
| Foo | Chinese | Chinese | Wong et al. |
| Fook | Chinese | Chinese | Wong et al. |
| Foon | Chinese | Chinese | Wong et al. |
| Foong | Chinese | Chinese | Wong et al. |
| Forcadela | Filipino | Other Asian | Wong et al. |
| Fordan | Filipino | Other Asian | Wong et al. |
| Formalejo | Filipino | Other Asian | Wong et al. |
| Formanes | Filipino | Other Asian | Wong et al. |
| Formantes | Filipino | Other Asian | Wong et al. |
| Formentera | Filipino | Other Asian | Wong et al. |
| Formoso | Filipino | Other Asian | Wong et al. |
| Fornillos | Filipino | Other Asian | Wong et al. |
| Fornoles | Filipino | Other Asian | Wong et al. |
| Foronda | Filipino | Other Asian | Wong et al. |
| Forral | Filipino | Other Asian | Wong et al. |
| Fortaleza | Filipino | Other Asian | Wong et al. |
| Fortes | Filipino | Other Asian | Wong et al. |
| Forteza | Filipino | Other Asian | Wong et al. |
| Fortich | Filipino | Other Asian | Wong et al. |
| Fortunado | Filipino | Other Asian | Wong et al. |
| Fortuno | Filipino | Other Asian | Wong et al. |
| Fot | Chinese | Chinese | Wong et al. |
| Fou | Chinese | Chinese | Wong et al. |
| Foun | Chinese | Chinese | Wong et al. |
| Foung | Chinese | Chinese | Wong et al. |
| Foyez |  | South Asian | Lim et al. |
| Foyn | Chinese | Chinese | Wong et al. |
| Foz | Filipino | Other Asian | Wong et al. |
| Fozdar | Asian Indian | South Asian | Wong et al. |
| Fragada | Filipino | Other Asian | Wong et al. |
| Fragante | Filipino | Other Asian | Wong et al. |
| Fragata | Filipino | Other Asian | Wong et al. |
| Fran | Filipino | Other Asian | Wong et al. |
| Franada | Filipino | Other Asian | Wong et al. |
| Francia | Filipino | Other Asian | Wong et al. |
| Francisco | Filipino | Other Asian | Kranick et al. |
| Franco | Filipino | Other Asian | Wong et al. |
| Frando | Filipino | Other Asian | Wong et al. |
| Frani | Filipino | Other Asian | Wong et al. |
| Frantilla | Filipino | Other Asian | Wong et al. |
| Franzuela | Filipino | Other Asian | Wong et al. |
| Frayna | Filipino | Other Asian | Wong et al. |
| Fredeluces | Filipino | Other Asian | Wong et al. |
| Fresnido | Filipino | Other Asian | Wong et al. |
| Fresnoza | Filipino | Other Asian | Wong et al. |
| Frial | Filipino | Other Asian | Wong et al. |
| Frialde | Filipino | Other Asian | Wong et al. |
| Frianeza | Filipino | Other Asian | Wong et al. |
| Frias | Filipino | Other Asian | Wong et al. |
| Frigillana | Filipino | Other Asian | Wong et al. |
| Friginal | Filipino | Other Asian | Wong et al. |
| Frijas | Filipino | Other Asian | Wong et al. |
| Frilles | Filipino | Other Asian | Wong et al. |
| Frivaldo | Filipino | Other Asian | Wong et al. |
| Froilan | Filipino | Other Asian | Wong et al. |
| Fronda | Filipino | Other Asian | Wong et al. |
| Frondarina | Filipino | Other Asian | Wong et al. |
| Frondozo | Filipino | Other Asian | Wong et al. |
| Fronteras | Filipino | Other Asian | Wong et al. |
| Fructuoso | Filipino | Other Asian | Wong et al. |
| Fruel | Filipino | Other Asian | Wong et al. |
| Fruto | Filipino | Other Asian | Wong et al. |
| Fteha | Filipino | Other Asian | Wong et al. |
| Fu | Chinese | Chinese | Kranick et al. |
| Fua | Filipino | Other Asian | Wong et al. |
| Fuan | Chinese | Chinese | Wong et al. |
| Fuchigami | Japanese | Other Asian | Wong et al. |
| Fuchiwaki | Japanese | Other Asian | Wong et al. |
| Fuen | Chinese | Chinese | Wong et al. |
| Fuentebella | Filipino | Other Asian | Wong et al. |
| Fuentecilla | Filipino | Other Asian | Wong et al. |
| Fuentes | Filipino | Other Asian | Wong et al. |
| Fuerte | Filipino | Other Asian | Wong et al. |
| Fuertes | Filipino | Other Asian | Wong et al. |
| Fugimoto | Japanese | Other Asian | Wong et al. |
| Fugita | Japanese | Other Asian | Wong et al. |
| Fuh | Chinese | Chinese | Wong et al. |
| Fui | Chinese | Chinese | Wong et al. |
| Fuji | Japanese | Other Asian | Wong et al. |
| Fujibayashi | Japanese | Other Asian | Wong et al. |
| Fujie | Japanese | Other Asian | Wong et al. |
| Fujieda | Japanese | Other Asian | Wong et al. |
| Fujihara | Japanese | Other Asian | Wong et al. |
| Fujihira | Japanese | Other Asian | Wong et al. |
| Fujii | Japanese | Other Asian | Wong et al. |
| Fujikake | Japanese | Other Asian | Wong et al. |
| Fujikawa | Japanese | Other Asian | Wong et al. |
| Fujiki | Japanese | Other Asian | Wong et al. |
| Fujikura | Japanese | Other Asian | Wong et al. |
| Fujima | Japanese | Other Asian | Wong et al. |
| Fujimaki | Japanese | Other Asian | Wong et al. |
| Fujimori | Japanese | Other Asian | Wong et al. |
| Fujimoto | Japanese | Other Asian | Wong et al. |
| Fujimura | Japanese | Other Asian | Wong et al. |
| Fujinaga | Japanese | Other Asian | Wong et al. |
| Fujinaka | Japanese | Other Asian | Wong et al. |
| Fujinami | Japanese | Other Asian | Wong et al. |
| Fujino | Japanese | Other Asian | Wong et al. |
| Fujinuma | Japanese | Other Asian | Wong et al. |
| Fujio | Japanese | Other Asian | Wong et al. |
| Fujioka | Japanese | Other Asian | Wong et al. |
| Fujisaka | Japanese | Other Asian | Wong et al. |
| Fujisaki | Japanese | Other Asian | Wong et al. |
| Fujisawa | Japanese | Other Asian | Wong et al. |
| Fujise | Japanese | Other Asian | Wong et al. |
| Fujishige | Japanese | Other Asian | Wong et al. |
| Fujishima | Japanese | Other Asian | Wong et al. |
| Fujishiro | Japanese | Other Asian | Wong et al. |
| Fujita | Japanese | Other Asian | Wong et al. |
| Fujitani | Japanese | Other Asian | Wong et al. |
| Fujiura | Japanese | Other Asian | Wong et al. |
| Fujiwara | Japanese | Other Asian | Wong et al. |
| Fujiya | Japanese | Other Asian | Wong et al. |
| Fujiyama | Japanese | Other Asian | Wong et al. |
| Fujiyoshi | Japanese | Other Asian | Wong et al. |
| Fuk | Chinese | Chinese | Wong et al. |
| Fukabori | Japanese | Other Asian | Wong et al. |
| Fukada | Japanese | Other Asian | Wong et al. |
| Fukagawa | Japanese | Other Asian | Wong et al. |
| Fukahori | Japanese | Other Asian | Wong et al. |
| Fukai | Japanese | Other Asian | Wong et al. |
| Fukamachi | Japanese | Other Asian | Wong et al. |
| Fukami | Japanese | Other Asian | Wong et al. |
| Fukano | Japanese | Other Asian | Wong et al. |
| Fukao | Japanese | Other Asian | Wong et al. |
| Fukasawa | Japanese | Other Asian | Wong et al. |
| Fukase | Japanese | Other Asian | Wong et al. |
| Fukatsu | Japanese | Other Asian | Wong et al. |
| Fukawa | Japanese | Other Asian | Wong et al. |
| Fukaya | Japanese | Other Asian | Wong et al. |
| Fukazawa | Japanese | Other Asian | Wong et al. |
| Fuku | Japanese | Other Asian | Wong et al. |
| Fukuba | Japanese | Other Asian | Wong et al. |
| Fukuchi | Japanese | Other Asian | Wong et al. |
| Fukuda | Japanese | Other Asian | Kranick et al. |
| Fukuhara | Japanese | Other Asian | Wong et al. |
| Fukui | Japanese | Other Asian | Wong et al. |
| Fukuma | Japanese | Other Asian | Wong et al. |
| Fukumitsu | Japanese | Other Asian | Wong et al. |
| Fukumori | Japanese | Other Asian | Wong et al. |
| Fukumoto | Japanese | Other Asian | Wong et al. |
| Fukumura | Japanese | Other Asian | Wong et al. |
| Fukunaga | Japanese | Other Asian | Wong et al. |
| Fukunishi | Japanese | Other Asian | Wong et al. |
| Fukuno | Japanese | Other Asian | Wong et al. |
| Fukuoka | Japanese | Other Asian | Wong et al. |
| Fukushi | Japanese | Other Asian | Wong et al. |
| Fukushima | Japanese | Other Asian | Wong et al. |
| Fukuta | Japanese | Other Asian | Wong et al. |
| Fukutani | Japanese | Other Asian | Wong et al. |
| Fukutomi | Japanese | Other Asian | Wong et al. |
| Fukuya | Japanese | Other Asian | Wong et al. |
| Fukuyama | Japanese | Other Asian | Wong et al. |
| Fukuzaki | Japanese | Other Asian | Wong et al. |
| Fukuzawa | Japanese | Other Asian | Wong et al. |
| Fukuzumi | Japanese | Other Asian | Wong et al. |
| Fulabhai | Asian Indian | South Asian | Wong et al. |
| Fulache | Filipino | Other Asian | Wong et al. |
| Fulgado | Filipino | Other Asian | Wong et al. |
| Fulgar | Filipino | Other Asian | Wong et al. |
| Fulgencio | Filipino | Other Asian | Wong et al. |
| Fulgueras | Filipino | Other Asian | Wong et al. |
| Fulinara | Filipino | Other Asian | Wong et al. |
| Fulo | Filipino | Other Asian | Wong et al. |
| Fumar | Filipino | Other Asian | Wong et al. |
| Fumio | Japanese | Other Asian | Wong et al. |
| Fun | Chinese | Chinese | Wong et al. |
| Funabashi | Japanese | Other Asian | Wong et al. |
| Funada | Japanese | Other Asian | Wong et al. |
| Funahashi | Japanese | Other Asian | Wong et al. |
| Funai | Japanese | Other Asian | Wong et al. |
| Funaki | Japanese | Other Asian | Wong et al. |
| Funakoshi | Japanese | Other Asian | Wong et al. |
| Funamoto | Japanese | Other Asian | Wong et al. |
| Funaoka | Japanese | Other Asian | Wong et al. |
| Funato | Japanese | Other Asian | Wong et al. |
| Funatsu | Japanese | Other Asian | Wong et al. |
| Funayama | Japanese | Other Asian | Wong et al. |
| Funcion | Filipino | Other Asian | Wong et al. |
| Fune | Filipino | Other Asian | Wong et al. |
| Funelas | Filipino | Other Asian | Wong et al. |
| Fung | Chinese | Chinese | Kranick et al. |
| Funiestas | Filipino | Other Asian | Wong et al. |
| Funtanilla | Filipino | Other Asian | Wong et al. |
| Funtera | Filipino | Other Asian | Wong et al. |
| Funtila | Filipino | Other Asian | Wong et al. |
| Furagganan | Filipino | Other Asian | Wong et al. |
| Furbeyre | Filipino | Other Asian | Wong et al. |
| Furigay | Filipino | Other Asian | Wong et al. |
| Furihata | Japanese | Other Asian | Wong et al. |
| Furugori | Japanese | Other Asian | Wong et al. |
| Furuhashi | Japanese | Other Asian | Wong et al. |
| Furuhata | Japanese | Other Asian | Wong et al. |
| Furui | Japanese | Other Asian | Wong et al. |
| Furuichi | Japanese | Other Asian | Wong et al. |
| Furukawa | Japanese | Other Asian | Wong et al. |
| Furuki | Japanese | Other Asian | Wong et al. |
| Furumoto | Japanese | Other Asian | Wong et al. |
| Furuno | Japanese | Other Asian | Wong et al. |
| Furusaki | Japanese | Other Asian | Wong et al. |
| Furusawa | Japanese | Other Asian | Wong et al. |
| Furusho | Japanese | Other Asian | Wong et al. |
| Furuta | Japanese | Other Asian | Wong et al. |
| Furutani | Japanese | Other Asian | Wong et al. |
| Furuuchi | Japanese | Other Asian | Wong et al. |
| Furuya | Japanese | Other Asian | Wong et al. |
| Furuyama | Japanese | Other Asian | Wong et al. |
| Furuzawa | Japanese | Other Asian | Wong et al. |
| Fuse | Japanese | Other Asian | Wong et al. |
| Fushimi | Japanese | Other Asian | Wong et al. |
| Fusilero | Filipino | Other Asian | Wong et al. |
| Fuster | Filipino | Other Asian | Wong et al. |
| Fut | Chinese | Chinese | Wong et al. |
| Futaba | Japanese | Other Asian | Wong et al. |
| Futami | Japanese | Other Asian | Wong et al. |
| Futamura | Japanese | Other Asian | Wong et al. |
| Fuwa | Japanese | Other Asian | Wong et al. |
| G | Chinese | Chinese | Wong et al. |
| G.C. |  | South Asian | Lim et al. |
| Ga | Filipino | Other Asian | Wong et al. |
| Gaa | Filipino | Other Asian | Wong et al. |
| Gaan | Chinese | Chinese | Wong et al. |
| Gaba | Filipino | Other Asian | Wong et al. |
| Gabagat | Filipino | Other Asian | Wong et al. |
| Gabaldon | Filipino | Other Asian | Wong et al. |
| Gabarda | Filipino | Other Asian | Wong et al. |
| Gabat | Filipino | Other Asian | Wong et al. |
| Gabatin | Filipino | Other Asian | Wong et al. |
| Gabato | Filipino | Other Asian | Wong et al. |
| Gabay | Filipino | Other Asian | Wong et al. |
| Gabayan | Filipino | Other Asian | Wong et al. |
| Gabertan | Filipino | Other Asian | Wong et al. |
| Gabiana | Filipino | Other Asian | Wong et al. |
| Gabilo | Filipino | Other Asian | Wong et al. |
| Gabiola | Filipino | Other Asian | Wong et al. |
| Gabisan | Filipino | Other Asian | Wong et al. |
| Gabison | Filipino | Other Asian | Wong et al. |
| Gabito | Filipino | Other Asian | Wong et al. |
| Gabo | Filipino | Other Asian | Wong et al. |
| Gabon | Filipino | Other Asian | Wong et al. |
| Gaborne | Filipino | Other Asian | Wong et al. |
| Gabot | Filipino | Other Asian | Wong et al. |
| Gaboya | Filipino | Other Asian | Wong et al. |
| Gabres | Filipino | Other Asian | Wong et al. |
| Gabriana | Filipino | Other Asian | Wong et al. |
| Gabrillo | Filipino | Other Asian | Wong et al. |
| Gabrino | Filipino | Other Asian | Wong et al. |
| Gabriola | Filipino | Other Asian | Wong et al. |
| Gabuat | Filipino | Other Asian | Wong et al. |
| Gabutin | Filipino | Other Asian | Wong et al. |
| Gabuya | Filipino | Other Asian | Wong et al. |
| Gabuyo | Filipino | Other Asian | Wong et al. |
| Gacad | Filipino | Other Asian | Wong et al. |
| Gacayan | Filipino | Other Asian | Wong et al. |
| Gaces | Filipino | Other Asian | Wong et al. |
| Gaceta | Filipino | Other Asian | Wong et al. |
| Gachallan | Filipino | Other Asian | Wong et al. |
| Gacho | Filipino | Other Asian | Wong et al. |
| Gacias | Filipino | Other Asian | Wong et al. |
| Gacis | Filipino | Other Asian | Wong et al. |
| Gaco | Filipino | Other Asian | Wong et al. |
| Gacos | Filipino | Other Asian | Wong et al. |
| Gacoscos | Filipino | Other Asian | Wong et al. |
| Gacula | Filipino | Other Asian | Wong et al. |
| Gacusan | Filipino | Other Asian | Wong et al. |
| Gacusana | Filipino | Other Asian | Wong et al. |
| Gacutan | Filipino | Other Asian | Wong et al. |
| Gacuya | Filipino | Other Asian | Wong et al. |
| Gaddam | Asian Indian | South Asian | Wong et al. |
| Gaddi | Filipino | Other Asian | Wong et al. |
| Gadgil | Asian Indian | South Asian | Wong et al. |
| Gadhia | Asian Indian | South Asian | Wong et al. |
| Gadhoke | Asian Indian | South Asian | Wong et al. |
| Gadi | Filipino | Other Asian | Wong et al. |
| Gadia | Filipino | Other Asian | Wong et al. |
| Gadiane | Filipino | Other Asian | Wong et al. |
| Gadiano | Filipino | Other Asian | Wong et al. |
| Gadingan | Filipino | Other Asian | Wong et al. |
| Gadkari | Asian Indian | South Asian | Wong et al. |
| Gadon | Filipino | Other Asian | Wong et al. |
| Gadong | Filipino | Other Asian | Wong et al. |
| Gador | Filipino | Other Asian | Wong et al. |
| Gadre | Asian Indian | South Asian | Wong et al. |
| Gaela | Filipino | Other Asian | Wong et al. |
| Gaerlan | Filipino | Other Asian | Wong et al. |
| Gaetos | Filipino | Other Asian | Wong et al. |
| Gaffar | Asian Indian | South Asian | Wong et al. |
| Gaffud | Filipino | Other Asian | Wong et al. |
| Gaffur | Asian Indian | South Asian | Wong et al. |
| Gafoor | Asian Indian | South Asian | Wong et al. |
| Gafur |  | South Asian | Lim et al. |
| Gagalang | Filipino | Other Asian | Wong et al. |
| Gagarin | Filipino | Other Asian | Wong et al. |
| Gaglani | Asian Indian | South Asian | Wong et al. |
| Gagni | Filipino | Other Asian | Wong et al. |
| Gago | Filipino | Other Asian | Wong et al. |
| Gagtan | Filipino | Other Asian | Wong et al. |
| Gagui | Filipino | Other Asian | Wong et al. |
| Gahol | Filipino | Other Asian | Wong et al. |
| Gahunia | Asian Indian | South Asian | Wong et al. |
| Gai | Chinese | Chinese | Wong et al. |
| Gaikwad | Asian Indian | South Asian | Wong et al. |
| Gaind | Asian Indian | South Asian | Wong et al. |
| Gaitan | Filipino | Other Asian | Wong et al. |
| Gaitonde | Asian Indian | South Asian | Wong et al. |
| Gajardo | Filipino | Other Asian | Wong et al. |
| Gajera | Asian Indian | South Asian | Wong et al. |
| Gajes | Filipino | Other Asian | Wong et al. |
| Gajete | Filipino | Other Asian | Wong et al. |
| Gajeton | Filipino | Other Asian | Wong et al. |
| Gajiwala | Asian Indian | South Asian | Wong et al. |
| Gajjar | Asian Indian | South Asian | Wong et al. |
| Gajo | Filipino | Other Asian | Wong et al. |
| Gajudo | Filipino | Other Asian | Wong et al. |
| Gakiya | Japanese | Other Asian | Wong et al. |
| Gako | Filipino | Other Asian | Wong et al. |
| Gala | Asian Indian | South Asian | Wong et al. |
| Galace | Filipino | Other Asian | Wong et al. |
| Galacgac | Filipino | Other Asian | Wong et al. |
| Galagar | Filipino | Other Asian | Wong et al. |
| Galam | Filipino | Other Asian | Wong et al. |
| Galamay | Filipino | Other Asian | Wong et al. |
| Galamgam | Filipino | Other Asian | Wong et al. |
| Galan | Filipino | Other Asian | Wong et al. |
| Galang | Filipino | Other Asian | Wong et al. |
| Galanga | Filipino | Other Asian | Wong et al. |
| Galano | Filipino | Other Asian | Wong et al. |
| Galanta | Filipino | Other Asian | Wong et al. |
| Galanto | Filipino | Other Asian | Wong et al. |
| Galanza | Filipino | Other Asian | Wong et al. |
| Galapate | Filipino | Other Asian | Wong et al. |
| Galapia | Filipino | Other Asian | Wong et al. |
| Galapin | Filipino | Other Asian | Wong et al. |
| Galapir | Filipino | Other Asian | Wong et al. |
| Galapon | Filipino | Other Asian | Wong et al. |
| Galario | Filipino | Other Asian | Wong et al. |
| Galarpe | Filipino | Other Asian | Wong et al. |
| Galasinao | Filipino | Other Asian | Wong et al. |
| Galbadores | Filipino | Other Asian | Wong et al. |
| Galban | Filipino | Other Asian | Wong et al. |
| Galdo | Filipino | Other Asian | Wong et al. |
| Galdones | Filipino | Other Asian | Wong et al. |
| Galedo | Filipino | Other Asian | Wong et al. |
| Galendez | Filipino | Other Asian | Wong et al. |
| Galeno | Filipino | Other Asian | Wong et al. |
| Galenzoga | Filipino | Other Asian | Wong et al. |
| Galeon | Filipino | Other Asian | Wong et al. |
| Galeos | Filipino | Other Asian | Wong et al. |
| Galera | Filipino | Other Asian | Wong et al. |
| Gali | Filipino | Other Asian | Wong et al. |
| Galicha | Filipino | Other Asian | Wong et al. |
| Galicia | Filipino | Other Asian | Wong et al. |
| Galicinao | Filipino | Other Asian | Wong et al. |
| Galido | Filipino | Other Asian | Wong et al. |
| Galigo | Filipino | Other Asian | Wong et al. |
| Galila | Filipino | Other Asian | Wong et al. |
| Galima | Filipino | Other Asian | Wong et al. |
| Galimba | Filipino | Other Asian | Wong et al. |
| Galinato | Filipino | Other Asian | Wong et al. |
| Galindez | Filipino | Other Asian | Wong et al. |
| Galindo | Filipino | Other Asian | Wong et al. |
| Galing | Filipino | Other Asian | Wong et al. |
| Galingan | Filipino | Other Asian | Wong et al. |
| Galino | Filipino | Other Asian | Wong et al. |
| Galiste | Filipino | Other Asian | Wong et al. |
| Galit | Filipino | Other Asian | Wong et al. |
| Galito | Filipino | Other Asian | Wong et al. |
| Galiza | Filipino | Other Asian | Wong et al. |
| Gallang | Filipino | Other Asian | Wong et al. |
| Gallano | Filipino | Other Asian | Wong et al. |
| Gallanosa | Filipino | Other Asian | Wong et al. |
| Gallarde | Filipino | Other Asian | Wong et al. |
| Gallardo | Filipino | Other Asian | Wong et al. |
| Gallarin | Filipino | Other Asian | Wong et al. |
| Gallego | Filipino | Other Asian | Wong et al. |
| Gallegos | Filipino | Other Asian | Wong et al. |
| Gallemit | Filipino | Other Asian | Wong et al. |
| Gallenero | Filipino | Other Asian | Wong et al. |
| Galleposo | Filipino | Other Asian | Wong et al. |
| Gallero | Filipino | Other Asian | Wong et al. |
| Galleto | Filipino | Other Asian | Wong et al. |
| Gallevo | Filipino | Other Asian | Wong et al. |
| Galliguez | Filipino | Other Asian | Wong et al. |
| Gallofin | Filipino | Other Asian | Wong et al. |
| Galman | Filipino | Other Asian | Wong et al. |
| Galo | Filipino | Other Asian | Wong et al. |
| Galolo | Filipino | Other Asian | Wong et al. |
| Galon | Filipino | Other Asian | Wong et al. |
| Galope | Filipino | Other Asian | Wong et al. |
| Galsim | Filipino | Other Asian | Wong et al. |
| Galsote | Filipino | Other Asian | Wong et al. |
| Galunas |  | South Asian | Lim et al. |
| Galupe | Filipino | Other Asian | Wong et al. |
| Galura | Filipino | Other Asian | Wong et al. |
| Galut | Filipino | Other Asian | Wong et al. |
| Galutera | Filipino | Other Asian | Wong et al. |
| Galutira | Filipino | Other Asian | Wong et al. |
| Galvan | Filipino | Other Asian | Wong et al. |
| Galvez | Filipino | Other Asian | Wong et al. |
| Galvizo | Filipino | Other Asian | Wong et al. |
| Galzote | Filipino | Other Asian | Wong et al. |
| Gam | Chinese | Chinese | Wong et al. |
| Gama | Asian Indian | South Asian | Wong et al. |
| Gamab | Filipino | Other Asian | Wong et al. |
| Gamad | Filipino | Other Asian | Wong et al. |
| Gamadia | Asian Indian | South Asian | Wong et al. |
| Gamalinda | Filipino | Other Asian | Wong et al. |
| Gamalo | Filipino | Other Asian | Wong et al. |
| Gamata | Filipino | Other Asian | Wong et al. |
| Gamatero | Filipino | Other Asian | Wong et al. |
| Gamayo | Filipino | Other Asian | Wong et al. |
| Gamayon | Filipino | Other Asian | Wong et al. |
| Gamazon | Filipino | Other Asian | Wong et al. |
| Gambalan | Filipino | Other Asian | Wong et al. |
| Gamban | Filipino | Other Asian | Wong et al. |
| Gambe | Filipino | Other Asian | Wong et al. |
| Gambito | Filipino | Other Asian | Wong et al. |
| Gamboa | Filipino | Other Asian | Wong et al. |
| Gambol | Filipino | Other Asian | Wong et al. |
| Gambon | Filipino | Other Asian | Wong et al. |
| Gamel |  | South Asian | Lim et al. |
| Gameng | Filipino | Other Asian | Wong et al. |
| Gamet | Filipino | Other Asian | Wong et al. |
| Gamez | Filipino | Other Asian | Wong et al. |
| Gami | Asian Indian | South Asian | Wong et al. |
| Gamiao | Filipino | Other Asian | Wong et al. |
| Gamido | Filipino | Other Asian | Wong et al. |
| Gamilla | Filipino | Other Asian | Wong et al. |
| Gamis | Filipino | Other Asian | Wong et al. |
| Gamit | Asian Indian | South Asian | Kranick et al. |
| Gammad | Filipino | Other Asian | Wong et al. |
| Gamo | Filipino | Other Asian | Wong et al. |
| Gamos | Filipino | Other Asian | Wong et al. |
| Gamoso | Filipino | Other Asian | Wong et al. |
| Gampon | Filipino | Other Asian | Wong et al. |
| Gampong | Filipino | Other Asian | Wong et al. |
| Gamponia | Filipino | Other Asian | Wong et al. |
| Gamueda | Filipino | Other Asian | Wong et al. |
| Gamulo | Filipino | Other Asian | Wong et al. |
| Gamundoy | Filipino | Other Asian | Wong et al. |
| Gamutan | Filipino | Other Asian | Wong et al. |
| Gan | Chinese | Chinese | Wong et al. |
| Ganaban | Filipino | Other Asian | Wong et al. |
| Ganac | Filipino | Other Asian | Wong et al. |
| Ganacias | Filipino | Other Asian | Wong et al. |
| Ganaden | Filipino | Other Asian | Wong et al. |
| Ganado | Filipino | Other Asian | Wong et al. |
| Ganal | Filipino | Other Asian | Wong et al. |
| Ganalon | Filipino | Other Asian | Wong et al. |
| Ganan | Filipino | Other Asian | Wong et al. |
| Ganancial | Filipino | Other Asian | Wong et al. |
| Ganapathi | Asian Indian | South Asian | Wong et al. |
| Ganapathy | Asian Indian | South Asian | Wong et al. |
| Ganarial | Filipino | Other Asian | Wong et al. |
| Ganatra | Asian Indian | South Asian | Wong et al. |
| Gancayco | Filipino | Other Asian | Wong et al. |
| Gancena | Filipino | Other Asian | Wong et al. |
| Gandeza | Filipino | Other Asian | Wong et al. |
| Gandhi | Asian Indian | South Asian | Wong et al. |
| Gandi | Asian Indian | South Asian | Wong et al. |
| Gandia | Filipino | Other Asian | Wong et al. |
| Ganding | Filipino | Other Asian | Wong et al. |
| Gandionco | Filipino | Other Asian | Wong et al. |
| Gandotra | Asian Indian | South Asian | Wong et al. |
| Ganeko | Japanese | Other Asian | Wong et al. |
| Ganesan | Asian Indian | South Asian | Wong et al. |
| Ganesh | Asian Indian | South Asian | Kranick et al. |
| Gangadeen |  | South Asian | Lim et al. |
| Gangadharan | Asian Indian | South Asian | Wong et al. |
| Gangal | Asian Indian | South Asian | Wong et al. |
| Gangano | Filipino | Other Asian | Wong et al. |
| Gangaram |  | South Asian | Lim et al. |
| Gangarum |  | South Asian | Lim et al. |
| Gangji | Asian Indian | South Asian | Wong et al. |
| Gangopadhyay | Asian Indian | South Asian | Wong et al. |
| Ganguli | Asian Indian | South Asian | Wong et al. |
| Ganguly | Asian Indian | South Asian | Wong et al. |
| Gani | Asian Indian | South Asian | Wong et al. |
| Gania | Filipino | Other Asian | Wong et al. |
| Ganiban | Filipino | Other Asian | Wong et al. |
| Ganibe | Filipino | Other Asian | Wong et al. |
| Ganibi | Filipino | Other Asian | Wong et al. |
| Ganigan | Filipino | Other Asian | Wong et al. |
| Ganio | Filipino | Other Asian | Wong et al. |
| Ganir | Filipino | Other Asian | Wong et al. |
| Ganiron | Filipino | Other Asian | Wong et al. |
| Ganitano | Filipino | Other Asian | Wong et al. |
| Ganju | Asian Indian | South Asian | Wong et al. |
| Gannaban | Filipino | Other Asian | Wong et al. |
| Ganny |  | South Asian | Lim et al. |
| Ganon | Filipino | Other Asian | Wong et al. |
| Ganotisi | Filipino | Other Asian | Wong et al. |
| Ganoy | Filipino | Other Asian | Wong et al. |
| Gantala | Filipino | Other Asian | Wong et al. |
| Gantan | Filipino | Other Asian | Wong et al. |
| Gante | Filipino | Other Asian | Wong et al. |
| Ganti | Asian Indian | South Asian | Wong et al. |
| Ganuelas | Filipino | Other Asian | Wong et al. |
| Ganzagan | Filipino | Other Asian | Wong et al. |
| Ganzon | Filipino | Other Asian | Wong et al. |
| Gao | Chinese | Chinese | Kranick et al. |
| Gaoat | Filipino | Other Asian | Wong et al. |
| Gaoay | Filipino | Other Asian | Wong et al. |
| Gaoiran | Filipino | Other Asian | Wong et al. |
| Gaor | Filipino | Other Asian | Wong et al. |
| Gapas | Filipino | Other Asian | Wong et al. |
| Gapasin | Filipino | Other Asian | Wong et al. |
| Gapero | Filipino | Other Asian | Wong et al. |
| Gapol | Filipino | Other Asian | Wong et al. |
| Gappi | Filipino | Other Asian | Wong et al. |
| Gapultos | Filipino | Other Asian | Wong et al. |
| Gapusan | Filipino | Other Asian | Wong et al. |
| Gapuz | Filipino | Other Asian | Wong et al. |
| Gapuzan | Filipino | Other Asian | Wong et al. |
| Gar | Chinese | Chinese | Wong et al. |
| Garabato | Filipino | Other Asian | Wong et al. |
| Garabiles | Filipino | Other Asian | Wong et al. |
| Garado | Filipino | Other Asian | Wong et al. |
| Garalde | Filipino | Other Asian | Wong et al. |
| Garan | Filipino | Other Asian | Wong et al. |
| Garay | Filipino | Other Asian | Wong et al. |
| Garaza | Filipino | Other Asian | Wong et al. |
| Garcellano | Filipino | Other Asian | Wong et al. |
| Garcera | Filipino | Other Asian | Wong et al. |
| Garces | Filipino | Other Asian | Wong et al. |
| Garcha | Asian Indian | South Asian | Wong et al. |
| Garchitorena | Filipino | Other Asian | Wong et al. |
| Garcia | Filipino | Other Asian | Wong et al. |
| Garciano | Filipino | Other Asian | Wong et al. |
| Garcines | Filipino | Other Asian | Wong et al. |
| Gardoce | Filipino | Other Asian | Wong et al. |
| Garduce | Filipino | Other Asian | Wong et al. |
| Garduno | Filipino | Other Asian | Wong et al. |
| Garduque | Filipino | Other Asian | Wong et al. |
| Garewal | Asian Indian | South Asian | Wong et al. |
| Garfil | Filipino | Other Asian | Wong et al. |
| Garfin | Filipino | Other Asian | Wong et al. |
| Garg | Asian Indian | South Asian | Wong et al. |
| Garganera | Filipino | Other Asian | Wong et al. |
| Garganta | Filipino | Other Asian | Wong et al. |
| Gargantiel | Filipino | Other Asian | Wong et al. |
| Gargarita | Filipino | Other Asian | Wong et al. |
| Gargaritano | Filipino | Other Asian | Wong et al. |
| Gariando | Filipino | Other Asian | Wong et al. |
| Garibay | Filipino | Other Asian | Wong et al. |
| Garilao | Filipino | Other Asian | Wong et al. |
| Garin | Filipino | Other Asian | Wong et al. |
| Garingalao | Filipino | Other Asian | Wong et al. |
| Garingan | Filipino | Other Asian | Wong et al. |
| Garingo | Filipino | Other Asian | Wong et al. |
| Garlejo | Filipino | Other Asian | Wong et al. |
| Garlit | Filipino | Other Asian | Wong et al. |
| Garlitos | Filipino | Other Asian | Wong et al. |
| Garma | Filipino | Other Asian | Wong et al. |
| Garo | Filipino | Other Asian | Wong et al. |
| Garong | Filipino | Other Asian | Wong et al. |
| Garrido | Filipino | Other Asian | Wong et al. |
| Garriz | Filipino | Other Asian | Wong et al. |
| Garrovillas | Filipino | Other Asian | Wong et al. |
| Garrovillo | Filipino | Other Asian | Wong et al. |
| Garudachar | Asian Indian | South Asian | Wong et al. |
| Garvida | Filipino | Other Asian | Wong et al. |
| Garza | Chinese | Chinese | Wong et al. |
| Garzon | Filipino | Other Asian | Wong et al. |
| Gasataya | Filipino | Other Asian | Wong et al. |
| Gascon | Filipino | Other Asian | Wong et al. |
| Gasgonia | Filipino | Other Asian | Wong et al. |
| Gasmen | Filipino | Other Asian | Wong et al. |
| Gasmin | Filipino | Other Asian | Wong et al. |
| Gaspar | Filipino | Other Asian | Wong et al. |
| Gat | Chinese | Chinese | Wong et al. |
| Gata | Filipino | Other Asian | Wong et al. |
| Gatal | Filipino | Other Asian | Wong et al. |
| Gatan | Filipino | Other Asian | Wong et al. |
| Gatbonton | Filipino | Other Asian | Wong et al. |
| Gatbunton | Filipino | Other Asian | Wong et al. |
| Gatchalian | Filipino | Other Asian | Kranick et al. |
| Gatdula | Filipino | Other Asian | Wong et al. |
| Gateb | Filipino | Other Asian | Wong et al. |
| Gatela | Filipino | Other Asian | Wong et al. |
| Gatmaitan | Filipino | Other Asian | Wong et al. |
| Gatmen | Filipino | Other Asian | Wong et al. |
| Gato | Filipino | Other Asian | Wong et al. |
| Gaton | Filipino | Other Asian | Wong et al. |
| Gatpandan | Filipino | Other Asian | Wong et al. |
| Gatus | Filipino | Other Asian | Wong et al. |
| Gaudia | Filipino | Other Asian | Wong et al. |
| Gaudinez | Filipino | Other Asian | Wong et al. |
| Gaur | Asian Indian | South Asian | Wong et al. |
| Gaurano | Filipino | Other Asian | Wong et al. |
| Gautam | Asian Indian | South Asian | Wong et al. |
| Gavankar | Asian Indian | South Asian | Wong et al. |
| Gavara | Asian Indian | South Asian | Kranick et al. |
| Gavieres | Filipino | Other Asian | Wong et al. |
| Gavilan | Filipino | Other Asian | Wong et al. |
| Gavina | Filipino | Other Asian | Wong et al. |
| Gavino | Filipino | Other Asian | Wong et al. |
| Gaviola | Filipino | Other Asian | Wong et al. |
| Gawaran | Filipino | Other Asian | Wong et al. |
| Gawat | Filipino | Other Asian | Wong et al. |
| Gayagoy | Filipino | Other Asian | Wong et al. |
| Gayamat | Filipino | Other Asian | Wong et al. |
| Gayanilo | Filipino | Other Asian | Wong et al. |
| Gayap | Filipino | Other Asian | Wong et al. |
| Gayares | Filipino | Other Asian | Wong et al. |
| Gayatin | Filipino | Other Asian | Wong et al. |
| Gayeta | Filipino | Other Asian | Wong et al. |
| Gayo | Filipino | Other Asian | Wong et al. |
| Gayon | Filipino | Other Asian | Wong et al. |
| Gayoso | Filipino | Other Asian | Wong et al. |
| Gayotin | Filipino | Other Asian | Wong et al. |
| Gaza | Filipino | Other Asian | Wong et al. |
| Gazi |  | South Asian | Lim et al. |
| Gazmen | Filipino | Other Asian | Wong et al. |
| Ge | Chinese | Chinese | Wong et al. |
| Geaga | Filipino | Other Asian | Wong et al. |
| Gee | Chinese | Chinese | Wong et al. |
| Geevarghese | Asian Indian | South Asian | Wong et al. |
| Geevarughese | Asian Indian | South Asian | Wong et al. |
| Gega | Filipino | Other Asian | Wong et al. |
| Gelacio | Filipino | Other Asian | Wong et al. |
| Gelera | Filipino | Other Asian | Wong et al. |
| Geli | Filipino | Other Asian | Wong et al. |
| Gelito | Filipino | Other Asian | Wong et al. |
| Gella | Filipino | Other Asian | Wong et al. |
| Gelle | Filipino | Other Asian | Wong et al. |
| Gellido | Filipino | Other Asian | Wong et al. |
| Geluz | Filipino | Other Asian | Wong et al. |
| Gelvezon | Filipino | Other Asian | Wong et al. |
| Gem | Chinese | Chinese | Wong et al. |
| Gemeniano | Filipino | Other Asian | Wong et al. |
| Gemina | Filipino | Other Asian | Wong et al. |
| Geminiano | Filipino | Other Asian | Wong et al. |
| Gemo | Filipino | Other Asian | Wong et al. |
| Gemora | Filipino | Other Asian | Wong et al. |
| Gempesaw | Filipino | Other Asian | Wong et al. |
| Gen | Chinese | Chinese | Wong et al. |
| Genato | Filipino | Other Asian | Wong et al. |
| Gendrano | Filipino | Other Asian | Wong et al. |
| Gene | Chinese | Chinese | Wong et al. |
| Geneciran | Filipino | Other Asian | Wong et al. |
| Gener | Filipino | Other Asian | Wong et al. |
| General | Filipino | Other Asian | Wong et al. |
| Generalao | Filipino | Other Asian | Wong et al. |
| Generao | Filipino | Other Asian | Wong et al. |
| Generosa | Filipino | Other Asian | Wong et al. |
| Generoso | Filipino | Other Asian | Wong et al. |
| Geneta | Filipino | Other Asian | Wong et al. |
| Genetiano | Filipino | Other Asian | Wong et al. |
| Geng | Chinese | Chinese | Kranick et al. |
| Genido | Filipino | Other Asian | Wong et al. |
| Genil | Filipino | Other Asian | Wong et al. |
| Genilo | Filipino | Other Asian | Wong et al. |
| Genio | Filipino | Other Asian | Wong et al. |
| Geniza | Filipino | Other Asian | Wong et al. |
| Genka | Japanese | Other Asian | Wong et al. |
| Genolaga | Filipino | Other Asian | Wong et al. |
| Genove | Filipino | Other Asian | Wong et al. |
| Genovia | Filipino | Other Asian | Wong et al. |
| Genuino | Filipino | Other Asian | Wong et al. |
| Geolingo | Filipino | Other Asian | Wong et al. |
| Geonzon | Filipino | Other Asian | Wong et al. |
| Georgekutty | Asian Indian | South Asian | Wong et al. |
| Geotina | Filipino | Other Asian | Wong et al. |
| Gepalaga | Filipino | Other Asian | Wong et al. |
| Gepuela | Filipino | Other Asian | Wong et al. |
| Gequillana | Filipino | Other Asian | Wong et al. |
| Ger | Chinese | Chinese | Wong et al. |
| Geralde | Filipino | Other Asian | Wong et al. |
| Geraldez | Filipino | Other Asian | Wong et al. |
| Geraldo | Filipino | Other Asian | Wong et al. |
| Gerali | Filipino | Other Asian | Wong et al. |
| Gerardo | Filipino | Other Asian | Wong et al. |
| Gerez | Filipino | Other Asian | Wong et al. |
| Germar | Filipino | Other Asian | Wong et al. |
| Germino | Filipino | Other Asian | Wong et al. |
| Germono | Filipino | Other Asian | Wong et al. |
| Gernale | Filipino | Other Asian | Wong et al. |
| Gerochi | Filipino | Other Asian | Wong et al. |
| Gerodias | Filipino | Other Asian | Wong et al. |
| Gerona | Filipino | Other Asian | Wong et al. |
| Geronca | Filipino | Other Asian | Wong et al. |
| Geronilla | Filipino | Other Asian | Wong et al. |
| Geronimo | Filipino | Other Asian | Wong et al. |
| Gerozaga | Filipino | Other Asian | Wong et al. |
| Gervacio | Filipino | Other Asian | Wong et al. |
| Gervado | Filipino | Other Asian | Wong et al. |
| Gesite | Filipino | Other Asian | Wong et al. |
| Geslani | Filipino | Other Asian | Wong et al. |
| Gesmundo | Filipino | Other Asian | Wong et al. |
| Gestiada | Filipino | Other Asian | Wong et al. |
| Gestopa | Filipino | Other Asian | Wong et al. |
| Gestoso | Filipino | Other Asian | Wong et al. |
| Gesulga | Filipino | Other Asian | Wong et al. |
| Get | Chinese | Chinese | Wong et al. |
| Getubig | Filipino | Other Asian | Wong et al. |
| Gevarghese | Asian Indian | South Asian | Wong et al. |
| Gew | Chinese | Chinese | Wong et al. |
| Ghadiali | Asian Indian | South Asian | Wong et al. |
| Ghadialy | Asian Indian | South Asian | Wong et al. |
| Ghaffar |  | South Asian | Lim et al. |
| Ghafoor | Asian Indian | South Asian | Wong et al. |
| Ghag | Asian Indian | South Asian | Wong et al. |
| Ghaghada |  | South Asian | Lim et al. |
| Ghai | Asian Indian | South Asian | Kranick et al. |
| Ghandhi | Asian Indian | South Asian | Wong et al. |
| Ghanekar | Asian Indian | South Asian | Wong et al. |
| Ghang | Korean | Other Asian | Wong et al. |
| Ghani |  | South Asian | Lim et al. |
| Ghanny |  | South Asian | Lim et al. |
| Gharda | Asian Indian | South Asian | Wong et al. |
| Ghartichhetry |  | South Asian | Lim et al. |
| Ghaswala | Asian Indian | South Asian | Wong et al. |
| Ghatak | Asian Indian | South Asian | Wong et al. |
| Ghatala | Asian Indian | South Asian | Wong et al. |
| Ghatalia | Asian Indian | South Asian | Wong et al. |
| Ghate | Asian Indian | South Asian | Wong et al. |
| Ghazala |  | South Asian | Lim et al. |
| Ghaznavi | Asian Indian | South Asian | Wong et al. |
| Ghee |  | South Asian | Lim et al. |
| Gheewala | Asian Indian | South Asian | Wong et al. |
| Ghelani | Asian Indian | South Asian | Wong et al. |
| Ghevarghese | Asian Indian | South Asian | Wong et al. |
| Ghia | Asian Indian | South Asian | Wong et al. |
| Ghim | Korean | Other Asian | Wong et al. |
| Ghimire |  | South Asian | Lim et al. |
| Ghisiawan |  | South Asian | Lim et al. |
| Ghiya | Asian Indian | South Asian | Wong et al. |
| Ghong | Korean | Other Asian | Wong et al. |
| Ghorai | Asian Indian | South Asian | Wong et al. |
| Ghori | Asian Indian | South Asian | Wong et al. |
| Ghosal | Asian Indian | South Asian | Wong et al. |
| Ghose | Asian Indian | South Asian | Wong et al. |
| Ghosh | Asian Indian; Bangladeshi | South Asian | Kranick et al. |
| Ghoshal | Asian Indian | South Asian | Kranick et al. |
| Ghotra |  | South Asian | Lim et al. |
| Ghouse | Asian Indian | South Asian | Wong et al. |
| Ghulam |  | South Asian | Lim et al. |
| Ghuman | Asian Indian | South Asian | Wong et al. |
| Ghumman | Asian Indian | South Asian | Wong et al. |
| Gi | Korean | Other Asian | Wong et al. |
| Gia | Vietnamese | Other Asian | Wong et al. |
| Giai | Vietnamese | Other Asian | Wong et al. |
| Giam | Vietnamese | Other Asian | Wong et al. |
| Gian | Vietnamese | Other Asian | Wong et al. |
| Gianan | Filipino | Other Asian | Wong et al. |
| Giang | Vietnamese | Other Asian | Wong et al. |
| Giao | Vietnamese | Other Asian | Wong et al. |
| Giap | Vietnamese | Other Asian | Wong et al. |
| Giau | Vietnamese | Other Asian | Wong et al. |
| Gibo | Japanese | Other Asian | Wong et al. |
| Gibu | Japanese | Other Asian | Wong et al. |
| Gica | Filipino | Other Asian | Wong et al. |
| Gidda | Asian Indian | South Asian | Wong et al. |
| Gieng | Vietnamese | Other Asian | Wong et al. |
| Giga | Asian Indian | South Asian | Wong et al. |
| Gil | Filipino | Other Asian | Wong et al. |
| Gilani | Asian Indian | South Asian | Wong et al. |
| Gilbuena | Filipino | Other Asian | Wong et al. |
| Gildore | Filipino | Other Asian | Wong et al. |
| Gilera | Filipino | Other Asian | Wong et al. |
| Gill |  | South Asian | Lim et al. |
| Gillego | Filipino | Other Asian | Wong et al. |
| Gilo | Filipino | Other Asian | Wong et al. |
| Gim | Korean | Other Asian | Kranick et al. |
| Gima | Japanese | Other Asian | Wong et al. |
| Gimena | Filipino | Other Asian | Wong et al. |
| Gimenez | Filipino | Other Asian | Wong et al. |
| Gimeno | Filipino | Other Asian | Wong et al. |
| Gimotea | Filipino | Other Asian | Wong et al. |
| Gin | Chinese | Chinese | Wong et al. |
| Giner | Filipino | Other Asian | Wong et al. |
| Gines | Filipino | Other Asian | Wong et al. |
| Ginete | Filipino | Other Asian | Wong et al. |
| Ginez | Filipino | Other Asian | Wong et al. |
| Ging | Chinese | Chinese | Wong et al. |
| Gingco | Filipino | Other Asian | Wong et al. |
| Ginoza | Japanese | Other Asian | Wong et al. |
| Gioi | Vietnamese | Other Asian | Wong et al. |
| Gip | Vietnamese | Other Asian | Wong et al. |
| Gir | Asian Indian | South Asian | Wong et al. |
| Girado | Filipino | Other Asian | Wong et al. |
| Girdhari |  | South Asian | Lim et al. |
| Girdharie |  | South Asian | Lim et al. |
| Girdharlal | Asian Indian | South Asian | Wong et al. |
| Giri | Asian Indian | South Asian | Wong et al. |
| Giron | Filipino | Other Asian | Wong et al. |
| Gironella | Filipino | Other Asian | Wong et al. |
| Gison | Filipino | Other Asian | Wong et al. |
| Git | Chinese | Chinese | Wong et al. |
| Gitana | Filipino | Other Asian | Wong et al. |
| Giu | Chinese | Chinese | Wong et al. |
| Glindro | Filipino | Other Asian | Wong et al. |
| Gloria | Filipino | Other Asian | Wong et al. |
| Gloriani | Filipino | Other Asian | Wong et al. |
| Go | Korean | Other Asian | Kranick et al. |
| Gobenciong | Filipino | Other Asian | Wong et al. |
| Gobin |  | South Asian | Lim et al. |
| Gobind,1 |  | South Asian | Lim et al. |
| Gobuyan | Filipino | Other Asian | Wong et al. |
| Goce | Filipino | Other Asian | Wong et al. |
| Gocho | Japanese | Other Asian | Wong et al. |
| Gock | Chinese | Chinese | Wong et al. |
| Goco | Filipino | Other Asian | Wong et al. |
| Goda | Japanese | Other Asian | Wong et al. |
| Godbole | Asian Indian | South Asian | Wong et al. |
| Godil | Asian Indian | South Asian | Wong et al. |
| Godinez | Filipino | Other Asian | Wong et al. |
| Godinho | Asian Indian | South Asian | Wong et al. |
| Godiwala | Asian Indian | South Asian | Wong et al. |
| Godoy | Filipino | Other Asian | Wong et al. |
| Godse | Asian Indian | South Asian | Wong et al. |
| Goduco | Filipino | Other Asian | Wong et al. |
| Goel | Asian Indian | South Asian | Wong et al. |
| Goenka | Asian Indian | South Asian | Wong et al. |
| Goey | Chinese | Chinese | Wong et al. |
| Gogineni | Asian Indian | South Asian | Wong et al. |
| Gogna | Asian Indian | South Asian | Wong et al. |
| Gogo | Filipino | Other Asian | Wong et al. |
| Goh | Chinese; Malaysian | Chinese | Kranick et al. |
| Gohel | Asian Indian | South Asian | Wong et al. |
| Gohil | Asian Indian | South Asian | Wong et al. |
| Goingco | Filipino | Other Asian | Wong et al. |
| Goitia | Filipino | Other Asian | Wong et al. |
| Gok | Chinese | Chinese | Wong et al. |
| Gokal | Asian Indian | South Asian | Wong et al. |
| Gokhale | Asian Indian | South Asian | Wong et al. |
| Gokita | Japanese | Other Asian | Wong et al. |
| Golam |  | South Asian | Lim et al. |
| Golder |  | South Asian | Lim et al. |
| Gole |  | South Asian | Lim et al. |
| Goleco | Filipino | Other Asian | Wong et al. |
| Goleta | Filipino | Other Asian | Wong et al. |
| Golez | Filipino | Other Asian | Wong et al. |
| Golfo | Filipino | Other Asian | Wong et al. |
| Golikeri | Asian Indian | South Asian | Wong et al. |
| Golingo | Filipino | Other Asian | Wong et al. |
| Gollakota | Asian Indian | South Asian | Wong et al. |
| Gollapudi | Asian Indian | South Asian | Wong et al. |
| Goloyugo | Filipino | Other Asian | Wong et al. |
| Goltiao | Filipino | Other Asian | Wong et al. |
| Golveo | Filipino | Other Asian | Wong et al. |
| Golwala | Asian Indian | South Asian | Wong et al. |
| Gom | Chinese | Chinese | Wong et al. |
| Gombio | Filipino | Other Asian | Wong et al. |
| Gomez | Filipino | Other Asian | Wong et al. |
| Gomi | Japanese | Other Asian | Wong et al. |
| Gon | Chinese | Chinese | Wong et al. |
| Gondal |  | South Asian | Lim et al. |
| Gondhalekar | Asian Indian | South Asian | Wong et al. |
| Gondo | Japanese | Other Asian | Wong et al. |
| Gone | Asian Indian | South Asian | Wong et al. |
| Gong | Chinese | Chinese | Wong et al. |
| Gongon | Filipino | Other Asian | Wong et al. |
| Gongora | Filipino | Other Asian | Wong et al. |
| Gono | Filipino | Other Asian | Wong et al. |
| Gonong | Filipino | Other Asian | Wong et al. |
| Gonora | Filipino | Other Asian | Wong et al. |
| Gonsales | Filipino | Other Asian | Wong et al. |
| Gonsalez | Filipino | Other Asian | Wong et al. |
| Gonsalves | Asian Indian | South Asian | Wong et al. |
| Gontang | Filipino | Other Asian | Wong et al. |
| Gonzaga | Filipino | Other Asian | Wong et al. |
| Gonzales | Filipino | Other Asian | Wong et al. |
| Gonzalez | Filipino | Other Asian | Wong et al. |
| Gonzalo | Filipino | Other Asian | Wong et al. |
| Gonzalves | Filipino | Other Asian | Wong et al. |
| Gonzalvo | Filipino | Other Asian | Wong et al. |
| Goo | Chinese | Chinese | Wong et al. |
| Gooey | Chinese | Chinese | Wong et al. |
| Gook | Chinese | Chinese | Wong et al. |
| Goon | Chinese | Chinese | Wong et al. |
| Goong | Chinese | Chinese | Wong et al. |
| Gopal | Asian Indian | South Asian | Wong et al. |
| Gopalakrishn | Asian Indian | South Asian | Wong et al. |
| Gopalan | Asian Indian | South Asian | Wong et al. |
| Gopalaswamy | Asian Indian | South Asian | Wong et al. |
| Gopaldas | Asian Indian | South Asian | Wong et al. |
| Gopalji | Asian Indian | South Asian | Wong et al. |
| Gopaul | Asian Indian | South Asian | Wong et al. |
| Gopez | Filipino | Other Asian | Wong et al. |
| Gopinath | Asian Indian | South Asian | Wong et al. |
| Gopinathan | Asian Indian | South Asian | Wong et al. |
| Gopual |  | South Asian | Lim et al. |
| Goradia | Asian Indian | South Asian | Wong et al. |
| Gorawara | Asian Indian | South Asian | Wong et al. |
| Goraya | Asian Indian | South Asian | Wong et al. |
| Gordhanbhai | Asian Indian | South Asian | Wong et al. |
| Gordo | Filipino | Other Asian | Wong et al. |
| Gordoncillo | Filipino | Other Asian | Wong et al. |
| Gorgod | Filipino | Other Asian | Wong et al. |
| Gorgonio | Filipino | Other Asian | Wong et al. |
| Goro | Japanese | Other Asian | Wong et al. |
| Gorospe | Filipino | Other Asian | Wong et al. |
| Gorostiza | Filipino | Other Asian | Wong et al. |
| Goroza | Filipino | Other Asian | Wong et al. |
| Gorre | Filipino | Other Asian | Wong et al. |
| Gorrez | Filipino | Other Asian | Wong et al. |
| Gorriceta | Filipino | Other Asian | Wong et al. |
| Gorsi | Asian Indian | South Asian | Wong et al. |
| Gosai | Asian Indian | South Asian | Wong et al. |
| Gosain | Asian Indian | South Asian | Wong et al. |
| Gosal | Asian Indian | South Asian | Wong et al. |
| Gosalia | Asian Indian | South Asian | Wong et al. |
| Goseco | Filipino | Other Asian | Wong et al. |
| Goshi | Japanese | Other Asian | Wong et al. |
| Goshima | Japanese | Other Asian | Wong et al. |
| Gosiengfiao | Filipino | Other Asian | Wong et al. |
| Gosioco | Filipino | Other Asian | Wong et al. |
| Goswami | Asian Indian | South Asian | Wong et al. |
| Got | Chinese | Chinese | Wong et al. |
| Gotengco | Filipino | Other Asian | Wong et al. |
| Gotera | Filipino | Other Asian | Wong et al. |
| Gotico | Filipino | Other Asian | Wong et al. |
| Gotidoc | Filipino | Other Asian | Wong et al. |
| Gotingco | Filipino | Other Asian | Wong et al. |
| Goto | Japanese | Other Asian | Wong et al. |
| Gotoh | Japanese | Other Asian | Wong et al. |
| Gotuaco | Filipino | Other Asian | Wong et al. |
| Gou | Chinese | Chinese | Wong et al. |
| Gounder | Asian Indian | South Asian | Wong et al. |
| Govani | Asian Indian | South Asian | Wong et al. |
| Govil | Asian Indian | South Asian | Wong et al. |
| Govind | Asian Indian | South Asian | Wong et al. |
| Govindan | Asian Indian | South Asian | Wong et al. |
| Govindarajan | Asian Indian | South Asian | Wong et al. |
| Govindarajul | Asian Indian | South Asian | Wong et al. |
| Govindaswamy | Asian Indian | South Asian | Wong et al. |
| Govindbhai | Asian Indian | South Asian | Wong et al. |
| Govindji | Asian Indian | South Asian | Wong et al. |
| Gow | Chinese | Chinese | Wong et al. |
| Gowda | Asian Indian | South Asian | Wong et al. |
| Goy | Chinese | Chinese | Wong et al. |
| Goya | Japanese | Other Asian | Wong et al. |
| Goyal | Asian Indian | South Asian | Wong et al. |
| Goyena | Filipino | Other Asian | Wong et al. |
| Gozar | Filipino | Other Asian | Wong et al. |
| Goze | Filipino | Other Asian | Wong et al. |
| Gozo | Filipino | Other Asian | Wong et al. |
| Gozon | Filipino | Other Asian | Wong et al. |
| Gozos | Filipino | Other Asian | Wong et al. |
| Gozum | Filipino | Other Asian | Wong et al. |
| Gozun | Filipino | Other Asian | Wong et al. |
| Gracia | Filipino | Other Asian | Wong et al. |
| Gracias | Asian Indian | South Asian | Wong et al. |
| Gracilla | Filipino | Other Asian | Wong et al. |
| Grado | Filipino | Other Asian | Wong et al. |
| Grafil | Filipino | Other Asian | Wong et al. |
| Gragasin | Filipino | Other Asian | Wong et al. |
| Grajo | Filipino | Other Asian | Wong et al. |
| Gramaje | Filipino | Other Asian | Wong et al. |
| Gramonte | Filipino | Other Asian | Wong et al. |
| Granada | Filipino | Other Asian | Wong et al. |
| Granado | Filipino | Other Asian | Wong et al. |
| Granados | Filipino | Other Asian | Wong et al. |
| Granda | Filipino | Other Asian | Wong et al. |
| Grandea | Filipino | Other Asian | Wong et al. |
| Grandhi | Asian Indian | South Asian | Wong et al. |
| Grapa | Filipino | Other Asian | Wong et al. |
| Gravador | Filipino | Other Asian | Wong et al. |
| Graycochea | Filipino | Other Asian | Wong et al. |
| Grecia | Filipino | Other Asian | Wong et al. |
| Gregorio | Filipino | Other Asian | Wong et al. |
| Greir |  | South Asian | Lim et al. |
| Grepo | Filipino | Other Asian | Wong et al. |
| Grewal | Asian Indian | South Asian | Kranick et al. |
| Grewall | Asian Indian | South Asian | Wong et al. |
| Griarte | Filipino | Other Asian | Wong et al. |
| Griffth |  | South Asian | Lim et al. |
| Grijaldo | Filipino | Other Asian | Wong et al. |
| Grino | Filipino | Other Asian | Wong et al. |
| Gromio | Filipino | Other Asian | Wong et al. |
| Grospe | Filipino | Other Asian | Wong et al. |
| Groyon | Filipino | Other Asian | Wong et al. |
| Gruspe | Filipino | Other Asian | Wong et al. |
| Gu | Chinese | Chinese | Wong et al. |
| Guadalupe | Filipino | Other Asian | Wong et al. |
| Guadiz | Filipino | Other Asian | Wong et al. |
| Gualberto | Filipino | Other Asian | Wong et al. |
| Guan | Chinese | Chinese | Kranick et al. |
| Guanco | Filipino | Other Asian | Wong et al. |
| Guang | Chinese | Chinese | Wong et al. |
| Guanga | Filipino | Other Asian | Wong et al. |
| Guanio | Filipino | Other Asian | Wong et al. |
| Guanlao | Filipino | Other Asian | Wong et al. |
| Guansing | Filipino | Other Asian | Wong et al. |
| Guanzon | Filipino | Other Asian | Wong et al. |
| Guardiano | Filipino | Other Asian | Wong et al. |
| Guarin | Filipino | Other Asian | Wong et al. |
| Guarina | Filipino | Other Asian | Wong et al. |
| Guarnes | Filipino | Other Asian | Wong et al. |
| Guazon | Filipino | Other Asian | Wong et al. |
| Gubac | Filipino | Other Asian | Wong et al. |
| Guballa | Filipino | Other Asian | Wong et al. |
| Gubatan | Filipino | Other Asian | Wong et al. |
| Gubaton | Filipino | Other Asian | Wong et al. |
| Gubbay | Asian Indian | South Asian | Wong et al. |
| Gube | Filipino | Other Asian | Wong et al. |
| Gubisch | Filipino | Other Asian | Wong et al. |
| Guce | Filipino | Other Asian | Wong et al. |
| Guda | Filipino | Other Asian | Wong et al. |
| Gudoy | Filipino | Other Asian | Wong et al. |
| Gueco | Filipino | Other Asian | Wong et al. |
| Guen | Chinese | Chinese | Wong et al. |
| Guerpo | Filipino | Other Asian | Wong et al. |
| Guerra | Filipino | Other Asian | Wong et al. |
| Guerrero | Filipino | Other Asian | Wong et al. |
| Guerzo | Filipino | Other Asian | Wong et al. |
| Guerzon | Filipino | Other Asian | Wong et al. |
| Guese | Filipino | Other Asian | Wong et al. |
| Gueta | Filipino | Other Asian | Wong et al. |
| Guevara | Filipino | Other Asian | Wong et al. |
| Guevarra | Filipino | Other Asian | Wong et al. |
| Guey | Chinese | Chinese | Wong et al. |
| Gugol | Filipino | Other Asian | Wong et al. |
| Guh | Chinese | Chinese | Wong et al. |
| Guha | Asian Indian; Bangladeshi | South Asian | Kranick et al. |
| Guhit | Filipino | Other Asian | Wong et al. |
| Gui | Chinese | Chinese | Wong et al. |
| Guia | Filipino | Other Asian | Wong et al. |
| Guiab | Filipino | Other Asian | Wong et al. |
| Guiam | Filipino | Other Asian | Wong et al. |
| Guianan | Filipino | Other Asian | Wong et al. |
| Guiang | Filipino | Other Asian | Wong et al. |
| Guiao | Filipino | Other Asian | Wong et al. |
| Guico | Filipino | Other Asian | Wong et al. |
| Guieb | Filipino | Other Asian | Wong et al. |
| Guilaran | Filipino | Other Asian | Wong et al. |
| Guilas | Filipino | Other Asian | Wong et al. |
| Guiling | Filipino | Other Asian | Wong et al. |
| Guillem | Filipino | Other Asian | Wong et al. |
| Guillen | Filipino | Other Asian | Wong et al. |
| Guillergan | Filipino | Other Asian | Wong et al. |
| Guillermo | Filipino | Other Asian | Wong et al. |
| Guina | Filipino | Other Asian | Wong et al. |
| Guingab | Filipino | Other Asian | Wong et al. |
| Guingon | Filipino | Other Asian | Wong et al. |
| Guingona | Filipino | Other Asian | Wong et al. |
| Guino | Filipino | Other Asian | Wong et al. |
| Guinoo | Filipino | Other Asian | Wong et al. |
| Guintivano | Filipino | Other Asian | Wong et al. |
| Guinto | Filipino | Other Asian | Wong et al. |
| Guintu | Filipino | Other Asian | Wong et al. |
| Guira | Filipino | Other Asian | Wong et al. |
| Guiriba | Filipino | Other Asian | Wong et al. |
| Guirnalda | Filipino | Other Asian | Wong et al. |
| Guirnela | Filipino | Other Asian | Wong et al. |
| Guisande | Filipino | Other Asian | Wong et al. |
| Guison | Filipino | Other Asian | Wong et al. |
| Guitang | Filipino | Other Asian | Wong et al. |
| Guittap | Filipino | Other Asian | Wong et al. |
| Guittard | Filipino | Other Asian | Wong et al. |
| Guiyab | Filipino | Other Asian | Wong et al. |
| Gujar | Asian Indian | South Asian | Wong et al. |
| Gujarathi | Asian Indian | South Asian | Wong et al. |
| Gulabbhai | Asian Indian | South Asian | Wong et al. |
| Gulam | Asian Indian | South Asian | Wong et al. |
| Gulapa | Filipino | Other Asian | Wong et al. |
| Guliani | Asian Indian | South Asian | Wong et al. |
| Gulinao | Filipino | Other Asian | Wong et al. |
| Gulle | Filipino | Other Asian | Wong et al. |
| Gulmatico | Filipino | Other Asian | Wong et al. |
| Guloy | Filipino | Other Asian | Wong et al. |
| Gultiano | Filipino | Other Asian | Wong et al. |
| Gulzar |  | South Asian | Lim et al. |
| Gum | Chinese | Chinese | Kranick et al. |
| Gumabay | Filipino | Other Asian | Wong et al. |
| Gumahad | Filipino | Other Asian | Wong et al. |
| Gumangan | Filipino | Other Asian | Wong et al. |
| Gumapac | Filipino | Other Asian | Wong et al. |
| Gumapas | Filipino | Other Asian | Wong et al. |
| Gumarang | Filipino | Other Asian | Wong et al. |
| Gumatay | Filipino | Other Asian | Wong et al. |
| Gumawid | Filipino | Other Asian | Wong et al. |
| Gumayagay | Filipino | Other Asian | Wong et al. |
| Gumba | Filipino | Other Asian | Wong et al. |
| Gumban | Filipino | Other Asian | Wong et al. |
| Gumboc | Filipino | Other Asian | Wong et al. |
| Gumera | Filipino | Other Asian | Wong et al. |
| Gumila | Filipino | Other Asian | Wong et al. |
| Gumiran | Filipino | Other Asian | Wong et al. |
| Gumpal | Filipino | Other Asian | Wong et al. |
| Gumtang | Filipino | Other Asian | Wong et al. |
| Gun | Chinese | Chinese | Wong et al. |
| Gunabe | Filipino | Other Asian | Wong et al. |
| Gunda | Filipino | Other Asian | Wong et al. |
| Gundayao | Filipino | Other Asian | Wong et al. |
| Gundran | Filipino | Other Asian | Wong et al. |
| Gung | Chinese | Chinese | Wong et al. |
| Gungab | Filipino | Other Asian | Wong et al. |
| Gungon | Filipino | Other Asian | Wong et al. |
| Gungor |  | South Asian | Lim et al. |
| Gunji | Japanese | Other Asian | Wong et al. |
| Guno | Filipino | Other Asian | Wong et al. |
| Guntang | Filipino | Other Asian | Wong et al. |
| Guo | Chinese | Chinese | Kranick et al. |
| Guong | Chinese | Chinese | Wong et al. |
| Gupta | Asian Indian | South Asian | Kranick et al. |
| Gupte | Asian Indian | South Asian | Wong et al. |
| Guram | Asian Indian | South Asian | Wong et al. |
| Gurango | Filipino | Other Asian | Wong et al. |
| Gurat | Filipino | Other Asian | Wong et al. |
| Guray | Filipino | Other Asian | Wong et al. |
| Guraya | Asian Indian | South Asian | Wong et al. |
| Gurion | Filipino | Other Asian | Wong et al. |
| Gurjar | Asian Indian | South Asian | Wong et al. |
| Guron | Filipino | Other Asian | Wong et al. |
| Gurtiza | Filipino | Other Asian | Wong et al. |
| Gurung | Nepali; Asian Indian | South Asian | Kranick et al. |
| Guruswamy | Asian Indian | South Asian | Wong et al. |
| Gushi | Japanese | Other Asian | Wong et al. |
| Gushiken | Japanese | Other Asian | Wong et al. |
| Gusman | Filipino | Other Asian | Wong et al. |
| Gustilo | Filipino | Other Asian | Wong et al. |
| Gusto | Filipino | Other Asian | Wong et al. |
| Gusukuma | Japanese | Other Asian | Wong et al. |
| Gutang | Filipino | Other Asian | Wong et al. |
| Guterres | Chinese | Chinese | Wong et al. |
| Gutierez | Filipino | Other Asian | Wong et al. |
| Gutierrez | Filipino | Other Asian | Wong et al. |
| Guting | Filipino | Other Asian | Wong et al. |
| Gutlay | Filipino | Other Asian | Wong et al. |
| Guttierrez | Filipino | Other Asian | Wong et al. |
| Guu | Chinese | Chinese | Wong et al. |
| Guyen | Vietnamese | Other Asian | Wong et al. |
| Guyos | Filipino | Other Asian | Wong et al. |
| Guzdar | Asian Indian | South Asian | Wong et al. |
| Guzman | Filipino | Other Asian | Wong et al. |
| Guzon | Filipino | Other Asian | Wong et al. |
| Gwak | Korean | Other Asian | Wong et al. |
| Gwon | Chinese | Chinese | Wong et al. |
| Gwong | Chinese | Chinese | Wong et al. |
| Gyani | Asian Indian | South Asian | Wong et al. |
| Gyatso | Chinese | Chinese | Wong et al. |
| Ha | Vietnamese; Korean; Chinese | Other Asian | Kranick et al. |
| Habacon | Filipino | Other Asian | Wong et al. |
| Habal | Filipino | Other Asian | Wong et al. |
| Haban | Filipino | Other Asian | Wong et al. |
| Habana | Filipino | Other Asian | Wong et al. |
| Habaradas | Filipino | Other Asian | Wong et al. |
| Habib | Asian Indian | South Asian | Wong et al. |
| Habiba |  | South Asian | Lim et al. |
| Habibulla | Asian Indian | South Asian | Wong et al. |
| Habibullah | Asian Indian | South Asian | Wong et al. |
| Habitan | Filipino | Other Asian | Wong et al. |
| Haboc | Filipino | Other Asian | Wong et al. |
| Habon | Filipino | Other Asian | Wong et al. |
| Habu | Japanese | Other Asian | Wong et al. |
| Hac | Vietnamese | Other Asian | Wong et al. |
| Hach | Vietnamese | Other Asian | Wong et al. |
| Hachero | Filipino | Other Asian | Wong et al. |
| Hachisuka | Japanese | Other Asian | Wong et al. |
| Hachiya | Japanese | Other Asian | Wong et al. |
| Hack |  | South Asian | Lim et al. |
| Hada | Japanese | Other Asian | Wong et al. |
| Hadap | Filipino | Other Asian | Wong et al. |
| Hadi |  | South Asian | Lim et al. |
| Hadloc | Filipino | Other Asian | Wong et al. |
| Haduca | Filipino | Other Asian | Wong et al. |
| Hae | Korean | Other Asian | Wong et al. |
| Haeng | Korean | Other Asian | Wong et al. |
| Hafalla | Filipino | Other Asian | Wong et al. |
| Hafeez | Pakistani | South Asian | Kranick et al. |
| Hafiz | Asian Indian | South Asian | Wong et al. |
| Hag | Asian Indian | South Asian | Wong et al. |
| Hagi | Japanese | Other Asian | Wong et al. |
| Hagihara | Japanese | Other Asian | Wong et al. |
| Hagino | Japanese | Other Asian | Wong et al. |
| Hagio | Japanese | Other Asian | Wong et al. |
| Hagiwara | Japanese | Other Asian | Wong et al. |
| Hagos | Filipino | Other Asian | Wong et al. |
| Haguisan | Filipino | Other Asian | Wong et al. |
| Hah | Korean | Other Asian | Wong et al. |
| Hahm | Korean | Other Asian | Wong et al. |
| Hahn | Korean | Other Asian | Kranick et al. |
| Hai | Chinese | Chinese | Wong et al. |
| Haidari | Asian Indian | South Asian | Wong et al. |
| Haider | Pakistani; Bangladeshi | South Asian | Kranick et al. |
| Haidermota | Asian Indian | South Asian | Wong et al. |
| Haidri | Asian Indian | South Asian | Wong et al. |
| Haing | Chinese | Chinese | Wong et al. |
| Hairman |  | South Asian | Lim et al. |
| Hajari | Asian Indian | South Asian | Wong et al. |
| Hajela | Asian Indian | South Asian | Wong et al. |
| Hajime | Japanese | Other Asian | Wong et al. |
| Hak | Korean | Other Asian | Wong et al. |
| Hakeem | Asian Indian | South Asian | Wong et al. |
| Hakim | Asian Indian | South Asian | Wong et al. |
| Hakomori | Japanese | Other Asian | Wong et al. |
| Hakozaki | Japanese | Other Asian | Wong et al. |
| Halago | Filipino | Other Asian | Wong et al. |
| Halasan | Filipino | Other Asian | Wong et al. |
| Halcon | Filipino | Other Asian | Wong et al. |
| Haldar | Asian Indian | South Asian | Wong et al. |
| Halder |  | South Asian | Lim et al. |
| Haleem | Asian Indian | South Asian | Wong et al. |
| Halili | Filipino | Other Asian | Wong et al. |
| Halim |  | South Asian | Lim et al. |
| Hallal |  | South Asian | Lim et al. |
| Hallare | Filipino | Other Asian | Wong et al. |
| Hallera | Filipino | Other Asian | Wong et al. |
| Halog | Filipino | Other Asian | Wong et al. |
| Ham | Korean | Other Asian | Kranick et al. |
| Hama | Japanese | Other Asian | Wong et al. |
| Hamabata | Japanese | Other Asian | Wong et al. |
| Hamabe | Japanese | Other Asian | Wong et al. |
| Hamachi | Japanese | Other Asian | Wong et al. |
| Hamada | Japanese | Other Asian | Wong et al. |
| Hamaguchi | Japanese | Other Asian | Wong et al. |
| Hamai | Japanese | Other Asian | Wong et al. |
| Hamaji | Japanese | Other Asian | Wong et al. |
| Hamakawa | Japanese | Other Asian | Wong et al. |
| Hamamoto | Japanese | Other Asian | Wong et al. |
| Hamamura | Japanese | Other Asian | Wong et al. |
| Hamanaka | Japanese | Other Asian | Wong et al. |
| Hamane | Japanese | Other Asian | Wong et al. |
| Hamano | Japanese | Other Asian | Wong et al. |
| Hamao | Japanese | Other Asian | Wong et al. |
| Hamaoka | Japanese | Other Asian | Wong et al. |
| Hamasaki | Japanese | Other Asian | Wong et al. |
| Hamashima | Japanese | Other Asian | Wong et al. |
| Hamaya | Japanese | Other Asian | Wong et al. |
| Hamdani | Asian Indian | South Asian | Wong et al. |
| Hamdulay |  | South Asian | Lim et al. |
| Hamed | Asian Indian | South Asian | Wong et al. |
| Hameed | Asian Indian | South Asian | Wong et al. |
| Hamid |  | South Asian | Lim et al. |
| Hamidani | Asian Indian | South Asian | Wong et al. |
| Hamirani | Asian Indian | South Asian | Wong et al. |
| Hamoy | Filipino | Other Asian | Wong et al. |
| Han | Chinese; Korean | Chinese | Kranick et al. |
| Hana |  | South Asian | Lim et al. |
| Hanabusa | Japanese | Other Asian | Wong et al. |
| Hanada | Japanese | Other Asian | Wong et al. |
| Hanafusa | Japanese | Other Asian | Wong et al. |
| Hanagata | Japanese | Other Asian | Wong et al. |
| Hanai | Japanese | Other Asian | Wong et al. |
| Hanamoto | Japanese | Other Asian | Wong et al. |
| Hanamura | Japanese | Other Asian | Wong et al. |
| Hanano | Japanese | Other Asian | Wong et al. |
| Hanaoka | Japanese | Other Asian | Wong et al. |
| Hanashiro | Japanese | Other Asian | Wong et al. |
| Hanawa | Japanese | Other Asian | Wong et al. |
| Hanazawa | Japanese | Other Asian | Wong et al. |
| Handa |  | South Asian | Lim et al. |
| Handog | Filipino | Other Asian | Wong et al. |
| Haneda | Japanese | Other Asian | Wong et al. |
| Haneishi | Japanese | Other Asian | Wong et al. |
| Hangad | Filipino | Other Asian | Wong et al. |
| Hanh | Vietnamese | Other Asian | Wong et al. |
| Hanif |  | South Asian | Lim et al. |
| Haniff |  | South Asian | Lim et al. |
| Hannan |  | South Asian | Lim et al. |
| Hano | Japanese | Other Asian | Wong et al. |
| Hansalia | Asian Indian | South Asian | Wong et al. |
| Hansotia | Asian Indian | South Asian | Wong et al. |
| Hansra | Asian Indian | South Asian | Wong et al. |
| Hansraj |  | South Asian | Lim et al. |
| Hanumanthapp | Asian Indian | South Asian | Wong et al. |
| Hanyu | Japanese | Other Asian | Wong et al. |
| Hanzawa | Japanese | Other Asian | Wong et al. |
| Hao | Chinese | Chinese | Wong et al. |
| Haong | Vietnamese | Other Asian | Wong et al. |
| Hapin | Filipino | Other Asian | Wong et al. |
| Hapitan | Filipino | Other Asian | Wong et al. |
| Haq | Bangladeshi; Pakistani | South Asian | Kranick et al. |
| Haqqi | Asian Indian | South Asian | Wong et al. |
| Haque | Bangladeshi | South Asian | Kranick et al. |
| Har | Chinese | Chinese | Wong et al. |
| Hara | Japanese | Other Asian | Wong et al. |
| Harada | Japanese | Other Asian | Kranick et al. |
| Haraguchi | Japanese | Other Asian | Wong et al. |
| Harai | Japanese | Other Asian | Wong et al. |
| Harakawa | Japanese | Other Asian | Wong et al. |
| Harano | Japanese | Other Asian | Wong et al. |
| Harashima | Japanese | Other Asian | Wong et al. |
| Harayama | Japanese | Other Asian | Wong et al. |
| Harbhajan |  | South Asian | Lim et al. |
| Hardikar | Asian Indian | South Asian | Wong et al. |
| Haresco | Filipino | Other Asian | Wong et al. |
| Hari | Asian Indian | South Asian | Wong et al. |
| Haribhai | Asian Indian | South Asian | Wong et al. |
| Hariharan | Asian Indian | South Asian | Wong et al. |
| Harilal | Asian Indian | South Asian | Wong et al. |
| Harima | Japanese | Other Asian | Wong et al. |
| Harimoto | Japanese | Other Asian | Wong et al. |
| Harinarine |  | South Asian | Lim et al. |
| Haripersaud |  | South Asian | Lim et al. |
| Harnandan |  | South Asian | Lim et al. |
| Haro | Filipino | Other Asian | Wong et al. |
| Haroon | Asian Indian | South Asian | Wong et al. |
| Harpaul |  | South Asian | Lim et al. |
| Harricharan |  | South Asian | Lim et al. |
| Harrilal |  | South Asian | Lim et al. |
| Harrimangal |  | South Asian | Lim et al. |
| Harrycharan |  | South Asian | Lim et al. |
| Harrysingh |  | South Asian | Lim et al. |
| Haruki | Japanese | Other Asian | Wong et al. |
| Harun |  | South Asian | Lim et al. |
| Haruna | Japanese | Other Asian | Wong et al. |
| Haruta | Japanese | Other Asian | Wong et al. |
| Haruyama | Japanese | Other Asian | Wong et al. |
| Hasagawa | Japanese | Other Asian | Wong et al. |
| Hasan | Bangladeshi | South Asian | Kranick et al. |
| Hasanali | Asian Indian | South Asian | Wong et al. |
| Hasebe | Japanese | Other Asian | Wong et al. |
| Hasegawa | Japanese | Other Asian | Wong et al. |
| Hasham | Asian Indian | South Asian | Wong et al. |
| Hasheem |  | South Asian | Lim et al. |
| Hashem |  | South Asian | Lim et al. |
| Hashi | Japanese | Other Asian | Wong et al. |
| Hashiba | Japanese | Other Asian | Wong et al. |
| Hashida | Japanese | Other Asian | Wong et al. |
| Hashigami | Japanese | Other Asian | Wong et al. |
| Hashiguchi | Japanese | Other Asian | Wong et al. |
| Hashim | Malaysian | Other Asian | Kranick et al. |
| Hashimi | Asian Indian | South Asian | Wong et al. |
| Hashimoto | Japanese | Other Asian | Kranick et al. |
| Hashimura | Japanese | Other Asian | Wong et al. |
| Hashizume | Japanese | Other Asian | Wong et al. |
| Hashmi | Asian Indian | South Asian | Wong et al. |
| Hashmy | Asian Indian | South Asian | Wong et al. |
| Hasija | Asian Indian | South Asian | Wong et al. |
| Hasildar |  | South Asian | Lim et al. |
| Hasim | Asian Indian | South Asian | Wong et al. |
| Hasin |  | South Asian | Lim et al. |
| Hasnain | Asian Indian | South Asian | Wong et al. |
| Hasni | Asian Indian | South Asian | Wong et al. |
| Hassain |  | South Asian | Lim et al. |
| Hassam | Asian Indian | South Asian | Wong et al. |
| Hassan | Bangladeshi | South Asian | Kranick et al. |
| Hassanali | Asian Indian | South Asian | Wong et al. |
| Hassanally | Asian Indian | South Asian | Wong et al. |
| Hassim |  | South Asian | Lim et al. |
| Hasumi | Japanese | Other Asian | Wong et al. |
| Hasunuma | Japanese | Other Asian | Wong et al. |
| Hasuo | Japanese | Other Asian | Wong et al. |
| Hat | Vietnamese | Other Asian | Wong et al. |
| Hata | Japanese | Other Asian | Wong et al. |
| Hatada | Japanese | Other Asian | Wong et al. |
| Hatae | Japanese | Other Asian | Wong et al. |
| Hatakeyama | Japanese | Other Asian | Wong et al. |
| Hatanaka | Japanese | Other Asian | Wong et al. |
| Hatano | Japanese | Other Asian | Wong et al. |
| Hatashita | Japanese | Other Asian | Wong et al. |
| Hatayama | Japanese | Other Asian | Wong et al. |
| Hatia | Asian Indian | South Asian | Wong et al. |
| Hatol | Filipino | Other Asian | Wong et al. |
| Hatori | Japanese | Other Asian | Wong et al. |
| Hatsushi | Japanese | Other Asian | Wong et al. |
| Hatta | Japanese | Other Asian | Wong et al. |
| Hattori | Japanese | Other Asian | Wong et al. |
| Hau | Vietnamese | Other Asian | Kranick et al. |
| Haung | Chinese | Chinese | Wong et al. |
| Hautea | Filipino | Other Asian | Wong et al. |
| Haw | Chinese | Chinese | Wong et al. |
| Haya | Japanese | Other Asian | Wong et al. |
| Hayag | Filipino | Other Asian | Wong et al. |
| Hayakawa | Japanese | Other Asian | Wong et al. |
| Hayama | Japanese | Other Asian | Wong et al. |
| Hayami | Japanese | Other Asian | Wong et al. |
| Hayano | Japanese | Other Asian | Wong et al. |
| Hayasaka | Japanese | Other Asian | Wong et al. |
| Hayasaki | Japanese | Other Asian | Wong et al. |
| Hayase | Japanese | Other Asian | Wong et al. |
| Hayashi | Japanese | Other Asian | Wong et al. |
| Hayashida | Japanese | Other Asian | Wong et al. |
| Hayasi | Japanese | Other Asian | Wong et al. |
| Hayata | Japanese | Other Asian | Wong et al. |
| Hayath | Asian Indian | South Asian | Wong et al. |
| Haydar |  | South Asian | Lim et al. |
| Hayder |  | South Asian | Lim et al. |
| Hazama | Japanese | Other Asian | Wong et al. |
| Hazari | Asian Indian | South Asian | Wong et al. |
| Hazarika | Asian Indian | South Asian | Wong et al. |
| Hazi |  | South Asian | Lim et al. |
| Hazra | Asian Indian | South Asian | Wong et al. |
| He | Chinese | Chinese | Kranick et al. |
| Hea | Korean | Other Asian | Wong et al. |
| Hebbar | Asian Indian | South Asian | Wong et al. |
| Hebreo | Filipino | Other Asian | Wong et al. |
| Hechanova | Filipino | Other Asian | Wong et al. |
| Hee | Chinese | Chinese | Wong et al. |
| Heen | Chinese | Chinese | Wong et al. |
| Heeralall |  | South Asian | Lim et al. |
| Heeraman |  | South Asian | Lim et al. |
| Hegde | Asian Indian | South Asian | Wong et al. |
| Heh | Chinese | Chinese | Wong et al. |
| Heir | Asian Indian | South Asian | Wong et al. |
| Helali |  | South Asian | Lim et al. |
| Hemani | Asian Indian | South Asian | Wong et al. |
| Hembrador | Filipino | Other Asian | Wong et al. |
| Hembrom | Asian Indian | South Asian | Wong et al. |
| Hemnani | Asian Indian | South Asian | Wong et al. |
| Hemnauth |  | South Asian | Lim et al. |
| Hen | Chinese | Chinese | Wong et al. |
| Hena |  | South Asian | Lim et al. |
| Henares | Filipino | Other Asian | Wong et al. |
| Henmi | Japanese | Other Asian | Wong et al. |
| Heo | Korean | Other Asian | Wong et al. |
| Heong | Chinese | Chinese | Wong et al. |
| Hep | Chinese | Chinese | Wong et al. |
| Heraldo | Filipino | Other Asian | Wong et al. |
| Heramia | Filipino | Other Asian | Wong et al. |
| Herana | Filipino | Other Asian | Wong et al. |
| Heredia | Filipino | Other Asian | Wong et al. |
| Heres | Filipino | Other Asian | Wong et al. |
| Herico | Filipino | Other Asian | Wong et al. |
| Hermano | Filipino | Other Asian | Wong et al. |
| Hermida | Filipino | Other Asian | Wong et al. |
| Hermogenes | Filipino | Other Asian | Wong et al. |
| Hermogeno | Filipino | Other Asian | Wong et al. |
| Hermogino | Filipino | Other Asian | Wong et al. |
| Hermosa | Filipino | Other Asian | Wong et al. |
| Hermosilla | Filipino | Other Asian | Wong et al. |
| Hermoso | Filipino | Other Asian | Wong et al. |
| Hermosura | Filipino | Other Asian | Wong et al. |
| Hernaez | Filipino | Other Asian | Wong et al. |
| Hernandes | Filipino | Other Asian | Wong et al. |
| Hernandez |  | South Asian | Lim et al. |
| Hernando | Filipino | Other Asian | Wong et al. |
| Herradura | Filipino | Other Asian | Wong et al. |
| Herras | Filipino | Other Asian | Wong et al. |
| Herrera | Filipino | Other Asian | Wong et al. |
| Herreria | Filipino | Other Asian | Wong et al. |
| Herrero | Filipino | Other Asian | Wong et al. |
| Hertez | Filipino | Other Asian | Wong et al. |
| Heruela | Filipino | Other Asian | Wong et al. |
| Hervas | Filipino | Other Asian | Wong et al. |
| Hervias | Filipino | Other Asian | Wong et al. |
| Heshiki | Japanese | Other Asian | Wong et al. |
| Heung | Chinese | Chinese | Wong et al. |
| Hew | Chinese | Chinese | Wong et al. |
| Hg | Chinese | Chinese | Wong et al. |
| Hgo | Vietnamese | Other Asian | Wong et al. |
| Hguyen | Vietnamese | Other Asian | Wong et al. |
| Hibi | Japanese | Other Asian | Wong et al. |
| Hibino | Japanese | Other Asian | Wong et al. |
| Hibionada | Filipino | Other Asian | Wong et al. |
| Hicarte | Filipino | Other Asian | Wong et al. |
| Hicban | Filipino | Other Asian | Wong et al. |
| Hida | Japanese | Other Asian | Wong et al. |
| Hidai | Japanese | Other Asian | Wong et al. |
| Hidaka | Japanese | Other Asian | Wong et al. |
| Hidalgo | Filipino | Other Asian | Wong et al. |
| Hidano | Japanese | Other Asian | Wong et al. |
| Hideo | Japanese | Other Asian | Wong et al. |
| Hideshima | Japanese | Other Asian | Wong et al. |
| Hidrosollo | Filipino | Other Asian | Wong et al. |
| Hieda | Japanese | Other Asian | Wong et al. |
| Hien | Vietnamese | Other Asian | Wong et al. |
| Hiep | Vietnamese | Other Asian | Wong et al. |
| Hieu | Vietnamese | Other Asian | Wong et al. |
| Higa | Japanese | Other Asian | Wong et al. |
| Higaki | Japanese | Other Asian | Wong et al. |
| Higashi | Japanese | Other Asian | Wong et al. |
| Higashida | Japanese | Other Asian | Wong et al. |
| Higashijima | Japanese | Other Asian | Wong et al. |
| Higashikawa | Japanese | Other Asian | Wong et al. |
| Higashimura | Japanese | Other Asian | Wong et al. |
| Higashino | Japanese | Other Asian | Wong et al. |
| Higashionna | Japanese | Other Asian | Wong et al. |
| Higashitani | Japanese | Other Asian | Wong et al. |
| Higashiyama | Japanese | Other Asian | Wong et al. |
| Higo | Japanese | Other Asian | Wong et al. |
| Higuchi | Japanese | Other Asian | Wong et al. |
| Higuma | Japanese | Other Asian | Wong et al. |
| Higurashi | Japanese | Other Asian | Wong et al. |
| Hijikata | Japanese | Other Asian | Wong et al. |
| Hiki | Japanese | Other Asian | Wong et al. |
| Hikichi | Japanese | Other Asian | Wong et al. |
| Hikida | Japanese | Other Asian | Wong et al. |
| Hikita | Japanese | Other Asian | Wong et al. |
| Hikosaka | Japanese | Other Asian | Wong et al. |
| Hilado | Filipino | Other Asian | Wong et al. |
| Hilao | Filipino | Other Asian | Wong et al. |
| Hilario | Filipino | Other Asian | Wong et al. |
| Hilo | Filipino | Other Asian | Wong et al. |
| Hilvano | Filipino | Other Asian | Wong et al. |
| Him | Chinese | Chinese | Wong et al. |
| Himan | Filipino | Other Asian | Wong et al. |
| Himatay | Filipino | Other Asian | Wong et al. |
| Himatlal | Asian Indian | South Asian | Wong et al. |
| Himeno | Japanese | Other Asian | Wong et al. |
| Hin | Chinese | Chinese | Wong et al. |
| Hinahon | Filipino | Other Asian | Wong et al. |
| Hinajon | Filipino | Other Asian | Wong et al. |
| Hinata | Japanese | Other Asian | Wong et al. |
| Hinaut | Filipino | Other Asian | Wong et al. |
| Hing | Cambodian | Other Asian | Kranick et al. |
| Hingada | Filipino | Other Asian | Wong et al. |
| Hinh | Vietnamese | Other Asian | Wong et al. |
| Hinlo | Filipino | Other Asian | Wong et al. |
| Hino | Japanese | Other Asian | Wong et al. |
| Hinola | Filipino | Other Asian | Wong et al. |
| Hinolan | Filipino | Other Asian | Wong et al. |
| Hioki | Japanese | Other Asian | Wong et al. |
| Hiong | Chinese | Chinese | Wong et al. |
| Hip | Chinese | Chinese | Wong et al. |
| Hipe | Filipino | Other Asian | Wong et al. |
| Hipol | Filipino | Other Asian | Wong et al. |
| Hipolito | Filipino | Other Asian | Wong et al. |
| Hipona | Filipino | Other Asian | Wong et al. |
| Hiquiana | Filipino | Other Asian | Wong et al. |
| Hirabayashi | Japanese | Other Asian | Wong et al. |
| Hiraga | Japanese | Other Asian | Wong et al. |
| Hirahara | Japanese | Other Asian | Wong et al. |
| Hirai | Japanese | Other Asian | Wong et al. |
| Hiraide | Japanese | Other Asian | Wong et al. |
| Hiraishi | Japanese | Other Asian | Wong et al. |
| Hiraiwa | Japanese | Other Asian | Wong et al. |
| Hirakata | Japanese | Other Asian | Wong et al. |
| Hirakawa | Japanese | Other Asian | Wong et al. |
| Hiraki | Japanese | Other Asian | Wong et al. |
| Hirako | Japanese | Other Asian | Wong et al. |
| Hiralal | Asian Indian | South Asian | Wong et al. |
| Hirama | Japanese | Other Asian | Wong et al. |
| Hiramatsu | Japanese | Other Asian | Wong et al. |
| Hiramine | Japanese | Other Asian | Wong et al. |
| Hiramitsu | Japanese | Other Asian | Wong et al. |
| Hiramoto | Japanese | Other Asian | Wong et al. |
| Hirang | Filipino | Other Asian | Wong et al. |
| Hirani | Asian Indian | South Asian | Wong et al. |
| Hirano | Japanese | Other Asian | Wong et al. |
| Hiranuma | Japanese | Other Asian | Wong et al. |
| Hirao | Japanese | Other Asian | Wong et al. |
| Hiraoka | Japanese | Other Asian | Wong et al. |
| Hirasa | Japanese | Other Asian | Wong et al. |
| Hirasawa | Japanese | Other Asian | Wong et al. |
| Hirase | Japanese | Other Asian | Wong et al. |
| Hirashima | Japanese | Other Asian | Wong et al. |
| Hirata | Japanese | Other Asian | Wong et al. |
| Hiratani | Japanese | Other Asian | Wong et al. |
| Hirato | Japanese | Other Asian | Wong et al. |
| Hiratsuka | Japanese | Other Asian | Wong et al. |
| Hirayama | Japanese | Other Asian | Wong et al. |
| Hiremath | Asian Indian | South Asian | Wong et al. |
| Hirji | Asian Indian | South Asian | Wong et al. |
| Hiro | Japanese | Other Asian | Wong et al. |
| Hirobe | Japanese | Other Asian | Wong et al. |
| Hiroe | Japanese | Other Asian | Wong et al. |
| Hirohashi | Japanese | Other Asian | Wong et al. |
| Hirohata | Japanese | Other Asian | Wong et al. |
| Hiroi | Japanese | Other Asian | Wong et al. |
| Hirokawa | Japanese | Other Asian | Wong et al. |
| Hiroki | Japanese | Other Asian | Wong et al. |
| Hiromoto | Japanese | Other Asian | Wong et al. |
| Hironaka | Japanese | Other Asian | Wong et al. |
| Hirono | Japanese | Other Asian | Wong et al. |
| Hiroo | Japanese | Other Asian | Wong et al. |
| Hirooka | Japanese | Other Asian | Wong et al. |
| Hirosawa | Japanese | Other Asian | Wong et al. |
| Hirose | Japanese | Other Asian | Wong et al. |
| Hiroshi | Japanese | Other Asian | Wong et al. |
| Hiroshige | Japanese | Other Asian | Wong et al. |
| Hiroshima | Japanese | Other Asian | Wong et al. |
| Hirota | Japanese | Other Asian | Wong et al. |
| Hirotsu | Japanese | Other Asian | Wong et al. |
| Hirpara | Asian Indian | South Asian | Wong et al. |
| Hiruta | Japanese | Other Asian | Wong et al. |
| Hisada | Japanese | Other Asian | Wong et al. |
| Hisamatsu | Japanese | Other Asian | Wong et al. |
| Hisamoto | Japanese | Other Asian | Wong et al. |
| Hisanaga | Japanese | Other Asian | Wong et al. |
| Hisano | Japanese | Other Asian | Wong et al. |
| Hisao | Japanese | Other Asian | Wong et al. |
| Hisatomi | Japanese | Other Asian | Wong et al. |
| Hisatsune | Japanese | Other Asian | Wong et al. |
| Hisayama | Japanese | Other Asian | Wong et al. |
| Hishida | Japanese | Other Asian | Wong et al. |
| Hishikawa | Japanese | Other Asian | Wong et al. |
| Hishinuma | Japanese | Other Asian | Wong et al. |
| Hishiyama | Japanese | Other Asian | Wong et al. |
| Hitomi | Japanese | Other Asian | Wong et al. |
| Hiu | Chinese | Chinese | Wong et al. |
| Hiura | Japanese | Other Asian | Wong et al. |
| Hiwale | Asian Indian | South Asian | Wong et al. |
| Hiwatashi | Japanese | Other Asian | Wong et al. |
| Hiyama | Japanese | Other Asian | Wong et al. |
| Hiyane | Japanese | Other Asian | Wong et al. |
| Hiyao | Filipino | Other Asian | Wong et al. |
| Hiyoshi | Japanese | Other Asian | Wong et al. |
| Hizon | Filipino | Other Asian | Wong et al. |
| Ho | Chinese; Vietnamese | Chinese | Kranick et al. |
| Hoa | Vietnamese | Other Asian | Wong et al. |
| Hoac | Vietnamese | Other Asian | Wong et al. |
| Hoach | Vietnamese | Other Asian | Wong et al. |
| Hoag | Vietnamese | Other Asian | Wong et al. |
| Hoai | Vietnamese | Other Asian | Wong et al. |
| Hoan | Vietnamese | Other Asian | Wong et al. |
| Hoang | Vietnamese | Other Asian | Wong et al. |
| Hoanh | Vietnamese | Other Asian | Wong et al. |
| Hoashi | Japanese | Other Asian | Wong et al. |
| Hoat | Vietnamese | Other Asian | Wong et al. |
| Hoc | Vietnamese | Other Asian | Wong et al. |
| Hocson | Filipino | Other Asian | Wong et al. |
| Hoe | Malaysian | Other Asian | Kranick et al. |
| Hofilena | Filipino | Other Asian | Wong et al. |
| Hoh | Chinese | Chinese | Wong et al. |
| Hoi | Chinese | Chinese | Wong et al. |
| Hojo | Japanese | Other Asian | Wong et al. |
| Hokama | Japanese | Other Asian | Wong et al. |
| Hokari | Japanese | Other Asian | Wong et al. |
| Holanday | Filipino | Other Asian | Wong et al. |
| Holgado | Filipino | Other Asian | Wong et al. |
| Holganza | Filipino | Other Asian | Wong et al. |
| Hollero | Filipino | Other Asian | Wong et al. |
| Hom | Burmese; Chinese | Other Asian | Kranick et al. |
| Homayra |  | South Asian | Lim et al. |
| Hombrebueno | Filipino | Other Asian | Wong et al. |
| Homeres | Filipino | Other Asian | Wong et al. |
| Homma | Japanese | Other Asian | Wong et al. |
| Hon | Chinese | Chinese | Wong et al. |
| Honculada | Filipino | Other Asian | Wong et al. |
| Honda | Japanese | Other Asian | Wong et al. |
| Hondo | Filipino | Other Asian | Wong et al. |
| Hong | Chinese; Vietnamese; Korean | Chinese | Kranick et al. |
| Hongo | Japanese | Other Asian | Wong et al. |
| Honjo | Japanese | Other Asian | Wong et al. |
| Honma | Japanese | Other Asian | Wong et al. |
| Honniball | Chinese | Chinese | Wong et al. |
| Honorio | Filipino | Other Asian | Wong et al. |
| Honra | Filipino | Other Asian | Wong et al. |
| Honrada | Filipino | Other Asian | Wong et al. |
| Honrado | Filipino | Other Asian | Wong et al. |
| Hontiveros | Filipino | Other Asian | Wong et al. |
| Hoo | Chinese | Chinese | Wong et al. |
| Hooda | Asian Indian | South Asian | Wong et al. |
| Hooi | Chinese | Chinese | Wong et al. |
| Hoosein |  | South Asian | Lim et al. |
| Hop | Chinese | Chinese | Wong et al. |
| Hoq |  | South Asian | Lim et al. |
| Hoque | Bangladeshi; Asian Indian | South Asian | Kranick et al. |
| Hor | Chinese | Chinese | Wong et al. |
| Horario | Filipino | Other Asian | Wong et al. |
| Horca | Filipino | Other Asian | Wong et al. |
| Hore |  | South Asian | Lim et al. |
| Hori | Japanese | Other Asian | Wong et al. |
| Horiba | Japanese | Other Asian | Wong et al. |
| Horibata | Japanese | Other Asian | Wong et al. |
| Horibe | Japanese | Other Asian | Wong et al. |
| Horie | Japanese | Other Asian | Wong et al. |
| Horiguchi | Japanese | Other Asian | Wong et al. |
| Horii | Japanese | Other Asian | Wong et al. |
| Horiike | Japanese | Other Asian | Wong et al. |
| Horikawa | Japanese | Other Asian | Wong et al. |
| Horiki | Japanese | Other Asian | Wong et al. |
| Horikiri | Japanese | Other Asian | Wong et al. |
| Horikoshi | Japanese | Other Asian | Wong et al. |
| Horimoto | Japanese | Other Asian | Wong et al. |
| Horino | Japanese | Other Asian | Wong et al. |
| Horinouchi | Japanese | Other Asian | Wong et al. |
| Horio | Japanese | Other Asian | Wong et al. |
| Horita | Japanese | Other Asian | Wong et al. |
| Horiuchi | Japanese | Other Asian | Wong et al. |
| Hormillosa | Filipino | Other Asian | Wong et al. |
| Hormitsang |  | South Asian | Lim et al. |
| Horng | Chinese | Chinese | Wong et al. |
| Hornilla | Filipino | Other Asian | Wong et al. |
| Hortaleza | Filipino | Other Asian | Wong et al. |
| Hortelano | Filipino | Other Asian | Wong et al. |
| Hortillosa | Filipino | Other Asian | Wong et al. |
| Hortinela | Filipino | Other Asian | Wong et al. |
| Hortizuela | Filipino | Other Asian | Wong et al. |
| Hosain | Asian Indian | South Asian | Wong et al. |
| Hosaka | Japanese | Other Asian | Wong et al. |
| Hosan |  | South Asian | Lim et al. |
| Hosein |  | South Asian | Lim et al. |
| Hoshi | Japanese | Other Asian | Wong et al. |
| Hoshida | Japanese | Other Asian | Wong et al. |
| Hoshika | Japanese | Other Asian | Wong et al. |
| Hoshikawa | Japanese | Other Asian | Wong et al. |
| Hoshiko | Japanese | Other Asian | Wong et al. |
| Hoshina | Japanese | Other Asian | Wong et al. |
| Hoshino | Japanese | Other Asian | Wong et al. |
| Hoshiyama | Japanese | Other Asian | Wong et al. |
| Hoshizaki | Japanese | Other Asian | Wong et al. |
| Hosobuchi | Japanese | Other Asian | Wong et al. |
| Hosoda | Japanese | Other Asian | Wong et al. |
| Hosoe | Japanese | Other Asian | Wong et al. |
| Hosoi | Japanese | Other Asian | Wong et al. |
| Hosokawa | Japanese | Other Asian | Wong et al. |
| Hosoki | Japanese | Other Asian | Wong et al. |
| Hosomi | Japanese | Other Asian | Wong et al. |
| Hosono | Japanese | Other Asian | Wong et al. |
| Hosotani | Japanese | Other Asian | Wong et al. |
| Hosoya | Japanese | Other Asian | Wong et al. |
| Hossain | Bangladeshi | South Asian | Kranick et al. |
| Hossan |  | South Asian | Lim et al. |
| Hossein | Bangladeshi | South Asian | Kranick et al. |
| Hossen |  | South Asian | Lim et al. |
| Hossin |  | South Asian | Lim et al. |
| Hossion |  | South Asian | Lim et al. |
| Hothi | Asian Indian | South Asian | Wong et al. |
| Hotta | Japanese | Other Asian | Wong et al. |
| Hotung | Chinese | Chinese | Wong et al. |
| Hou | Chinese | Chinese | Kranick et al. |
| Houang | Vietnamese | Other Asian | Wong et al. |
| Houng | Chinese | Chinese | Wong et al. |
| How | Chinese | Chinese | Wong et al. |
| Howladar |  | South Asian | Lim et al. |
| Howlader |  | South Asian | Lim et al. |
| Hoyla | Filipino | Other Asian | Wong et al. |
| Hozumi | Japanese | Other Asian | Wong et al. |
| Hsa | Chinese | Chinese | Wong et al. |
| Hsang | Chinese | Chinese | Wong et al. |
| Hse | Chinese | Chinese | Wong et al. |
| Hsei | Chinese | Chinese | Wong et al. |
| Hsi | Chinese | Chinese | Wong et al. |
| Hsia | Chinese | Chinese | Kranick et al. |
| Hsiad | Chinese | Chinese | Wong et al. |
| Hsian | Chinese | Chinese | Wong et al. |
| Hsiang | Chinese | Chinese | Wong et al. |
| Hsiao | Chinese | Chinese | Kranick et al. |
| Hsich | Chinese | Chinese | Wong et al. |
| Hsie | Chinese | Chinese | Wong et al. |
| Hsieh | Chinese | Chinese | Kranick et al. |
| Hsien | Chinese | Chinese | Wong et al. |
| Hsigh | Chinese | Chinese | Wong et al. |
| Hsih | Chinese | Chinese | Wong et al. |
| Hsin | Chinese | Chinese | Wong et al. |
| Hsing | Chinese | Chinese | Wong et al. |
| Hsiu | Chinese | Chinese | Wong et al. |
| Hsiun | Chinese | Chinese | Wong et al. |
| Hsiung | Chinese | Chinese | Wong et al. |
| Hsu | Chinese | Chinese | Kranick et al. |
| Hsuan | Chinese | Chinese | Wong et al. |
| Hsue | Chinese | Chinese | Wong et al. |
| Hsueh | Chinese | Chinese | Wong et al. |
| Hsuen | Chinese | Chinese | Wong et al. |
| Hsui | Chinese | Chinese | Wong et al. |
| Hsun | Chinese | Chinese | Wong et al. |
| Hsung | Chinese | Chinese | Wong et al. |
| Hu | Chinese | Chinese | Kranick et al. |
| Hua | Chinese | Chinese | Kranick et al. |
| Huai | Chinese | Chinese | Wong et al. |
| Huan | Chinese | Chinese | Wong et al. |
| Huana | Chinese | Chinese | Wong et al. |
| Huang | Chinese | Chinese | Kranick et al. |
| Hubahib | Filipino | Other Asian | Wong et al. |
| Hubayan | Filipino | Other Asian | Wong et al. |
| Hubilla | Filipino | Other Asian | Wong et al. |
| Huda | Asian Indian | South Asian | Wong et al. |
| Huei | Chinese | Chinese | Wong et al. |
| Huelar | Filipino | Other Asian | Wong et al. |
| Huen | Chinese | Chinese | Wong et al. |
| Hueng | Chinese | Chinese | Wong et al. |
| Huerta | Filipino | Other Asian | Wong et al. |
| Huertas | Filipino | Other Asian | Wong et al. |
| Huerto | Filipino | Other Asian | Wong et al. |
| Huey | Chinese | Chinese | Wong et al. |
| Hufalar | Filipino | Other Asian | Wong et al. |
| Hufana | Filipino | Other Asian | Wong et al. |
| Hufano | Filipino | Other Asian | Wong et al. |
| Hugh | Chinese | Chinese | Wong et al. |
| Huh | Korean | Other Asian | Kranick et al. |
| Hui | Chinese | Chinese | Kranick et al. |
| Huibonhoa | Vietnamese | Other Asian | Wong et al. |
| Huie | Chinese | Chinese | Wong et al. |
| Hulguin | Filipino | Other Asian | Wong et al. |
| Huliganga | Filipino | Other Asian | Wong et al. |
| Hullana | Filipino | Other Asian | Wong et al. |
| Hulog | Filipino | Other Asian | Wong et al. |
| Hum | Cambodian | Other Asian | Kranick et al. |
| Humayun | Pakistani; Bangladeshi | South Asian | Kranick et al. |
| Hunag | Chinese | Chinese | Wong et al. |
| Hundal | Asian Indian | South Asian | Wong et al. |
| Hung | Chinese; Vietnamese | Chinese | Kranick et al. |
| Hunjan | Asian Indian | South Asian | Wong et al. |
| Hunnex | Chinese | Chinese | Wong et al. |
| Huo | Chinese | Chinese | Kranick et al. |
| Huong | Vietnamese | Other Asian | Wong et al. |
| Huq |  | South Asian | Lim et al. |
| Huque |  | South Asian | Lim et al. |
| Hur | Korean | Other Asian | Wong et al. |
| Hurdle |  | South Asian | Lim et al. |
| Hurh | Korean | Other Asian | Wong et al. |
| Hurtado | Filipino | Other Asian | Wong et al. |
| Husain |  | South Asian | Lim et al. |
| Husaini | Asian Indian | South Asian | Wong et al. |
| Husan | Asian Indian | South Asian | Wong et al. |
| Husein | Asian Indian | South Asian | Wong et al. |
| Hushen |  | South Asian | Lim et al. |
| Husmillo | Filipino | Other Asian | Wong et al. |
| Husneara |  | South Asian | Lim et al. |
| Hussain | Pakistani; Asian Indian; Bangladeshi | South Asian | Kranick et al. |
| Hussaini | Asian Indian | South Asian | Wong et al. |
| Hussan | Asian Indian | South Asian | Wong et al. |
| Hussein |  | South Asian | Lim et al. |
| Hussian | Asian Indian | South Asian | Wong et al. |
| Huu | Vietnamese | Other Asian | Wong et al. |
| Huy | Vietnamese | Other Asian | Wong et al. |
| Huyah | Vietnamese | Other Asian | Wong et al. |
| Huyen | Vietnamese | Other Asian | Wong et al. |
| Huyenh | Vietnamese | Other Asian | Wong et al. |
| Huyhn | Vietnamese | Other Asian | Wong et al. |
| Huyng | Vietnamese | Other Asian | Wong et al. |
| Huynh | Vietnamese | Other Asian | Kranick et al. |
| Hwa | Chinese | Chinese | Wong et al. |
| Hwai | Chinese | Chinese | Wong et al. |
| Hwan | Korean | Other Asian | Wong et al. |
| Hwang | Korean; Chinese | Other Asian | Kranick et al. |
| Hwangbo | Korean | Other Asian | Wong et al. |
| Hwangpo | Korean | Other Asian | Wong et al. |
| Hwe | Chinese | Chinese | Wong et al. |
| Hwei | Chinese | Chinese | Wong et al. |
| Hwi | Korean | Other Asian | Wong et al. |
| Hwo | Chinese | Chinese | Wong et al. |
| Hwong | Chinese | Chinese | Wong et al. |
| Hwu | Chinese | Chinese | Wong et al. |
| Hwung | Chinese | Chinese | Wong et al. |
| Hy | Vietnamese | Other Asian | Wong et al. |
| Hyan | Korean | Other Asian | Wong et al. |
| Hyang | Korean | Other Asian | Wong et al. |
| Hyare | Asian Indian | South Asian | Wong et al. |
| Hyeon | Korean | Other Asian | Wong et al. |
| Hyodo | Japanese | Other Asian | Wong et al. |
| Hyon | Korean | Other Asian | Wong et al. |
| Hyong | Korean | Other Asian | Wong et al. |
| Hyuk | Korean | Other Asian | Wong et al. |
| Hyun | Korean | Other Asian | Wong et al. |
| Hyung | Korean | Other Asian | Wong et al. |
| I | Chinese | Chinese | Wong et al. |
| Iam | Chinese | Chinese | Wong et al. |
| Iasmin |  | South Asian | Lim et al. |
| Iau | Chinese | Chinese | Wong et al. |
| Iba | Japanese | Other Asian | Wong et al. |
| Ibabao | Filipino | Other Asian | Wong et al. |
| Ibale | Filipino | Other Asian | Wong et al. |
| Ibalio | Filipino | Other Asian | Wong et al. |
| Iban | Filipino | Other Asian | Wong et al. |
| Ibana | Filipino | Other Asian | Wong et al. |
| Ibanez | Filipino | Other Asian | Wong et al. |
| Ibaraki | Japanese | Other Asian | Wong et al. |
| Ibardolaza | Filipino | Other Asian | Wong et al. |
| Ibarra | Filipino | Other Asian | Wong et al. |
| Ibarreta | Filipino | Other Asian | Wong et al. |
| Ibarrola | Filipino | Other Asian | Wong et al. |
| Ibasan | Filipino | Other Asian | Wong et al. |
| Ibasco | Filipino | Other Asian | Wong et al. |
| Ibatuan | Filipino | Other Asian | Wong et al. |
| Ibay | Filipino | Other Asian | Wong et al. |
| Ibe | Filipino | Other Asian | Wong et al. |
| Ibea | Filipino | Other Asian | Wong et al. |
| Ibera | Filipino | Other Asian | Wong et al. |
| Ibon | Filipino | Other Asian | Wong et al. |
| Ibrahim |  | South Asian | Lim et al. |
| Ibrar |  | South Asian | Lim et al. |
| Ibuan | Filipino | Other Asian | Wong et al. |
| Ibuki | Japanese | Other Asian | Wong et al. |
| Ibus | Filipino | Other Asian | Wong et al. |
| Icalla | Filipino | Other Asian | Wong et al. |
| Icarangal | Filipino | Other Asian | Wong et al. |
| Icarro | Filipino | Other Asian | Wong et al. |
| Icasiano | Filipino | Other Asian | Wong et al. |
| Icay | Filipino | Other Asian | Wong et al. |
| Icban | Filipino | Other Asian | Wong et al. |
| Ichiba | Japanese | Other Asian | Wong et al. |
| Ichida | Japanese | Other Asian | Wong et al. |
| Ichige | Japanese | Other Asian | Wong et al. |
| Ichihara | Japanese | Other Asian | Wong et al. |
| Ichihashi | Japanese | Other Asian | Wong et al. |
| Ichii | Japanese | Other Asian | Wong et al. |
| Ichijo | Japanese | Other Asian | Wong et al. |
| Ichikawa | Japanese | Other Asian | Wong et al. |
| Ichiki | Japanese | Other Asian | Wong et al. |
| Ichimaru | Japanese | Other Asian | Wong et al. |
| Ichimura | Japanese | Other Asian | Wong et al. |
| Ichino | Japanese | Other Asian | Wong et al. |
| Ichinohe | Japanese | Other Asian | Wong et al. |
| Ichinose | Japanese | Other Asian | Wong et al. |
| Ichioka | Japanese | Other Asian | Wong et al. |
| Ichiro | Japanese | Other Asian | Wong et al. |
| Ichise | Japanese | Other Asian | Wong et al. |
| Ichishima | Japanese | Other Asian | Wong et al. |
| Ichiyama | Japanese | Other Asian | Wong et al. |
| Ichiyanagi | Japanese | Other Asian | Wong et al. |
| Ick | Filipino | Other Asian | Wong et al. |
| Ico | Filipino | Other Asian | Wong et al. |
| Ida | Japanese | Other Asian | Wong et al. |
| Idanan | Filipino | Other Asian | Wong et al. |
| Ide | Japanese | Other Asian | Wong et al. |
| Idea | Filipino | Other Asian | Wong et al. |
| Ideta | Japanese | Other Asian | Wong et al. |
| Idian | Filipino | Other Asian | Wong et al. |
| Idica | Filipino | Other Asian | Wong et al. |
| Idichandy | Asian Indian | South Asian | Wong et al. |
| Idicula | Asian Indian | South Asian | Wong et al. |
| Idiculla | Asian Indian | South Asian | Wong et al. |
| Idio | Filipino | Other Asian | Wong et al. |
| Ido | Japanese | Other Asian | Wong et al. |
| Idos | Filipino | Other Asian | Wong et al. |
| Idrees |  | South Asian | Lim et al. |
| Idris |  | South Asian | Lim et al. |
| Iengar | Asian Indian | South Asian | Wong et al. |
| Ieong | Chinese | Chinese | Wong et al. |
| Iftikhar | Pakistani | South Asian | Kranick et al. |
| Ifuku | Japanese | Other Asian | Wong et al. |
| Ifurung | Filipino | Other Asian | Wong et al. |
| Iga | Japanese | Other Asian | Wong et al. |
| Igama | Filipino | Other Asian | Wong et al. |
| Igana | Filipino | Other Asian | Wong et al. |
| Igarashi | Japanese | Other Asian | Wong et al. |
| Igari | Japanese | Other Asian | Wong et al. |
| Igarta | Filipino | Other Asian | Wong et al. |
| Igata | Japanese | Other Asian | Wong et al. |
| Igawa | Japanese | Other Asian | Wong et al. |
| Igaya | Filipino | Other Asian | Wong et al. |
| Igayac | Filipino | Other Asian | Wong et al. |
| Ige | Japanese | Other Asian | Wong et al. |
| Igei | Japanese | Other Asian | Wong et al. |
| Igeta | Japanese | Other Asian | Wong et al. |
| Igharas | Filipino | Other Asian | Wong et al. |
| Iglesia | Filipino | Other Asian | Wong et al. |
| Iglesias | Filipino | Other Asian | Wong et al. |
| Ignacio | Filipino | Other Asian | Wong et al. |
| Ignao | Filipino | Other Asian | Wong et al. |
| Igne | Filipino | Other Asian | Wong et al. |
| Igno | Filipino | Other Asian | Wong et al. |
| Igtanloc | Filipino | Other Asian | Wong et al. |
| Igtiben | Filipino | Other Asian | Wong et al. |
| Iguban | Filipino | Other Asian | Wong et al. |
| Iguchi | Japanese | Other Asian | Wong et al. |
| Iha | Japanese | Other Asian | Wong et al. |
| Ihara | Japanese | Other Asian | Wong et al. |
| Ihm | Korean | Other Asian | Wong et al. |
| Ii | Japanese | Other Asian | Wong et al. |
| Iida | Japanese | Other Asian | Wong et al. |
| Iidaka | Japanese | Other Asian | Wong et al. |
| Iijima | Japanese | Other Asian | Wong et al. |
| Iimori | Japanese | Other Asian | Wong et al. |
| Iimura | Japanese | Other Asian | Wong et al. |
| Iino | Japanese | Other Asian | Wong et al. |
| Iinuma | Japanese | Other Asian | Wong et al. |
| Iio | Japanese | Other Asian | Wong et al. |
| Iiyama | Japanese | Other Asian | Wong et al. |
| Iizuka | Japanese | Other Asian | Wong et al. |
| Iizumi | Japanese | Other Asian | Wong et al. |
| Ijichi | Japanese | Other Asian | Wong et al. |
| Ijima | Japanese | Other Asian | Wong et al. |
| Ijuin | Japanese | Other Asian | Wong et al. |
| Ik | Korean | Other Asian | Wong et al. |
| Ikada | Japanese | Other Asian | Wong et al. |
| Ikai | Japanese | Other Asian | Wong et al. |
| Ikari | Japanese | Other Asian | Wong et al. |
| Ikawa | Japanese | Other Asian | Wong et al. |
| Ike | Japanese | Other Asian | Wong et al. |
| Ikebe | Japanese | Other Asian | Wong et al. |
| Ikebuchi | Japanese | Other Asian | Wong et al. |
| Ikeda | Japanese | Other Asian | Wong et al. |
| Ikegami | Japanese | Other Asian | Wong et al. |
| Ikehara | Japanese | Other Asian | Wong et al. |
| Ikehata | Japanese | Other Asian | Wong et al. |
| Ikejiri | Japanese | Other Asian | Wong et al. |
| Ikemi | Japanese | Other Asian | Wong et al. |
| Ikemiya | Japanese | Other Asian | Wong et al. |
| Ikemoto | Japanese | Other Asian | Wong et al. |
| Ikemura | Japanese | Other Asian | Wong et al. |
| Ikenaga | Japanese | Other Asian | Wong et al. |
| Ikeno | Japanese | Other Asian | Wong et al. |
| Ikenoue | Japanese | Other Asian | Wong et al. |
| Ikeo | Japanese | Other Asian | Wong et al. |
| Ikeoka | Japanese | Other Asian | Wong et al. |
| Iketani | Japanese | Other Asian | Wong et al. |
| Ikeuchi | Japanese | Other Asian | Wong et al. |
| Ikeya | Japanese | Other Asian | Wong et al. |
| Ikezawa | Japanese | Other Asian | Wong et al. |
| Iki | Japanese | Other Asian | Wong et al. |
| Ikoma | Japanese | Other Asian | Wong et al. |
| Ikram |  | South Asian | Lim et al. |
| Ikuno | Japanese | Other Asian | Wong et al. |
| Ikushima | Japanese | Other Asian | Wong et al. |
| Ikuta | Japanese | Other Asian | Wong et al. |
| Ilaban | Filipino | Other Asian | Wong et al. |
| Ilac | Filipino | Other Asian | Wong et al. |
| Ilacas | Filipino | Other Asian | Wong et al. |
| Ilag | Filipino | Other Asian | Wong et al. |
| Ilagan | Filipino | Other Asian | Wong et al. |
| Ilan | Filipino | Other Asian | Wong et al. |
| Ilangilang | Filipino | Other Asian | Wong et al. |
| Ilano | Filipino | Other Asian | Wong et al. |
| Ilao | Filipino | Other Asian | Wong et al. |
| Ilar | Filipino | Other Asian | Wong et al. |
| Ilarde | Filipino | Other Asian | Wong et al. |
| Ilas | Filipino | Other Asian | Wong et al. |
| Ilasco | Filipino | Other Asian | Wong et al. |
| Ilasin | Filipino | Other Asian | Wong et al. |
| Ilaw | Filipino | Other Asian | Wong et al. |
| Ildefonso | Filipino | Other Asian | Wong et al. |
| Ildefonzo | Filipino | Other Asian | Wong et al. |
| Iledan | Filipino | Other Asian | Wong et al. |
| Ilejay | Filipino | Other Asian | Wong et al. |
| Ileto | Filipino | Other Asian | Wong et al. |
| Ilias | Bangladeshi | South Asian | Kranick et al. |
| Ilio | Filipino | Other Asian | Wong et al. |
| Illescas | Filipino | Other Asian | Wong et al. |
| Ilog | Filipino | Other Asian | Wong et al. |
| Iloreta | Filipino | Other Asian | Wong et al. |
| Ilumin | Filipino | Other Asian | Wong et al. |
| Ilusorio | Filipino | Other Asian | Wong et al. |
| Ilustre | Filipino | Other Asian | Wong et al. |
| Ilustrisimo | Filipino | Other Asian | Wong et al. |
| Iluzada | Filipino | Other Asian | Wong et al. |
| Ilyas | Asian Indian | South Asian | Wong et al. |
| Im | Korean | Other Asian | Wong et al. |
| Ima | Japanese | Other Asian | Wong et al. |
| Imabayashi | Japanese | Other Asian | Wong et al. |
| Imada | Japanese | Other Asian | Wong et al. |
| Imaeda | Japanese | Other Asian | Wong et al. |
| Imagawa | Japanese | Other Asian | Wong et al. |
| Imahashi | Japanese | Other Asian | Wong et al. |
| Imai | Japanese | Other Asian | Wong et al. |
| Imaizumi | Japanese | Other Asian | Wong et al. |
| Imal | Japanese | Other Asian | Wong et al. |
| Imam | Asian Indian | South Asian | Wong et al. |
| Imamiya | Japanese | Other Asian | Wong et al. |
| Imamoto | Japanese | Other Asian | Wong et al. |
| Imamura | Japanese | Other Asian | Wong et al. |
| Iman |  | South Asian | Lim et al. |
| Imanaga | Japanese | Other Asian | Wong et al. |
| Imanaka | Japanese | Other Asian | Wong et al. |
| Imanil | Filipino | Other Asian | Wong et al. |
| Imanishi | Japanese | Other Asian | Wong et al. |
| Imaoka | Japanese | Other Asian | Wong et al. |
| Imasa | Filipino | Other Asian | Wong et al. |
| Imazawa | Japanese | Other Asian | Wong et al. |
| Imazeki | Japanese | Other Asian | Wong et al. |
| Imazu | Japanese | Other Asian | Wong et al. |
| Imbat | Filipino | Other Asian | Wong et al. |
| Imbo | Filipino | Other Asian | Wong et al. |
| Imbuido | Filipino | Other Asian | Wong et al. |
| Imoto | Japanese | Other Asian | Wong et al. |
| Impelido | Filipino | Other Asian | Wong et al. |
| Imperial | Filipino | Other Asian | Wong et al. |
| Imperio | Filipino | Other Asian | Wong et al. |
| Importante | Filipino | Other Asian | Wong et al. |
| Imran | Pakistani; Bangladeshi | South Asian | Kranick et al. |
| Imson | Filipino | Other Asian | Wong et al. |
| Imura | Japanese | Other Asian | Wong et al. |
| Imuta | Japanese | Other Asian | Wong et al. |
| In | Korean | Other Asian | Wong et al. |
| Ina | Japanese | Other Asian | Wong et al. |
| Inaba | Japanese | Other Asian | Wong et al. |
| Inacay | Filipino | Other Asian | Wong et al. |
| Inada | Japanese | Other Asian | Wong et al. |
| Inafuku | Japanese | Other Asian | Wong et al. |
| Inagaki | Japanese | Other Asian | Wong et al. |
| Inagawa | Japanese | Other Asian | Wong et al. |
| Inage | Japanese | Other Asian | Wong et al. |
| Inaldo | Filipino | Other Asian | Wong et al. |
| Inamasu | Japanese | Other Asian | Wong et al. |
| Inamdar | Asian Indian | South Asian | Wong et al. |
| Inami | Japanese | Other Asian | Wong et al. |
| Inamine | Japanese | Other Asian | Wong et al. |
| Inamori | Japanese | Other Asian | Wong et al. |
| Inamoto | Japanese | Other Asian | Wong et al. |
| Inamura | Japanese | Other Asian | Wong et al. |
| Inao | Japanese | Other Asian | Wong et al. |
| Inaoka | Japanese | Other Asian | Wong et al. |
| Inarda | Filipino | Other Asian | Wong et al. |
| Inatome | Japanese | Other Asian | Wong et al. |
| Inay | Filipino | Other Asian | Wong et al. |
| Inayama | Japanese | Other Asian | Wong et al. |
| Inayoshi | Japanese | Other Asian | Wong et al. |
| Inazu | Japanese | Other Asian | Wong et al. |
| Inciong | Filipino | Other Asian | Wong et al. |
| Incoy | Filipino | Other Asian | Wong et al. |
| Inda | Filipino | Other Asian | Wong et al. |
| Indar |  | South Asian | Lim et al. |
| Indarjit |  | South Asian | Lim et al. |
| Indiongco | Filipino | Other Asian | Wong et al. |
| Inductivo | Filipino | Other Asian | Wong et al. |
| Ines | Filipino | Other Asian | Wong et al. |
| Inez | Filipino | Other Asian | Wong et al. |
| Infantado | Filipino | Other Asian | Wong et al. |
| Infante | Filipino | Other Asian | Wong et al. |
| Infiel | Filipino | Other Asian | Wong et al. |
| Infiesto | Filipino | Other Asian | Wong et al. |
| Ing | Chinese | Chinese | Wong et al. |
| Ingalla | Filipino | Other Asian | Wong et al. |
| Ingco | Filipino | Other Asian | Wong et al. |
| Ingel | Filipino | Other Asian | Wong et al. |
| Ingente | Filipino | Other Asian | Wong et al. |
| Inguito | Filipino | Other Asian | Wong et al. |
| Iniba | Filipino | Other Asian | Wong et al. |
| Inigo | Filipino | Other Asian | Wong et al. |
| Inis | Filipino | Other Asian | Wong et al. |
| Ino | Japanese | Other Asian | Wong et al. |
| Inocelda | Filipino | Other Asian | Wong et al. |
| Inocencio | Filipino | Other Asian | Wong et al. |
| Inocente | Filipino | Other Asian | Wong et al. |
| Inocentes | Filipino | Other Asian | Wong et al. |
| Inohara | Japanese | Other Asian | Wong et al. |
| Inoki | Japanese | Other Asian | Wong et al. |
| Inokuchi | Japanese | Other Asian | Wong et al. |
| Inomata | Japanese | Other Asian | Wong et al. |
| Inomoto | Japanese | Other Asian | Wong et al. |
| Inong | Filipino | Other Asian | Wong et al. |
| Inopea | Filipino | Other Asian | Wong et al. |
| Inosanto | Filipino | Other Asian | Wong et al. |
| Inose | Japanese | Other Asian | Wong et al. |
| Inoshita | Japanese | Other Asian | Wong et al. |
| Inot | Filipino | Other Asian | Wong et al. |
| Inoue | Japanese | Other Asian | Wong et al. |
| Inouye | Japanese | Other Asian | Wong et al. |
| Inove | Japanese | Other Asian | Wong et al. |
| Inovejas | Filipino | Other Asian | Wong et al. |
| Inserto | Filipino | Other Asian | Wong et al. |
| Inshanally |  | South Asian | Lim et al. |
| Insular | Filipino | Other Asian | Wong et al. |
| Intal | Filipino | Other Asian | Wong et al. |
| Interior | Filipino | Other Asian | Wong et al. |
| Intia | Filipino | Other Asian | Wong et al. |
| Inting | Filipino | Other Asian | Wong et al. |
| Inton | Filipino | Other Asian | Wong et al. |
| Intong | Filipino | Other Asian | Wong et al. |
| Intwala | Asian Indian | South Asian | Wong et al. |
| Inui | Japanese | Other Asian | Wong et al. |
| Inukai | Japanese | Other Asian | Wong et al. |
| Inumaru | Japanese | Other Asian | Wong et al. |
| Inumerable | Filipino | Other Asian | Wong et al. |
| Inuzuka | Japanese | Other Asian | Wong et al. |
| Invencion | Filipino | Other Asian | Wong et al. |
| Inventor | Filipino | Other Asian | Wong et al. |
| Ip | Chinese | Chinese | Kranick et al. |
| Ipac | Filipino | Other Asian | Wong et al. |
| Ipalari | Filipino | Other Asian | Wong et al. |
| Ipanag | Filipino | Other Asian | Wong et al. |
| Ipapo | Filipino | Other Asian | Wong et al. |
| Iqbal | Pakistani | South Asian | Kranick et al. |
| Irabagon | Filipino | Other Asian | Wong et al. |
| Iraha | Japanese | Other Asian | Wong et al. |
| Irani | Asian Indian | South Asian | Wong et al. |
| Irao | Filipino | Other Asian | Wong et al. |
| Irei | Japanese | Other Asian | Wong et al. |
| Irfan |  | South Asian | Lim et al. |
| Iriarte | Filipino | Other Asian | Wong et al. |
| Irie | Japanese | Other Asian | Wong et al. |
| Irikura | Japanese | Other Asian | Wong et al. |
| Irimajiri | Japanese | Other Asian | Wong et al. |
| Iritani | Japanese | Other Asian | Wong et al. |
| Iriyama | Japanese | Other Asian | Wong et al. |
| Iriye | Japanese | Other Asian | Wong et al. |
| Irlandez | Filipino | Other Asian | Wong et al. |
| Irshad | Pakistani | South Asian | Kranick et al. |
| Iruguin | Filipino | Other Asian | Wong et al. |
| Isa | Japanese | Other Asian | Wong et al. |
| Isaac |  | South Asian | Lim et al. |
| Isabelo | Filipino | Other Asian | Wong et al. |
| Isac | Asian Indian | South Asian | Wong et al. |
| Isada | Filipino | Other Asian | Wong et al. |
| Isago | Japanese | Other Asian | Wong et al. |
| Isaguirre | Filipino | Other Asian | Wong et al. |
| Isaka | Japanese | Other Asian | Wong et al. |
| Isamu | Japanese | Other Asian | Wong et al. |
| Isang | Chinese | Chinese | Wong et al. |
| Isao | Chinese | Chinese | Wong et al. |
| Isardat |  | South Asian | Lim et al. |
| Isawa | Japanese | Other Asian | Wong et al. |
| Isberto | Filipino | Other Asian | Wong et al. |
| Ise | Japanese | Other Asian | Wong et al. |
| Iseda | Japanese | Other Asian | Wong et al. |
| Iseki | Japanese | Other Asian | Wong et al. |
| Iseri | Japanese | Other Asian | Wong et al. |
| Ishak | Asian Indian | South Asian | Wong et al. |
| Ishaq |  | South Asian | Lim et al. |
| Ishi | Japanese | Other Asian | Wong et al. |
| Ishibashi | Japanese | Other Asian | Kranick et al. |
| Ishibe | Japanese | Other Asian | Wong et al. |
| Ishida | Japanese | Other Asian | Kranick et al. |
| Ishido | Japanese | Other Asian | Wong et al. |
| Ishigaki | Japanese | Other Asian | Wong et al. |
| Ishigami | Japanese | Other Asian | Wong et al. |
| Ishige | Japanese | Other Asian | Wong et al. |
| Ishigure | Japanese | Other Asian | Wong et al. |
| Ishiguro | Japanese | Other Asian | Wong et al. |
| Ishihama | Japanese | Other Asian | Wong et al. |
| Ishihara | Japanese | Other Asian | Wong et al. |
| Ishii | Japanese | Other Asian | Wong et al. |
| Ishijima | Japanese | Other Asian | Wong et al. |
| Ishikawa | Japanese | Other Asian | Kranick et al. |
| Ishiki | Japanese | Other Asian | Wong et al. |
| Ishikura | Japanese | Other Asian | Wong et al. |
| Ishil | Japanese | Other Asian | Wong et al. |
| Ishima | Japanese | Other Asian | Wong et al. |
| Ishimaru | Japanese | Other Asian | Wong et al. |
| Ishimatsu | Japanese | Other Asian | Wong et al. |
| Ishimine | Japanese | Other Asian | Wong et al. |
| Ishimori | Japanese | Other Asian | Wong et al. |
| Ishimoto | Japanese | Other Asian | Wong et al. |
| Ishimura | Japanese | Other Asian | Wong et al. |
| Ishino | Japanese | Other Asian | Wong et al. |
| Ishioka | Japanese | Other Asian | Wong et al. |
| Ishitani | Japanese | Other Asian | Wong et al. |
| Ishiwata | Japanese | Other Asian | Wong et al. |
| Ishiwatari | Japanese | Other Asian | Wong et al. |
| Ishiyama | Japanese | Other Asian | Wong et al. |
| Ishizaka | Japanese | Other Asian | Wong et al. |
| Ishizaki | Japanese | Other Asian | Wong et al. |
| Ishizawa | Japanese | Other Asian | Kranick et al. |
| Ishizu | Japanese | Other Asian | Wong et al. |
| Ishizuka | Japanese | Other Asian | Wong et al. |
| Ishizuki | Japanese | Other Asian | Wong et al. |
| Ishll | Japanese | Other Asian | Wong et al. |
| Ishmael |  | South Asian | Lim et al. |
| Isidoro | Filipino | Other Asian | Wong et al. |
| Isidro | Filipino | Other Asian | Wong et al. |
| Isip | Filipino | Other Asian | Wong et al. |
| Isla | Filipino | Other Asian | Wong et al. |
| Islam | Bangladeshi | South Asian | Kranick et al. |
| Islamnibir |  | South Asian | Lim et al. |
| Islaya | Filipino | Other Asian | Wong et al. |
| Isleta | Filipino | Other Asian | Wong et al. |
| Ismael | Filipino | Other Asian | Wong et al. |
| Ismail |  | South Asian | Lim et al. |
| Iso | Japanese | Other Asian | Wong et al. |
| Isobe | Japanese | Other Asian | Wong et al. |
| Isoda | Japanese | Other Asian | Wong et al. |
| Isogai | Japanese | Other Asian | Wong et al. |
| Isokawa | Japanese | Other Asian | Wong et al. |
| Isomura | Japanese | Other Asian | Wong et al. |
| Isono | Japanese | Other Asian | Wong et al. |
| Isorena | Filipino | Other Asian | Wong et al. |
| Isozaki | Japanese | Other Asian | Wong et al. |
| Israr |  | South Asian | Lim et al. |
| Issa | Asian Indian | South Asian | Wong et al. |
| Issac | Asian Indian | South Asian | Wong et al. |
| Isshiki | Japanese | Other Asian | Wong et al. |
| Isteak |  | South Asian | Lim et al. |
| Isturis | Filipino | Other Asian | Wong et al. |
| Itaas | Filipino | Other Asian | Wong et al. |
| Itabashi | Japanese | Other Asian | Wong et al. |
| Itagaki | Japanese | Other Asian | Wong et al. |
| Itahashi | Japanese | Other Asian | Wong et al. |
| Itakura | Japanese | Other Asian | Wong et al. |
| Itami | Japanese | Other Asian | Wong et al. |
| Itani | Japanese | Other Asian | Wong et al. |
| Itano | Japanese | Other Asian | Wong et al. |
| Itaya | Japanese | Other Asian | Wong et al. |
| Itchon | Filipino | Other Asian | Wong et al. |
| Itliong | Filipino | Other Asian | Wong et al. |
| Ito | Japanese | Other Asian | Kranick et al. |
| Itoh | Japanese | Other Asian | Kranick et al. |
| Itoi | Japanese | Other Asian | Wong et al. |
| Itokawa | Japanese | Other Asian | Wong et al. |
| Itokazu | Japanese | Other Asian | Wong et al. |
| Itonaga | Japanese | Other Asian | Wong et al. |
| Itoses | Filipino | Other Asian | Wong et al. |
| Itow | Japanese | Other Asian | Wong et al. |
| Itty | Asian Indian | South Asian | Wong et al. |
| Ituriaga | Filipino | Other Asian | Wong et al. |
| Iturralde | Filipino | Other Asian | Wong et al. |
| Itwaru |  | South Asian | Lim et al. |
| Iu | Chinese | Chinese | Wong et al. |
| Iuchi | Japanese | Other Asian | Wong et al. |
| Ivanoff | Chinese | Chinese | Wong et al. |
| Ivanov | Chinese | Chinese | Wong et al. |
| Iwa | Japanese | Other Asian | Wong et al. |
| Iwabuchi | Japanese | Other Asian | Wong et al. |
| Iwahara | Japanese | Other Asian | Wong et al. |
| Iwahashi | Japanese | Other Asian | Wong et al. |
| Iwahori | Japanese | Other Asian | Wong et al. |
| Iwai | Japanese | Other Asian | Wong et al. |
| Iwakami | Japanese | Other Asian | Wong et al. |
| Iwakata | Japanese | Other Asian | Wong et al. |
| Iwakawa | Japanese | Other Asian | Wong et al. |
| Iwaki | Japanese | Other Asian | Wong et al. |
| Iwakiri | Japanese | Other Asian | Wong et al. |
| Iwakura | Japanese | Other Asian | Wong et al. |
| Iwama | Japanese | Other Asian | Wong et al. |
| Iwamatsu | Japanese | Other Asian | Wong et al. |
| Iwami | Japanese | Other Asian | Wong et al. |
| Iwamori | Japanese | Other Asian | Wong et al. |
| Iwamoto | Japanese | Other Asian | Wong et al. |
| Iwamura | Japanese | Other Asian | Wong et al. |
| Iwanaga | Japanese | Other Asian | Wong et al. |
| Iwanami | Japanese | Other Asian | Wong et al. |
| Iwane | Japanese | Other Asian | Wong et al. |
| Iwano | Japanese | Other Asian | Wong et al. |
| Iwao | Japanese | Other Asian | Wong et al. |
| Iwasa | Japanese | Other Asian | Wong et al. |
| Iwasaki | Japanese | Other Asian | Wong et al. |
| Iwasawa | Japanese | Other Asian | Wong et al. |
| Iwase | Japanese | Other Asian | Wong et al. |
| Iwashima | Japanese | Other Asian | Wong et al. |
| Iwashiro | Japanese | Other Asian | Kranick et al. |
| Iwashita | Japanese | Other Asian | Wong et al. |
| Iwata | Japanese | Other Asian | Wong et al. |
| Iwatani | Japanese | Other Asian | Wong et al. |
| Iwato | Japanese | Other Asian | Wong et al. |
| Iwatsu | Japanese | Other Asian | Wong et al. |
| Iwatsubo | Japanese | Other Asian | Wong et al. |
| Iwatsuki | Japanese | Other Asian | Wong et al. |
| Iwawaki | Japanese | Other Asian | Wong et al. |
| Iway | Filipino | Other Asian | Wong et al. |
| Iwaya | Japanese | Other Asian | Wong et al. |
| Iwayama | Japanese | Other Asian | Wong et al. |
| Iwazaki | Japanese | Other Asian | Wong et al. |
| Iyama | Japanese | Other Asian | Wong et al. |
| Iyengar | Asian Indian | South Asian | Wong et al. |
| Iyer | Asian Indian | South Asian | Wong et al. |
| Iype | Asian Indian | South Asian | Wong et al. |
| Izaki | Japanese | Other Asian | Wong et al. |
| Izawa | Japanese | Other Asian | Wong et al. |
| Izeki | Japanese | Other Asian | Wong et al. |
| Izon | Filipino | Other Asian | Wong et al. |
| Izu | Japanese | Other Asian | Wong et al. |
| Izuhara | Japanese | Other Asian | Wong et al. |
| Izuka | Japanese | Other Asian | Wong et al. |
| Izumi | Japanese | Other Asian | Wong et al. |
| Izumida | Japanese | Other Asian | Wong et al. |
| Izumisawa | Japanese | Other Asian | Wong et al. |
| Izumiya | Japanese | Other Asian | Wong et al. |
| Izuno | Japanese | Other Asian | Wong et al. |
| Izutsu | Japanese | Other Asian | Wong et al. |
| Ja | Korean | Other Asian | Wong et al. |
| Jabagat | Filipino | Other Asian | Wong et al. |
| Jaballas | Filipino | Other Asian | Wong et al. |
| Jaban | Filipino | Other Asian | Wong et al. |
| Jabbar | Asian Indian | South Asian | Wong et al. |
| Jabidando | Filipino | Other Asian | Wong et al. |
| Jabier | Filipino | Other Asian | Wong et al. |
| Jabines | Filipino | Other Asian | Wong et al. |
| Jabola | Filipino | Other Asian | Wong et al. |
| Jabonillo | Filipino | Other Asian | Wong et al. |
| Jabson | Filipino | Other Asian | Wong et al. |
| Jaca | Filipino | Other Asian | Wong et al. |
| Jacala | Filipino | Other Asian | Wong et al. |
| Jacalan | Filipino | Other Asian | Wong et al. |
| Jacalne | Filipino | Other Asian | Wong et al. |
| Jacildo | Filipino | Other Asian | Wong et al. |
| Jacildone | Filipino | Other Asian | Wong et al. |
| Jacinto | Filipino | Other Asian | Wong et al. |
| Jacoba | Filipino | Other Asian | Wong et al. |
| Jacobe | Filipino | Other Asian | Wong et al. |
| Jacobo | Filipino | Other Asian | Wong et al. |
| Jacosalem | Filipino | Other Asian | Wong et al. |
| Jadav | Asian Indian | South Asian | Wong et al. |
| Jadavji | Asian Indian | South Asian | Wong et al. |
| Jadeja | Asian Indian | South Asian | Wong et al. |
| Jadhav | Asian Indian | South Asian | Wong et al. |
| Jae | Korean | Other Asian | Wong et al. |
| Jaen | Filipino | Other Asian | Wong et al. |
| Jaena | Filipino | Other Asian | Wong et al. |
| Jafar | Asian Indian | South Asian | Wong et al. |
| Jafari | Asian Indian | South Asian | Wong et al. |
| Jafarullah |  | South Asian | Lim et al. |
| Jaffer | Asian Indian | South Asian | Wong et al. |
| Jafferi | Asian Indian | South Asian | Wong et al. |
| Jaffery | Asian Indian | South Asian | Wong et al. |
| Jaffri | Asian Indian | South Asian | Wong et al. |
| Jafri | Asian Indian | South Asian | Wong et al. |
| Jafry | Asian Indian | South Asian | Wong et al. |
| Jagannath | Asian Indian | South Asian | Wong et al. |
| Jagannathan | Asian Indian | South Asian | Wong et al. |
| Jagat |  | South Asian | Lim et al. |
| Jagdeo |  | South Asian | Lim et al. |
| Jagga | Asian Indian | South Asian | Wong et al. |
| Jagiah |  | South Asian | Lim et al. |
| Jagmohan |  | South Asian | Lim et al. |
| Jagnarine |  | South Asian | Lim et al. |
| Jagsarran |  | South Asian | Lim et al. |
| Jahagirdar | Asian Indian | South Asian | Wong et al. |
| Jahan | Asian Indian; Bangladeshi | South Asian | Kranick et al. |
| Jahangir | Bangladeshi; Pakistani | South Asian | Kranick et al. |
| Jahawir |  | South Asian | Lim et al. |
| Jahir | Asian Indian; Bangladeshi | South Asian | Kranick et al. |
| Jahng | Korean | Other Asian | Wong et al. |
| Jai | Chinese | Chinese | Wong et al. |
| Jaidka | Asian Indian | South Asian | Wong et al. |
| Jaikaran |  | South Asian | Lim et al. |
| Jaikavan |  | South Asian | Lim et al. |
| Jailall |  | South Asian | Lim et al. |
| Jaimangaldebra |  | South Asian | Lim et al. |
| Jaime | Filipino | Other Asian | Wong et al. |
| Jain | Asian Indian | South Asian | Kranick et al. |
| Jaing | Chinese | Chinese | Wong et al. |
| Jair | Chinese | Chinese | Wong et al. |
| Jairam | Asian Indian | South Asian | Wong et al. |
| Jaiswal | Asian Indian | South Asian | Wong et al. |
| Jaitly | Asian Indian | South Asian | Wong et al. |
| Jajal | Asian Indian | South Asian | Wong et al. |
| Jakaria |  | South Asian | Lim et al. |
| Jakosalem | Filipino | Other Asian | Wong et al. |
| Jala | Filipino | Other Asian | Wong et al. |
| Jalal | Pakistani; Bangladeshi | South Asian | Kranick et al. |
| Jalaluddin | Asian Indian | South Asian | Wong et al. |
| Jalan | Asian Indian | South Asian | Wong et al. |
| Jalandoni | Filipino | Other Asian | Wong et al. |
| Jalandoon | Filipino | Other Asian | Wong et al. |
| Jalbuena | Filipino | Other Asian | Wong et al. |
| Jalbuna | Filipino | Other Asian | Wong et al. |
| Jaleco | Filipino | Other Asian | Wong et al. |
| Jaleel | Asian Indian | South Asian | Wong et al. |
| Jalil | Asian Indian | South Asian | Wong et al. |
| Jallorina | Filipino | Other Asian | Wong et al. |
| Jalota | Asian Indian | South Asian | Wong et al. |
| Jam | Chinese | Chinese | Wong et al. |
| Jamal | Asian Indian | South Asian | Wong et al. |
| Jamaluddin | Asian Indian | South Asian | Wong et al. |
| Jamaly |  | South Asian | Lim et al. |
| Jamandre | Filipino | Other Asian | Wong et al. |
| Jambalos | Filipino | Other Asian | Wong et al. |
| Jambaro | Filipino | Other Asian | Wong et al. |
| Jambotkar | Asian Indian | South Asian | Wong et al. |
| Jambunathan | Asian Indian | South Asian | Wong et al. |
| Jambusaria | Asian Indian | South Asian | Wong et al. |
| Jameel | Asian Indian | South Asian | Wong et al. |
| Jamelo | Filipino | Other Asian | Wong et al. |
| Jamero | Filipino | Other Asian | Wong et al. |
| Jamias | Filipino | Other Asian | Wong et al. |
| Jamil | Pakistani | South Asian | Kranick et al. |
| Jamila | Filipino | Other Asian | Wong et al. |
| Jamilal |  | South Asian | Lim et al. |
| Jamilano | Filipino | Other Asian | Wong et al. |
| Jamili | Filipino | Other Asian | Wong et al. |
| Jamilla | Filipino | Other Asian | Wong et al. |
| Jamilosa | Filipino | Other Asian | Wong et al. |
| Jaminal | Filipino | Other Asian | Wong et al. |
| Jaminola | Filipino | Other Asian | Wong et al. |
| Jamir | Filipino | Other Asian | Wong et al. |
| Jamito | Filipino | Other Asian | Wong et al. |
| Jamlig | Filipino | Other Asian | Wong et al. |
| Jamnadas | Asian Indian | South Asian | Wong et al. |
| Jamoles | Filipino | Other Asian | Wong et al. |
| Jamolin | Filipino | Other Asian | Wong et al. |
| Jamon | Filipino | Other Asian | Wong et al. |
| Jamora | Filipino | Other Asian | Wong et al. |
| Jamorabon | Filipino | Other Asian | Wong et al. |
| Jan | Pakistani | South Asian | Kranick et al. |
| Janakiraman | Asian Indian | South Asian | Wong et al. |
| Jandoc | Filipino | Other Asian | Wong et al. |
| Jandu | Asian Indian | South Asian | Wong et al. |
| Janea | Filipino | Other Asian | Wong et al. |
| Jang | Korean | Other Asian | Kranick et al. |
| Jangaon | Filipino | Other Asian | Wong et al. |
| Jangda | Asian Indian | South Asian | Wong et al. |
| Jani | Asian Indian | South Asian | Kranick et al. |
| Janjua | Pakistani | South Asian | Kranick et al. |
| Jannat |  | South Asian | Lim et al. |
| Janolino | Filipino | Other Asian | Wong et al. |
| Janolo | Filipino | Other Asian | Wong et al. |
| Jao | Chinese | Chinese | Wong et al. |
| Jaochico | Filipino | Other Asian | Wong et al. |
| Jaojoco | Filipino | Other Asian | Wong et al. |
| Japan | Japanese | Other Asian | Wong et al. |
| Japitana | Filipino | Other Asian | Wong et al. |
| Japlit | Filipino | Other Asian | Wong et al. |
| Japson | Filipino | Other Asian | Wong et al. |
| Japzon | Filipino | Other Asian | Wong et al. |
| Jaquias | Filipino | Other Asian | Wong et al. |
| Jar | Chinese | Chinese | Wong et al. |
| Jara | Filipino | Other Asian | Wong et al. |
| Jarabelo | Filipino | Other Asian | Wong et al. |
| Jaramilla | Filipino | Other Asian | Wong et al. |
| Jaramillo | Filipino | Other Asian | Wong et al. |
| Jaranilla | Filipino | Other Asian | Wong et al. |
| Jarasa | Filipino | Other Asian | Wong et al. |
| Jaravata | Filipino | Other Asian | Wong et al. |
| Jardeleza | Filipino | Other Asian | Wong et al. |
| Jardenil | Filipino | Other Asian | Wong et al. |
| Jardiel | Filipino | Other Asian | Wong et al. |
| Jardin | Filipino | Other Asian | Kranick et al. |
| Jardiniano | Filipino | Other Asian | Wong et al. |
| Jardinico | Filipino | Other Asian | Wong et al. |
| Jardiolin | Filipino | Other Asian | Wong et al. |
| Jareno | Filipino | Other Asian | Wong et al. |
| Jarin | Filipino | Other Asian | Wong et al. |
| Jaring | Filipino | Other Asian | Wong et al. |
| Jariwala | Asian Indian | South Asian | Wong et al. |
| Jaro | Filipino | Other Asian | Wong et al. |
| Jarque | Filipino | Other Asian | Wong et al. |
| Jarquio | Filipino | Other Asian | Wong et al. |
| Jarumay | Filipino | Other Asian | Wong et al. |
| Jasani | Asian Indian | South Asian | Wong et al. |
| Jashbhai | Asian Indian | South Asian | Wong et al. |
| Jasmin |  | South Asian | Lim et al. |
| Jaspal | Asian Indian | South Asian | Wong et al. |
| Jassal | Asian Indian | South Asian | Wong et al. |
| Jassan |  | South Asian | Lim et al. |
| Jasti | Asian Indian | South Asian | Wong et al. |
| Jastillana | Filipino | Other Asian | Wong et al. |
| Jaswal | Asian Indian | South Asian | Wong et al. |
| Jatico | Filipino | Other Asian | Wong et al. |
| Jau | Chinese | Chinese | Wong et al. |
| Jaucian | Filipino | Other Asian | Wong et al. |
| Jauco | Filipino | Other Asian | Wong et al. |
| Jauhar | Asian Indian | South Asian | Wong et al. |
| Jaundoo |  | South Asian | Lim et al. |
| Jaurigue | Filipino | Other Asian | Wong et al. |
| Javan | Filipino | Other Asian | Wong et al. |
| Javar | Filipino | Other Asian | Wong et al. |
| Javate | Filipino | Other Asian | Wong et al. |
| Javed |  | South Asian | Lim et al. |
| Javeid |  | South Asian | Lim et al. |
| Javellana | Filipino | Other Asian | Wong et al. |
| Javelosa | Filipino | Other Asian | Wong et al. |
| Javeri | Asian Indian | South Asian | Wong et al. |
| Javia | Asian Indian | South Asian | Wong et al. |
| Javien | Filipino | Other Asian | Wong et al. |
| Javier | Filipino | Other Asian | Wong et al. |
| Javierto | Filipino | Other Asian | Wong et al. |
| Javillo | Filipino | Other Asian | Wong et al. |
| Javillonar | Filipino | Other Asian | Wong et al. |
| Javinar | Filipino | Other Asian | Wong et al. |
| Javines | Filipino | Other Asian | Wong et al. |
| Javonillo | Filipino | Other Asian | Wong et al. |
| Jaw | Chinese | Chinese | Wong et al. |
| Jawa | Asian Indian | South Asian | Wong et al. |
| Jawad | Asian Indian | South Asian | Wong et al. |
| Jawanda | Asian Indian | South Asian | Wong et al. |
| Jawandha | Asian Indian | South Asian | Wong et al. |
| Jay |  | South Asian | Lim et al. |
| Jayakar | Asian Indian | South Asian | Wong et al. |
| Jayantilal | Asian Indian | South Asian | Wong et al. |
| Jayaram | Asian Indian | South Asian | Wong et al. |
| Jayaraman | Asian Indian | South Asian | Wong et al. |
| Jayaswal | Asian Indian | South Asian | Wong et al. |
| Jayco | Filipino | Other Asian | Wong et al. |
| Jayme | Filipino | Other Asian | Wong et al. |
| Jazmin | Filipino | Other Asian | Wong et al. |
| Je | Korean | Other Asian | Wong et al. |
| Jea | Chinese | Chinese | Wong et al. |
| Jeang | Chinese | Chinese | Wong et al. |
| Jebunnahar |  | South Asian | Lim et al. |
| Jeddy | Asian Indian | South Asian | Wong et al. |
| Jee | Pakistani; Chinese | South Asian | Kranick et al. |
| Jeelani | Asian Indian | South Asian | Wong et al. |
| Jeet |  | South Asian | Lim et al. |
| Jehan | Asian Indian | South Asian | Wong et al. |
| Jei | Chinese | Chinese | Wong et al. |
| Jen | Chinese | Chinese | Wong et al. |
| Jeng | Chinese | Chinese | Wong et al. |
| Jeon | Korean | Other Asian | Kranick et al. |
| Jeong | Korean | Other Asian | Kranick et al. |
| Jequinto | Filipino | Other Asian | Wong et al. |
| Jer | Chinese | Chinese | Wong et al. |
| Jerath | Asian Indian | South Asian | Wong et al. |
| Jereos | Filipino | Other Asian | Wong et al. |
| Jerez | Filipino | Other Asian | Wong et al. |
| Jereza | Filipino | Other Asian | Wong et al. |
| Jerusalem | Filipino | Other Asian | Wong et al. |
| Jervoso | Filipino | Other Asian | Wong et al. |
| Jesalva | Filipino | Other Asian | Wong et al. |
| Jesena | Filipino | Other Asian | Wong et al. |
| Jesmin |  | South Asian | Lim et al. |
| Jesudas | Asian Indian | South Asian | Wong et al. |
| Jesudasan | Asian Indian | South Asian | Wong et al. |
| Jesudason | Asian Indian | South Asian | Wong et al. |
| Jesuitas | Filipino | Other Asian | Wong et al. |
| Jesus | Filipino | Other Asian | Wong et al. |
| Jethabhai | Asian Indian | South Asian | Wong et al. |
| Jethalal | Asian Indian | South Asian | Wong et al. |
| Jethva | Asian Indian | South Asian | Wong et al. |
| Jethwa | Asian Indian | South Asian | Wong et al. |
| Jeu | Chinese | Chinese | Wong et al. |
| Jeun | Korean | Other Asian | Wong et al. |
| Jeung | Chinese | Chinese | Wong et al. |
| Jew | Chinese | Chinese | Wong et al. |
| Jewel |  | South Asian | Lim et al. |
| Jha | Asian Indian | South Asian | Kranick et al. |
| Jhaj | Asian Indian | South Asian | Wong et al. |
| Jhala | Asian Indian | South Asian | Wong et al. |
| Jhamb | Asian Indian | South Asian | Wong et al. |
| Jhanda | Chinese | Chinese | Wong et al. |
| Jhanji | Asian Indian | South Asian | Wong et al. |
| Jhaveri | Asian Indian | South Asian | Wong et al. |
| Jhawar | Asian Indian | South Asian | Wong et al. |
| Jhee | Korean | Other Asian | Wong et al. |
| Jhin | Korean | Other Asian | Wong et al. |
| Jho | Korean | Other Asian | Wong et al. |
| Jhon | Korean | Other Asian | Wong et al. |
| Jhong | Korean | Other Asian | Wong et al. |
| Jhun | Korean | Other Asian | Wong et al. |
| Jhung | Korean | Other Asian | Wong et al. |
| Jhunu |  | South Asian | Lim et al. |
| Ji | Chinese | Chinese | Kranick et al. |
| Jia | Chinese | Chinese | Wong et al. |
| Jian | Chinese | Chinese | Wong et al. |
| Jiang | Chinese | Chinese | Kranick et al. |
| Jiao | Chinese | Chinese | Kranick et al. |
| Jick | Chinese | Chinese | Wong et al. |
| Jie | Chinese | Chinese | Wong et al. |
| Jien | Chinese | Chinese | Wong et al. |
| Jih | Chinese | Chinese | Wong et al. |
| Jijina | Asian Indian | South Asian | Wong et al. |
| Jik | Korean | Other Asian | Wong et al. |
| Jilani | Asian Indian | South Asian | Wong et al. |
| Jim | Chinese | Chinese | Wong et al. |
| Jimbo | Japanese | Other Asian | Wong et al. |
| Jimena | Filipino | Other Asian | Wong et al. |
| Jimenea | Filipino | Other Asian | Wong et al. |
| Jimenez | Filipino | Other Asian | Wong et al. |
| Jimeno | Filipino | Other Asian | Wong et al. |
| Jiminez | Filipino | Other Asian | Wong et al. |
| Jin | Chinese | Chinese | Kranick et al. |
| Jinbo | Japanese | Other Asian | Wong et al. |
| Jindal | Asian Indian | South Asian | Wong et al. |
| Jing | Chinese | Chinese | Wong et al. |
| Jingco | Filipino | Other Asian | Wong et al. |
| Jinnah | Asian Indian | South Asian | Wong et al. |
| Jinno | Japanese | Other Asian | Wong et al. |
| Jinon | Filipino | Other Asian | Wong et al. |
| Jinwala | Asian Indian | South Asian | Wong et al. |
| Jiongco | Filipino | Other Asian | Wong et al. |
| Jip | Chinese | Chinese | Wong et al. |
| Jiro | Japanese | Other Asian | Wong et al. |
| Jison | Filipino | Other Asian | Wong et al. |
| Jiu | Chinese | Chinese | Wong et al. |
| Jiva | Asian Indian | South Asian | Wong et al. |
| Jivabhai | Asian Indian | South Asian | Wong et al. |
| Jivan | Asian Indian | South Asian | Wong et al. |
| Jivani | Asian Indian | South Asian | Wong et al. |
| Jivanlal | Asian Indian | South Asian | Wong et al. |
| Jiwa | Asian Indian | South Asian | Wong et al. |
| Jiwani | Asian Indian | South Asian | Wong et al. |
| Jo | Korean | Other Asian | Wong et al. |
| Joa | Chinese | Chinese | Wong et al. |
| Joanino | Filipino | Other Asian | Wong et al. |
| Joaquin | Filipino | Other Asian | Wong et al. |
| Joaquino | Filipino | Other Asian | Wong et al. |
| Jobalia | Asian Indian | South Asian | Wong et al. |
| Jobanputra | Asian Indian | South Asian | Wong et al. |
| Jobeda |  | South Asian | Lim et al. |
| Jock | Chinese | Chinese | Wong et al. |
| Joco | Filipino | Other Asian | Wong et al. |
| Jocom | Filipino | Other Asian | Wong et al. |
| Jocson | Filipino | Other Asian | Wong et al. |
| Jodha |  | South Asian | Lim et al. |
| Joe | Chinese | Chinese | Wong et al. |
| Jog | Asian Indian | South Asian | Wong et al. |
| Joglekar | Asian Indian | South Asian | Wong et al. |
| Joh | Korean | Other Asian | Kranick et al. |
| Johal | Asian Indian | South Asian | Wong et al. |
| Johar | Pakistani | South Asian | Kranick et al. |
| Johl | Asian Indian | South Asian | Wong et al. |
| John | Asian Indian | South Asian | Kranick et al. |
| Johng | Korean | Other Asian | Wong et al. |
| Johri | Asian Indian | South Asian | Wong et al. |
| Jok | Chinese | Chinese | Wong et al. |
| Jom | Chinese | Chinese | Wong et al. |
| Jon | Chinese | Chinese | Wong et al. |
| Jonayed |  | South Asian | Lim et al. |
| Jone | Chinese | Chinese | Wong et al. |
| Jong | Chinese | Chinese | Wong et al. |
| Jongco | Filipino | Other Asian | Wong et al. |
| Jonnalagadda | Asian Indian | South Asian | Wong et al. |
| Jono | Japanese | Other Asian | Wong et al. |
| Joo | Korean | Other Asian | Wong et al. |
| Jooma | Asian Indian | South Asian | Wong et al. |
| Joon |  | South Asian | Lim et al. |
| Joong | Korean | Other Asian | Wong et al. |
| Jor | Chinese | Chinese | Wong et al. |
| Joree |  | South Asian | Lim et al. |
| Jorge | Filipino | Other Asian | Wong et al. |
| Jornacion | Filipino | Other Asian | Wong et al. |
| Jornales | Filipino | Other Asian | Wong et al. |
| Jorque | Filipino | Other Asian | Wong et al. |
| Jorvina | Filipino | Other Asian | Wong et al. |
| Josafat | Filipino | Other Asian | Wong et al. |
| Josan | Asian Indian | South Asian | Wong et al. |
| Jose | Filipino | Other Asian | Wong et al. |
| Josef | Filipino | Other Asian | Wong et al. |
| Joseph | Asian Indian | South Asian | Kranick et al. |
| Joseph-Pesnell |  | South Asian | Lim et al. |
| Joshi | Asian Indian; Nepali | South Asian | Kranick et al. |
| Joshna |  | South Asian | Lim et al. |
| Joshua | Asian Indian | South Asian | Wong et al. |
| Josol | Filipino | Other Asian | Wong et al. |
| Joson | Filipino | Other Asian | Wong et al. |
| Josue | Filipino | Other Asian | Wong et al. |
| Jota | Filipino | Other Asian | Wong et al. |
| Jothi |  | South Asian | Lim et al. |
| Jotie | Filipino | Other Asian | Wong et al. |
| Jotojot | Filipino | Other Asian | Wong et al. |
| Jou | Chinese | Chinese | Wong et al. |
| Jouhal | Asian Indian | South Asian | Wong et al. |
| Joun | Korean | Other Asian | Wong et al. |
| Joung | Korean | Other Asian | Wong et al. |
| Jove | Filipino | Other Asian | Wong et al. |
| Jovellano | Filipino | Other Asian | Wong et al. |
| Jovellanos | Filipino | Other Asian | Wong et al. |
| Joven | Filipino | Other Asian | Wong et al. |
| Jovenal | Filipino | Other Asian | Wong et al. |
| Jover | Filipino | Other Asian | Wong et al. |
| Jovero | Filipino | Other Asian | Wong et al. |
| Joves | Filipino | Other Asian | Wong et al. |
| Jow | Chinese | Chinese | Wong et al. |
| Joya | Filipino | Other Asian | Wong et al. |
| Jse | Chinese | Chinese | Wong et al. |
| Ju | Chinese | Chinese | Wong et al. |
| Juachon | Filipino | Other Asian | Wong et al. |
| Juadiong | Filipino | Other Asian | Wong et al. |
| Juan | Filipino | Other Asian | Wong et al. |
| Juaner | Filipino | Other Asian | Wong et al. |
| Juaneza | Filipino | Other Asian | Wong et al. |
| Juang | Chinese | Chinese | Wong et al. |
| Juangco | Filipino | Other Asian | Wong et al. |
| Juanico | Filipino | Other Asian | Wong et al. |
| Juanillo | Filipino | Other Asian | Wong et al. |
| Juanitas | Filipino | Other Asian | Wong et al. |
| Juarez | Filipino | Other Asian | Wong et al. |
| Juario | Filipino | Other Asian | Wong et al. |
| Juat | Filipino | Other Asian | Wong et al. |
| Jubilo | Filipino | Other Asian | Wong et al. |
| Jucaban | Filipino | Other Asian | Wong et al. |
| Jucar | Filipino | Other Asian | Wong et al. |
| Juco | Filipino | Other Asian | Wong et al. |
| Jucutan | Filipino | Other Asian | Wong et al. |
| Judal | Filipino | Other Asian | Wong et al. |
| Judalena | Filipino | Other Asian | Wong et al. |
| Judan | Filipino | Other Asian | Wong et al. |
| Judilla | Filipino | Other Asian | Wong et al. |
| Judistir |  | South Asian | Lim et al. |
| Jue | Chinese | Chinese | Wong et al. |
| Jueco | Filipino | Other Asian | Wong et al. |
| Juen | Chinese | Chinese | Wong et al. |
| Juezan | Filipino | Other Asian | Wong et al. |
| Jugal | Filipino | Other Asian | Wong et al. |
| Jugo | Filipino | Other Asian | Wong et al. |
| Jugueta | Filipino | Other Asian | Wong et al. |
| Juguilon | Filipino | Other Asian | Wong et al. |
| Juhn | Korean | Other Asian | Wong et al. |
| Jui | Chinese | Chinese | Wong et al. |
| Juico | Filipino | Other Asian | Wong et al. |
| Juinio | Filipino | Other Asian | Wong et al. |
| Julao | Filipino | Other Asian | Wong et al. |
| Jularbal | Filipino | Other Asian | Wong et al. |
| Julaton | Filipino | Other Asian | Wong et al. |
| Julio | Filipino | Other Asian | Wong et al. |
| Jum | Chinese | Chinese | Wong et al. |
| Juma | Asian Indian | South Asian | Wong et al. |
| Jumai |  | South Asian | Lim et al. |
| Jumalon | Filipino | Other Asian | Wong et al. |
| Jumamil | Filipino | Other Asian | Wong et al. |
| Jumangit | Filipino | Other Asian | Wong et al. |
| Jumaoas | Filipino | Other Asian | Wong et al. |
| Jumaquio | Filipino | Other Asian | Wong et al. |
| Jumarang | Filipino | Other Asian | Wong et al. |
| Jumawan | Filipino | Other Asian | Wong et al. |
| Jumsheed |  | South Asian | Lim et al. |
| Jun | Chinese; Korean | Chinese | Kranick et al. |
| Junaid | Asian Indian | South Asian | Wong et al. |
| Junayed |  | South Asian | Lim et al. |
| Junco | Filipino | Other Asian | Wong et al. |
| Juneja | Asian Indian | South Asian | Wong et al. |
| Junejo | Asian Indian | South Asian | Wong et al. |
| Jung | Korean | Other Asian | Kranick et al. |
| Juni | Filipino | Other Asian | Wong et al. |
| Junio | Filipino | Other Asian | Wong et al. |
| Junnarkar | Asian Indian | South Asian | Wong et al. |
| Junsay | Filipino | Other Asian | Wong et al. |
| Juntado | Filipino | Other Asian | Wong et al. |
| Juntereal | Filipino | Other Asian | Wong et al. |
| Juntilla | Filipino | Other Asian | Wong et al. |
| Juo | Chinese | Chinese | Wong et al. |
| Juong | Vietnamese | Other Asian | Wong et al. |
| Jurado | Filipino | Other Asian | Wong et al. |
| Jurilla | Filipino | Other Asian | Wong et al. |
| Jurwan |  | South Asian | Lim et al. |
| Jusay | Filipino | Other Asian | Wong et al. |
| Jusayan | Filipino | Other Asian | Wong et al. |
| Jusi | Filipino | Other Asian | Wong et al. |
| Juson | Filipino | Other Asian | Wong et al. |
| Justiniani | Filipino | Other Asian | Wong et al. |
| Justiniano | Filipino | Other Asian | Wong et al. |
| Justo | Filipino | Other Asian | Wong et al. |
| Jusufi |  | South Asian | Lim et al. |
| Juta | Filipino | Other Asian | Wong et al. |
| Juvida | Filipino | Other Asian | Wong et al. |
| Juynh | Vietnamese | Other Asian | Wong et al. |
| Jwarchan |  | South Asian | Lim et al. |
| Jyo | Japanese | Other Asian | Wong et al. |
| Ka | Chinese | Chinese | Wong et al. |
| Kaba | Japanese | Other Asian | Wong et al. |
| Kabahit | Filipino | Other Asian | Wong et al. |
| Kabani | Asian Indian | South Asian | Wong et al. |
| Kabaria | Asian Indian | South Asian | Wong et al. |
| Kabasawa | Japanese | Other Asian | Wong et al. |
| Kabashima | Japanese | Other Asian | Wong et al. |
| Kabaya | Japanese | Other Asian | Wong et al. |
| Kabayama | Japanese | Other Asian | Wong et al. |
| Kabayashi | Japanese | Other Asian | Wong et al. |
| Kaber |  | South Asian | Lim et al. |
| Kabigting | Filipino | Other Asian | Kranick et al. |
| Kabil |  | South Asian | Lim et al. |
| Kabiling | Filipino | Other Asian | Wong et al. |
| Kabir | Bangladeshi | South Asian | Kranick et al. |
| Kabirkhan |  | South Asian | Lim et al. |
| Kachroo | Asian Indian | South Asian | Wong et al. |
| Kacker | Asian Indian | South Asian | Wong et al. |
| Kada | Japanese | Other Asian | Wong et al. |
| Kadaba | Asian Indian | South Asian | Wong et al. |
| Kadakia | Asian Indian | South Asian | Wong et al. |
| Kadar |  | South Asian | Lim et al. |
| Kadavil | Asian Indian | South Asian | Wong et al. |
| Kadel |  | South Asian | Lim et al. |
| Kader |  | South Asian | Lim et al. |
| Kaderuzzaman |  | South Asian | Lim et al. |
| Kadery |  | South Asian | Lim et al. |
| Kadir | Asian Indian | South Asian | Wong et al. |
| Kadirgamar | Sri Lankan | South Asian | Kranick et al. |
| Kadirmuhammad |  | South Asian | Lim et al. |
| Kado | Japanese | Other Asian | Wong et al. |
| Kadokura | Japanese | Other Asian | Wong et al. |
| Kadono | Japanese | Other Asian | Wong et al. |
| Kadota | Japanese | Other Asian | Wong et al. |
| Kadowaki | Japanese | Other Asian | Wong et al. |
| Kadoya | Japanese | Other Asian | Wong et al. |
| Kadri | Asian Indian | South Asian | Wong et al. |
| Kae | Korean | Other Asian | Wong et al. |
| Kaga | Japanese | Other Asian | Wong et al. |
| Kagahastian | Filipino | Other Asian | Wong et al. |
| Kagal | Asian Indian | South Asian | Wong et al. |
| Kagami | Japanese | Other Asian | Wong et al. |
| Kagaoan | Filipino | Other Asian | Wong et al. |
| Kagawa | Japanese | Other Asian | Wong et al. |
| Kagaya | Japanese | Other Asian | Wong et al. |
| Kagayama | Japanese | Other Asian | Wong et al. |
| Kageyama | Japanese | Other Asian | Wong et al. |
| Kagi | Japanese | Other Asian | Wong et al. |
| Kagimoto | Japanese | Other Asian | Wong et al. |
| Kagiyama | Japanese | Other Asian | Wong et al. |
| Kagoshima | Japanese | Other Asian | Wong et al. |
| Kaguyutan | Filipino | Other Asian | Wong et al. |
| Kahng | Korean | Other Asian | Wong et al. |
| Kai | Chinese | Chinese | Wong et al. |
| Kaibara | Japanese | Other Asian | Wong et al. |
| Kaichi | Japanese | Other Asian | Wong et al. |
| Kaida | Japanese | Other Asian | Wong et al. |
| Kaihara | Japanese | Other Asian | Wong et al. |
| Kaiklian | Filipino | Other Asian | Wong et al. |
| Kailasam | Asian Indian | South Asian | Wong et al. |
| Kaino | Japanese | Other Asian | Wong et al. |
| Kainth | Asian Indian | South Asian | Wong et al. |
| Kainuma | Japanese | Other Asian | Wong et al. |
| Kaiser |  | South Asian | Lim et al. |
| Kaito | Japanese | Other Asian | Wong et al. |
| Kaizaki | Japanese | Other Asian | Wong et al. |
| Kaizuka | Japanese | Other Asian | Wong et al. |
| Kaji | Japanese | Other Asian | Wong et al. |
| Kajihara | Japanese | Other Asian | Wong et al. |
| Kajii | Japanese | Other Asian | Wong et al. |
| Kajikawa | Japanese | Other Asian | Wong et al. |
| Kajimura | Japanese | Other Asian | Wong et al. |
| Kajita | Japanese | Other Asian | Wong et al. |
| Kajitani | Japanese | Other Asian | Wong et al. |
| Kajiura | Japanese | Other Asian | Wong et al. |
| Kajiwara | Japanese | Other Asian | Wong et al. |
| Kajiya | Japanese | Other Asian | Wong et al. |
| Kajiyama | Japanese | Other Asian | Wong et al. |
| Kak | Korean | Other Asian | Wong et al. |
| Kaka | Asian Indian | South Asian | Wong et al. |
| Kakadia | Asian Indian | South Asian | Wong et al. |
| Kakani | Asian Indian | South Asian | Wong et al. |
| Kakar | Pakistani; Afghani; | South Asian | Kranick et al. |
| Kakarala | Asian Indian | South Asian | Wong et al. |
| Kakarla | Asian Indian | South Asian | Wong et al. |
| Kakazu | Japanese | Other Asian | Wong et al. |
| Kakegawa | Japanese | Other Asian | Wong et al. |
| Kakehashi | Japanese | Other Asian | Wong et al. |
| Kakehi | Japanese | Other Asian | Wong et al. |
| Kakihana | Japanese | Other Asian | Wong et al. |
| Kakihara | Japanese | Other Asian | Wong et al. |
| Kakimi | Japanese | Other Asian | Wong et al. |
| Kakimoto | Japanese | Other Asian | Wong et al. |
| Kakinuma | Japanese | Other Asian | Wong et al. |
| Kakita | Japanese | Other Asian | Wong et al. |
| Kakiuchi | Japanese | Other Asian | Wong et al. |
| Kakizaki | Japanese | Other Asian | Wong et al. |
| Kakkar | Asian Indian | South Asian | Wong et al. |
| Kakli | Asian Indian | South Asian | Wong et al. |
| Kakniya |  | South Asian | Lim et al. |
| Kako | Japanese | Other Asian | Wong et al. |
| Kaku | Japanese | Other Asian | Wong et al. |
| Kakuda | Japanese | Other Asian | Wong et al. |
| Kakuta | Japanese | Other Asian | Wong et al. |
| Kakutani | Japanese | Other Asian | Wong et al. |
| Kala | Asian Indian | South Asian | Wong et al. |
| Kalagayan | Filipino | Other Asian | Wong et al. |
| Kalam |  | South Asian | Lim et al. |
| Kalambakal | Filipino | Other Asian | Wong et al. |
| Kalani | Asian Indian | South Asian | Wong et al. |
| Kalaria | Asian Indian | South Asian | Wong et al. |
| Kalathil | Asian Indian | South Asian | Wong et al. |
| Kalaw | Filipino | Other Asian | Wong et al. |
| Kalayil | Asian Indian | South Asian | Wong et al. |
| Kaleka | Asian Indian | South Asian | Wong et al. |
| Kalia | Asian Indian | South Asian | Wong et al. |
| Kalidas | Asian Indian | South Asian | Wong et al. |
| Kalirai | Asian Indian | South Asian | Wong et al. |
| Kalkat | Asian Indian | South Asian | Wong et al. |
| Kalluri | Asian Indian | South Asian | Wong et al. |
| Kalpoo |  | South Asian | Lim et al. |
| Kalra | Asian Indian | South Asian | Kranick et al. |
| Kalsi | Asian Indian | South Asian | Wong et al. |
| Kalsoon |  | South Asian | Lim et al. |
| Kalu |  | South Asian | Lim et al. |
| Kalyan | Asian Indian | South Asian | Wong et al. |
| Kalyanasunda | Asian Indian | South Asian | Wong et al. |
| Kalyani | Asian Indian | South Asian | Wong et al. |
| Kam | Chinese | Chinese | Kranick et al. |
| Kama | Japanese | Other Asian | Wong et al. |
| Kamachi | Japanese | Other Asian | Wong et al. |
| Kamada | Japanese | Other Asian | Wong et al. |
| Kamakura | Japanese | Other Asian | Wong et al. |
| Kamal | Bangladeshi; Pakistani | South Asian | Kranick et al. |
| Kamalodeen |  | South Asian | Lim et al. |
| Kamaluddin | Asian Indian | South Asian | Wong et al. |
| Kamani | Asian Indian | South Asian | Wong et al. |
| Kamantigue | Filipino | Other Asian | Wong et al. |
| Kamaruddin | Asian Indian | South Asian | Wong et al. |
| Kamata | Japanese | Other Asian | Wong et al. |
| Kamath | Asian Indian | South Asian | Wong et al. |
| Kamatoy | Filipino | Other Asian | Wong et al. |
| Kambara | Japanese | Other Asian | Wong et al. |
| Kambayashi | Japanese | Other Asian | Wong et al. |
| Kambe | Japanese | Other Asian | Wong et al. |
| Kamboj | Asian Indian | South Asian | Wong et al. |
| Kamdar | Asian Indian | South Asian | Wong et al. |
| Kame | Japanese | Other Asian | Wong et al. |
| Kameda | Japanese | Other Asian | Wong et al. |
| Kamei | Japanese | Other Asian | Wong et al. |
| Kamekawa | Japanese | Other Asian | Wong et al. |
| Kamemoto | Japanese | Other Asian | Wong et al. |
| Kameoka | Japanese | Other Asian | Wong et al. |
| Kametani | Japanese | Other Asian | Wong et al. |
| Kameya | Japanese | Other Asian | Wong et al. |
| Kameyama | Japanese | Other Asian | Wong et al. |
| Kami | Japanese | Other Asian | Wong et al. |
| Kamibayashi | Japanese | Other Asian | Wong et al. |
| Kamihara | Japanese | Other Asian | Wong et al. |
| Kamijo | Japanese | Other Asian | Wong et al. |
| Kamikawa | Japanese | Other Asian | Wong et al. |
| Kamimoto | Japanese | Other Asian | Wong et al. |
| Kamimura | Japanese | Other Asian | Wong et al. |
| Kaminaga | Japanese | Other Asian | Wong et al. |
| Kamino | Japanese | Other Asian | Wong et al. |
| Kamio | Japanese | Other Asian | Wong et al. |
| Kamioka | Japanese | Other Asian | Wong et al. |
| Kamishima | Japanese | Other Asian | Wong et al. |
| Kamiya | Japanese | Other Asian | Wong et al. |
| Kamiyama | Japanese | Other Asian | Kranick et al. |
| Kamlani | Asian Indian | South Asian | Wong et al. |
| Kamo | Japanese | Other Asian | Wong et al. |
| Kamogawa | Japanese | Other Asian | Wong et al. |
| Kamoshida | Japanese | Other Asian | Wong et al. |
| Kampani | Asian Indian | South Asian | Wong et al. |
| Kamra | Asian Indian | South Asian | Wong et al. |
| Kamran | Pakistani | South Asian | Kranick et al. |
| Kamrujjaman |  | South Asian | Lim et al. |
| Kamrul |  | South Asian | Lim et al. |
| Kamruzzaman |  | South Asian | Lim et al. |
| Kamura | Japanese | Other Asian | Wong et al. |
| Kan | Chinese; Uzbekistan | Chinese | Kranick et al. |
| Kanada | Japanese | Other Asian | Wong et al. |
| Kanade | Asian Indian | South Asian | Wong et al. |
| Kanagawa | Japanese | Other Asian | Wong et al. |
| Kanai | Japanese | Other Asian | Wong et al. |
| Kanakubo | Japanese | Other Asian | Wong et al. |
| Kanal | Asian Indian | South Asian | Wong et al. |
| Kanamaru | Japanese | Other Asian | Kranick et al. |
| Kanamori | Japanese | Other Asian | Wong et al. |
| Kanamoto | Japanese | Other Asian | Wong et al. |
| Kanani | Asian Indian | South Asian | Wong et al. |
| Kanao | Japanese | Other Asian | Wong et al. |
| Kanaoka | Japanese | Other Asian | Wong et al. |
| Kanapi | Filipino | Other Asian | Wong et al. |
| Kanashiro | Japanese | Other Asian | Wong et al. |
| Kanaya | Japanese | Other Asian | Wong et al. |
| Kanayama | Japanese | Other Asian | Wong et al. |
| Kanazashi | Japanese | Other Asian | Wong et al. |
| Kanazawa | Japanese | Other Asian | Wong et al. |
| Kanbara | Japanese | Other Asian | Wong et al. |
| Kanbayashi | Japanese | Other Asian | Wong et al. |
| Kanbe | Japanese | Other Asian | Wong et al. |
| Kanda | Japanese | Other Asian | Wong et al. |
| Kandasamy | Asian Indian | South Asian | Wong et al. |
| Kandaswami | Asian Indian | South Asian | Wong et al. |
| Kandaswamy | Asian Indian | South Asian | Wong et al. |
| Kandathil | Asian Indian | South Asian | Wong et al. |
| Kanden |  | South Asian | Lim et al. |
| Kandhari | Asian Indian | South Asian | Wong et al. |
| Kandola | Asian Indian | South Asian | Wong et al. |
| Kaneda | Japanese | Other Asian | Wong et al. |
| Kanegae | Japanese | Other Asian | Wong et al. |
| Kanehira | Japanese | Other Asian | Wong et al. |
| Kaneishi | Japanese | Other Asian | Wong et al. |
| Kaneko | Japanese | Other Asian | Wong et al. |
| Kanemaru | Japanese | Other Asian | Wong et al. |
| Kanematsu | Japanese | Other Asian | Wong et al. |
| Kanemitsu | Japanese | Other Asian | Wong et al. |
| Kanemoto | Japanese | Other Asian | Wong et al. |
| Kanemura | Japanese | Other Asian | Wong et al. |
| Kaneria | Asian Indian | South Asian | Wong et al. |
| Kaneshige | Japanese | Other Asian | Wong et al. |
| Kaneshima | Japanese | Other Asian | Wong et al. |
| Kaneshiro | Japanese | Other Asian | Wong et al. |
| Kaneta | Japanese | Other Asian | Wong et al. |
| Kanetkar | Asian Indian | South Asian | Wong et al. |
| Kang | Chinese; Korean | Chinese | Kranick et al. |
| Kanga | Asian Indian | South Asian | Wong et al. |
| Kangleon | Filipino | Other Asian | Wong et al. |
| Kanh | Vietnamese | Other Asian | Wong et al. |
| Kani | Japanese | Other Asian | Wong et al. |
| Kaniz |  | South Asian | Lim et al. |
| Kanji | Asian Indian | South Asian | Wong et al. |
| Kanki | Japanese | Other Asian | Wong et al. |
| Kanna | Japanese | Other Asian | Wong et al. |
| Kanno | Japanese | Other Asian | Wong et al. |
| Kano | Japanese | Other Asian | Wong et al. |
| Kanoh | Japanese | Other Asian | Wong et al. |
| Kanojia | Asian Indian | South Asian | Wong et al. |
| Kansagara | Asian Indian | South Asian | Wong et al. |
| Kansal | Asian Indian | South Asian | Wong et al. |
| Kansara | Asian Indian | South Asian | Wong et al. |
| Kanta | Asian Indian; Bangladeshi | South Asian | Kranick et al. |
| Kantawala | Asian Indian | South Asian | Wong et al. |
| Kantere | Filipino | Other Asian | Wong et al. |
| Kantharia | Asian Indian | South Asian | Wong et al. |
| Kantilal | Asian Indian | South Asian | Wong et al. |
| Kanwal | Asian Indian | South Asian | Wong et al. |
| Kanwar | Asian Indian | South Asian | Wong et al. |
| Kanzaki | Japanese | Other Asian | Wong et al. |
| Kao | Chinese | Chinese | Wong et al. |
| Kapadia | Asian Indian | South Asian | Wong et al. |
| Kapasi | Asian Indian | South Asian | Wong et al. |
| Kapil | Asian Indian | South Asian | Wong et al. |
| Kapila | Asian Indian | South Asian | Wong et al. |
| Kapoor | Asian Indian | South Asian | Kranick et al. |
| Kapunan | Filipino | Other Asian | Wong et al. |
| Kapur | Asian Indian | South Asian | Wong et al. |
| Kapuria | Asian Indian | South Asian | Wong et al. |
| Kar | Asian Indian | South Asian | Wong et al. |
| Karaan | Filipino | Other Asian | Wong et al. |
| Karakas |  | South Asian | Lim et al. |
| Karamali | Asian Indian | South Asian | Wong et al. |
| Karandikar | Asian Indian | South Asian | Wong et al. |
| Karani | Asian Indian | South Asian | Wong et al. |
| Karanjia | Asian Indian | South Asian | Wong et al. |
| Karasawa | Japanese | Other Asian | Wong et al. |
| Karashima | Japanese | Other Asian | Wong et al. |
| Karatsu | Japanese | Other Asian | Wong et al. |
| Karchatsang |  | South Asian | Lim et al. |
| Kare | Filipino | Other Asian | Wong et al. |
| Kareem | Asian Indian | South Asian | Wong et al. |
| Karganilla | Filipino | Other Asian | Wong et al. |
| Karibe | Japanese | Other Asian | Wong et al. |
| Karim | Bangladeshi; Pakistani | South Asian | Kranick et al. |
| Karimata | Japanese | Other Asian | Wong et al. |
| Karimi | Asian Indian | South Asian | Wong et al. |
| Karino | Japanese | Other Asian | Wong et al. |
| Kariya | Japanese | Other Asian | Wong et al. |
| Karkhanis | Asian Indian | South Asian | Wong et al. |
| Karki |  | South Asian | Lim et al. |
| Karmali | Asian Indian | South Asian | Wong et al. |
| Karmarkar | Asian Indian | South Asian | Wong et al. |
| Karmoker |  | South Asian | Lim et al. |
| Karnad | Asian Indian | South Asian | Wong et al. |
| Karnati | Asian Indian | South Asian | Kranick et al. |
| Karnik | Asian Indian | South Asian | Wong et al. |
| Karpen |  | South Asian | Lim et al. |
| Karra | Asian Indian | South Asian | Wong et al. |
| Kartha | Asian Indian | South Asian | Wong et al. |
| Karube | Japanese | Other Asian | Wong et al. |
| Karunakaran | Asian Indian | South Asian | Wong et al. |
| Karve | Asian Indian | South Asian | Wong et al. |
| Karwa | Asian Indian | South Asian | Wong et al. |
| Kasagi | Japanese | Other Asian | Wong et al. |
| Kasahara | Japanese | Other Asian | Wong et al. |
| Kasai | Japanese | Other Asian | Wong et al. |
| Kasama | Japanese | Other Asian | Wong et al. |
| Kasamatsu | Japanese | Other Asian | Wong et al. |
| Kasaoka | Japanese | Other Asian | Wong et al. |
| Kasbekar | Asian Indian | South Asian | Wong et al. |
| Kasham |  | South Asian | Lim et al. |
| Kashem |  | South Asian | Lim et al. |
| Kashihara | Japanese | Other Asian | Wong et al. |
| Kashiki | Japanese | Other Asian | Wong et al. |
| Kashima | Japanese | Other Asian | Wong et al. |
| Kashimoto | Japanese | Other Asian | Wong et al. |
| Kashio | Japanese | Other Asian | Wong et al. |
| Kashiwa | Japanese | Other Asian | Wong et al. |
| Kashiwabara | Japanese | Other Asian | Wong et al. |
| Kashiwagi | Japanese | Other Asian | Wong et al. |
| Kashiwamura | Japanese | Other Asian | Wong et al. |
| Kashiwaya | Japanese | Other Asian | Wong et al. |
| Kashiyama | Japanese | Other Asian | Wong et al. |
| Kashyap | Asian Indian | South Asian | Wong et al. |
| Kasi | Asian Indian | South Asian | Wong et al. |
| Kasilag | Filipino | Other Asian | Wong et al. |
| Kasim | Asian Indian | South Asian | Wong et al. |
| Kasinath |  | South Asian | Lim et al. |
| Kassam | Asian Indian | South Asian | Wong et al. |
| Kassim |  | South Asian | Lim et al. |
| Kasturi | Asian Indian | South Asian | Wong et al. |
| Kasuga | Japanese | Other Asian | Wong et al. |
| Kasugai | Japanese | Other Asian | Wong et al. |
| Kasukawa | Japanese | Other Asian | Wong et al. |
| Kasuya | Japanese | Other Asian | Wong et al. |
| Kat | Chinese | Chinese | Wong et al. |
| Katada | Japanese | Other Asian | Wong et al. |
| Katagiri | Japanese | Other Asian | Wong et al. |
| Katahira | Japanese | Other Asian | Wong et al. |
| Katakura | Japanese | Other Asian | Wong et al. |
| Katano | Japanese | Other Asian | Wong et al. |
| Kataoka | Japanese | Other Asian | Wong et al. |
| Katayama | Japanese | Other Asian | Wong et al. |
| Kathawala | Asian Indian | South Asian | Wong et al. |
| Kathiria | Asian Indian | South Asian | Wong et al. |
| Kathrotia | Asian Indian | South Asian | Wong et al. |
| Kathuria | Asian Indian | South Asian | Wong et al. |
| Katigbak | Filipino | Other Asian | Wong et al. |
| Katindig | Filipino | Other Asian | Wong et al. |
| Katipunan | Filipino | Other Asian | Wong et al. |
| Katiyar | Asian Indian | South Asian | Wong et al. |
| Kato | Japanese | Other Asian | Wong et al. |
| Katoh | Japanese | Other Asian | Wong et al. |
| Katori | Japanese | Other Asian | Wong et al. |
| Katow | Japanese | Other Asian | Wong et al. |
| Katragadda | Asian Indian | South Asian | Wong et al. |
| Katsu | Japanese | Other Asian | Wong et al. |
| Katsube | Japanese | Other Asian | Wong et al. |
| Katsuda | Japanese | Other Asian | Wong et al. |
| Katsuki | Japanese | Other Asian | Wong et al. |
| Katsuma | Japanese | Other Asian | Wong et al. |
| Katsumata | Japanese | Other Asian | Wong et al. |
| Katsumi | Japanese | Other Asian | Wong et al. |
| Katsumori | Japanese | Other Asian | Wong et al. |
| Katsumoto | Japanese | Other Asian | Wong et al. |
| Katsumura | Japanese | Other Asian | Wong et al. |
| Katsuno | Japanese | Other Asian | Wong et al. |
| Katsunuma | Japanese | Other Asian | Wong et al. |
| Katsura | Japanese | Other Asian | Wong et al. |
| Katsuragi | Japanese | Other Asian | Wong et al. |
| Katsuren | Japanese | Other Asian | Wong et al. |
| Katsushima | Japanese | Other Asian | Wong et al. |
| Katsuta | Japanese | Other Asian | Wong et al. |
| Katsuura | Japanese | Other Asian | Wong et al. |
| Katsuya | Japanese | Other Asian | Wong et al. |
| Katsuyama | Japanese | Other Asian | Wong et al. |
| Katta | Asian Indian | South Asian | Wong et al. |
| Katwala | Asian Indian | South Asian | Wong et al. |
| Katwaru |  | South Asian | Lim et al. |
| Katyal | Asian Indian | South Asian | Wong et al. |
| Kau | Chinese | Chinese | Wong et al. |
| Kauh | Korean | Other Asian | Wong et al. |
| Kaung | Chinese | Chinese | Wong et al. |
| Kaur | Asian Indian | South Asian | Kranick et al. |
| Kaura | Asian Indian | South Asian | Wong et al. |
| Kausar |  | South Asian | Lim et al. |
| Kaushal | Asian Indian | South Asian | Wong et al. |
| Kaushik | Asian Indian | South Asian | Wong et al. |
| Kavan | Vietnamese | Other Asian | Wong et al. |
| Kawabata | Japanese | Other Asian | Wong et al. |
| Kawabe | Japanese | Other Asian | Wong et al. |
| Kawabuchi | Japanese | Other Asian | Wong et al. |
| Kawachi | Japanese | Other Asian | Wong et al. |
| Kawada | Japanese | Other Asian | Wong et al. |
| Kawade | Japanese | Other Asian | Wong et al. |
| Kawae | Japanese | Other Asian | Wong et al. |
| Kawagishi | Japanese | Other Asian | Wong et al. |
| Kawagoe | Japanese | Other Asian | Wong et al. |
| Kawaguchi | Japanese | Other Asian | Wong et al. |
| Kawahara | Japanese | Other Asian | Wong et al. |
| Kawaharada | Japanese | Other Asian | Wong et al. |
| Kawahata | Japanese | Other Asian | Wong et al. |
| Kawahira | Japanese | Other Asian | Wong et al. |
| Kawai | Japanese | Other Asian | Wong et al. |
| Kawaji | Japanese | Other Asian | Wong et al. |
| Kawajiri | Japanese | Other Asian | Wong et al. |
| Kawakami | Japanese | Other Asian | Wong et al. |
| Kawakatsu | Japanese | Other Asian | Wong et al. |
| Kawakita | Japanese | Other Asian | Wong et al. |
| Kawakubo | Japanese | Other Asian | Wong et al. |
| Kawamata | Japanese | Other Asian | Wong et al. |
| Kawamitsu | Japanese | Other Asian | Wong et al. |
| Kawamoto | Japanese | Other Asian | Wong et al. |
| Kawamura | Japanese | Other Asian | Wong et al. |
| Kawana | Japanese | Other Asian | Wong et al. |
| Kawanabe | Japanese | Other Asian | Wong et al. |
| Kawanaka | Japanese | Other Asian | Wong et al. |
| Kawanami | Japanese | Other Asian | Wong et al. |
| Kawane | Japanese | Other Asian | Wong et al. |
| Kawanishi | Japanese | Other Asian | Wong et al. |
| Kawano | Japanese | Other Asian | Wong et al. |
| Kawaoka | Japanese | Other Asian | Wong et al. |
| Kawarada | Japanese | Other Asian | Wong et al. |
| Kawaragi | Japanese | Other Asian | Wong et al. |
| Kawasaki | Japanese | Other Asian | Wong et al. |
| Kawase | Japanese | Other Asian | Wong et al. |
| Kawashima | Japanese | Other Asian | Wong et al. |
| Kawasumi | Japanese | Other Asian | Wong et al. |
| Kawata | Japanese | Other Asian | Wong et al. |
| Kawatani | Japanese | Other Asian | Wong et al. |
| Kawate | Japanese | Other Asian | Wong et al. |
| Kawato | Japanese | Other Asian | Wong et al. |
| Kawatsu | Japanese | Other Asian | Wong et al. |
| Kawauchi | Japanese | Other Asian | Wong et al. |
| Kawazoe | Japanese | Other Asian | Wong et al. |
| Kawazu | Japanese | Other Asian | Wong et al. |
| Kawcher |  | South Asian | Lim et al. |
| Kawlra | Asian Indian | South Asian | Wong et al. |
| Kawn | Chinese | Chinese | Wong et al. |
| Kawsar |  | South Asian | Lim et al. |
| Kawser |  | South Asian | Lim et al. |
| Kaya | Japanese | Other Asian | Wong et al. |
| Kayaba | Japanese | Other Asian | Wong et al. |
| Kayaban | Filipino | Other Asian | Wong et al. |
| Kayahara | Japanese | Other Asian | Wong et al. |
| Kayama | Japanese | Other Asian | Wong et al. |
| Kayano | Japanese | Other Asian | Wong et al. |
| Kayas | Filipino | Other Asian | Wong et al. |
| Kayashima | Japanese | Other Asian | Wong et al. |
| Kayastha | Asian Indian | South Asian | Wong et al. |
| Kayo | Japanese | Other Asian | Wong et al. |
| Kazama | Japanese | Other Asian | Wong et al. |
| Kazi | Bangladeshi | South Asian | Kranick et al. |
| Kazim | Asian Indian | South Asian | Wong et al. |
| Kazmi | Asian Indian | South Asian | Wong et al. |
| Kazumi | Japanese | Other Asian | Wong et al. |
| Kazuo | Japanese | Other Asian | Wong et al. |
| Kc |  | South Asian | Lim et al. |
| Ke | Chinese | Chinese | Wong et al. |
| Kedia | Asian Indian | South Asian | Wong et al. |
| Kee | Chinese | Chinese | Wong et al. |
| Keh | Chinese | Chinese | Wong et al. |
| Kei | Chinese | Chinese | Wong et al. |
| Kelkar | Asian Indian | South Asian | Wong et al. |
| Kempis | Filipino | Other Asian | Wong et al. |
| Ken | Chinese | Chinese | Wong et al. |
| Keng | Chinese | Chinese | Wong et al. |
| Kenjo | Japanese | Other Asian | Wong et al. |
| Kenmotsu | Japanese | Other Asian | Wong et al. |
| Keong | Chinese | Chinese | Wong et al. |
| Kermani | Asian Indian | South Asian | Wong et al. |
| Kerr |  | South Asian | Lim et al. |
| Kesavan | Asian Indian | South Asian | Wong et al. |
| Keshavlal | Asian Indian | South Asian | Wong et al. |
| Ketkar | Asian Indian | South Asian | Wong et al. |
| Keu | Chinese | Chinese | Wong et al. |
| Keum | Korean | Other Asian | Wong et al. |
| Keun | Korean | Other Asian | Wong et al. |
| Keung | Chinese | Chinese | Wong et al. |
| Kew | Chinese | Chinese | Wong et al. |
| Kezuka | Japanese | Other Asian | Wong et al. |
| Khac | Vietnamese | Other Asian | Wong et al. |
| Khader | Asian Indian | South Asian | Wong et al. |
| Khadilkar | Asian Indian | South Asian | Wong et al. |
| Khadka |  | South Asian | Lim et al. |
| Khai | Vietnamese | Other Asian | Wong et al. |
| Khair | Bangladeshi | South Asian | Kranick et al. |
| Khaira | Asian Indian | South Asian | Wong et al. |
| Khalasi | Asian Indian | South Asian | Wong et al. |
| Khaleel | Asian Indian | South Asian | Wong et al. |
| Khaleque |  | South Asian | Lim et al. |
| Khalequzzaman |  | South Asian | Lim et al. |
| Khalid |  | South Asian | Lim et al. |
| Khalifa | Asian Indian | South Asian | Wong et al. |
| Khalil | Asian Indian | South Asian | Wong et al. |
| Khaloo |  | South Asian | Lim et al. |
| Khalsa | Asian Indian | South Asian | Wong et al. |
| Khamba | Asian Indian | South Asian | Wong et al. |
| Khambanonda | Thai | Other Asian | Kranick et al. |
| Khambata | Asian Indian | South Asian | Wong et al. |
| Khambatta | Asian Indian | South Asian | Wong et al. |
| Khambaty | Asian Indian | South Asian | Wong et al. |
| Khan | Pakistani; Bangladeshi | South Asian | Kranick et al. |
| Khana | Asian Indian | South Asian | Wong et al. |
| Khanai |  | South Asian | Lim et al. |
| Khanam |  | South Asian | Lim et al. |
| Khanani | Asian Indian | South Asian | Wong et al. |
| Khandaker | Bangladeshi | South Asian | Kranick et al. |
| Khandekar | Asian Indian | South Asian | Wong et al. |
| Khandelwal | Asian Indian | South Asian | Wong et al. |
| Khanderia | Asian Indian | South Asian | Wong et al. |
| Khandhar | Asian Indian | South Asian | Wong et al. |
| Khandkar | Asian Indian | South Asian | Wong et al. |
| Khandokar |  | South Asian | Lim et al. |
| Khandoker | Bangladeshi | South Asian | Kranick et al. |
| Khandpur | Asian Indian | South Asian | Wong et al. |
| Khandwala | Asian Indian | South Asian | Wong et al. |
| Khandwalla | Asian Indian | South Asian | Wong et al. |
| Khangura | Asian Indian | South Asian | Wong et al. |
| Khanh | Vietnamese | Other Asian | Wong et al. |
| Khanm |  | South Asian | Lim et al. |
| Khanmohammad |  | South Asian | Lim et al. |
| Khanna | Asian Indian | South Asian | Wong et al. |
| Khanom |  | South Asian | Lim et al. |
| Khanoom |  | South Asian | Lim et al. |
| Khanum | Pakistani | South Asian | Kranick et al. |
| Khanzada | Asian Indian | South Asian | Wong et al. |
| Khara | Asian Indian | South Asian | Wong et al. |
| Kharanja |  | South Asian | Lim et al. |
| Kharbanda | Asian Indian | South Asian | Wong et al. |
| Khare | Asian Indian | South Asian | Wong et al. |
| Khargie |  | South Asian | Lim et al. |
| Kharkar | Asian Indian | South Asian | Wong et al. |
| Kharm |  | South Asian | Lim et al. |
| Kharod | Asian Indian | South Asian | Wong et al. |
| Khasru |  | South Asian | Lim et al. |
| Khatib | Asian Indian | South Asian | Wong et al. |
| Khatiwala | Asian Indian | South Asian | Wong et al. |
| Khatkhate | Asian Indian | South Asian | Wong et al. |
| Khaton |  | South Asian | Lim et al. |
| Khatoon | Asian Indian | South Asian | Kranick et al. |
| Khatra | Asian Indian | South Asian | Wong et al. |
| Khatri | Asian Indian | South Asian | Wong et al. |
| Khatun | Asian Indian | South Asian | Kranick et al. |
| Khawaja | Pakistani | South Asian | Kranick et al. |
| Khay | Chinese | Chinese | Wong et al. |
| Khela | Asian Indian | South Asian | Wong et al. |
| Khem |  | South Asian | Lim et al. |
| Khemchandani | Asian Indian | South Asian | Wong et al. |
| Khemka | Asian Indian | South Asian | Wong et al. |
| Khemraj |  | South Asian | Lim et al. |
| Kher | Asian Indian | South Asian | Wong et al. |
| Khera | Asian Indian | South Asian | Wong et al. |
| Kherani | Asian Indian | South Asian | Wong et al. |
| Khetani | Asian Indian | South Asian | Wong et al. |
| Khetarpal | Asian Indian | South Asian | Wong et al. |
| Khi | Vietnamese | Other Asian | Wong et al. |
| Khiam | Chinese | Chinese | Wong et al. |
| Khiem | Vietnamese | Other Asian | Wong et al. |
| Khiet | Vietnamese | Other Asian | Wong et al. |
| Khieu | Vietnamese | Other Asian | Wong et al. |
| Khilji |  | South Asian | Lim et al. |
| Khimani | Asian Indian | South Asian | Wong et al. |
| Khimji | Asian Indian | South Asian | Wong et al. |
| Khoa | Vietnamese | Other Asian | Wong et al. |
| Khoai | Vietnamese | Other Asian | Wong et al. |
| Khoan | Vietnamese | Other Asian | Wong et al. |
| Khoda |  | South Asian | Lim et al. |
| Khodadad |  | South Asian | Lim et al. |
| Khode | Asian Indian | South Asian | Wong et al. |
| Khoi | Vietnamese | Other Asian | Wong et al. |
| Khoja | Asian Indian | South Asian | Wong et al. |
| Khokhar |  | South Asian | Lim et al. |
| Khokon |  | South Asian | Lim et al. |
| Khona | Asian Indian | South Asian | Wong et al. |
| Khondaker |  | South Asian | Lim et al. |
| Khondker |  | South Asian | Lim et al. |
| Khondokar |  | South Asian | Lim et al. |
| Khondoker |  | South Asian | Lim et al. |
| Khong | Vietnamese | Other Asian | Wong et al. |
| Khorshed |  | South Asian | Lim et al. |
| Khosa | Asian Indian | South Asian | Wong et al. |
| Khosla | Asian Indian | South Asian | Wong et al. |
| Khot | Asian Indian | South Asian | Wong et al. |
| Khuat | Vietnamese | Other Asian | Wong et al. |
| Khubchandani | Asian Indian | South Asian | Wong et al. |
| Khuc | Vietnamese | Other Asian | Wong et al. |
| Khue | Vietnamese | Other Asian | Wong et al. |
| Khuki |  | South Asian | Lim et al. |
| Khullar | Asian Indian | South Asian | Wong et al. |
| Khung | Vietnamese | Other Asian | Wong et al. |
| Khunkhun | Asian Indian | South Asian | Wong et al. |
| Khuong | Vietnamese | Other Asian | Wong et al. |
| Khurshed |  | South Asian | Lim et al. |
| Khuu | Vietnamese | Other Asian | Kranick et al. |
| Khuyen | Vietnamese | Other Asian | Wong et al. |
| Ki | Korean | Other Asian | Kranick et al. |
| Kiamco | Filipino | Other Asian | Wong et al. |
| Kiang | Chinese | Chinese | Wong et al. |
| Kiani |  | South Asian | Lim et al. |
| Kiao | Chinese | Chinese | Wong et al. |
| Kiba | Japanese | Other Asian | Wong et al. |
| Kibe | Japanese | Other Asian | Wong et al. |
| Kibria |  | South Asian | Lim et al. |
| Kida | Japanese | Other Asian | Wong et al. |
| Kidani | Japanese | Other Asian | Wong et al. |
| Kido | Japanese | Other Asian | Wong et al. |
| Kidokoro | Japanese | Other Asian | Wong et al. |
| Kidwai | Asian Indian | South Asian | Wong et al. |
| Kiem | Vietnamese | Other Asian | Wong et al. |
| Kien | Vietnamese | Other Asian | Wong et al. |
| Kierulf | Filipino | Other Asian | Wong et al. |
| Kiet | Vietnamese | Other Asian | Wong et al. |
| Kieu | Vietnamese | Other Asian | Wong et al. |
| Kiew | Chinese | Chinese | Wong et al. |
| Kigawa | Japanese | Other Asian | Wong et al. |
| Kigoshi | Japanese | Other Asian | Wong et al. |
| Kiguchi | Japanese | Other Asian | Wong et al. |
| Kihara | Japanese | Other Asian | Wong et al. |
| Kii | Japanese | Other Asian | Wong et al. |
| Kijima | Japanese | Other Asian | Wong et al. |
| Kikani | Asian Indian | South Asian | Wong et al. |
| Kikkawa | Japanese | Other Asian | Wong et al. |
| Kikuchi | Japanese | Other Asian | Wong et al. |
| Kikugawa | Japanese | Other Asian | Wong et al. |
| Kikuma | Japanese | Other Asian | Wong et al. |
| Kikumoto | Japanese | Other Asian | Wong et al. |
| Kikunaga | Japanese | Other Asian | Wong et al. |
| Kikuno | Japanese | Other Asian | Wong et al. |
| Kikuoka | Japanese | Other Asian | Wong et al. |
| Kikushima | Japanese | Other Asian | Wong et al. |
| Kikuta | Japanese | Other Asian | Wong et al. |
| Kikutani | Japanese | Other Asian | Wong et al. |
| Kikuya | Japanese | Other Asian | Wong et al. |
| Kil | Korean | Other Asian | Wong et al. |
| Kilachand | Asian Indian | South Asian | Wong et al. |
| Kilala | Filipino | Other Asian | Wong et al. |
| Kilayko | Filipino | Other Asian | Wong et al. |
| Kim | Korean | Other Asian | Kranick et al. |
| Kimata | Japanese | Other Asian | Wong et al. |
| Kimizuka | Japanese | Other Asian | Wong et al. |
| Kimm | Korean | Other Asian | Wong et al. |
| Kimoto | Japanese | Other Asian | Wong et al. |
| Kimpo | Filipino | Other Asian | Wong et al. |
| Kimura | Japanese | Other Asian | Wong et al. |
| Kin | Chinese | Chinese | Wong et al. |
| Kina | Japanese | Other Asian | Wong et al. |
| Kinariwala | Asian Indian | South Asian | Wong et al. |
| Kingra | Asian Indian | South Asian | Wong et al. |
| Kinh | Vietnamese | Other Asian | Wong et al. |
| Kini | Asian Indian | South Asian | Wong et al. |
| Kinjo | Japanese | Other Asian | Wong et al. |
| Kino | Japanese | Other Asian | Wong et al. |
| Kinoshita | Japanese | Other Asian | Wong et al. |
| Kintanar | Filipino | Other Asian | Wong et al. |
| Kinugasa | Japanese | Other Asian | Wong et al. |
| Kinugawa | Japanese | Other Asian | Wong et al. |
| Kiocho | Filipino | Other Asian | Wong et al. |
| Kira | Japanese | Other Asian | Wong et al. |
| Kirihara | Japanese | Other Asian | Wong et al. |
| Kirisawa | Japanese | Other Asian | Wong et al. |
| Kirishima | Japanese | Other Asian | Wong et al. |
| Kirita | Japanese | Other Asian | Wong et al. |
| Kiritani | Japanese | Other Asian | Wong et al. |
| Kiriu | Japanese | Other Asian | Wong et al. |
| Kiriyama | Japanese | Other Asian | Wong et al. |
| Kirmani | Asian Indian | South Asian | Wong et al. |
| Kishaba | Japanese | Other Asian | Wong et al. |
| Kishi | Japanese | Other Asian | Wong et al. |
| Kishida | Japanese | Other Asian | Wong et al. |
| Kishii | Japanese | Other Asian | Wong et al. |
| Kishikawa | Japanese | Other Asian | Wong et al. |
| Kishima | Japanese | Other Asian | Wong et al. |
| Kishimoto | Japanese | Other Asian | Wong et al. |
| Kishiyama | Japanese | Other Asian | Wong et al. |
| Kishore | Asian Indian | South Asian | Wong et al. |
| Kiso | Japanese | Other Asian | Wong et al. |
| Kisoon |  | South Asian | Lim et al. |
| Kisson |  | South Asian | Lim et al. |
| Kissoom |  | South Asian | Lim et al. |
| Kissoon | Asian Indian | South Asian | Kranick et al. |
| Kisu | Japanese | Other Asian | Wong et al. |
| Kit | Chinese | Chinese | Wong et al. |
| Kita | Japanese | Other Asian | Wong et al. |
| Kitabatake | Japanese | Other Asian | Wong et al. |
| Kitabayashi | Japanese | Other Asian | Wong et al. |
| Kitada | Japanese | Other Asian | Wong et al. |
| Kitagawa | Japanese | Other Asian | Wong et al. |
| Kitaguchi | Japanese | Other Asian | Wong et al. |
| Kitahara | Japanese | Other Asian | Wong et al. |
| Kitahata | Japanese | Other Asian | Wong et al. |
| Kitai | Japanese | Other Asian | Wong et al. |
| Kitajima | Japanese | Other Asian | Wong et al. |
| Kitami | Japanese | Other Asian | Wong et al. |
| Kitamori | Japanese | Other Asian | Wong et al. |
| Kitamoto | Japanese | Other Asian | Wong et al. |
| Kitamura | Japanese | Other Asian | Wong et al. |
| Kitani | Japanese | Other Asian | Wong et al. |
| Kitano | Japanese | Other Asian | Wong et al. |
| Kitao | Japanese | Other Asian | Wong et al. |
| Kitaoka | Japanese | Other Asian | Wong et al. |
| Kitashima | Japanese | Other Asian | Wong et al. |
| Kitatani | Japanese | Other Asian | Wong et al. |
| Kitaura | Japanese | Other Asian | Wong et al. |
| Kitayama | Japanese | Other Asian | Wong et al. |
| Kitazato | Japanese | Other Asian | Wong et al. |
| Kitazawa | Japanese | Other Asian | Wong et al. |
| Kitazume | Japanese | Other Asian | Wong et al. |
| Kitazumi | Japanese | Other Asian | Wong et al. |
| Kito | Japanese | Other Asian | Kranick et al. |
| Kittaka | Japanese | Other Asian | Wong et al. |
| Kiu | Chinese | Chinese | Wong et al. |
| Kiuchi | Japanese | Other Asian | Wong et al. |
| Kiya | Japanese | Other Asian | Wong et al. |
| Kiyabu | Japanese | Other Asian | Wong et al. |
| Kiyama | Japanese | Other Asian | Wong et al. |
| Kiyan | Japanese | Other Asian | Wong et al. |
| Kiyohara | Japanese | Other Asian | Wong et al. |
| Kiyokawa | Japanese | Other Asian | Wong et al. |
| Kiyonaga | Japanese | Other Asian | Wong et al. |
| Kiyono | Japanese | Other Asian | Wong et al. |
| Kiyooka | Japanese | Other Asian | Wong et al. |
| Kiyosawa | Japanese | Other Asian | Wong et al. |
| Kiyoshi | Japanese | Other Asian | Wong et al. |
| Kiyota | Japanese | Other Asian | Wong et al. |
| Kiyuna | Japanese | Other Asian | Wong et al. |
| Kizaki | Japanese | Other Asian | Wong et al. |
| Kizu | Japanese | Other Asian | Wong et al. |
| Kliatchko | Filipino | Other Asian | Wong et al. |
| Ko | Korean; Burmese | Other Asian | Kranick et al. |
| Koa | Chinese | Chinese | Wong et al. |
| Koan | Chinese | Chinese | Wong et al. |
| Koba | Japanese | Other Asian | Wong et al. |
| Kobari | Japanese | Other Asian | Wong et al. |
| Kobashi | Japanese | Other Asian | Wong et al. |
| Kobashigawa | Japanese | Other Asian | Wong et al. |
| Kobata | Japanese | Other Asian | Wong et al. |
| Kobatake | Japanese | Other Asian | Wong et al. |
| Kobayakawa | Japanese | Other Asian | Wong et al. |
| Kobayashi | Japanese | Other Asian | Wong et al. |
| Kobayaski | Japanese | Other Asian | Wong et al. |
| Kobayshi | Japanese | Other Asian | Wong et al. |
| Kobori | Japanese | Other Asian | Wong et al. |
| Kobuchi | Japanese | Other Asian | Wong et al. |
| Kobuke | Japanese | Other Asian | Wong et al. |
| Koc | Chinese | Chinese | Wong et al. |
| Kochar | Asian Indian | South Asian | Wong et al. |
| Kochhar | Asian Indian | South Asian | Wong et al. |
| Kochi | Japanese | Other Asian | Wong et al. |
| Kochukunju | Asian Indian | South Asian | Wong et al. |
| Kochuparampi | Asian Indian | South Asian | Wong et al. |
| Koda | Japanese | Other Asian | Wong et al. |
| Kodaira | Japanese | Other Asian | Wong et al. |
| Kodaka | Japanese | Other Asian | Wong et al. |
| Kodali | Asian Indian | South Asian | Wong et al. |
| Kodama | Japanese | Other Asian | Wong et al. |
| Kodani | Japanese | Other Asian | Wong et al. |
| Kodera | Japanese | Other Asian | Wong et al. |
| Koga | Japanese | Other Asian | Wong et al. |
| Kogima | Japanese | Other Asian | Wong et al. |
| Kogo | Japanese | Other Asian | Wong et al. |
| Koguchi | Japanese | Other Asian | Wong et al. |
| Kogure | Japanese | Other Asian | Wong et al. |
| Koh | Korean; Chinese | Other Asian | Kranick et al. |
| Kohagura | Japanese | Other Asian | Wong et al. |
| Kohama | Japanese | Other Asian | Wong et al. |
| Kohara | Japanese | Other Asian | Wong et al. |
| Kohashi | Japanese | Other Asian | Wong et al. |
| Kohatsu | Japanese | Other Asian | Wong et al. |
| Kohli | Asian Indian | South Asian | Kranick et al. |
| Kohno | Japanese | Other Asian | Wong et al. |
| Kohra | Japanese | Other Asian | Wong et al. |
| Kohtiao | Filipino | Other Asian | Wong et al. |
| Kohyama | Japanese | Other Asian | Wong et al. |
| Koichi | Japanese | Other Asian | Wong et al. |
| Koide | Japanese | Other Asian | Wong et al. |
| Koike | Japanese | Other Asian | Wong et al. |
| Koinuma | Japanese | Other Asian | Wong et al. |
| Koishi | Japanese | Other Asian | Wong et al. |
| Koiso | Japanese | Other Asian | Wong et al. |
| Koizumi | Japanese | Other Asian | Wong et al. |
| Koja | Japanese | Other Asian | Wong et al. |
| Kojima | Japanese | Other Asian | Wong et al. |
| Kok | Chinese | Chinese | Wong et al. |
| Kokubo | Japanese | Other Asian | Wong et al. |
| Kokubu | Japanese | Other Asian | Wong et al. |
| Kokubun | Japanese | Other Asian | Wong et al. |
| Kolhatkar | Asian Indian | South Asian | Wong et al. |
| Kolla | Asian Indian | South Asian | Wong et al. |
| Kolli | Asian Indian | South Asian | Wong et al. |
| Kollipara | Asian Indian | South Asian | Wong et al. |
| Kolluri | Asian Indian | South Asian | Wong et al. |
| Komai | Japanese | Other Asian | Wong et al. |
| Komaki | Japanese | Other Asian | Wong et al. |
| Komata | Japanese | Other Asian | Wong et al. |
| Komatsu | Japanese | Other Asian | Wong et al. |
| Komatsubara | Japanese | Other Asian | Wong et al. |
| Komatsuzaki | Japanese | Other Asian | Wong et al. |
| Komazawa | Japanese | Other Asian | Wong et al. |
| Komesu | Japanese | Other Asian | Wong et al. |
| Kometani | Japanese | Other Asian | Wong et al. |
| Kominami | Japanese | Other Asian | Wong et al. |
| Komine | Japanese | Other Asian | Wong et al. |
| Komiya | Japanese | Other Asian | Wong et al. |
| Komiyama | Japanese | Other Asian | Wong et al. |
| Komoda | Japanese | Other Asian | Wong et al. |
| Komori | Japanese | Other Asian | Wong et al. |
| Komorita | Japanese | Other Asian | Wong et al. |
| Komoriya | Japanese | Other Asian | Wong et al. |
| Komoto | Japanese | Other Asian | Wong et al. |
| Komukai | Japanese | Other Asian | Wong et al. |
| Komura | Japanese | Other Asian | Wong et al. |
| Komuro | Japanese | Other Asian | Wong et al. |
| Konaka | Japanese | Other Asian | Wong et al. |
| Konda | Japanese | Other Asian | Wong et al. |
| Kondo | Japanese | Other Asian | Wong et al. |
| Kondoh | Japanese | Other Asian | Wong et al. |
| Kong | Chinese | Chinese | Kranick et al. |
| Konishi | Japanese | Other Asian | Wong et al. |
| Konno | Japanese | Other Asian | Wong et al. |
| Kono | Japanese | Other Asian | Wong et al. |
| Konoki | Japanese | Other Asian | Wong et al. |
| Koo | Chinese; Korean | Chinese | Kranick et al. |
| Kooner | Asian Indian | South Asian | Wong et al. |
| Koong | Chinese | Chinese | Wong et al. |
| Koongie |  | South Asian | Lim et al. |
| Koppikar | Asian Indian | South Asian | Wong et al. |
| Kora | Asian Indian | South Asian | Wong et al. |
| Koradia | Asian Indian | South Asian | Wong et al. |
| Korah | Asian Indian | South Asian | Wong et al. |
| Korde | Asian Indian | South Asian | Wong et al. |
| Kori | Japanese | Other Asian | Wong et al. |
| Koriyama | Japanese | Other Asian | Wong et al. |
| Koruthu | Asian Indian | South Asian | Wong et al. |
| Kosai | Japanese | Other Asian | Wong et al. |
| Kosaka | Japanese | Other Asian | Wong et al. |
| Kosaki | Japanese | Other Asian | Wong et al. |
| Kosar |  | South Asian | Lim et al. |
| Kosaraju | Asian Indian | South Asian | Wong et al. |
| Kose | Japanese | Other Asian | Wong et al. |
| Koseki | Japanese | Other Asian | Wong et al. |
| Koser |  | South Asian | Lim et al. |
| Koshi | Japanese | Other Asian | Wong et al. |
| Koshiba | Japanese | Other Asian | Wong et al. |
| Koshida | Japanese | Other Asian | Wong et al. |
| Koshiishi | Japanese | Other Asian | Wong et al. |
| Koshikawa | Japanese | Other Asian | Wong et al. |
| Koshimizu | Japanese | Other Asian | Wong et al. |
| Koshino | Japanese | Other Asian | Wong et al. |
| Koshiro | Japanese | Other Asian | Wong et al. |
| Koshiyama | Japanese | Other Asian | Wong et al. |
| Koshy | Asian Indian | South Asian | Kranick et al. |
| Kosuga | Japanese | Other Asian | Wong et al. |
| Kosuge | Japanese | Other Asian | Wong et al. |
| Kosugi | Japanese | Other Asian | Wong et al. |
| Kotadia | Asian Indian | South Asian | Wong et al. |
| Kotak | Asian Indian | South Asian | Wong et al. |
| Kotaka | Japanese | Other Asian | Wong et al. |
| Kotake | Japanese | Other Asian | Wong et al. |
| Kotaki | Japanese | Other Asian | Wong et al. |
| Kotani | Japanese | Other Asian | Wong et al. |
| Kotecha | Asian Indian | South Asian | Wong et al. |
| Kothandarama | Asian Indian | South Asian | Wong et al. |
| Kothari | Asian Indian | South Asian | Wong et al. |
| Kothawala | Asian Indian | South Asian | Wong et al. |
| Kotiah |  | South Asian | Lim et al. |
| Kotima | Japanese | Other Asian | Wong et al. |
| Koto | Japanese | Other Asian | Wong et al. |
| Kotsubo | Japanese | Other Asian | Wong et al. |
| Kottarathil | Asian Indian | South Asian | Wong et al. |
| Kotwal | Asian Indian | South Asian | Wong et al. |
| Kotwani | Asian Indian | South Asian | Wong et al. |
| Kou | Chinese | Chinese | Wong et al. |
| Kouchi | Japanese | Other Asian | Wong et al. |
| Kouda | Japanese | Other Asian | Wong et al. |
| Koun | Chinese | Chinese | Wong et al. |
| Koung | Chinese | Chinese | Wong et al. |
| Kouno | Japanese | Other Asian | Wong et al. |
| Kouo | Vietnamese | Other Asian | Wong et al. |
| Koura | Japanese | Other Asian | Wong et al. |
| Kousar |  | South Asian | Lim et al. |
| Kow | Chinese | Chinese | Wong et al. |
| Kowk | Chinese | Chinese | Wong et al. |
| Kowlessar |  | South Asian | Lim et al. |
| Kown | Korean | Other Asian | Wong et al. |
| Kowng | Chinese | Chinese | Wong et al. |
| Koya | Japanese | Other Asian | Wong et al. |
| Koyama | Japanese | Other Asian | Wong et al. |
| Koyanagi | Japanese | Other Asian | Kranick et al. |
| Koyano | Japanese | Other Asian | Wong et al. |
| Koyasu | Japanese | Other Asian | Wong et al. |
| Kozai | Japanese | Other Asian | Wong et al. |
| Kozaki | Japanese | Other Asian | Wong et al. |
| Kozakura | Japanese | Other Asian | Wong et al. |
| Kozawa | Japanese | Other Asian | Wong et al. |
| Kozen | Japanese | Other Asian | Wong et al. |
| Kozono | Japanese | Other Asian | Wong et al. |
| Kozu | Japanese | Other Asian | Wong et al. |
| Kozuka | Japanese | Other Asian | Wong et al. |
| Kozuma | Japanese | Other Asian | Wong et al. |
| Kraskov | Chinese | Chinese | Wong et al. |
| Krishna | Asian Indian | South Asian | Wong et al. |
| Krishnaiah | Asian Indian | South Asian | Wong et al. |
| Krishnaiyeng | Asian Indian | South Asian | Wong et al. |
| Krishnaiyer | Asian Indian | South Asian | Wong et al. |
| Krishnamacha | Asian Indian | South Asian | Wong et al. |
| Krishnamoort | Asian Indian | South Asian | Wong et al. |
| Krishnamurt | Asian Indian | South Asian | Wong et al. |
| Krishnamurth | Asian Indian | South Asian | Wong et al. |
| Krishnamurti | Asian Indian | South Asian | Wong et al. |
| Krishnamurty | Asian Indian | South Asian | Wong et al. |
| Krishnan | Asian Indian | South Asian | Wong et al. |
| Krishnappa | Asian Indian | South Asian | Wong et al. |
| Krishnarao | Asian Indian | South Asian | Wong et al. |
| Krishnasamy | Asian Indian | South Asian | Wong et al. |
| Krishnaswami | Asian Indian | South Asian | Wong et al. |
| Krishnaswamy | Asian Indian | South Asian | Wong et al. |
| Krishnayya | Asian Indian | South Asian | Wong et al. |
| Kritonia |  | South Asian | Lim et al. |
| Kshatriya | Asian Indian | South Asian | Wong et al. |
| Kshetri |  | South Asian | Lim et al. |
| Kshirsagar | Asian Indian | South Asian | Wong et al. |
| Ku | Chinese | Chinese | Kranick et al. |
| Kua | Chinese | Chinese | Wong et al. |
| Kuai | Chinese | Chinese | Wong et al. |
| Kuan | Chinese | Chinese | Wong et al. |
| Kuang | Chinese | Chinese | Wong et al. |
| Kuay | Chinese | Chinese | Wong et al. |
| Kuba | Japanese | Other Asian | Wong et al. |
| Kubo | Japanese | Other Asian | Wong et al. |
| Kubokawa | Japanese | Other Asian | Wong et al. |
| Kuboki | Japanese | Other Asian | Wong et al. |
| Kubota | Japanese | Other Asian | Wong et al. |
| Kubotera | Japanese | Other Asian | Wong et al. |
| Kuboyama | Japanese | Other Asian | Wong et al. |
| Kubra |  | South Asian | Lim et al. |
| Kuchipudi | Asian Indian | South Asian | Wong et al. |
| Kuda | Japanese | Other Asian | Wong et al. |
| Kudaka | Japanese | Other Asian | Wong et al. |
| Kudilil | Asian Indian | South Asian | Wong et al. |
| Kudo | Japanese | Other Asian | Wong et al. |
| Kudva | Asian Indian | South Asian | Wong et al. |
| Kuei | Chinese | Chinese | Wong et al. |
| Kuen | Chinese | Chinese | Wong et al. |
| Kuey | Chinese | Chinese | Wong et al. |
| Kuga | Japanese | Other Asian | Wong et al. |
| Kuge | Japanese | Other Asian | Wong et al. |
| Kuh | Chinese | Chinese | Wong et al. |
| Kuhara | Japanese | Other Asian | Wong et al. |
| Kui | Chinese | Chinese | Wong et al. |
| Kuit | Chinese | Chinese | Wong et al. |
| Kuizon | Filipino | Other Asian | Wong et al. |
| Kuji | Japanese | Other Asian | Wong et al. |
| Kujiraoka | Japanese | Other Asian | Wong et al. |
| Kuki | Japanese | Other Asian | Wong et al. |
| Kukita | Japanese | Other Asian | Wong et al. |
| Kulangara | Asian Indian | South Asian | Wong et al. |
| Kulkarni | Asian Indian | South Asian | Wong et al. |
| Kulsum |  | South Asian | Lim et al. |
| Kum | Chinese | Chinese | Wong et al. |
| Kuma | Japanese | Other Asian | Wong et al. |
| Kumabe | Japanese | Other Asian | Wong et al. |
| Kumada | Japanese | Other Asian | Wong et al. |
| Kumagae | Japanese | Other Asian | Wong et al. |
| Kumagai | Japanese | Other Asian | Wong et al. |
| Kumai | Japanese | Other Asian | Wong et al. |
| Kumaki | Japanese | Other Asian | Wong et al. |
| Kumakura | Japanese | Other Asian | Wong et al. |
| Kumamoto | Japanese | Other Asian | Wong et al. |
| Kumano | Japanese | Other Asian | Wong et al. |
| Kumar | Asian Indian | South Asian | Kranick et al. |
| Kumaran | Asian Indian | South Asian | Wong et al. |
| Kumari | Asian Indian | South Asian | Wong et al. |
| Kumasaka | Japanese | Other Asian | Wong et al. |
| Kumashiro | Japanese | Other Asian | Wong et al. |
| Kumata | Japanese | Other Asian | Wong et al. |
| Kumazawa | Japanese | Other Asian | Wong et al. |
| Kumbhani | Asian Indian | South Asian | Wong et al. |
| Kume | Japanese | Other Asian | Wong et al. |
| Kumra | Asian Indian | South Asian | Wong et al. |
| Kunchandy | Asian Indian | South Asian | Wong et al. |
| Kundi | Asian Indian | South Asian | Wong et al. |
| Kundu | Asian Indian | South Asian | Wong et al. |
| Kung | Chinese | Chinese | Kranick et al. |
| Kunieda | Japanese | Other Asian | Wong et al. |
| Kunihiro | Japanese | Other Asian | Wong et al. |
| Kunii | Japanese | Other Asian | Wong et al. |
| Kunimatsu | Japanese | Other Asian | Wong et al. |
| Kunimitsu | Japanese | Other Asian | Wong et al. |
| Kunimoto | Japanese | Other Asian | Wong et al. |
| Kunimura | Japanese | Other Asian | Wong et al. |
| Kunisada | Japanese | Other Asian | Wong et al. |
| Kunisawa | Japanese | Other Asian | Wong et al. |
| Kunishige | Japanese | Other Asian | Kranick et al. |
| Kunita | Japanese | Other Asian | Wong et al. |
| Kunitake | Japanese | Other Asian | Wong et al. |
| Kunitomo | Japanese | Other Asian | Wong et al. |
| Kuniyasu | Japanese | Other Asian | Wong et al. |
| Kuniyoshi | Japanese | Other Asian | Wong et al. |
| Kuniyuki | Japanese | Other Asian | Wong et al. |
| Kunj-Ramen |  | South Asian | Lim et al. |
| Kunju | Asian Indian | South Asian | Wong et al. |
| Kunjummen | Asian Indian | South Asian | Wong et al. |
| Kunnel | Asian Indian | South Asian | Wong et al. |
| Kuno | Japanese | Other Asian | Wong et al. |
| Kunugi | Japanese | Other Asian | Wong et al. |
| Kunwor |  | South Asian | Lim et al. |
| Kuo | Chinese | Chinese | Kranick et al. |
| Kuok | Chinese | Chinese | Wong et al. |
| Kuong | Chinese | Chinese | Wong et al. |
| Kuppusamy | Asian Indian | South Asian | Wong et al. |
| Kuppuswamy | Asian Indian | South Asian | Wong et al. |
| Kupsammy |  | South Asian | Lim et al. |
| Kurabayashi | Japanese | Other Asian | Wong et al. |
| Kurachi | Japanese | Other Asian | Wong et al. |
| Kurahara | Japanese | Other Asian | Wong et al. |
| Kurahashi | Japanese | Other Asian | Wong et al. |
| Kuraishi | Japanese | Other Asian | Wong et al. |
| Kuramochi | Japanese | Other Asian | Wong et al. |
| Kuramoto | Japanese | Other Asian | Wong et al. |
| Kuramura | Japanese | Other Asian | Wong et al. |
| Kuranishi | Japanese | Other Asian | Wong et al. |
| Kurano | Japanese | Other Asian | Wong et al. |
| Kuraoka | Japanese | Other Asian | Wong et al. |
| Kurasawa | Japanese | Other Asian | Wong et al. |
| Kurashige | Japanese | Other Asian | Wong et al. |
| Kurashima | Japanese | Other Asian | Wong et al. |
| Kurata | Japanese | Other Asian | Wong et al. |
| Kuratomi | Japanese | Other Asian | Wong et al. |
| Kurauchi | Japanese | Other Asian | Wong et al. |
| Kurebayashi | Japanese | Other Asian | Wong et al. |
| Kureishy | Asian Indian | South Asian | Wong et al. |
| Kureshi | Asian Indian | South Asian | Wong et al. |
| Kuri |  | South Asian | Lim et al. |
| Kuriakose | Asian Indian | South Asian | Wong et al. |
| Kurian | Asian Indian | South Asian | Kranick et al. |
| Kuribayashi | Japanese | Other Asian | Wong et al. |
| Kurien | Asian Indian | South Asian | Wong et al. |
| Kurihara | Japanese | Other Asian | Kranick et al. |
| Kuriki | Japanese | Other Asian | Wong et al. |
| Kurimoto | Japanese | Other Asian | Wong et al. |
| Kurino | Japanese | Other Asian | Wong et al. |
| Kurioka | Japanese | Other Asian | Wong et al. |
| Kurisu | Japanese | Other Asian | Wong et al. |
| Kurita | Japanese | Other Asian | Wong et al. |
| Kuriyama | Japanese | Other Asian | Wong et al. |
| Kurji | Asian Indian | South Asian | Wong et al. |
| Kuroda | Japanese | Other Asian | Wong et al. |
| Kurogi | Japanese | Other Asian | Wong et al. |
| Kuroiwa | Japanese | Other Asian | Wong et al. |
| Kurokawa | Japanese | Other Asian | Wong et al. |
| Kuroki | Japanese | Other Asian | Wong et al. |
| Kurosaka | Japanese | Other Asian | Wong et al. |
| Kurosaki | Japanese | Other Asian | Wong et al. |
| Kurosawa | Japanese | Other Asian | Wong et al. |
| Kurose | Japanese | Other Asian | Wong et al. |
| Kuroshima | Japanese | Other Asian | Wong et al. |
| Kurosu | Japanese | Other Asian | Wong et al. |
| Kurotaki | Japanese | Other Asian | Wong et al. |
| Kuroyanagi | Japanese | Other Asian | Wong et al. |
| Kurozumi | Japanese | Other Asian | Wong et al. |
| Kurup | Asian Indian | South Asian | Wong et al. |
| Kurut | Asian Indian | South Asian | Wong et al. |
| Kuruvila | Asian Indian | South Asian | Wong et al. |
| Kuruvilla | Asian Indian | South Asian | Wong et al. |
| Kusaba | Japanese | Other Asian | Wong et al. |
| Kusada | Japanese | Other Asian | Wong et al. |
| Kusaka | Japanese | Other Asian | Wong et al. |
| Kusakabe | Japanese | Other Asian | Wong et al. |
| Kusakari | Japanese | Other Asian | Wong et al. |
| Kusama | Japanese | Other Asian | Wong et al. |
| Kusanagi | Japanese | Other Asian | Wong et al. |
| Kusano | Japanese | Other Asian | Wong et al. |
| Kusayanagi | Japanese | Other Asian | Wong et al. |
| Kushi | Japanese | Other Asian | Wong et al. |
| Kushida | Japanese | Other Asian | Wong et al. |
| Kushiro | Japanese | Other Asian | Wong et al. |
| Kushwaha | Asian Indian | South Asian | Wong et al. |
| Kusuda | Japanese | Other Asian | Wong et al. |
| Kusuhara | Japanese | Other Asian | Wong et al. |
| Kusumi | Japanese | Other Asian | Wong et al. |
| Kusumoto | Japanese | Other Asian | Wong et al. |
| Kusunoki | Japanese | Other Asian | Wong et al. |
| Kusuyama | Japanese | Other Asian | Wong et al. |
| Kutty | Asian Indian | South Asian | Wong et al. |
| Kuwabara | Japanese | Other Asian | Wong et al. |
| Kuwada | Japanese | Other Asian | Wong et al. |
| Kuwae | Japanese | Other Asian | Wong et al. |
| Kuwahara | Japanese | Other Asian | Wong et al. |
| Kuwahata | Japanese | Other Asian | Wong et al. |
| Kuwamura | Japanese | Other Asian | Wong et al. |
| Kuwana | Japanese | Other Asian | Wong et al. |
| Kuwano | Japanese | Other Asian | Wong et al. |
| Kuwasaki | Japanese | Other Asian | Wong et al. |
| Kuwashima | Japanese | Other Asian | Wong et al. |
| Kuwata | Japanese | Other Asian | Wong et al. |
| Kuwayama | Japanese | Other Asian | Wong et al. |
| Kuyama | Japanese | Other Asian | Wong et al. |
| Kuze | Japanese | Other Asian | Wong et al. |
| Kuzuhara | Japanese | Other Asian | Wong et al. |
| Kwack | Korean | Other Asian | Wong et al. |
| Kwai | Chinese | Chinese | Wong et al. |
| Kwak | Korean; Chinese | Other Asian | Kranick et al. |
| Kwan | Chinese | Chinese | Kranick et al. |
| Kwang | Chinese | Chinese | Wong et al. |
| Kwatra | Asian Indian | South Asian | Wong et al. |
| Kwei | Chinese | Chinese | Wong et al. |
| Kwen | Chinese | Chinese | Wong et al. |
| Kweon | Korean | Other Asian | Wong et al. |
| Kwn | Korean | Other Asian | Wong et al. |
| Kwo | Chinese | Chinese | Wong et al. |
| Kwock | Chinese | Chinese | Wong et al. |
| Kwoh | Chinese | Chinese | Wong et al. |
| Kwok | Chinese | Chinese | Kranick et al. |
| Kwon | Korean | Other Asian | Kranick et al. |
| Kwong | Chinese | Chinese | Kranick et al. |
| Kwoon | Chinese | Chinese | Wong et al. |
| Kwoun | Korean | Other Asian | Wong et al. |
| Kwun | Korean | Other Asian | Wong et al. |
| Kwung | Chinese | Chinese | Wong et al. |
| Ky | Vietnamese | Other Asian | Wong et al. |
| Kye | Korean | Other Asian | Wong et al. |
| Kyi | Burmese | Other Asian | Kranick et al. |
| Kyogoku | Japanese | Other Asian | Wong et al. |
| Kyon | Korean | Other Asian | Wong et al. |
| Kyong | Korean | Other Asian | Wong et al. |
| Kyono | Japanese | Other Asian | Wong et al. |
| Kyoo | Korean | Other Asian | Wong et al. |
| Kyoung | Korean | Other Asian | Wong et al. |
| Kyu | Korean | Other Asian | Wong et al. |
| Kyum | Korean | Other Asian | Wong et al. |
| Kyun | Korean | Other Asian | Wong et al. |
| Kyung | Korean | Other Asian | Wong et al. |
| Kyutoku | Japanese | Other Asian | Wong et al. |
| La | Vietnamese; Burmese | Other Asian | Kranick et al. |
| Lababit | Filipino | Other Asian | Wong et al. |
| Labad | Filipino | Other Asian | Wong et al. |
| Labadan | Filipino | Other Asian | Wong et al. |
| Labador | Filipino | Other Asian | Wong et al. |
| Labajo | Filipino | Other Asian | Wong et al. |
| Labampa | Filipino | Other Asian | Wong et al. |
| Labao | Filipino | Other Asian | Wong et al. |
| Labarda | Filipino | Other Asian | Wong et al. |
| Labaria | Filipino | Other Asian | Wong et al. |
| Labaro | Filipino | Other Asian | Wong et al. |
| Labasan | Filipino | Other Asian | Wong et al. |
| Labastida | Filipino | Other Asian | Wong et al. |
| Labata | Filipino | Other Asian | Wong et al. |
| Labayan | Filipino | Other Asian | Wong et al. |
| Labayen | Filipino | Other Asian | Wong et al. |
| Labayog | Filipino | Other Asian | Wong et al. |
| Laberinto | Filipino | Other Asian | Wong et al. |
| Labiaga | Filipino | Other Asian | Wong et al. |
| Labiano | Filipino | Other Asian | Wong et al. |
| Labio | Filipino | Other Asian | Wong et al. |
| Labios | Filipino | Other Asian | Wong et al. |
| Labis | Filipino | Other Asian | Wong et al. |
| Labit | Filipino | Other Asian | Wong et al. |
| Labitoria | Filipino | Other Asian | Wong et al. |
| Lablu |  | South Asian | Lim et al. |
| Labog | Filipino | Other Asian | Wong et al. |
| Labony |  | South Asian | Lim et al. |
| Labor | Filipino | Other Asian | Wong et al. |
| Laborte | Filipino | Other Asian | Wong et al. |
| Labos | Filipino | Other Asian | Wong et al. |
| Labra | Filipino | Other Asian | Wong et al. |
| Labrado | Filipino | Other Asian | Wong et al. |
| Labrador | Filipino | Other Asian | Wong et al. |
| Labsan | Filipino | Other Asian | Wong et al. |
| Labson | Filipino | Other Asian | Wong et al. |
| Labuanan | Filipino | Other Asian | Wong et al. |
| Labucay | Filipino | Other Asian | Wong et al. |
| Labuguen | Filipino | Other Asian | Wong et al. |
| Labuni | Filipino | Other Asian | Wong et al. |
| Labutong | Filipino | Other Asian | Wong et al. |
| Lac | Vietnamese | Other Asian | Wong et al. |
| Laca | Filipino | Other Asian | Wong et al. |
| Lacaba | Filipino | Other Asian | Wong et al. |
| Lacaden | Filipino | Other Asian | Wong et al. |
| Lacambacal | Filipino | Other Asian | Wong et al. |
| Lacambra | Filipino | Other Asian | Wong et al. |
| Lacandola | Filipino | Other Asian | Wong et al. |
| Lacanienta | Filipino | Other Asian | Wong et al. |
| Lacanilao | Filipino | Other Asian | Wong et al. |
| Lacanlale | Filipino | Other Asian | Wong et al. |
| Lacao | Filipino | Other Asian | Wong et al. |
| Lacap | Filipino | Other Asian | Wong et al. |
| Lacar | Filipino | Other Asian | Wong et al. |
| Lacasandile | Filipino | Other Asian | Wong et al. |
| Lacaste | Filipino | Other Asian | Wong et al. |
| Lacayanga | Filipino | Other Asian | Wong et al. |
| Lacdan | Filipino | Other Asian | Wong et al. |
| Lacdoo | Filipino | Other Asian | Wong et al. |
| Lacebal | Filipino | Other Asian | Wong et al. |
| Laceda | Filipino | Other Asian | Wong et al. |
| Lacerna | Filipino | Other Asian | Wong et al. |
| Lachhman |  | South Asian | Lim et al. |
| Lachica | Filipino | Other Asian | Wong et al. |
| Laciste | Filipino | Other Asian | Wong et al. |
| Laconico | Filipino | Other Asian | Wong et al. |
| Laconsay | Filipino | Other Asian | Wong et al. |
| Lacsa | Filipino | Other Asian | Wong et al. |
| Lacsam | Filipino | Other Asian | Wong et al. |
| Lacsamana | Filipino | Other Asian | Wong et al. |
| Lacsina | Filipino | Other Asian | Wong et al. |
| Lacson | Filipino | Other Asian | Wong et al. |
| Lactaoen | Filipino | Other Asian | Wong et al. |
| Lactaotao | Filipino | Other Asian | Wong et al. |
| Lacuata | Filipino | Other Asian | Wong et al. |
| Lacuesta | Filipino | Other Asian | Wong et al. |
| Lacuin | Filipino | Other Asian | Wong et al. |
| Lacuna | Filipino | Other Asian | Wong et al. |
| Lad | Asian Indian | South Asian | Wong et al. |
| Ladaga | Filipino | Other Asian | Wong et al. |
| Ladao | Filipino | Other Asian | Wong et al. |
| Laddaran | Filipino | Other Asian | Wong et al. |
| Ladera | Filipino | Other Asian | Wong et al. |
| Laderas | Filipino | Other Asian | Wong et al. |
| Ladha | Asian Indian | South Asian | Wong et al. |
| Ladhani | Asian Indian | South Asian | Wong et al. |
| Ladia | Filipino | Other Asian | Wong et al. |
| Ladiero | Filipino | Other Asian | Wong et al. |
| Ladines | Filipino | Other Asian | Wong et al. |
| Ladion | Filipino | Other Asian | Wong et al. |
| Ladisla | Filipino | Other Asian | Wong et al. |
| Ladislao | Filipino | Other Asian | Wong et al. |
| Ladkani |  | South Asian | Lim et al. |
| Ladores | Filipino | Other Asian | Wong et al. |
| Ladra | Filipino | Other Asian | Wong et al. |
| Ladrera | Filipino | Other Asian | Wong et al. |
| Ladrido | Filipino | Other Asian | Wong et al. |
| Ladrillo | Filipino | Other Asian | Wong et al. |
| Ladrillono | Filipino | Other Asian | Wong et al. |
| Ladringan | Filipino | Other Asian | Wong et al. |
| Laeno | Filipino | Other Asian | Wong et al. |
| Laforga | Filipino | Other Asian | Wong et al. |
| Laforteza | Filipino | Other Asian | Wong et al. |
| Lafrades | Filipino | Other Asian | Wong et al. |
| Lafradez | Filipino | Other Asian | Wong et al. |
| Lafuente | Filipino | Other Asian | Wong et al. |
| Lagac | Filipino | Other Asian | Wong et al. |
| Lagado | Filipino | Other Asian | Wong et al. |
| Lagahit | Filipino | Other Asian | Wong et al. |
| Lagang | Filipino | Other Asian | Wong et al. |
| Lagao | Filipino | Other Asian | Wong et al. |
| Lagapa | Filipino | Other Asian | Wong et al. |
| Lagario | Filipino | Other Asian | Wong et al. |
| Lagarto | Filipino | Other Asian | Wong et al. |
| Lagasca | Filipino | Other Asian | Wong et al. |
| Lagat | Filipino | Other Asian | Wong et al. |
| Lagatuz | Filipino | Other Asian | Wong et al. |
| Lagaya | Filipino | Other Asian | Wong et al. |
| Lagazo | Filipino | Other Asian | Wong et al. |
| Lagda | Filipino | Other Asian | Wong et al. |
| Lagdaan | Filipino | Other Asian | Wong et al. |
| Lagdamen | Filipino | Other Asian | Wong et al. |
| Lagdameo | Filipino | Other Asian | Wong et al. |
| Lagera | Filipino | Other Asian | Wong et al. |
| Laggui | Filipino | Other Asian | Wong et al. |
| Lagleva | Filipino | Other Asian | Wong et al. |
| Lagman | Filipino | Other Asian | Wong et al. |
| Lagmay | Filipino | Other Asian | Wong et al. |
| Lagniton | Filipino | Other Asian | Wong et al. |
| Lago | Filipino | Other Asian | Wong et al. |
| Lagoc | Filipino | Other Asian | Wong et al. |
| Lagon | Filipino | Other Asian | Wong et al. |
| Lagonoy | Filipino | Other Asian | Wong et al. |
| Lagos | Filipino | Other Asian | Wong et al. |
| Lagoy | Filipino | Other Asian | Wong et al. |
| Lagpacan | Filipino | Other Asian | Wong et al. |
| Lagria | Filipino | Other Asian | Wong et al. |
| Lagrimas | Filipino | Other Asian | Wong et al. |
| Lagrisola | Filipino | Other Asian | Wong et al. |
| Lagrosa | Filipino | Other Asian | Wong et al. |
| Lagtapon | Filipino | Other Asian | Wong et al. |
| Lagua | Filipino | Other Asian | Wong et al. |
| Laguador | Filipino | Other Asian | Wong et al. |
| Laguardia | Filipino | Other Asian | Wong et al. |
| Laguatan | Filipino | Other Asian | Wong et al. |
| Lagud | Filipino | Other Asian | Wong et al. |
| Laguda | Filipino | Other Asian | Wong et al. |
| Laguer | Filipino | Other Asian | Wong et al. |
| Laguerta | Filipino | Other Asian | Wong et al. |
| Laguesma | Filipino | Other Asian | Wong et al. |
| Laguio | Filipino | Other Asian | Wong et al. |
| Laguisma | Filipino | Other Asian | Wong et al. |
| Laguit | Filipino | Other Asian | Wong et al. |
| Laguitan | Filipino | Other Asian | Wong et al. |
| Lagula | Filipino | Other Asian | Wong et al. |
| Lagumbay | Filipino | Other Asian | Wong et al. |
| Laguna | Filipino | Other Asian | Wong et al. |
| Lagunday | Filipino | Other Asian | Wong et al. |
| Lagundino | Filipino | Other Asian | Wong et al. |
| Lagunero | Filipino | Other Asian | Wong et al. |
| Lagunilla | Filipino | Other Asian | Wong et al. |
| Lagunte | Filipino | Other Asian | Wong et al. |
| Lagunzad | Filipino | Other Asian | Wong et al. |
| Lagura | Filipino | Other Asian | Wong et al. |
| Lagutan | Filipino | Other Asian | Wong et al. |
| Lah | Korean | Other Asian | Wong et al. |
| Lahip | Filipino | Other Asian | Wong et al. |
| Lahiri | Asian Indian | South Asian | Wong et al. |
| Lahorra | Filipino | Other Asian | Wong et al. |
| Lahoti | Asian Indian | South Asian | Wong et al. |
| Lahoz | Filipino | Other Asian | Wong et al. |
| Lai | Chinese | Chinese | Kranick et al. |
| Laigo | Filipino | Other Asian | Wong et al. |
| Laila |  | South Asian | Lim et al. |
| Laily |  | South Asian | Lim et al. |
| Lainez | Filipino | Other Asian | Wong et al. |
| Laitan | Filipino | Other Asian | Wong et al. |
| Laiz | Filipino | Other Asian | Wong et al. |
| Laja | Filipino | Other Asian | Wong et al. |
| Lajom | Filipino | Other Asian | Wong et al. |
| Lakdawala | Asian Indian | South Asian | Wong et al. |
| Lakdawalla | Asian Indian | South Asian | Wong et al. |
| Lakeram |  | South Asian | Lim et al. |
| Lakhan |  | South Asian | Lim et al. |
| Lakhani | Asian Indian | South Asian | Wong et al. |
| Lakhanpal | Asian Indian | South Asian | Wong et al. |
| Lakhany | Asian Indian | South Asian | Wong et al. |
| Lakhdhir | Asian Indian | South Asian | Wong et al. |
| Lakhia | Asian Indian | South Asian | Wong et al. |
| Lakhwara | Asian Indian | South Asian | Wong et al. |
| Lakra | Asian Indian | South Asian | Wong et al. |
| Lakshman | Asian Indian | South Asian | Wong et al. |
| Lakshmanan | Asian Indian | South Asian | Wong et al. |
| Lakshminaray | Asian Indian | South Asian | Wong et al. |
| Lal | Asian Indian | South Asian | Wong et al. |
| Lala |  | South Asian | Lim et al. |
| Lalan | Asian Indian | South Asian | Wong et al. |
| Lalangan | Filipino | Other Asian | Wong et al. |
| Lalani | Pakistani | South Asian | Kranick et al. |
| Lalas | Filipino | Other Asian | Wong et al. |
| Lalata | Filipino | Other Asian | Wong et al. |
| Lalbhai | Asian Indian | South Asian | Wong et al. |
| Lalic | Filipino | Other Asian | Wong et al. |
| Lalisan | Filipino | Other Asian | Wong et al. |
| Lalji | Asian Indian | South Asian | Wong et al. |
| Lall |  | South Asian | Lim et al. |
| Lallana | Filipino | Other Asian | Wong et al. |
| Lallave | Filipino | Other Asian | Wong et al. |
| Lallubhai | Asian Indian | South Asian | Wong et al. |
| Lalman |  | South Asian | Lim et al. |
| Lalmansingh |  | South Asian | Lim et al. |
| Lalog | Filipino | Other Asian | Wong et al. |
| Laluan | Filipino | Other Asian | Wong et al. |
| Lalucis | Filipino | Other Asian | Wong et al. |
| Lam | Chinese; Vietnamese | Chinese | Kranick et al. |
| Lama |  | South Asian | Lim et al. |
| Lamadrid | Filipino | Other Asian | Wong et al. |
| Lamangan | Filipino | Other Asian | Wong et al. |
| Lamano | Filipino | Other Asian | Wong et al. |
| Lambinicio | Filipino | Other Asian | Wong et al. |
| Lambino | Filipino | Other Asian | Wong et al. |
| Lamera | Filipino | Other Asian | Wong et al. |
| Lamichhane |  | South Asian | Lim et al. |
| Lamigo | Filipino | Other Asian | Wong et al. |
| Lamorena | Filipino | Other Asian | Wong et al. |
| Lamoste | Filipino | Other Asian | Wong et al. |
| Lampa | Filipino | Other Asian | Wong et al. |
| Lampitoc | Filipino | Other Asian | Wong et al. |
| Lamsen | Filipino | Other Asian | Wong et al. |
| Lamug | Filipino | Other Asian | Wong et al. |
| Lamzon | Filipino | Other Asian | Wong et al. |
| Lan | Chinese | Chinese | Wong et al. |
| Lanada | Filipino | Other Asian | Wong et al. |
| Lanaria | Filipino | Other Asian | Wong et al. |
| Lanceta | Filipino | Other Asian | Wong et al. |
| Landas | Filipino | Other Asian | Wong et al. |
| Landayan | Filipino | Other Asian | Wong et al. |
| Landazabal | Filipino | Other Asian | Wong et al. |
| Landero | Filipino | Other Asian | Wong et al. |
| Landicho | Filipino | Other Asian | Wong et al. |
| Landingin | Filipino | Other Asian | Wong et al. |
| Landinguin | Filipino | Other Asian | Wong et al. |
| Landrito | Filipino | Other Asian | Wong et al. |
| Langaman | Filipino | Other Asian | Wong et al. |
| Langi | Filipino | Other Asian | Wong et al. |
| Langit | Filipino | Other Asian | Wong et al. |
| Languido | Filipino | Other Asian | Wong et al. |
| Lanh | Vietnamese | Other Asian | Wong et al. |
| Lanot | Filipino | Other Asian | Wong et al. |
| Lanoza | Filipino | Other Asian | Wong et al. |
| Lansang | Filipino | Other Asian | Wong et al. |
| Lansangan | Filipino | Other Asian | Wong et al. |
| Lansigan | Filipino | Other Asian | Wong et al. |
| Lantano | Filipino | Other Asian | Wong et al. |
| Lantican | Filipino | Other Asian | Wong et al. |
| Lantin | Filipino | Other Asian | Wong et al. |
| Lanting | Filipino | Other Asian | Wong et al. |
| Lanuevo | Filipino | Other Asian | Wong et al. |
| Lanuza | Filipino | Other Asian | Wong et al. |
| Lanuzo | Filipino | Other Asian | Wong et al. |
| Lanzona | Filipino | Other Asian | Wong et al. |
| Lao | Chinese | Chinese | Kranick et al. |
| Laoang | Filipino | Other Asian | Wong et al. |
| Laolao | Filipino | Other Asian | Wong et al. |
| Laoyan | Filipino | Other Asian | Wong et al. |
| Lap | Chinese | Chinese | Wong et al. |
| Lapa | Filipino | Other Asian | Wong et al. |
| Lapada | Filipino | Other Asian | Wong et al. |
| Laparan | Filipino | Other Asian | Wong et al. |
| Lapastora | Filipino | Other Asian | Wong et al. |
| Lapaz | Filipino | Other Asian | Wong et al. |
| Lapena | Filipino | Other Asian | Wong et al. |
| Lapenia | Filipino | Other Asian | Wong et al. |
| Lapez | Filipino | Other Asian | Wong et al. |
| Lapid | Filipino | Other Asian | Wong et al. |
| Lapidario | Filipino | Other Asian | Wong et al. |
| Lapina | Filipino | Other Asian | Wong et al. |
| Lapinig | Filipino | Other Asian | Wong et al. |
| Lapitan | Filipino | Other Asian | Wong et al. |
| Lapiz | Filipino | Other Asian | Wong et al. |
| Laplana | Filipino | Other Asian | Wong et al. |
| Lapuebla | Filipino | Other Asian | Wong et al. |
| Lapurga | Filipino | Other Asian | Wong et al. |
| Lapus | Filipino | Other Asian | Wong et al. |
| Laput | Filipino | Other Asian | Wong et al. |
| Lapuz | Filipino | Other Asian | Wong et al. |
| Laqui | Filipino | Other Asian | Wong et al. |
| Laquian | Filipino | Other Asian | Wong et al. |
| Laquindanum | Filipino | Other Asian | Wong et al. |
| Lara | Filipino | Other Asian | Wong et al. |
| Laracas | Filipino | Other Asian | Wong et al. |
| Laranang | Filipino | Other Asian | Wong et al. |
| Laraya | Filipino | Other Asian | Wong et al. |
| Lardizabal | Filipino | Other Asian | Wong et al. |
| Larena | Filipino | Other Asian | Wong et al. |
| Lareza | Filipino | Other Asian | Wong et al. |
| Largo | Filipino | Other Asian | Wong et al. |
| Largoza | Filipino | Other Asian | Wong et al. |
| Larican | Filipino | Other Asian | Wong et al. |
| Larin | Filipino | Other Asian | Wong et al. |
| Larios | Filipino | Other Asian | Wong et al. |
| Lariosa | Filipino | Other Asian | Wong et al. |
| Larioza | Filipino | Other Asian | Wong et al. |
| Larita | Filipino | Other Asian | Wong et al. |
| Laroco | Filipino | Other Asian | Wong et al. |
| Laroia | Asian Indian | South Asian | Wong et al. |
| Laron | Filipino | Other Asian | Wong et al. |
| Larot | Filipino | Other Asian | Wong et al. |
| Laroya | Filipino | Other Asian | Wong et al. |
| Laroza | Filipino | Other Asian | Wong et al. |
| Larracas | Filipino | Other Asian | Wong et al. |
| Larraga | Filipino | Other Asian | Wong et al. |
| Larrarte | Filipino | Other Asian | Wong et al. |
| Larroza | Filipino | Other Asian | Wong et al. |
| Lasa | Filipino | Other Asian | Wong et al. |
| Lasam | Filipino | Other Asian | Wong et al. |
| Lascano | Filipino | Other Asian | Wong et al. |
| Lascuna | Filipino | Other Asian | Wong et al. |
| Laserna | Filipino | Other Asian | Wong et al. |
| Lashari |  | South Asian | Lim et al. |
| Lasin | Filipino | Other Asian | Wong et al. |
| Lasker |  | South Asian | Lim et al. |
| Lasmarias | Filipino | Other Asian | Wong et al. |
| Lasola | Filipino | Other Asian | Wong et al. |
| Laspinas | Filipino | Other Asian | Wong et al. |
| Lasquete | Filipino | Other Asian | Wong et al. |
| Lasquety | Filipino | Other Asian | Wong et al. |
| Lastima | Filipino | Other Asian | Wong et al. |
| Lastimosa | Filipino | Other Asian | Wong et al. |
| Lastimoso | Filipino | Other Asian | Wong et al. |
| Lastimoza | Filipino | Other Asian | Wong et al. |
| Lastra | Filipino | Other Asian | Wong et al. |
| Lastrella | Filipino | Other Asian | Wong et al. |
| Lastrilla | Filipino | Other Asian | Wong et al. |
| Lat | Filipino | Other Asian | Wong et al. |
| Lata |  | South Asian | Lim et al. |
| Latada | Filipino | Other Asian | Wong et al. |
| Latag | Filipino | Other Asian | Wong et al. |
| Lataquin | Filipino | Other Asian | Wong et al. |
| Latchaman |  | South Asian | Lim et al. |
| Latchman |  | South Asian | Lim et al. |
| Lateef | Asian Indian | South Asian | Wong et al. |
| Lathi | Asian Indian | South Asian | Wong et al. |
| Lathia | Asian Indian | South Asian | Wong et al. |
| Latif | Pakistani; Bangladeshi | South Asian | Kranick et al. |
| Latifa |  | South Asian | Lim et al. |
| Latiff | Asian Indian | South Asian | Wong et al. |
| Latonio | Filipino | Other Asian | Wong et al. |
| Latosa | Filipino | Other Asian | Wong et al. |
| Latoza | Filipino | Other Asian | Wong et al. |
| Lau | Chinese | Chinese | Kranick et al. |
| Lauchengco | Filipino | Other Asian | Wong et al. |
| Laud | Filipino | Other Asian | Wong et al. |
| Laudencia | Filipino | Other Asian | Wong et al. |
| Lauigan | Filipino | Other Asian | Wong et al. |
| Laungayan | Filipino | Other Asian | Wong et al. |
| Laure | Filipino | Other Asian | Wong et al. |
| Laurea | Filipino | Other Asian | Wong et al. |
| Laureano | Filipino | Other Asian | Wong et al. |
| Laurel | Filipino | Other Asian | Wong et al. |
| Laurena | Filipino | Other Asian | Wong et al. |
| Laurencio | Filipino | Other Asian | Wong et al. |
| Laurente | Filipino | Other Asian | Wong et al. |
| Laureta | Filipino | Other Asian | Wong et al. |
| Laureto | Filipino | Other Asian | Wong et al. |
| Lauricio | Filipino | Other Asian | Wong et al. |
| Laurio | Filipino | Other Asian | Wong et al. |
| Lauron | Filipino | Other Asian | Wong et al. |
| Laus | Filipino | Other Asian | Wong et al. |
| Lauyan | Filipino | Other Asian | Wong et al. |
| Lav | Chinese | Chinese | Wong et al. |
| Lavadia | Filipino | Other Asian | Wong et al. |
| Lavado | Filipino | Other Asian | Wong et al. |
| Lavares | Filipino | Other Asian | Wong et al. |
| Lavarez | Filipino | Other Asian | Wong et al. |
| Lavarias | Filipino | Other Asian | Wong et al. |
| Lavaro | Filipino | Other Asian | Wong et al. |
| Lavarro | Filipino | Other Asian | Wong et al. |
| Lavides | Filipino | Other Asian | Wong et al. |
| Lavilla | Filipino | Other Asian | Wong et al. |
| Lavina | Filipino | Other Asian | Wong et al. |
| Lavitoria | Filipino | Other Asian | Wong et al. |
| Law | Chinese | Chinese | Kranick et al. |
| Lawas | Filipino | Other Asian | Wong et al. |
| Laxa | Filipino | Other Asian | Wong et al. |
| Laxamana | Filipino | Other Asian | Wong et al. |
| Laxina | Filipino | Other Asian | Wong et al. |
| Laxminarayan | Asian Indian | South Asian | Wong et al. |
| Laya | Filipino | Other Asian | Wong et al. |
| Layacan | Filipino | Other Asian | Wong et al. |
| Layag | Filipino | Other Asian | Wong et al. |
| Layamana | Filipino | Other Asian | Wong et al. |
| Layao | Filipino | Other Asian | Wong et al. |
| Layaoen | Filipino | Other Asian | Wong et al. |
| Layco | Filipino | Other Asian | Wong et al. |
| Laygo | Filipino | Other Asian | Wong et al. |
| Layla |  | South Asian | Lim et al. |
| Layno | Filipino | Other Asian | Wong et al. |
| Layog | Filipino | Other Asian | Wong et al. |
| Layola | Filipino | Other Asian | Wong et al. |
| Layon | Filipino | Other Asian | Wong et al. |
| Layos | Filipino | Other Asian | Wong et al. |
| Layson | Filipino | Other Asian | Wong et al. |
| Layug | Filipino | Other Asian | Wong et al. |
| Layugan | Filipino | Other Asian | Wong et al. |
| Layus | Filipino | Other Asian | Wong et al. |
| Laza | Filipino | Other Asian | Wong et al. |
| Lazaga | Filipino | Other Asian | Wong et al. |
| Lazam | Filipino | Other Asian | Wong et al. |
| Lazarito | Filipino | Other Asian | Wong et al. |
| Lazaro | Filipino | Other Asian | Wong et al. |
| Lazarte | Filipino | Other Asian | Wong et al. |
| Lazatin | Filipino | Other Asian | Wong et al. |
| Lazo | Filipino | Other Asian | Wong et al. |
| Le | Vietnamese | Other Asian | Kranick et al. |
| Leabres | Filipino | Other Asian | Wong et al. |
| Leachon | Filipino | Other Asian | Wong et al. |
| Leal | Filipino | Other Asian | Wong et al. |
| Leang | Chinese | Chinese | Wong et al. |
| Leano | Filipino | Other Asian | Wong et al. |
| Leao | Chinese | Chinese | Wong et al. |
| Leba | Chinese | Chinese | Wong et al. |
| Lebrilla | Filipino | Other Asian | Wong et al. |
| Lebumfacil | Filipino | Other Asian | Wong et al. |
| Lecaros | Filipino | Other Asian | Wong et al. |
| Lechuga | Filipino | Other Asian | Wong et al. |
| Lectura | Filipino | Other Asian | Wong et al. |
| Ledda | Filipino | Other Asian | Wong et al. |
| Ledesma | Filipino | Other Asian | Wong et al. |
| Ledina | Filipino | Other Asian | Wong et al. |
| Leduc | Vietnamese | Other Asian | Wong et al. |
| Lee | Chinese; Korean | Chinese | Kranick et al. |
| Leelasingh |  | South Asian | Lim et al. |
| Leelin | Filipino | Other Asian | Wong et al. |
| Leem | Korean | Other Asian | Wong et al. |
| Leer |  | South Asian | Lim et al. |
| Leeton |  | South Asian | Lim et al. |
| Leg | Chinese | Chinese | Wong et al. |
| Legarda | Filipino | Other Asian | Wong et al. |
| Legarde | Filipino | Other Asian | Wong et al. |
| Legaspi | Filipino | Other Asian | Wong et al. |
| Legasto | Filipino | Other Asian | Wong et al. |
| Legion | Filipino | Other Asian | Wong et al. |
| Legislador | Filipino | Other Asian | Wong et al. |
| Lehal | Asian Indian | South Asian | Wong et al. |
| Lei | Chinese | Chinese | Kranick et al. |
| Leing | Chinese | Chinese | Wong et al. |
| Leiva | Chinese | Chinese | Wong et al. |
| Lejano | Filipino | Other Asian | Wong et al. |
| Lejarde | Filipino | Other Asian | Wong et al. |
| Lele | Asian Indian | South Asian | Wong et al. |
| Lelina | Filipino | Other Asian | Wong et al. |
| Lelis | Filipino | Other Asian | Wong et al. |
| Lem | Cambodian; Vietnamese | Other Asian | Kranick et al. |
| Lemi | Filipino | Other Asian | Wong et al. |
| Lemoncito | Filipino | Other Asian | Wong et al. |
| Lemos | Filipino | Other Asian | Wong et al. |
| Len | Chinese | Chinese | Wong et al. |
| Lena | Filipino | Other Asian | Wong et al. |
| Leng | Chinese | Chinese | Wong et al. |
| Lenh | Vietnamese | Other Asian | Wong et al. |
| Leo | Chinese | Chinese | Kranick et al. |
| Leocadio | Filipino | Other Asian | Wong et al. |
| Leomo | Filipino | Other Asian | Wong et al. |
| Leon | Chinese | Chinese | Wong et al. |
| Leonardo | Filipino | Other Asian | Wong et al. |
| Leoncio | Filipino | Other Asian | Wong et al. |
| Leonen | Filipino | Other Asian | Wong et al. |
| Leones | Filipino | Other Asian | Wong et al. |
| Leong | Chinese | Chinese | Kranick et al. |
| Leonida | Filipino | Other Asian | Wong et al. |
| Leonin | Filipino | Other Asian | Wong et al. |
| Leonor | Filipino | Other Asian | Wong et al. |
| Leopando | Filipino | Other Asian | Wong et al. |
| Leopoldo | Filipino | Other Asian | Wong et al. |
| Lep | Chinese | Chinese | Wong et al. |
| Leprozo | Filipino | Other Asian | Wong et al. |
| Lequang | Vietnamese | Other Asian | Wong et al. |
| Lerias | Filipino | Other Asian | Wong et al. |
| Lerio | Filipino | Other Asian | Wong et al. |
| Lerit | Filipino | Other Asian | Wong et al. |
| Lerma | Filipino | Other Asian | Wong et al. |
| Lerum | Filipino | Other Asian | Wong et al. |
| Lesaca | Filipino | Other Asian | Wong et al. |
| Lescano | Filipino | Other Asian | Wong et al. |
| Let | Chinese | Chinese | Wong et al. |
| Letrero | Filipino | Other Asian | Wong et al. |
| Leu | Chinese | Chinese | Wong et al. |
| Leuang | Vietnamese | Other Asian | Wong et al. |
| Leun | Chinese | Chinese | Wong et al. |
| Leung | Chinese | Chinese | Kranick et al. |
| Leus | Filipino | Other Asian | Wong et al. |
| Leuterio | Filipino | Other Asian | Wong et al. |
| Levan | Vietnamese | Other Asian | Wong et al. |
| Leveriza | Filipino | Other Asian | Wong et al. |
| Leviste | Filipino | Other Asian | Wong et al. |
| Levng | Chinese | Chinese | Wong et al. |
| Lew | Chinese | Chinese | Kranick et al. |
| Lewe | Korean | Other Asian | Wong et al. |
| Lewis |  | South Asian | Lim et al. |
| Leyba | Filipino | Other Asian | Wong et al. |
| Leyco | Filipino | Other Asian | Wong et al. |
| Leyesa | Filipino | Other Asian | Wong et al. |
| Leygo | Filipino | Other Asian | Wong et al. |
| Leynes | Filipino | Other Asian | Wong et al. |
| Leyno | Filipino | Other Asian | Wong et al. |
| Leyran | Filipino | Other Asian | Wong et al. |
| Leyson | Filipino | Other Asian | Wong et al. |
| Leyva | Filipino | Other Asian | Wong et al. |
| Lezada | Filipino | Other Asian | Wong et al. |
| Lhamo |  | South Asian | Lim et al. |
| Li | Chinese | Chinese | Kranick et al. |
| Liad | Chinese | Chinese | Wong et al. |
| Lian | Chinese | Chinese | Wong et al. |
| Liang | Chinese | Chinese | Kranick et al. |
| Liao | Chinese | Chinese | Kranick et al. |
| Liau | Chinese | Chinese | Wong et al. |
| Liaw | Chinese | Chinese | Wong et al. |
| Libadia | Filipino | Other Asian | Wong et al. |
| Liban | Filipino | Other Asian | Wong et al. |
| Libanan | Filipino | Other Asian | Wong et al. |
| Libao | Filipino | Other Asian | Wong et al. |
| Libarnes | Filipino | Other Asian | Wong et al. |
| Libatique | Filipino | Other Asian | Wong et al. |
| Libay | Filipino | Other Asian | Wong et al. |
| Libed | Filipino | Other Asian | Wong et al. |
| Liberato | Filipino | Other Asian | Wong et al. |
| Libiran | Filipino | Other Asian | Wong et al. |
| Liboon | Filipino | Other Asian | Wong et al. |
| Liboro | Filipino | Other Asian | Wong et al. |
| Librado | Filipino | Other Asian | Wong et al. |
| Libranda | Filipino | Other Asian | Wong et al. |
| Libre | Filipino | Other Asian | Wong et al. |
| Librea | Filipino | Other Asian | Wong et al. |
| Libres | Filipino | Other Asian | Wong et al. |
| Librojo | Filipino | Other Asian | Wong et al. |
| Libuit | Filipino | Other Asian | Wong et al. |
| Libunao | Filipino | Other Asian | Wong et al. |
| Libut | Filipino | Other Asian | Wong et al. |
| Licad | Filipino | Other Asian | Wong et al. |
| Licardo | Filipino | Other Asian | Wong et al. |
| Licayan | Filipino | Other Asian | Wong et al. |
| Licerio | Filipino | Other Asian | Wong et al. |
| Lichauco | Filipino | Other Asian | Wong et al. |
| Liclican | Filipino | Other Asian | Wong et al. |
| Licop | Filipino | Other Asian | Wong et al. |
| Lictao | Filipino | Other Asian | Wong et al. |
| Licuan | Filipino | Other Asian | Wong et al. |
| Licuanan | Filipino | Other Asian | Wong et al. |
| Licud | Filipino | Other Asian | Wong et al. |
| Licudan | Filipino | Other Asian | Wong et al. |
| Licudine | Filipino | Other Asian | Wong et al. |
| Licup | Filipino | Other Asian | Wong et al. |
| Lieh | Chinese | Chinese | Wong et al. |
| Lieng | Vietnamese | Other Asian | Wong et al. |
| Lieou | Vietnamese | Other Asian | Wong et al. |
| Lieu | Chinese | Chinese | Wong et al. |
| Liew | Chinese; Malaysian | Chinese | Kranick et al. |
| Liganor | Filipino | Other Asian | Wong et al. |
| Ligason | Filipino | Other Asian | Wong et al. |
| Ligaya | Filipino | Other Asian | Wong et al. |
| Ligeralde | Filipino | Other Asian | Wong et al. |
| Liggayu | Filipino | Other Asian | Wong et al. |
| Ligh | Chinese | Chinese | Wong et al. |
| Ligot | Filipino | Other Asian | Wong et al. |
| Ligsay | Filipino | Other Asian | Wong et al. |
| Liguid | Filipino | Other Asian | Wong et al. |
| Ligutom | Filipino | Other Asian | Wong et al. |
| Lii | Chinese | Chinese | Wong et al. |
| Lijauco | Filipino | Other Asian | Wong et al. |
| Lil | Chinese | Chinese | Wong et al. |
| Lila |  | South Asian | Lim et al. |
| Lilagan | Filipino | Other Asian | Wong et al. |
| Lilani | Asian Indian | South Asian | Wong et al. |
| Lili |  | South Asian | Lim et al. |
| Lim | Korean; Malaysian; Chinese | Other Asian | Kranick et al. |
| Limaye | Asian Indian | South Asian | Wong et al. |
| Limb | Korean | Other Asian | Wong et al. |
| Limbachia | Asian Indian | South Asian | Wong et al. |
| Limbag | Filipino | Other Asian | Wong et al. |
| Limbaga | Filipino | Other Asian | Wong et al. |
| Limbo | Filipino | Other Asian | Wong et al. |
| Limcaco | Filipino | Other Asian | Wong et al. |
| Limcangco | Filipino | Other Asian | Wong et al. |
| Limcaoco | Filipino | Other Asian | Wong et al. |
| Limcolioc | Filipino | Other Asian | Wong et al. |
| Limcuando | Filipino | Other Asian | Wong et al. |
| Limdico | Filipino | Other Asian | Wong et al. |
| Lime | Filipino | Other Asian | Wong et al. |
| Limen | Filipino | Other Asian | Wong et al. |
| Limin | Filipino | Other Asian | Wong et al. |
| Limjap | Filipino | Other Asian | Wong et al. |
| Limjoco | Filipino | Other Asian | Wong et al. |
| Limjuco | Filipino | Other Asian | Wong et al. |
| Limlingan | Filipino | Other Asian | Wong et al. |
| Limon | Filipino | Other Asian | Wong et al. |
| Limos | Filipino | Other Asian | Wong et al. |
| Limosnero | Filipino | Other Asian | Wong et al. |
| Limpin | Filipino | Other Asian | Wong et al. |
| Limsico | Chinese | Chinese | Wong et al. |
| Limson | Filipino | Other Asian | Wong et al. |
| Limuaco | Filipino | Other Asian | Wong et al. |
| Lin | Chinese | Chinese | Kranick et al. |
| Lina | Filipino | Other Asian | Wong et al. |
| Linan | Filipino | Other Asian | Wong et al. |
| Linao | Filipino | Other Asian | Wong et al. |
| Linco | Filipino | Other Asian | Wong et al. |
| Lindayen | Filipino | Other Asian | Wong et al. |
| Lindo | Filipino | Other Asian | Wong et al. |
| Ling | Chinese; Malaysian | Chinese | Kranick et al. |
| Linga | Filipino | Other Asian | Wong et al. |
| Lingad | Filipino | Other Asian | Wong et al. |
| Lingam | Asian Indian | South Asian | Wong et al. |
| Lingan | Filipino | Other Asian | Wong et al. |
| Lingao | Filipino | Other Asian | Wong et al. |
| Lingat | Filipino | Other Asian | Wong et al. |
| Linh | Vietnamese | Other Asian | Wong et al. |
| Lino | Filipino | Other Asian | Wong et al. |
| Linsangan | Filipino | Other Asian | Wong et al. |
| Linsao | Filipino | Other Asian | Wong et al. |
| Lintag | Filipino | Other Asian | Wong et al. |
| Lintao | Filipino | Other Asian | Wong et al. |
| Lio | Chinese | Chinese | Wong et al. |
| Liongson | Filipino | Other Asian | Wong et al. |
| Liou | Chinese | Chinese | Wong et al. |
| Lip | Chinese | Chinese | Wong et al. |
| Lipana | Filipino | Other Asian | Wong et al. |
| Lipi |  | South Asian | Lim et al. |
| Liquete | Filipino | Other Asian | Wong et al. |
| Liquigan | Filipino | Other Asian | Wong et al. |
| Lira | Filipino | Other Asian | Wong et al. |
| Lirag | Filipino | Other Asian | Wong et al. |
| Lirazan | Filipino | Other Asian | Wong et al. |
| Lirio | Filipino | Other Asian | Wong et al. |
| Lirios | Filipino | Other Asian | Wong et al. |
| Lisan | Filipino | Other Asian | Wong et al. |
| Lisboa | Filipino | Other Asian | Wong et al. |
| Liscano | Filipino | Other Asian | Wong et al. |
| Lising | Filipino | Other Asian | Wong et al. |
| Lisondra | Filipino | Other Asian | Wong et al. |
| Lista | Filipino | Other Asian | Wong et al. |
| Lit | Chinese | Chinese | Wong et al. |
| Liton |  | South Asian | Lim et al. |
| Litonjua | Filipino | Other Asian | Wong et al. |
| Litorja | Filipino | Other Asian | Wong et al. |
| Littaua | Filipino | Other Asian | Wong et al. |
| Liu | Chinese | Chinese | Kranick et al. |
| Liv | Chinese | Chinese | Wong et al. |
| Lived | Filipino | Other Asian | Wong et al. |
| Livelo | Filipino | Other Asian | Wong et al. |
| Liwag | Filipino | Other Asian | Wong et al. |
| Liwanag | Filipino | Other Asian | Wong et al. |
| Liy | Chinese | Chinese | Wong et al. |
| Liza |  | South Asian | Lim et al. |
| Lizada | Filipino | Other Asian | Wong et al. |
| Lizarda | Filipino | Other Asian | Wong et al. |
| Lizardo | Filipino | Other Asian | Wong et al. |
| Lizares | Filipino | Other Asian | Wong et al. |
| Lizaso | Filipino | Other Asian | Wong et al. |
| Llaban | Filipino | Other Asian | Wong et al. |
| Llabres | Filipino | Other Asian | Wong et al. |
| Llacer | Filipino | Other Asian | Wong et al. |
| Llacuna | Filipino | Other Asian | Wong et al. |
| Lladoc | Filipino | Other Asian | Wong et al. |
| Llagas | Filipino | Other Asian | Wong et al. |
| Llaguno | Filipino | Other Asian | Wong et al. |
| Llamado | Filipino | Other Asian | Wong et al. |
| Llamanzares | Filipino | Other Asian | Wong et al. |
| Llamas | Filipino | Other Asian | Wong et al. |
| Llames | Filipino | Other Asian | Wong et al. |
| Llamido | Filipino | Other Asian | Wong et al. |
| Llamoso | Filipino | Other Asian | Wong et al. |
| Llamzon | Filipino | Other Asian | Wong et al. |
| Llana | Filipino | Other Asian | Wong et al. |
| Llanera | Filipino | Other Asian | Wong et al. |
| Llanes | Filipino | Other Asian | Wong et al. |
| Llaneta | Filipino | Other Asian | Wong et al. |
| Llaneza | Filipino | Other Asian | Wong et al. |
| Llanillo | Filipino | Other Asian | Wong et al. |
| Llanos | Filipino | Other Asian | Wong et al. |
| Llantada | Filipino | Other Asian | Wong et al. |
| Llantero | Filipino | Other Asian | Wong et al. |
| Llantino | Filipino | Other Asian | Wong et al. |
| Llanto | Filipino | Other Asian | Wong et al. |
| Llapitan | Filipino | Other Asian | Wong et al. |
| Llarena | Filipino | Other Asian | Wong et al. |
| Llarenas | Filipino | Other Asian | Wong et al. |
| Llarinas | Filipino | Other Asian | Wong et al. |
| Llave | Filipino | Other Asian | Wong et al. |
| Llavore | Filipino | Other Asian | Wong et al. |
| Lledo | Filipino | Other Asian | Wong et al. |
| Llego | Filipino | Other Asian | Wong et al. |
| Llemos | Filipino | Other Asian | Wong et al. |
| Llena | Filipino | Other Asian | Wong et al. |
| Llenado | Filipino | Other Asian | Wong et al. |
| Llenos | Filipino | Other Asian | Wong et al. |
| Llerena | Filipino | Other Asian | Wong et al. |
| Llereza | Filipino | Other Asian | Wong et al. |
| Lleva | Filipino | Other Asian | Wong et al. |
| Llido | Filipino | Other Asian | Wong et al. |
| Llobrera | Filipino | Other Asian | Wong et al. |
| Llopis | Filipino | Other Asian | Wong et al. |
| Llora | Filipino | Other Asian | Wong et al. |
| Llorca | Filipino | Other Asian | Wong et al. |
| Lloren | Filipino | Other Asian | Wong et al. |
| Llorente | Filipino | Other Asian | Wong et al. |
| Llorin | Filipino | Other Asian | Wong et al. |
| Lluch | Filipino | Other Asian | Wong et al. |
| Lo | Chinese | Chinese | Kranick et al. |
| Loan | Vietnamese | Other Asian | Wong et al. |
| Loanzon | Filipino | Other Asian | Wong et al. |
| Lobaton | Filipino | Other Asian | Wong et al. |
| Lobitana | Filipino | Other Asian | Wong et al. |
| Lobo | Asian Indian | South Asian | Wong et al. |
| Lobrigo | Filipino | Other Asian | Wong et al. |
| Lobrin | Filipino | Other Asian | Wong et al. |
| Lobrio | Filipino | Other Asian | Wong et al. |
| Lobusta | Filipino | Other Asian | Wong et al. |
| Loc | Vietnamese | Other Asian | Wong et al. |
| Lock | Chinese | Chinese | Wong et al. |
| Locquiao | Filipino | Other Asian | Wong et al. |
| Locsin | Filipino | Other Asian | Wong et al. |
| Lodaya | Asian Indian | South Asian | Wong et al. |
| Lodevico | Filipino | Other Asian | Wong et al. |
| Lodhi | Asian Indian | South Asian | Wong et al. |
| Lodhia | Asian Indian | South Asian | Wong et al. |
| Lodhie | Asian Indian | South Asian | Wong et al. |
| Lodi |  | South Asian | Lim et al. |
| Lofranco | Filipino | Other Asian | Wong et al. |
| Log | Chinese | Chinese | Wong et al. |
| Loganathan | Asian Indian | South Asian | Wong et al. |
| Logarta | Filipino | Other Asian | Wong et al. |
| Logero | Filipino | Other Asian | Wong et al. |
| Logo | Filipino | Other Asian | Wong et al. |
| Logronio | Filipino | Other Asian | Wong et al. |
| Loh | Chinese; Malaysian | Chinese | Kranick et al. |
| Lojo | Filipino | Other Asian | Wong et al. |
| Lok | Chinese | Chinese | Kranick et al. |
| Loke | Chinese | Chinese | Wong et al. |
| Lokenauth |  | South Asian | Lim et al. |
| Lokhande | Asian Indian | South Asian | Wong et al. |
| Lokhandwala | Asian Indian | South Asian | Wong et al. |
| Lolarga | Filipino | Other Asian | Wong et al. |
| Loleng | Filipino | Other Asian | Wong et al. |
| Lom | Chinese | Chinese | Wong et al. |
| Loma | Filipino | Other Asian | Wong et al. |
| Lomahan | Filipino | Other Asian | Wong et al. |
| Lomarda | Filipino | Other Asian | Wong et al. |
| Lombos | Filipino | Other Asian | Wong et al. |
| Lomboy | Filipino | Other Asian | Wong et al. |
| Lomentigar | Filipino | Other Asian | Wong et al. |
| Lomibao | Filipino | Other Asian | Wong et al. |
| Lomongo | Filipino | Other Asian | Wong et al. |
| Lomotan | Filipino | Other Asian | Wong et al. |
| Lomuntad | Filipino | Other Asian | Wong et al. |
| Lon | Chinese | Chinese | Wong et al. |
| Londres | Filipino | Other Asian | Wong et al. |
| Longa | Filipino | Other Asian | Wong et al. |
| Longakit | Filipino | Other Asian | Wong et al. |
| Longboy | Filipino | Other Asian | Wong et al. |
| Longgat | Filipino | Other Asian | Wong et al. |
| Lontoc | Filipino | Other Asian | Wong et al. |
| Lontok | Filipino | Other Asian | Wong et al. |
| Lonzanida | Filipino | Other Asian | Wong et al. |
| Loo | Chinese | Chinese | Kranick et al. |
| Looc | Vietnamese | Other Asian | Wong et al. |
| Look | Chinese | Chinese | Wong et al. |
| Looknauth |  | South Asian | Lim et al. |
| Loomba | Asian Indian | South Asian | Wong et al. |
| Loon | Chinese | Chinese | Wong et al. |
| Loong | Chinese | Chinese | Wong et al. |
| Lope | Filipino | Other Asian | Wong et al. |
| Lopena | Filipino | Other Asian | Wong et al. |
| Lopez | Filipino | Other Asian | Wong et al. |
| Loque | Filipino | Other Asian | Wong et al. |
| Loquellano | Filipino | Other Asian | Wong et al. |
| Lora | Filipino | Other Asian | Wong et al. |
| Lorca | Filipino | Other Asian | Wong et al. |
| Loredo | Filipino | Other Asian | Wong et al. |
| Lorenzana | Filipino | Other Asian | Wong et al. |
| Lorenzo | Filipino | Other Asian | Wong et al. |
| Loresco | Filipino | Other Asian | Wong et al. |
| Loresto | Filipino | Other Asian | Wong et al. |
| Loreto | Filipino | Other Asian | Wong et al. |
| Lorica | Filipino | Other Asian | Wong et al. |
| Lorico | Filipino | Other Asian | Wong et al. |
| Loriega | Filipino | Other Asian | Wong et al. |
| Lorilla | Filipino | Other Asian | Wong et al. |
| Loristo | Filipino | Other Asian | Wong et al. |
| Lorzano | Filipino | Other Asian | Wong et al. |
| Losa | Filipino | Other Asian | Wong et al. |
| Losbanes | Filipino | Other Asian | Wong et al. |
| Losbanos | Filipino | Other Asian | Wong et al. |
| Lota | Filipino | Other Asian | Wong et al. |
| Lotia | Pakistani | South Asian | Kranick et al. |
| Lotuaco | Filipino | Other Asian | Wong et al. |
| Lou | Chinese | Chinese | Wong et al. |
| Louey | Chinese | Chinese | Wong et al. |
| Loui | Chinese | Chinese | Wong et al. |
| Louie | Chinese | Chinese | Wong et al. |
| Louis | Chinese | Chinese | Wong et al. |
| Loung | Chinese | Chinese | Wong et al. |
| Lovan | Vietnamese | Other Asian | Wong et al. |
| Lovie | Chinese | Chinese | Wong et al. |
| Low | Chinese; Malaysian | Chinese | Kranick et al. |
| Loyola | Filipino | Other Asian | Wong et al. |
| Loza | Filipino | Other Asian | Wong et al. |
| Lozada | Filipino | Other Asian | Wong et al. |
| Lozano | Filipino | Other Asian | Wong et al. |
| Lozares | Filipino | Other Asian | Wong et al. |
| Lu | Chinese; Vietnamese | Chinese | Kranick et al. |
| Luague | Filipino | Other Asian | Wong et al. |
| Lualhati | Filipino | Other Asian | Wong et al. |
| Luan | Chinese | Chinese | Wong et al. |
| Luansing | Filipino | Other Asian | Wong et al. |
| Luarca | Filipino | Other Asian | Wong et al. |
| Luat | Filipino | Other Asian | Wong et al. |
| Lubag | Filipino | Other Asian | Wong et al. |
| Lubang | Filipino | Other Asian | Wong et al. |
| Lubao | Filipino | Other Asian | Wong et al. |
| Lubaton | Filipino | Other Asian | Wong et al. |
| Lubiano | Filipino | Other Asian | Wong et al. |
| Lubina | Filipino | Other Asian | Wong et al. |
| Lubong | Filipino | Other Asian | Wong et al. |
| Lubos | Filipino | Other Asian | Wong et al. |
| Lubrica | Filipino | Other Asian | Wong et al. |
| Lubrico | Filipino | Other Asian | Wong et al. |
| Lubrin | Filipino | Other Asian | Wong et al. |
| Lubuguin | Filipino | Other Asian | Wong et al. |
| Luc | Vietnamese | Other Asian | Wong et al. |
| Lucena | Filipino | Other Asian | Wong et al. |
| Lucenara | Filipino | Other Asian | Wong et al. |
| Lucero | Filipino | Other Asian | Wong et al. |
| Luces | Filipino | Other Asian | Wong et al. |
| Luchangco | Filipino | Other Asian | Wong et al. |
| Lucila | Filipino | Other Asian | Wong et al. |
| Lucina | Filipino | Other Asian | Wong et al. |
| Lucindo | Filipino | Other Asian | Wong et al. |
| Lucio | Filipino | Other Asian | Wong et al. |
| Lucky |  | South Asian | Lim et al. |
| Luczon | Filipino | Other Asian | Wong et al. |
| Ludan | Filipino | Other Asian | Wong et al. |
| Ludovice | Filipino | Other Asian | Wong et al. |
| Ludovico | Filipino | Other Asian | Wong et al. |
| Lue | Chinese | Chinese | Wong et al. |
| Luen | Chinese | Chinese | Wong et al. |
| Lueng | Chinese | Chinese | Wong et al. |
| Luey | Chinese | Chinese | Wong et al. |
| Luga | Filipino | Other Asian | Wong et al. |
| Lugay | Filipino | Other Asian | Wong et al. |
| Lugo | Filipino | Other Asian | Wong et al. |
| Lugod | Filipino | Other Asian | Wong et al. |
| Lugtu | Filipino | Other Asian | Wong et al. |
| Lugue | Filipino | Other Asian | Wong et al. |
| Luh | Chinese | Chinese | Wong et al. |
| Luhar | Asian Indian | South Asian | Wong et al. |
| Lui | Chinese | Chinese | Kranick et al. |
| Luib | Filipino | Other Asian | Wong et al. |
| Luis | Filipino | Other Asian | Wong et al. |
| Luistro | Filipino | Other Asian | Wong et al. |
| Luk | Chinese | Chinese | Wong et al. |
| Luka | Asian Indian | South Asian | Wong et al. |
| Lukban | Filipino | Other Asian | Wong et al. |
| Luke | Chinese | Chinese | Wong et al. |
| Lukose | Asian Indian | South Asian | Wong et al. |
| Lulu | Filipino | Other Asian | Wong et al. |
| Lum | Malaysian; Chinese | Other Asian | Kranick et al. |
| Lumaban | Filipino | Other Asian | Wong et al. |
| Lumabao | Filipino | Other Asian | Wong et al. |
| Lumabas | Filipino | Other Asian | Wong et al. |
| Lumagbas | Filipino | Other Asian | Wong et al. |
| Lumague | Filipino | Other Asian | Wong et al. |
| Lumagui | Filipino | Other Asian | Wong et al. |
| Lumampao | Filipino | Other Asian | Wong et al. |
| Lumang | Filipino | Other Asian | Wong et al. |
| Lumanlan | Filipino | Other Asian | Wong et al. |
| Lumanog | Filipino | Other Asian | Wong et al. |
| Lumantas | Filipino | Other Asian | Wong et al. |
| Lumapas | Filipino | Other Asian | Wong et al. |
| Lumasag | Filipino | Other Asian | Wong et al. |
| Lumauig | Filipino | Other Asian | Wong et al. |
| Lumawig | Filipino | Other Asian | Wong et al. |
| Lumayag | Filipino | Other Asian | Wong et al. |
| Lumba | Filipino | Other Asian | Wong et al. |
| Lumbab | Filipino | Other Asian | Wong et al. |
| Lumbang | Filipino | Other Asian | Wong et al. |
| Lumbania | Filipino | Other Asian | Wong et al. |
| Lumbao | Filipino | Other Asian | Wong et al. |
| Lumbera | Filipino | Other Asian | Wong et al. |
| Lumbo | Filipino | Other Asian | Wong et al. |
| Lumbre | Filipino | Other Asian | Wong et al. |
| Lumbres | Filipino | Other Asian | Wong et al. |
| Lumibao | Filipino | Other Asian | Wong et al. |
| Lumicao | Filipino | Other Asian | Wong et al. |
| Luminarias | Filipino | Other Asian | Wong et al. |
| Lumio | Filipino | Other Asian | Wong et al. |
| Lun | Chinese | Chinese | Wong et al. |
| Luna |  | South Asian | Lim et al. |
| Lunar | Filipino | Other Asian | Wong et al. |
| Lunasco | Filipino | Other Asian | Wong et al. |
| Lunasin | Filipino | Other Asian | Wong et al. |
| Lunat | Asian Indian | South Asian | Wong et al. |
| Lunes | Filipino | Other Asian | Wong et al. |
| Lung | Chinese | Chinese | Wong et al. |
| Lungay | Filipino | Other Asian | Wong et al. |
| Luo | Chinese | Chinese | Wong et al. |
| Luon | Vietnamese | Other Asian | Wong et al. |
| Luong | Chinese | Chinese | Kranick et al. |
| Luongvan | Vietnamese | Other Asian | Wong et al. |
| Lupisan | Filipino | Other Asian | Wong et al. |
| Luque | Filipino | Other Asian | Wong et al. |
| Lusoc | Filipino | Other Asian | Wong et al. |
| Luspo | Filipino | Other Asian | Wong et al. |
| Lustado | Filipino | Other Asian | Wong et al. |
| Lustan | Filipino | Other Asian | Wong et al. |
| Lusterio | Filipino | Other Asian | Wong et al. |
| Lustina | Filipino | Other Asian | Wong et al. |
| Lustre | Filipino | Other Asian | Wong et al. |
| Lusung | Filipino | Other Asian | Wong et al. |
| Lutao | Filipino | Other Asian | Wong et al. |
| Lutap | Filipino | Other Asian | Wong et al. |
| Lutero | Filipino | Other Asian | Wong et al. |
| Lutfunnesa |  | South Asian | Lim et al. |
| Luthra | Asian Indian | South Asian | Wong et al. |
| Luu | Vietnamese | Other Asian | Kranick et al. |
| Luyen | Vietnamese | Other Asian | Wong et al. |
| Luz | Filipino | Other Asian | Wong et al. |
| Luzano | Filipino | Other Asian | Wong et al. |
| Luzod | Filipino | Other Asian | Wong et al. |
| Luzon | Filipino | Other Asian | Wong et al. |
| Luzuriaga | Filipino | Other Asian | Wong et al. |
| Ly | Vietnamese | Other Asian | Kranick et al. |
| Lyen | Chinese | Chinese | Wong et al. |
| Lym | Korean | Other Asian | Wong et al. |
| Lyn | Chinese | Chinese | Wong et al. |
| Lyo | Korean | Other Asian | Wong et al. |
| Lyu | Korean | Other Asian | Wong et al. |
| Ma | Chinese | Chinese | Kranick et al. |
| Maa | Chinese | Chinese | Wong et al. |
| Maala | Filipino | Other Asian | Wong et al. |
| Maan | Asian Indian | South Asian | Wong et al. |
| Maano | Filipino | Other Asian | Wong et al. |
| Mabagos | Filipino | Other Asian | Wong et al. |
| Mabalay | Filipino | Other Asian | Wong et al. |
| Mabalot | Filipino | Other Asian | Wong et al. |
| Mabanag | Filipino | Other Asian | Wong et al. |
| Mabanglo | Filipino | Other Asian | Wong et al. |
| Mabansag | Filipino | Other Asian | Wong et al. |
| Mabanta | Filipino | Other Asian | Wong et al. |
| Mabaquiao | Filipino | Other Asian | Wong et al. |
| Mabasa | Filipino | Other Asian | Wong et al. |
| Mabborang | Filipino | Other Asian | Wong et al. |
| Mabesa | Filipino | Other Asian | Wong et al. |
| Mabilangan | Filipino | Other Asian | Wong et al. |
| Mabilog | Filipino | Other Asian | Wong et al. |
| Mabin |  | South Asian | Lim et al. |
| Mabini | Filipino | Other Asian | Wong et al. |
| Mabuchi | Japanese | Other Asian | Wong et al. |
| Mabulay | Filipino | Other Asian | Wong et al. |
| Mabunay | Filipino | Other Asian | Wong et al. |
| Mabunga | Filipino | Other Asian | Wong et al. |
| Mabutas | Filipino | Other Asian | Wong et al. |
| Mabuti | Filipino | Other Asian | Wong et al. |
| Mac | Vietnamese | Other Asian | Wong et al. |
| Macaalay | Filipino | Other Asian | Wong et al. |
| Macababbad | Filipino | Other Asian | Wong et al. |
| Macabagdal | Filipino | Other Asian | Wong et al. |
| Macabales | Filipino | Other Asian | Wong et al. |
| Macabasco | Filipino | Other Asian | Wong et al. |
| Macabata | Filipino | Other Asian | Wong et al. |
| Macabenta | Filipino | Other Asian | Wong et al. |
| Macabeo | Filipino | Other Asian | Wong et al. |
| Macabio | Filipino | Other Asian | Wong et al. |
| Macabiog | Filipino | Other Asian | Wong et al. |
| Macabitas | Filipino | Other Asian | Wong et al. |
| Macabuhay | Filipino | Other Asian | Wong et al. |
| Macabulos | Filipino | Other Asian | Wong et al. |
| Macadaan | Filipino | Other Asian | Wong et al. |
| Macadaeg | Filipino | Other Asian | Wong et al. |
| Macadamia | Filipino | Other Asian | Wong et al. |
| Macadangdang | Filipino | Other Asian | Wong et al. |
| Macagba | Filipino | Other Asian | Wong et al. |
| Macahia | Filipino | Other Asian | Wong et al. |
| Macahilas | Filipino | Other Asian | Wong et al. |
| Macahilig | Filipino | Other Asian | Wong et al. |
| Macaibay | Filipino | Other Asian | Wong et al. |
| Macairan | Filipino | Other Asian | Wong et al. |
| Macaisa | Filipino | Other Asian | Wong et al. |
| Macalalad | Filipino | Other Asian | Wong et al. |
| Macalalag | Filipino | Other Asian | Wong et al. |
| Macalanda | Filipino | Other Asian | Wong et al. |
| Macale | Filipino | Other Asian | Wong et al. |
| Macalinao | Filipino | Other Asian | Wong et al. |
| Macalincag | Filipino | Other Asian | Wong et al. |
| Macalindong | Filipino | Other Asian | Wong et al. |
| Macalino | Filipino | Other Asian | Wong et al. |
| Macalintal | Filipino | Other Asian | Wong et al. |
| Macalisang | Filipino | Other Asian | Wong et al. |
| Macalma | Filipino | Other Asian | Wong et al. |
| Macaltao | Filipino | Other Asian | Wong et al. |
| Macam | Filipino | Other Asian | Wong et al. |
| Macamay | Filipino | Other Asian | Wong et al. |
| Macanas | Filipino | Other Asian | Wong et al. |
| Macandili | Filipino | Other Asian | Wong et al. |
| Macandog | Filipino | Other Asian | Wong et al. |
| Macaoay | Filipino | Other Asian | Wong et al. |
| Macapagal | Filipino | Other Asian | Wong et al. |
| Macapanas | Filipino | Other Asian | Wong et al. |
| Macapinlac | Filipino | Other Asian | Wong et al. |
| Macapugay | Filipino | Other Asian | Wong et al. |
| Macapulay | Filipino | Other Asian | Wong et al. |
| Macaraeg | Filipino | Other Asian | Wong et al. |
| Macaraig | Filipino | Other Asian | Wong et al. |
| Macaranas | Filipino | Other Asian | Wong et al. |
| Macariola | Filipino | Other Asian | Wong et al. |
| Macasa | Filipino | Other Asian | Wong et al. |
| Macasaet | Filipino | Other Asian | Wong et al. |
| Macasero | Filipino | Other Asian | Wong et al. |
| Macasieb | Filipino | Other Asian | Wong et al. |
| Macasinag | Filipino | Other Asian | Wong et al. |
| Macasio | Filipino | Other Asian | Wong et al. |
| Macaso | Filipino | Other Asian | Wong et al. |
| Macaspac | Filipino | Other Asian | Wong et al. |
| Macatangay | Filipino | Other Asian | Wong et al. |
| Macatbag | Filipino | Other Asian | Wong et al. |
| Macatiag | Filipino | Other Asian | Wong et al. |
| Macato | Filipino | Other Asian | Wong et al. |
| Macatula | Filipino | Other Asian | Wong et al. |
| Macatumbas | Filipino | Other Asian | Wong et al. |
| Macavinta | Filipino | Other Asian | Wong et al. |
| Macawile | Filipino | Other Asian | Wong et al. |
| Macawili | Filipino | Other Asian | Wong et al. |
| Macayan | Filipino | Other Asian | Wong et al. |
| Maceda | Filipino | Other Asian | Wong et al. |
| Maceren | Filipino | Other Asian | Wong et al. |
| Mach | Vietnamese | Other Asian | Wong et al. |
| Machhar | Asian Indian | South Asian | Wong et al. |
| Machida | Japanese | Other Asian | Wong et al. |
| Machii | Japanese | Other Asian | Wong et al. |
| Machitar | Filipino | Other Asian | Wong et al. |
| Machuca | Chinese | Chinese | Wong et al. |
| Macias | Filipino | Other Asian | Wong et al. |
| Macion | Filipino | Other Asian | Wong et al. |
| Maclang | Filipino | Other Asian | Wong et al. |
| Macorol | Filipino | Other Asian | Wong et al. |
| Macrohon | Filipino | Other Asian | Wong et al. |
| Mactal | Filipino | Other Asian | Wong et al. |
| Macugay | Filipino | Other Asian | Wong et al. |
| Macusi | Filipino | Other Asian | Wong et al. |
| Macwan | Asian Indian | South Asian | Wong et al. |
| Madahar | Asian Indian | South Asian | Wong et al. |
| Madali | Filipino | Other Asian | Wong et al. |
| Madalipay | Filipino | Other Asian | Wong et al. |
| Madamba | Filipino | Other Asian | Wong et al. |
| Madan | Asian Indian | South Asian | Wong et al. |
| Madani | Asian Indian | South Asian | Wong et al. |
| Madarang | Filipino | Other Asian | Wong et al. |
| Madariaga | Filipino | Other Asian | Wong et al. |
| Madayag | Filipino | Other Asian | Wong et al. |
| Maddagan | Filipino | Other Asian | Wong et al. |
| Maddali | Asian Indian | South Asian | Wong et al. |
| Maddela | Asian Indian | South Asian | Kranick et al. |
| Madduri | Asian Indian | South Asian | Wong et al. |
| Madeja | Filipino | Other Asian | Wong et al. |
| Madela | Filipino | Other Asian | Wong et al. |
| Madelo | Filipino | Other Asian | Wong et al. |
| Madera | Filipino | Other Asian | Wong et al. |
| Maderazo | Filipino | Other Asian | Wong et al. |
| Madhani | Asian Indian | South Asian | Wong et al. |
| Madhav | Asian Indian | South Asian | Wong et al. |
| Madhavan | Asian Indian | South Asian | Wong et al. |
| Madhavbhai | Asian Indian | South Asian | Wong et al. |
| Madhu |  | South Asian | Lim et al. |
| Madla | Filipino | Other Asian | Wong et al. |
| Madlambayan | Filipino | Other Asian | Wong et al. |
| Madlangbayan | Filipino | Other Asian | Wong et al. |
| Madlangsakay | Filipino | Other Asian | Wong et al. |
| Madlansacay | Filipino | Other Asian | Wong et al. |
| Madolora | Filipino | Other Asian | Wong et al. |
| Madray |  | South Asian | Lim et al. |
| Madria | Filipino | Other Asian | Wong et al. |
| Madriaga | Filipino | Other Asian | Wong et al. |
| Madrid | Filipino | Other Asian | Wong et al. |
| Madridejos | Filipino | Other Asian | Wong et al. |
| Madrigal | Filipino | Other Asian | Wong et al. |
| Madrilejo | Filipino | Other Asian | Wong et al. |
| Madrilejos | Filipino | Other Asian | Wong et al. |
| Madrinan | Filipino | Other Asian | Wong et al. |
| Madrona | Filipino | Other Asian | Wong et al. |
| Madronio | Filipino | Other Asian | Wong et al. |
| Maducdoc | Filipino | Other Asian | Wong et al. |
| Madula | Filipino | Other Asian | Wong et al. |
| Madulid | Filipino | Other Asian | Wong et al. |
| Maduro | Filipino | Other Asian | Wong et al. |
| Maeda | Japanese | Other Asian | Wong et al. |
| Maegawa | Japanese | Other Asian | Wong et al. |
| Maehara | Japanese | Other Asian | Wong et al. |
| Maehira | Japanese | Other Asian | Wong et al. |
| Maejima | Japanese | Other Asian | Wong et al. |
| Maekawa | Japanese | Other Asian | Kranick et al. |
| Maemoto | Japanese | Other Asian | Wong et al. |
| Maemura | Japanese | Other Asian | Wong et al. |
| Maeng | Korean | Other Asian | Wong et al. |
| Maeno | Japanese | Other Asian | Wong et al. |
| Maeshima | Japanese | Other Asian | Wong et al. |
| Maeshiro | Japanese | Other Asian | Wong et al. |
| Maestre | Filipino | Other Asian | Wong et al. |
| Maeyama | Japanese | Other Asian | Wong et al. |
| Maezawa | Japanese | Other Asian | Wong et al. |
| Mafune | Japanese | Other Asian | Wong et al. |
| Magabo | Filipino | Other Asian | Wong et al. |
| Magada | Filipino | Other Asian | Wong et al. |
| Magadan | Filipino | Other Asian | Wong et al. |
| Magadia | Filipino | Other Asian | Wong et al. |
| Magahis | Filipino | Other Asian | Wong et al. |
| Magale | Filipino | Other Asian | Wong et al. |
| Magaling | Filipino | Other Asian | Wong et al. |
| Magalit | Filipino | Other Asian | Wong et al. |
| Magallanes | Filipino | Other Asian | Wong et al. |
| Magallano | Filipino | Other Asian | Wong et al. |
| Magallon | Filipino | Other Asian | Wong et al. |
| Magallones | Filipino | Other Asian | Wong et al. |
| Magalona | Filipino | Other Asian | Wong et al. |
| Magalong | Filipino | Other Asian | Wong et al. |
| Magana | Filipino | Other Asian | Wong et al. |
| Maganbhai | Asian Indian | South Asian | Wong et al. |
| Maganlal | Asian Indian | South Asian | Wong et al. |
| Magante | Filipino | Other Asian | Wong et al. |
| Maganti | Asian Indian | South Asian | Wong et al. |
| Magaoay | Filipino | Other Asian | Wong et al. |
| Magarin | Filipino | Other Asian | Wong et al. |
| Magarro | Filipino | Other Asian | Wong et al. |
| Magat | Filipino | Other Asian | Wong et al. |
| Magata | Japanese | Other Asian | Wong et al. |
| Magaway | Filipino | Other Asian | Wong et al. |
| Magayanes | Filipino | Other Asian | Wong et al. |
| Magbag | Filipino | Other Asian | Wong et al. |
| Magbaleta | Filipino | Other Asian | Wong et al. |
| Magbanua | Filipino | Other Asian | Wong et al. |
| Magbitang | Filipino | Other Asian | Wong et al. |
| Magbojos | Filipino | Other Asian | Wong et al. |
| Magboo | Filipino | Other Asian | Wong et al. |
| Magbual | Filipino | Other Asian | Wong et al. |
| Magbuhat | Filipino | Other Asian | Wong et al. |
| Magcalas | Filipino | Other Asian | Wong et al. |
| Magcale | Filipino | Other Asian | Wong et al. |
| Magdael | Filipino | Other Asian | Wong et al. |
| Magdaleno | Filipino | Other Asian | Wong et al. |
| Magdales | Filipino | Other Asian | Wong et al. |
| Magdaluyo | Filipino | Other Asian | Wong et al. |
| Magdamo | Filipino | Other Asian | Wong et al. |
| Magdangal | Filipino | Other Asian | Wong et al. |
| Magdaraog | Filipino | Other Asian | Wong et al. |
| Magdato | Filipino | Other Asian | Wong et al. |
| Magday | Filipino | Other Asian | Wong et al. |
| Magdirila | Filipino | Other Asian | Wong et al. |
| Magdongon | Filipino | Other Asian | Wong et al. |
| Maggay | Filipino | Other Asian | Wong et al. |
| Maghanoy | Filipino | Other Asian | Wong et al. |
| Maghari | Filipino | Other Asian | Wong et al. |
| Maghinay | Filipino | Other Asian | Wong et al. |
| Maghirang | Filipino | Other Asian | Wong et al. |
| Maghuyop | Filipino | Other Asian | Wong et al. |
| Magisa | Filipino | Other Asian | Wong et al. |
| Magistrado | Filipino | Other Asian | Wong et al. |
| Maglalang | Filipino | Other Asian | Wong et al. |
| Maglangit | Filipino | Other Asian | Wong et al. |
| Maglaque | Filipino | Other Asian | Wong et al. |
| Maglaqui | Filipino | Other Asian | Wong et al. |
| Maglasang | Filipino | Other Asian | Wong et al. |
| Maglaya | Filipino | Other Asian | Wong et al. |
| Maglente | Filipino | Other Asian | Wong et al. |
| Magleo | Filipino | Other Asian | Wong et al. |
| Maglinte | Filipino | Other Asian | Wong et al. |
| Magnabijon | Filipino | Other Asian | Wong et al. |
| Magnaye | Filipino | Other Asian | Wong et al. |
| Magnayon | Filipino | Other Asian | Wong et al. |
| Magno | Filipino | Other Asian | Wong et al. |
| Mago | Filipino | Other Asian | Wong et al. |
| Magos | Filipino | Other Asian | Wong et al. |
| Magpale | Filipino | Other Asian | Wong et al. |
| Magpali | Filipino | Other Asian | Wong et al. |
| Magpantay | Filipino | Other Asian | Wong et al. |
| Magpayo | Filipino | Other Asian | Wong et al. |
| Magpile | Filipino | Other Asian | Wong et al. |
| Magpily | Filipino | Other Asian | Wong et al. |
| Magpiong | Filipino | Other Asian | Wong et al. |
| Magpoc | Filipino | Other Asian | Wong et al. |
| Magpuri | Filipino | Other Asian | Wong et al. |
| Magpusao | Filipino | Other Asian | Wong et al. |
| Magracia | Filipino | Other Asian | Wong et al. |
| Magsakay | Filipino | Other Asian | Wong et al. |
| Magsalay | Filipino | Other Asian | Wong et al. |
| Magsalin | Filipino | Other Asian | Wong et al. |
| Magsanay | Filipino | Other Asian | Wong et al. |
| Magsanoc | Filipino | Other Asian | Wong et al. |
| Magsarili | Filipino | Other Asian | Wong et al. |
| Magsayo | Filipino | Other Asian | Wong et al. |
| Magsaysay | Filipino | Other Asian | Wong et al. |
| Magsino | Filipino | Other Asian | Wong et al. |
| Magsipoc | Filipino | Other Asian | Wong et al. |
| Magsombol | Filipino | Other Asian | Wong et al. |
| Magsumbol | Filipino | Other Asian | Wong et al. |
| Magtalas | Filipino | Other Asian | Wong et al. |
| Magtanong | Filipino | Other Asian | Wong et al. |
| Magtibay | Filipino | Other Asian | Wong et al. |
| Magtira | Filipino | Other Asian | Wong et al. |
| Magtoto | Filipino | Other Asian | Wong et al. |
| Magtulis | Filipino | Other Asian | Wong et al. |
| Magturo | Filipino | Other Asian | Wong et al. |
| Maguad | Filipino | Other Asian | Wong et al. |
| Maguigad | Filipino | Other Asian | Wong et al. |
| Magwili | Filipino | Other Asian | Wong et al. |
| Mah | Chinese; Pakistani | Chinese | Kranick et al. |
| Mahabir |  | South Asian | Lim et al. |
| Mahabubalam |  | South Asian | Lim et al. |
| Mahadeo |  | South Asian | Lim et al. |
| Mahadevan | Asian Indian | South Asian | Wong et al. |
| Mahadevappa | Asian Indian | South Asian | Wong et al. |
| Mahadevia | Asian Indian | South Asian | Wong et al. |
| Mahagamage |  | South Asian | Lim et al. |
| Mahajan | Asian Indian | South Asian | Kranick et al. |
| Mahal | Asian Indian | South Asian | Wong et al. |
| Mahale | Asian Indian | South Asian | Wong et al. |
| Mahalingam | Asian Indian | South Asian | Wong et al. |
| Mahamed | Asian Indian | South Asian | Wong et al. |
| Mahanti | Asian Indian | South Asian | Wong et al. |
| Mahanty | Asian Indian | South Asian | Wong et al. |
| Mahapatra | Asian Indian | South Asian | Wong et al. |
| Maharaj | Asian Indian | South Asian | Kranick et al. |
| Mahboob | Asian Indian | South Asian | Wong et al. |
| Mahbub |  | South Asian | Lim et al. |
| Mahbubani | Asian Indian | South Asian | Wong et al. |
| Mahendra | Asian Indian | South Asian | Wong et al. |
| Mahendru | Asian Indian | South Asian | Wong et al. |
| Maheshwari | Asian Indian | South Asian | Wong et al. |
| Maheswaran | Sri Lankan | South Asian | Kranick et al. |
| Mahfuz |  | South Asian | Lim et al. |
| Mahida | Asian Indian | South Asian | Wong et al. |
| Mahil | Asian Indian | South Asian | Wong et al. |
| Mahilum | Filipino | Other Asian | Wong et al. |
| Mahinay | Filipino | Other Asian | Wong et al. |
| Mahindru | Asian Indian | South Asian | Wong et al. |
| Mahmood | Bangladeshi | South Asian | Kranick et al. |
| Mahmud | Bangladeshi | South Asian | Kranick et al. |
| Mahmuda |  | South Asian | Lim et al. |
| Mahomed | Asian Indian | South Asian | Wong et al. |
| Mahor | Filipino | Other Asian | Wong et al. |
| Mahroof | Pakistani; Sri Lankan | South Asian | Kranick et al. |
| Mahtab | Asian Indian | South Asian | Wong et al. |
| Mahusay | Filipino | Other Asian | Wong et al. |
| Mahzabin |  | South Asian | Lim et al. |
| Mai | Chinese | Chinese | Kranick et al. |
| Maico | Filipino | Other Asian | Wong et al. |
| Maigue | Filipino | Other Asian | Wong et al. |
| Maikal |  | South Asian | Lim et al. |
| Mainkar | Asian Indian | South Asian | Wong et al. |
| Maiquez | Filipino | Other Asian | Wong et al. |
| Maitra | Asian Indian | South Asian | Wong et al. |
| Maity | Asian Indian | South Asian | Wong et al. |
| Maiz | Filipino | Other Asian | Wong et al. |
| Maja | Japanese | Other Asian | Wong et al. |
| Majam | Filipino | Other Asian | Wong et al. |
| Majeed | Asian Indian | South Asian | Wong et al. |
| Majima | Japanese | Other Asian | Wong et al. |
| Majithia | Asian Indian | South Asian | Wong et al. |
| Majmudar | Asian Indian | South Asian | Wong et al. |
| Majmundar | Asian Indian | South Asian | Wong et al. |
| Majul | Filipino | Other Asian | Wong et al. |
| Majumdar | Asian Indian | South Asian | Wong et al. |
| Majumder | Bangladeshi; Asian Indian | South Asian | Kranick et al. |
| Mak | Chinese | Chinese | Kranick et al. |
| Makabali | Filipino | Other Asian | Kranick et al. |
| Makabe | Japanese | Other Asian | Wong et al. |
| Makabenta | Filipino | Other Asian | Wong et al. |
| Makadia | Asian Indian | South Asian | Wong et al. |
| Makalintal | Filipino | Other Asian | Wong et al. |
| Makam | Asian Indian | South Asian | Wong et al. |
| Makanbhai | Asian Indian | South Asian | Wong et al. |
| Makani | Asian Indian | South Asian | Wong et al. |
| Makanji | Asian Indian | South Asian | Wong et al. |
| Makapugay | Filipino | Other Asian | Wong et al. |
| Makarchand |  | South Asian | Lim et al. |
| Makati | Asian Indian | South Asian | Wong et al. |
| Makayan | Filipino | Other Asian | Wong et al. |
| Makda | Asian Indian | South Asian | Wong et al. |
| Makhani | Asian Indian | South Asian | Wong et al. |
| Makihara | Japanese | Other Asian | Wong et al. |
| Makiling | Filipino | Other Asian | Wong et al. |
| Makim | Asian Indian | South Asian | Wong et al. |
| Makimoto | Japanese | Other Asian | Wong et al. |
| Makimura | Japanese | Other Asian | Wong et al. |
| Makino | Japanese | Other Asian | Wong et al. |
| Makise | Japanese | Other Asian | Wong et al. |
| Makishi | Japanese | Other Asian | Wong et al. |
| Makishima | Japanese | Other Asian | Wong et al. |
| Makita | Japanese | Other Asian | Wong et al. |
| Makiuchi | Japanese | Other Asian | Wong et al. |
| Makiyama | Japanese | Other Asian | Wong et al. |
| Maknojia | Asian Indian | South Asian | Wong et al. |
| Maknun |  | South Asian | Lim et al. |
| Makwana | Asian Indian | South Asian | Wong et al. |
| Mal | Asian Indian | South Asian | Wong et al. |
| Malabanan | Filipino | Other Asian | Wong et al. |
| Malabed | Filipino | Other Asian | Wong et al. |
| Malabrigo | Filipino | Other Asian | Wong et al. |
| Malaca | Filipino | Other Asian | Wong et al. |
| Malacad | Filipino | Other Asian | Wong et al. |
| Malacas | Filipino | Other Asian | Wong et al. |
| Maladwala | Asian Indian | South Asian | Wong et al. |
| Malag | Filipino | Other Asian | Wong et al. |
| Malaga | Filipino | Other Asian | Wong et al. |
| Malagar | Filipino | Other Asian | Wong et al. |
| Malaki | Filipino | Other Asian | Wong et al. |
| Malalis | Filipino | Other Asian | Wong et al. |
| Malaluan | Filipino | Other Asian | Wong et al. |
| Malamug | Filipino | Other Asian | Wong et al. |
| Malana | Filipino | Other Asian | Wong et al. |
| Malanay | Filipino | Other Asian | Wong et al. |
| Malang | Filipino | Other Asian | Wong et al. |
| Malani | Asian Indian | South Asian | Wong et al. |
| Malano | Filipino | Other Asian | Wong et al. |
| Malantic | Filipino | Other Asian | Wong et al. |
| Malanum | Filipino | Other Asian | Wong et al. |
| Malapira | Filipino | Other Asian | Wong et al. |
| Malapit | Filipino | Other Asian | Wong et al. |
| Malapitan | Filipino | Other Asian | Wong et al. |
| Malaque | Filipino | Other Asian | Wong et al. |
| Malaqui | Filipino | Other Asian | Wong et al. |
| Malasaga | Filipino | Other Asian | Wong et al. |
| Malasig | Filipino | Other Asian | Wong et al. |
| Malata | Filipino | Other Asian | Wong et al. |
| Malatag | Filipino | Other Asian | Wong et al. |
| Malate | Filipino | Other Asian | Wong et al. |
| Malaviya | Asian Indian | South Asian | Wong et al. |
| Malayang | Filipino | Other Asian | Wong et al. |
| Malayil | Asian Indian | South Asian | Wong et al. |
| Malazarte | Filipino | Other Asian | Wong et al. |
| Malazo | Filipino | Other Asian | Wong et al. |
| Malbari | Asian Indian | South Asian | Wong et al. |
| Malbas | Filipino | Other Asian | Wong et al. |
| Malbog | Filipino | Other Asian | Wong et al. |
| Maldia | Filipino | Other Asian | Wong et al. |
| Maldonado | Filipino | Other Asian | Wong et al. |
| Malek | Asian Indian | South Asian | Kranick et al. |
| Malgapo | Filipino | Other Asian | Wong et al. |
| Malhan | Asian Indian | South Asian | Wong et al. |
| Malhi | Asian Indian | South Asian | Wong et al. |
| Malhotra | Asian Indian | South Asian | Wong et al. |
| Malibago | Filipino | Other Asian | Wong et al. |
| Malibiran | Filipino | Other Asian | Wong et al. |
| Malibunas | Filipino | Other Asian | Wong et al. |
| Malic | Filipino | Other Asian | Wong et al. |
| Malicad | Filipino | Other Asian | Wong et al. |
| Malicay | Filipino | Other Asian | Wong et al. |
| Malicdem | Filipino | Other Asian | Wong et al. |
| Malicsi | Filipino | Other Asian | Wong et al. |
| Malig | Filipino | Other Asian | Kranick et al. |
| Maligalig | Filipino | Other Asian | Wong et al. |
| Maligaso | Filipino | Other Asian | Wong et al. |
| Maligaya | Filipino | Other Asian | Wong et al. |
| Malijan | Filipino | Other Asian | Wong et al. |
| Malik | Asian Indian; Bangladeshi; Indonesian | South Asian | Kranick et al. |
| Maliksi | Filipino | Other Asian | Wong et al. |
| Malilay | Filipino | Other Asian | Wong et al. |
| Malimban | Filipino | Other Asian | Wong et al. |
| Malinao | Filipino | Other Asian | Wong et al. |
| Malinay | Filipino | Other Asian | Wong et al. |
| Malinis | Filipino | Other Asian | Wong et al. |
| Malinit | Filipino | Other Asian | Wong et al. |
| Malino | Filipino | Other Asian | Wong et al. |
| Malique |  | South Asian | Lim et al. |
| Malit | Filipino | Other Asian | Wong et al. |
| Maliwanag | Filipino | Other Asian | Wong et al. |
| Maliwat | Filipino | Other Asian | Wong et al. |
| Malixi | Filipino | Other Asian | Wong et al. |
| Malkan | Asian Indian | South Asian | Wong et al. |
| Malla |  | South Asian | Lim et al. |
| Malladi | Asian Indian | South Asian | Wong et al. |
| Mallare | Filipino | Other Asian | Wong et al. |
| Mallari | Filipino | Other Asian | Wong et al. |
| Malliik |  | South Asian | Lim et al. |
| Mallik | Asian Indian | South Asian | Wong et al. |
| Mallillin | Filipino | Other Asian | Wong et al. |
| Mallonga | Filipino | Other Asian | Wong et al. |
| Mallorca | Filipino | Other Asian | Wong et al. |
| Mallya | Asian Indian | South Asian | Wong et al. |
| Malmotra | Asian Indian | South Asian | Wong et al. |
| Maloco | Filipino | Other Asian | Wong et al. |
| Maloles | Filipino | Other Asian | Wong et al. |
| Malolos | Filipino | Other Asian | Wong et al. |
| Malolot | Filipino | Other Asian | Wong et al. |
| Malonda | Filipino | Other Asian | Wong et al. |
| Malong | Filipino | Other Asian | Wong et al. |
| Malonzo | Filipino | Other Asian | Wong et al. |
| Maloyo | Filipino | Other Asian | Wong et al. |
| Maltezo | Filipino | Other Asian | Wong et al. |
| Malto | Asian Indian | South Asian | Kranick et al. |
| Malubag | Filipino | Other Asian | Wong et al. |
| Malubay | Filipino | Other Asian | Wong et al. |
| Malunao | Filipino | Other Asian | Wong et al. |
| Malunes | Filipino | Other Asian | Wong et al. |
| Malupa | Filipino | Other Asian | Wong et al. |
| Maluto | Filipino | Other Asian | Wong et al. |
| Maluyo | Filipino | Other Asian | Wong et al. |
| Malvar | Filipino | Other Asian | Wong et al. |
| Malvas | Filipino | Other Asian | Wong et al. |
| Malveda | Filipino | Other Asian | Wong et al. |
| Mamaat | Filipino | Other Asian | Wong et al. |
| Mamaclay | Filipino | Other Asian | Wong et al. |
| Mamada | Japanese | Other Asian | Wong et al. |
| Mamaed | Filipino | Other Asian | Wong et al. |
| Mamangon | Filipino | Other Asian | Wong et al. |
| Mamangun | Filipino | Other Asian | Wong et al. |
| Mamaradlo | Filipino | Other Asian | Wong et al. |
| Mamaril | Filipino | Other Asian | Wong et al. |
| Mamasig | Filipino | Other Asian | Wong et al. |
| Mamauag | Filipino | Other Asian | Wong et al. |
| Mamawal | Filipino | Other Asian | Wong et al. |
| Mamdani | Asian Indian | South Asian | Wong et al. |
| Mamenta | Filipino | Other Asian | Wong et al. |
| Mamerto | Filipino | Other Asian | Wong et al. |
| Maminta | Filipino | Other Asian | Wong et al. |
| Mamiya | Japanese | Other Asian | Wong et al. |
| Mamon | Filipino | Other Asian | Wong et al. |
| Mamoon |  | South Asian | Lim et al. |
| Mamuad | Filipino | Other Asian | Wong et al. |
| Mamucod | Filipino | Other Asian | Wong et al. |
| Mamun |  | South Asian | Lim et al. |
| Mamuri | Filipino | Other Asian | Wong et al. |
| Mamuyac | Filipino | Other Asian | Wong et al. |
| Man | Chinese | Chinese | Wong et al. |
| Mana | Filipino | Other Asian | Wong et al. |
| Manaay | Filipino | Other Asian | Wong et al. |
| Manabat | Filipino | Other Asian | Wong et al. |
| Manabe | Japanese | Other Asian | Wong et al. |
| Manacap | Filipino | Other Asian | Wong et al. |
| Manacop | Filipino | Other Asian | Wong et al. |
| Manacsa | Filipino | Other Asian | Wong et al. |
| Managbanag | Filipino | Other Asian | Wong et al. |
| Manago | Filipino | Other Asian | Wong et al. |
| Manahan | Filipino | Other Asian | Wong et al. |
| Manaig | Filipino | Other Asian | Wong et al. |
| Manaka | Japanese | Other Asian | Wong et al. |
| Manaktala | Asian Indian | South Asian | Wong et al. |
| Manalac | Filipino | Other Asian | Wong et al. |
| Manalad | Filipino | Other Asian | Wong et al. |
| Manalang | Filipino | Other Asian | Wong et al. |
| Manalansan | Filipino | Other Asian | Wong et al. |
| Manalastas | Filipino | Other Asian | Wong et al. |
| Manalaysay | Filipino | Other Asian | Wong et al. |
| Manalese | Filipino | Other Asian | Wong et al. |
| Manaligod | Filipino | Other Asian | Wong et al. |
| Manalili | Filipino | Other Asian | Wong et al. |
| Manalo | Filipino | Other Asian | Wong et al. |
| Manalon | Filipino | Other Asian | Wong et al. |
| Manaloto | Filipino | Other Asian | Wong et al. |
| Manangan | Filipino | Other Asian | Wong et al. |
| Mananghaya | Filipino | Other Asian | Wong et al. |
| Manankil | Filipino | Other Asian | Wong et al. |
| Mananquil | Filipino | Other Asian | Wong et al. |
| Manansala | Filipino | Other Asian | Wong et al. |
| Manantan | Filipino | Other Asian | Wong et al. |
| Mananzan | Filipino | Other Asian | Wong et al. |
| Manaoat | Filipino | Other Asian | Wong et al. |
| Manaog | Filipino | Other Asian | Wong et al. |
| Manaois | Filipino | Other Asian | Wong et al. |
| Manapat | Filipino | Other Asian | Wong et al. |
| Manapol | Filipino | Other Asian | Wong et al. |
| Manapsal | Filipino | Other Asian | Wong et al. |
| Manarang | Filipino | Other Asian | Wong et al. |
| Manares | Filipino | Other Asian | Wong et al. |
| Manarpaac | Filipino | Other Asian | Wong et al. |
| Manas | Filipino | Other Asian | Wong et al. |
| Manasan | Filipino | Other Asian | Wong et al. |
| Manatad | Filipino | Other Asian | Wong et al. |
| Manay | Filipino | Other Asian | Wong et al. |
| Manayan | Filipino | Other Asian | Wong et al. |
| Mancao | Filipino | Other Asian | Wong et al. |
| Mancenido | Filipino | Other Asian | Wong et al. |
| Manchanda | Asian Indian | South Asian | Wong et al. |
| Mancilla | Filipino | Other Asian | Wong et al. |
| Mandac | Filipino | Other Asian | Wong et al. |
| Mandai | Japanese | Other Asian | Wong et al. |
| Mandal | Asian Indian | South Asian | Wong et al. |
| Mandalaywala | Asian Indian | South Asian | Wong et al. |
| Mandalia | Asian Indian | South Asian | Wong et al. |
| Mandanas | Filipino | Other Asian | Wong et al. |
| Mandani | Filipino | Other Asian | Wong et al. |
| Mandap | Filipino | Other Asian | Wong et al. |
| Mandapat | Filipino | Other Asian | Wong et al. |
| Mandava | Asian Indian | South Asian | Wong et al. |
| Mandawe | Filipino | Other Asian | Wong et al. |
| Mandigma | Filipino | Other Asian | Wong et al. |
| Mandilag | Filipino | Other Asian | Wong et al. |
| Mandin | Filipino | Other Asian | Wong et al. |
| Manding | Filipino | Other Asian | Wong et al. |
| Mandocdoc | Filipino | Other Asian | Wong et al. |
| Mandol |  | South Asian | Lim et al. |
| Mandrekar | Asian Indian | South Asian | Kranick et al. |
| Mandrial | Filipino | Other Asian | Wong et al. |
| Mane | Asian Indian | South Asian | Kranick et al. |
| Maneclang | Filipino | Other Asian | Wong et al. |
| Manegdeg | Filipino | Other Asian | Wong et al. |
| Maneja | Filipino | Other Asian | Wong et al. |
| Manek | Asian Indian | South Asian | Wong et al. |
| Manela | Filipino | Other Asian | Wong et al. |
| Manera | Filipino | Other Asian | Wong et al. |
| Maneru | Filipino | Other Asian | Wong et al. |
| Manese | Filipino | Other Asian | Wong et al. |
| Manga | Filipino | Other Asian | Wong et al. |
| Mangabat | Filipino | Other Asian | Wong et al. |
| Mangaccat | Filipino | Other Asian | Wong et al. |
| Mangahas | Filipino | Other Asian | Wong et al. |
| Mangahis | Filipino | Other Asian | Wong et al. |
| Mangal | Asian Indian | South Asian | Wong et al. |
| Mangali | Filipino | Other Asian | Wong et al. |
| Mangaliag | Filipino | Other Asian | Wong et al. |
| Mangaliman | Filipino | Other Asian | Wong et al. |
| Mangalindan | Filipino | Other Asian | Wong et al. |
| Manganaan | Filipino | Other Asian | Wong et al. |
| Manganti | Filipino | Other Asian | Wong et al. |
| Mangao | Filipino | Other Asian | Wong et al. |
| Mangaoang | Filipino | Other Asian | Wong et al. |
| Mangaoil | Filipino | Other Asian | Wong et al. |
| Mangapit | Filipino | Other Asian | Wong et al. |
| Mangar |  | South Asian | Lim et al. |
| Mangaser | Filipino | Other Asian | Wong et al. |
| Mangasi | Filipino | Other Asian | Wong et al. |
| Mangat | Asian Indian | South Asian | Wong et al. |
| Mangawang | Filipino | Other Asian | Wong et al. |
| Mangayayam | Filipino | Other Asian | Wong et al. |
| Mangibin | Filipino | Other Asian | Wong et al. |
| Mangila | Filipino | Other Asian | Wong et al. |
| Mangiliman | Filipino | Other Asian | Wong et al. |
| Mangilit | Filipino | Other Asian | Wong et al. |
| Mangio | Filipino | Other Asian | Wong et al. |
| Mangla | Asian Indian | South Asian | Wong et al. |
| Manglallan | Filipino | Other Asian | Wong et al. |
| Manglicmot | Filipino | Other Asian | Wong et al. |
| Mangligot | Filipino | Other Asian | Wong et al. |
| Manglinong | Filipino | Other Asian | Wong et al. |
| Mangoba | Filipino | Other Asian | Wong et al. |
| Mangohig | Filipino | Other Asian | Wong et al. |
| Mangona | Filipino | Other Asian | Wong et al. |
| Mangonon | Filipino | Other Asian | Wong et al. |
| Mangosing | Filipino | Other Asian | Wong et al. |
| Mangrobang | Filipino | Other Asian | Wong et al. |
| Mangrubang | Filipino | Other Asian | Wong et al. |
| Mangrulkar |  | South Asian | Lim et al. |
| Mangsat | Filipino | Other Asian | Wong et al. |
| Mangubat | Filipino | Other Asian | Wong et al. |
| Manguera | Filipino | Other Asian | Wong et al. |
| Manguerra | Filipino | Other Asian | Wong et al. |
| Manguiat | Filipino | Other Asian | Wong et al. |
| Mangulabnan | Filipino | Other Asian | Wong et al. |
| Mangundayao | Filipino | Other Asian | Wong et al. |
| Mangune | Filipino | Other Asian | Wong et al. |
| Manh | Vietnamese | Other Asian | Wong et al. |
| Manhas | Asian Indian | South Asian | Wong et al. |
| Mani | Asian Indian | South Asian | Wong et al. |
| Maniago | Filipino | Other Asian | Wong et al. |
| Maniar | Asian Indian | South Asian | Wong et al. |
| Maniaul | Filipino | Other Asian | Wong et al. |
| Manibhai | Asian Indian | South Asian | Wong et al. |
| Manibo | Filipino | Other Asian | Wong et al. |
| Manibog | Filipino | Other Asian | Wong et al. |
| Manickam | Asian Indian | South Asian | Wong et al. |
| Maniego | Filipino | Other Asian | Wong et al. |
| Manigbas | Filipino | Other Asian | Wong et al. |
| Manik |  | South Asian | Lim et al. |
| Manikan | Filipino | Other Asian | Wong et al. |
| Manikis | Filipino | Other Asian | Wong et al. |
| Manila | Filipino | Other Asian | Wong et al. |
| Manilal | Asian Indian | South Asian | Wong et al. |
| Manilall |  | South Asian | Lim et al. |
| Manimala | Asian Indian | South Asian | Wong et al. |
| Manimtim | Filipino | Other Asian | Wong et al. |
| Maninang | Filipino | Other Asian | Wong et al. |
| Maningas | Filipino | Other Asian | Wong et al. |
| Maningat | Filipino | Other Asian | Wong et al. |
| Maningding | Filipino | Other Asian | Wong et al. |
| Maningo | Filipino | Other Asian | Wong et al. |
| Manio | Filipino | Other Asian | Wong et al. |
| Manipis | Filipino | Other Asian | Wong et al. |
| Manipol | Filipino | Other Asian | Wong et al. |
| Manipon | Filipino | Other Asian | Wong et al. |
| Manipula | Filipino | Other Asian | Wong et al. |
| Maniquis | Filipino | Other Asian | Wong et al. |
| Maniquiz | Filipino | Other Asian | Wong et al. |
| Maniruzzaman |  | South Asian | Lim et al. |
| Maniti | Filipino | Other Asian | Wong et al. |
| Maniulit | Filipino | Other Asian | Wong et al. |
| Maniwa | Japanese | Other Asian | Wong et al. |
| Maniyar | Asian Indian | South Asian | Wong et al. |
| Manjares | Filipino | Other Asian | Wong et al. |
| Manji | Asian Indian | South Asian | Wong et al. |
| Manjrekar | Asian Indian | South Asian | Wong et al. |
| Manju |  | South Asian | Lim et al. |
| Mankad | Asian Indian | South Asian | Wong et al. |
| Mankikar | Asian Indian | South Asian | Wong et al. |
| Manku | Asian Indian | South Asian | Wong et al. |
| Manlangit | Filipino | Other Asian | Wong et al. |
| Manlapas | Filipino | Other Asian | Wong et al. |
| Manlapaz | Filipino | Other Asian | Wong et al. |
| Manlapig | Filipino | Other Asian | Wong et al. |
| Manlavi | Filipino | Other Asian | Wong et al. |
| Manliclic | Filipino | Other Asian | Wong et al. |
| Manligas | Filipino | Other Asian | Wong et al. |
| Manlogon | Filipino | Other Asian | Wong et al. |
| Manlongat | Filipino | Other Asian | Wong et al. |
| Manlosa | Filipino | Other Asian | Wong et al. |
| Manlucu | Filipino | Other Asian | Wong et al. |
| Manlulu | Filipino | Other Asian | Wong et al. |
| Manlunas | Filipino | Other Asian | Wong et al. |
| Manlutac | Filipino | Other Asian | Wong et al. |
| Manmano | Filipino | Other Asian | Wong et al. |
| Mann |  | South Asian | Lim et al. |
| Mannaf |  | South Asian | Lim et al. |
| Mannan |  | South Asian | Lim et al. |
| Mano | Japanese | Other Asian | Wong et al. |
| Manocha | Asian Indian | South Asian | Wong et al. |
| Manohar | Asian Indian | South Asian | Wong et al. |
| Manoharan | Asian Indian | South Asian | Wong et al. |
| Manoharlall |  | South Asian | Lim et al. |
| Manongdo | Filipino | Other Asian | Wong et al. |
| Manongsong | Filipino | Other Asian | Wong et al. |
| Manosa | Filipino | Other Asian | Wong et al. |
| Manosca | Filipino | Other Asian | Wong et al. |
| Manoto | Filipino | Other Asian | Wong et al. |
| Manrique | Filipino | Other Asian | Wong et al. |
| Manriquez | Filipino | Other Asian | Wong et al. |
| Mansharamani | Asian Indian | South Asian | Wong et al. |
| Mansilla | Filipino | Other Asian | Wong et al. |
| Mansuri | Asian Indian | South Asian | Wong et al. |
| Mantaring | Filipino | Other Asian | Wong et al. |
| Mante | Filipino | Other Asian | Wong et al. |
| Mantes | Filipino | Other Asian | Wong et al. |
| Mantilla | Filipino | Other Asian | Wong et al. |
| Manto | Filipino | Other Asian | Wong et al. |
| Mantri | Asian Indian | South Asian | Wong et al. |
| Mantua | Filipino | Other Asian | Wong et al. |
| Mantuano | Filipino | Other Asian | Wong et al. |
| Manubag | Filipino | Other Asian | Wong et al. |
| Manubay | Filipino | Other Asian | Wong et al. |
| Manucal | Filipino | Other Asian | Wong et al. |
| Manucdoc | Filipino | Other Asian | Wong et al. |
| Manuel | Filipino | Other Asian | Wong et al. |
| Manugo | Filipino | Other Asian | Wong et al. |
| Manuit | Filipino | Other Asian | Wong et al. |
| Manuja | Asian Indian | South Asian | Wong et al. |
| Manulat | Filipino | Other Asian | Wong et al. |
| Manulid | Filipino | Other Asian | Wong et al. |
| Manuntag | Filipino | Other Asian | Wong et al. |
| Manuud | Filipino | Other Asian | Wong et al. |
| Manuyag | Filipino | Other Asian | Wong et al. |
| Manuzon | Filipino | Other Asian | Wong et al. |
| Manyam | Asian Indian | South Asian | Wong et al. |
| Manzan | Filipino | Other Asian | Wong et al. |
| Manzanal | Filipino | Other Asian | Wong et al. |
| Manzanares | Filipino | Other Asian | Wong et al. |
| Manzanero | Filipino | Other Asian | Wong et al. |
| Manzanilla | Filipino | Other Asian | Wong et al. |
| Manzanillo | Filipino | Other Asian | Wong et al. |
| Manzano | Filipino | Other Asian | Wong et al. |
| Manzo | Filipino | Other Asian | Wong et al. |
| Manzon | Filipino | Other Asian | Wong et al. |
| Manzoor | Asian Indian | South Asian | Wong et al. |
| Mao | Chinese | Chinese | Kranick et al. |
| Maola |  | South Asian | Lim et al. |
| Mapa | Filipino | Other Asian | Wong et al. |
| Mapalad | Filipino | Other Asian | Wong et al. |
| Mapalo | Filipino | Other Asian | Wong et al. |
| Mapanao | Filipino | Other Asian | Wong et al. |
| Mapanoo | Filipino | Other Asian | Wong et al. |
| Mapara | Asian Indian | South Asian | Wong et al. |
| Mapaye | Filipino | Other Asian | Wong et al. |
| Mape | Filipino | Other Asian | Wong et al. |
| Mapeso | Filipino | Other Asian | Wong et al. |
| Mapile | Filipino | Other Asian | Wong et al. |
| Mapili | Filipino | Other Asian | Wong et al. |
| Mapote | Filipino | Other Asian | Wong et al. |
| Mapoy | Filipino | Other Asian | Wong et al. |
| Mappala | Filipino | Other Asian | Wong et al. |
| Mapua | Filipino | Other Asian | Wong et al. |
| Maputi | Filipino | Other Asian | Wong et al. |
| Maqbool | Asian Indian | South Asian | Wong et al. |
| Maque | Filipino | Other Asian | Wong et al. |
| Maquera | Filipino | Other Asian | Wong et al. |
| Maqui | Filipino | Other Asian | Wong et al. |
| Maquilan | Filipino | Other Asian | Wong et al. |
| Maquiling | Filipino | Other Asian | Wong et al. |
| Mar | Chinese | Chinese | Wong et al. |
| Maraan | Filipino | Other Asian | Wong et al. |
| Maradia | Asian Indian | South Asian | Wong et al. |
| Maragay | Filipino | Other Asian | Wong et al. |
| Maraj |  | South Asian | Lim et al. |
| Marajas | Filipino | Other Asian | Wong et al. |
| Maralit | Filipino | Other Asian | Wong et al. |
| Maramag | Filipino | Other Asian | Wong et al. |
| Maramba | Filipino | Other Asian | Wong et al. |
| Marana | Filipino | Other Asian | Wong et al. |
| Maranan | Filipino | Other Asian | Wong et al. |
| Maranion | Filipino | Other Asian | Wong et al. |
| Maranon | Filipino | Other Asian | Wong et al. |
| Marantan | Filipino | Other Asian | Wong et al. |
| Marapao | Filipino | Other Asian | Wong et al. |
| Mararac | Filipino | Other Asian | Wong et al. |
| Marasigan | Filipino | Other Asian | Kranick et al. |
| Maratas | Filipino | Other Asian | Wong et al. |
| Marathe | Asian Indian | South Asian | Wong et al. |
| Marave | Filipino | Other Asian | Wong et al. |
| Maravilla | Filipino | Other Asian | Wong et al. |
| Maravillas | Filipino | Other Asian | Wong et al. |
| Maraya | Filipino | Other Asian | Wong et al. |
| Marayag | Filipino | Other Asian | Wong et al. |
| Marbas | Filipino | Other Asian | Wong et al. |
| Marbella | Filipino | Other Asian | Kranick et al. |
| Marcaida | Filipino | Other Asian | Wong et al. |
| Marcelino | Filipino | Other Asian | Wong et al. |
| Marcellana | Filipino | Other Asian | Wong et al. |
| Marcelo | Filipino | Other Asian | Kranick et al. |
| Marces | Filipino | Other Asian | Wong et al. |
| Marchadesch | Filipino | Other Asian | Wong et al. |
| Marchan | Filipino | Other Asian | Wong et al. |
| Marcia | Filipino | Other Asian | Wong et al. |
| Marcial | Filipino | Other Asian | Wong et al. |
| Marcos | Filipino | Other Asian | Wong et al. |
| Maredia | Asian Indian | South Asian | Wong et al. |
| Marfa | Filipino | Other Asian | Wong et al. |
| Marfatia | Asian Indian | South Asian | Wong et al. |
| Marfil | Filipino | Other Asian | Wong et al. |
| Marfori | Filipino | Other Asian | Wong et al. |
| Margallo | Filipino | Other Asian | Wong et al. |
| Margarejo | Filipino | Other Asian | Wong et al. |
| Margate | Filipino | Other Asian | Wong et al. |
| Marguez | Filipino | Other Asian | Wong et al. |
| Mari | Filipino | Other Asian | Wong et al. |
| Maria | Filipino | Other Asian | Wong et al. |
| Mariano | Filipino | Other Asian | Kranick et al. |
| Maribao | Filipino | Other Asian | Wong et al. |
| Marifosque | Filipino | Other Asian | Wong et al. |
| Marigmen | Filipino | Other Asian | Wong et al. |
| Marigomen | Filipino | Other Asian | Wong et al. |
| Marigza | Filipino | Other Asian | Wong et al. |
| Marilao | Filipino | Other Asian | Wong et al. |
| Marilla | Filipino | Other Asian | Wong et al. |
| Marimla | Filipino | Other Asian | Wong et al. |
| Marimon | Filipino | Other Asian | Wong et al. |
| Marimuthu |  | South Asian | Lim et al. |
| Marin | Filipino | Other Asian | Wong et al. |
| Marina |  | South Asian | Lim et al. |
| Marinas | Filipino | Other Asian | Kranick et al. |
| Marinda | Filipino | Other Asian | Wong et al. |
| Marinduque | Filipino | Other Asian | Wong et al. |
| Mario | Filipino | Other Asian | Wong et al. |
| Maristela | Filipino | Other Asian | Wong et al. |
| Mariveles | Filipino | Other Asian | Wong et al. |
| Mark | Chinese | Chinese | Wong et al. |
| Markar | Asian Indian | South Asian | Wong et al. |
| Markose | Asian Indian | South Asian | Wong et al. |
| Marmeto | Filipino | Other Asian | Wong et al. |
| Marmito | Filipino | Other Asian | Wong et al. |
| Marmol | Filipino | Other Asian | Wong et al. |
| Marmolejo | Filipino | Other Asian | Wong et al. |
| Marpa | Filipino | Other Asian | Wong et al. |
| Marqueses | Filipino | Other Asian | Wong et al. |
| Marquez | Filipino | Other Asian | Wong et al. |
| Marquina | Filipino | Other Asian | Wong et al. |
| Marquinez | Filipino | Other Asian | Wong et al. |
| Marrero | Filipino | Other Asian | Wong et al. |
| Marte | Filipino | Other Asian | Wong et al. |
| Marteja | Filipino | Other Asian | Wong et al. |
| Martelino | Filipino | Other Asian | Wong et al. |
| Martija | Filipino | Other Asian | Wong et al. |
| Martillano | Filipino | Other Asian | Wong et al. |
| Martines | Filipino | Other Asian | Wong et al. |
| Martinez | Filipino | Other Asian | Wong et al. |
| Martir | Filipino | Other Asian | Wong et al. |
| Martires | Filipino | Other Asian | Wong et al. |
| Martirez | Filipino | Other Asian | Wong et al. |
| Martos | Filipino | Other Asian | Wong et al. |
| Maru | Asian Indian | South Asian | Wong et al. |
| Marubayashi | Japanese | Other Asian | Wong et al. |
| Marucut | Filipino | Other Asian | Wong et al. |
| Maruhashi | Japanese | Other Asian | Wong et al. |
| Marui | Japanese | Other Asian | Wong et al. |
| Marukawa | Japanese | Other Asian | Wong et al. |
| Marumo | Japanese | Other Asian | Wong et al. |
| Marumoto | Japanese | Other Asian | Wong et al. |
| Maruo | Japanese | Other Asian | Wong et al. |
| Maruoka | Japanese | Other Asian | Wong et al. |
| Maruta | Japanese | Other Asian | Wong et al. |
| Marutani | Japanese | Other Asian | Wong et al. |
| Maruyama | Japanese | Other Asian | Wong et al. |
| Marwah | Asian Indian | South Asian | Wong et al. |
| Marwaha | Asian Indian | South Asian | Wong et al. |
| Marzan |  | South Asian | Lim et al. |
| Marzo | Filipino | Other Asian | Wong et al. |
| Masa | Filipino | Other Asian | Wong et al. |
| Masada | Japanese | Other Asian | Wong et al. |
| Masagca | Filipino | Other Asian | Wong et al. |
| Masago | Japanese | Other Asian | Wong et al. |
| Masai | Japanese | Other Asian | Wong et al. |
| Masaki | Japanese | Other Asian | Wong et al. |
| Masana | Filipino | Other Asian | Wong et al. |
| Masangcay | Filipino | Other Asian | Wong et al. |
| Masangkay | Filipino | Other Asian | Wong et al. |
| Masani | Asian Indian | South Asian | Wong et al. |
| Masanque | Filipino | Other Asian | Wong et al. |
| Masao | Japanese | Other Asian | Wong et al. |
| Masaoay | Filipino | Other Asian | Wong et al. |
| Masaoka | Japanese | Other Asian | Wong et al. |
| Masaoy | Filipino | Other Asian | Wong et al. |
| Masaquel | Filipino | Other Asian | Wong et al. |
| Masaru | Japanese | Other Asian | Wong et al. |
| Masayoshi | Japanese | Other Asian | Wong et al. |
| Mascardo | Filipino | Other Asian | Wong et al. |
| Mascarenhas | Asian Indian | South Asian | Wong et al. |
| Mascarina | Filipino | Other Asian | Wong et al. |
| Mascarinas | Filipino | Other Asian | Wong et al. |
| Masculino | Filipino | Other Asian | Wong et al. |
| Mascunana | Filipino | Other Asian | Wong et al. |
| Masecampo | Filipino | Other Asian | Wong et al. |
| Mashimo | Japanese | Other Asian | Wong et al. |
| Mashita | Japanese | Other Asian | Wong et al. |
| Mashruwala | Asian Indian | South Asian | Wong et al. |
| Mashud | Bangladeshi | South Asian | Kranick et al. |
| Masias | Filipino | Other Asian | Wong et al. |
| Masibay | Filipino | Other Asian | Wong et al. |
| Masiclat | Filipino | Other Asian | Wong et al. |
| Masigla | Filipino | Other Asian | Wong et al. |
| Masiglat | Filipino | Other Asian | Wong et al. |
| Masih |  | South Asian | Lim et al. |
| Masilamani | Asian Indian | South Asian | Wong et al. |
| Masilang | Filipino | Other Asian | Wong et al. |
| Masinas | Filipino | Other Asian | Wong et al. |
| Maskarino | Filipino | Other Asian | Wong et al. |
| Masocol | Filipino | Other Asian | Wong et al. |
| Masongsong | Filipino | Other Asian | Wong et al. |
| Masood | Asian Indian | South Asian | Wong et al. |
| Mastana | Asian Indian | South Asian | Wong et al. |
| Mastay |  | South Asian | Lim et al. |
| Masubuchi | Japanese | Other Asian | Wong et al. |
| Masud |  | South Asian | Lim et al. |
| Masuda | Japanese | Other Asian | Wong et al. |
| Masuhara | Japanese | Other Asian | Wong et al. |
| Masui | Japanese | Other Asian | Wong et al. |
| Masukawa | Japanese | Other Asian | Wong et al. |
| Masuki |  | South Asian | Lim et al. |
| Masuko | Japanese | Other Asian | Wong et al. |
| Masulit | Filipino | Other Asian | Wong et al. |
| Masum |  | South Asian | Lim et al. |
| Masumoto | Japanese | Other Asian | Kranick et al. |
| Masumura | Japanese | Other Asian | Wong et al. |
| Masunaga | Japanese | Other Asian | Wong et al. |
| Masuno | Japanese | Other Asian | Wong et al. |
| Masuo | Japanese | Other Asian | Wong et al. |
| Masuoka | Japanese | Other Asian | Wong et al. |
| Masutani | Japanese | Other Asian | Wong et al. |
| Masuya | Japanese | Other Asian | Wong et al. |
| Masuyama | Japanese | Other Asian | Wong et al. |
| Masuzawa | Japanese | Other Asian | Wong et al. |
| Mata | Filipino | Other Asian | Wong et al. |
| Mataban | Filipino | Other Asian | Wong et al. |
| Matabang | Filipino | Other Asian | Wong et al. |
| Matadeen |  | South Asian | Lim et al. |
| Matalia | Asian Indian | South Asian | Wong et al. |
| Matanguihan | Filipino | Other Asian | Wong et al. |
| Matano | Japanese | Other Asian | Wong et al. |
| Matas | Filipino | Other Asian | Wong et al. |
| Matawaran | Filipino | Other Asian | Wong et al. |
| Matayoshi | Japanese | Other Asian | Wong et al. |
| Matela | Filipino | Other Asian | Wong et al. |
| Mateo | Filipino | Other Asian | Wong et al. |
| Mateu | Filipino | Other Asian | Wong et al. |
| Mathai | Asian Indian | South Asian | Wong et al. |
| Mathal | Asian Indian | South Asian | Wong et al. |
| Matharu | Asian Indian | South Asian | Wong et al. |
| Mathen | Asian Indian | South Asian | Wong et al. |
| Mathew | Asian Indian | South Asian | Kranick et al. |
| Mathrani | Asian Indian | South Asian | Wong et al. |
| Mathulla | Asian Indian | South Asian | Wong et al. |
| Mathunny | Asian Indian | South Asian | Wong et al. |
| Mathur | Asian Indian | South Asian | Wong et al. |
| Mathuraprasad |  | South Asian | Lim et al. |
| Mati | Filipino | Other Asian | Wong et al. |
| Matias | Filipino | Other Asian | Wong et al. |
| Matibag | Filipino | Other Asian | Wong et al. |
| Matic | Filipino | Other Asian | Wong et al. |
| Matienzo | Filipino | Other Asian | Wong et al. |
| Matiga | Filipino | Other Asian | Wong et al. |
| Matilla | Filipino | Other Asian | Wong et al. |
| Matillano | Filipino | Other Asian | Wong et al. |
| Matin |  | South Asian | Lim et al. |
| Mationg | Filipino | Other Asian | Wong et al. |
| Matira | Filipino | Other Asian | Wong et al. |
| Mato | Japanese | Other Asian | Wong et al. |
| Matoba | Japanese | Other Asian | Wong et al. |
| Matriano | Filipino | Other Asian | Wong et al. |
| Matro | Filipino | Other Asian | Wong et al. |
| Matsu | Japanese | Other Asian | Wong et al. |
| Matsuba | Japanese | Other Asian | Wong et al. |
| Matsubara | Japanese | Other Asian | Wong et al. |
| Matsubayashi | Japanese | Other Asian | Wong et al. |
| Matsuda | Japanese | Other Asian | Wong et al. |
| Matsudaira | Japanese | Other Asian | Wong et al. |
| Matsue | Japanese | Other Asian | Wong et al. |
| Matsueda | Japanese | Other Asian | Wong et al. |
| Matsuhashi | Japanese | Other Asian | Wong et al. |
| Matsui | Japanese | Other Asian | Wong et al. |
| Matsukado | Japanese | Other Asian | Wong et al. |
| Matsukawa | Japanese | Other Asian | Wong et al. |
| Matsuki | Japanese | Other Asian | Wong et al. |
| Matsukuma | Japanese | Other Asian | Wong et al. |
| Matsukura | Japanese | Other Asian | Wong et al. |
| Matsumaru | Japanese | Other Asian | Wong et al. |
| Matsumato | Japanese | Other Asian | Wong et al. |
| Matsumiya | Japanese | Other Asian | Wong et al. |
| Matsumori | Japanese | Other Asian | Wong et al. |
| Matsumoto | Japanese | Other Asian | Wong et al. |
| Matsumura | Japanese | Other Asian | Wong et al. |
| Matsunaga | Japanese | Other Asian | Wong et al. |
| Matsunami | Japanese | Other Asian | Wong et al. |
| Matsuno | Japanese | Other Asian | Wong et al. |
| Matsunobu | Japanese | Other Asian | Wong et al. |
| Matsuo | Japanese | Other Asian | Wong et al. |
| Matsuoka | Japanese | Other Asian | Wong et al. |
| Matsura | Japanese | Other Asian | Wong et al. |
| Matsusaka | Japanese | Other Asian | Wong et al. |
| Matsushige | Japanese | Other Asian | Wong et al. |
| Matsushima | Japanese | Other Asian | Wong et al. |
| Matsushita | Japanese | Other Asian | Wong et al. |
| Matsuta | Japanese | Other Asian | Wong et al. |
| Matsutani | Japanese | Other Asian | Wong et al. |
| Matsuura | Japanese | Other Asian | Wong et al. |
| Matsuya | Japanese | Other Asian | Wong et al. |
| Matsuyama | Japanese | Other Asian | Wong et al. |
| Matsuzaka | Japanese | Other Asian | Wong et al. |
| Matsuzaki | Japanese | Other Asian | Wong et al. |
| Matsuzawa | Japanese | Other Asian | Wong et al. |
| Matti | Filipino | Other Asian | Wong et al. |
| Mattu | Asian Indian | South Asian | Wong et al. |
| Matudio | Filipino | Other Asian | Wong et al. |
| Matuguina | Filipino | Other Asian | Wong et al. |
| Matulac | Filipino | Other Asian | Wong et al. |
| Matumoto | Japanese | Other Asian | Wong et al. |
| Matundan | Filipino | Other Asian | Wong et al. |
| Maturan | Filipino | Other Asian | Wong et al. |
| Maturgo | Filipino | Other Asian | Wong et al. |
| Matushima | Japanese | Other Asian | Wong et al. |
| Matute | Filipino | Other Asian | Wong et al. |
| Matutina | Filipino | Other Asian | Wong et al. |
| Matutino | Filipino | Other Asian | Wong et al. |
| Matuzaki | Japanese | Other Asian | Wong et al. |
| Mau | Chinese | Chinese | Wong et al. |
| Mauhay | Filipino | Other Asian | Wong et al. |
| Mauleon | Filipino | Other Asian | Wong et al. |
| Maulit | Filipino | Other Asian | Wong et al. |
| Maullon | Filipino | Other Asian | Wong et al. |
| Maunes | Filipino | Other Asian | Wong et al. |
| Mauricio | Filipino | Other Asian | Wong et al. |
| Mausisa | Filipino | Other Asian | Wong et al. |
| Mavani | Asian Indian | South Asian | Wong et al. |
| Mawatari | Japanese | Other Asian | Wong et al. |
| Maximo | Filipino | Other Asian | Wong et al. |
| Maxino | Filipino | Other Asian | Wong et al. |
| Maxion | Filipino | Other Asian | Wong et al. |
| Maya | Filipino | Other Asian | Wong et al. |
| Mayama | Japanese | Other Asian | Wong et al. |
| Mayeda | Japanese | Other Asian | Wong et al. |
| Mayehara | Japanese | Other Asian | Wong et al. |
| Mayemura | Japanese | Other Asian | Wong et al. |
| Maylad | Filipino | Other Asian | Wong et al. |
| Maynigo | Filipino | Other Asian | Wong et al. |
| Mayo | Filipino | Other Asian | Kranick et al. |
| Mayol | Filipino | Other Asian | Wong et al. |
| Mayor | Filipino | Other Asian | Wong et al. |
| Mayoralgo | Filipino | Other Asian | Wong et al. |
| Maypa | Filipino | Other Asian | Wong et al. |
| Mayrina | Filipino | Other Asian | Wong et al. |
| Mayuga | Filipino | Other Asian | Wong et al. |
| Mayugba | Filipino | Other Asian | Wong et al. |
| Mayumi | Japanese | Other Asian | Wong et al. |
| Mayuzumi | Japanese | Other Asian | Wong et al. |
| Maywahlall |  | South Asian | Lim et al. |
| Maywalall |  | South Asian | Lim et al. |
| Maza | Filipino | Other Asian | Wong et al. |
| Mazda | Asian Indian | South Asian | Wong et al. |
| Mazed |  | South Asian | Lim et al. |
| Mazhar | Asian Indian | South Asian | Wong et al. |
| Mazid | Bangladeshi | South Asian | Kranick et al. |
| Mazon | Filipino | Other Asian | Wong et al. |
| Mazumdar | Asian Indian | South Asian | Wong et al. |
| Mazumder | Asian Indian | South Asian | Wong et al. |
| Mbegum |  | South Asian | Lim et al. |
| Md |  | South Asian | Lim et al. |
| Meah | Asian Indian | South Asian | Wong et al. |
| Meana | Filipino | Other Asian | Wong et al. |
| Mecenas | Filipino | Other Asian | Wong et al. |
| Mecua | Filipino | Other Asian | Wong et al. |
| Meda | Asian Indian | South Asian | Wong et al. |
| Medalla | Filipino | Other Asian | Wong et al. |
| Medalle | Filipino | Other Asian | Wong et al. |
| Medel | Filipino | Other Asian | Wong et al. |
| Medellin | Filipino | Other Asian | Wong et al. |
| Medenilla | Filipino | Other Asian | Wong et al. |
| Mediana | Filipino | Other Asian | Wong et al. |
| Medina | Filipino | Other Asian | Kranick et al. |
| Medino | Filipino | Other Asian | Wong et al. |
| Mediodia | Filipino | Other Asian | Wong et al. |
| Medios | Filipino | Other Asian | Wong et al. |
| Mediran | Filipino | Other Asian | Wong et al. |
| Medrano | Filipino | Other Asian | Wong et al. |
| Medriano | Filipino | Other Asian | Wong et al. |
| Meerazackyn |  | South Asian | Lim et al. |
| Meerza | Asian Indian | South Asian | Wong et al. |
| Megallon | Filipino | Other Asian | Wong et al. |
| Meghani | Asian Indian | South Asian | Wong et al. |
| Megia | Filipino | Other Asian | Wong et al. |
| Megino | Filipino | Other Asian | Wong et al. |
| Megu | Filipino | Other Asian | Wong et al. |
| Megumi | Japanese | Other Asian | Wong et al. |
| Meguro | Japanese | Other Asian | Wong et al. |
| Mehdi | Asian Indian | South Asian | Wong et al. |
| Mehedi |  | South Asian | Lim et al. |
| Mehia | Asian Indian | South Asian | Wong et al. |
| Mehmood | Asian Indian | South Asian | Wong et al. |
| Mehra | Asian Indian | South Asian | Kranick et al. |
| Mehrok | Asian Indian | South Asian | Wong et al. |
| Mehrotra | Asian Indian | South Asian | Wong et al. |
| Mehta | Asian Indian | South Asian | Kranick et al. |
| Mehtani | Asian Indian | South Asian | Wong et al. |
| Mei | Chinese | Chinese | Kranick et al. |
| Meim | Filipino | Other Asian | Wong et al. |
| Meimban | Filipino | Other Asian | Wong et al. |
| Mejares | Filipino | Other Asian | Wong et al. |
| Mejia | Filipino | Other Asian | Wong et al. |
| Mejica | Filipino | Other Asian | Wong et al. |
| Mejorada | Filipino | Other Asian | Wong et al. |
| Mekhuri |  | South Asian | Lim et al. |
| Melad | Filipino | Other Asian | Wong et al. |
| Melano | Filipino | Other Asian | Wong et al. |
| Melchor | Filipino | Other Asian | Wong et al. |
| Melecio | Filipino | Other Asian | Wong et al. |
| Melegrito | Filipino | Other Asian | Wong et al. |
| Melencio | Filipino | Other Asian | Wong et al. |
| Melendez | Filipino | Other Asian | Wong et al. |
| Melendres | Filipino | Other Asian | Wong et al. |
| Melendrez | Filipino | Other Asian | Wong et al. |
| Melgar | Filipino | Other Asian | Wong et al. |
| Melgarejo | Filipino | Other Asian | Wong et al. |
| Melicor | Filipino | Other Asian | Wong et al. |
| Melitante | Filipino | Other Asian | Wong et al. |
| Meliton | Filipino | Other Asian | Wong et al. |
| Mella | Filipino | Other Asian | Wong et al. |
| Melliza | Filipino | Other Asian | Wong et al. |
| Melo | Filipino | Other Asian | Wong et al. |
| Melocoton | Filipino | Other Asian | Wong et al. |
| Membreno |  | South Asian | Lim et al. |
| Membrere | Filipino | Other Asian | Wong et al. |
| Memije | Filipino | Other Asian | Wong et al. |
| Memita | Filipino | Other Asian | Wong et al. |
| Memon | Pakistani | South Asian | Kranick et al. |
| Mempin | Filipino | Other Asian | Wong et al. |
| Memta | Asian Indian | South Asian | Wong et al. |
| Men | Chinese | Chinese | Wong et al. |
| Mena | Filipino | Other Asian | Wong et al. |
| Menchaca | Filipino | Other Asian | Wong et al. |
| Menchavez | Filipino | Other Asian | Wong et al. |
| Mencias | Filipino | Other Asian | Wong et al. |
| Menda | Asian Indian | South Asian | Wong et al. |
| Mendaros | Filipino | Other Asian | Wong et al. |
| Mendez | Filipino | Other Asian | Wong et al. |
| Mendezona | Filipino | Other Asian | Wong et al. |
| Mendieta | Filipino | Other Asian | Wong et al. |
| Mendigorin | Filipino | Other Asian | Wong et al. |
| Mendinueto | Filipino | Other Asian | Wong et al. |
| Mendiola | Filipino | Other Asian | Wong et al. |
| Mendioro | Filipino | Other Asian | Wong et al. |
| Mendiratta | Asian Indian | South Asian | Wong et al. |
| Mendones | Filipino | Other Asian | Wong et al. |
| Mendosa | Filipino | Other Asian | Wong et al. |
| Mendoza | Filipino | Other Asian | Wong et al. |
| Menes | Filipino | Other Asian | Wong et al. |
| Meneses | Filipino | Other Asian | Wong et al. |
| Menez | Filipino | Other Asian | Wong et al. |
| Menezes | Asian Indian | South Asian | Wong et al. |
| Meng | Chinese | Chinese | Wong et al. |
| Menguita | Filipino | Other Asian | Wong et al. |
| Menguito | Filipino | Other Asian | Wong et al. |
| Menon | Asian Indian | South Asian | Wong et al. |
| Menor | Filipino | Other Asian | Wong et al. |
| Menorca | Filipino | Other Asian | Wong et al. |
| Mensalvas | Filipino | Other Asian | Wong et al. |
| Merano | Filipino | Other Asian | Wong et al. |
| Mercader | Filipino | Other Asian | Wong et al. |
| Mercado | Filipino | Other Asian | Kranick et al. |
| Merced | Filipino | Other Asian | Wong et al. |
| Mercene | Filipino | Other Asian | Wong et al. |
| Merchan | Filipino | Other Asian | Wong et al. |
| Meregillano | Filipino | Other Asian | Wong et al. |
| Merete | Filipino | Other Asian | Wong et al. |
| Meria | Filipino | Other Asian | Wong et al. |
| Meriales | Filipino | Other Asian | Wong et al. |
| Merida | Filipino | Other Asian | Wong et al. |
| Merilles | Filipino | Other Asian | Wong et al. |
| Merilos | Filipino | Other Asian | Wong et al. |
| Merina |  | South Asian | Lim et al. |
| Merino | Filipino | Other Asian | Wong et al. |
| Meris | Filipino | Other Asian | Wong et al. |
| Merjudio | Filipino | Other Asian | Wong et al. |
| Merlan | Filipino | Other Asian | Wong et al. |
| Mero |  | South Asian | Lim et al. |
| Merto | Filipino | Other Asian | Wong et al. |
| Merza | Filipino | Other Asian | Wong et al. |
| Mesa | Filipino | Other Asian | Wong et al. |
| Mesia | Filipino | Other Asian | Wong et al. |
| Mesias | Filipino | Other Asian | Wong et al. |
| Mesina | Filipino | Other Asian | Wong et al. |
| Mesinas | Filipino | Other Asian | Wong et al. |
| Metiam | Filipino | Other Asian | Wong et al. |
| Metra | Filipino | Other Asian | Wong et al. |
| Metul |  | South Asian | Lim et al. |
| Mew | Chinese | Chinese | Wong et al. |
| Mg | Chinese | Chinese | Wong et al. |
| Mguyen | Vietnamese | Other Asian | Wong et al. |
| Mhamud |  | South Asian | Lim et al. |
| Mhatre | Asian Indian | South Asian | Wong et al. |
| Mi | Chinese | Chinese | Wong et al. |
| Mia |  | South Asian | Lim et al. |
| Miah | Bangladeshi; Asian Indian | South Asian | Kranick et al. |
| Mian | Pakistani | South Asian | Kranick et al. |
| Miana | Filipino | Other Asian | Wong et al. |
| Miao | Chinese | Chinese | Wong et al. |
| Miau | Chinese | Chinese | Wong et al. |
| Miazi |  | South Asian | Lim et al. |
| Micabalo | Filipino | Other Asian | Wong et al. |
| Miciano | Filipino | Other Asian | Wong et al. |
| Miclat | Filipino | Other Asian | Wong et al. |
| Mico | Filipino | Other Asian | Wong et al. |
| Micu | Filipino | Other Asian | Wong et al. |
| Micua | Filipino | Other Asian | Wong et al. |
| Miculob | Filipino | Other Asian | Wong et al. |
| Midorikawa | Japanese | Other Asian | Wong et al. |
| Mie | Chinese | Chinese | Wong et al. |
| Mien | Vietnamese | Other Asian | Wong et al. |
| Mier | Filipino | Other Asian | Wong et al. |
| Mieu | Vietnamese | Other Asian | Wong et al. |
| Mifune | Japanese | Other Asian | Wong et al. |
| Migaki | Japanese | Other Asian | Wong et al. |
| Migallos | Filipino | Other Asian | Wong et al. |
| Migita | Japanese | Other Asian | Wong et al. |
| Migrino | Filipino | Other Asian | Wong et al. |
| Miguel | Filipino | Other Asian | Kranick et al. |
| Miguelino | Filipino | Other Asian | Wong et al. |
| Mih | Chinese | Chinese | Wong et al. |
| Miha | Asian Indian | South Asian | Wong et al. |
| Mihara | Japanese | Other Asian | Wong et al. |
| Mihashi | Japanese | Other Asian | Wong et al. |
| Miho | Japanese | Other Asian | Wong et al. |
| Mijares | Filipino | Other Asian | Wong et al. |
| Mikami | Japanese | Other Asian | Wong et al. |
| Mikamo | Japanese | Other Asian | Wong et al. |
| Mikasa | Japanese | Other Asian | Wong et al. |
| Mikawa | Japanese | Other Asian | Wong et al. |
| Miki | Japanese | Other Asian | Wong et al. |
| Mikkilineni | Asian Indian | South Asian | Wong et al. |
| Mikoshiba | Japanese | Other Asian | Wong et al. |
| Mikumo | Japanese | Other Asian | Wong et al. |
| Mikuni | Japanese | Other Asian | Wong et al. |
| Mikuriya | Japanese | Other Asian | Wong et al. |
| Mil | Filipino | Other Asian | Wong et al. |
| Milambiling | Filipino | Other Asian | Wong et al. |
| Milana | Filipino | Other Asian | Wong et al. |
| Milanes | Filipino | Other Asian | Wong et al. |
| Milay | Filipino | Other Asian | Wong et al. |
| Militante | Filipino | Other Asian | Wong et al. |
| Militar | Filipino | Other Asian | Wong et al. |
| Milki |  | South Asian | Lim et al. |
| Milla | Filipino | Other Asian | Wong et al. |
| Millado | Filipino | Other Asian | Wong et al. |
| Millan | Filipino | Other Asian | Wong et al. |
| Millanes | Filipino | Other Asian | Wong et al. |
| Millano | Filipino | Other Asian | Wong et al. |
| Millare | Filipino | Other Asian | Wong et al. |
| Millares | Filipino | Other Asian | Wong et al. |
| Millarez | Filipino | Other Asian | Wong et al. |
| Millena | Filipino | Other Asian | Wong et al. |
| Millora | Filipino | Other Asian | Wong et al. |
| Mim | Korean | Other Asian | Wong et al. |
| Mimaki | Japanese | Other Asian | Wong et al. |
| Mimura | Japanese | Other Asian | Wong et al. |
| Min | Korean; Chinese; Burmese | Other Asian | Kranick et al. |
| Mina | Filipino | Other Asian | Wong et al. |
| Minagawa | Japanese | Other Asian | Wong et al. |
| Minakata | Japanese | Other Asian | Wong et al. |
| Minakawa | Japanese | Other Asian | Wong et al. |
| Minakuchi | Japanese | Other Asian | Wong et al. |
| Minami | Japanese | Other Asian | Wong et al. |
| Minamide | Japanese | Other Asian | Wong et al. |
| Minamoto | Japanese | Other Asian | Kranick et al. |
| Minas | Filipino | Other Asian | Wong et al. |
| Minato | Japanese | Other Asian | Wong et al. |
| Minatoya | Japanese | Other Asian | Wong et al. |
| Minawa | Japanese | Other Asian | Wong et al. |
| Minaya | Filipino | Other Asian | Wong et al. |
| Minayoshi | Japanese | Other Asian | Wong et al. |
| Mindanao | Filipino | Other Asian | Wong et al. |
| Mindo | Filipino | Other Asian | Wong et al. |
| Mine | Japanese | Other Asian | Wong et al. |
| Minegishi | Japanese | Other Asian | Wong et al. |
| Minei | Japanese | Other Asian | Wong et al. |
| Minel | Filipino | Other Asian | Wong et al. |
| Minemura | Japanese | Other Asian | Wong et al. |
| Mineshima | Japanese | Other Asian | Wong et al. |
| Ming | Chinese | Chinese | Wong et al. |
| Minglana | Filipino | Other Asian | Wong et al. |
| Mingming | Filipino | Other Asian | Wong et al. |
| Minh | Vietnamese | Other Asian | Wong et al. |
| Minhajuddin | Asian Indian | South Asian | Wong et al. |
| Mini |  | South Asian | Lim et al. |
| Miniano | Filipino | Other Asian | Wong et al. |
| Minimo | Filipino | Other Asian | Wong et al. |
| Minn | Burmese | Other Asian | Kranick et al. |
| Mino | Japanese | Other Asian | Wong et al. |
| Minobe | Japanese | Other Asian | Wong et al. |
| Minocha | Asian Indian | South Asian | Wong et al. |
| Minoda | Japanese | Other Asian | Wong et al. |
| Minoshima | Japanese | Other Asian | Wong et al. |
| Minoura | Japanese | Other Asian | Wong et al. |
| Minowa | Japanese | Other Asian | Wong et al. |
| Minoza | Filipino | Other Asian | Wong et al. |
| Mio | Japanese | Other Asian | Wong et al. |
| Mique | Filipino | Other Asian | Wong et al. |
| Mir |  | South Asian | Lim et al. |
| Mira | Filipino | Other Asian | Wong et al. |
| Mirabel | Filipino | Other Asian | Wong et al. |
| Mirabueno | Filipino | Other Asian | Wong et al. |
| Mirador | Filipino | Other Asian | Wong et al. |
| Miraflor | Filipino | Other Asian | Wong et al. |
| Miraflores | Filipino | Other Asian | Wong et al. |
| Mirafuentes | Filipino | Other Asian | Wong et al. |
| Miral | Filipino | Other Asian | Wong et al. |
| Miralles | Filipino | Other Asian | Wong et al. |
| Miran | Filipino | Other Asian | Wong et al. |
| Miranda | Filipino | Other Asian | Wong et al. |
| Mirani | Asian Indian | South Asian | Wong et al. |
| Mirano | Filipino | Other Asian | Wong et al. |
| Miras | Filipino | Other Asian | Wong et al. |
| Mirasol | Filipino | Other Asian | Wong et al. |
| Miravite | Filipino | Other Asian | Wong et al. |
| Mirdha |  | South Asian | Lim et al. |
| Miro | Filipino | Other Asian | Wong et al. |
| Miroy | Filipino | Other Asian | Wong et al. |
| Mirza | Asian Indian | South Asian | Wong et al. |
| Misa | Filipino | Other Asian | Wong et al. |
| Misajon | Filipino | Other Asian | Wong et al. |
| Misaka | Japanese | Other Asian | Wong et al. |
| Misaki | Japanese | Other Asian | Wong et al. |
| Misawa | Japanese | Other Asian | Wong et al. |
| Misenas | Filipino | Other Asian | Wong et al. |
| Mishima | Japanese | Other Asian | Wong et al. |
| Mishina | Japanese | Other Asian | Wong et al. |
| Mishra | Asian Indian | South Asian | Wong et al. |
| Mislang | Filipino | Other Asian | Wong et al. |
| Misola | Filipino | Other Asian | Wong et al. |
| Misono | Japanese | Other Asian | Wong et al. |
| Misquitta | Asian Indian | South Asian | Wong et al. |
| Misra | Asian Indian | South Asian | Wong et al. |
| Mission | Filipino | Other Asian | Wong et al. |
| Mistica | Filipino | Other Asian | Wong et al. |
| Mistri | Asian Indian | South Asian | Wong et al. |
| Mistry | Asian Indian | South Asian | Wong et al. |
| Misu | Japanese | Other Asian | Wong et al. |
| Misumi | Japanese | Other Asian | Wong et al. |
| Mita | Japanese | Other Asian | Wong et al. |
| Mital | Asian Indian | South Asian | Wong et al. |
| Mitamura | Japanese | Other Asian | Wong et al. |
| Mitani | Japanese | Other Asian | Wong et al. |
| Mitarai | Japanese | Other Asian | Wong et al. |
| Mitha | Asian Indian | South Asian | Wong et al. |
| Mithal | Asian Indian | South Asian | Wong et al. |
| Mithani | Asian Indian | South Asian | Wong et al. |
| Mito |  | South Asian | Lim et al. |
| Mitobe | Japanese | Other Asian | Wong et al. |
| Mitoma | Japanese | Other Asian | Wong et al. |
| Mitome | Japanese | Other Asian | Wong et al. |
| Mitomi | Japanese | Other Asian | Wong et al. |
| Mitra | Asian Indian; Bangladeshi | South Asian | Kranick et al. |
| Mitre | Filipino | Other Asian | Wong et al. |
| Mitsuda | Japanese | Other Asian | Wong et al. |
| Mitsufuji | Japanese | Other Asian | Wong et al. |
| Mitsugi | Japanese | Other Asian | Wong et al. |
| Mitsuhashi | Japanese | Other Asian | Wong et al. |
| Mitsui | Japanese | Other Asian | Wong et al. |
| Mitsuishi | Japanese | Other Asian | Wong et al. |
| Mitsumori | Japanese | Other Asian | Wong et al. |
| Mitsumoto | Japanese | Other Asian | Wong et al. |
| Mitsunaga | Japanese | Other Asian | Wong et al. |
| Mitsuoka | Japanese | Other Asian | Wong et al. |
| Mitsuta | Japanese | Other Asian | Wong et al. |
| Mitsuya | Japanese | Other Asian | Wong et al. |
| Mitsuyasu | Japanese | Other Asian | Wong et al. |
| Mittal | Asian Indian | South Asian | Wong et al. |
| Mittra | Asian Indian | South Asian | Wong et al. |
| Mitu |  | South Asian | Lim et al. |
| Miu | Vietnamese; Japanese | Other Asian | Kranick et al. |
| Miura | Japanese | Other Asian | Wong et al. |
| Miwa | Japanese | Other Asian | Wong et al. |
| Miya | Japanese | Other Asian | Wong et al. |
| Miyabara | Japanese | Other Asian | Wong et al. |
| Miyabayashi | Japanese | Other Asian | Wong et al. |
| Miyabe | Japanese | Other Asian | Wong et al. |
| Miyachi | Japanese | Other Asian | Wong et al. |
| Miyadera | Japanese | Other Asian | Wong et al. |
| Miyagawa | Japanese | Other Asian | Wong et al. |
| Miyagi | Japanese | Other Asian | Wong et al. |
| Miyagishima | Japanese | Other Asian | Wong et al. |
| Miyaguchi | Japanese | Other Asian | Kranick et al. |
| Miyahara | Japanese | Other Asian | Wong et al. |
| Miyahira | Japanese | Other Asian | Wong et al. |
| Miyai | Japanese | Other Asian | Wong et al. |
| Miyaji | Japanese | Other Asian | Wong et al. |
| Miyajima | Japanese | Other Asian | Wong et al. |
| Miyakawa | Japanese | Other Asian | Wong et al. |
| Miyake | Japanese | Other Asian | Wong et al. |
| Miyaki | Japanese | Other Asian | Wong et al. |
| Miyakoshi | Japanese | Other Asian | Wong et al. |
| Miyama | Japanese | Other Asian | Wong et al. |
| Miyamori | Japanese | Other Asian | Wong et al. |
| Miyamoto | Japanese | Other Asian | Wong et al. |
| Miyamura | Japanese | Other Asian | Wong et al. |
| Miyanaga | Japanese | Other Asian | Wong et al. |
| Miyanishi | Japanese | Other Asian | Wong et al. |
| Miyano | Japanese | Other Asian | Wong et al. |
| Miyao | Japanese | Other Asian | Wong et al. |
| Miyaoka | Japanese | Other Asian | Wong et al. |
| Miyasaka | Japanese | Other Asian | Wong et al. |
| Miyasaki | Japanese | Other Asian | Wong et al. |
| Miyasato | Japanese | Other Asian | Wong et al. |
| Miyasawa | Japanese | Other Asian | Wong et al. |
| Miyashiro | Japanese | Other Asian | Wong et al. |
| Miyashita | Japanese | Other Asian | Wong et al. |
| Miyata | Japanese | Other Asian | Wong et al. |
| Miyatake | Japanese | Other Asian | Wong et al. |
| Miyatani | Japanese | Other Asian | Wong et al. |
| Miyauchi | Japanese | Other Asian | Wong et al. |
| Miyawaki | Japanese | Other Asian | Wong et al. |
| Miyayama | Japanese | Other Asian | Wong et al. |
| Miyazaki | Japanese | Other Asian | Wong et al. |
| Miyazato | Japanese | Other Asian | Wong et al. |
| Miyazawa | Japanese | Other Asian | Wong et al. |
| Miyazono | Japanese | Other Asian | Wong et al. |
| Miyokawa | Japanese | Other Asian | Wong et al. |
| Miyoshi | Japanese | Other Asian | Wong et al. |
| Mizan |  | South Asian | Lim et al. |
| Mizi |  | South Asian | Lim et al. |
| Mizobe | Japanese | Other Asian | Wong et al. |
| Mizobuchi | Japanese | Other Asian | Wong et al. |
| Mizoguchi | Japanese | Other Asian | Wong et al. |
| Mizokami | Japanese | Other Asian | Wong et al. |
| Mizota | Japanese | Other Asian | Wong et al. |
| Mizrahi | Asian Indian | South Asian | Wong et al. |
| Mizugaki | Japanese | Other Asian | Wong et al. |
| Mizuguchi | Japanese | Other Asian | Wong et al. |
| Mizuhara | Japanese | Other Asian | Wong et al. |
| Mizukami | Japanese | Other Asian | Wong et al. |
| Mizuki | Japanese | Other Asian | Wong et al. |
| Mizukoshi | Japanese | Other Asian | Wong et al. |
| Mizumachi | Japanese | Other Asian | Wong et al. |
| Mizumoto | Japanese | Other Asian | Wong et al. |
| Mizumura | Japanese | Other Asian | Wong et al. |
| Mizuno | Japanese | Other Asian | Wong et al. |
| Mizunuma | Japanese | Other Asian | Wong et al. |
| Mizusaki | Japanese | Other Asian | Wong et al. |
| Mizusawa | Japanese | Other Asian | Wong et al. |
| Mizushima | Japanese | Other Asian | Wong et al. |
| Mizushina | Japanese | Other Asian | Wong et al. |
| Mizuta | Japanese | Other Asian | Wong et al. |
| Mizutani | Japanese | Other Asian | Kranick et al. |
| Mo | Chinese | Chinese | Kranick et al. |
| Moa | Chinese | Chinese | Wong et al. |
| Mobo | Filipino | Other Asian | Wong et al. |
| Moc | Vietnamese | Other Asian | Wong et al. |
| Mochida | Japanese | Other Asian | Wong et al. |
| Mochimaru | Japanese | Other Asian | Wong et al. |
| Mochinaga | Japanese | Other Asian | Wong et al. |
| Mochizuki | Japanese | Other Asian | Wong et al. |
| Mocorro | Filipino | Other Asian | Wong et al. |
| Modelo | Filipino | Other Asian | Wong et al. |
| Modequillo | Filipino | Other Asian | Wong et al. |
| Modesto | Filipino | Other Asian | Wong et al. |
| Modh | Asian Indian | South Asian | Wong et al. |
| Modha | Asian Indian | South Asian | Wong et al. |
| Modi | Asian Indian | South Asian | Wong et al. |
| Modina | Filipino | Other Asian | Wong et al. |
| Mody | Asian Indian | South Asian | Wong et al. |
| Moey | Chinese | Chinese | Wong et al. |
| Mogami | Japanese | Other Asian | Wong et al. |
| Moghe | Asian Indian | South Asian | Wong et al. |
| Mogi | Japanese | Other Asian | Wong et al. |
| Moh |  | South Asian | Lim et al. |
| Mohabeer |  | South Asian | Lim et al. |
| Mohabir |  | South Asian | Lim et al. |
| Mohabir,2 |  | South Asian | Lim et al. |
| Mohajir | Asian Indian | South Asian | Wong et al. |
| Mohal |  | South Asian | Lim et al. |
| Mohamad | Asian Indian | South Asian | Wong et al. |
| Mohamed | Pakistani; Malaysian | South Asian | Kranick et al. |
| Mohamedali | Asian Indian | South Asian | Wong et al. |
| Mohammad | Asian Indian; Bangladeshi; Pakistani | South Asian | Kranick et al. |
| Mohammed |  | South Asian | Lim et al. |
| Mohammedkhan |  | South Asian | Lim et al. |
| Mohan | Asian Indian | South Asian | Kranick et al. |
| Mohandas | Asian Indian | South Asian | Wong et al. |
| Mohanlal | Asian Indian | South Asian | Wong et al. |
| Mohanlall |  | South Asian | Lim et al. |
| Mohanrao | Asian Indian | South Asian | Wong et al. |
| Mohanty | Asian Indian | South Asian | Wong et al. |
| Mohapatra | Asian Indian | South Asian | Kranick et al. |
| Mohd |  | South Asian | Lim et al. |
| Moheet |  | South Asian | Lim et al. |
| Mohideen | Asian Indian | South Asian | Wong et al. |
| Mohile | Asian Indian | South Asian | Wong et al. |
| Mohindra | Asian Indian | South Asian | Wong et al. |
| Mohiuddin | Asian Indian | South Asian | Wong et al. |
| Mohomed | Asian Indian | South Asian | Wong et al. |
| Mohri | Japanese | Other Asian | Wong et al. |
| Mohsin | Asian Indian | South Asian | Wong et al. |
| Moi | Chinese | Chinese | Wong et al. |
| Moin |  | South Asian | Lim et al. |
| Moinuddin | Asian Indian | South Asian | Wong et al. |
| Moises | Filipino | Other Asian | Wong et al. |
| Moitra | Asian Indian | South Asian | Wong et al. |
| Moizuddin | Asian Indian | South Asian | Wong et al. |
| Mojado | Filipino | Other Asian | Wong et al. |
| Mojar | Filipino | Other Asian | Wong et al. |
| Mojares | Filipino | Other Asian | Wong et al. |
| Mojica | Filipino | Other Asian | Wong et al. |
| Mojumder |  | South Asian | Lim et al. |
| Mok | Chinese; Korean | Chinese | Kranick et al. |
| Moktader |  | South Asian | Lim et al. |
| Moktadir |  | South Asian | Lim et al. |
| Molano | Filipino | Other Asian | Wong et al. |
| Molapour |  | South Asian | Lim et al. |
| Molas | Filipino | Other Asian | Wong et al. |
| Molato | Filipino | Other Asian | Wong et al. |
| Molera | Filipino | Other Asian | Wong et al. |
| Molina | Filipino | Other Asian | Wong et al. |
| Molinos | Filipino | Other Asian | Wong et al. |
| Molla | Asian Indian; Bangladeshi | South Asian | Kranick et al. |
| Mollah |  | South Asian | Lim et al. |
| Molleno | Filipino | Other Asian | Wong et al. |
| Mollik |  | South Asian | Lim et al. |
| Molo | Filipino | Other Asian | Wong et al. |
| Molon | Filipino | Other Asian | Wong et al. |
| Momen |  | South Asian | Lim et al. |
| Momena |  | South Asian | Lim et al. |
| Momi | Asian Indian | South Asian | Wong et al. |
| Momii | Japanese | Other Asian | Wong et al. |
| Momin | Asian Indian; Bangladeshi | South Asian | Kranick et al. |
| Momiyama | Japanese | Other Asian | Wong et al. |
| Momoi | Japanese | Other Asian | Wong et al. |
| Momongan | Filipino | Other Asian | Wong et al. |
| Momosaki | Japanese | Other Asian | Wong et al. |
| Momose | Japanese | Other Asian | Wong et al. |
| Momota | Japanese | Other Asian | Wong et al. |
| Mon | Chinese | Chinese | Wong et al. |
| Monar | Filipino | Other Asian | Wong et al. |
| Monares | Filipino | Other Asian | Wong et al. |
| Monasterial | Filipino | Other Asian | Wong et al. |
| Monasterio | Filipino | Other Asian | Wong et al. |
| Monato | Filipino | Other Asian | Wong et al. |
| Monayem |  | South Asian | Lim et al. |
| Moncada | Filipino | Other Asian | Wong et al. |
| Moncayo | Filipino | Other Asian | Wong et al. |
| Monces | Filipino | Other Asian | Wong et al. |
| Mondal |  | South Asian | Lim et al. |
| Mondala | Filipino | Other Asian | Wong et al. |
| Mondares | Filipino | Other Asian | Wong et al. |
| Mondejar | Filipino | Other Asian | Wong et al. |
| Monden | Japanese | Other Asian | Wong et al. |
| Mondero | Filipino | Other Asian | Wong et al. |
| Mondido | Filipino | Other Asian | Wong et al. |
| Mondina | Filipino | Other Asian | Wong et al. |
| Mondol |  | South Asian | Lim et al. |
| Mondonedo | Filipino | Other Asian | Wong et al. |
| Mondragon | Filipino | Other Asian | Wong et al. |
| Moneda | Filipino | Other Asian | Wong et al. |
| Mones | Filipino | Other Asian | Wong et al. |
| Monfero | Filipino | Other Asian | Wong et al. |
| Mong | Chinese | Chinese | Wong et al. |
| Monga | Asian Indian | South Asian | Wong et al. |
| Monge |  | South Asian | Lim et al. |
| Moni |  | South Asian | Lim et al. |
| Monillas | Filipino | Other Asian | Wong et al. |
| Monir |  | South Asian | Lim et al. |
| Monira |  | South Asian | Lim et al. |
| Moniruzzaman |  | South Asian | Lim et al. |
| Monis | Filipino | Other Asian | Wong et al. |
| Monje | Filipino | Other Asian | Wong et al. |
| Monji | Japanese | Other Asian | Wong et al. |
| Monma | Japanese | Other Asian | Wong et al. |
| Monnan |  | South Asian | Lim et al. |
| Monreal | Filipino | Other Asian | Wong et al. |
| Monroy | Filipino | Other Asian | Wong et al. |
| Monsada | Filipino | Other Asian | Wong et al. |
| Monsale | Filipino | Other Asian | Wong et al. |
| Monsalud | Filipino | Other Asian | Wong et al. |
| Monsanto | Filipino | Other Asian | Wong et al. |
| Monsayac | Filipino | Other Asian | Wong et al. |
| Monserate | Filipino | Other Asian | Wong et al. |
| Monserrat | Filipino | Other Asian | Wong et al. |
| Monsod | Filipino | Other Asian | Wong et al. |
| Monta | Filipino | Other Asian | Wong et al. |
| Montajes | Filipino | Other Asian | Wong et al. |
| Montalban | Filipino | Other Asian | Wong et al. |
| Montalbo | Filipino | Other Asian | Wong et al. |
| Montales | Filipino | Other Asian | Wong et al. |
| Montalla | Filipino | Other Asian | Wong et al. |
| Montallana | Filipino | Other Asian | Wong et al. |
| Montalvo | Filipino | Other Asian | Wong et al. |
| Montanano | Filipino | Other Asian | Wong et al. |
| Montaner | Filipino | Other Asian | Wong et al. |
| Montanez | Filipino | Other Asian | Wong et al. |
| Montano | Filipino | Other Asian | Wong et al. |
| Montante | Filipino | Other Asian | Wong et al. |
| Montaos | Filipino | Other Asian | Wong et al. |
| Monteagudo | Filipino | Other Asian | Wong et al. |
| Montealegre | Filipino | Other Asian | Wong et al. |
| Montebon | Filipino | Other Asian | Wong et al. |
| Montecer | Filipino | Other Asian | Wong et al. |
| Montecillo | Filipino | Other Asian | Kranick et al. |
| Monteclaro | Filipino | Other Asian | Wong et al. |
| Montefalcon | Filipino | Other Asian | Wong et al. |
| Montehermoso | Filipino | Other Asian | Wong et al. |
| Montejo | Filipino | Other Asian | Wong et al. |
| Montelibano | Filipino | Other Asian | Wong et al. |
| Montellano | Filipino | Other Asian | Wong et al. |
| Montemayor | Filipino | Other Asian | Wong et al. |
| Montenegro | Filipino | Other Asian | Wong et al. |
| Monterey | Filipino | Other Asian | Wong et al. |
| Montero | Filipino | Other Asian | Wong et al. |
| Monterola | Filipino | Other Asian | Wong et al. |
| Montes | Filipino | Other Asian | Wong et al. |
| Montesa | Filipino | Other Asian | Wong et al. |
| Montesclaros | Filipino | Other Asian | Wong et al. |
| Monteverde | Filipino | Other Asian | Wong et al. |
| Montevirgen | Filipino | Other Asian | Wong et al. |
| Montez | Filipino | Other Asian | Wong et al. |
| Monteza | Filipino | Other Asian | Wong et al. |
| Montiague | Filipino | Other Asian | Wong et al. |
| Montiero | Filipino | Other Asian | Wong et al. |
| Montilla | Filipino | Other Asian | Wong et al. |
| Montillano | Filipino | Other Asian | Wong et al. |
| Montinola | Filipino | Other Asian | Wong et al. |
| Montojo | Filipino | Other Asian | Wong et al. |
| Monton | Filipino | Other Asian | Wong et al. |
| Montoya | Filipino | Other Asian | Wong et al. |
| Montu | Bangladeshi | South Asian | Kranick et al. |
| Montuya | Filipino | Other Asian | Wong et al. |
| Monzon | Filipino | Other Asian | Wong et al. |
| Monzones | Filipino | Other Asian | Wong et al. |
| Moo | Chinese | Chinese | Wong et al. |
| Mooc | Vietnamese | Other Asian | Wong et al. |
| Mookerjee | Asian Indian | South Asian | Wong et al. |
| Mookherjee | Asian Indian | South Asian | Wong et al. |
| Moolchand | Filipino | Other Asian | Wong et al. |
| Moom | Chinese | Chinese | Wong et al. |
| Moon | Korean | Other Asian | Kranick et al. |
| Moonasar |  | South Asian | Lim et al. |
| Moonessar |  | South Asian | Lim et al. |
| Moonilal |  | South Asian | Lim et al. |
| Moorthy | Asian Indian | South Asian | Wong et al. |
| Moosa | Asian Indian | South Asian | Wong et al. |
| Moosvi | Asian Indian | South Asian | Wong et al. |
| Mootoo |  | South Asian | Lim et al. |
| Mopera | Filipino | Other Asian | Wong et al. |
| Mora | Filipino | Other Asian | Wong et al. |
| Morabe | Filipino | Other Asian | Wong et al. |
| Morad |  | South Asian | Lim et al. |
| Morada | Filipino | Other Asian | Wong et al. |
| Morado | Filipino | Other Asian | Wong et al. |
| Morados | Filipino | Other Asian | Wong et al. |
| Moraes | Asian Indian | South Asian | Wong et al. |
| Moraga | Filipino | Other Asian | Wong et al. |
| Moral | Filipino | Other Asian | Wong et al. |
| Moralde | Filipino | Other Asian | Wong et al. |
| Moraleda | Filipino | Other Asian | Wong et al. |
| Morales | Filipino | Other Asian | Wong et al. |
| Morani | Asian Indian | South Asian | Wong et al. |
| Morano | Filipino | Other Asian | Kranick et al. |
| Morante | Filipino | Other Asian | Wong et al. |
| Morata | Filipino | Other Asian | Wong et al. |
| Morato | Filipino | Other Asian | Wong et al. |
| Morco | Filipino | Other Asian | Wong et al. |
| Mordeno | Filipino | Other Asian | Wong et al. |
| Morelos | Filipino | Other Asian | Wong et al. |
| Morena | Filipino | Other Asian | Wong et al. |
| Moreno | Filipino | Other Asian | Wong et al. |
| Morente | Filipino | Other Asian | Wong et al. |
| Moreto | Filipino | Other Asian | Wong et al. |
| Morga | Filipino | Other Asian | Wong et al. |
| Morgado | Filipino | Other Asian | Wong et al. |
| Morgia | Filipino | Other Asian | Wong et al. |
| Mori | Japanese | Other Asian | Wong et al. |
| Moriam |  | South Asian | Lim et al. |
| Moribe | Japanese | Other Asian | Wong et al. |
| Morigaki | Japanese | Other Asian | Wong et al. |
| Moriguchi | Japanese | Other Asian | Wong et al. |
| Morihara | Japanese | Other Asian | Wong et al. |
| Morihiro | Japanese | Other Asian | Wong et al. |
| Morii | Japanese | Other Asian | Wong et al. |
| Morikawa | Japanese | Other Asian | Kranick et al. |
| Morilla | Filipino | Other Asian | Wong et al. |
| Morillo | Filipino | Other Asian | Wong et al. |
| Morimoto | Japanese | Other Asian | Wong et al. |
| Morimura | Japanese | Other Asian | Wong et al. |
| Morinaga | Japanese | Other Asian | Wong et al. |
| Morinaka | Japanese | Other Asian | Wong et al. |
| Morino | Japanese | Other Asian | Wong et al. |
| Morioka | Japanese | Other Asian | Wong et al. |
| Morisaki | Japanese | Other Asian | Wong et al. |
| Morisako | Japanese | Other Asian | Wong et al. |
| Morisawa | Japanese | Other Asian | Wong et al. |
| Morishige | Japanese | Other Asian | Wong et al. |
| Morishima | Japanese | Other Asian | Wong et al. |
| Morishita | Japanese | Other Asian | Wong et al. |
| Morita | Japanese | Other Asian | Wong et al. |
| Moritani | Japanese | Other Asian | Wong et al. |
| Morito | Japanese | Other Asian | Wong et al. |
| Moriuchi | Japanese | Other Asian | Wong et al. |
| Moriwaki | Japanese | Other Asian | Wong et al. |
| Moriya | Japanese | Other Asian | Wong et al. |
| Moriyama | Japanese | Other Asian | Wong et al. |
| Moriyasu | Japanese | Other Asian | Wong et al. |
| Morizono | Japanese | Other Asian | Wong et al. |
| Morla | Filipino | Other Asian | Wong et al. |
| Morohashi | Japanese | Other Asian | Wong et al. |
| Morohoshi | Japanese | Other Asian | Wong et al. |
| Morooka | Japanese | Other Asian | Wong et al. |
| Morota | Filipino | Other Asian | Wong et al. |
| Moroto | Japanese | Other Asian | Wong et al. |
| Morozumi | Japanese | Other Asian | Wong et al. |
| Morshed |  | South Asian | Lim et al. |
| Morshid |  | South Asian | Lim et al. |
| Morta | Filipino | Other Asian | Wong et al. |
| Mortaza |  | South Asian | Lim et al. |
| Mortel | Filipino | Other Asian | Wong et al. |
| Mortela | Filipino | Other Asian | Wong et al. |
| Mortera | Filipino | Other Asian | Wong et al. |
| Moscoso | Filipino | Other Asian | Wong et al. |
| Mose | Filipino | Other Asian | Wong et al. |
| Moselina | Filipino | Other Asian | Wong et al. |
| Moskito | Filipino | Other Asian | Wong et al. |
| Mosqueda | Filipino | Other Asian | Wong et al. |
| Mosquera | Filipino | Other Asian | Wong et al. |
| Mostafa | Bangladeshi | South Asian | Kranick et al. |
| Mostajo | Filipino | Other Asian | Wong et al. |
| Mostoles | Filipino | Other Asian | Wong et al. |
| Mostrales | Filipino | Other Asian | Wong et al. |
| Mosuela | Filipino | Other Asian | Wong et al. |
| Mot | Vietnamese | Other Asian | Wong et al. |
| Mota | Filipino | Other Asian | Wong et al. |
| Motalab |  | South Asian | Lim et al. |
| Motaleb |  | South Asian | Lim et al. |
| Motas | Filipino | Other Asian | Wong et al. |
| Motegi | Japanese | Other Asian | Wong et al. |
| Moten |  | South Asian | Lim et al. |
| Moti | Asian Indian | South Asian | Wong et al. |
| Motibhai | Asian Indian | South Asian | Wong et al. |
| Motilal | Asian Indian | South Asian | Wong et al. |
| Motilla | Filipino | Other Asian | Wong et al. |
| Motin |  | South Asian | Lim et al. |
| Motiram | Asian Indian | South Asian | Wong et al. |
| Motivala | Asian Indian | South Asian | Wong et al. |
| Motiwala | Asian Indian | South Asian | Wong et al. |
| Motiwalla | Asian Indian | South Asian | Wong et al. |
| Moto | Japanese | Other Asian | Wong et al. |
| Motobayashi | Japanese | Other Asian | Wong et al. |
| Motohashi | Japanese | Other Asian | Wong et al. |
| Motoi | Japanese | Other Asian | Wong et al. |
| Motojima | Japanese | Other Asian | Wong et al. |
| Motokawa | Japanese | Other Asian | Wong et al. |
| Motoki | Japanese | Other Asian | Wong et al. |
| Motomal | Filipino | Other Asian | Wong et al. |
| Motomura | Japanese | Other Asian | Wong et al. |
| Motooka | Japanese | Other Asian | Wong et al. |
| Motos | Filipino | Other Asian | Wong et al. |
| Motoyama | Japanese | Other Asian | Wong et al. |
| Motoyoshi | Japanese | Other Asian | Wong et al. |
| Motus | Filipino | Other Asian | Wong et al. |
| Motwane | Asian Indian | South Asian | Wong et al. |
| Mou | Chinese | Chinese | Wong et al. |
| Moudgil | Asian Indian | South Asian | Wong et al. |
| Moulic | Filipino | Other Asian | Wong et al. |
| Mouri | Japanese | Other Asian | Wong et al. |
| Moushomi |  | South Asian | Lim et al. |
| Movida | Filipino | Other Asian | Wong et al. |
| Movido | Filipino | Other Asian | Wong et al. |
| Movilla | Filipino | Other Asian | Wong et al. |
| Mow | Chinese | Chinese | Wong et al. |
| Mowla |  | South Asian | Lim et al. |
| Moy | Chinese | Chinese | Kranick et al. |
| Moya | Filipino | Other Asian | Wong et al. |
| Moyeen |  | South Asian | Lim et al. |
| Mozaffar | Asian Indian | South Asian | Wong et al. |
| Mozeb |  | South Asian | Lim et al. |
| Mozib |  | South Asian | Lim et al. |
| Mozo | Filipino | Other Asian | Wong et al. |
| Mozumder |  | South Asian | Lim et al. |
| Mrida |  | South Asian | Lim et al. |
| Mridha |  | South Asian | Lim et al. |
| Mruthyunjaya | Asian Indian | South Asian | Wong et al. |
| Mu | Chinese | Chinese | Wong et al. |
| Muana | Filipino | Other Asian | Wong et al. |
| Muc | Vietnamese | Other Asian | Wong et al. |
| Mudaliar | Asian Indian | South Asian | Wong et al. |
| Mudassar |  | South Asian | Lim et al. |
| Mudzinski | Filipino | Other Asian | Wong et al. |
| Mue | Chinese | Chinese | Wong et al. |
| Mueca | Filipino | Other Asian | Wong et al. |
| Muego | Filipino | Other Asian | Wong et al. |
| Mufti |  | South Asian | Lim et al. |
| Mughal |  | South Asian | Lim et al. |
| Mugol | Filipino | Other Asian | Wong et al. |
| Muh | Chinese | Chinese | Wong et al. |
| Muhammad |  | South Asian | Lim et al. |
| Muhiuddin |  | South Asian | Lim et al. |
| Mui | Chinese; Vietnamese | Chinese | Kranick et al. |
| Mujaffar |  | South Asian | Lim et al. |
| Mujeeb | Asian Indian | South Asian | Wong et al. |
| Mujtaba | Asian Indian | South Asian | Wong et al. |
| Mujumdar | Asian Indian | South Asian | Wong et al. |
| Muk | Chinese | Chinese | Wong et al. |
| Mukae | Japanese | Other Asian | Wong et al. |
| Mukai | Japanese | Other Asian | Wong et al. |
| Mukaida | Japanese | Other Asian | Wong et al. |
| Mukalel | Asian Indian | South Asian | Wong et al. |
| Mukasa | Japanese | Other Asian | Wong et al. |
| Mukawa | Japanese | Other Asian | Wong et al. |
| Mukerjee | Asian Indian | South Asian | Wong et al. |
| Mukerji | Asian Indian | South Asian | Wong et al. |
| Mukherjee | Asian Indian | South Asian | Kranick et al. |
| Mukherji | Asian Indian | South Asian | Wong et al. |
| Mukhopadhyay |  | South Asian | Lim et al. |
| Mukhtar | Pakistani | South Asian | Kranick et al. |
| Mukim |  | South Asian | Lim et al. |
| Mukkamala | Asian Indian | South Asian | Wong et al. |
| Mukoyama | Japanese | Other Asian | Wong et al. |
| Mulani | Asian Indian | South Asian | Wong et al. |
| Mulato | Filipino | Other Asian | Wong et al. |
| Mulay | Asian Indian | South Asian | Wong et al. |
| Muldong | Filipino | Other Asian | Wong et al. |
| Mulic |  | South Asian | Lim et al. |
| Mulingtapang | Filipino | Other Asian | Wong et al. |
| Mulji | Asian Indian | South Asian | Wong et al. |
| Muljibhai | Asian Indian | South Asian | Wong et al. |
| Mulla | Asian Indian | South Asian | Wong et al. |
| Mullick | Asian Indian | South Asian | Wong et al. |
| Multani | Asian Indian | South Asian | Wong et al. |
| Mulye | Asian Indian | South Asian | Wong et al. |
| Mumin |  | South Asian | Lim et al. |
| Mumith |  | South Asian | Lim et al. |
| Mumtaz | Pakistani | South Asian | Kranick et al. |
| Mun | Korean | Other Asian | Wong et al. |
| Munakata | Japanese | Other Asian | Wong et al. |
| Munar | Filipino | Other Asian | Wong et al. |
| Munawar | Asian Indian | South Asian | Wong et al. |
| Muncal | Filipino | Other Asian | Wong et al. |
| Mundala | Filipino | Other Asian | Wong et al. |
| Mundkur | Asian Indian | South Asian | Wong et al. |
| Mundo | Filipino | Other Asian | Wong et al. |
| Munekata | Japanese | Other Asian | Wong et al. |
| Muneoka | Japanese | Other Asian | Wong et al. |
| Munez | Filipino | Other Asian | Wong et al. |
| Mungcal | Filipino | Other Asian | Wong et al. |
| Munim |  | South Asian | Lim et al. |
| Munir |  | South Asian | Lim et al. |
| Muniyappa |  | South Asian | Lim et al. |
| Muniz | Filipino | Other Asian | Wong et al. |
| Munjal | Asian Indian | South Asian | Wong et al. |
| Munmun |  | South Asian | Lim et al. |
| Munoa | Filipino | Other Asian | Wong et al. |
| Munoz | Filipino | Other Asian | Wong et al. |
| Munsayac | Filipino | Other Asian | Wong et al. |
| Munshi | Asian Indian | South Asian | Wong et al. |
| Munsif | Pakistani | South Asian | Kranick et al. |
| Munukutla | Asian Indian | South Asian | Wong et al. |
| Muoi | Vietnamese | Other Asian | Wong et al. |
| Muolic | Filipino | Other Asian | Wong et al. |
| Muon | Vietnamese | Other Asian | Wong et al. |
| Mupas | Filipino | Other Asian | Wong et al. |
| Mura | Japanese | Other Asian | Wong et al. |
| Murad |  | South Asian | Lim et al. |
| Muragishi | Japanese | Other Asian | Wong et al. |
| Murai | Japanese | Other Asian | Wong et al. |
| Muraishi | Japanese | Other Asian | Wong et al. |
| Murakami | Japanese | Other Asian | Wong et al. |
| Murakawa | Japanese | Other Asian | Wong et al. |
| Muraki | Japanese | Other Asian | Wong et al. |
| Murakoshi | Japanese | Other Asian | Wong et al. |
| Murallon | Filipino | Other Asian | Wong et al. |
| Muramatsu | Japanese | Other Asian | Wong et al. |
| Muramoto | Japanese | Other Asian | Wong et al. |
| Muranaka | Japanese | Other Asian | Wong et al. |
| Murao | Japanese | Other Asian | Wong et al. |
| Muraoka | Japanese | Other Asian | Wong et al. |
| Murarka | Asian Indian | South Asian | Wong et al. |
| Murase | Japanese | Other Asian | Wong et al. |
| Murashige | Japanese | Other Asian | Wong et al. |
| Murashima | Japanese | Other Asian | Wong et al. |
| Murata | Japanese | Other Asian | Wong et al. |
| Murayama | Japanese | Other Asian | Wong et al. |
| Murcia | Filipino | Other Asian | Wong et al. |
| Murgai | Asian Indian | South Asian | Wong et al. |
| Muriel | Filipino | Other Asian | Wong et al. |
| Murillo | Filipino | Other Asian | Wong et al. |
| Murofushi | Japanese | Other Asian | Wong et al. |
| Muroga | Japanese | Other Asian | Wong et al. |
| Muroi | Japanese | Other Asian | Wong et al. |
| Muromoto | Japanese | Other Asian | Wong et al. |
| Murota | Japanese | Other Asian | Wong et al. |
| Murotani | Japanese | Other Asian | Wong et al. |
| Muroya | Japanese | Other Asian | Wong et al. |
| Mursalini |  | South Asian | Lim et al. |
| Murshaed |  | South Asian | Lim et al. |
| Murshid |  | South Asian | Lim et al. |
| Murtaza | Pakistani | South Asian | Kranick et al. |
| Murthi | Asian Indian | South Asian | Wong et al. |
| Murthy | Asian Indian | South Asian | Wong et al. |
| Murti | Asian Indian | South Asian | Wong et al. |
| Murtuza | Asian Indian | South Asian | Wong et al. |
| Musa |  | South Asian | Lim et al. |
| Mushtaq |  | South Asian | Lim et al. |
| Musico | Filipino | Other Asian | Wong et al. |
| Musngi | Filipino | Other Asian | Wong et al. |
| Musni | Filipino | Other Asian | Wong et al. |
| Mustafa |  | South Asian | Lim et al. |
| Mustafoski |  | South Asian | Lim et al. |
| Mustapha |  | South Asian | Lim et al. |
| Muta | Japanese | Other Asian | Wong et al. |
| Mutaguchi | Japanese | Other Asian | Wong et al. |
| Mutalik | Asian Indian | South Asian | Wong et al. |
| Muthiah | Asian Indian | South Asian | Wong et al. |
| Muthktum |  | South Asian | Lim et al. |
| Muthu | Asian Indian | South Asian | Wong et al. |
| Muthukrishna | Asian Indian | South Asian | Wong et al. |
| Muthuswami | Asian Indian | South Asian | Wong et al. |
| Muthuswamy | Asian Indian | South Asian | Wong et al. |
| Mutia | Indonesian; Filipino | Other Asian | Kranick et al. |
| Muto | Japanese | Other Asian | Wong et al. |
| Mutoh | Japanese | Other Asian | Wong et al. |
| Mutuc | Filipino | Other Asian | Wong et al. |
| Muya | Filipino | Other Asian | Wong et al. |
| Muyco | Filipino | Other Asian | Wong et al. |
| Muyot | Filipino | Other Asian | Wong et al. |
| Muzahid |  | South Asian | Lim et al. |
| Muzammel |  | South Asian | Lim et al. |
| Muzones | Filipino | Other Asian | Wong et al. |
| My | Vietnamese | Other Asian | Wong et al. |
| Mycoosingh |  | South Asian | Lim et al. |
| Myong | Korean | Other Asian | Wong et al. |
| Myoung | Korean | Other Asian | Wong et al. |
| Myung | Korean | Other Asian | Kranick et al. |
| N | Asian Indian | South Asian | Wong et al. |
| Na | Korean | Other Asian | Kranick et al. |
| Naanep | Filipino | Other Asian | Wong et al. |
| Nabeshima | Japanese | Other Asian | Wong et al. |
| Nabeta | Japanese | Other Asian | Wong et al. |
| Nable | Filipino | Other Asian | Wong et al. |
| Naboa | Filipino | Other Asian | Wong et al. |
| Nabong | Filipino | Other Asian | Wong et al. |
| Nabor | Filipino | Other Asian | Wong et al. |
| Nabua | Filipino | Other Asian | Wong et al. |
| Nabung | Filipino | Other Asian | Wong et al. |
| Nacapuy | Filipino | Other Asian | Wong et al. |
| Nacar | Filipino | Other Asian | Wong et al. |
| Nacario | Filipino | Other Asian | Wong et al. |
| Naces | Filipino | Other Asian | Wong et al. |
| Nachor | Filipino | Other Asian | Wong et al. |
| Nacianceno | Filipino | Other Asian | Wong et al. |
| Nacino | Filipino | Other Asian | Wong et al. |
| Nacion | Filipino | Other Asian | Wong et al. |
| Nacional | Filipino | Other Asian | Wong et al. |
| Nacionales | Filipino | Other Asian | Wong et al. |
| Nacis | Filipino | Other Asian | Wong et al. |
| Nacnac | Filipino | Other Asian | Wong et al. |
| Nacor | Filipino | Other Asian | Wong et al. |
| Nacorda | Filipino | Other Asian | Wong et al. |
| Nacpil | Filipino | Other Asian | Wong et al. |
| Nacu | Filipino | Other Asian | Wong et al. |
| Nacua | Filipino | Other Asian | Wong et al. |
| Nada | Japanese | Other Asian | Wong et al. |
| Nadal | Filipino | Other Asian | Wong et al. |
| Nadeem |  | South Asian | Lim et al. |
| Nadela | Filipino | Other Asian | Wong et al. |
| Nadkarni | Asian Indian | South Asian | Wong et al. |
| Nadonga | Filipino | Other Asian | Wong et al. |
| Nadres | Filipino | Other Asian | Wong et al. |
| Nadura | Filipino | Other Asian | Wong et al. |
| Nadurata | Filipino | Other Asian | Wong et al. |
| Nadvi | Asian Indian | South Asian | Wong et al. |
| Nae | Korean | Other Asian | Wong et al. |
| Naeem |  | South Asian | Lim et al. |
| Naelgas | Filipino | Other Asian | Wong et al. |
| Naemura | Japanese | Other Asian | Wong et al. |
| Nafarrete | Filipino | Other Asian | Wong et al. |
| Nafrada | Filipino | Other Asian | Wong et al. |
| Nag |  | South Asian | Lim et al. |
| Naga | Japanese | Other Asian | Wong et al. |
| Nagabhushana | Asian Indian | South Asian | Wong et al. |
| Nagae | Japanese | Other Asian | Wong et al. |
| Nagafuchi | Japanese | Other Asian | Wong et al. |
| Nagahama | Japanese | Other Asian | Wong et al. |
| Nagahara | Japanese | Other Asian | Wong et al. |
| Nagahashi | Japanese | Other Asian | Wong et al. |
| Nagahiro | Japanese | Other Asian | Wong et al. |
| Nagahisa | Japanese | Other Asian | Wong et al. |
| Nagai | Japanese | Other Asian | Wong et al. |
| Nagaishi | Japanese | Other Asian | Wong et al. |
| Nagaki | Japanese | Other Asian | Wong et al. |
| Nagakubo | Japanese | Other Asian | Wong et al. |
| Nagakura | Japanese | Other Asian | Wong et al. |
| Nagal | Filipino | Other Asian | Wong et al. |
| Nagales | Filipino | Other Asian | Wong et al. |
| Nagamatsu | Japanese | Other Asian | Wong et al. |
| Nagami | Japanese | Other Asian | Wong et al. |
| Nagamine | Japanese | Other Asian | Wong et al. |
| Nagamori | Japanese | Other Asian | Wong et al. |
| Nagamoto | Japanese | Other Asian | Wong et al. |
| Nagamura | Japanese | Other Asian | Wong et al. |
| Nagano | Japanese | Other Asian | Wong et al. |
| Naganuma | Japanese | Other Asian | Wong et al. |
| Nagao | Japanese | Other Asian | Wong et al. |
| Nagaoka | Japanese | Other Asian | Wong et al. |
| Nagar | Asian Indian | South Asian | Wong et al. |
| Nagara | Japanese | Other Asian | Wong et al. |
| Nagaraj | Asian Indian | South Asian | Wong et al. |
| Nagaraja | Asian Indian | South Asian | Wong et al. |
| Nagarajan | Asian Indian | South Asian | Wong et al. |
| Nagaratnam | Asian Indian | South Asian | Wong et al. |
| Nagarkar | Asian Indian | South Asian | Wong et al. |
| Nagarsheth | Asian Indian | South Asian | Wong et al. |
| Nagasaka | Japanese | Other Asian | Wong et al. |
| Nagasaki | Japanese | Other Asian | Wong et al. |
| Nagasawa | Japanese | Other Asian | Wong et al. |
| Nagase | Japanese | Other Asian | Wong et al. |
| Nagashima | Japanese | Other Asian | Wong et al. |
| Nagata | Japanese | Other Asian | Wong et al. |
| Nagataki | Japanese | Other Asian | Wong et al. |
| Nagatani | Japanese | Other Asian | Wong et al. |
| Nagato | Japanese | Other Asian | Wong et al. |
| Nagatomi | Japanese | Other Asian | Wong et al. |
| Nagatomo | Japanese | Other Asian | Wong et al. |
| Nagatoshi | Japanese | Other Asian | Wong et al. |
| Nagatsu | Japanese | Other Asian | Wong et al. |
| Nagatsuka | Japanese | Other Asian | Wong et al. |
| Nagaya | Japanese | Other Asian | Wong et al. |
| Nagayama | Japanese | Other Asian | Wong et al. |
| Nagayo | Japanese | Other Asian | Wong et al. |
| Nagayoshi | Japanese | Other Asian | Wong et al. |
| Nagi | Asian Indian | South Asian | Kranick et al. |
| Nagindas | Asian Indian | South Asian | Wong et al. |
| Nagji | Asian Indian | South Asian | Wong et al. |
| Nago | Japanese | Other Asian | Wong et al. |
| Nagoshi | Japanese | Other Asian | Wong et al. |
| Nagpaul | Asian Indian | South Asian | Wong et al. |
| Nagra | Asian Indian | South Asian | Wong et al. |
| Nagrampa | Filipino | Other Asian | Wong et al. |
| Nagtalon | Filipino | Other Asian | Wong et al. |
| Nagui | Filipino | Other Asian | Wong et al. |
| Naguiat | Filipino | Other Asian | Wong et al. |
| Naguit | Filipino | Other Asian | Wong et al. |
| Nagumo | Japanese | Other Asian | Wong et al. |
| Nagun | Filipino | Other Asian | Wong et al. |
| Nagura | Japanese | Other Asian | Wong et al. |
| Nah | Korean | Other Asian | Wong et al. |
| Nahal | Asian Indian | South Asian | Wong et al. |
| Nahar | Bangladeshi | South Asian | Kranick et al. |
| Naher |  | South Asian | Lim et al. |
| Nahm | Korean | Other Asian | Wong et al. |
| Nai | Chinese | Chinese | Wong et al. |
| Naidas | Filipino | Other Asian | Wong et al. |
| Naido | Asian Indian | South Asian | Wong et al. |
| Naidu | Asian Indian | South Asian | Wong et al. |
| Naik | Asian Indian | South Asian | Wong et al. |
| Naiki | Japanese | Other Asian | Wong et al. |
| Naim | Asian Indian | South Asian | Wong et al. |
| Naipaul |  | South Asian | Lim et al. |
| Nair | Asian Indian | South Asian | Kranick et al. |
| Naithani | Asian Indian | South Asian | Kranick et al. |
| Naito | Japanese | Other Asian | Kranick et al. |
| Najam | Pakistani | South Asian | Kranick et al. |
| Najarro | Filipino | Other Asian | Wong et al. |
| Najera | Filipino | Other Asian | Wong et al. |
| Naji |  | South Asian | Lim et al. |
| Najima | Japanese | Other Asian | Wong et al. |
| Najmi | Asian Indian | South Asian | Wong et al. |
| Najmuddin | Asian Indian | South Asian | Wong et al. |
| Najneen |  | South Asian | Lim et al. |
| Naka | Japanese | Other Asian | Wong et al. |
| Nakabayashi | Japanese | Other Asian | Wong et al. |
| Nakachi | Japanese | Other Asian | Wong et al. |
| Nakada | Japanese | Other Asian | Wong et al. |
| Nakadate | Japanese | Other Asian | Wong et al. |
| Nakade | Japanese | Other Asian | Wong et al. |
| Nakae | Japanese | Other Asian | Wong et al. |
| Nakagaki | Japanese | Other Asian | Wong et al. |
| Nakagami | Japanese | Other Asian | Wong et al. |
| Nakagawa | Japanese | Other Asian | Wong et al. |
| Nakagima | Japanese | Other Asian | Wong et al. |
| Nakagome | Japanese | Other Asian | Wong et al. |
| Nakagoshi | Japanese | Other Asian | Wong et al. |
| Nakahama | Japanese | Other Asian | Wong et al. |
| Nakahara | Japanese | Other Asian | Wong et al. |
| Nakahira | Japanese | Other Asian | Wong et al. |
| Nakahodo | Japanese | Other Asian | Wong et al. |
| Nakai | Japanese | Other Asian | Wong et al. |
| Nakaishi | Japanese | Other Asian | Wong et al. |
| Nakaji | Japanese | Other Asian | Wong et al. |
| Nakajima | Japanese | Other Asian | Wong et al. |
| Nakajo | Japanese | Other Asian | Wong et al. |
| Nakakura | Japanese | Other Asian | Wong et al. |
| Nakama | Japanese | Other Asian | Wong et al. |
| Nakamae | Japanese | Other Asian | Wong et al. |
| Nakamara | Japanese | Other Asian | Wong et al. |
| Nakamaru | Japanese | Other Asian | Wong et al. |
| Nakamatsu | Japanese | Other Asian | Wong et al. |
| Nakamichi | Japanese | Other Asian | Wong et al. |
| Nakamine | Japanese | Other Asian | Wong et al. |
| Nakamizo | Japanese | Other Asian | Wong et al. |
| Nakamori | Japanese | Other Asian | Wong et al. |
| Nakamoto | Japanese | Other Asian | Wong et al. |
| Nakamura | Japanese | Other Asian | Kranick et al. |
| Nakamuta | Japanese | Other Asian | Wong et al. |
| Nakandakari | Japanese | Other Asian | Wong et al. |
| Nakane | Japanese | Other Asian | Wong et al. |
| Nakanishi | Japanese | Other Asian | Wong et al. |
| Nakano | Japanese | Other Asian | Wong et al. |
| Nakao | Japanese | Other Asian | Wong et al. |
| Nakaoka | Japanese | Other Asian | Wong et al. |
| Nakarmi |  | South Asian | Lim et al. |
| Nakasato | Japanese | Other Asian | Wong et al. |
| Nakase | Japanese | Other Asian | Wong et al. |
| Nakashima | Japanese | Other Asian | Wong et al. |
| Nakasone | Japanese | Other Asian | Wong et al. |
| Nakasuji | Japanese | Other Asian | Wong et al. |
| Nakata | Japanese | Other Asian | Wong et al. |
| Nakatani | Japanese | Other Asian | Wong et al. |
| Nakato | Japanese | Other Asian | Wong et al. |
| Nakatsu | Japanese | Other Asian | Wong et al. |
| Nakatsuji | Japanese | Other Asian | Wong et al. |
| Nakatsuka | Japanese | Other Asian | Wong et al. |
| Nakatsukasa | Japanese | Other Asian | Wong et al. |
| Nakauchi | Japanese | Other Asian | Wong et al. |
| Nakawatase | Japanese | Other Asian | Wong et al. |
| Nakaya | Japanese | Other Asian | Wong et al. |
| Nakayama | Japanese | Other Asian | Wong et al. |
| Nakayasu | Japanese | Other Asian | Wong et al. |
| Nakayoshi | Japanese | Other Asian | Wong et al. |
| Nakaza | Japanese | Other Asian | Wong et al. |
| Nakazaki | Japanese | Other Asian | Wong et al. |
| Nakazato | Japanese | Other Asian | Wong et al. |
| Nakazawa | Japanese | Other Asian | Wong et al. |
| Nakazima | Japanese | Other Asian | Wong et al. |
| Nakazono | Japanese | Other Asian | Wong et al. |
| Nakhuda | Asian Indian | South Asian | Wong et al. |
| Nako | Japanese | Other Asian | Wong et al. |
| Nakpil | Filipino | Other Asian | Wong et al. |
| Nakrani | Asian Indian | South Asian | Wong et al. |
| Nalagan | Filipino | Other Asian | Wong et al. |
| Nalam | Filipino | Other Asian | Wong et al. |
| Nalapo | Filipino | Other Asian | Wong et al. |
| Nalda | Filipino | Other Asian | Wong et al. |
| Naldo | Filipino | Other Asian | Wong et al. |
| Naldoza | Filipino | Other Asian | Wong et al. |
| Naljit |  | South Asian | Lim et al. |
| Nalla | Asian Indian | South Asian | Kranick et al. |
| Nalupa | Filipino | Other Asian | Wong et al. |
| Naluz | Filipino | Other Asian | Wong et al. |
| Nalwa | Asian Indian | South Asian | Wong et al. |
| Nam | Korean | Other Asian | Kranick et al. |
| Namba | Japanese | Other Asian | Wong et al. |
| Nambiar | Asian Indian | South Asian | Wong et al. |
| Namboodiri | Asian Indian | South Asian | Wong et al. |
| Nambu | Japanese | Other Asian | Wong et al. |
| Name | Filipino | Other Asian | Wong et al. |
| Named | Vietnamese | Other Asian | Wong et al. |
| Namekawa | Japanese | Other Asian | Wong et al. |
| Nameraj |  | South Asian | Lim et al. |
| Namgoong | Korean | Other Asian | Wong et al. |
| Namgung | Korean | Other Asian | Wong et al. |
| Namikawa | Japanese | Other Asian | Wong et al. |
| Namiki | Japanese | Other Asian | Wong et al. |
| Namin | Filipino | Other Asian | Wong et al. |
| Namit | Filipino | Other Asian | Wong et al. |
| Namkoong | Korean | Other Asian | Wong et al. |
| Namkung | Korean | Other Asian | Wong et al. |
| Namnama | Filipino | Other Asian | Wong et al. |
| Namoc | Filipino | Other Asian | Wong et al. |
| Namoca | Filipino | Other Asian | Wong et al. |
| Nan | Chinese | Chinese | Wong et al. |
| Nana | Filipino | Other Asian | Wong et al. |
| Nanagas | Filipino | Other Asian | Wong et al. |
| Nanavati | Asian Indian | South Asian | Wong et al. |
| Nanavaty | Asian Indian | South Asian | Wong et al. |
| Nanba | Japanese | Other Asian | Wong et al. |
| Nanbu | Japanese | Other Asian | Wong et al. |
| Nanca | Filipino | Other Asian | Wong et al. |
| Nand | Asian Indian | South Asian | Wong et al. |
| Nanda | Asian Indian | South Asian | Wong et al. |
| Nandalall |  | South Asian | Lim et al. |
| Nandi | Asian Indian | South Asian | Wong et al. |
| Nandial |  | South Asian | Lim et al. |
| Nandkumar |  | South Asian | Lim et al. |
| Nandlal |  | South Asian | Lim et al. |
| Nandy | Bangladeshi; Asian Indian | South Asian | Kranick et al. |
| Nanez | Filipino | Other Asian | Wong et al. |
| Naniong | Filipino | Other Asian | Wong et al. |
| Nanjappa | Asian Indian | South Asian | Wong et al. |
| Nanji | Asian Indian | South Asian | Wong et al. |
| Nanjo | Japanese | Other Asian | Wong et al. |
| Nanjundiah | Asian Indian | South Asian | Wong et al. |
| Nano | Filipino | Other Asian | Wong et al. |
| Nanola | Filipino | Other Asian | Wong et al. |
| Nanquil | Filipino | Other Asian | Wong et al. |
| Nanri | Japanese | Other Asian | Wong et al. |
| Nantes | Filipino | Other Asian | Wong et al. |
| Naoe | Filipino | Other Asian | Wong et al. |
| Naoi | Japanese | Other Asian | Wong et al. |
| Napalan | Filipino | Other Asian | Wong et al. |
| Napenas | Filipino | Other Asian | Wong et al. |
| Napitupulu | Indonesian | Other Asian | Kranick et al. |
| Napiza | Filipino | Other Asian | Wong et al. |
| Napoles | Filipino | Other Asian | Wong et al. |
| Napuli | Filipino | Other Asian | Wong et al. |
| Naqi | Asian Indian | South Asian | Wong et al. |
| Naqui | Asian Indian | South Asian | Wong et al. |
| Naqvi | Pakistani | South Asian | Kranick et al. |
| Nara | Japanese | Other Asian | Wong et al. |
| Narag | Filipino | Other Asian | Wong et al. |
| Narahara | Japanese | Other Asian | Wong et al. |
| Narain | Asian Indian | South Asian | Kranick et al. |
| Narainasami |  | South Asian | Lim et al. |
| Naraindas | Asian Indian | South Asian | Wong et al. |
| Naraine |  | South Asian | Lim et al. |
| Naraja | Filipino | Other Asian | Wong et al. |
| Naran | Asian Indian | South Asian | Wong et al. |
| Narandas | Asian Indian | South Asian | Wong et al. |
| Narang |  | South Asian | Lim et al. |
| Naranja | Filipino | Other Asian | Wong et al. |
| Naranji | Asian Indian | South Asian | Wong et al. |
| Naranjo | Filipino | Other Asian | Wong et al. |
| Narasimhacha | Asian Indian | South Asian | Wong et al. |
| Narasimham | Asian Indian | South Asian | Wong et al. |
| Narasimhan | Asian Indian | South Asian | Wong et al. |
| Narasingarao | Asian Indian | South Asian | Wong et al. |
| Naraval | Filipino | Other Asian | Wong et al. |
| Narayan | Asian Indian | South Asian | Wong et al. |
| Narayana | Asian Indian | South Asian | Kranick et al. |
| Narayanan | Asian Indian; Malaysian | South Asian | Kranick et al. |
| Narayanarao | Asian Indian | South Asian | Wong et al. |
| Narayanasamy | Asian Indian | South Asian | Wong et al. |
| Narayanaswam | Asian Indian | South Asian | Wong et al. |
| Narayanswami | Asian Indian | South Asian | Wong et al. |
| Narazaki | Japanese | Other Asian | Wong et al. |
| Narcelles | Filipino | Other Asian | Wong et al. |
| Narcida | Filipino | Other Asian | Wong et al. |
| Narciso | Filipino | Other Asian | Wong et al. |
| Naredo | Filipino | Other Asian | Wong et al. |
| Nargis |  | South Asian | Lim et al. |
| Narido | Filipino | Other Asian | Wong et al. |
| Narielwala | Asian Indian | South Asian | Wong et al. |
| Narikawa | Japanese | Other Asian | Wong et al. |
| Narimatsu | Japanese | Other Asian | Wong et al. |
| Narine |  | South Asian | Lim et al. |
| Nario | Filipino | Other Asian | Wong et al. |
| Narisawa | Japanese | Other Asian | Wong et al. |
| Narisma | Filipino | Other Asian | Wong et al. |
| Narita | Japanese | Other Asian | Wong et al. |
| Narito | Filipino | Other Asian | Wong et al. |
| Narrido | Filipino | Other Asian | Wong et al. |
| Narsai | Asian Indian | South Asian | Wong et al. |
| Nartates | Filipino | Other Asian | Wong et al. |
| Nartatez | Filipino | Other Asian | Wong et al. |
| Narte | Filipino | Other Asian | Wong et al. |
| Nartea | Filipino | Other Asian | Wong et al. |
| Narula | Asian Indian | South Asian | Kranick et al. |
| Narumi | Japanese | Other Asian | Wong et al. |
| Narusawa | Japanese | Other Asian | Wong et al. |
| Naruse | Japanese | Other Asian | Wong et al. |
| Narushima | Japanese | Other Asian | Wong et al. |
| Narvaez | Filipino | Other Asian | Wong et al. |
| Narvarte | Filipino | Other Asian | Wong et al. |
| Narvas | Filipino | Other Asian | Wong et al. |
| Narvasa | Filipino | Other Asian | Wong et al. |
| Narvaza | Filipino | Other Asian | Wong et al. |
| Narvios | Filipino | Other Asian | Wong et al. |
| Narwal | Asian Indian | South Asian | Wong et al. |
| Narwani | Asian Indian | South Asian | Wong et al. |
| Nasa |  | South Asian | Lim et al. |
| Naseem | Pakistani | South Asian | Kranick et al. |
| Naseer | Asian Indian | South Asian | Wong et al. |
| Naseeruddin | Asian Indian | South Asian | Wong et al. |
| Naser |  | South Asian | Lim et al. |
| Nasib |  | South Asian | Lim et al. |
| Nasif |  | South Asian | Lim et al. |
| Nasim |  | South Asian | Lim et al. |
| Nasima |  | South Asian | Lim et al. |
| Nasir |  | South Asian | Lim et al. |
| Nasiruddin | Asian Indian | South Asian | Wong et al. |
| Nasis | Filipino | Other Asian | Wong et al. |
| Nasol | Filipino | Other Asian | Wong et al. |
| Nasreen |  | South Asian | Lim et al. |
| Nasrin | Bangladeshi | South Asian | Kranick et al. |
| Nastor | Filipino | Other Asian | Wong et al. |
| Nasu | Japanese | Other Asian | Wong et al. |
| Nat | Asian Indian | South Asian | Wong et al. |
| Natac | Filipino | Other Asian | Wong et al. |
| Natalio | Filipino | Other Asian | Wong et al. |
| Natan | Filipino | Other Asian | Wong et al. |
| Natanauan | Filipino | Other Asian | Wong et al. |
| Nataraj | Asian Indian | South Asian | Wong et al. |
| Natarajan | Asian Indian | South Asian | Kranick et al. |
| Natesan | Asian Indian | South Asian | Wong et al. |
| Nath | Asian Indian | South Asian | Kranick et al. |
| Nathabhai | Asian Indian | South Asian | Wong et al. |
| Nathaniel | Asian Indian | South Asian | Wong et al. |
| Nathoo | Asian Indian | South Asian | Wong et al. |
| Nathu | Asian Indian | South Asian | Wong et al. |
| Nathubhai | Asian Indian | South Asian | Wong et al. |
| Nathwani | Asian Indian | South Asian | Wong et al. |
| Natino | Filipino | Other Asian | Wong et al. |
| Natividad | Filipino | Other Asian | Kranick et al. |
| Natnat | Filipino | Other Asian | Wong et al. |
| Nato | Filipino | Other Asian | Wong et al. |
| Nator | Filipino | Other Asian | Wong et al. |
| Natori | Japanese | Other Asian | Wong et al. |
| Natsume | Japanese | Other Asian | Wong et al. |
| Naui | Filipino | Other Asian | Wong et al. |
| Nauman | Pakistani | South Asian | Kranick et al. |
| Naungayan | Filipino | Other Asian | Wong et al. |
| Nauyen | Vietnamese | Other Asian | Wong et al. |
| Nava | Filipino | Other Asian | Wong et al. |
| Navaja | Filipino | Other Asian | Wong et al. |
| Naval | Filipino | Other Asian | Wong et al. |
| Navales | Filipino | Other Asian | Wong et al. |
| Navalkar | Asian Indian | South Asian | Kranick et al. |
| Navallo | Filipino | Other Asian | Wong et al. |
| Navalta | Filipino | Other Asian | Wong et al. |
| Navares | Filipino | Other Asian | Wong et al. |
| Navarette | Filipino | Other Asian | Wong et al. |
| Navarez | Filipino | Other Asian | Wong et al. |
| Navarrete | Filipino | Other Asian | Wong et al. |
| Navarro | Filipino | Other Asian | Wong et al. |
| Navarroza | Filipino | Other Asian | Wong et al. |
| Navas | Filipino | Other Asian | Wong et al. |
| Navasca | Filipino | Other Asian | Wong et al. |
| Navat | Filipino | Other Asian | Wong et al. |
| Navata | Filipino | Other Asian | Wong et al. |
| Navato | Filipino | Other Asian | Wong et al. |
| Navea | Filipino | Other Asian | Wong et al. |
| Navera | Filipino | Other Asian | Wong et al. |
| Navia | Filipino | Other Asian | Wong et al. |
| Navidad | Filipino | Other Asian | Wong et al. |
| Navo | Filipino | Other Asian | Wong et al. |
| Navoa | Filipino | Other Asian | Wong et al. |
| Navor | Filipino | Other Asian | Wong et al. |
| Nawa | Japanese | Other Asian | Wong et al. |
| Nawab | Asian Indian | South Asian | Wong et al. |
| Nawata | Japanese | Other Asian | Wong et al. |
| Nawaz | Pakistani | South Asian | Kranick et al. |
| Nayak | Asian Indian | South Asian | Wong et al. |
| Nayampalli | Asian Indian | South Asian | Wong et al. |
| Nayar | Asian Indian | South Asian | Wong et al. |
| Nayee | Asian Indian | South Asian | Wong et al. |
| Nayeem | Asian Indian | South Asian | Wong et al. |
| Nayem |  | South Asian | Lim et al. |
| Nayve | Filipino | Other Asian | Wong et al. |
| Nayyar | Pakistani | South Asian | Kranick et al. |
| Nayyer | Pakistani | South Asian | Kranick et al. |
| Naz | Pakistani | South Asian | Kranick et al. |
| Nazaire | Filipino | Other Asian | Wong et al. |
| Nazal | Filipino | Other Asian | Wong et al. |
| Nazarali | Asian Indian | South Asian | Wong et al. |
| Nazareno | Filipino | Other Asian | Wong et al. |
| Nazario | Filipino | Other Asian | Wong et al. |
| Nazarro | Filipino | Other Asian | Wong et al. |
| Nazibullah |  | South Asian | Lim et al. |
| Nazim | Asian Indian | South Asian | Wong et al. |
| Nazir |  | South Asian | Lim et al. |
| Nazmun |  | South Asian | Lim et al. |
| Naznin |  | South Asian | Lim et al. |
| Nazrul |  | South Asian | Lim et al. |
| Nebalasca | Filipino | Other Asian | Wong et al. |
| Nebre | Filipino | Other Asian | Wong et al. |
| Nebres | Filipino | Other Asian | Wong et al. |
| Nebriaga | Filipino | Other Asian | Wong et al. |
| Nebrida | Filipino | Other Asian | Wong et al. |
| Nebril | Filipino | Other Asian | Wong et al. |
| Necesario | Filipino | Other Asian | Wong et al. |
| Necesito | Filipino | Other Asian | Wong et al. |
| Nedic | Filipino | Other Asian | Wong et al. |
| Nee | Chinese | Chinese | Wong et al. |
| Neelakantan | Asian Indian | South Asian | Wong et al. |
| Negami | Japanese | Other Asian | Wong et al. |
| Negandhi | Asian Indian | South Asian | Wong et al. |
| Negi | Asian Indian | South Asian | Wong et al. |
| Negishi | Japanese | Other Asian | Wong et al. |
| Negoro | Japanese | Other Asian | Wong et al. |
| Negranza | Filipino | Other Asian | Wong et al. |
| Negre | Filipino | Other Asian | Wong et al. |
| Negrillo | Filipino | Other Asian | Wong et al. |
| Nehra | Asian Indian | South Asian | Wong et al. |
| Nehru | Asian Indian | South Asian | Wong et al. |
| Nejal | Filipino | Other Asian | Wong et al. |
| Nellas | Filipino | Other Asian | Wong et al. |
| Nelmida | Filipino | Other Asian | Wong et al. |
| Nelufa |  | South Asian | Lim et al. |
| Nemani | Asian Indian | South Asian | Wong et al. |
| Nembangkhanal |  | South Asian | Lim et al. |
| Nemoto | Japanese | Other Asian | Wong et al. |
| Nen | Chinese | Chinese | Wong et al. |
| Neogi | Asian Indian | South Asian | Wong et al. |
| Nepal |  | South Asian | Lim et al. |
| Nepalgautam |  | South Asian | Lim et al. |
| Nepomuceno | Filipino | Other Asian | Wong et al. |
| Ner | Filipino | Other Asian | Wong et al. |
| Nera | Filipino | Other Asian | Wong et al. |
| Neral | Filipino | Other Asian | Wong et al. |
| Neri | Filipino | Other Asian | Wong et al. |
| Neria | Filipino | Other Asian | Wong et al. |
| Nerida | Filipino | Other Asian | Wong et al. |
| Nerit | Filipino | Other Asian | Wong et al. |
| Nerona | Filipino | Other Asian | Wong et al. |
| Nerpio | Filipino | Other Asian | Wong et al. |
| Nerpiol | Filipino | Other Asian | Wong et al. |
| Nerurkar | Asian Indian | South Asian | Wong et al. |
| Nerveza | Filipino | Other Asian | Wong et al. |
| Nery | Filipino | Other Asian | Wong et al. |
| Nesa |  | South Asian | Lim et al. |
| Nesan |  | South Asian | Lim et al. |
| Nesha |  | South Asian | Lim et al. |
| Nessa | Asian Indian; Bangladeshi | South Asian | Kranick et al. |
| Nessia | Filipino | Other Asian | Wong et al. |
| Neti | Asian Indian | South Asian | Kranick et al. |
| Neupane |  | South Asian | Lim et al. |
| Neuyen | Vietnamese | Other Asian | Wong et al. |
| Nevado | Filipino | Other Asian | Wong et al. |
| Neverida | Filipino | Other Asian | Wong et al. |
| Neville |  | South Asian | Lim et al. |
| Neypes | Filipino | Other Asian | Wong et al. |
| Neyra | Filipino | Other Asian | Wong et al. |
| Nezu | Japanese | Other Asian | Wong et al. |
| Ng | Chinese | Chinese | Kranick et al. |
| Ngac | Vietnamese | Other Asian | Wong et al. |
| Ngai | Chinese | Chinese | Kranick et al. |
| Ngan | Chinese | Chinese | Wong et al. |
| Ngao | Chinese | Chinese | Wong et al. |
| Ngar | Chinese | Chinese | Wong et al. |
| Ngau | Chinese | Chinese | Wong et al. |
| Ngay | Chinese | Chinese | Wong et al. |
| Ngayan | Filipino | Other Asian | Wong et al. |
| Ngee | Chinese | Chinese | Wong et al. |
| Ngew | Chinese | Chinese | Wong et al. |
| Nghe | Vietnamese | Other Asian | Wong et al. |
| Nghi | Vietnamese | Other Asian | Kranick et al. |
| Nghia | Vietnamese | Other Asian | Wong et al. |
| Nghiem | Vietnamese | Other Asian | Wong et al. |
| Nghien | Vietnamese | Other Asian | Wong et al. |
| Nghiep | Vietnamese | Other Asian | Wong et al. |
| Nghieu | Vietnamese | Other Asian | Wong et al. |
| Ngi | Chinese | Chinese | Wong et al. |
| Ngim | Chinese | Chinese | Wong et al. |
| Ngin | Chinese | Chinese | Wong et al. |
| Nging | Chinese | Chinese | Wong et al. |
| Ngip | Chinese | Chinese | Wong et al. |
| Ngo | Vietnamese | Other Asian | Kranick et al. |
| Ngoc | Vietnamese | Other Asian | Wong et al. |
| Ngoi | Vietnamese | Other Asian | Wong et al. |
| Ngok | Chinese | Chinese | Wong et al. |
| Ngon | Chinese | Chinese | Wong et al. |
| Ngong | Chinese | Chinese | Wong et al. |
| Ngook | Chinese | Chinese | Wong et al. |
| Ngoon | Chinese | Chinese | Wong et al. |
| Ngor | Chinese | Chinese | Wong et al. |
| Ngow | Chinese | Chinese | Wong et al. |
| Ngoyen | Vietnamese | Other Asian | Wong et al. |
| Ngu | Vietnamese | Other Asian | Kranick et al. |
| Nguen | Vietnamese | Other Asian | Wong et al. |
| Nguey | Chinese | Chinese | Wong et al. |
| Ngueyn | Vietnamese | Other Asian | Wong et al. |
| Ngugen | Vietnamese | Other Asian | Wong et al. |
| Ngui | Malaysian | Other Asian | Kranick et al. |
| Ngun | Chinese | Chinese | Wong et al. |
| Nguu | Vietnamese | Other Asian | Wong et al. |
| Nguy | Vietnamese | Other Asian | Wong et al. |
| Nguyan | Vietnamese | Other Asian | Wong et al. |
| Nguye | Vietnamese | Other Asian | Wong et al. |
| Nguyen | Vietnamese | Other Asian | Kranick et al. |
| Nguyenvan | Vietnamese | Other Asian | Wong et al. |
| Nguyet | Vietnamese | Other Asian | Wong et al. |
| Nguyne | Vietnamese | Other Asian | Wong et al. |
| Ngyen | Vietnamese | Other Asian | Wong et al. |
| Ngyuen | Vietnamese | Other Asian | Wong et al. |
| Nha | Vietnamese | Other Asian | Wong et al. |
| Nham | Vietnamese | Other Asian | Wong et al. |
| Nhan | Vietnamese | Other Asian | Wong et al. |
| Nhat | Vietnamese | Other Asian | Wong et al. |
| Nhi | Vietnamese | Other Asian | Wong et al. |
| Nhiem | Vietnamese | Other Asian | Wong et al. |
| Nhien | Vietnamese | Other Asian | Wong et al. |
| Nhieu | Vietnamese | Other Asian | Wong et al. |
| Nhin | Vietnamese | Other Asian | Wong et al. |
| Nho | Vietnamese | Other Asian | Wong et al. |
| Nhon | Vietnamese | Other Asian | Wong et al. |
| Nhu | Vietnamese | Other Asian | Wong et al. |
| Nhuan | Vietnamese | Other Asian | Wong et al. |
| Nhung | Vietnamese | Other Asian | Wong et al. |
| Nhuong | Vietnamese | Other Asian | Wong et al. |
| Nhut | Vietnamese | Other Asian | Wong et al. |
| Nhuyen | Vietnamese | Other Asian | Wong et al. |
| Ni | Chinese | Chinese | Wong et al. |
| Niazi | Pakistani | South Asian | Kranick et al. |
| Nicandro | Filipino | Other Asian | Wong et al. |
| Nicanor | Filipino | Other Asian | Wong et al. |
| Nicart | Filipino | Other Asian | Wong et al. |
| Nicasio | Filipino | Other Asian | Wong et al. |
| Nicdao | Filipino | Other Asian | Wong et al. |
| Nicio | Filipino | Other Asian | Wong et al. |
| Nicolas | Filipino | Other Asian | Wong et al. |
| Nicolasora | Filipino | Other Asian | Wong et al. |
| Nicomedes | Filipino | Other Asian | Wong et al. |
| Nicomedez | Filipino | Other Asian | Wong et al. |
| Nidea | Filipino | Other Asian | Wong et al. |
| Nido | Filipino | Other Asian | Wong et al. |
| Nidoy | Filipino | Other Asian | Wong et al. |
| Niduaza | Filipino | Other Asian | Wong et al. |
| Nie | Chinese | Chinese | Wong et al. |
| Niebres | Filipino | Other Asian | Wong et al. |
| Niedo | Filipino | Other Asian | Wong et al. |
| Nieh | Chinese | Chinese | Wong et al. |
| Nien | Chinese | Chinese | Wong et al. |
| Niere | Filipino | Other Asian | Wong et al. |
| Nierras | Filipino | Other Asian | Wong et al. |
| Nierva | Filipino | Other Asian | Wong et al. |
| Nietes | Filipino | Other Asian | Wong et al. |
| Nieto | Filipino | Other Asian | Wong et al. |
| Nieva | Filipino | Other Asian | Wong et al. |
| Nievera | Filipino | Other Asian | Wong et al. |
| Nieveras | Filipino | Other Asian | Wong et al. |
| Nieves | Filipino | Other Asian | Wong et al. |
| Nigam | Asian Indian | South Asian | Wong et al. |
| Nigo | Japanese | Other Asian | Wong et al. |
| Nigos | Filipino | Other Asian | Wong et al. |
| Niguidula | Filipino | Other Asian | Wong et al. |
| Nihei | Japanese | Other Asian | Wong et al. |
| Niher |  | South Asian | Lim et al. |
| Nihira | Japanese | Other Asian | Wong et al. |
| Nii | Japanese | Other Asian | Wong et al. |
| Niida | Japanese | Other Asian | Wong et al. |
| Niijima | Japanese | Other Asian | Wong et al. |
| Niikura | Japanese | Other Asian | Wong et al. |
| Niimi | Japanese | Other Asian | Wong et al. |
| Niimoto | Japanese | Other Asian | Wong et al. |
| Niimura | Japanese | Other Asian | Wong et al. |
| Niino | Japanese | Other Asian | Wong et al. |
| Niioka | Japanese | Other Asian | Wong et al. |
| Niiyama | Japanese | Other Asian | Wong et al. |
| Niizawa | Japanese | Other Asian | Wong et al. |
| Niizeki | Japanese | Other Asian | Wong et al. |
| Niizuma | Japanese | Other Asian | Wong et al. |
| Nijhawan | Asian Indian | South Asian | Wong et al. |
| Nijjar | Asian Indian | South Asian | Wong et al. |
| Nijjer | Asian Indian | South Asian | Wong et al. |
| Nikaido | Japanese | Other Asian | Wong et al. |
| Niki | Japanese | Other Asian | Wong et al. |
| Nilakantan | Asian Indian | South Asian | Wong et al. |
| Nillo | Filipino | Other Asian | Wong et al. |
| Nillos | Filipino | Other Asian | Wong et al. |
| Nilo | Filipino | Other Asian | Wong et al. |
| Nim | Vietnamese | Other Asian | Wong et al. |
| Nimes | Filipino | Other Asian | Wong et al. |
| Nimmagadda | Asian Indian | South Asian | Wong et al. |
| Nimo | Filipino | Other Asian | Wong et al. |
| Nimura | Japanese | Other Asian | Wong et al. |
| Nin | Chinese | Chinese | Wong et al. |
| Ninal | Filipino | Other Asian | Wong et al. |
| Ninan | Asian Indian | South Asian | Wong et al. |
| Ning | Chinese | Chinese | Wong et al. |
| Ninh | Vietnamese | Other Asian | Wong et al. |
| Nino | Filipino | Other Asian | Wong et al. |
| Ninobla | Filipino | Other Asian | Wong et al. |
| Ninomiya | Japanese | Other Asian | Wong et al. |
| Ninonuevo | Filipino | Other Asian | Wong et al. |
| Nip | Chinese | Chinese | Wong et al. |
| Nipales | Filipino | Other Asian | Wong et al. |
| Nipay | Filipino | Other Asian | Wong et al. |
| Nirei | Japanese | Other Asian | Wong et al. |
| Nirona | Filipino | Other Asian | Wong et al. |
| Niroula |  | South Asian | Lim et al. |
| Nirula | Asian Indian | South Asian | Wong et al. |
| Nisa | Asian Indian; Indonesian; Pakistani | South Asian | Kranick et al. |
| Nisha |  | South Asian | Lim et al. |
| Nishi | Japanese | Other Asian | Wong et al. |
| Nishibata | Japanese | Other Asian | Wong et al. |
| Nishibayashi | Japanese | Other Asian | Wong et al. |
| Nishibe | Japanese | Other Asian | Wong et al. |
| Nishibori | Japanese | Other Asian | Wong et al. |
| Nishida | Japanese | Other Asian | Wong et al. |
| Nishide | Japanese | Other Asian | Wong et al. |
| Nishie | Japanese | Other Asian | Wong et al. |
| Nishigaki | Japanese | Other Asian | Wong et al. |
| Nishigori | Japanese | Other Asian | Wong et al. |
| Nishiguchi | Japanese | Other Asian | Wong et al. |
| Nishihama | Japanese | Other Asian | Wong et al. |
| Nishihara | Japanese | Other Asian | Wong et al. |
| Nishihata | Japanese | Other Asian | Wong et al. |
| Nishihira | Japanese | Other Asian | Wong et al. |
| Nishii | Japanese | Other Asian | Wong et al. |
| Nishijima | Japanese | Other Asian | Wong et al. |
| Nishikata | Japanese | Other Asian | Wong et al. |
| Nishikawa | Japanese | Other Asian | Wong et al. |
| Nishiki | Japanese | Other Asian | Wong et al. |
| Nishikiori | Japanese | Other Asian | Wong et al. |
| Nishimaki | Japanese | Other Asian | Wong et al. |
| Nishimatsu | Japanese | Other Asian | Wong et al. |
| Nishimiya | Japanese | Other Asian | Wong et al. |
| Nishimori | Japanese | Other Asian | Wong et al. |
| Nishimoto | Japanese | Other Asian | Wong et al. |
| Nishimura | Japanese | Other Asian | Wong et al. |
| Nishina | Japanese | Other Asian | Wong et al. |
| Nishinaka | Japanese | Other Asian | Wong et al. |
| Nishino | Japanese | Other Asian | Wong et al. |
| Nishio | Japanese | Other Asian | Wong et al. |
| Nishioka | Japanese | Other Asian | Wong et al. |
| Nishishita | Japanese | Other Asian | Wong et al. |
| Nishita | Japanese | Other Asian | Wong et al. |
| Nishitani | Japanese | Other Asian | Wong et al. |
| Nishiuchi | Japanese | Other Asian | Wong et al. |
| Nishiura | Japanese | Other Asian | Wong et al. |
| Nishiwaki | Japanese | Other Asian | Wong et al. |
| Nishiyama | Japanese | Other Asian | Wong et al. |
| Nishiyori | Japanese | Other Asian | Wong et al. |
| Nishizaka | Japanese | Other Asian | Wong et al. |
| Nishizaki | Japanese | Other Asian | Wong et al. |
| Nishizawa | Japanese | Other Asian | Wong et al. |
| Nisnisan | Filipino | Other Asian | Wong et al. |
| Nisperos | Filipino | Other Asian | Wong et al. |
| Nitafan | Filipino | Other Asian | Wong et al. |
| Nitao | Japanese | Other Asian | Wong et al. |
| Nite | Filipino | Other Asian | Wong et al. |
| Nito | Filipino | Other Asian | Wong et al. |
| Nitta | Japanese | Other Asian | Wong et al. |
| Nitura | Filipino | Other Asian | Wong et al. |
| Niu | Chinese | Chinese | Wong et al. |
| Nivera | Filipino | Other Asian | Wong et al. |
| Niwa | Japanese | Other Asian | Wong et al. |
| Niyogi | Asian Indian | South Asian | Wong et al. |
| Nizami | Asian Indian | South Asian | Wong et al. |
| Nizamuddin |  | South Asian | Lim et al. |
| Nizamudeen |  | South Asian | Lim et al. |
| Nknown | Vietnamese | Other Asian | Wong et al. |
| No | Korean | Other Asian | Wong et al. |
| Nobata | Japanese | Other Asian | Wong et al. |
| Nobe | Japanese | Other Asian | Wong et al. |
| Nobello | Filipino | Other Asian | Wong et al. |
| Nobi |  | South Asian | Lim et al. |
| Nobida | Filipino | Other Asian | Wong et al. |
| Noblejas | Filipino | Other Asian | Wong et al. |
| Nobleza | Filipino | Other Asian | Wong et al. |
| Noboru | Japanese | Other Asian | Wong et al. |
| Nobuhara | Japanese | Other Asian | Wong et al. |
| Nobuo | Japanese | Other Asian | Wong et al. |
| Noceda | Filipino | Other Asian | Wong et al. |
| Noces | Filipino | Other Asian | Wong et al. |
| Noche | Filipino | Other Asian | Wong et al. |
| Nocom | Filipino | Other Asian | Wong et al. |
| Nocon | Filipino | Other Asian | Wong et al. |
| Nocos | Filipino | Other Asian | Wong et al. |
| Nocum | Filipino | Other Asian | Wong et al. |
| Noda | Japanese | Other Asian | Kranick et al. |
| Nodado | Filipino | Other Asian | Wong et al. |
| Nodora | Filipino | Other Asian | Wong et al. |
| Nogami | Japanese | Other Asian | Wong et al. |
| Nogawa | Japanese | Other Asian | Wong et al. |
| Nogoy | Filipino | Other Asian | Wong et al. |
| Nogra | Filipino | Other Asian | Wong et al. |
| Noguchi | Japanese | Other Asian | Wong et al. |
| Noguera | Filipino | Other Asian | Wong et al. |
| Noh | Korean | Other Asian | Kranick et al. |
| Nohara | Japanese | Other Asian | Wong et al. |
| Nojadera | Filipino | Other Asian | Wong et al. |
| Noji | Japanese | Other Asian | Wong et al. |
| Nojima | Japanese | Other Asian | Wong et al. |
| Nojiri | Japanese | Other Asian | Wong et al. |
| Nok | Korean | Other Asian | Wong et al. |
| Nolasco | Filipino | Other Asian | Wong et al. |
| Nolastname | Asian Indian | South Asian | Wong et al. |
| Nollido | Filipino | Other Asian | Wong et al. |
| Nom | Chinese | Chinese | Wong et al. |
| Noma | Japanese | Other Asian | Wong et al. |
| Nomachi | Japanese | Other Asian | Wong et al. |
| Nomaguchi | Japanese | Other Asian | Wong et al. |
| Noman |  | South Asian | Lim et al. |
| Nomi | Japanese | Other Asian | Wong et al. |
| Nomiyama | Japanese | Other Asian | Wong et al. |
| Nomoto | Japanese | Other Asian | Wong et al. |
| Nomura | Japanese | Other Asian | Wong et al. |
| Non | Chinese | Chinese | Wong et al. |
| Nonaka | Japanese | Other Asian | Wong et al. |
| Noname | Asian Indian | South Asian | Wong et al. |
| Nonan | Filipino | Other Asian | Wong et al. |
| Nonato | Filipino | Other Asian | Wong et al. |
| Nones | Filipino | Other Asian | Wong et al. |
| Nonisa | Filipino | Other Asian | Wong et al. |
| Nono | Filipino | Other Asian | Wong et al. |
| Nonog | Filipino | Other Asian | Wong et al. |
| Nonomura | Japanese | Other Asian | Wong et al. |
| Nonoyama | Japanese | Other Asian | Wong et al. |
| Nool | Filipino | Other Asian | Wong et al. |
| Noor |  | South Asian | Lim et al. |
| Noora | Filipino | Other Asian | Wong et al. |
| Noorani | Asian Indian | South Asian | Wong et al. |
| Nooruddin | Asian Indian | South Asian | Wong et al. |
| Norberte | Filipino | Other Asian | Wong et al. |
| Norico | Filipino | Other Asian | Wong et al. |
| Noriega | Filipino | Other Asian | Wong et al. |
| Noriel | Filipino | Other Asian | Wong et al. |
| Norimatsu | Japanese | Other Asian | Wong et al. |
| Noro | Japanese | Other Asian | Wong et al. |
| Norona | Filipino | Other Asian | Wong et al. |
| Noronha | Asian Indian | South Asian | Wong et al. |
| Norte | Filipino | Other Asian | Wong et al. |
| Nosa | Filipino | Other Asian | Wong et al. |
| Nosaka | Japanese | Other Asian | Wong et al. |
| Nose | Japanese | Other Asian | Wong et al. |
| Nostratis | Filipino | Other Asian | Wong et al. |
| Notario | Filipino | Other Asian | Wong et al. |
| Notarte | Filipino | Other Asian | Wong et al. |
| Notomi | Japanese | Other Asian | Wong et al. |
| Noval | Filipino | Other Asian | Wong et al. |
| Novales | Filipino | Other Asian | Wong et al. |
| Nove |  | South Asian | Lim et al. |
| Noveda | Filipino | Other Asian | Wong et al. |
| Novelo | Filipino | Other Asian | Wong et al. |
| Noveloso | Filipino | Other Asian | Wong et al. |
| Novenario | Filipino | Other Asian | Wong et al. |
| Novencido | Filipino | Other Asian | Wong et al. |
| Noveno | Filipino | Other Asian | Wong et al. |
| Novero | Filipino | Other Asian | Wong et al. |
| Novesteras | Filipino | Other Asian | Wong et al. |
| Novicio | Filipino | Other Asian | Wong et al. |
| Novida | Filipino | Other Asian | Wong et al. |
| Novilla | Filipino | Other Asian | Wong et al. |
| Novio | Filipino | Other Asian | Wong et al. |
| Nowab |  | South Asian | Lim et al. |
| Nozaki | Japanese | Other Asian | Wong et al. |
| Nozawa | Japanese | Other Asian | Wong et al. |
| Nozoe | Japanese | Other Asian | Wong et al. |
| Nozu | Japanese | Other Asian | Wong et al. |
| Nquyen | Vietnamese | Other Asian | Wong et al. |
| Nubla | Filipino | Other Asian | Wong et al. |
| Nucum | Filipino | Other Asian | Wong et al. |
| Nucup | Filipino | Other Asian | Wong et al. |
| Nuen | Chinese | Chinese | Wong et al. |
| Nuesa | Filipino | Other Asian | Wong et al. |
| Nuesca | Filipino | Other Asian | Wong et al. |
| Nuestro | Filipino | Other Asian | Wong et al. |
| Nueva | Filipino | Other Asian | Wong et al. |
| Nuevo | Filipino | Other Asian | Wong et al. |
| Nuez | Filipino | Other Asian | Wong et al. |
| Nuezca | Filipino | Other Asian | Wong et al. |
| Nufable | Filipino | Other Asian | Wong et al. |
| Nuguid | Filipino | Other Asian | Wong et al. |
| Nugyen | Vietnamese | Other Asian | Wong et al. |
| Nui | Chinese | Chinese | Wong et al. |
| Nukaya | Japanese | Other Asian | Wong et al. |
| Nukui | Japanese | Other Asian | Wong et al. |
| Nulud | Filipino | Other Asian | Wong et al. |
| Num | Chinese | Chinese | Wong et al. |
| Numa | Japanese | Other Asian | Wong et al. |
| Numajiri | Japanese | Other Asian | Wong et al. |
| Numakura | Japanese | Other Asian | Wong et al. |
| Numano | Japanese | Other Asian | Wong et al. |
| Numata | Japanese | Other Asian | Wong et al. |
| Numazaki | Japanese | Other Asian | Wong et al. |
| Nunag | Filipino | Other Asian | Wong et al. |
| Nunez | Filipino | Other Asian | Wong et al. |
| Nung | Chinese | Chinese | Wong et al. |
| Nunga | Filipino | Other Asian | Wong et al. |
| Nunokawa | Japanese | Other Asian | Wong et al. |
| Nunome | Japanese | Other Asian | Wong et al. |
| Nunotani | Japanese | Other Asian | Wong et al. |
| Nupur |  | South Asian | Lim et al. |
| Nuque | Filipino | Other Asian | Wong et al. |
| Nuqui | Filipino | Other Asian | Wong et al. |
| Nurnabi |  | South Asian | Lim et al. |
| Nuruddin |  | South Asian | Lim et al. |
| Nurulhoque |  | South Asian | Lim et al. |
| Nurunnabi |  | South Asian | Lim et al. |
| Nuruzzaman | Bangladeshi | South Asian | Kranick et al. |
| Nuval | Filipino | Other Asian | Wong et al. |
| Nuyda | Filipino | Other Asian | Wong et al. |
| Nuyen | Vietnamese | Other Asian | Wong et al. |
| Nuynh | Vietnamese | Other Asian | Wong et al. |
| Nyi | Chinese | Chinese | Wong et al. |
| O | Korean | Other Asian | Wong et al. |
| Oades | Filipino | Other Asian | Wong et al. |
| Oai | Vietnamese | Other Asian | Wong et al. |
| Oamil | Filipino | Other Asian | Wong et al. |
| Oana | Filipino | Other Asian | Wong et al. |
| Oandasan | Filipino | Other Asian | Wong et al. |
| Oanh | Vietnamese | Other Asian | Wong et al. |
| Oasay | Filipino | Other Asian | Wong et al. |
| Oba | Japanese | Other Asian | Wong et al. |
| Obado | Filipino | Other Asian | Wong et al. |
| Obaldo | Filipino | Other Asian | Wong et al. |
| Obana | Filipino | Other Asian | Wong et al. |
| Obando | Filipino | Other Asian | Wong et al. |
| Obara | Japanese | Other Asian | Wong et al. |
| Obata | Japanese | Other Asian | Wong et al. |
| Obayashi | Japanese | Other Asian | Wong et al. |
| Obcemea | Filipino | Other Asian | Wong et al. |
| Obedencio | Filipino | Other Asian | Wong et al. |
| Obedoza | Filipino | Other Asian | Wong et al. |
| Obena | Filipino | Other Asian | Wong et al. |
| Obenario | Filipino | Other Asian | Wong et al. |
| Obenza | Filipino | Other Asian | Wong et al. |
| Oberoi | Asian Indian | South Asian | Kranick et al. |
| Obi | Japanese | Other Asian | Wong et al. |
| Obial | Filipino | Other Asian | Wong et al. |
| Obias | Filipino | Other Asian | Wong et al. |
| Obien | Filipino | Other Asian | Wong et al. |
| Obillo | Filipino | Other Asian | Wong et al. |
| Obina | Filipino | Other Asian | Wong et al. |
| Obinata | Japanese | Other Asian | Wong et al. |
| Obispo | Filipino | Other Asian | Wong et al. |
| Oblea | Filipino | Other Asian | Wong et al. |
| Oblefias | Filipino | Other Asian | Wong et al. |
| Oblepias | Filipino | Other Asian | Wong et al. |
| Obligacion | Filipino | Other Asian | Wong et al. |
| Obligar | Filipino | Other Asian | Wong et al. |
| Obong | Filipino | Other Asian | Wong et al. |
| Oboza | Filipino | Other Asian | Wong et al. |
| Obra | Filipino | Other Asian | Wong et al. |
| Obregon | Filipino | Other Asian | Wong et al. |
| Obrero | Filipino | Other Asian | Wong et al. |
| Obsuna | Filipino | Other Asian | Wong et al. |
| Obtera | Filipino | Other Asian | Wong et al. |
| Obuchi | Japanese | Other Asian | Wong et al. |
| Obungen | Filipino | Other Asian | Wong et al. |
| Obusan | Filipino | Other Asian | Wong et al. |
| Obydullah |  | South Asian | Lim et al. |
| Oca | Filipino | Other Asian | Wong et al. |
| Ocampo | Filipino | Other Asian | Kranick et al. |
| Ocampos | Filipino | Other Asian | Wong et al. |
| Ocana | Filipino | Other Asian | Wong et al. |
| Ocariza | Filipino | Other Asian | Wong et al. |
| Ocasion | Filipino | Other Asian | Wong et al. |
| Ocava | Filipino | Other Asian | Wong et al. |
| Ocbina | Filipino | Other Asian | Wong et al. |
| Occena | Filipino | Other Asian | Wong et al. |
| Occeno | Filipino | Other Asian | Wong et al. |
| Ocenar | Filipino | Other Asian | Wong et al. |
| Ocera | Filipino | Other Asian | Wong et al. |
| Ocfemia | Filipino | Other Asian | Wong et al. |
| Ochangco | Filipino | Other Asian | Wong et al. |
| Ochave | Filipino | Other Asian | Wong et al. |
| Ochhavlal | Asian Indian | South Asian | Wong et al. |
| Ochi | Japanese | Other Asian | Wong et al. |
| Ochiai | Japanese | Other Asian | Wong et al. |
| Ochinang | Filipino | Other Asian | Wong et al. |
| Ochoa | Filipino | Other Asian | Wong et al. |
| Ochoco | Filipino | Other Asian | Wong et al. |
| Ochosa | Filipino | Other Asian | Wong et al. |
| Ochotorena | Filipino | Other Asian | Wong et al. |
| Ock | Chinese | Chinese | Wong et al. |
| Oclarino | Filipino | Other Asian | Wong et al. |
| Oco | Filipino | Other Asian | Wong et al. |
| Ocol | Filipino | Other Asian | Wong et al. |
| Ocoma | Filipino | Other Asian | Wong et al. |
| Ocomen | Filipino | Other Asian | Wong et al. |
| Ocon | Filipino | Other Asian | Wong et al. |
| Oconer | Filipino | Other Asian | Wong et al. |
| Ocreto | Filipino | Other Asian | Wong et al. |
| Ocsan | Filipino | Other Asian | Wong et al. |
| Octaviano | Filipino | Other Asian | Wong et al. |
| Octavio | Filipino | Other Asian | Wong et al. |
| Octavo | Filipino | Other Asian | Wong et al. |
| Oculto | Filipino | Other Asian | Wong et al. |
| Ocumen | Filipino | Other Asian | Wong et al. |
| Oczon | Filipino | Other Asian | Wong et al. |
| Oda | Japanese | Other Asian | Wong et al. |
| Odagiri | Japanese | Other Asian | Wong et al. |
| Odahara | Japanese | Other Asian | Wong et al. |
| Odaira | Japanese | Other Asian | Wong et al. |
| Odajima | Japanese | Other Asian | Wong et al. |
| Odaka | Japanese | Other Asian | Wong et al. |
| Odake | Japanese | Other Asian | Wong et al. |
| Odal | Filipino | Other Asian | Wong et al. |
| Odanaka | Japanese | Other Asian | Wong et al. |
| Odanga | Filipino | Other Asian | Wong et al. |
| Odani | Japanese | Other Asian | Wong et al. |
| Odashima | Japanese | Other Asian | Wong et al. |
| Odavar | Filipino | Other Asian | Wong et al. |
| Odawara | Japanese | Other Asian | Wong et al. |
| Odedra | Asian Indian | South Asian | Wong et al. |
| Odo | Japanese | Other Asian | Wong et al. |
| Odtohan | Filipino | Other Asian | Wong et al. |
| Oduca | Filipino | Other Asian | Wong et al. |
| Oducayen | Filipino | Other Asian | Wong et al. |
| Odulio | Filipino | Other Asian | Wong et al. |
| Odviar | Filipino | Other Asian | Wong et al. |
| Odvina | Filipino | Other Asian | Wong et al. |
| Oe | Japanese | Other Asian | Wong et al. |
| Ofiana | Filipino | Other Asian | Wong et al. |
| Ofilada | Filipino | Other Asian | Wong et al. |
| Ofina | Filipino | Other Asian | Wong et al. |
| Ofrancia | Filipino | Other Asian | Wong et al. |
| Ofrecio | Filipino | Other Asian | Wong et al. |
| Ofreneo | Filipino | Other Asian | Wong et al. |
| Ofuji | Japanese | Other Asian | Wong et al. |
| Oga | Japanese | Other Asian | Wong et al. |
| Ogahara | Japanese | Other Asian | Wong et al. |
| Ogai | Japanese | Other Asian | Wong et al. |
| Ogaki | Japanese | Other Asian | Wong et al. |
| Ogalino | Filipino | Other Asian | Wong et al. |
| Ogami | Japanese | Other Asian | Wong et al. |
| Ogana | Filipino | Other Asian | Wong et al. |
| Ogario | Filipino | Other Asian | Wong et al. |
| Ogasawara | Japanese | Other Asian | Wong et al. |
| Ogata | Japanese | Other Asian | Wong et al. |
| Ogawa | Japanese | Other Asian | Wong et al. |
| Ogawara | Japanese | Other Asian | Wong et al. |
| Ogaya | Filipino | Other Asian | Wong et al. |
| Ogi | Japanese | Other Asian | Wong et al. |
| Ogihara | Japanese | Other Asian | Wong et al. |
| Ogino | Japanese | Other Asian | Wong et al. |
| Ogishima | Japanese | Other Asian | Wong et al. |
| Ogiso | Japanese | Other Asian | Wong et al. |
| Ogita | Japanese | Other Asian | Wong et al. |
| Ogiwara | Japanese | Other Asian | Wong et al. |
| Ogo | Japanese | Other Asian | Wong et al. |
| Ogoshi | Japanese | Other Asian | Wong et al. |
| Ogoy | Filipino | Other Asian | Wong et al. |
| Ogtong | Filipino | Other Asian | Wong et al. |
| Oguchi | Japanese | Other Asian | Wong et al. |
| Oguma | Japanese | Other Asian | Wong et al. |
| Ogura | Japanese | Other Asian | Wong et al. |
| Oguri | Japanese | Other Asian | Wong et al. |
| Oguro | Japanese | Other Asian | Wong et al. |
| Ogushi | Japanese | Other Asian | Wong et al. |
| Oh | Korean | Other Asian | Kranick et al. |
| Ohama | Japanese | Other Asian | Wong et al. |
| Ohashi | Japanese | Other Asian | Wong et al. |
| Ohata | Japanese | Other Asian | Wong et al. |
| Ohba | Japanese | Other Asian | Wong et al. |
| Ohga | Japanese | Other Asian | Wong et al. |
| Ohgi | Japanese | Other Asian | Wong et al. |
| Ohhashi | Japanese | Other Asian | Wong et al. |
| Ohi | Japanese | Other Asian | Wong et al. |
| Ohid | Bangladeshi; Asian Indian | South Asian | Kranick et al. |
| Ohigashi | Japanese | Other Asian | Wong et al. |
| Ohinata | Japanese | Other Asian | Wong et al. |
| Ohira | Japanese | Other Asian | Wong et al. |
| Ohishi | Japanese | Other Asian | Wong et al. |
| Ohkawa | Japanese | Other Asian | Wong et al. |
| Ohkawara | Japanese | Other Asian | Wong et al. |
| Ohki | Japanese | Other Asian | Wong et al. |
| Ohkubo | Japanese | Other Asian | Wong et al. |
| Ohkuma | Japanese | Other Asian | Wong et al. |
| Ohkura | Japanese | Other Asian | Wong et al. |
| Ohmi | Japanese | Other Asian | Wong et al. |
| Ohmiya | Japanese | Other Asian | Wong et al. |
| Ohmori | Japanese | Other Asian | Wong et al. |
| Ohmoto | Japanese | Other Asian | Wong et al. |
| Ohmura | Japanese | Other Asian | Wong et al. |
| Ohnishi | Japanese | Other Asian | Wong et al. |
| Ohno | Japanese | Other Asian | Wong et al. |
| Ohnuki | Japanese | Other Asian | Wong et al. |
| Ohnuma | Japanese | Other Asian | Wong et al. |
| Oho | Japanese | Other Asian | Wong et al. |
| Ohori | Japanese | Other Asian | Wong et al. |
| Ohri | Asian Indian | South Asian | Wong et al. |
| Ohsaki | Japanese | Other Asian | Wong et al. |
| Ohsato | Japanese | Other Asian | Wong et al. |
| Ohsawa | Japanese | Other Asian | Wong et al. |
| Ohshima | Japanese | Other Asian | Wong et al. |
| Ohshiro | Japanese | Other Asian | Wong et al. |
| Ohta | Japanese | Other Asian | Wong et al. |
| Ohtake | Japanese | Other Asian | Wong et al. |
| Ohtaki | Japanese | Other Asian | Wong et al. |
| Ohtani | Japanese | Other Asian | Wong et al. |
| Ohto | Japanese | Other Asian | Wong et al. |
| Ohtomo | Japanese | Other Asian | Wong et al. |
| Ohtsu | Japanese | Other Asian | Wong et al. |
| Ohtsubo | Japanese | Other Asian | Wong et al. |
| Ohtsuka | Japanese | Other Asian | Wong et al. |
| Ohtsuki | Japanese | Other Asian | Wong et al. |
| Ohuchi | Japanese | Other Asian | Wong et al. |
| Ohwada | Japanese | Other Asian | Wong et al. |
| Ohya | Japanese | Other Asian | Wong et al. |
| Ohyama | Japanese | Other Asian | Wong et al. |
| Oi | Chinese | Chinese | Wong et al. |
| Oida | Filipino | Other Asian | Wong et al. |
| Oikawa | Japanese | Other Asian | Wong et al. |
| Oike | Japanese | Other Asian | Wong et al. |
| Oishi | Japanese | Other Asian | Wong et al. |
| Oiwa | Japanese | Other Asian | Wong et al. |
| Oizumi | Japanese | Other Asian | Wong et al. |
| Ojano | Filipino | Other Asian | Wong et al. |
| Ojastro | Filipino | Other Asian | Wong et al. |
| Ojeda | Filipino | Other Asian | Wong et al. |
| Ojha | Asian Indian | South Asian | Wong et al. |
| Oji | Japanese | Other Asian | Wong et al. |
| Ojima | Japanese | Other Asian | Wong et al. |
| Ok | Korean | Other Asian | Wong et al. |
| Oka | Japanese | Other Asian | Wong et al. |
| Okabayashi | Japanese | Other Asian | Wong et al. |
| Okabe | Japanese | Other Asian | Wong et al. |
| Okada | Japanese | Other Asian | Wong et al. |
| Okado | Japanese | Other Asian | Wong et al. |
| Okahara | Japanese | Other Asian | Wong et al. |
| Okai | Japanese | Other Asian | Wong et al. |
| Okajima | Japanese | Other Asian | Wong et al. |
| Okamatsu | Japanese | Other Asian | Wong et al. |
| Okami | Japanese | Other Asian | Wong et al. |
| Okamoto | Japanese | Other Asian | Wong et al. |
| Okamura | Japanese | Other Asian | Wong et al. |
| Okaniwa | Japanese | Other Asian | Wong et al. |
| Okano | Japanese | Other Asian | Wong et al. |
| Okashita | Japanese | Other Asian | Wong et al. |
| Okatani | Japanese | Other Asian | Wong et al. |
| Okawa | Japanese | Other Asian | Wong et al. |
| Okawara | Japanese | Other Asian | Wong et al. |
| Okaya | Japanese | Other Asian | Wong et al. |
| Okayama | Japanese | Other Asian | Wong et al. |
| Okazaki | Japanese | Other Asian | Wong et al. |
| Okazawa | Japanese | Other Asian | Wong et al. |
| Oki | Japanese | Other Asian | Wong et al. |
| Okihara | Japanese | Other Asian | Wong et al. |
| Okimoto | Japanese | Other Asian | Wong et al. |
| Okimura | Japanese | Other Asian | Wong et al. |
| Okinaga | Japanese | Other Asian | Wong et al. |
| Okinaka | Japanese | Other Asian | Wong et al. |
| Okino | Japanese | Other Asian | Wong et al. |
| Okita | Japanese | Other Asian | Wong et al. |
| Okitsu | Japanese | Other Asian | Wong et al. |
| Okiyama | Japanese | Other Asian | Wong et al. |
| Okochi | Japanese | Other Asian | Wong et al. |
| Okoshi | Japanese | Other Asian | Wong et al. |
| Oku | Japanese | Other Asian | Wong et al. |
| Okubo | Japanese | Other Asian | Wong et al. |
| Okuda | Japanese | Other Asian | Wong et al. |
| Okudaira | Japanese | Other Asian | Wong et al. |
| Okuhara | Japanese | Other Asian | Wong et al. |
| Okui | Japanese | Other Asian | Wong et al. |
| Okuma | Japanese | Other Asian | Wong et al. |
| Okumoto | Japanese | Other Asian | Wong et al. |
| Okumura | Japanese | Other Asian | Wong et al. |
| Okuni | Japanese | Other Asian | Wong et al. |
| Okuno | Japanese | Other Asian | Wong et al. |
| Okura | Japanese | Other Asian | Wong et al. |
| Okutani | Japanese | Other Asian | Wong et al. |
| Okutsu | Japanese | Other Asian | Wong et al. |
| Okuyama | Japanese | Other Asian | Wong et al. |
| Okuzawa | Japanese | Other Asian | Wong et al. |
| Ola | Filipino | Other Asian | Wong et al. |
| Olaer | Filipino | Other Asian | Wong et al. |
| Olaes | Filipino | Other Asian | Wong et al. |
| Olaguer | Filipino | Other Asian | Wong et al. |
| Olais | Filipino | Other Asian | Wong et al. |
| Olalia | Filipino | Other Asian | Wong et al. |
| Olan | Filipino | Other Asian | Wong et al. |
| Olanday | Filipino | Other Asian | Wong et al. |
| Olano | Filipino | Other Asian | Wong et al. |
| Olario | Filipino | Other Asian | Wong et al. |
| Olarte | Filipino | Other Asian | Wong et al. |
| Olaso | Filipino | Other Asian | Wong et al. |
| Olavario | Filipino | Other Asian | Wong et al. |
| Olavere | Filipino | Other Asian | Wong et al. |
| Olaya | Filipino | Other Asian | Wong et al. |
| Olayvar | Filipino | Other Asian | Wong et al. |
| Olazo | Filipino | Other Asian | Wong et al. |
| Olbes | Filipino | Other Asian | Wong et al. |
| Olea | Filipino | Other Asian | Wong et al. |
| Oledan | Filipino | Other Asian | Wong et al. |
| Olegario | Filipino | Other Asian | Wong et al. |
| Oleta | Filipino | Other Asian | Wong et al. |
| Olfato | Filipino | Other Asian | Wong et al. |
| Olfindo | Filipino | Other Asian | Wong et al. |
| Oli |  | South Asian | Lim et al. |
| Olid | Filipino | Other Asian | Wong et al. |
| Olidan | Filipino | Other Asian | Wong et al. |
| Oligario | Filipino | Other Asian | Wong et al. |
| Oligo | Filipino | Other Asian | Wong et al. |
| Olila | Filipino | Other Asian | Wong et al. |
| Olimba | Filipino | Other Asian | Wong et al. |
| Olimpo | Filipino | Other Asian | Wong et al. |
| Olinares | Filipino | Other Asian | Wong et al. |
| Olipares | Filipino | Other Asian | Wong et al. |
| Olis | Filipino | Other Asian | Wong et al. |
| Olita | Filipino | Other Asian | Wong et al. |
| Olitan | Filipino | Other Asian | Wong et al. |
| Oliullah |  | South Asian | Lim et al. |
| Oliva | Filipino | Other Asian | Wong et al. |
| Olivar | Filipino | Other Asian | Wong et al. |
| Olivares | Filipino | Other Asian | Wong et al. |
| Olivarez | Filipino | Other Asian | Wong et al. |
| Olivas | Filipino | Other Asian | Wong et al. |
| Olivera | Filipino | Other Asian | Wong et al. |
| Oliveria | Filipino | Other Asian | Wong et al. |
| Oliveros | Filipino | Other Asian | Wong et al. |
| Olivo | Filipino | Other Asian | Wong et al. |
| Ollano | Filipino | Other Asian | Wong et al. |
| Ollero | Filipino | Other Asian | Wong et al. |
| Olmedo | Filipino | Other Asian | Wong et al. |
| Olmo | Filipino | Other Asian | Wong et al. |
| Olmos | Filipino | Other Asian | Wong et al. |
| Olonan | Filipino | Other Asian | Wong et al. |
| Olpindo | Filipino | Other Asian | Wong et al. |
| Olympia | Filipino | Other Asian | Wong et al. |
| Om | Korean | Other Asian | Wong et al. |
| Omachi | Japanese | Other Asian | Wong et al. |
| Omae | Japanese | Other Asian | Wong et al. |
| Omagari | Japanese | Other Asian | Wong et al. |
| Omana | Filipino | Other Asian | Wong et al. |
| Omandam | Filipino | Other Asian | Wong et al. |
| Omar | Malaysian; Bangladeshi | Other Asian | Kranick et al. |
| Omata | Japanese | Other Asian | Wong et al. |
| Ombac | Filipino | Other Asian | Wong et al. |
| Ombao | Filipino | Other Asian | Wong et al. |
| Omed |  | South Asian | Lim et al. |
| Omega | Filipino | Other Asian | Wong et al. |
| Omi | Japanese | Other Asian | Wong et al. |
| Omichi | Japanese | Other Asian | Wong et al. |
| Omine | Japanese | Other Asian | Wong et al. |
| Omitted | Chinese | Chinese | Wong et al. |
| Omiya | Japanese | Other Asian | Wong et al. |
| Omnes | Filipino | Other Asian | Wong et al. |
| Omo | Filipino | Other Asian | Wong et al. |
| Omori | Japanese | Other Asian | Wong et al. |
| Omote | Japanese | Other Asian | Wong et al. |
| Omoto | Japanese | Other Asian | Wong et al. |
| Ompad | Filipino | Other Asian | Wong et al. |
| Omura | Japanese | Other Asian | Wong et al. |
| On | Chinese | Chinese | Wong et al. |
| Ona | Filipino | Other Asian | Wong et al. |
| Onaga | Japanese | Other Asian | Wong et al. |
| Onaka | Japanese | Other Asian | Wong et al. |
| Onas | Filipino | Other Asian | Wong et al. |
| Onate | Filipino | Other Asian | Wong et al. |
| Ondoy | Filipino | Other Asian | Wong et al. |
| Ong | Chinese; Malaysian; Singapore | Chinese | Kranick et al. |
| Ongaco | Filipino | Other Asian | Wong et al. |
| Ongchangco | Filipino | Other Asian | Wong et al. |
| Ongcoy | Filipino | Other Asian | Wong et al. |
| Ongjoco | Filipino | Other Asian | Wong et al. |
| Ongkeko | Filipino | Other Asian | Wong et al. |
| Ongkiko | Filipino | Other Asian | Wong et al. |
| Ongkingco | Filipino | Other Asian | Wong et al. |
| Ongoco | Filipino | Other Asian | Wong et al. |
| Ongos | Filipino | Other Asian | Wong et al. |
| Ongpin | Filipino | Other Asian | Wong et al. |
| Ongsiako | Filipino | Other Asian | Wong et al. |
| Ongsioco | Filipino | Other Asian | Wong et al. |
| Ongtangco | Filipino | Other Asian | Wong et al. |
| Ongtengco | Filipino | Other Asian | Wong et al. |
| Ongveloso | Filipino | Other Asian | Wong et al. |
| Onia | Filipino | Other Asian | Wong et al. |
| Oniki | Japanese | Other Asian | Wong et al. |
| Onishi | Japanese | Other Asian | Kranick et al. |
| Onitsuka | Japanese | Other Asian | Wong et al. |
| Onizuka | Japanese | Other Asian | Wong et al. |
| Onn | Chinese | Chinese | Wong et al. |
| Onnagan | Filipino | Other Asian | Wong et al. |
| Ono | Japanese | Other Asian | Wong et al. |
| Onoda | Japanese | Other Asian | Wong et al. |
| Onodera | Japanese | Other Asian | Wong et al. |
| Onoe | Japanese | Other Asian | Wong et al. |
| Onogi | Japanese | Other Asian | Wong et al. |
| Onose | Japanese | Other Asian | Wong et al. |
| Onoue | Japanese | Other Asian | Wong et al. |
| Onoyama | Japanese | Other Asian | Wong et al. |
| Onozato | Japanese | Other Asian | Wong et al. |
| Onozawa | Japanese | Other Asian | Wong et al. |
| Onozuka | Japanese | Other Asian | Wong et al. |
| Onrubia | Filipino | Other Asian | Wong et al. |
| Ontoy | Filipino | Other Asian | Wong et al. |
| Onuki | Japanese | Other Asian | Wong et al. |
| Onuma | Japanese | Other Asian | Wong et al. |
| Onza | Filipino | Other Asian | Wong et al. |
| Ooka | Japanese | Other Asian | Wong et al. |
| Oomer | Asian Indian | South Asian | Wong et al. |
| Oommen | Asian Indian | South Asian | Wong et al. |
| Oong | Chinese | Chinese | Wong et al. |
| Oosman |  | South Asian | Lim et al. |
| Oota | Japanese | Other Asian | Wong et al. |
| Ootaki | Japanese | Other Asian | Wong et al. |
| Opalec | Filipino | Other Asian | Wong et al. |
| Opena | Filipino | Other Asian | Wong et al. |
| Opendo | Filipino | Other Asian | Wong et al. |
| Operana | Filipino | Other Asian | Wong et al. |
| Operario | Filipino | Other Asian | Wong et al. |
| Opiana | Filipino | Other Asian | Wong et al. |
| Opilas | Filipino | Other Asian | Wong et al. |
| Opina | Filipino | Other Asian | Wong et al. |
| Opinaldo | Filipino | Other Asian | Wong et al. |
| Opiniano | Filipino | Other Asian | Wong et al. |
| Opinion | Filipino | Other Asian | Wong et al. |
| Ople | Filipino | Other Asian | Wong et al. |
| Oppus | Filipino | Other Asian | Wong et al. |
| Opulencia | Filipino | Other Asian | Wong et al. |
| Oquendo | Filipino | Other Asian | Wong et al. |
| Oquias | Filipino | Other Asian | Wong et al. |
| Oquindo | Filipino | Other Asian | Wong et al. |
| Or | Chinese | Chinese | Wong et al. |
| Ora | Filipino | Other Asian | Wong et al. |
| Oracion | Filipino | Other Asian | Wong et al. |
| Orais | Filipino | Other Asian | Wong et al. |
| Orallo | Filipino | Other Asian | Wong et al. |
| Orante | Filipino | Other Asian | Wong et al. |
| Orap | Filipino | Other Asian | Wong et al. |
| Orara | Filipino | Other Asian | Wong et al. |
| Orata | Filipino | Other Asian | Wong et al. |
| Orate | Filipino | Other Asian | Wong et al. |
| Orbase | Filipino | Other Asian | Wong et al. |
| Orbe | Filipino | Other Asian | Wong et al. |
| Orbeta | Filipino | Other Asian | Kranick et al. |
| Orbita | Filipino | Other Asian | Wong et al. |
| Orca | Filipino | Other Asian | Wong et al. |
| Orcales | Filipino | Other Asian | Wong et al. |
| Orcine | Filipino | Other Asian | Wong et al. |
| Orcino | Filipino | Other Asian | Wong et al. |
| Orcullo | Filipino | Other Asian | Wong et al. |
| Orda | Filipino | Other Asian | Wong et al. |
| Ordanes | Filipino | Other Asian | Wong et al. |
| Ordanez | Filipino | Other Asian | Wong et al. |
| Ordaniel | Filipino | Other Asian | Wong et al. |
| Ordanza | Filipino | Other Asian | Wong et al. |
| Ordas | Filipino | Other Asian | Wong et al. |
| Orden | Filipino | Other Asian | Wong et al. |
| Ordiales | Filipino | Other Asian | Wong et al. |
| Ordillas | Filipino | Other Asian | Wong et al. |
| Ordinario | Filipino | Other Asian | Wong et al. |
| Ordiz | Filipino | Other Asian | Wong et al. |
| Ordona | Filipino | Other Asian | Kranick et al. |
| Ordonez | Filipino | Other Asian | Wong et al. |
| Ordonia | Filipino | Other Asian | Wong et al. |
| Ordonio | Filipino | Other Asian | Wong et al. |
| Ordono | Filipino | Other Asian | Wong et al. |
| Ordoveza | Filipino | Other Asian | Wong et al. |
| Orduna | Filipino | Other Asian | Wong et al. |
| Oreiro | Filipino | Other Asian | Wong et al. |
| Orejola | Filipino | Other Asian | Wong et al. |
| Orejudos | Filipino | Other Asian | Wong et al. |
| Orellana | Chinese | Chinese | Wong et al. |
| Orencia | Filipino | Other Asian | Wong et al. |
| Orendain | Filipino | Other Asian | Wong et al. |
| Orense | Filipino | Other Asian | Wong et al. |
| Oreta | Filipino | Other Asian | Wong et al. |
| Orfiano | Filipino | Other Asian | Wong et al. |
| Organo | Filipino | Other Asian | Wong et al. |
| Oria | Filipino | Other Asian | Wong et al. |
| Orial | Filipino | Other Asian | Wong et al. |
| Orias | Filipino | Other Asian | Wong et al. |
| Oribello | Filipino | Other Asian | Wong et al. |
| Oriel | Filipino | Other Asian | Wong et al. |
| Orig | Filipino | Other Asian | Wong et al. |
| Origenes | Filipino | Other Asian | Wong et al. |
| Orihara | Japanese | Other Asian | Wong et al. |
| Orii | Japanese | Other Asian | Wong et al. |
| Orikasa | Japanese | Other Asian | Wong et al. |
| Orilla | Filipino | Other Asian | Wong et al. |
| Orillaza | Filipino | Other Asian | Wong et al. |
| Orille | Filipino | Other Asian | Wong et al. |
| Orillo | Filipino | Other Asian | Wong et al. |
| Orimoto | Japanese | Other Asian | Wong et al. |
| Orina | Filipino | Other Asian | Wong et al. |
| Orines | Filipino | Other Asian | Wong et al. |
| Orinion | Filipino | Other Asian | Wong et al. |
| Orio | Filipino | Other Asian | Wong et al. |
| Orita | Japanese | Other Asian | Wong et al. |
| Orjalo | Filipino | Other Asian | Wong et al. |
| Orlanda | Filipino | Other Asian | Wong et al. |
| Orlanes | Filipino | Other Asian | Wong et al. |
| Orlina | Filipino | Other Asian | Wong et al. |
| Orlino | Filipino | Other Asian | Wong et al. |
| Ormeo | Filipino | Other Asian | Wong et al. |
| Ormita | Filipino | Other Asian | Wong et al. |
| Ornedo | Filipino | Other Asian | Wong et al. |
| Ornopia | Filipino | Other Asian | Wong et al. |
| Ornum | Filipino | Other Asian | Wong et al. |
| Oro | Filipino | Other Asian | Wong et al. |
| Orobia | Filipino | Other Asian | Wong et al. |
| Oroc | Filipino | Other Asian | Wong et al. |
| Orodio | Filipino | Other Asian | Wong et al. |
| Orogo | Filipino | Other Asian | Wong et al. |
| Orola | Filipino | Other Asian | Wong et al. |
| Orolfo | Filipino | Other Asian | Wong et al. |
| Oronce | Filipino | Other Asian | Wong et al. |
| Oronos | Filipino | Other Asian | Wong et al. |
| Oropesa | Filipino | Other Asian | Wong et al. |
| Oropilla | Filipino | Other Asian | Wong et al. |
| Orosa | Filipino | Other Asian | Wong et al. |
| Orosco | Filipino | Other Asian | Wong et al. |
| Orozco | Filipino | Other Asian | Wong et al. |
| Orpia | Filipino | Other Asian | Wong et al. |
| Orpiano | Filipino | Other Asian | Wong et al. |
| Orpilla | Filipino | Other Asian | Wong et al. |
| Orprecio | Filipino | Other Asian | Wong et al. |
| Orque | Filipino | Other Asian | Wong et al. |
| Orquiola | Filipino | Other Asian | Wong et al. |
| Orquiza | Filipino | Other Asian | Wong et al. |
| Ortal | Filipino | Other Asian | Wong et al. |
| Ortaleza | Filipino | Other Asian | Wong et al. |
| Ortaliz | Filipino | Other Asian | Wong et al. |
| Ortaliza | Filipino | Other Asian | Wong et al. |
| Ortanez | Filipino | Other Asian | Wong et al. |
| Ortega | Filipino | Other Asian | Wong et al. |
| Ortencio | Filipino | Other Asian | Wong et al. |
| Ortez | Filipino | Other Asian | Wong et al. |
| Orteza | Filipino | Other Asian | Wong et al. |
| Ortigas | Filipino | Other Asian | Wong et al. |
| Ortigoza | Filipino | Other Asian | Wong et al. |
| Ortiguero | Filipino | Other Asian | Wong et al. |
| Ortiguerra | Filipino | Other Asian | Wong et al. |
| Ortilla | Filipino | Other Asian | Wong et al. |
| Ortillo | Filipino | Other Asian | Wong et al. |
| Ortin | Filipino | Other Asian | Wong et al. |
| Ortiz | Filipino | Other Asian | Wong et al. |
| Ortizluis | Filipino | Other Asian | Wong et al. |
| Ortizo | Filipino | Other Asian | Wong et al. |
| Ortonio | Filipino | Other Asian | Wong et al. |
| Ortua | Filipino | Other Asian | Wong et al. |
| Orzame | Filipino | Other Asian | Wong et al. |
| Osa | Japanese | Other Asian | Wong et al. |
| Osabe | Japanese | Other Asian | Wong et al. |
| Osada | Japanese | Other Asian | Wong et al. |
| Osafune | Japanese | Other Asian | Wong et al. |
| Osaka | Japanese | Other Asian | Wong et al. |
| Osakabe | Japanese | Other Asian | Wong et al. |
| Osaki | Japanese | Other Asian | Wong et al. |
| Osako | Japanese | Other Asian | Wong et al. |
| Osana | Filipino | Other Asian | Wong et al. |
| Osanai | Japanese | Other Asian | Wong et al. |
| Osano | Filipino | Other Asian | Wong et al. |
| Osato | Japanese | Other Asian | Wong et al. |
| Osawa | Japanese | Other Asian | Wong et al. |
| Osea | Filipino | Other Asian | Wong et al. |
| Oshida | Japanese | Other Asian | Wong et al. |
| Oshige | Japanese | Other Asian | Wong et al. |
| Oshika | Japanese | Other Asian | Wong et al. |
| Oshikawa | Japanese | Other Asian | Wong et al. |
| Oshima | Japanese | Other Asian | Wong et al. |
| Oshimo | Japanese | Other Asian | Wong et al. |
| Oshino | Japanese | Other Asian | Wong et al. |
| Oshio | Japanese | Other Asian | Wong et al. |
| Oshiro | Japanese | Other Asian | Wong et al. |
| Oshita | Japanese | Other Asian | Wong et al. |
| Oshiyama | Japanese | Other Asian | Wong et al. |
| Osias | Filipino | Other Asian | Wong et al. |
| Osilla | Filipino | Other Asian | Wong et al. |
| Osio | Filipino | Other Asian | Wong et al. |
| Osit | Filipino | Other Asian | Wong et al. |
| Osman |  | South Asian | Lim et al. |
| Osmani | Asian Indian | South Asian | Wong et al. |
| Osmena | Filipino | Other Asian | Wong et al. |
| Osorio | Filipino | Other Asian | Wong et al. |
| Osoteo | Filipino | Other Asian | Wong et al. |
| Ossorio | Filipino | Other Asian | Wong et al. |
| Ostil | Filipino | Other Asian | Wong et al. |
| Ostowari | Asian Indian | South Asian | Wong et al. |
| Ostrea | Filipino | Other Asian | Wong et al. |
| Osuga | Japanese | Other Asian | Wong et al. |
| Osugi | Japanese | Other Asian | Wong et al. |
| Osumi | Japanese | Other Asian | Wong et al. |
| Ota | Japanese | Other Asian | Wong et al. |
| Otabe | Japanese | Other Asian | Wong et al. |
| Otaka | Japanese | Other Asian | Wong et al. |
| Otake | Japanese | Other Asian | Wong et al. |
| Otaki | Japanese | Other Asian | Wong et al. |
| Otanes | Filipino | Other Asian | Wong et al. |
| Otani | Japanese | Other Asian | Wong et al. |
| Otero | Filipino | Other Asian | Wong et al. |
| Oteyza | Filipino | Other Asian | Wong et al. |
| Oto | Japanese | Other Asian | Wong et al. |
| Otomo | Japanese | Other Asian | Wong et al. |
| Otonari | Japanese | Other Asian | Wong et al. |
| Otoshi | Japanese | Other Asian | Wong et al. |
| Otsu | Japanese | Other Asian | Wong et al. |
| Otsubo | Japanese | Other Asian | Wong et al. |
| Otsuji | Japanese | Other Asian | Wong et al. |
| Otsuka | Japanese | Other Asian | Wong et al. |
| Otsuki | Japanese | Other Asian | Wong et al. |
| Ou | Chinese | Chinese | Kranick et al. |
| Ouan | Chinese | Chinese | Wong et al. |
| Ouano | Filipino | Other Asian | Wong et al. |
| Ouchi | Japanese | Other Asian | Wong et al. |
| Ouchida | Japanese | Other Asian | Wong et al. |
| Oue | Japanese | Other Asian | Wong et al. |
| Ouei | Chinese | Chinese | Wong et al. |
| Ouh | Korean | Other Asian | Wong et al. |
| Ouong | Chinese | Chinese | Wong et al. |
| Oura | Japanese | Other Asian | Wong et al. |
| Ousaka | Japanese | Other Asian | Kranick et al. |
| Ouseph | Asian Indian | South Asian | Wong et al. |
| Ouyang | Chinese | Chinese | Kranick et al. |
| Ouye | Japanese | Other Asian | Wong et al. |
| Ouyoung | Chinese | Chinese | Wong et al. |
| Ovalles | Filipino | Other Asian | Wong et al. |
| Oviedo | Filipino | Other Asian | Wong et al. |
| Ow | Chinese | Chinese | Wong et al. |
| Owa | Japanese | Other Asian | Wong et al. |
| Owada | Japanese | Other Asian | Wong et al. |
| Owan | Japanese | Other Asian | Wong et al. |
| Own | Chinese | Chinese | Wong et al. |
| Owyang | Chinese | Chinese | Wong et al. |
| Owyoung | Chinese | Chinese | Wong et al. |
| Oxales | Filipino | Other Asian | Wong et al. |
| Oy | Chinese | Chinese | Wong et al. |
| Oya | Japanese | Other Asian | Wong et al. |
| Oyabu | Japanese | Other Asian | Wong et al. |
| Oyadomari | Japanese | Other Asian | Wong et al. |
| Oyakawa | Japanese | Other Asian | Wong et al. |
| Oyama | Japanese | Other Asian | Wong et al. |
| Oyamada | Japanese | Other Asian | Wong et al. |
| Oyanagi | Japanese | Other Asian | Wong et al. |
| Oyang | Chinese | Chinese | Wong et al. |
| Oye | Japanese | Other Asian | Wong et al. |
| Oyoung | Chinese | Chinese | Wong et al. |
| Oyzon | Filipino | Other Asian | Wong et al. |
| Oza | Asian Indian | South Asian | Wong et al. |
| Ozaeta | Filipino | Other Asian | Wong et al. |
| Ozaki | Japanese | Other Asian | Wong et al. |
| Ozaraga | Filipino | Other Asian | Wong et al. |
| Ozasa | Japanese | Other Asian | Wong et al. |
| Ozawa | Japanese | Other Asian | Wong et al. |
| Ozeki | Japanese | Other Asian | Wong et al. |
| Ozoa | Filipino | Other Asian | Wong et al. |
| Ozono | Japanese | Other Asian | Wong et al. |
| Ozorio | Chinese | Chinese | Wong et al. |
| Ozu | Japanese | Other Asian | Wong et al. |
| Pa | Chinese | Chinese | Wong et al. |
| Paa | Filipino | Other Asian | Wong et al. |
| Paak | Chinese | Chinese | Wong et al. |
| Paas | Filipino | Other Asian | Wong et al. |
| Paat | Filipino | Other Asian | Wong et al. |
| Pabalan | Filipino | Other Asian | Wong et al. |
| Pabalate | Filipino | Other Asian | Wong et al. |
| Pabbi | Asian Indian | South Asian | Wong et al. |
| Pabelico | Filipino | Other Asian | Wong et al. |
| Pabellano | Filipino | Other Asian | Wong et al. |
| Pabello | Filipino | Other Asian | Wong et al. |
| Pabellon | Filipino | Other Asian | Wong et al. |
| Pabia | Filipino | Other Asian | Wong et al. |
| Pabilona | Filipino | Other Asian | Wong et al. |
| Pabilonia | Filipino | Other Asian | Wong et al. |
| Pabinguit | Filipino | Other Asian | Wong et al. |
| Pabiona | Filipino | Other Asian | Wong et al. |
| Pabla | Asian Indian | South Asian | Wong et al. |
| Pableo | Filipino | Other Asian | Wong et al. |
| Pablico | Filipino | Other Asian | Wong et al. |
| Pablo | Filipino | Other Asian | Wong et al. |
| Pabo | Filipino | Other Asian | Wong et al. |
| Pabon | Filipino | Other Asian | Wong et al. |
| Pabro | Filipino | Other Asian | Wong et al. |
| Pabros | Filipino | Other Asian | Wong et al. |
| Pabustan | Filipino | Other Asian | Wong et al. |
| Paca | Filipino | Other Asian | Wong et al. |
| Pacada | Filipino | Other Asian | Wong et al. |
| Pacaldo | Filipino | Other Asian | Wong et al. |
| Pacana | Filipino | Other Asian | Wong et al. |
| Pacardo | Filipino | Other Asian | Wong et al. |
| Pacariem | Filipino | Other Asian | Wong et al. |
| Pacete | Filipino | Other Asian | Wong et al. |
| Pacheco | Filipino | Other Asian | Wong et al. |
| Pachikara | Asian Indian | South Asian | Wong et al. |
| Pacho | Filipino | Other Asian | Wong et al. |
| Pacia | Filipino | Other Asian | Wong et al. |
| Paciente | Filipino | Other Asian | Wong et al. |
| Pacificar | Filipino | Other Asian | Wong et al. |
| Pacio | Filipino | Other Asian | Wong et al. |
| Pacis | Filipino | Other Asian | Wong et al. |
| Pack | Korean | Other Asian | Kranick et al. |
| Packing | Filipino | Other Asian | Wong et al. |
| Pacleb | Filipino | Other Asian | Wong et al. |
| Paclebar | Filipino | Other Asian | Wong et al. |
| Paclibar | Filipino | Other Asian | Wong et al. |
| Paco | Filipino | Other Asian | Wong et al. |
| Pacol | Filipino | Other Asian | Wong et al. |
| Pacot | Filipino | Other Asian | Wong et al. |
| Pacpaco | Filipino | Other Asian | Wong et al. |
| Pacquing | Filipino | Other Asian | Wong et al. |
| Pacris | Filipino | Other Asian | Wong et al. |
| Pacubas | Filipino | Other Asian | Wong et al. |
| Paculan | Filipino | Other Asian | Wong et al. |
| Pacumio | Filipino | Other Asian | Wong et al. |
| Pacunayen | Filipino | Other Asian | Wong et al. |
| Pacupac | Filipino | Other Asian | Wong et al. |
| Pada | Filipino | Other Asian | Wong et al. |
| Padaca | Filipino | Other Asian | Wong et al. |
| Padagas | Filipino | Other Asian | Wong et al. |
| Padam | Asian Indian | South Asian | Wong et al. |
| Padama | Filipino | Other Asian | Wong et al. |
| Padaoan | Filipino | Other Asian | Wong et al. |
| Padaong | Filipino | Other Asian | Wong et al. |
| Padasas | Filipino | Other Asian | Wong et al. |
| Padasdao | Filipino | Other Asian | Wong et al. |
| Padayao | Filipino | Other Asian | Wong et al. |
| Padayhag | Filipino | Other Asian | Wong et al. |
| Padda | Asian Indian | South Asian | Wong et al. |
| Pader | Filipino | Other Asian | Wong et al. |
| Paderanga | Filipino | Other Asian | Wong et al. |
| Paderes | Filipino | Other Asian | Wong et al. |
| Paderna | Filipino | Other Asian | Wong et al. |
| Padernal | Filipino | Other Asian | Wong et al. |
| Padero | Filipino | Other Asian | Wong et al. |
| Padhiar | Asian Indian | South Asian | Wong et al. |
| Padhye | Asian Indian | South Asian | Wong et al. |
| Padia | Asian Indian | South Asian | Wong et al. |
| Padiernos | Filipino | Other Asian | Wong et al. |
| Padigos | Filipino | Other Asian | Wong et al. |
| Padilla | Filipino | Other Asian | Kranick et al. |
| Padillo | Filipino | Other Asian | Wong et al. |
| Padillon | Filipino | Other Asian | Wong et al. |
| Padin | Filipino | Other Asian | Wong et al. |
| Padlan | Filipino | Other Asian | Wong et al. |
| Padmanabha | Asian Indian | South Asian | Wong et al. |
| Padmanabhan | Asian Indian | South Asian | Wong et al. |
| Padolina | Filipino | Other Asian | Wong et al. |
| Pador | Filipino | Other Asian | Wong et al. |
| Padre | Filipino | Other Asian | Wong et al. |
| Padrigo | Filipino | Other Asian | Wong et al. |
| Padrigon | Filipino | Other Asian | Wong et al. |
| Padrique | Filipino | Other Asian | Wong et al. |
| Padron | Filipino | Other Asian | Wong et al. |
| Padrones | Filipino | Other Asian | Wong et al. |
| Padua | Filipino | Other Asian | Wong et al. |
| Paduganan | Filipino | Other Asian | Wong et al. |
| Pae | Korean | Other Asian | Kranick et al. |
| Paed | Filipino | Other Asian | Wong et al. |
| Paek | Korean | Other Asian | Wong et al. |
| Pael | Asian Indian | South Asian | Wong et al. |
| Paelmo | Filipino | Other Asian | Wong et al. |
| Paeng | Korean | Other Asian | Wong et al. |
| Paeste | Filipino | Other Asian | Wong et al. |
| Paet | Filipino | Other Asian | Wong et al. |
| Paeveen |  | South Asian | Lim et al. |
| Paez | Filipino | Other Asian | Wong et al. |
| Pagador | Filipino | Other Asian | Wong et al. |
| Pagadora | Filipino | Other Asian | Wong et al. |
| Pagaduan | Filipino | Other Asian | Wong et al. |
| Pagal | Filipino | Other Asian | Wong et al. |
| Pagala | Filipino | Other Asian | Wong et al. |
| Pagalilauan | Filipino | Other Asian | Wong et al. |
| Pagaling | Filipino | Other Asian | Wong et al. |
| Pagao | Filipino | Other Asian | Wong et al. |
| Pagar | Filipino | Other Asian | Wong et al. |
| Pagaran | Filipino | Other Asian | Wong et al. |
| Pagarigan | Filipino | Other Asian | Wong et al. |
| Pagat | Filipino | Other Asian | Wong et al. |
| Pagatpatan | Filipino | Other Asian | Wong et al. |
| Pagay | Filipino | Other Asian | Wong et al. |
| Pagayon | Filipino | Other Asian | Wong et al. |
| Pagayonan | Filipino | Other Asian | Wong et al. |
| Pagba | Filipino | Other Asian | Wong et al. |
| Pagdanganan | Filipino | Other Asian | Wong et al. |
| Pagdilao | Filipino | Other Asian | Wong et al. |
| Pagente | Filipino | Other Asian | Wong et al. |
| Paggao | Filipino | Other Asian | Wong et al. |
| Pagilagan | Filipino | Other Asian | Wong et al. |
| Pagkalinawan | Filipino | Other Asian | Wong et al. |
| Pagkaliwanga | Filipino | Other Asian | Wong et al. |
| Pagkatipunan | Filipino | Other Asian | Wong et al. |
| Paglinawan | Filipino | Other Asian | Wong et al. |
| Paglomutan | Filipino | Other Asian | Wong et al. |
| Pagsanjan | Filipino | Other Asian | Wong et al. |
| Pagsibigan | Filipino | Other Asian | Wong et al. |
| Pagsisihan | Filipino | Other Asian | Wong et al. |
| Pagsolingan | Filipino | Other Asian | Wong et al. |
| Pagtakhan | Filipino | Other Asian | Kranick et al. |
| Pagtalunan | Filipino | Other Asian | Wong et al. |
| Pagtama | Filipino | Other Asian | Wong et al. |
| Pagtanac | Filipino | Other Asian | Wong et al. |
| Pagtulingan | Filipino | Other Asian | Wong et al. |
| Paguia | Filipino | Other Asian | Wong et al. |
| Paguiligan | Filipino | Other Asian | Wong et al. |
| Paguio | Filipino | Other Asian | Wong et al. |
| Paguirigan | Filipino | Other Asian | Wong et al. |
| Pagulayan | Filipino | Other Asian | Wong et al. |
| Pagunsan | Filipino | Other Asian | Wong et al. |
| Paguntalan | Filipino | Other Asian | Wong et al. |
| Paguyo | Filipino | Other Asian | Wong et al. |
| Pah | Chinese | Chinese | Wong et al. |
| Pahalan |  | South Asian | Lim et al. |
| Pahang | Filipino | Other Asian | Wong et al. |
| Pahati | Filipino | Other Asian | Wong et al. |
| Pahed | Filipino | Other Asian | Wong et al. |
| Pahinag | Filipino | Other Asian | Wong et al. |
| Pahk | Korean | Other Asian | Wong et al. |
| Pahm | Vietnamese | Other Asian | Wong et al. |
| Pahuja | Asian Indian | South Asian | Wong et al. |
| Pai | Chinese; Asian Indian | Chinese | Kranick et al. |
| Paik | Korean | Other Asian | Kranick et al. |
| Paily | Asian Indian | South Asian | Wong et al. |
| Pain | Asian Indian | South Asian | Wong et al. |
| Paintal | Asian Indian | South Asian | Wong et al. |
| Paiso | Filipino | Other Asian | Wong et al. |
| Paiste | Filipino | Other Asian | Wong et al. |
| Pajardo | Filipino | Other Asian | Wong et al. |
| Pajares | Filipino | Other Asian | Wong et al. |
| Pajarillaga | Filipino | Other Asian | Wong et al. |
| Pajarillo | Filipino | Other Asian | Wong et al. |
| Pajarito | Filipino | Other Asian | Wong et al. |
| Pajaro | Filipino | Other Asian | Wong et al. |
| Pajaron | Filipino | Other Asian | Wong et al. |
| Pajatin | Filipino | Other Asian | Wong et al. |
| Paje | Filipino | Other Asian | Wong et al. |
| Pajel | Filipino | Other Asian | Wong et al. |
| Pajela | Filipino | Other Asian | Wong et al. |
| Pajinag | Filipino | Other Asian | Wong et al. |
| Pajo | Filipino | Other Asian | Wong et al. |
| Pak | Korean; Chinese | Other Asian | Kranick et al. |
| Pakingan | Filipino | Other Asian | Wong et al. |
| Pal |  | South Asian | Lim et al. |
| Palabay | Filipino | Other Asian | Wong et al. |
| Palabrica | Filipino | Other Asian | Wong et al. |
| Palac | Filipino | Other Asian | Wong et al. |
| Palaca | Filipino | Other Asian | Wong et al. |
| Palacay | Filipino | Other Asian | Wong et al. |
| Palacio | Filipino | Other Asian | Wong et al. |
| Palacios | Filipino | Other Asian | Wong et al. |
| Palacol | Filipino | Other Asian | Wong et al. |
| Palacpac | Filipino | Other Asian | Wong et al. |
| Palad | Filipino | Other Asian | Wong et al. |
| Palada | Filipino | Other Asian | Wong et al. |
| Paladio | Filipino | Other Asian | Wong et al. |
| Palado | Filipino | Other Asian | Wong et al. |
| Paladugu | Asian Indian | South Asian | Wong et al. |
| Palafox | Filipino | Other Asian | Wong et al. |
| Palaganas | Filipino | Other Asian | Wong et al. |
| Palahang | Filipino | Other Asian | Wong et al. |
| Palalay | Filipino | Other Asian | Wong et al. |
| Palaming | Filipino | Other Asian | Wong et al. |
| Palamos | Filipino | Other Asian | Wong et al. |
| Palana | Filipino | Other Asian | Wong et al. |
| Palanca | Filipino | Other Asian | Wong et al. |
| Palang | Filipino | Other Asian | Wong et al. |
| Palaniappan | Asian Indian; Malaysian | South Asian | Kranick et al. |
| Palao | Filipino | Other Asian | Wong et al. |
| Palarca | Filipino | Other Asian | Wong et al. |
| Palaroan | Filipino | Other Asian | Wong et al. |
| Palaruan | Filipino | Other Asian | Wong et al. |
| Palas | Filipino | Other Asian | Wong et al. |
| Palasigue | Filipino | Other Asian | Wong et al. |
| Palattao | Filipino | Other Asian | Wong et al. |
| Palaylay | Filipino | Other Asian | Wong et al. |
| Palaypay | Filipino | Other Asian | Wong et al. |
| Palconit | Filipino | Other Asian | Wong et al. |
| Paldon |  | South Asian | Lim et al. |
| Palec | Filipino | Other Asian | Wong et al. |
| Paleja | Asian Indian | South Asian | Wong et al. |
| Palekar | Asian Indian | South Asian | Wong et al. |
| Palencia | Filipino | Other Asian | Wong et al. |
| Palenzuela | Filipino | Other Asian | Wong et al. |
| Paler | Filipino | Other Asian | Wong et al. |
| Paleracio | Filipino | Other Asian | Wong et al. |
| Palicte | Filipino | Other Asian | Wong et al. |
| Palileo | Filipino | Other Asian | Wong et al. |
| Palima | Filipino | Other Asian | Wong et al. |
| Palines | Filipino | Other Asian | Wong et al. |
| Palis | Filipino | Other Asian | Wong et al. |
| Palisoc | Filipino | Other Asian | Wong et al. |
| Palk | Korean | Other Asian | Wong et al. |
| Palkhiwala | Asian Indian | South Asian | Wong et al. |
| Palle | Filipino | Other Asian | Wong et al. |
| Pallera | Filipino | Other Asian | Wong et al. |
| Pallones | Filipino | Other Asian | Wong et al. |
| Pallorina | Filipino | Other Asian | Wong et al. |
| Palma | Filipino | Other Asian | Kranick et al. |
| Palmares | Filipino | Other Asian | Wong et al. |
| Palmario | Filipino | Other Asian | Wong et al. |
| Palmejar | Filipino | Other Asian | Wong et al. |
| Palmera | Filipino | Other Asian | Wong et al. |
| Palmero | Filipino | Other Asian | Wong et al. |
| Palmon | Filipino | Other Asian | Wong et al. |
| Palmos | Filipino | Other Asian | Wong et al. |
| Palogan | Filipino | Other Asian | Wong et al. |
| Paloma | Filipino | Other Asian | Wong et al. |
| Palomar | Filipino | Other Asian | Wong et al. |
| Palomares | Filipino | Other Asian | Kranick et al. |
| Palomaria | Filipino | Other Asian | Wong et al. |
| Palomo | Filipino | Other Asian | Wong et al. |
| Palompo | Filipino | Other Asian | Wong et al. |
| Palor | Filipino | Other Asian | Wong et al. |
| Palos | Filipino | Other Asian | Wong et al. |
| Palpallatoc | Filipino | Other Asian | Wong et al. |
| Palta | Asian Indian | South Asian | Wong et al. |
| Palting | Filipino | Other Asian | Wong et al. |
| Palugod | Filipino | Other Asian | Wong et al. |
| Palustre | Filipino | Other Asian | Wong et al. |
| Pama | Filipino | Other Asian | Wong et al. |
| Paman | Filipino | Other Asian | Wong et al. |
| Pamandanan | Filipino | Other Asian | Wong et al. |
| Pamani | Filipino | Other Asian | Wong et al. |
| Pamanian | Filipino | Other Asian | Wong et al. |
| Pamaran | Filipino | Other Asian | Wong et al. |
| Pamarang | Filipino | Other Asian | Wong et al. |
| Pamatian | Filipino | Other Asian | Wong et al. |
| Pamatmat | Filipino | Other Asian | Wong et al. |
| Pambid | Filipino | Other Asian | Wong et al. |
| Pamilar | Filipino | Other Asian | Wong et al. |
| Paminsan | Filipino | Other Asian | Wong et al. |
| Pamintuan | Filipino | Other Asian | Wong et al. |
| Pamittan | Filipino | Other Asian | Wong et al. |
| Pammit | Filipino | Other Asian | Wong et al. |
| Pamor | Filipino | Other Asian | Wong et al. |
| Pamplona | Filipino | Other Asian | Wong et al. |
| Pampo | Filipino | Other Asian | Wong et al. |
| Pan | Chinese | Chinese | Kranick et al. |
| Pana | Filipino | Other Asian | Wong et al. |
| Panadero | Filipino | Other Asian | Wong et al. |
| Panag | Filipino | Other Asian | Wong et al. |
| Panagsagan | Filipino | Other Asian | Wong et al. |
| Panaguiton | Filipino | Other Asian | Wong et al. |
| Panahon | Filipino | Other Asian | Wong et al. |
| Panajon | Filipino | Other Asian | Wong et al. |
| Panal | Filipino | Other Asian | Wong et al. |
| Panaligan | Filipino | Other Asian | Wong et al. |
| Panares | Filipino | Other Asian | Wong et al. |
| Panchal | Asian Indian | South Asian | Wong et al. |
| Panchapakesa | Asian Indian | South Asian | Wong et al. |
| Pancho | Filipino | Other Asian | Wong et al. |
| Pancholi | Asian Indian | South Asian | Wong et al. |
| Pancholy | Asian Indian | South Asian | Wong et al. |
| Panda | Asian Indian | South Asian | Wong et al. |
| Panday | Filipino | Other Asian | Wong et al. |
| Pande | Asian Indian | South Asian | Wong et al. |
| Pandes | Filipino | Other Asian | Wong et al. |
| Pandey | Asian Indian | South Asian | Wong et al. |
| Pandeya | Asian Indian | South Asian | Wong et al. |
| Pandeykarki |  | South Asian | Lim et al. |
| Pandher | Asian Indian | South Asian | Wong et al. |
| Pandhi | Asian Indian | South Asian | Wong et al. |
| Pandian | Asian Indian | South Asian | Wong et al. |
| Pandit | Asian Indian | South Asian | Kranick et al. |
| Pandohie |  | South Asian | Lim et al. |
| Pandy | Filipino | Other Asian | Wong et al. |
| Pandya | Asian Indian | South Asian | Wong et al. |
| Paneda | Filipino | Other Asian | Wong et al. |
| Panela | Filipino | Other Asian | Wong et al. |
| Panelo | Filipino | Other Asian | Wong et al. |
| Panem | Filipino | Other Asian | Wong et al. |
| Paner | Filipino | Other Asian | Wong et al. |
| Panergo | Filipino | Other Asian | Wong et al. |
| Panerio | Filipino | Other Asian | Wong et al. |
| Panes | Filipino | Other Asian | Kranick et al. |
| Panesar | Asian Indian | South Asian | Wong et al. |
| Pang | Chinese; Malaysian; Singapore | Chinese | Kranick et al. |
| Pangalangan | Filipino | Other Asian | Wong et al. |
| Pangan | Filipino | Other Asian | Wong et al. |
| Panganiban | Filipino | Other Asian | Wong et al. |
| Pangco | Filipino | Other Asian | Wong et al. |
| Pangdan | Filipino | Other Asian | Wong et al. |
| Pangilinan | Filipino | Other Asian | Wong et al. |
| Pangindian | Filipino | Other Asian | Wong et al. |
| Panicker | Asian Indian | South Asian | Wong et al. |
| Panigrahi | Asian Indian | South Asian | Wong et al. |
| Panikkar | Asian Indian | South Asian | Wong et al. |
| Panilag | Filipino | Other Asian | Wong et al. |
| Paningbatan | Filipino | Other Asian | Wong et al. |
| Panis | Filipino | Other Asian | Wong et al. |
| Panisa | Filipino | Other Asian | Wong et al. |
| Panit | Filipino | Other Asian | Wong et al. |
| Panjwani | Asian Indian | South Asian | Wong et al. |
| Panlaqui | Filipino | Other Asian | Wong et al. |
| Panlasigui | Filipino | Other Asian | Wong et al. |
| Panlilio | Filipino | Other Asian | Wong et al. |
| Panni |  | South Asian | Lim et al. |
| Pannu | Asian Indian | South Asian | Wong et al. |
| Pano | Filipino | Other Asian | Wong et al. |
| Panopio | Filipino | Other Asian | Wong et al. |
| Panotes | Filipino | Other Asian | Wong et al. |
| Pansacola | Filipino | Other Asian | Wong et al. |
| Pansoy | Filipino | Other Asian | Wong et al. |
| Panta |  | South Asian | Lim et al. |
| Pantaleon | Filipino | Other Asian | Wong et al. |
| Pantangco | Filipino | Other Asian | Wong et al. |
| Pante | Filipino | Other Asian | Wong et al. |
| Panti | Filipino | Other Asian | Wong et al. |
| Pantig | Filipino | Other Asian | Wong et al. |
| Pantil | Filipino | Other Asian | Wong et al. |
| Pantoja | Filipino | Other Asian | Wong et al. |
| Pantonial | Filipino | Other Asian | Wong et al. |
| Pantorilla | Filipino | Other Asian | Wong et al. |
| Pantua | Filipino | Other Asian | Wong et al. |
| Panuelos | Filipino | Other Asian | Wong et al. |
| Panuncio | Filipino | Other Asian | Wong et al. |
| Panwala | Asian Indian | South Asian | Wong et al. |
| Panwar | Asian Indian | South Asian | Wong et al. |
| Pao | Chinese | Chinese | Wong et al. |
| Papagayo | Filipino | Other Asian | Wong et al. |
| Papasin | Filipino | Other Asian | Wong et al. |
| Papelera | Filipino | Other Asian | Wong et al. |
| Papica | Filipino | Other Asian | Wong et al. |
| Papilla | Filipino | Other Asian | Wong et al. |
| Pappan | Asian Indian | South Asian | Wong et al. |
| Pappu | Asian Indian | South Asian | Wong et al. |
| Pappy | Asian Indian | South Asian | Wong et al. |
| Par | Filipino | Other Asian | Wong et al. |
| Paraan | Filipino | Other Asian | Wong et al. |
| Paradela | Filipino | Other Asian | Wong et al. |
| Paradero | Filipino | Other Asian | Wong et al. |
| Paradeza | Filipino | Other Asian | Wong et al. |
| Parado | Filipino | Other Asian | Wong et al. |
| Parafina | Filipino | Other Asian | Wong et al. |
| Paragas | Filipino | Other Asian | Wong et al. |
| Paragua | Filipino | Other Asian | Wong et al. |
| Paraguas | Filipino | Other Asian | Wong et al. |
| Paraguya | Filipino | Other Asian | Wong et al. |
| Paraiso | Filipino | Other Asian | Wong et al. |
| Parakh | Asian Indian | South Asian | Wong et al. |
| Parales | Filipino | Other Asian | Wong et al. |
| Param | Filipino | Other Asian | Wong et al. |
| Parameswaran | Asian Indian | South Asian | Wong et al. |
| Paramio | Filipino | Other Asian | Wong et al. |
| Paran | Filipino | Other Asian | Wong et al. |
| Paranada | Filipino | Other Asian | Wong et al. |
| Paranal | Filipino | Other Asian | Wong et al. |
| Parangan | Filipino | Other Asian | Wong et al. |
| Parani | Filipino | Other Asian | Wong et al. |
| Paranjape | Asian Indian | South Asian | Wong et al. |
| Paranjpe | Asian Indian | South Asian | Wong et al. |
| Paraoan | Filipino | Other Asian | Wong et al. |
| Paraon | Filipino | Other Asian | Wong et al. |
| Paras | Filipino | Other Asian | Wong et al. |
| Parashar | Asian Indian | South Asian | Wong et al. |
| Paraso | Filipino | Other Asian | Wong et al. |
| Parasram |  | South Asian | Lim et al. |
| Parawan | Filipino | Other Asian | Wong et al. |
| Parayao | Filipino | Other Asian | Wong et al. |
| Parayil | Asian Indian | South Asian | Wong et al. |
| Parayno | Filipino | Other Asian | Wong et al. |
| Paraz | Filipino | Other Asian | Wong et al. |
| Parazo | Filipino | Other Asian | Wong et al. |
| Parbhakar | Asian Indian | South Asian | Wong et al. |
| Parbhoo | Asian Indian | South Asian | Wong et al. |
| Parbhubhai | Asian Indian | South Asian | Wong et al. |
| Parbin |  | South Asian | Lim et al. |
| Parbo | Filipino | Other Asian | Wong et al. |
| Parcasio | Filipino | Other Asian | Wong et al. |
| Parcero | Filipino | Other Asian | Wong et al. |
| Parchamento | Filipino | Other Asian | Wong et al. |
| Parco | Filipino | Other Asian | Wong et al. |
| Parcon | Filipino | Other Asian | Wong et al. |
| Pardilla | Filipino | Other Asian | Wong et al. |
| Pardillo | Filipino | Other Asian | Wong et al. |
| Pardo | Filipino | Other Asian | Wong et al. |
| Parducho | Filipino | Other Asian | Wong et al. |
| Pardumansingh |  | South Asian | Lim et al. |
| Paredes | Filipino | Other Asian | Wong et al. |
| Pareja | Filipino | Other Asian | Wong et al. |
| Parekh | Asian Indian | South Asian | Kranick et al. |
| Parel | Filipino | Other Asian | Wong et al. |
| Pareno | Filipino | Other Asian | Wong et al. |
| Parfan | Filipino | Other Asian | Wong et al. |
| Parghi | Asian Indian | South Asian | Wong et al. |
| Parhar | Asian Indian | South Asian | Wong et al. |
| Parial | Filipino | Other Asian | Wong et al. |
| Parian | Filipino | Other Asian | Wong et al. |
| Parihar | Asian Indian | South Asian | Wong et al. |
| Parikh | Asian Indian | South Asian | Wong et al. |
| Parikm | Asian Indian | South Asian | Wong et al. |
| Parilla | Filipino | Other Asian | Wong et al. |
| Parimi | Asian Indian | South Asian | Wong et al. |
| Parinas | Filipino | Other Asian | Wong et al. |
| Paringit | Filipino | Other Asian | Wong et al. |
| Park | Korean | Other Asian | Kranick et al. |
| Parkash | Asian Indian | South Asian | Wong et al. |
| Parkhani | Asian Indian | South Asian | Wong et al. |
| Parlade | Filipino | Other Asian | Wong et al. |
| Parlan | Filipino | Other Asian | Wong et al. |
| Parmar | Asian Indian | South Asian | Wong et al. |
| Parocha | Filipino | Other Asian | Wong et al. |
| Paronable | Filipino | Other Asian | Wong et al. |
| Parong | Filipino | Other Asian | Wong et al. |
| Parpan | Filipino | Other Asian | Wong et al. |
| Parpana | Filipino | Other Asian | Wong et al. |
| Parra | Filipino | Other Asian | Wong et al. |
| Parras | Filipino | Other Asian | Wong et al. |
| Parrenas | Filipino | Other Asian | Wong et al. |
| Parreno | Filipino | Other Asian | Wong et al. |
| Parrilla | Filipino | Other Asian | Wong et al. |
| Parrocha | Filipino | Other Asian | Wong et al. |
| Parshad | Asian Indian | South Asian | Wong et al. |
| Parsuram |  | South Asian | Lim et al. |
| Parthasarath | Asian Indian | South Asian | Wong et al. |
| Parthasarathy | Asian Indian | South Asian | Kranick et al. |
| Partible | Filipino | Other Asian | Wong et al. |
| Partido | Filipino | Other Asian | Wong et al. |
| Partosa | Filipino | Other Asian | Wong et al. |
| Partosan | Filipino | Other Asian | Wong et al. |
| Partoza | Filipino | Other Asian | Wong et al. |
| Parubrub | Filipino | Other Asian | Wong et al. |
| Paruchuri | Asian Indian | South Asian | Wong et al. |
| Parulan | Filipino | Other Asian | Wong et al. |
| Parulekar | Asian Indian | South Asian | Wong et al. |
| Parumog | Filipino | Other Asian | Wong et al. |
| Parungao | Filipino | Other Asian | Wong et al. |
| Parvati |  | South Asian | Lim et al. |
| Parvaz |  | South Asian | Lim et al. |
| Parveen |  | South Asian | Lim et al. |
| Parvez |  | South Asian | Lim et al. |
| Parvin |  | South Asian | Lim et al. |
| Pasa | Filipino | Other Asian | Wong et al. |
| Pasag | Filipino | Other Asian | Wong et al. |
| Pasalo | Filipino | Other Asian | Wong et al. |
| Pasamba | Filipino | Other Asian | Wong et al. |
| Pasamonte | Filipino | Other Asian | Wong et al. |
| Pasana | Filipino | Other Asian | Wong et al. |
| Pasao | Filipino | Other Asian | Wong et al. |
| Pasaoa | Filipino | Other Asian | Wong et al. |
| Pasaol | Filipino | Other Asian | Wong et al. |
| Pasaporte | Filipino | Other Asian | Wong et al. |
| Pasatiempo | Filipino | Other Asian | Wong et al. |
| Pasay | Filipino | Other Asian | Wong et al. |
| Pascasio | Filipino | Other Asian | Wong et al. |
| Pascion | Filipino | Other Asian | Wong et al. |
| Pasco | Filipino | Other Asian | Wong et al. |
| Pascua | Filipino | Other Asian | Wong et al. |
| Pascual | Filipino | Other Asian | Wong et al. |
| Pascubillo | Filipino | Other Asian | Wong et al. |
| Pasetes | Filipino | Other Asian | Wong et al. |
| Pasha | Asian Indian | South Asian | Wong et al. |
| Pasia | Filipino | Other Asian | Wong et al. |
| Pasibe | Filipino | Other Asian | Wong et al. |
| Pasig | Filipino | Other Asian | Wong et al. |
| Pasiliao | Filipino | Other Asian | Wong et al. |
| Pasimio | Filipino | Other Asian | Wong et al. |
| Pasion | Filipino | Other Asian | Wong et al. |
| Pasol | Filipino | Other Asian | Wong et al. |
| Pasquin | Filipino | Other Asian | Wong et al. |
| Pasricha | Asian Indian | South Asian | Wong et al. |
| Passi | Asian Indian | South Asian | Wong et al. |
| Pastakia | Asian Indian | South Asian | Wong et al. |
| Pastolero | Filipino | Other Asian | Wong et al. |
| Pastor | Filipino | Other Asian | Wong et al. |
| Pastoral | Filipino | Other Asian | Wong et al. |
| Pastores | Filipino | Other Asian | Wong et al. |
| Pastorfide | Filipino | Other Asian | Wong et al. |
| Pastoriza | Filipino | Other Asian | Wong et al. |
| Pastrana | Filipino | Other Asian | Wong et al. |
| Pastrano | Filipino | Other Asian | Wong et al. |
| Pasumbal | Filipino | Other Asian | Wong et al. |
| Patac | Filipino | Other Asian | Wong et al. |
| Patacsil | Filipino | Other Asian | Wong et al. |
| Patadia | Asian Indian | South Asian | Wong et al. |
| Patag | Filipino | Other Asian | Wong et al. |
| Patague | Filipino | Other Asian | Wong et al. |
| Patajo | Filipino | Other Asian | Wong et al. |
| Patalinghug | Filipino | Other Asian | Wong et al. |
| Patalinjug | Filipino | Other Asian | Wong et al. |
| Patangan | Filipino | Other Asian | Wong et al. |
| Patankar | Asian Indian | South Asian | Wong et al. |
| Patano | Filipino | Other Asian | Wong et al. |
| Patao | Filipino | Other Asian | Wong et al. |
| Patawaran | Filipino | Other Asian | Wong et al. |
| Patayan | Filipino | Other Asian | Wong et al. |
| Patdu | Filipino | Other Asian | Wong et al. |
| Patel | Asian Indian | South Asian | Kranick et al. |
| Patell | Asian Indian | South Asian | Wong et al. |
| Patez | Asian Indian | South Asian | Wong et al. |
| Pathak | Asian Indian | South Asian | Wong et al. |
| Pathan |  | South Asian | Lim et al. |
| Pathare | Asian Indian | South Asian | Wong et al. |
| Pathmasiri | Sri Lankan | South Asian | Kranick et al. |
| Pathrose | Asian Indian | South Asian | Wong et al. |
| Pati | Asian Indian | South Asian | Wong et al. |
| Patiag | Filipino | Other Asian | Wong et al. |
| Patibandla | Asian Indian | South Asian | Wong et al. |
| Patidar | Asian Indian | South Asian | Wong et al. |
| Patil | Asian Indian | South Asian | Wong et al. |
| Pating | Filipino | Other Asian | Wong et al. |
| Patino | Filipino | Other Asian | Wong et al. |
| Patio | Filipino | Other Asian | Wong et al. |
| Patkar | Asian Indian | South Asian | Wong et al. |
| Patle | Asian Indian | South Asian | Wong et al. |
| Patnaik | Asian Indian | South Asian | Wong et al. |
| Patney | Asian Indian | South Asian | Wong et al. |
| Patni | Asian Indian | South Asian | Wong et al. |
| Patoc | Filipino | Other Asian | Wong et al. |
| Patole | Asian Indian | South Asian | Wong et al. |
| Patolot | Filipino | Other Asian | Wong et al. |
| Patrawala | Asian Indian | South Asian | Wong et al. |
| Patriarca | Filipino | Other Asian | Wong et al. |
| Patricio | Filipino | Other Asian | Wong et al. |
| Patrimonio | Filipino | Other Asian | Wong et al. |
| Patrocinio | Filipino | Other Asian | Wong et al. |
| Patron | Filipino | Other Asian | Wong et al. |
| Pattanayak | Asian Indian | South Asian | Wong et al. |
| Pattani | Asian Indian | South Asian | Wong et al. |
| Pattar | Asian Indian | South Asian | Wong et al. |
| Patti |  | South Asian | Lim et al. |
| Pattugalan | Filipino | Other Asian | Wong et al. |
| Patubo | Filipino | Other Asian | Wong et al. |
| Patulot | Filipino | Other Asian | Wong et al. |
| Patungan | Filipino | Other Asian | Wong et al. |
| Paturi | Asian Indian | South Asian | Wong et al. |
| Patwa | Asian Indian | South Asian | Wong et al. |
| Patwardhan | Asian Indian | South Asian | Wong et al. |
| Patwary | Bangladeshi | South Asian | Kranick et al. |
| Pau | Chinese | Chinese | Wong et al. |
| Pauig | Filipino | Other Asian | Wong et al. |
| Paul |  | South Asian | Lim et al. |
| Paule | Filipino | Other Asian | Wong et al. |
| Paulino | Filipino | Other Asian | Wong et al. |
| Paulite | Filipino | Other Asian | Wong et al. |
| Paulmino | Filipino | Other Asian | Wong et al. |
| Paulo | Filipino | Other Asian | Wong et al. |
| Paulose | Asian Indian | South Asian | Wong et al. |
| Paunan | Filipino | Other Asian | Wong et al. |
| Paunil | Filipino | Other Asian | Wong et al. |
| Pautin | Filipino | Other Asian | Wong et al. |
| Pavico | Filipino | Other Asian | Wong et al. |
| Pavo | Filipino | Other Asian | Wong et al. |
| Pavon | Filipino | Other Asian | Wong et al. |
| Pawar | Asian Indian | South Asian | Wong et al. |
| Payabyab | Filipino | Other Asian | Wong et al. |
| Payad | Filipino | Other Asian | Wong et al. |
| Payawal | Filipino | Other Asian | Wong et al. |
| Payo | Filipino | Other Asian | Wong et al. |
| Payomo | Filipino | Other Asian | Wong et al. |
| Payon | Filipino | Other Asian | Wong et al. |
| Payongayong | Filipino | Other Asian | Wong et al. |
| Payopay | Filipino | Other Asian | Wong et al. |
| Payot | Filipino | Other Asian | Wong et al. |
| Payoyo | Filipino | Other Asian | Wong et al. |
| Payumo | Filipino | Other Asian | Wong et al. |
| Payuran | Filipino | Other Asian | Wong et al. |
| Paz | Filipino | Other Asian | Wong et al. |
| Pebenito | Filipino | Other Asian | Wong et al. |
| Pecache | Filipino | Other Asian | Wong et al. |
| Pecho | Filipino | Other Asian | Wong et al. |
| Pecjo | Filipino | Other Asian | Wong et al. |
| Pecson | Filipino | Other Asian | Wong et al. |
| Ped | Filipino | Other Asian | Wong et al. |
| Peddada | Asian Indian | South Asian | Wong et al. |
| Pedeglorio | Filipino | Other Asian | Wong et al. |
| Pednekar | Asian Indian | South Asian | Wong et al. |
| Pedrajas | Filipino | Other Asian | Wong et al. |
| Pedralvez | Filipino | Other Asian | Wong et al. |
| Pedraza | Filipino | Other Asian | Wong et al. |
| Pedregosa | Filipino | Other Asian | Wong et al. |
| Pedrigal | Filipino | Other Asian | Wong et al. |
| Pedro | Filipino | Other Asian | Wong et al. |
| Pedroche | Filipino | Other Asian | Wong et al. |
| Pedron | Filipino | Other Asian | Wong et al. |
| Pedronan | Filipino | Other Asian | Wong et al. |
| Pedrosa | Filipino | Other Asian | Wong et al. |
| Pedroso | Filipino | Other Asian | Wong et al. |
| Pedrozo | Filipino | Other Asian | Wong et al. |
| Peeran | Asian Indian | South Asian | Wong et al. |
| Peganyee | Asian Indian | South Asian | Wong et al. |
| Pei | Chinese | Chinese | Wong et al. |
| Peji | Filipino | Other Asian | Wong et al. |
| Pekson | Filipino | Other Asian | Wong et al. |
| Pelaez | Filipino | Other Asian | Wong et al. |
| Pelagio | Filipino | Other Asian | Wong et al. |
| Pelayo | Filipino | Other Asian | Wong et al. |
| Pelea | Filipino | Other Asian | Wong et al. |
| Pelejo | Filipino | Other Asian | Wong et al. |
| Pelen | Filipino | Other Asian | Wong et al. |
| Peleo | Filipino | Other Asian | Wong et al. |
| Pelina | Filipino | Other Asian | Wong et al. |
| Pelingon | Filipino | Other Asian | Wong et al. |
| Pelobello | Filipino | Other Asian | Wong et al. |
| Pelonio | Filipino | Other Asian | Wong et al. |
| Pena | Filipino | Other Asian | Wong et al. |
| Penacerrada | Filipino | Other Asian | Wong et al. |
| Penafiel | Filipino | Other Asian | Wong et al. |
| Penaflor | Filipino | Other Asian | Wong et al. |
| Penaflorida | Filipino | Other Asian | Wong et al. |
| Penafuerte | Filipino | Other Asian | Wong et al. |
| Penalba | Filipino | Other Asian | Wong et al. |
| Penales | Filipino | Other Asian | Wong et al. |
| Penalosa | Filipino | Other Asian | Wong et al. |
| Penaloza | Filipino | Other Asian | Wong et al. |
| Penamante | Filipino | Other Asian | Wong et al. |
| Penano | Filipino | Other Asian | Wong et al. |
| Penaranda | Filipino | Other Asian | Wong et al. |
| Penaredondo | Filipino | Other Asian | Wong et al. |
| Penarubia | Filipino | Other Asian | Wong et al. |
| Penas | Filipino | Other Asian | Wong et al. |
| Pendharkar | Asian Indian | South Asian | Wong et al. |
| Pendon | Filipino | Other Asian | Wong et al. |
| Pendse | Asian Indian | South Asian | Wong et al. |
| Penera | Filipino | Other Asian | Wong et al. |
| Penetrante | Filipino | Other Asian | Wong et al. |
| Peneyra | Filipino | Other Asian | Wong et al. |
| Peng | Chinese | Chinese | Wong et al. |
| Pengson | Filipino | Other Asian | Wong et al. |
| Penolio | Filipino | Other Asian | Wong et al. |
| Penserga | Filipino | Other Asian | Wong et al. |
| Pensotes | Filipino | Other Asian | Wong et al. |
| Pentecostes | Filipino | Other Asian | Wong et al. |
| Penuela | Filipino | Other Asian | Wong et al. |
| Penuliar | Filipino | Other Asian | Wong et al. |
| Penullar | Filipino | Other Asian | Wong et al. |
| Pepa | Filipino | Other Asian | Wong et al. |
| Pepito | Filipino | Other Asian | Wong et al. |
| Perales | Filipino | Other Asian | Wong et al. |
| Peralta | Filipino | Other Asian | Wong et al. |
| Peras | Filipino | Other Asian | Wong et al. |
| Perdido | Filipino | Other Asian | Wong et al. |
| Perdigon | Filipino | Other Asian | Wong et al. |
| Perdon | Filipino | Other Asian | Wong et al. |
| Perea | Filipino | Other Asian | Wong et al. |
| Pereda | Filipino | Other Asian | Wong et al. |
| Peredo | Filipino | Other Asian | Wong et al. |
| Peregil | Filipino | Other Asian | Wong et al. |
| Peregrino | Filipino | Other Asian | Wong et al. |
| Pereira | Asian Indian | South Asian | Wong et al. |
| Perena | Filipino | Other Asian | Wong et al. |
| Perera |  | South Asian | Lim et al. |
| Pereras | Filipino | Other Asian | Wong et al. |
| Peres | Filipino | Other Asian | Wong et al. |
| Perey | Filipino | Other Asian | Wong et al. |
| Pereyra | Filipino | Other Asian | Wong et al. |
| Perez | Filipino | Other Asian | Wong et al. |
| Perfecto | Filipino | Other Asian | Wong et al. |
| Peria | Filipino | Other Asian | Wong et al. |
| Perico | Filipino | Other Asian | Wong et al. |
| Perido | Filipino | Other Asian | Wong et al. |
| Periquet | Filipino | Other Asian | Wong et al. |
| Peris |  | South Asian | Lim et al. |
| Perlas | Filipino | Other Asian | Wong et al. |
| Permaul |  | South Asian | Lim et al. |
| Permejo | Filipino | Other Asian | Wong et al. |
| Permito | Filipino | Other Asian | Wong et al. |
| Perng | Chinese | Chinese | Wong et al. |
| Pernia | Filipino | Other Asian | Wong et al. |
| Pernito | Filipino | Other Asian | Wong et al. |
| Perocho | Filipino | Other Asian | Wong et al. |
| Perolina | Filipino | Other Asian | Wong et al. |
| Peros | Filipino | Other Asian | Wong et al. |
| Perreras | Filipino | Other Asian | Wong et al. |
| Persaud | Asian Indian | South Asian | Kranick et al. |
| Pershad | Asian Indian | South Asian | Wong et al. |
| Pertierra | Filipino | Other Asian | Wong et al. |
| Peru | Filipino | Other Asian | Wong et al. |
| Perucho | Filipino | Other Asian | Wong et al. |
| Perumal | Asian Indian | South Asian | Wong et al. |
| Perveen |  | South Asian | Lim et al. |
| Pervej |  | South Asian | Lim et al. |
| Pervez | Pakistani; Bangladeshi | South Asian | Kranick et al. |
| Pervin | Bangladeshi | South Asian | Kranick et al. |
| Pesado | Filipino | Other Asian | Wong et al. |
| Pescador | Filipino | Other Asian | Wong et al. |
| Peshimam | Asian Indian | South Asian | Wong et al. |
| Pesigan | Filipino | Other Asian | Wong et al. |
| Pestanas | Filipino | Other Asian | Wong et al. |
| Pestano | Filipino | Other Asian | Kranick et al. |
| Pestonji | Asian Indian | South Asian | Wong et al. |
| Pet | Filipino | Other Asian | Wong et al. |
| Petalver | Filipino | Other Asian | Wong et al. |
| Petel | Asian Indian | South Asian | Wong et al. |
| Petelo | Filipino | Other Asian | Wong et al. |
| Peter |  | South Asian | Lim et al. |
| Pethe | Asian Indian | South Asian | Wong et al. |
| Petilla | Filipino | Other Asian | Wong et al. |
| Petines | Filipino | Other Asian | Wong et al. |
| Phadke | Asian Indian | South Asian | Wong et al. |
| Phadnis | Asian Indian | South Asian | Wong et al. |
| Phai | Vietnamese | Other Asian | Wong et al. |
| Phak | Korean | Other Asian | Wong et al. |
| Phaliea |  | South Asian | Lim et al. |
| Pham | Vietnamese | Other Asian | Kranick et al. |
| Phan | Vietnamese | Other Asian | Kranick et al. |
| Phansalkar | Asian Indian | South Asian | Wong et al. |
| Phap | Vietnamese | Other Asian | Wong et al. |
| Phat | Vietnamese | Other Asian | Wong et al. |
| Phatak | Asian Indian | South Asian | Wong et al. |
| Phee | Filipino | Other Asian | Wong et al. |
| Phi | Vietnamese | Other Asian | Wong et al. |
| Phien | Vietnamese | Other Asian | Wong et al. |
| Philip | Asian Indian | South Asian | Wong et al. |
| Philipose | Asian Indian | South Asian | Wong et al. |
| Phillip | Asian Indian | South Asian | Wong et al. |
| Pho | Vietnamese | Other Asian | Wong et al. |
| Phojanakong | Thai | Other Asian | Kranick et al. |
| Phong | Vietnamese | Other Asian | Wong et al. |
| Phoon | Chinese | Chinese | Wong et al. |
| Phou | Cambodian | Other Asian | Kranick et al. |
| Phu | Vietnamese | Other Asian | Wong et al. |
| Phua | Chinese | Chinese | Wong et al. |
| Phuc | Vietnamese | Other Asian | Wong et al. |
| Phun | Vietnamese | Other Asian | Wong et al. |
| Phung | Vietnamese | Other Asian | Wong et al. |
| Phuoc | Vietnamese | Other Asian | Wong et al. |
| Phuong | Vietnamese | Other Asian | Wong et al. |
| Pi | Chinese | Chinese | Wong et al. |
| Pia | Filipino | Other Asian | Wong et al. |
| Piak | Korean | Other Asian | Wong et al. |
| Piamonte | Filipino | Other Asian | Wong et al. |
| Pian | Chinese | Chinese | Wong et al. |
| Piano | Filipino | Other Asian | Wong et al. |
| Piansay | Filipino | Other Asian | Wong et al. |
| Piao | Chinese | Chinese | Kranick et al. |
| Piau | Chinese | Chinese | Wong et al. |
| Picache | Filipino | Other Asian | Wong et al. |
| Picar | Filipino | Other Asian | Wong et al. |
| Picardal | Filipino | Other Asian | Wong et al. |
| Picardo | Filipino | Other Asian | Wong et al. |
| Picart | Filipino | Other Asian | Wong et al. |
| Picazo | Filipino | Other Asian | Wong et al. |
| Piccio | Filipino | Other Asian | Wong et al. |
| Pichay | Filipino | Other Asian | Wong et al. |
| Pico | Filipino | Other Asian | Wong et al. |
| Piczon | Filipino | Other Asian | Wong et al. |
| Pidlaoan | Filipino | Other Asian | Wong et al. |
| Pido | Filipino | Other Asian | Wong et al. |
| Piedad | Filipino | Other Asian | Wong et al. |
| Pielago | Filipino | Other Asian | Wong et al. |
| Pien | Chinese | Chinese | Wong et al. |
| Piezas | Filipino | Other Asian | Wong et al. |
| Piga | Filipino | Other Asian | Wong et al. |
| Pigao | Filipino | Other Asian | Wong et al. |
| Piguing | Filipino | Other Asian | Wong et al. |
| Pih | Chinese | Chinese | Wong et al. |
| Pijuan | Filipino | Other Asian | Wong et al. |
| Pil | Korean | Other Asian | Wong et al. |
| Pila | Filipino | Other Asian | Wong et al. |
| Pilao | Filipino | Other Asian | Wong et al. |
| Pilapil | Filipino | Other Asian | Wong et al. |
| Pilar | Filipino | Other Asian | Wong et al. |
| Pilares | Filipino | Other Asian | Wong et al. |
| Pili | Filipino | Other Asian | Wong et al. |
| Pilien | Filipino | Other Asian | Wong et al. |
| Pillai | Asian Indian | South Asian | Wong et al. |
| Pillay | Asian Indian | South Asian | Wong et al. |
| Pillos | Filipino | Other Asian | Wong et al. |
| Pilotin | Filipino | Other Asian | Wong et al. |
| Pilpa | Filipino | Other Asian | Wong et al. |
| Pimentel | Filipino | Other Asian | Kranick et al. |
| Pimentil | Filipino | Other Asian | Wong et al. |
| Pin | Chinese | Chinese | Wong et al. |
| Pinacate | Filipino | Other Asian | Wong et al. |
| Pinaroc | Filipino | Other Asian | Wong et al. |
| Pinat | Filipino | Other Asian | Wong et al. |
| Pinca | Filipino | Other Asian | Wong et al. |
| Pineda | Filipino | Other Asian | Kranick et al. |
| Pinera | Filipino | Other Asian | Wong et al. |
| Pinero | Filipino | Other Asian | Wong et al. |
| Ping | Chinese | Chinese | Wong et al. |
| Pinga | Filipino | Other Asian | Wong et al. |
| Pingol | Filipino | Other Asian | Wong et al. |
| Pingul | Filipino | Other Asian | Wong et al. |
| Pinili | Filipino | Other Asian | Wong et al. |
| Piniones | Filipino | Other Asian | Wong et al. |
| Pinlac | Filipino | Other Asian | Wong et al. |
| Pinnock | Japanese | Other Asian | Wong et al. |
| Pino | Filipino | Other Asian | Wong et al. |
| Pinol | Filipino | Other Asian | Wong et al. |
| Pinoliar | Filipino | Other Asian | Wong et al. |
| Pinon | Filipino | Other Asian | Wong et al. |
| Pinote | Filipino | Other Asian | Wong et al. |
| Pinpin | Filipino | Other Asian | Wong et al. |
| Pintado | Filipino | Other Asian | Wong et al. |
| Pintang | Filipino | Other Asian | Wong et al. |
| Pintor | Filipino | Other Asian | Wong et al. |
| Pinzon | Filipino | Other Asian | Wong et al. |
| Piocos | Filipino | Other Asian | Wong et al. |
| Piol | Filipino | Other Asian | Wong et al. |
| Piolo | Filipino | Other Asian | Wong et al. |
| Pioquinto | Filipino | Other Asian | Wong et al. |
| Pipalia | Asian Indian | South Asian | Wong et al. |
| Pipitone |  | South Asian | Lim et al. |
| Piplani | Asian Indian | South Asian | Wong et al. |
| Pira |  | South Asian | Lim et al. |
| Pirani | Asian Indian | South Asian | Wong et al. |
| Pirbhai | Asian Indian | South Asian | Wong et al. |
| Pires | Asian Indian | South Asian | Wong et al. |
| Piring | Filipino | Other Asian | Wong et al. |
| Piros | Filipino | Other Asian | Wong et al. |
| Pirote | Filipino | Other Asian | Wong et al. |
| Pirzada | Asian Indian | South Asian | Wong et al. |
| Pisco | Filipino | Other Asian | Wong et al. |
| Pisharody | Asian Indian | South Asian | Wong et al. |
| Pisig | Filipino | Other Asian | Wong et al. |
| Piso | Filipino | Other Asian | Wong et al. |
| Pita | Filipino | Other Asian | Wong et al. |
| Pithadia | Asian Indian | South Asian | Wong et al. |
| Pitogo | Filipino | Other Asian | Wong et al. |
| Pitpit | Filipino | Other Asian | Wong et al. |
| Pittiman |  | South Asian | Lim et al. |
| Piu | Chinese | Chinese | Wong et al. |
| Pius |  | South Asian | Lim et al. |
| Pizana | Filipino | Other Asian | Wong et al. |
| Pizarro | Filipino | Other Asian | Wong et al. |
| Placido | Filipino | Other Asian | Wong et al. |
| Plagata | Filipino | Other Asian | Wong et al. |
| Plamenco | Filipino | Other Asian | Wong et al. |
| Plana | Filipino | Other Asian | Wong et al. |
| Planas | Filipino | Other Asian | Wong et al. |
| Planta | Filipino | Other Asian | Wong et al. |
| Plantilla | Filipino | Other Asian | Kranick et al. |
| Plata | Filipino | Other Asian | Wong et al. |
| Platero | Filipino | Other Asian | Wong et al. |
| Platon | Filipino | Other Asian | Wong et al. |
| Plaza | Filipino | Other Asian | Wong et al. |
| Plenos | Filipino | Other Asian | Wong et al. |
| Po | Burmese; Filipino | Other Asian | Kranick et al. |
| Poa | Chinese | Chinese | Wong et al. |
| Poblacion | Filipino | Other Asian | Wong et al. |
| Poblador | Filipino | Other Asian | Wong et al. |
| Poblete | Filipino | Other Asian | Wong et al. |
| Pobre | Filipino | Other Asian | Wong et al. |
| Poddar | Asian Indian | South Asian | Wong et al. |
| Podder |  | South Asian | Lim et al. |
| Poh | Chinese | Chinese | Wong et al. |
| Poi | Chinese | Chinese | Wong et al. |
| Pojas | Filipino | Other Asian | Wong et al. |
| Polanco | Filipino | Other Asian | Wong et al. |
| Polavarapu | Asian Indian | South Asian | Wong et al. |
| Polendey | Filipino | Other Asian | Wong et al. |
| Policar | Filipino | Other Asian | Wong et al. |
| Policarpio | Filipino | Other Asian | Wong et al. |
| Polido | Filipino | Other Asian | Wong et al. |
| Polinar | Filipino | Other Asian | Wong et al. |
| Polintan | Filipino | Other Asian | Wong et al. |
| Poliquit | Filipino | Other Asian | Wong et al. |
| Polistico | Filipino | Other Asian | Wong et al. |
| Polo |  | South Asian | Lim et al. |
| Polvorosa | Filipino | Other Asian | Wong et al. |
| Poly |  | South Asian | Lim et al. |
| Pomar | Filipino | Other Asian | Wong et al. |
| Pon | Chinese | Chinese | Wong et al. |
| Ponce | Filipino | Other Asian | Wong et al. |
| Poncedeleon | Filipino | Other Asian | Wong et al. |
| Ponciano | Filipino | Other Asian | Wong et al. |
| Ponda | Asian Indian | South Asian | Wong et al. |
| Pondoc | Filipino | Other Asian | Wong et al. |
| Ponferrada | Filipino | Other Asian | Wong et al. |
| Pong | Chinese | Chinese | Wong et al. |
| Pongan | Filipino | Other Asian | Wong et al. |
| Pongco | Filipino | Other Asian | Wong et al. |
| Pongos | Filipino | Other Asian | Wong et al. |
| Poniente | Filipino | Other Asian | Wong et al. |
| Ponio | Filipino | Other Asian | Kranick et al. |
| Ponniah | Asian Indian | South Asian | Wong et al. |
| Ponnusamy | Asian Indian | South Asian | Wong et al. |
| Ponnuswamy | Asian Indian | South Asian | Wong et al. |
| Ponnuthurai |  | South Asian | Lim et al. |
| Pono | Filipino | Other Asian | Wong et al. |
| Ponseca | Filipino | Other Asian | Wong et al. |
| Pontanilla | Filipino | Other Asian | Wong et al. |
| Pontejos | Filipino | Other Asian | Wong et al. |
| Ponteras | Filipino | Other Asian | Wong et al. |
| Pontillas | Filipino | Other Asian | Wong et al. |
| Pontilo | Filipino | Other Asian | Wong et al. |
| Pontino | Filipino | Other Asian | Wong et al. |
| Poo | Chinese | Chinese | Wong et al. |
| Poola | Asian Indian | South Asian | Kranick et al. |
| Poon | Chinese | Chinese | Wong et al. |
| Poonawala | Asian Indian | South Asian | Wong et al. |
| Poonawalla | Asian Indian | South Asian | Wong et al. |
| Poong | Korean | Other Asian | Wong et al. |
| Poonja | Asian Indian | South Asian | Wong et al. |
| Popat | Asian Indian | South Asian | Wong et al. |
| Popatia | Asian Indian | South Asian | Wong et al. |
| Popatlal | Asian Indian | South Asian | Wong et al. |
| Poquiz | Filipino | Other Asian | Wong et al. |
| Por | Chinese | Chinese | Wong et al. |
| Poral | Filipino | Other Asian | Wong et al. |
| Porbandarwal | Asian Indian | South Asian | Wong et al. |
| Porcincula | Filipino | Other Asian | Wong et al. |
| Porcioncula | Filipino | Other Asian | Wong et al. |
| Porciuncula | Filipino | Other Asian | Wong et al. |
| Porcuna | Filipino | Other Asian | Wong et al. |
| Pornela | Filipino | Other Asian | Wong et al. |
| Porral | Filipino | Other Asian | Wong et al. |
| Porras | Filipino | Other Asian | Wong et al. |
| Portacio | Filipino | Other Asian | Wong et al. |
| Portal | Filipino | Other Asian | Wong et al. |
| Portes | Filipino | Other Asian | Wong et al. |
| Portilla | Filipino | Other Asian | Wong et al. |
| Portillo | Filipino | Other Asian | Wong et al. |
| Portugal | Filipino | Other Asian | Wong et al. |
| Portugues | Filipino | Other Asian | Wong et al. |
| Porwal | Asian Indian | South Asian | Wong et al. |
| Posadas | Filipino | Other Asian | Wong et al. |
| Posas | Filipino | Other Asian | Wong et al. |
| Poscablo | Filipino | Other Asian | Wong et al. |
| Posidio | Filipino | Other Asian | Wong et al. |
| Posis | Filipino | Other Asian | Wong et al. |
| Postadan | Filipino | Other Asian | Wong et al. |
| Potdar | Asian Indian | South Asian | Wong et al. |
| Potenciano | Filipino | Other Asian | Wong et al. |
| Potencion | Filipino | Other Asian | Wong et al. |
| Potente | Filipino | Other Asian | Wong et al. |
| Potes | Filipino | Other Asian | Wong et al. |
| Potha | Asian Indian | South Asian | Wong et al. |
| Pothan | Asian Indian | South Asian | Wong et al. |
| Pothen | Asian Indian | South Asian | Wong et al. |
| Pothuri | Asian Indian | South Asian | Kranick et al. |
| Potluri | Asian Indian | South Asian | Wong et al. |
| Potnis | Asian Indian | South Asian | Wong et al. |
| Poulose | Asian Indian | South Asian | Wong et al. |
| Pow | Chinese | Chinese | Wong et al. |
| Poy | Chinese | Chinese | Wong et al. |
| Poyaoan | Filipino | Other Asian | Wong et al. |
| Pozas | Filipino | Other Asian | Wong et al. |
| Pozon | Filipino | Other Asian | Wong et al. |
| Prabhakar | Asian Indian | South Asian | Wong et al. |
| Prabhakara | Asian Indian | South Asian | Wong et al. |
| Prabhakaran | Asian Indian | South Asian | Wong et al. |
| Prabhaker | Asian Indian | South Asian | Wong et al. |
| Prabhoo |  | South Asian | Lim et al. |
| Prabhu | Asian Indian | South Asian | Wong et al. |
| Prabhudas | Asian Indian | South Asian | Wong et al. |
| Prabhudesai | Asian Indian | South Asian | Wong et al. |
| Prades | Filipino | Other Asian | Wong et al. |
| Pradhan | Asian Indian | South Asian | Wong et al. |
| Prado | Filipino | Other Asian | Wong et al. |
| Prajapati | Asian Indian | South Asian | Wong et al. |
| Prakash | Asian Indian | South Asian | Kranick et al. |
| Pranada | Filipino | Other Asian | Wong et al. |
| Prasad | Asian Indian | South Asian | Kranick et al. |
| Prasadarao | Asian Indian | South Asian | Wong et al. |
| Prasai |  | South Asian | Lim et al. |
| Prashad | Asian Indian | South Asian | Kranick et al. |
| Prashadi |  | South Asian | Lim et al. |
| Prashar | Asian Indian | South Asian | Wong et al. |
| Prasla | Asian Indian | South Asian | Wong et al. |
| Pratap | Asian Indian | South Asian | Wong et al. |
| Pre | Filipino | Other Asian | Wong et al. |
| Preclaro | Filipino | Other Asian | Wong et al. |
| Premani | Asian Indian | South Asian | Wong et al. |
| Premi | Asian Indian | South Asian | Wong et al. |
| Premjee | Asian Indian | South Asian | Wong et al. |
| Premji | Asian Indian | South Asian | Wong et al. |
| Premwatee |  | South Asian | Lim et al. |
| Prepose | Filipino | Other Asian | Wong et al. |
| Presa | Filipino | Other Asian | Wong et al. |
| Presas | Filipino | Other Asian | Wong et al. |
| Presto | Filipino | Other Asian | Wong et al. |
| Prestosa | Filipino | Other Asian | Wong et al. |
| Prestoza | Filipino | Other Asian | Wong et al. |
| Priela | Filipino | Other Asian | Wong et al. |
| Prieto | Filipino | Other Asian | Wong et al. |
| Primero | Filipino | Other Asian | Wong et al. |
| Primicias | Filipino | Other Asian | Wong et al. |
| Primitivo | Filipino | Other Asian | Wong et al. |
| Prodigalidad | Filipino | Other Asian | Wong et al. |
| Promarak | Thai | Other Asian | Kranick et al. |
| Pronto | Filipino | Other Asian | Wong et al. |
| Prospero | Filipino | Other Asian | Wong et al. |
| Protacio | Filipino | Other Asian | Wong et al. |
| Provido | Filipino | Other Asian | Wong et al. |
| Prudencio | Filipino | Other Asian | Wong et al. |
| Prudente | Filipino | Other Asian | Wong et al. |
| Pruthi | Asian Indian | South Asian | Wong et al. |
| Ptel | Asian Indian | South Asian | Wong et al. |
| Pu | Chinese | Chinese | Wong et al. |
| Pua | Filipino | Other Asian | Wong et al. |
| Puangco | Filipino | Other Asian | Wong et al. |
| Puatu | Filipino | Other Asian | Wong et al. |
| Publico | Filipino | Other Asian | Wong et al. |
| Pucay | Filipino | Other Asian | Wong et al. |
| Pudiquet | Filipino | Other Asian | Wong et al. |
| Pueblo | Filipino | Other Asian | Wong et al. |
| Pueblos | Filipino | Other Asian | Wong et al. |
| Puen | Chinese | Chinese | Wong et al. |
| Puerto | Filipino | Other Asian | Wong et al. |
| Puertollano | Filipino | Other Asian | Wong et al. |
| Puey | Filipino | Other Asian | Wong et al. |
| Pueyo | Filipino | Other Asian | Wong et al. |
| Pugal | Filipino | Other Asian | Wong et al. |
| Pugay | Filipino | Other Asian | Wong et al. |
| Pugeda | Filipino | Other Asian | Wong et al. |
| Pui | Chinese | Chinese | Wong et al. |
| Pujalte | Filipino | Other Asian | Wong et al. |
| Pujara | Asian Indian | South Asian | Wong et al. |
| Pujari | Asian Indian | South Asian | Wong et al. |
| Pula | Filipino | Other Asian | Wong et al. |
| Pulanco | Filipino | Other Asian | Wong et al. |
| Pulchan |  | South Asian | Lim et al. |
| Pulgar | Filipino | Other Asian | Wong et al. |
| Pulido | Filipino | Other Asian | Kranick et al. |
| Pullido | Filipino | Other Asian | Wong et al. |
| Pulmano | Filipino | Other Asian | Wong et al. |
| Pulmones | Filipino | Other Asian | Wong et al. |
| Pulumbarit | Filipino | Other Asian | Wong et al. |
| Pumarada | Filipino | Other Asian | Wong et al. |
| Pumaras | Filipino | Other Asian | Wong et al. |
| Pumaren | Filipino | Other Asian | Wong et al. |
| Pun | Chinese | Chinese | Wong et al. |
| Punay | Filipino | Other Asian | Wong et al. |
| Pundavela | Filipino | Other Asian | Wong et al. |
| Pung | Chinese | Chinese | Wong et al. |
| Pungtilan | Filipino | Other Asian | Wong et al. |
| Punia | Asian Indian | South Asian | Wong et al. |
| Punj | Asian Indian | South Asian | Wong et al. |
| Punjani | Pakistani | South Asian | Kranick et al. |
| Punjwani | Asian Indian | South Asian | Wong et al. |
| Punla | Filipino | Other Asian | Wong et al. |
| Punn | Asian Indian | South Asian | Wong et al. |
| Punnoose | Asian Indian | South Asian | Wong et al. |
| Puno | Filipino | Other Asian | Wong et al. |
| Punongbayan | Filipino | Other Asian | Wong et al. |
| Punsal | Filipino | Other Asian | Wong et al. |
| Punsalan | Filipino | Other Asian | Wong et al. |
| Punsalang | Filipino | Other Asian | Wong et al. |
| Punzal | Filipino | Other Asian | Wong et al. |
| Punzalan | Filipino | Other Asian | Wong et al. |
| Pura | Filipino | Other Asian | Wong et al. |
| Purani | Asian Indian | South Asian | Wong et al. |
| Puray | Filipino | Other Asian | Wong et al. |
| Purba | Asian Indian | South Asian | Wong et al. |
| Pureuuel |  | South Asian | Lim et al. |
| Purewal | Asian Indian | South Asian | Wong et al. |
| Pureza | Filipino | Other Asian | Wong et al. |
| Purganan | Filipino | Other Asian | Wong et al. |
| Puri | Asian Indian | South Asian | Kranick et al. |
| Purificacion | Filipino | Other Asian | Wong et al. |
| Purisima | Filipino | Other Asian | Wong et al. |
| Purnima |  | South Asian | Lim et al. |
| Purohit | Asian Indian | South Asian | Wong et al. |
| Puruganan | Filipino | Other Asian | Wong et al. |
| Purugganan | Filipino | Other Asian | Wong et al. |
| Purushottam | Asian Indian | South Asian | Wong et al. |
| Puspo |  | South Asian | Lim et al. |
| Puthenparamp | Asian Indian | South Asian | Wong et al. |
| Puthenpurack | Asian Indian | South Asian | Wong et al. |
| Puthenpurayi | Asian Indian | South Asian | Wong et al. |
| Puthumana | Asian Indian | South Asian | Wong et al. |
| Puthuparampi | Asian Indian | South Asian | Wong et al. |
| Puttaswamy | Asian Indian | South Asian | Wong et al. |
| Putti | Asian Indian | South Asian | Kranick et al. |
| Puyaoan | Filipino | Other Asian | Wong et al. |
| Puyat | Filipino | Other Asian | Wong et al. |
| Puyot | Filipino | Other Asian | Wong et al. |
| Puzon | Filipino | Other Asian | Wong et al. |
| Pyo | Korean | Other Asian | Wong et al. |
| Pyon | Korean | Other Asian | Wong et al. |
| Pyong | Korean | Other Asian | Wong et al. |
| Pysadee |  | South Asian | Lim et al. |
| Pyun | Korean | Other Asian | Wong et al. |
| Pyung | Korean | Other Asian | Wong et al. |
| Qadeer | Asian Indian | South Asian | Wong et al. |
| Qader | Asian Indian | South Asian | Wong et al. |
| Qadir | Asian Indian | South Asian | Wong et al. |
| Qadri | Asian Indian | South Asian | Wong et al. |
| Qaiser | Asian Indian | South Asian | Wong et al. |
| Qasim | Pakistani | South Asian | Kranick et al. |
| Qayyum |  | South Asian | Lim et al. |
| Qazi |  | South Asian | Lim et al. |
| Qi | Chinese | Chinese | Kranick et al. |
| Qian | Chinese | Chinese | Kranick et al. |
| Qiang | Chinese | Chinese | Wong et al. |
| Qiao | Chinese | Chinese | Wong et al. |
| Qin | Chinese | Chinese | Wong et al. |
| Qing | Chinese | Chinese | Wong et al. |
| Qiu | Chinese | Chinese | Kranick et al. |
| Qu | Chinese | Chinese | Kranick et al. |
| Quach | Vietnamese | Other Asian | Wong et al. |
| Quadery |  | South Asian | Lim et al. |
| Quadir |  | South Asian | Lim et al. |
| Quadra | Filipino | Other Asian | Wong et al. |
| Quadri | Asian Indian | South Asian | Wong et al. |
| Quai | Chinese | Chinese | Wong et al. |
| Quaiyum |  | South Asian | Lim et al. |
| Quan | Chinese | Chinese | Kranick et al. |
| Quang | Vietnamese | Other Asian | Wong et al. |
| Quangvan | Vietnamese | Other Asian | Wong et al. |
| Quanico | Filipino | Other Asian | Wong et al. |
| Quant | Chinese | Chinese | Wong et al. |
| Quasem |  | South Asian | Lim et al. |
| Quat | Vietnamese | Other Asian | Wong et al. |
| Quazi |  | South Asian | Lim et al. |
| Qubra |  | South Asian | Lim et al. |
| Quddus |  | South Asian | Lim et al. |
| Quebada | Filipino | Other Asian | Wong et al. |
| Quebec | Filipino | Other Asian | Wong et al. |
| Quebral | Filipino | Other Asian | Wong et al. |
| Quedado | Filipino | Other Asian | Wong et al. |
| Queddeng | Filipino | Other Asian | Wong et al. |
| Quei | Chinese | Chinese | Wong et al. |
| Queja | Filipino | Other Asian | Wong et al. |
| Quejada | Filipino | Other Asian | Wong et al. |
| Quejado | Filipino | Other Asian | Wong et al. |
| Quelnan | Filipino | Other Asian | Wong et al. |
| Quema | Filipino | Other Asian | Wong et al. |
| Quemado | Filipino | Other Asian | Wong et al. |
| Quemuel | Filipino | Other Asian | Wong et al. |
| Quen | Chinese | Chinese | Wong et al. |
| Queral | Filipino | Other Asian | Wong et al. |
| Querido | Filipino | Other Asian | Wong et al. |
| Querijero | Filipino | Other Asian | Wong et al. |
| Querimit | Filipino | Other Asian | Wong et al. |
| Quero | Filipino | Other Asian | Wong et al. |
| Querol | Filipino | Other Asian | Wong et al. |
| Quershi | Asian Indian | South Asian | Wong et al. |
| Querubin | Filipino | Other Asian | Wong et al. |
| Quesada | Filipino | Other Asian | Wong et al. |
| Quevedo | Filipino | Other Asian | Wong et al. |
| Quey | Chinese | Chinese | Wong et al. |
| Queypo | Filipino | Other Asian | Wong et al. |
| Queyquep | Filipino | Other Asian | Wong et al. |
| Quezada | Filipino | Other Asian | Wong et al. |
| Quezon | Filipino | Other Asian | Wong et al. |
| Qui | Chinese | Chinese | Wong et al. |
| Quia | Filipino | Other Asian | Wong et al. |
| Quiachon | Filipino | Other Asian | Wong et al. |
| Quial | Filipino | Other Asian | Wong et al. |
| Quiambao | Filipino | Other Asian | Kranick et al. |
| Quiamco | Filipino | Other Asian | Wong et al. |
| Quiamzon | Filipino | Other Asian | Wong et al. |
| Quianio | Filipino | Other Asian | Wong et al. |
| Quiaoit | Filipino | Other Asian | Wong et al. |
| Quiaot | Filipino | Other Asian | Wong et al. |
| Quiapo | Filipino | Other Asian | Wong et al. |
| Quias | Filipino | Other Asian | Wong et al. |
| Quiason | Filipino | Other Asian | Wong et al. |
| Quiatchon | Filipino | Other Asian | Wong et al. |
| Quiazon | Filipino | Other Asian | Wong et al. |
| Quiba | Filipino | Other Asian | Wong et al. |
| Quibael | Filipino | Other Asian | Wong et al. |
| Quiban | Filipino | Other Asian | Wong et al. |
| Quibilan | Filipino | Other Asian | Wong et al. |
| Quibin | Filipino | Other Asian | Wong et al. |
| Quibuyen | Filipino | Other Asian | Wong et al. |
| Quicho | Filipino | Other Asian | Wong et al. |
| Quidato | Filipino | Other Asian | Wong et al. |
| Quiday | Filipino | Other Asian | Wong et al. |
| Quides | Filipino | Other Asian | Wong et al. |
| Quidilla | Filipino | Other Asian | Wong et al. |
| Quidlat | Filipino | Other Asian | Wong et al. |
| Quijada | Filipino | Other Asian | Wong et al. |
| Quijalvo | Filipino | Other Asian | Wong et al. |
| Quijano | Filipino | Other Asian | Kranick et al. |
| Quijote | Filipino | Other Asian | Wong et al. |
| Quilala | Filipino | Other Asian | Wong et al. |
| Quilalang | Filipino | Other Asian | Wong et al. |
| Quilang | Filipino | Other Asian | Wong et al. |
| Quilantang | Filipino | Other Asian | Wong et al. |
| Quilantip | Filipino | Other Asian | Wong et al. |
| Quilao | Filipino | Other Asian | Wong et al. |
| Quilatan | Filipino | Other Asian | Wong et al. |
| Quilaton | Filipino | Other Asian | Wong et al. |
| Quilenderino | Filipino | Other Asian | Wong et al. |
| Quiling | Filipino | Other Asian | Wong et al. |
| Quilit | Filipino | Other Asian | Wong et al. |
| Quiliza | Filipino | Other Asian | Wong et al. |
| Quillopo | Filipino | Other Asian | Wong et al. |
| Quilon | Filipino | Other Asian | Wong et al. |
| Quilop | Filipino | Other Asian | Wong et al. |
| Quimada | Filipino | Other Asian | Wong et al. |
| Quimado | Filipino | Other Asian | Wong et al. |
| Quimbo | Filipino | Other Asian | Wong et al. |
| Quimen | Filipino | Other Asian | Wong et al. |
| Quiming | Filipino | Other Asian | Wong et al. |
| Quimino | Filipino | Other Asian | Wong et al. |
| Quimosing | Filipino | Other Asian | Wong et al. |
| Quimoyog | Filipino | Other Asian | Wong et al. |
| Quimpo | Filipino | Other Asian | Wong et al. |
| Quimsing | Filipino | Other Asian | Wong et al. |
| Quimson | Filipino | Other Asian | Wong et al. |
| Quimzon | Filipino | Other Asian | Wong et al. |
| Quinabo | Filipino | Other Asian | Wong et al. |
| Quinagon | Filipino | Other Asian | Wong et al. |
| Quinal | Filipino | Other Asian | Wong et al. |
| Quinanola | Filipino | Other Asian | Wong et al. |
| Quincena | Filipino | Other Asian | Wong et al. |
| Quindara | Filipino | Other Asian | Wong et al. |
| Quindiagan | Filipino | Other Asian | Wong et al. |
| Quindipan | Filipino | Other Asian | Wong et al. |
| Quindoy | Filipino | Other Asian | Wong et al. |
| Quines | Filipino | Other Asian | Wong et al. |
| Quing | Chinese | Chinese | Wong et al. |
| Quinio | Filipino | Other Asian | Wong et al. |
| Quiniones | Filipino | Other Asian | Wong et al. |
| Quinit | Filipino | Other Asian | Wong et al. |
| Quino | Filipino | Other Asian | Wong et al. |
| Quinola | Filipino | Other Asian | Wong et al. |
| Quinones | Filipino | Other Asian | Wong et al. |
| Quinsaat | Filipino | Other Asian | Wong et al. |
| Quinsay | Filipino | Other Asian | Wong et al. |
| Quintana | Filipino | Other Asian | Wong et al. |
| Quintanilla | Filipino | Other Asian | Wong et al. |
| Quintans | Filipino | Other Asian | Wong et al. |
| Quintero | Filipino | Other Asian | Wong et al. |
| Quintia | Filipino | Other Asian | Wong et al. |
| Quinto | Filipino | Other Asian | Kranick et al. |
| Quintong | Filipino | Other Asian | Wong et al. |
| Quintos | Filipino | Other Asian | Wong et al. |
| Quintua | Filipino | Other Asian | Wong et al. |
| Quinzon | Filipino | Other Asian | Wong et al. |
| Quioc | Filipino | Other Asian | Wong et al. |
| Quiocho | Filipino | Other Asian | Wong et al. |
| Quiogue | Filipino | Other Asian | Wong et al. |
| Quion | Filipino | Other Asian | Wong et al. |
| Quirante | Filipino | Other Asian | Wong et al. |
| Quiray | Filipino | Other Asian | Wong et al. |
| Quirimit | Filipino | Other Asian | Wong et al. |
| Quirino | Filipino | Other Asian | Wong et al. |
| Quirit | Filipino | Other Asian | Wong et al. |
| Quiroga | Filipino | Other Asian | Wong et al. |
| Quirong | Filipino | Other Asian | Wong et al. |
| Quiros | Filipino | Other Asian | Wong et al. |
| Quiroz | Filipino | Other Asian | Wong et al. |
| Quisano | Filipino | Other Asian | Wong et al. |
| Quisao | Filipino | Other Asian | Wong et al. |
| Quisay | Filipino | Other Asian | Wong et al. |
| Quisisem | Filipino | Other Asian | Wong et al. |
| Quismorio | Filipino | Other Asian | Wong et al. |
| Quismundo | Filipino | Other Asian | Wong et al. |
| Quisol | Filipino | Other Asian | Wong et al. |
| Quisumbing | Filipino | Other Asian | Wong et al. |
| Quita | Filipino | Other Asian | Wong et al. |
| Quitaleg | Filipino | Other Asian | Wong et al. |
| Quitania | Filipino | Other Asian | Wong et al. |
| Quitasol | Filipino | Other Asian | Wong et al. |
| Quitazol | Filipino | Other Asian | Wong et al. |
| Quitevis | Filipino | Other Asian | Wong et al. |
| Quitilen | Filipino | Other Asian | Wong et al. |
| Quitiquit | Filipino | Other Asian | Wong et al. |
| Quitlong | Filipino | Other Asian | Wong et al. |
| Quito | Filipino | Other Asian | Wong et al. |
| Quiton | Filipino | Other Asian | Wong et al. |
| Quitoriano | Filipino | Other Asian | Wong et al. |
| Quitzon | Filipino | Other Asian | Wong et al. |
| Quiwa | Filipino | Other Asian | Wong et al. |
| Quiza | Filipino | Other Asian | Wong et al. |
| Quizon | Filipino | Other Asian | Wong et al. |
| Qun | Chinese | Chinese | Wong et al. |
| Qung | Chinese | Chinese | Wong et al. |
| Quo | Chinese | Chinese | Wong et al. |
| Quoc | Vietnamese | Other Asian | Wong et al. |
| Quock | Chinese | Chinese | Wong et al. |
| Quoi | Vietnamese | Other Asian | Wong et al. |
| Quon | Chinese | Chinese | Wong et al. |
| Quong | Chinese | Chinese | Wong et al. |
| Quoork | Chinese | Chinese | Wong et al. |
| Quraishi | Pakistani | South Asian | Kranick et al. |
| Qurashi |  | South Asian | Lim et al. |
| Qureishi | Asian Indian | South Asian | Wong et al. |
| Qureshi | Pakistani | South Asian | Kranick et al. |
| Qureshy | Asian Indian | South Asian | Wong et al. |
| Qutubuddin | Asian Indian | South Asian | Wong et al. |
| Quy | Vietnamese | Other Asian | Wong et al. |
| Quyen | Vietnamese | Other Asian | Wong et al. |
| Quynh | Vietnamese | Other Asian | Wong et al. |
| R | Asian Indian | South Asian | Wong et al. |
| Ra | Korean | Other Asian | Wong et al. |
| Raagas | Filipino | Other Asian | Wong et al. |
| Rab |  | South Asian | Lim et al. |
| Raba |  | South Asian | Lim et al. |
| Rabaca | Filipino | Other Asian | Wong et al. |
| Rabacal | Filipino | Other Asian | Wong et al. |
| Rabadia | Asian Indian | South Asian | Wong et al. |
| Rabago | Filipino | Other Asian | Wong et al. |
| Rabaino | Filipino | Other Asian | Wong et al. |
| Rabaja | Filipino | Other Asian | Wong et al. |
| Rabanal | Filipino | Other Asian | Wong et al. |
| Rabanera | Filipino | Other Asian | Wong et al. |
| Rabanes | Filipino | Other Asian | Wong et al. |
| Rabang | Filipino | Other Asian | Wong et al. |
| Rabano | Filipino | Other Asian | Wong et al. |
| Rabara | Filipino | Other Asian | Wong et al. |
| Rabaya | Filipino | Other Asian | Wong et al. |
| Rabbani |  | South Asian | Lim et al. |
| Rabbi |  | South Asian | Lim et al. |
| Rabbon | Filipino | Other Asian | Wong et al. |
| Rabby |  | South Asian | Lim et al. |
| Rabena | Filipino | Other Asian | Wong et al. |
| Rabina | Filipino | Other Asian | Wong et al. |
| Rabindranauth |  | South Asian | Lim et al. |
| Rabino | Filipino | Other Asian | Wong et al. |
| Rabo | Filipino | Other Asian | Wong et al. |
| Rabor | Filipino | Other Asian | Wong et al. |
| Rabot | Filipino | Other Asian | Wong et al. |
| Raboy | Filipino | Other Asian | Wong et al. |
| Rabuco | Filipino | Other Asian | Wong et al. |
| Rabulan | Filipino | Other Asian | Wong et al. |
| Rabut | Filipino | Other Asian | Wong et al. |
| Racadio | Filipino | Other Asian | Wong et al. |
| Racaza | Filipino | Other Asian | Wong et al. |
| Racca | Filipino | Other Asian | Wong et al. |
| Racela | Filipino | Other Asian | Wong et al. |
| Racelis | Filipino | Other Asian | Wong et al. |
| Racho | Filipino | Other Asian | Wong et al. |
| Racimo | Filipino | Other Asian | Wong et al. |
| Racines | Filipino | Other Asian | Wong et al. |
| Racinez | Filipino | Other Asian | Wong et al. |
| Racoma | Filipino | Other Asian | Wong et al. |
| Racoya | Filipino | Other Asian | Wong et al. |
| Racpan | Filipino | Other Asian | Wong et al. |
| Racsa | Filipino | Other Asian | Wong et al. |
| Racuya | Filipino | Other Asian | Wong et al. |
| Rada | Filipino | Other Asian | Wong et al. |
| Radadia | Asian Indian | South Asian | Wong et al. |
| Radam | Filipino | Other Asian | Wong et al. |
| Radaza | Filipino | Other Asian | Wong et al. |
| Radhakrishna | Asian Indian | South Asian | Wong et al. |
| Radia | Asian Indian | South Asian | Wong et al. |
| Radoc | Filipino | Other Asian | Wong et al. |
| Radona | Filipino | Other Asian | Wong et al. |
| Radovan | Filipino | Other Asian | Wong et al. |
| Rafael | Filipino | Other Asian | Wong et al. |
| Rafal | Filipino | Other Asian | Wong et al. |
| Rafanan | Filipino | Other Asian | Wong et al. |
| Rafer | Filipino | Other Asian | Wong et al. |
| Rafi | Asian Indian | South Asian | Wong et al. |
| Rafique |  | South Asian | Lim et al. |
| Rafiuddin | Asian Indian | South Asian | Wong et al. |
| Rafol | Filipino | Other Asian | Wong et al. |
| Rafols | Filipino | Other Asian | Wong et al. |
| Raga | Filipino | Other Asian | Wong et al. |
| Ragadio | Filipino | Other Asian | Wong et al. |
| Ragamat | Filipino | Other Asian | Wong et al. |
| Raganit | Filipino | Other Asian | Wong et al. |
| Ragasa | Filipino | Other Asian | Wong et al. |
| Ragat | Filipino | Other Asian | Wong et al. |
| Ragaza | Filipino | Other Asian | Wong et al. |
| Ragel | Filipino | Other Asian | Wong et al. |
| Raghava | Asian Indian | South Asian | Wong et al. |
| Raghavachar | Asian Indian | South Asian | Wong et al. |
| Raghavachari | Asian Indian | South Asian | Wong et al. |
| Raghavaiah | Asian Indian | South Asian | Wong et al. |
| Raghavan | Asian Indian | South Asian | Wong et al. |
| Raghunath | Asian Indian | South Asian | Kranick et al. |
| Ragmat | Filipino | Other Asian | Wong et al. |
| Ragonton | Filipino | Other Asian | Wong et al. |
| Ragoonanan |  | South Asian | Lim et al. |
| Ragos | Filipino | Other Asian | Wong et al. |
| Ragsac | Filipino | Other Asian | Wong et al. |
| Ragual | Filipino | Other Asian | Wong et al. |
| Ragudo | Filipino | Other Asian | Wong et al. |
| Ragudos | Filipino | Other Asian | Wong et al. |
| Raguindin | Filipino | Other Asian | Wong et al. |
| Raguine | Filipino | Other Asian | Wong et al. |
| Raguini | Filipino | Other Asian | Wong et al. |
| Ragunton | Filipino | Other Asian | Wong et al. |
| Raguro | Filipino | Other Asian | Wong et al. |
| Ragusante | Filipino | Other Asian | Wong et al. |
| Rah | Korean | Other Asian | Wong et al. |
| Raha | Asian Indian | South Asian | Wong et al. |
| Rahaman | Asian Indian | South Asian | Wong et al. |
| Raheem | Asian Indian | South Asian | Wong et al. |
| Rahi | Asian Indian | South Asian | Wong et al. |
| Rahim | Asian Indian | South Asian | Wong et al. |
| Rahima |  | South Asian | Lim et al. |
| Rahimtoola | Asian Indian | South Asian | Wong et al. |
| Rahman | Bangladeshi; Asian Indian; Pakistani; Indonesian | South Asian | Kranick et al. |
| Rahmani | Asian Indian | South Asian | Wong et al. |
| Rahmatullah | Asian Indian | South Asian | Wong et al. |
| Rai | Asian Indian; Nepali | South Asian | Kranick et al. |
| Raian |  | South Asian | Lim et al. |
| Raihan |  | South Asian | Lim et al. |
| Raina | Asian Indian | South Asian | Wong et al. |
| Raiz | Filipino | Other Asian | Wong et al. |
| Raizada | Asian Indian | South Asian | Wong et al. |
| Raj | Asian Indian; Bangladeshi | South Asian | Kranick et al. |
| Raja |  | South Asian | Lim et al. |
| Rajabali | Asian Indian | South Asian | Wong et al. |
| Rajagopal | Asian Indian | South Asian | Wong et al. |
| Rajagopalach | Asian Indian | South Asian | Wong et al. |
| Rajagopalan | Asian Indian | South Asian | Wong et al. |
| Rajaiah | Asian Indian | South Asian | Kranick et al. |
| Rajamani | Asian Indian | South Asian | Wong et al. |
| Rajan | Asian Indian | South Asian | Kranick et al. |
| Rajani | Asian Indian | South Asian | Wong et al. |
| Rajanna | Asian Indian | South Asian | Wong et al. |
| Rajappa | Asian Indian | South Asian | Wong et al. |
| Rajaram | Asian Indian | South Asian | Wong et al. |
| Rajarao | Asian Indian | South Asian | Wong et al. |
| Rajaratnam | Sri Lankan | South Asian | Kranick et al. |
| Rajbanshi | Asian Indian | South Asian | Kranick et al. |
| Rajbhandari |  | South Asian | Lim et al. |
| Raje | Asian Indian | South Asian | Wong et al. |
| Rajeev | Asian Indian; Bangladeshi | South Asian | Kranick et al. |
| Rajendran | Asian Indian | South Asian | Wong et al. |
| Rajgopal | Asian Indian | South Asian | Wong et al. |
| Rajguru | Asian Indian | South Asian | Wong et al. |
| Rajiv |  | South Asian | Lim et al. |
| Rajkumar | Asian Indian | South Asian | Kranick et al. |
| Rajkunar |  | South Asian | Lim et al. |
| Rajpal |  | South Asian | Lim et al. |
| Rajpat |  | South Asian | Lim et al. |
| Rajpaulsingh |  | South Asian | Lim et al. |
| Rajput | Pakistani; Asian Indian | South Asian | Kranick et al. |
| Raju | Asian Indian | South Asian | Wong et al. |
| Rajwani | Asian Indian | South Asian | Wong et al. |
| Rajwany | Asian Indian | South Asian | Wong et al. |
| Rala | Filipino | Other Asian | Wong et al. |
| Ralleta | Filipino | Other Asian | Wong et al. |
| Rallos | Filipino | Other Asian | Wong et al. |
| Ralutin | Filipino | Other Asian | Wong et al. |
| Ram |  | South Asian | Lim et al. |
| Rama | Filipino | Other Asian | Wong et al. |
| Ramac | Filipino | Other Asian | Wong et al. |
| Ramachandra | Asian Indian | South Asian | Wong et al. |
| Ramachandran | Asian Indian | South Asian | Wong et al. |
| Ramachandrar | Asian Indian | South Asian | Wong et al. |
| Ramadas | Asian Indian | South Asian | Wong et al. |
| Ramadhar | Asian Indian | South Asian | Kranick et al. |
| Ramaiah | Asian Indian | South Asian | Wong et al. |
| Ramaiyer | Asian Indian | South Asian | Wong et al. |
| Ramajo | Filipino | Other Asian | Wong et al. |
| Ramakrishna | Asian Indian | South Asian | Wong et al. |
| Ramakrishnan | Asian Indian | South Asian | Wong et al. |
| Ramales | Filipino | Other Asian | Wong et al. |
| Ramalingam | Asian Indian | South Asian | Wong et al. |
| Ramamoorthy | Asian Indian | South Asian | Kranick et al. |
| Ramamurthi | Asian Indian | South Asian | Wong et al. |
| Ramamurthy | Asian Indian | South Asian | Wong et al. |
| Ramamurti | Asian Indian | South Asian | Wong et al. |
| Raman | Asian Indian | South Asian | Wong et al. |
| Ramana | Asian Indian | South Asian | Wong et al. |
| Ramanathan | Asian Indian | South Asian | Kranick et al. |
| Ramani | Asian Indian | South Asian | Wong et al. |
| Ramanjulu | Asian Indian | South Asian | Wong et al. |
| Ramanlal | Asian Indian | South Asian | Wong et al. |
| Ramanna | Asian Indian | South Asian | Wong et al. |
| Ramanujam | Asian Indian | South Asian | Kranick et al. |
| Ramarao | Asian Indian | South Asian | Wong et al. |
| Ramarathnam | Asian Indian | South Asian | Wong et al. |
| Ramas | Filipino | Other Asian | Wong et al. |
| Ramasamy | Asian Indian | South Asian | Wong et al. |
| Ramasubraman | Asian Indian | South Asian | Wong et al. |
| Ramaswami | Asian Indian | South Asian | Wong et al. |
| Ramaswamy | Asian Indian | South Asian | Wong et al. |
| Ramat | Filipino | Other Asian | Wong et al. |
| Ramatalli |  | South Asian | Lim et al. |
| Ramawadh |  | South Asian | Lim et al. |
| Ramayya | Asian Indian | South Asian | Wong et al. |
| Rambac | Filipino | Other Asian | Wong et al. |
| Rambahal |  | South Asian | Lim et al. |
| Rambarran |  | South Asian | Lim et al. |
| Rambarrat |  | South Asian | Lim et al. |
| Rambaud | Filipino | Other Asian | Wong et al. |
| Rambayon | Filipino | Other Asian | Wong et al. |
| Rambhai | Asian Indian | South Asian | Wong et al. |
| Ramboyong | Filipino | Other Asian | Wong et al. |
| Ramchand | Asian Indian | South Asian | Wong et al. |
| Ramchandani | Asian Indian | South Asian | Kranick et al. |
| Ramchandran | Asian Indian | South Asian | Wong et al. |
| Ramcharit |  | South Asian | Lim et al. |
| Ramcharran |  | South Asian | Lim et al. |
| Ramdas | Asian Indian | South Asian | Kranick et al. |
| Ramdeen |  | South Asian | Lim et al. |
| Ramdeo |  | South Asian | Lim et al. |
| Ramdhan |  | South Asian | Lim et al. |
| Ramdhani | Indonesian | Other Asian | Kranick et al. |
| Ramdihal |  | South Asian | Lim et al. |
| Ramel | Filipino | Other Asian | Wong et al. |
| Ramelb | Filipino | Other Asian | Wong et al. |
| Ramento | Filipino | Other Asian | Wong et al. |
| Ramgobin |  | South Asian | Lim et al. |
| Ramgoolam |  | South Asian | Lim et al. |
| Ramgoonhinton |  | South Asian | Lim et al. |
| Rami | Asian Indian | South Asian | Wong et al. |
| Ramiah | Asian Indian | South Asian | Wong et al. |
| Ramil | Filipino | Other Asian | Wong et al. |
| Ramilla | Filipino | Other Asian | Wong et al. |
| Ramilo | Filipino | Other Asian | Wong et al. |
| Ramires | Filipino | Other Asian | Wong et al. |
| Ramirez | Filipino | Other Asian | Wong et al. |
| Ramiro | Filipino | Other Asian | Wong et al. |
| Ramis | Filipino | Other Asian | Wong et al. |
| Ramiscal | Filipino | Other Asian | Wong et al. |
| Ramjas |  | South Asian | Lim et al. |
| Ramjattan |  | South Asian | Lim et al. |
| Ramjeawan |  | South Asian | Lim et al. |
| Ramji | Asian Indian | South Asian | Wong et al. |
| Ramjohn |  | South Asian | Lim et al. |
| Ramkaran |  | South Asian | Lim et al. |
| Ramkhalawan |  | South Asian | Lim et al. |
| Ramkhalawen |  | South Asian | Lim et al. |
| Ramkissoon |  | South Asian | Lim et al. |
| Ramlagan |  | South Asian | Lim et al. |
| Ramlal | Asian Indian | South Asian | Wong et al. |
| Ramlall |  | South Asian | Lim et al. |
| Ramnanan |  | South Asian | Lim et al. |
| Ramnarace |  | South Asian | Lim et al. |
| Ramnarain |  | South Asian | Lim et al. |
| Ramnaraine |  | South Asian | Lim et al. |
| Ramnarine |  | South Asian | Lim et al. |
| Ramnath |  | South Asian | Lim et al. |
| Ramnauth |  | South Asian | Lim et al. |
| Ramo | Filipino | Other Asian | Wong et al. |
| Ramolete | Filipino | Other Asian | Wong et al. |
| Ramon | Filipino | Other Asian | Wong et al. |
| Ramones | Filipino | Other Asian | Kranick et al. |
| Ramoran | Filipino | Other Asian | Wong et al. |
| Ramos | Filipino | Other Asian | Wong et al. |
| Ramoso | Filipino | Other Asian | Wong et al. |
| Ramotar |  | South Asian | Lim et al. |
| Ramoutar |  | South Asian | Lim et al. |
| Ramowad |  | South Asian | Lim et al. |
| Rampal | Asian Indian | South Asian | Wong et al. |
| Rampersad |  | South Asian | Lim et al. |
| Rampersaud |  | South Asian | Lim et al. |
| Ramphal |  | South Asian | Lim et al. |
| Ramratan |  | South Asian | Lim et al. |
| Ramroo |  | South Asian | Lim et al. |
| Ramroop |  | South Asian | Lim et al. |
| Ramsackal |  | South Asian | Lim et al. |
| Ramsammy |  | South Asian | Lim et al. |
| Ramsaran |  | South Asian | Lim et al. |
| Ramsarran |  | South Asian | Lim et al. |
| Ramsarup |  | South Asian | Lim et al. |
| Ramsawak |  | South Asian | Lim et al. |
| Ramsingh |  | South Asian | Lim et al. |
| Ramsubhag |  | South Asian | Lim et al. |
| Ramsunder |  | South Asian | Lim et al. |
| Ran | Chinese | Chinese | Wong et al. |
| Rana | Pakistani | South Asian | Kranick et al. |
| Ranada | Filipino | Other Asian | Wong et al. |
| Ranade | Asian Indian | South Asian | Wong et al. |
| Ranadive | Asian Indian | South Asian | Wong et al. |
| Ranara | Filipino | Other Asian | Wong et al. |
| Ranario | Filipino | Other Asian | Wong et al. |
| Ranas | Filipino | Other Asian | Wong et al. |
| Ranat | Asian Indian | South Asian | Wong et al. |
| Ranay | Filipino | Other Asian | Wong et al. |
| Rances | Filipino | Other Asian | Wong et al. |
| Ranches | Filipino | Other Asian | Wong et al. |
| Ranchez | Filipino | Other Asian | Wong et al. |
| Ranchhodbhai | Asian Indian | South Asian | Wong et al. |
| Ranchod | Asian Indian | South Asian | Wong et al. |
| Rancudo | Filipino | Other Asian | Wong et al. |
| Randeria | Asian Indian | South Asian | Wong et al. |
| Randhawa | Asian Indian | South Asian | Wong et al. |
| Raneses | Filipino | Other Asian | Wong et al. |
| Ranga | Filipino | Other Asian | Wong et al. |
| Rangachar | Asian Indian | South Asian | Wong et al. |
| Rangachari | Asian Indian | South Asian | Wong et al. |
| Rangan | Asian Indian | South Asian | Wong et al. |
| Ranganathan | Asian Indian | South Asian | Wong et al. |
| Rangarajan | Asian Indian | South Asian | Wong et al. |
| Rangasammy |  | South Asian | Lim et al. |
| Rangasamy | Asian Indian | South Asian | Wong et al. |
| Rangaswami | Asian Indian | South Asian | Wong et al. |
| Rangaswamy | Asian Indian | South Asian | Wong et al. |
| Rangel | Filipino | Other Asian | Wong et al. |
| Rangi | Asian Indian | South Asian | Wong et al. |
| Rangnekar | Asian Indian | South Asian | Wong et al. |
| Rangoonwala | Asian Indian | South Asian | Wong et al. |
| Rangwala | Asian Indian | South Asian | Wong et al. |
| Rani | Asian Indian | South Asian | Kranick et al. |
| Rania | Filipino | Other Asian | Wong et al. |
| Ranido | Filipino | Other Asian | Wong et al. |
| Ranin | Filipino | Other Asian | Wong et al. |
| Ranjit | Asian Indian | South Asian | Wong et al. |
| Ranjo | Filipino | Other Asian | Wong et al. |
| Ranju |  | South Asian | Lim et al. |
| Ranoa | Filipino | Other Asian | Wong et al. |
| Ranola | Filipino | Other Asian | Kranick et al. |
| Ranon | Filipino | Other Asian | Wong et al. |
| Rante | Filipino | Other Asian | Wong et al. |
| Rao | Chinese; Indian; Pakistani | Chinese | Kranick et al. |
| Raojibhai | Asian Indian | South Asian | Wong et al. |
| Raol | Asian Indian | South Asian | Wong et al. |
| Raoof | Asian Indian | South Asian | Wong et al. |
| Raouf |  | South Asian | Lim et al. |
| Rapacon | Filipino | Other Asian | Wong et al. |
| Rapada | Filipino | Other Asian | Wong et al. |
| Rapadas | Filipino | Other Asian | Wong et al. |
| Rapanan | Filipino | Other Asian | Wong et al. |
| Rapanot | Filipino | Other Asian | Wong et al. |
| Rapanut | Filipino | Other Asian | Wong et al. |
| Rapinan | Filipino | Other Asian | Wong et al. |
| Rapisura | Filipino | Other Asian | Wong et al. |
| Rapiz | Filipino | Other Asian | Wong et al. |
| Raposas | Filipino | Other Asian | Wong et al. |
| Raquedan | Filipino | Other Asian | Wong et al. |
| Raquel | Filipino | Other Asian | Wong et al. |
| Raqueno | Filipino | Other Asian | Wong et al. |
| Raquepo | Filipino | Other Asian | Wong et al. |
| Raquidan | Filipino | Other Asian | Wong et al. |
| Raquindin | Filipino | Other Asian | Wong et al. |
| Raquinio | Filipino | Other Asian | Wong et al. |
| Raquino | Filipino | Other Asian | Wong et al. |
| Raquipiso | Filipino | Other Asian | Wong et al. |
| Raquiza | Filipino | Other Asian | Wong et al. |
| Rara | Filipino | Other Asian | Wong et al. |
| Rarama | Filipino | Other Asian | Wong et al. |
| Rarang | Filipino | Other Asian | Wong et al. |
| Rarangol | Filipino | Other Asian | Wong et al. |
| Raras | Filipino | Other Asian | Wong et al. |
| Rarela | Filipino | Other Asian | Wong et al. |
| Rarogal | Filipino | Other Asian | Wong et al. |
| Raroque | Filipino | Other Asian | Wong et al. |
| Ras | Filipino | Other Asian | Wong et al. |
| Rasalan | Filipino | Other Asian | Kranick et al. |
| Rasay | Filipino | Other Asian | Wong et al. |
| Rasel |  | South Asian | Lim et al. |
| Rashed |  | South Asian | Lim et al. |
| Rasheed |  | South Asian | Lim et al. |
| Rashid | Bangladeshi; Pakistani | South Asian | Kranick et al. |
| Rasid |  | South Asian | Lim et al. |
| Rasiklal | Asian Indian | South Asian | Wong et al. |
| Rasing | Filipino | Other Asian | Wong et al. |
| Rasmi |  | South Asian | Lim et al. |
| Rasool | Pakistani | South Asian | Kranick et al. |
| Rasos | Filipino | Other Asian | Wong et al. |
| Raspado | Filipino | Other Asian | Wong et al. |
| Rasquinha | Asian Indian | South Asian | Wong et al. |
| Rastogi | Asian Indian | South Asian | Wong et al. |
| Rasuda |  | South Asian | Lim et al. |
| Rasul |  | South Asian | Lim et al. |
| Rat | Vietnamese | Other Asian | Wong et al. |
| Ratansi | Asian Indian | South Asian | Wong et al. |
| Raterta | Filipino | Other Asian | Kranick et al. |
| Rathi | Asian Indian | South Asian | Wong et al. |
| Rathnam | Asian Indian | South Asian | Wong et al. |
| Rathod | Asian Indian | South Asian | Kranick et al. |
| Rathor | Asian Indian | South Asian | Wong et al. |
| Rathore | Asian Indian | South Asian | Wong et al. |
| Ratilal | Asian Indian | South Asian | Wong et al. |
| Ratilla | Filipino | Other Asian | Wong et al. |
| Ratnam | Asian Indian | South Asian | Wong et al. |
| Ratnani | Asian Indian | South Asian | Wong et al. |
| Raton |  | South Asian | Lim et al. |
| Ratonel | Filipino | Other Asian | Wong et al. |
| Ratuita | Filipino | Other Asian | Wong et al. |
| Ratunil | Filipino | Other Asian | Wong et al. |
| Rauf | Pakistani | South Asian | Kranick et al. |
| Rausa | Filipino | Other Asian | Wong et al. |
| Raut | Asian Indian | South Asian | Wong et al. |
| Ravago | Filipino | Other Asian | Wong et al. |
| Raval | Asian Indian | South Asian | Wong et al. |
| Ravalo | Filipino | Other Asian | Wong et al. |
| Ravana | Filipino | Other Asian | Wong et al. |
| Ravancho | Filipino | Other Asian | Wong et al. |
| Ravanes | Filipino | Other Asian | Wong et al. |
| Ravani | Asian Indian | South Asian | Wong et al. |
| Ravara | Filipino | Other Asian | Wong et al. |
| Ravarra | Filipino | Other Asian | Wong et al. |
| Ravel | Asian Indian | South Asian | Wong et al. |
| Ravello | Filipino | Other Asian | Wong et al. |
| Ravelo | Filipino | Other Asian | Wong et al. |
| Ravi | Asian Indian | South Asian | Wong et al. |
| Ravichandran | Asian Indian | South Asian | Kranick et al. |
| Ravina | Filipino | Other Asian | Wong et al. |
| Ravindran | Asian Indian | South Asian | Wong et al. |
| Ravindranath | Asian Indian | South Asian | Wong et al. |
| Ravji | Asian Indian | South Asian | Wong et al. |
| Rawal | Asian Indian | South Asian | Wong et al. |
| Rawat | Asian Indian | South Asian | Wong et al. |
| Rawjee | Asian Indian | South Asian | Wong et al. |
| Rawji | Asian Indian | South Asian | Wong et al. |
| Raya | Filipino | Other Asian | Wong et al. |
| Rayala | Filipino | Other Asian | Wong et al. |
| Rayani | Asian Indian | South Asian | Wong et al. |
| Raychaudhuri | Asian Indian | South Asian | Wong et al. |
| Rayco | Filipino | Other Asian | Wong et al. |
| Rayman |  | South Asian | Lim et al. |
| Raymond |  | South Asian | Lim et al. |
| Raymondo | Filipino | Other Asian | Wong et al. |
| Raymundo | Filipino | Other Asian | Kranick et al. |
| Rayo | Filipino | Other Asian | Wong et al. |
| Rayos | Filipino | Other Asian | Wong et al. |
| Rayray | Filipino | Other Asian | Wong et al. |
| Raz | Bangladeshi | South Asian | Kranick et al. |
| Raza | Pakistani | South Asian | Kranick et al. |
| Razack | Asian Indian | South Asian | Wong et al. |
| Razak |  | South Asian | Lim et al. |
| Razal | Filipino | Other Asian | Wong et al. |
| Razalan | Filipino | Other Asian | Wong et al. |
| Razdan | Asian Indian | South Asian | Wong et al. |
| Razia |  | South Asian | Lim et al. |
| Razinoor |  | South Asian | Lim et al. |
| Razo | Filipino | Other Asian | Kranick et al. |
| Razon | Filipino | Other Asian | Wong et al. |
| Razonable | Filipino | Other Asian | Wong et al. |
| Razote | Filipino | Other Asian | Wong et al. |
| Razvi | Asian Indian | South Asian | Wong et al. |
| Razzack | Asian Indian | South Asian | Wong et al. |
| Razzak | Bangladeshi; Pakistani | South Asian | Kranick et al. |
| Razzaq | Pakistani | South Asian | Kranick et al. |
| Razzaque |  | South Asian | Lim et al. |
| Real | Filipino | Other Asian | Wong et al. |
| Reales | Filipino | Other Asian | Wong et al. |
| Realica | Filipino | Other Asian | Wong et al. |
| Realin | Filipino | Other Asian | Wong et al. |
| Realista | Filipino | Other Asian | Wong et al. |
| Realiza | Filipino | Other Asian | Wong et al. |
| Realubit | Filipino | Other Asian | Wong et al. |
| Reano | Filipino | Other Asian | Wong et al. |
| Reantaso | Filipino | Other Asian | Wong et al. |
| Reasonda | Filipino | Other Asian | Wong et al. |
| Reaz |  | South Asian | Lim et al. |
| Rebanal | Filipino | Other Asian | Wong et al. |
| Rebancos | Filipino | Other Asian | Wong et al. |
| Rebantad |  | South Asian | Lim et al. |
| Rebeiro | Asian Indian | South Asian | Wong et al. |
| Rebibis | Filipino | Other Asian | Wong et al. |
| Reblando | Filipino | Other Asian | Wong et al. |
| Reboja | Filipino | Other Asian | Wong et al. |
| Reboldela | Filipino | Other Asian | Wong et al. |
| Rebolledo | Filipino | Other Asian | Wong et al. |
| Rebollido | Filipino | Other Asian | Wong et al. |
| Rebong | Filipino | Other Asian | Wong et al. |
| Rebosura | Filipino | Other Asian | Wong et al. |
| Rebucas | Filipino | Other Asian | Wong et al. |
| Rebueno | Filipino | Other Asian | Wong et al. |
| Rebugio | Filipino | Other Asian | Wong et al. |
| Rebujio | Filipino | Other Asian | Wong et al. |
| Rebuldela | Filipino | Other Asian | Wong et al. |
| Rebullar | Filipino | Other Asian | Wong et al. |
| Rebultan | Filipino | Other Asian | Wong et al. |
| Reburiano | Filipino | Other Asian | Wong et al. |
| Recaido | Filipino | Other Asian | Wong et al. |
| Recalde | Filipino | Other Asian | Wong et al. |
| Recato | Filipino | Other Asian | Wong et al. |
| Recile | Filipino | Other Asian | Wong et al. |
| Recinto | Filipino | Other Asian | Wong et al. |
| Recio | Filipino | Other Asian | Wong et al. |
| Reclosado | Filipino | Other Asian | Wong et al. |
| Reclusado | Filipino | Other Asian | Wong et al. |
| Recodo | Filipino | Other Asian | Wong et al. |
| Recometa | Filipino | Other Asian | Wong et al. |
| Recreo | Filipino | Other Asian | Wong et al. |
| Recto | Filipino | Other Asian | Wong et al. |
| Rectra | Filipino | Other Asian | Wong et al. |
| Redaja | Filipino | Other Asian | Wong et al. |
| Redaus | Filipino | Other Asian | Wong et al. |
| Reddi | Asian Indian | South Asian | Wong et al. |
| Redila | Filipino | Other Asian | Wong et al. |
| Redito | Filipino | Other Asian | Wong et al. |
| Redoblado | Filipino | Other Asian | Wong et al. |
| Redoble | Filipino | Other Asian | Wong et al. |
| Redolme | Filipino | Other Asian | Wong et al. |
| Redona | Filipino | Other Asian | Wong et al. |
| Redondo | Filipino | Other Asian | Wong et al. |
| Redor | Filipino | Other Asian | Wong et al. |
| Redulla | Filipino | Other Asian | Wong et al. |
| Reduque | Filipino | Other Asian | Wong et al. |
| Ree | Korean | Other Asian | Wong et al. |
| Reehal | Asian Indian | South Asian | Wong et al. |
| Reforma | Filipino | Other Asian | Wong et al. |
| Reformado | Filipino | Other Asian | Wong et al. |
| Refuerzo | Filipino | Other Asian | Wong et al. |
| Refugio | Filipino | Other Asian | Wong et al. |
| Reg | Filipino | Other Asian | Wong et al. |
| Regacho | Filipino | Other Asian | Wong et al. |
| Regala | Filipino | Other Asian | Wong et al. |
| Regalado | Filipino | Other Asian | Wong et al. |
| Reganit | Filipino | Other Asian | Wong et al. |
| Regaspi | Filipino | Other Asian | Wong et al. |
| Regencia | Filipino | Other Asian | Wong et al. |
| Regidor | Filipino | Other Asian | Wong et al. |
| Reginald |  | South Asian | Lim et al. |
| Reginaldo | Filipino | Other Asian | Wong et al. |
| Regines | Filipino | Other Asian | Wong et al. |
| Regino | Filipino | Other Asian | Wong et al. |
| Regio | Filipino | Other Asian | Wong et al. |
| Reglos | Filipino | Other Asian | Wong et al. |
| Regondola | Filipino | Other Asian | Wong et al. |
| Regua | Filipino | Other Asian | Wong et al. |
| Regualos | Filipino | Other Asian | Wong et al. |
| Reguindin | Filipino | Other Asian | Wong et al. |
| Rehana |  | South Asian | Lim et al. |
| Rehman | Pakistani | South Asian | Kranick et al. |
| Rehmani | Asian Indian | South Asian | Wong et al. |
| Rehmatullah | Asian Indian | South Asian | Wong et al. |
| Reinoso | Filipino | Other Asian | Wong et al. |
| Reintegrado | Filipino | Other Asian | Wong et al. |
| Reja |  | South Asian | Lim et al. |
| Rejano | Filipino | Other Asian | Wong et al. |
| Rejuso | Filipino | Other Asian | Wong et al. |
| Relampagos | Filipino | Other Asian | Wong et al. |
| Relano | Filipino | Other Asian | Wong et al. |
| Relayson | Filipino | Other Asian | Wong et al. |
| Relente | Filipino | Other Asian | Wong et al. |
| Relevo | Filipino | Other Asian | Wong et al. |
| Religioso | Filipino | Other Asian | Wong et al. |
| Rellamas | Filipino | Other Asian | Wong et al. |
| Rellin | Filipino | Other Asian | Wong et al. |
| Rellora | Filipino | Other Asian | Wong et al. |
| Rellosa | Filipino | Other Asian | Wong et al. |
| Reloj | Filipino | Other Asian | Wong et al. |
| Relos | Filipino | Other Asian | Wong et al. |
| Relosa | Filipino | Other Asian | Wong et al. |
| Relova | Filipino | Other Asian | Wong et al. |
| Relucio | Filipino | Other Asian | Wong et al. |
| Remedio | Filipino | Other Asian | Wong et al. |
| Remedios | Chinese | Chinese | Wong et al. |
| Rementilla | Filipino | Other Asian | Wong et al. |
| Remigio | Filipino | Other Asian | Wong et al. |
| Remo | Filipino | Other Asian | Wong et al. |
| Remolacio | Filipino | Other Asian | Wong et al. |
| Remolador | Filipino | Other Asian | Wong et al. |
| Remollino | Filipino | Other Asian | Wong et al. |
| Remolona | Filipino | Other Asian | Wong et al. |
| Remonte | Filipino | Other Asian | Wong et al. |
| Remorin | Filipino | Other Asian | Wong et al. |
| Remos | Filipino | Other Asian | Wong et al. |
| Remotigue | Filipino | Other Asian | Wong et al. |
| Rempillo | Filipino | Other Asian | Wong et al. |
| Remular | Filipino | Other Asian | Wong et al. |
| Remulla | Filipino | Other Asian | Wong et al. |
| Ren | Chinese | Chinese | Kranick et al. |
| Rendal | Filipino | Other Asian | Wong et al. |
| Rendon | Filipino | Other Asian | Wong et al. |
| Renegado | Filipino | Other Asian | Wong et al. |
| Rengasamy | Asian Indian | South Asian | Wong et al. |
| Renojo | Filipino | Other Asian | Wong et al. |
| Renon | Filipino | Other Asian | Wong et al. |
| Rentino | Filipino | Other Asian | Wong et al. |
| Rentoria | Filipino | Other Asian | Wong et al. |
| Reodica | Filipino | Other Asian | Wong et al. |
| Reodique | Filipino | Other Asian | Wong et al. |
| Reola | Filipino | Other Asian | Wong et al. |
| Reolada | Filipino | Other Asian | Wong et al. |
| Reonal | Filipino | Other Asian | Wong et al. |
| Reototar | Filipino | Other Asian | Wong et al. |
| Repaso | Filipino | Other Asian | Wong et al. |
| Repato | Filipino | Other Asian | Wong et al. |
| Repaul |  | South Asian | Lim et al. |
| Repiedad | Filipino | Other Asian | Wong et al. |
| Repique | Filipino | Other Asian | Wong et al. |
| Repollo | Filipino | Other Asian | Wong et al. |
| Reponte | Filipino | Other Asian | Wong et al. |
| Repunte | Filipino | Other Asian | Wong et al. |
| Requejo | Filipino | Other Asian | Wong et al. |
| Requierme | Filipino | Other Asian | Wong et al. |
| Requilman | Filipino | Other Asian | Wong et al. |
| Requina | Filipino | Other Asian | Wong et al. |
| Reroma | Filipino | Other Asian | Wong et al. |
| Rescober | Filipino | Other Asian | Wong et al. |
| Reshamwala | Asian Indian | South Asian | Wong et al. |
| Reside | Filipino | Other Asian | Wong et al. |
| Resma | Filipino | Other Asian | Wong et al. |
| Resngit | Filipino | Other Asian | Wong et al. |
| Resolme | Filipino | Other Asian | Wong et al. |
| Respicio | Filipino | Other Asian | Wong et al. |
| Resplandor | Filipino | Other Asian | Wong et al. |
| Restauro | Filipino | Other Asian | Wong et al. |
| Restua | Filipino | Other Asian | Wong et al. |
| Resuello | Filipino | Other Asian | Wong et al. |
| Resultay | Filipino | Other Asian | Wong et al. |
| Resuma | Filipino | Other Asian | Wong et al. |
| Resurreccion | Filipino | Other Asian | Wong et al. |
| Resurrecion | Filipino | Other Asian | Wong et al. |
| Resus | Filipino | Other Asian | Wong et al. |
| Reta | Filipino | Other Asian | Wong et al. |
| Retiro | Filipino | Other Asian | Wong et al. |
| Retuta | Filipino | Other Asian | Wong et al. |
| Retuya | Filipino | Other Asian | Wong et al. |
| Reuben | Asian Indian | South Asian | Wong et al. |
| Revankar | Asian Indian | South Asian | Wong et al. |
| Reveche | Filipino | Other Asian | Wong et al. |
| Revelo | Filipino | Other Asian | Wong et al. |
| Revera | Filipino | Other Asian | Wong et al. |
| Revereza | Filipino | Other Asian | Wong et al. |
| Revil | Filipino | Other Asian | Wong et al. |
| Revilla | Filipino | Other Asian | Wong et al. |
| Revilleza | Filipino | Other Asian | Wong et al. |
| Revita | Filipino | Other Asian | Wong et al. |
| Revives | Filipino | Other Asian | Wong et al. |
| Rey | Filipino | Other Asian | Wong et al. |
| Reyes | Filipino | Other Asian | Wong et al. |
| Reyesreyes | Filipino | Other Asian | Wong et al. |
| Reyla | Filipino | Other Asian | Wong et al. |
| Reyles | Filipino | Other Asian | Wong et al. |
| Reymundo | Filipino | Other Asian | Wong et al. |
| Reyna | Filipino | Other Asian | Wong et al. |
| Reynado | Filipino | Other Asian | Wong et al. |
| Reynaldo | Filipino | Other Asian | Wong et al. |
| Reynante | Filipino | Other Asian | Wong et al. |
| Reynera | Filipino | Other Asian | Wong et al. |
| Reynes | Filipino | Other Asian | Wong et al. |
| Reyno | Filipino | Other Asian | Wong et al. |
| Reynon | Filipino | Other Asian | Wong et al. |
| Reynoso | Filipino | Other Asian | Wong et al. |
| Reyta | Filipino | Other Asian | Wong et al. |
| Reza | Asian Indian | South Asian | Wong et al. |
| Rezauddula |  | South Asian | Lim et al. |
| Rha | Korean | Other Asian | Wong et al. |
| Rhaman |  | South Asian | Lim et al. |
| Rhee | Korean | Other Asian | Kranick et al. |
| Rhie | Korean | Other Asian | Wong et al. |
| Rhim | Korean | Other Asian | Wong et al. |
| Rhiman |  | South Asian | Lim et al. |
| Rho | Korean | Other Asian | Wong et al. |
| Rhyu | Korean | Other Asian | Wong et al. |
| Riad |  | South Asian | Lim et al. |
| Riano | Filipino | Other Asian | Wong et al. |
| Riar | Asian Indian | South Asian | Wong et al. |
| Riaz | Pakistani; Bangladeshi | South Asian | Kranick et al. |
| Ribac | Filipino | Other Asian | Wong et al. |
| Ribao | Filipino | Other Asian | Wong et al. |
| Ribay | Filipino | Other Asian | Wong et al. |
| Ribaya | Filipino | Other Asian | Wong et al. |
| Riberal | Filipino | Other Asian | Wong et al. |
| Ribuca | Filipino | Other Asian | Wong et al. |
| Ricafort | Filipino | Other Asian | Wong et al. |
| Ricaforte | Filipino | Other Asian | Wong et al. |
| Ricafrente | Filipino | Other Asian | Wong et al. |
| Ricalde | Filipino | Other Asian | Wong et al. |
| Ricana | Filipino | Other Asian | Wong et al. |
| Ricardo | Filipino | Other Asian | Wong et al. |
| Ricardos | Filipino | Other Asian | Wong et al. |
| Ricarte | Filipino | Other Asian | Wong et al. |
| Ricasa | Filipino | Other Asian | Wong et al. |
| Ricasata | Filipino | Other Asian | Wong et al. |
| Rico | Filipino | Other Asian | Wong et al. |
| Ricohermoso | Filipino | Other Asian | Wong et al. |
| Ricolcol | Filipino | Other Asian | Wong et al. |
| Ridad | Filipino | Other Asian | Wong et al. |
| Ridao | Filipino | Other Asian | Wong et al. |
| Ridon | Filipino | Other Asian | Wong et al. |
| Riego | Filipino | Other Asian | Wong et al. |
| Riegodedio | Filipino | Other Asian | Wong et al. |
| Riegodedios | Filipino | Other Asian | Wong et al. |
| Rieta | Filipino | Other Asian | Wong et al. |
| Rieza | Filipino | Other Asian | Wong et al. |
| Rifareal | Filipino | Other Asian | Wong et al. |
| Rifat |  | South Asian | Lim et al. |
| Rigodon | Filipino | Other Asian | Wong et al. |
| Rigonan | Filipino | Other Asian | Wong et al. |
| Rigor | Filipino | Other Asian | Wong et al. |
| Rigos | Filipino | Other Asian | Wong et al. |
| Rigunay | Filipino | Other Asian | Wong et al. |
| Rihal | Asian Indian | South Asian | Wong et al. |
| Rikhi | Asian Indian | South Asian | Wong et al. |
| Rikhy |  | South Asian | Lim et al. |
| Rikitake | Japanese | Other Asian | Wong et al. |
| Rili | Filipino | Other Asian | Wong et al. |
| Rillamas | Filipino | Other Asian | Wong et al. |
| Rillera | Filipino | Other Asian | Wong et al. |
| Rillo | Filipino | Other Asian | Wong et al. |
| Rillon | Filipino | Other Asian | Wong et al. |
| Rilloraza | Filipino | Other Asian | Wong et al. |
| Rillorta | Filipino | Other Asian | Wong et al. |
| Rilveria | Filipino | Other Asian | Wong et al. |
| Rim | Korean | Other Asian | Kranick et al. |
| Rimando | Filipino | Other Asian | Wong et al. |
| Rimas | Filipino | Other Asian | Wong et al. |
| Rimorin | Filipino | Other Asian | Wong et al. |
| Rina | Asian Indian; Indonesian | South Asian | Kranick et al. |
| Rinen | Filipino | Other Asian | Wong et al. |
| Ringor | Filipino | Other Asian | Wong et al. |
| Rinon | Filipino | Other Asian | Wong et al. |
| Rinonos | Filipino | Other Asian | Wong et al. |
| Rinosa | Filipino | Other Asian | Wong et al. |
| Rint | Filipino | Other Asian | Wong et al. |
| Riola | Filipino | Other Asian | Wong et al. |
| Riopta | Filipino | Other Asian | Wong et al. |
| Rios | Filipino | Other Asian | Wong et al. |
| Riosa | Filipino | Other Asian | Wong et al. |
| Ripalda | Filipino | Other Asian | Wong et al. |
| Riparip | Filipino | Other Asian | Wong et al. |
| Ripon |  | South Asian | Lim et al. |
| Ripoyla | Filipino | Other Asian | Wong et al. |
| Rishi | Asian Indian | South Asian | Kranick et al. |
| Ritarita | Filipino | Other Asian | Wong et al. |
| Rito | Filipino | Other Asian | Wong et al. |
| Ritualo | Filipino | Other Asian | Wong et al. |
| Ritumalta | Filipino | Other Asian | Wong et al. |
| Riu | Korean | Other Asian | Wong et al. |
| Rivada | Filipino | Other Asian | Wong et al. |
| Rivadelo | Filipino | Other Asian | Wong et al. |
| Rivamonte | Filipino | Other Asian | Wong et al. |
| Rivas |  | South Asian | Lim et al. |
| Rivera | Filipino | Other Asian | Wong et al. |
| Riveral | Filipino | Other Asian | Wong et al. |
| Rivero | Filipino | Other Asian | Wong et al. |
| Rivetna | Asian Indian | South Asian | Wong et al. |
| Rivo | Filipino | Other Asian | Wong et al. |
| Riyadh | Bangladeshi | South Asian | Kranick et al. |
| Rizal | Filipino | Other Asian | Wong et al. |
| Rizo | Filipino | Other Asian | Wong et al. |
| Rizon | Filipino | Other Asian | Wong et al. |
| Rizui | Asian Indian | South Asian | Wong et al. |
| Rizvi | Asian Indian | South Asian | Wong et al. |
| Rizwan |  | South Asian | Lim et al. |
| Ro | Korean | Other Asian | Wong et al. |
| Roa | Filipino | Other Asian | Wong et al. |
| Roallos | Filipino | Other Asian | Wong et al. |
| Roaquin | Filipino | Other Asian | Wong et al. |
| Roasa | Filipino | Other Asian | Wong et al. |
| Rob |  | South Asian | Lim et al. |
| Robancho | Filipino | Other Asian | Wong et al. |
| Robbani |  | South Asian | Lim et al. |
| Robeniol | Filipino | Other Asian | Wong et al. |
| Robi |  | South Asian | Lim et al. |
| Robianes | Filipino | Other Asian | Wong et al. |
| Robillos | Filipino | Other Asian | Wong et al. |
| Robin |  | South Asian | Lim et al. |
| Robina | Filipino | Other Asian | Wong et al. |
| Robino | Filipino | Other Asian | Wong et al. |
| Robinol | Filipino | Other Asian | Wong et al. |
| Robinos | Filipino | Other Asian | Wong et al. |
| Roble | Filipino | Other Asian | Wong et al. |
| Robledo | Filipino | Other Asian | Wong et al. |
| Robles | Filipino | Other Asian | Wong et al. |
| Robleza | Filipino | Other Asian | Wong et al. |
| Robrigado | Filipino | Other Asian | Wong et al. |
| Roc | Filipino | Other Asian | Wong et al. |
| Roca | Filipino | Other Asian | Wong et al. |
| Rocaberte | Filipino | Other Asian | Wong et al. |
| Rocabo | Filipino | Other Asian | Wong et al. |
| Rocamora | Filipino | Other Asian | Wong et al. |
| Rocero | Filipino | Other Asian | Wong et al. |
| Roces | Filipino | Other Asian | Wong et al. |
| Rocha | Filipino | Other Asian | Wong et al. |
| Rocillo | Filipino | Other Asian | Wong et al. |
| Rocket |  | South Asian | Lim et al. |
| Roco | Filipino | Other Asian | Wong et al. |
| Rodas | Filipino | Other Asian | Wong et al. |
| Rodelas | Filipino | Other Asian | Wong et al. |
| Rodenas | Filipino | Other Asian | Wong et al. |
| Roderos | Filipino | Other Asian | Wong et al. |
| Rodica | Filipino | Other Asian | Wong et al. |
| Rodil | Filipino | Other Asian | Wong et al. |
| Rodillas | Filipino | Other Asian | Wong et al. |
| Rodis | Filipino | Other Asian | Wong et al. |
| Rodolfo | Filipino | Other Asian | Wong et al. |
| Rodricks | Asian Indian | South Asian | Wong et al. |
| Rodrigo | Filipino | Other Asian | Wong et al. |
| Rodriguera | Filipino | Other Asian | Wong et al. |
| Rodrigues | Asian Indian | South Asian | Wong et al. |
| Rodriguez | Filipino | Other Asian | Wong et al. |
| Rodriques | Asian Indian | South Asian | Wong et al. |
| Rodriquez | Filipino | Other Asian | Wong et al. |
| Rodulfa | Filipino | Other Asian | Wong et al. |
| Rodulfo | Filipino | Other Asian | Wong et al. |
| Rofiquzzaman |  | South Asian | Lim et al. |
| Rogacion | Filipino | Other Asian | Wong et al. |
| Rogado | Filipino | Other Asian | Wong et al. |
| Rogayan | Filipino | Other Asian | Wong et al. |
| Rogel | Filipino | Other Asian | Wong et al. |
| Rogue | Filipino | Other Asian | Wong et al. |
| Roh | Korean | Other Asian | Wong et al. |
| Rohatgi | Asian Indian | South Asian | Wong et al. |
| Rohila | Asian Indian | South Asian | Wong et al. |
| Rohim |  | South Asian | Lim et al. |
| Rojales | Filipino | Other Asian | Wong et al. |
| Rojas | Filipino | Other Asian | Wong et al. |
| Rojo | Filipino | Other Asian | Wong et al. |
| Rokanouzzaman |  | South Asian | Lim et al. |
| Rokerya | Asian Indian | South Asian | Wong et al. |
| Rokey |  | South Asian | Lim et al. |
| Rokia |  | South Asian | Lim et al. |
| Rokkaku | Japanese | Other Asian | Wong et al. |
| Rol | Filipino | Other Asian | Wong et al. |
| Rola | Filipino | Other Asian | Wong et al. |
| Roldan | Filipino | Other Asian | Kranick et al. |
| Roliz | Chinese | Chinese | Wong et al. |
| Rollan | Filipino | Other Asian | Wong et al. |
| Rollolazo | Filipino | Other Asian | Wong et al. |
| Rollon | Filipino | Other Asian | Wong et al. |
| Roman | Filipino | Other Asian | Wong et al. |
| Romana | Filipino | Other Asian | Wong et al. |
| Romance |  | South Asian | Lim et al. |
| Romasanta | Filipino | Other Asian | Wong et al. |
| Romasoc | Filipino | Other Asian | Wong et al. |
| Rombaoa | Filipino | Other Asian | Wong et al. |
| Rombawa | Filipino | Other Asian | Wong et al. |
| Romen | Filipino | Other Asian | Wong et al. |
| Romena | Filipino | Other Asian | Wong et al. |
| Romero | Filipino | Other Asian | Wong et al. |
| Romerosalas | Filipino | Other Asian | Wong et al. |
| Romillo | Filipino | Other Asian | Wong et al. |
| Romo | Filipino | Other Asian | Wong et al. |
| Romos | Filipino | Other Asian | Wong et al. |
| Romua | Filipino | Other Asian | Wong et al. |
| Romualdez | Filipino | Other Asian | Wong et al. |
| Romualdo | Filipino | Other Asian | Wong et al. |
| Romuar | Filipino | Other Asian | Wong et al. |
| Romulo | Filipino | Other Asian | Wong et al. |
| Rona | Filipino | Other Asian | Wong et al. |
| Ronas | Filipino | Other Asian | Wong et al. |
| Ronato | Filipino | Other Asian | Wong et al. |
| Roncal | Filipino | Other Asian | Wong et al. |
| Rondario | Filipino | Other Asian | Wong et al. |
| Rondaris | Filipino | Other Asian | Wong et al. |
| Rondero | Filipino | Other Asian | Wong et al. |
| Rondez | Filipino | Other Asian | Wong et al. |
| Rondilla | Filipino | Other Asian | Wong et al. |
| Rondolo | Filipino | Other Asian | Wong et al. |
| Rondolos | Filipino | Other Asian | Wong et al. |
| Rondon | Filipino | Other Asian | Wong et al. |
| Ronduen | Filipino | Other Asian | Wong et al. |
| Rong | Chinese | Chinese | Wong et al. |
| Rongavilla | Filipino | Other Asian | Wong et al. |
| Roni |  | South Asian | Lim et al. |
| Rono | Filipino | Other Asian | Wong et al. |
| Ronquillo | Filipino | Other Asian | Kranick et al. |
| Roopchand |  | South Asian | Lim et al. |
| Roopnarain |  | South Asian | Lim et al. |
| Roopnarine |  | South Asian | Lim et al. |
| Roopnautt | Indonesian | Other Asian | Kranick et al. |
| Roque | Filipino | Other Asian | Wong et al. |
| Rosa | Filipino | Other Asian | Wong et al. |
| Rosacia | Filipino | Other Asian | Wong et al. |
| Rosagas | Filipino | Other Asian | Wong et al. |
| Rosal | Filipino | Other Asian | Wong et al. |
| Rosaldo | Filipino | Other Asian | Wong et al. |
| Rosales | Filipino | Other Asian | Wong et al. |
| Rosalin | Filipino | Other Asian | Wong et al. |
| Rosana | Filipino | Other Asian | Wong et al. |
| Rosanes | Filipino | Other Asian | Wong et al. |
| Rosario | Filipino | Other Asian | Wong et al. |
| Rosaro | Filipino | Other Asian | Wong et al. |
| Rosas | Filipino | Other Asian | Wong et al. |
| Rosauro | Filipino | Other Asian | Wong et al. |
| Roscom | Filipino | Other Asian | Wong et al. |
| Rosel | Filipino | Other Asian | Wong et al. |
| Rosello | Filipino | Other Asian | Wong et al. |
| Rosendo | Filipino | Other Asian | Wong et al. |
| Rosero | Filipino | Other Asian | Wong et al. |
| Rosete | Filipino | Other Asian | Wong et al. |
| Rosetes | Filipino | Other Asian | Wong et al. |
| Rosillo | Filipino | Other Asian | Wong et al. |
| Rosiri | Thai | Other Asian | Kranick et al. |
| Rosqueta | Filipino | Other Asian | Wong et al. |
| Rostrata | Filipino | Other Asian | Wong et al. |
| Rosy |  | South Asian | Lim et al. |
| Rot | Vietnamese | Other Asian | Wong et al. |
| Rotairo | Filipino | Other Asian | Wong et al. |
| Rotea | Filipino | Other Asian | Wong et al. |
| Rotor | Filipino | Other Asian | Wong et al. |
| Rouf |  | South Asian | Lim et al. |
| Rovillos | Filipino | Other Asian | Wong et al. |
| Roxas | Filipino | Other Asian | Wong et al. |
| Roy | Asian Indian | South Asian | Kranick et al. |
| Roychoudhur | Asian Indian | South Asian | Wong et al. |
| Roychoudhury | Asian Indian | South Asian | Wong et al. |
| Roychowdhur | Asian Indian | South Asian | Wong et al. |
| Roychowdhury | Asian Indian | South Asian | Wong et al. |
| Royo | Filipino | Other Asian | Wong et al. |
| Rozal | Filipino | Other Asian | Wong et al. |
| Rozario |  | South Asian | Lim et al. |
| Rozul | Filipino | Other Asian | Wong et al. |
| Ru | Chinese | Chinese | Wong et al. |
| Ruaburo | Filipino | Other Asian | Wong et al. |
| Rualo | Filipino | Other Asian | Wong et al. |
| Ruan | Chinese | Chinese | Kranick et al. |
| Ruano | Filipino | Other Asian | Wong et al. |
| Ruanto | Filipino | Other Asian | Wong et al. |
| Ruaro | Filipino | Other Asian | Wong et al. |
| Ruas | Chinese | Chinese | Wong et al. |
| Ruazol | Filipino | Other Asian | Wong et al. |
| Rubang | Filipino | Other Asian | Wong et al. |
| Rubel |  | South Asian | Lim et al. |
| Rubi | Filipino | Other Asian | Wong et al. |
| Rubia | Filipino | Other Asian | Wong et al. |
| Rubiano | Filipino | Other Asian | Wong et al. |
| Rubic | Filipino | Other Asian | Wong et al. |
| Rubina | Filipino | Other Asian | Wong et al. |
| Rubio | Filipino | Other Asian | Wong et al. |
| Rubrico | Filipino | Other Asian | Wong et al. |
| Rudher |  | South Asian | Lim et al. |
| Rudio | Filipino | Other Asian | Wong et al. |
| Rudra | Asian Indian | South Asian | Wong et al. |
| Rueca | Filipino | Other Asian | Wong et al. |
| Rueda | Filipino | Other Asian | Wong et al. |
| Ruedas | Filipino | Other Asian | Wong et al. |
| Ruelan | Filipino | Other Asian | Wong et al. |
| Ruelos | Filipino | Other Asian | Wong et al. |
| Ruffy | Filipino | Other Asian | Wong et al. |
| Rufino | Filipino | Other Asian | Wong et al. |
| Rufo | Filipino | Other Asian | Wong et al. |
| Rufon | Filipino | Other Asian | Wong et al. |
| Rugnao | Filipino | Other Asian | Wong et al. |
| Ruhul |  | South Asian | Lim et al. |
| Rui | Chinese | Chinese | Wong et al. |
| Ruidera | Filipino | Other Asian | Wong et al. |
| Ruivivar | Filipino | Other Asian | Wong et al. |
| Ruiz | Filipino | Other Asian | Wong et al. |
| Rull | Filipino | Other Asian | Wong et al. |
| Rullamas | Filipino | Other Asian | Wong et al. |
| Rullan | Filipino | Other Asian | Wong et al. |
| Rulloda | Filipino | Other Asian | Wong et al. |
| Rulona | Filipino | Other Asian | Wong et al. |
| Ruma | Filipino | Other Asian | Wong et al. |
| Rumbaoa | Filipino | Other Asian | Wong et al. |
| Rumbaua | Filipino | Other Asian | Wong et al. |
| Rumi |  | South Asian | Lim et al. |
| Rumjahn | Chinese | Chinese | Wong et al. |
| Run | Chinese | Chinese | Wong et al. |
| Runas | Filipino | Other Asian | Wong et al. |
| Runes | Filipino | Other Asian | Wong et al. |
| Runez | Filipino | Other Asian | Wong et al. |
| Runia |  | South Asian | Lim et al. |
| Rupa | Filipino | Other Asian | Wong et al. |
| Rupani | Asian Indian | South Asian | Wong et al. |
| Ruparel | Asian Indian | South Asian | Wong et al. |
| Ruperto | Filipino | Other Asian | Wong et al. |
| Rupisan | Filipino | Other Asian | Wong et al. |
| Rusiana | Filipino | Other Asian | Wong et al. |
| Rusit | Filipino | Other Asian | Wong et al. |
| Russel |  | South Asian | Lim et al. |
| Rustia |  | South Asian | Lim et al. |
| Rutab | Filipino | Other Asian | Wong et al. |
| Rutaquio | Filipino | Other Asian | Wong et al. |
| Ruz | Filipino | Other Asian | Wong et al. |
| Ryang | Korean | Other Asian | Wong et al. |
| Ryong | Korean | Other Asian | Wong et al. |
| Ryono | Japanese | Other Asian | Wong et al. |
| Ryoo | Korean | Other Asian | Wong et al. |
| Ryou | Korean | Other Asian | Kranick et al. |
| Ryu | Korean | Other Asian | Kranick et al. |
| Ryuzaki | Japanese | Other Asian | Wong et al. |
| S | Chinese | Chinese | Wong et al. |
| Saa | Filipino | Other Asian | Wong et al. |
| Saavedra | Filipino | Other Asian | Wong et al. |
| Sabado | Filipino | Other Asian | Wong et al. |
| Sabal | Filipino | Other Asian | Wong et al. |
| Sabala | Filipino | Other Asian | Wong et al. |
| Sabalboro | Filipino | Other Asian | Wong et al. |
| Sabalburo | Filipino | Other Asian | Wong et al. |
| Sabale | Filipino | Other Asian | Wong et al. |
| Saballa | Filipino | Other Asian | Wong et al. |
| Sabalvaro | Filipino | Other Asian | Wong et al. |
| Sabana |  | South Asian | Lim et al. |
| Sabanal | Filipino | Other Asian | Wong et al. |
| Sabandal | Filipino | Other Asian | Wong et al. |
| Sabando | Filipino | Other Asian | Wong et al. |
| Sabangan | Filipino | Other Asian | Wong et al. |
| Sabanpan | Filipino | Other Asian | Wong et al. |
| Sabarre | Filipino | Other Asian | Wong et al. |
| Sabarwal | Asian Indian | South Asian | Wong et al. |
| Sabas | Filipino | Other Asian | Wong et al. |
| Sabate | Filipino | Other Asian | Wong et al. |
| Sabater | Filipino | Other Asian | Wong et al. |
| Sabatin | Filipino | Other Asian | Wong et al. |
| Sabay | Filipino | Other Asian | Wong et al. |
| Sabayle | Filipino | Other Asian | Wong et al. |
| Sabejon | Filipino | Other Asian | Wong et al. |
| Sabellano | Filipino | Other Asian | Wong et al. |
| Saberon | Filipino | Other Asian | Wong et al. |
| Sabharwal | Asian Indian | South Asian | Wong et al. |
| Sabherwal | Asian Indian | South Asian | Wong et al. |
| Sabido | Filipino | Other Asian | Wong et al. |
| Sabijon | Filipino | Other Asian | Wong et al. |
| Sabile | Filipino | Other Asian | Wong et al. |
| Sabillo | Filipino | Other Asian | Wong et al. |
| Sabinay | Filipino | Other Asian | Wong et al. |
| Sabiniano | Filipino | Other Asian | Wong et al. |
| Sabino | Filipino | Other Asian | Wong et al. |
| Sabio | Filipino | Other Asian | Wong et al. |
| Sabir | Asian Indian | South Asian | Wong et al. |
| Sablad | Filipino | Other Asian | Wong et al. |
| Sablan | Filipino | Other Asian | Wong et al. |
| Sablay | Filipino | Other Asian | Wong et al. |
| Sabnis | Asian Indian | South Asian | Wong et al. |
| Sabong | Filipino | Other Asian | Wong et al. |
| Sabordo | Filipino | Other Asian | Wong et al. |
| Sabri | Asian Indian | South Asian | Wong et al. |
| Sabugo | Filipino | Other Asian | Wong et al. |
| Sabuj |  | South Asian | Lim et al. |
| Sabur |  | South Asian | Lim et al. |
| Saburi | Japanese | Other Asian | Wong et al. |
| Saburo | Japanese | Other Asian | Wong et al. |
| Sabuz |  | South Asian | Lim et al. |
| Sacaguing | Filipino | Other Asian | Wong et al. |
| Sacay | Filipino | Other Asian | Wong et al. |
| Sacayanan | Filipino | Other Asian | Wong et al. |
| Sacdalan | Filipino | Other Asian | Wong et al. |
| Saceda | Filipino | Other Asian | Wong et al. |
| Sach | Vietnamese | Other Asian | Wong et al. |
| Sachariah | Asian Indian | South Asian | Wong et al. |
| Sachdev | Asian Indian | South Asian | Wong et al. |
| Sachdeva | Asian Indian | South Asian | Kranick et al. |
| Saclayan | Filipino | Other Asian | Wong et al. |
| Saclolo | Filipino | Other Asian | Wong et al. |
| Sacluti | Filipino | Other Asian | Wong et al. |
| Sacoco | Filipino | Other Asian | Wong et al. |
| Sacramento | Filipino | Other Asian | Wong et al. |
| Sacro | Filipino | Other Asian | Wong et al. |
| Saculles | Filipino | Other Asian | Wong et al. |
| Sada | Japanese | Other Asian | Wong et al. |
| Sadang | Filipino | Other Asian | Wong et al. |
| Sadasivan | Asian Indian | South Asian | Wong et al. |
| Sadaya | Filipino | Other Asian | Wong et al. |
| Saddana |  | South Asian | Lim et al. |
| Saddi | Filipino | Other Asian | Wong et al. |
| Sadhu | Asian Indian | South Asian | Wong et al. |
| Sadhukhan | Asian Indian | South Asian | Wong et al. |
| Sadia | Filipino | Other Asian | Wong et al. |
| Sadian | Filipino | Other Asian | Wong et al. |
| Sadiasa | Filipino | Other Asian | Wong et al. |
| Sadie | Filipino | Other Asian | Wong et al. |
| Sadio | Filipino | Other Asian | Wong et al. |
| Sadique |  | South Asian | Lim et al. |
| Sado | Filipino | Other Asian | Wong et al. |
| Sadorra | Filipino | Other Asian | Wong et al. |
| Sadsad | Filipino | Other Asian | Wong et al. |
| Sadueste | Filipino | Other Asian | Wong et al. |
| Sadumiano | Filipino | Other Asian | Wong et al. |
| Sadural | Filipino | Other Asian | Wong et al. |
| Sae | Korean | Other Asian | Wong et al. |
| Saebae | Chinese | Chinese | Wong et al. |
| Saeed | Pakistani | South Asian | Kranick et al. |
| Saegusa | Japanese | Other Asian | Wong et al. |
| Saeki | Japanese | Other Asian | Wong et al. |
| Saenz | Filipino | Other Asian | Wong et al. |
| Saet | Filipino | Other Asian | Wong et al. |
| Saeung | Chinese | Chinese | Wong et al. |
| Saez | Filipino | Other Asian | Wong et al. |
| Safa | Japanese | Other Asian | Wong et al. |
| Safiullah |  | South Asian | Lim et al. |
| Saflor | Filipino | Other Asian | Wong et al. |
| Saga | Japanese | Other Asian | Wong et al. |
| Sagabaen | Filipino | Other Asian | Wong et al. |
| Sagadraca | Filipino | Other Asian | Wong et al. |
| Sagala | Filipino | Other Asian | Wong et al. |
| Sagami | Japanese | Other Asian | Wong et al. |
| Sagaoinit | Filipino | Other Asian | Wong et al. |
| Sagara | Japanese | Other Asian | Wong et al. |
| Sagario | Filipino | Other Asian | Wong et al. |
| Sagarwala | Asian Indian | South Asian | Wong et al. |
| Sagawa | Japanese | Other Asian | Wong et al. |
| Sagayadoro | Filipino | Other Asian | Wong et al. |
| Sagayaga | Filipino | Other Asian | Wong et al. |
| Sagaysay | Filipino | Other Asian | Wong et al. |
| Sagcal | Filipino | Other Asian | Wong et al. |
| Saggar | Asian Indian | South Asian | Wong et al. |
| Saggu | Asian Indian | South Asian | Wong et al. |
| Sagisi | Filipino | Other Asian | Wong et al. |
| Sagmaquen | Filipino | Other Asian | Wong et al. |
| Sagmit | Filipino | Other Asian | Wong et al. |
| Sagon | Filipino | Other Asian | Wong et al. |
| Sagoo | Asian Indian | South Asian | Wong et al. |
| Sagpao | Filipino | Other Asian | Wong et al. |
| Sagrado | Filipino | Other Asian | Kranick et al. |
| Sagritalo | Filipino | Other Asian | Wong et al. |
| Sagucio | Filipino | Other Asian | Wong et al. |
| Saguibo | Filipino | Other Asian | Wong et al. |
| Saguid | Filipino | Other Asian | Wong et al. |
| Saguiguit | Filipino | Other Asian | Wong et al. |
| Saguil | Filipino | Other Asian | Wong et al. |
| Saguinsin | Filipino | Other Asian | Wong et al. |
| Saguiped | Filipino | Other Asian | Wong et al. |
| Sagullo | Filipino | Other Asian | Wong et al. |
| Sagum | Filipino | Other Asian | Wong et al. |
| Sagun | Filipino | Other Asian | Wong et al. |
| Sah | Chinese | Chinese | Wong et al. |
| Saha | Asian Indian; Bangladeshi | South Asian | Kranick et al. |
| Sahab |  | South Asian | Lim et al. |
| Sahabuddin |  | South Asian | Lim et al. |
| Sahadeo |  | South Asian | Lim et al. |
| Sahagun | Filipino | Other Asian | Wong et al. |
| Sahai | Asian Indian | South Asian | Wong et al. |
| Sahajahan |  | South Asian | Lim et al. |
| Sahara | Japanese | Other Asian | Wong et al. |
| Sahashi | Japanese | Other Asian | Wong et al. |
| Sahay | Asian Indian | South Asian | Wong et al. |
| Sahdev | Asian Indian | South Asian | Kranick et al. |
| Saheba | Asian Indian | South Asian | Wong et al. |
| Saheeda |  | South Asian | Lim et al. |
| Saheki | Japanese | Other Asian | Wong et al. |
| Sahgal | Asian Indian | South Asian | Wong et al. |
| Sahi | Asian Indian | South Asian | Wong et al. |
| Sahid |  | South Asian | Lim et al. |
| Sahney | Asian Indian | South Asian | Wong et al. |
| Sahni |  | South Asian | Lim et al. |
| Sahota | Asian Indian | South Asian | Wong et al. |
| Sahu | Asian Indian | South Asian | Wong et al. |
| Sai | Chinese | Chinese | Wong et al. |
| Saida | Japanese | Other Asian | Wong et al. |
| Saidullah |  | South Asian | Lim et al. |
| Saifee | Asian Indian | South Asian | Wong et al. |
| Saifi |  | South Asian | Lim et al. |
| Saifuddin |  | South Asian | Lim et al. |
| Saiful |  | South Asian | Lim et al. |
| Saifullah |  | South Asian | Lim et al. |
| Saiga | Japanese | Other Asian | Wong et al. |
| Saigo | Japanese | Other Asian | Wong et al. |
| Saigusa | Japanese | Other Asian | Wong et al. |
| Saijo | Japanese | Other Asian | Wong et al. |
| Saika | Japanese | Other Asian | Wong et al. |
| Saikawa | Japanese | Other Asian | Wong et al. |
| Saiki | Japanese | Other Asian | Wong et al. |
| Saikia | Asian Indian | South Asian | Wong et al. |
| Saima |  | South Asian | Lim et al. |
| Saini | Asian Indian | South Asian | Wong et al. |
| Sainz | Filipino | Other Asian | Wong et al. |
| Saisho | Japanese | Other Asian | Wong et al. |
| Sait | Asian Indian | South Asian | Wong et al. |
| Saita | Japanese | Other Asian | Wong et al. |
| Saito | Japanese | Other Asian | Wong et al. |
| Saitoh | Japanese | Other Asian | Wong et al. |
| Saiyed | Asian Indian | South Asian | Wong et al. |
| Saizuddin |  | South Asian | Lim et al. |
| Saji | Japanese | Other Asian | Wong et al. |
| Sajib |  | South Asian | Lim et al. |
| Sajid |  | South Asian | Lim et al. |
| Sajja | Asian Indian | South Asian | Wong et al. |
| Sajo | Filipino | Other Asian | Wong et al. |
| Sajol | Filipino | Other Asian | Wong et al. |
| Sajonas | Filipino | Other Asian | Wong et al. |
| Sajor | Filipino | Other Asian | Wong et al. |
| Saka | Japanese | Other Asian | Wong et al. |
| Sakabe | Japanese | Other Asian | Wong et al. |
| Sakae | Japanese | Other Asian | Wong et al. |
| Sakagami | Japanese | Other Asian | Wong et al. |
| Sakaguchi | Japanese | Other Asian | Wong et al. |
| Sakai | Japanese | Other Asian | Wong et al. |
| Sakaida | Japanese | Other Asian | Wong et al. |
| Sakairi | Japanese | Other Asian | Wong et al. |
| Sakaki | Japanese | Other Asian | Wong et al. |
| Sakakibara | Japanese | Other Asian | Wong et al. |
| Sakakihara | Japanese | Other Asian | Wong et al. |
| Sakakura | Japanese | Other Asian | Wong et al. |
| Sakamaki | Japanese | Other Asian | Wong et al. |
| Sakamoto | Japanese | Other Asian | Wong et al. |
| Sakane | Japanese | Other Asian | Wong et al. |
| Sakano | Japanese | Other Asian | Wong et al. |
| Sakaria | Asian Indian | South Asian | Wong et al. |
| Sakashita | Japanese | Other Asian | Kranick et al. |
| Sakata | Japanese | Other Asian | Wong et al. |
| Sakauchi | Japanese | Other Asian | Wong et al. |
| Sakaue | Japanese | Other Asian | Wong et al. |
| Sakay | Filipino | Other Asian | Wong et al. |
| Sakazaki | Japanese | Other Asian | Wong et al. |
| Saki | Japanese | Other Asian | Wong et al. |
| Sakihama | Japanese | Other Asian | Wong et al. |
| Sakihara | Japanese | Other Asian | Wong et al. |
| Sakima | Japanese | Other Asian | Wong et al. |
| Sakimoto | Japanese | Other Asian | Wong et al. |
| Sakimura | Japanese | Other Asian | Wong et al. |
| Sakioka | Japanese | Other Asian | Wong et al. |
| Sakita | Japanese | Other Asian | Wong et al. |
| Sakiyama | Japanese | Other Asian | Wong et al. |
| Sakkari |  | South Asian | Lim et al. |
| Sako | Japanese | Other Asian | Wong et al. |
| Sakoda | Japanese | Other Asian | Wong et al. |
| Sakoor |  | South Asian | Lim et al. |
| Sakore | Asian Indian | South Asian | Kranick et al. |
| Saksena | Asian Indian | South Asian | Wong et al. |
| Sakuda | Japanese | Other Asian | Wong et al. |
| Sakugawa | Japanese | Other Asian | Wong et al. |
| Sakuma | Japanese | Other Asian | Kranick et al. |
| Sakumoto | Japanese | Other Asian | Wong et al. |
| Sakura | Japanese | Other Asian | Wong et al. |
| Sakuraba | Japanese | Other Asian | Wong et al. |
| Sakurada | Japanese | Other Asian | Wong et al. |
| Sakuragi | Japanese | Other Asian | Wong et al. |
| Sakurai | Japanese | Other Asian | Wong et al. |
| Salabao | Filipino | Other Asian | Wong et al. |
| Salac | Filipino | Other Asian | Wong et al. |
| Salacup | Filipino | Other Asian | Wong et al. |
| Saladar | Filipino | Other Asian | Wong et al. |
| Salagubang | Filipino | Other Asian | Wong et al. |
| Salahdin |  | South Asian | Lim et al. |
| Salahuddin |  | South Asian | Lim et al. |
| Salalila | Filipino | Other Asian | Wong et al. |
| Salam | Bangladeshi; Asian Indian; Pakistani; Indonesian | South Asian | Kranick et al. |
| Salamanca | Filipino | Other Asian | Wong et al. |
| Salamante | Filipino | Other Asian | Wong et al. |
| Salamat | Filipino | Other Asian | Wong et al. |
| Salameda | Filipino | Other Asian | Wong et al. |
| Salamero | Filipino | Other Asian | Wong et al. |
| Salandanan | Filipino | Other Asian | Wong et al. |
| Salang | Filipino | Other Asian | Wong et al. |
| Salanga | Filipino | Other Asian | Wong et al. |
| Salangad | Filipino | Other Asian | Wong et al. |
| Salango | Filipino | Other Asian | Wong et al. |
| Salangsang | Filipino | Other Asian | Wong et al. |
| Salanguit | Filipino | Other Asian | Wong et al. |
| Salanio | Filipino | Other Asian | Wong et al. |
| Salantes | Filipino | Other Asian | Wong et al. |
| Salao | Filipino | Other Asian | Wong et al. |
| Salapare | Filipino | Other Asian | Wong et al. |
| Salapong | Filipino | Other Asian | Wong et al. |
| Salarda | Filipino | Other Asian | Wong et al. |
| Salaria | Filipino | Other Asian | Wong et al. |
| Salas |  | South Asian | Lim et al. |
| Salatan | Filipino | Other Asian | Wong et al. |
| Salauddin |  | South Asian | Lim et al. |
| Salaver | Filipino | Other Asian | Wong et al. |
| Salaveria | Filipino | Other Asian | Wong et al. |
| Salayog | Filipino | Other Asian | Wong et al. |
| Salaysay | Filipino | Other Asian | Wong et al. |
| Salazar | Filipino | Other Asian | Kranick et al. |
| Salceda | Filipino | Other Asian | Wong et al. |
| Salcedo | Filipino | Other Asian | Wong et al. |
| Saldajeno | Filipino | Other Asian | Wong et al. |
| Saldana | Filipino | Other Asian | Wong et al. |
| Saldanha | Asian Indian | South Asian | Wong et al. |
| Salde | Filipino | Other Asian | Wong et al. |
| Saldevar | Filipino | Other Asian | Wong et al. |
| Saldivar | Filipino | Other Asian | Wong et al. |
| Saldua | Filipino | Other Asian | Wong et al. |
| Saleem | Pakistani | South Asian | Kranick et al. |
| Saleh | Bangladeshi | South Asian | Kranick et al. |
| Salenga | Filipino | Other Asian | Wong et al. |
| Sales | Filipino | Other Asian | Wong et al. |
| Salgado | Filipino | Other Asian | Wong et al. |
| Salguero | Filipino | Other Asian | Wong et al. |
| Salibad | Filipino | Other Asian | Wong et al. |
| Salido | Filipino | Other Asian | Wong et al. |
| Saliganan | Filipino | Other Asian | Wong et al. |
| Saligumba | Filipino | Other Asian | Wong et al. |
| Salik | Pakistani | South Asian | Kranick et al. |
| Salim |  | South Asian | Lim et al. |
| Salinas | Filipino | Other Asian | Wong et al. |
| Salinda | Sri Lankan | South Asian | Kranick et al. |
| Salindo | Filipino | Other Asian | Wong et al. |
| Salindong | Filipino | Other Asian | Wong et al. |
| Salis | Filipino | Other Asian | Wong et al. |
| Salisi | Filipino | Other Asian | Wong et al. |
| Salita | Filipino | Other Asian | Wong et al. |
| Salma |  | South Asian | Lim et al. |
| Salmasan | Filipino | Other Asian | Wong et al. |
| Salmo | Filipino | Other Asian | Wong et al. |
| Salmorin | Filipino | Other Asian | Wong et al. |
| Salom | Filipino | Other Asian | Wong et al. |
| Salon | Filipino | Other Asian | Wong et al. |
| Salonga | Filipino | Other Asian | Wong et al. |
| Salta | Filipino | Other Asian | Wong et al. |
| Salting | Filipino | Other Asian | Wong et al. |
| Salud | Filipino | Other Asian | Wong et al. |
| Saludares | Filipino | Other Asian | Wong et al. |
| Saludes | Filipino | Other Asian | Wong et al. |
| Saludez | Filipino | Other Asian | Wong et al. |
| Saludo | Filipino | Other Asian | Wong et al. |
| Salugao | Filipino | Other Asian | Wong et al. |
| Saluja |  | South Asian | Lim et al. |
| Salum | Filipino | Other Asian | Wong et al. |
| Salumbides | Filipino | Other Asian | Wong et al. |
| Salunga | Filipino | Other Asian | Wong et al. |
| Saluta | Filipino | Other Asian | Wong et al. |
| Salva | Filipino | Other Asian | Wong et al. |
| Salvacion | Filipino | Other Asian | Wong et al. |
| Salvacruz | Filipino | Other Asian | Wong et al. |
| Salvado | Filipino | Other Asian | Wong et al. |
| Salvador | Filipino | Other Asian | Kranick et al. |
| Salvadora | Filipino | Other Asian | Wong et al. |
| Salvaleon | Filipino | Other Asian | Wong et al. |
| Salvan | Filipino | Other Asian | Wong et al. |
| Salvanera | Filipino | Other Asian | Wong et al. |
| Salvani | Filipino | Other Asian | Wong et al. |
| Salvante | Filipino | Other Asian | Wong et al. |
| Salvatera | Filipino | Other Asian | Wong et al. |
| Salvatierra | Filipino | Other Asian | Wong et al. |
| Salvatus | Filipino | Other Asian | Wong et al. |
| Salviejo | Filipino | Other Asian | Wong et al. |
| Salvilla | Filipino | Other Asian | Wong et al. |
| Salvosa | Filipino | Other Asian | Wong et al. |
| Salwan | Asian Indian | South Asian | Wong et al. |
| Sam | Chinese | Chinese | Wong et al. |
| Sama |  | South Asian | Lim et al. |
| Samaco | Filipino | Other Asian | Wong et al. |
| Samad | Asian Indian | South Asian | Wong et al. |
| Samaddar |  | South Asian | Lim et al. |
| Samala | Filipino | Other Asian | Wong et al. |
| Samaniego | Filipino | Other Asian | Wong et al. |
| Samano | Filipino | Other Asian | Wong et al. |
| Samant | Asian Indian | South Asian | Wong et al. |
| Samanta | Asian Indian | South Asian | Wong et al. |
| Samaroo |  | South Asian | Lim et al. |
| Samatra | Filipino | Other Asian | Wong et al. |
| Sambajon | Filipino | Other Asian | Wong et al. |
| Sambamurti | Asian Indian | South Asian | Wong et al. |
| Sambas | Filipino | Other Asian | Wong et al. |
| Sambasivan | Asian Indian | South Asian | Wong et al. |
| Sambat | Filipino | Other Asian | Wong et al. |
| Sambhi | Asian Indian | South Asian | Wong et al. |
| Sambilay | Filipino | Other Asian | Wong et al. |
| Sambile | Filipino | Other Asian | Wong et al. |
| Sambo | Filipino | Other Asian | Wong et al. |
| Sambrano | Filipino | Other Asian | Wong et al. |
| Samdani |  | South Asian | Lim et al. |
| Samejima | Japanese | Other Asian | Wong et al. |
| Samera | Filipino | Other Asian | Wong et al. |
| Sameshima | Japanese | Other Asian | Wong et al. |
| Sami | Asian Indian | South Asian | Wong et al. |
| Samia | Filipino | Other Asian | Wong et al. |
| Samiano | Filipino | Other Asian | Wong et al. |
| Samilin | Filipino | Other Asian | Wong et al. |
| Samina |  | South Asian | Lim et al. |
| Samlall |  | South Asian | Lim et al. |
| Sammy |  | South Asian | Lim et al. |
| Samodal | Filipino | Other Asian | Wong et al. |
| Samodio | Filipino | Other Asian | Wong et al. |
| Samonte | Filipino | Other Asian | Wong et al. |
| Samoy | Filipino | Other Asian | Wong et al. |
| Sampaga | Filipino | Other Asian | Wong et al. |
| Sampana | Filipino | Other Asian | Wong et al. |
| Sampang | Filipino | Other Asian | Wong et al. |
| Sampat | Asian Indian | South Asian | Wong et al. |
| Sampath | Asian Indian | South Asian | Wong et al. |
| Sampathkumar | Asian Indian | South Asian | Wong et al. |
| Sampayan | Filipino | Other Asian | Wong et al. |
| Sampilo | Filipino | Other Asian | Wong et al. |
| Sampson |  | South Asian | Lim et al. |
| Samra | Asian Indian | South Asian | Wong et al. |
| Samson | Filipino | Other Asian | Wong et al. |
| Samukawa | Japanese | Other Asian | Wong et al. |
| Samy | Asian Indian | South Asian | Wong et al. |
| San | Chinese | Chinese | Wong et al. |
| San Jose | Filipino | Other Asian | Kranick et al. |
| Sana |  | South Asian | Lim et al. |
| Sanada | Japanese | Other Asian | Wong et al. |
| Sanagustin | Filipino | Other Asian | Wong et al. |
| Sanandres | Filipino | Other Asian | Wong et al. |
| Sanantonio | Filipino | Other Asian | Wong et al. |
| Sanares | Filipino | Other Asian | Wong et al. |
| Sanbuenaven | Filipino | Other Asian | Wong et al. |
| Sanbuenavent | Filipino | Other Asian | Wong et al. |
| Sanches | Filipino | Other Asian | Wong et al. |
| Sanchez | Filipino | Other Asian | Wong et al. |
| Sancho | Filipino | Other Asian | Wong et al. |
| Sanciangco | Filipino | Other Asian | Wong et al. |
| Sandaga | Filipino | Other Asian | Wong et al. |
| Sandagan | Filipino | Other Asian | Wong et al. |
| Sandan | Filipino | Other Asian | Wong et al. |
| Sandejas | Filipino | Other Asian | Wong et al. |
| Sandhar | Asian Indian | South Asian | Wong et al. |
| Sandhu |  | South Asian | Lim et al. |
| Sandi | Filipino | Other Asian | Wong et al. |
| Sandico | Filipino | Other Asian | Wong et al. |
| Sandiego | Filipino | Other Asian | Wong et al. |
| Sando | Japanese | Other Asian | Wong et al. |
| Sandoval | Filipino | Other Asian | Wong et al. |
| Sanez | Filipino | Other Asian | Wong et al. |
| Sanfelipe | Filipino | Other Asian | Wong et al. |
| Sang | Chinese; Vietnamese; Korean | Chinese | Kranick et al. |
| Sanga | Filipino | Other Asian | Wong et al. |
| Sangabriel | Filipino | Other Asian | Wong et al. |
| Sangalang | Filipino | Other Asian | Wong et al. |
| Sangani | Asian Indian | South Asian | Wong et al. |
| Sangar | Asian Indian | South Asian | Wong et al. |
| Sangaralingam |  | South Asian | Lim et al. |
| Sangarapillai | Sri Lankan | South Asian | Kranick et al. |
| Sangco | Filipino | Other Asian | Wong et al. |
| Sangel | Filipino | Other Asian | Wong et al. |
| Sangha | Asian Indian | South Asian | Wong et al. |
| Sanghani | Asian Indian | South Asian | Wong et al. |
| Sanghavi | Asian Indian | South Asian | Kranick et al. |
| Sanghera | Asian Indian | South Asian | Wong et al. |
| Sanghi | Asian Indian | South Asian | Wong et al. |
| Sanghui | Asian Indian | South Asian | Wong et al. |
| Sanghvi | Asian Indian | South Asian | Kranick et al. |
| Sangil | Filipino | Other Asian | Wong et al. |
| Sanglay | Filipino | Other Asian | Wong et al. |
| Sangoi | Asian Indian | South Asian | Kranick et al. |
| Sangria | Filipino | Other Asian | Wong et al. |
| Sanguyo | Filipino | Other Asian | Wong et al. |
| Sanh | Vietnamese | Other Asian | Wong et al. |
| Sani | Indonesian; Pakistani; Bangladeshi | Other Asian | Kranick et al. |
| Saniatan | Filipino | Other Asian | Wong et al. |
| Sanico | Filipino | Other Asian | Wong et al. |
| Sanidad | Filipino | Other Asian | Wong et al. |
| Saniel | Filipino | Other Asian | Wong et al. |
| Sanjose | Filipino | Other Asian | Wong et al. |
| Sanjuan | Filipino | Other Asian | Kranick et al. |
| Sankar |  | South Asian | Lim et al. |
| Sankaran | Asian Indian | South Asian | Wong et al. |
| Sanluis | Filipino | Other Asian | Wong et al. |
| Sanmateo | Filipino | Other Asian | Wong et al. |
| Sanmiguel | Filipino | Other Asian | Wong et al. |
| Sannomiya | Japanese | Other Asian | Wong et al. |
| Sano | Japanese | Other Asian | Wong et al. |
| Sanoria | Filipino | Other Asian | Wong et al. |
| Sanosa | Filipino | Other Asian | Wong et al. |
| Sanoy | Filipino | Other Asian | Wong et al. |
| Sanpablo | Filipino | Other Asian | Wong et al. |
| Sanpascual | Filipino | Other Asian | Wong et al. |
| Sanpedro | Filipino | Other Asian | Wong et al. |
| Sanqui | Filipino | Other Asian | Wong et al. |
| Sansano | Filipino | Other Asian | Wong et al. |
| Santaana | Filipino | Other Asian | Wong et al. |
| Santacruz | Filipino | Other Asian | Wong et al. |
| Santamaria | Filipino | Other Asian | Wong et al. |
| Santana | Filipino | Other Asian | Wong et al. |
| Santander | Filipino | Other Asian | Wong et al. |
| Santarin | Filipino | Other Asian | Wong et al. |
| Santarina | Filipino | Other Asian | Wong et al. |
| Santarita | Filipino | Other Asian | Wong et al. |
| Santaromana | Filipino | Other Asian | Wong et al. |
| Santarosa | Filipino | Other Asian | Wong et al. |
| Santarromana | Filipino | Other Asian | Wong et al. |
| Santeco | Filipino | Other Asian | Wong et al. |
| Santelices | Filipino | Other Asian | Wong et al. |
| Santera | Filipino | Other Asian | Wong et al. |
| Santero | Filipino | Other Asian | Wong et al. |
| Santhanam | Asian Indian | South Asian | Wong et al. |
| Santi | Indonesian | Other Asian | Kranick et al. |
| Santiago | Filipino | Other Asian | Wong et al. |
| Santiaguel | Filipino | Other Asian | Wong et al. |
| Santiangco | Filipino | Other Asian | Wong et al. |
| Santiano | Filipino | Other Asian | Wong et al. |
| Santibanez | Filipino | Other Asian | Wong et al. |
| Santillan | Filipino | Other Asian | Wong et al. |
| Santillana | Filipino | Other Asian | Wong et al. |
| Santillano | Filipino | Other Asian | Wong et al. |
| Santisteban | Filipino | Other Asian | Wong et al. |
| Santodoming | Filipino | Other Asian | Wong et al. |
| Santonil | Filipino | Other Asian | Wong et al. |
| Santos | Filipino | Other Asian | Wong et al. |
| Santossanto | Filipino | Other Asian | Wong et al. |
| Santra | Asian Indian | South Asian | Kranick et al. |
| Santulan | Filipino | Other Asian | Wong et al. |
| Sanuki | Japanese | Other Asian | Wong et al. |
| Sanvicente | Filipino | Other Asian | Wong et al. |
| Sanvictores | Filipino | Other Asian | Wong et al. |
| Sanwal | Asian Indian | South Asian | Wong et al. |
| Sanyal | Asian Indian | South Asian | Kranick et al. |
| Sanz | Filipino | Other Asian | Wong et al. |
| Sanzar |  | South Asian | Lim et al. |
| Saoit | Filipino | Other Asian | Wong et al. |
| Saotome | Japanese | Other Asian | Wong et al. |
| Sapaden | Filipino | Other Asian | Wong et al. |
| Sapalasan | Filipino | Other Asian | Wong et al. |
| Sapalo | Filipino | Other Asian | Wong et al. |
| Sapang | Filipino | Other Asian | Wong et al. |
| Sapasap | Filipino | Other Asian | Wong et al. |
| Sapiandante | Filipino | Other Asian | Wong et al. |
| Sapico | Filipino | Other Asian | Wong et al. |
| Sapida | Filipino | Other Asian | Wong et al. |
| Sapiera | Filipino | Other Asian | Wong et al. |
| Sapigao | Filipino | Other Asian | Wong et al. |
| Sapin | Filipino | Other Asian | Wong et al. |
| Sapinoro | Filipino | Other Asian | Wong et al. |
| Sapinoso | Filipino | Other Asian | Wong et al. |
| Sapitan | Filipino | Other Asian | Wong et al. |
| Sapla | Filipino | Other Asian | Wong et al. |
| Saplala | Filipino | Other Asian | Wong et al. |
| Saplan | Filipino | Other Asian | Wong et al. |
| Sapnu | Filipino | Other Asian | Wong et al. |
| Sapon | Filipino | Other Asian | Wong et al. |
| Saporsantos | Filipino | Other Asian | Wong et al. |
| Sapre | Asian Indian | South Asian | Wong et al. |
| Saprid | Filipino | Other Asian | Wong et al. |
| Sapru | Asian Indian | South Asian | Wong et al. |
| Sapuay | Filipino | Other Asian | Wong et al. |
| Sapungan | Filipino | Other Asian | Wong et al. |
| Sapuriada | Filipino | Other Asian | Wong et al. |
| Saqib | Asian Indian | South Asian | Wong et al. |
| Saqueton | Filipino | Other Asian | Wong et al. |
| Saqui |  | South Asian | Lim et al. |
| Saquilayan | Filipino | Other Asian | Wong et al. |
| Saquin | Filipino | Other Asian | Wong et al. |
| Saquing | Filipino | Other Asian | Wong et al. |
| Sarabia | Filipino | Other Asian | Wong et al. |
| Saradpon | Filipino | Other Asian | Wong et al. |
| Saraf | Asian Indian | South Asian | Wong et al. |
| Saraiya | Asian Indian | South Asian | Wong et al. |
| Saran | Asian Indian | South Asian | Wong et al. |
| Sarandi | Filipino | Other Asian | Wong et al. |
| Sarang | Asian Indian | South Asian | Wong et al. |
| Sarangapani | Asian Indian | South Asian | Wong et al. |
| Sarangay | Filipino | Other Asian | Wong et al. |
| Sarangaya | Filipino | Other Asian | Wong et al. |
| Sarasammaamma |  | South Asian | Lim et al. |
| Saraswat | Asian Indian | South Asian | Wong et al. |
| Saraswati | Asian Indian | South Asian | Wong et al. |
| Saravanamuthu |  | South Asian | Lim et al. |
| Sarawar |  | South Asian | Lim et al. |
| Sarayba | Filipino | Other Asian | Wong et al. |
| Saraza | Filipino | Other Asian | Wong et al. |
| Sarceda | Filipino | Other Asian | Wong et al. |
| Sardana | Asian Indian | South Asian | Wong et al. |
| Sardar |  | South Asian | Lim et al. |
| Sardea | Filipino | Other Asian | Wong et al. |
| Sardena | Filipino | Other Asian | Wong et al. |
| Sarder |  | South Asian | Lim et al. |
| Sardesai | Asian Indian | South Asian | Wong et al. |
| Sardillo | Filipino | Other Asian | Wong et al. |
| Sardon | Filipino | Other Asian | Wong et al. |
| Sareen | Asian Indian | South Asian | Wong et al. |
| Saren | Filipino | Other Asian | Wong et al. |
| Sarenas | Filipino | Other Asian | Wong et al. |
| Sarfuddin |  | South Asian | Lim et al. |
| Sargento | Filipino | Other Asian | Wong et al. |
| Saria | Filipino | Other Asian | Wong et al. |
| Sarian | Filipino | Other Asian | Wong et al. |
| Saribay | Filipino | Other Asian | Wong et al. |
| Sarigumba | Filipino | Other Asian | Wong et al. |
| Sarin | Asian Indian | South Asian | Wong et al. |
| Sarinas | Filipino | Other Asian | Wong et al. |
| Sarino | Filipino | Other Asian | Wong et al. |
| Sario | Filipino | Other Asian | Wong et al. |
| Sarita | Filipino | Other Asian | Wong et al. |
| Sarjuprashad |  | South Asian | Lim et al. |
| Sarkar | Asian Indian | South Asian | Wong et al. |
| Sarkari | Asian Indian | South Asian | Wong et al. |
| Sarkaria | Asian Indian | South Asian | Wong et al. |
| Sarker | Bangladeshi | South Asian | Kranick et al. |
| Sarma | Asian Indian | South Asian | Wong et al. |
| Sarmenta | Filipino | Other Asian | Wong et al. |
| Sarmiento | Filipino | Other Asian | Wong et al. |
| Sarmin |  | South Asian | Lim et al. |
| Sarnalata |  | South Asian | Lim et al. |
| Sarne | Filipino | Other Asian | Wong et al. |
| Sarol | Filipino | Other Asian | Wong et al. |
| Sarong | Filipino | Other Asian | Wong et al. |
| Sarowar |  | South Asian | Lim et al. |
| Saroya | Asian Indian | South Asian | Wong et al. |
| Sarreal | Filipino | Other Asian | Wong et al. |
| Sarroca | Filipino | Other Asian | Wong et al. |
| Sarsadias | Filipino | Other Asian | Wong et al. |
| Sarsonas | Filipino | Other Asian | Wong et al. |
| Sarsoza | Filipino | Other Asian | Wong et al. |
| Sarte | Filipino | Other Asian | Wong et al. |
| Sarthou | Filipino | Other Asian | Wong et al. |
| Saruca | Filipino | Other Asian | Wong et al. |
| Sarup | Asian Indian | South Asian | Wong et al. |
| Saruwatari | Japanese | Other Asian | Wong et al. |
| Sarvaiya | Asian Indian | South Asian | Wong et al. |
| Sarwal | Asian Indian | South Asian | Wong et al. |
| Sarwar |  | South Asian | Lim et al. |
| Sarwat |  | South Asian | Lim et al. |
| Sarza | Filipino | Other Asian | Wong et al. |
| Sasa | Japanese | Other Asian | Wong et al. |
| Sasabe | Japanese | Other Asian | Wong et al. |
| Sasada | Japanese | Other Asian | Wong et al. |
| Sasagawa | Japanese | Other Asian | Wong et al. |
| Sasahara | Japanese | Other Asian | Wong et al. |
| Sasai | Japanese | Other Asian | Wong et al. |
| Sasajima | Japanese | Other Asian | Wong et al. |
| Sasakawa | Japanese | Other Asian | Wong et al. |
| Sasaki | Japanese | Other Asian | Wong et al. |
| Sasakura | Japanese | Other Asian | Wong et al. |
| Sasamori | Japanese | Other Asian | Wong et al. |
| Sasamoto | Japanese | Other Asian | Wong et al. |
| Sasan | Filipino | Other Asian | Wong et al. |
| Sasano | Japanese | Other Asian | Wong et al. |
| Sasao | Japanese | Other Asian | Wong et al. |
| Sasaoka | Japanese | Other Asian | Wong et al. |
| Sasayama | Japanese | Other Asian | Wong et al. |
| Sase | Japanese | Other Asian | Wong et al. |
| Sasis | Filipino | Other Asian | Wong et al. |
| Sason | Filipino | Other Asian | Wong et al. |
| Sastri | Asian Indian | South Asian | Wong et al. |
| Sastry | Asian Indian | South Asian | Kranick et al. |
| Sasuman | Filipino | Other Asian | Wong et al. |
| Sata | Japanese | Other Asian | Wong et al. |
| Satake | Japanese | Other Asian | Wong et al. |
| Sathe | Asian Indian | South Asian | Wong et al. |
| Sathi |  | South Asian | Lim et al. |
| Sathianathan | Asian Indian | South Asian | Wong et al. |
| Sathyanaraya | Asian Indian | South Asian | Wong et al. |
| Sato | Japanese | Other Asian | Kranick et al. |
| Satoh | Japanese | Other Asian | Wong et al. |
| Satomi | Japanese | Other Asian | Wong et al. |
| Satoo | Japanese | Other Asian | Wong et al. |
| Satorre | Filipino | Other Asian | Wong et al. |
| Satow | Japanese | Other Asian | Wong et al. |
| Satsatin | Filipino | Other Asian | Wong et al. |
| Satsuma | Japanese | Other Asian | Wong et al. |
| Sattar |  | South Asian | Lim et al. |
| Sattari |  | South Asian | Lim et al. |
| Saturnino | Filipino | Other Asian | Wong et al. |
| Saturnio | Filipino | Other Asian | Wong et al. |
| Saturno | Filipino | Other Asian | Wong et al. |
| Satyadevi |  | South Asian | Lim et al. |
| Satyanarayan | Asian Indian | South Asian | Wong et al. |
| Sau | Chinese | Chinese | Wong et al. |
| Saucelo | Filipino | Other Asian | Wong et al. |
| Sauco | Filipino | Other Asian | Wong et al. |
| Saulo | Filipino | Other Asian | Wong et al. |
| Saulog | Filipino | Other Asian | Wong et al. |
| Saunar | Filipino | Other Asian | Wong et al. |
| Saupan | Filipino | Other Asian | Wong et al. |
| Saura | Filipino | Other Asian | Wong et al. |
| Sauz | Filipino | Other Asian | Wong et al. |
| Sauza | Filipino | Other Asian | Wong et al. |
| Savalia | Asian Indian | South Asian | Wong et al. |
| Savani | Asian Indian | South Asian | Wong et al. |
| Savarimuthu | Asian Indian | South Asian | Wong et al. |
| Savdharia | Asian Indian | South Asian | Wong et al. |
| Savedra | Filipino | Other Asian | Wong et al. |
| Savella | Filipino | Other Asian | Wong et al. |
| Savellano | Filipino | Other Asian | Wong et al. |
| Savet | Filipino | Other Asian | Wong et al. |
| Savillo | Filipino | Other Asian | Wong et al. |
| Savkoor | Asian Indian | South Asian | Wong et al. |
| Savla | Asian Indian | South Asian | Wong et al. |
| Sawa | Japanese | Other Asian | Wong et al. |
| Sawabe | Japanese | Other Asian | Wong et al. |
| Sawada | Japanese | Other Asian | Wong et al. |
| Sawaguchi | Japanese | Other Asian | Wong et al. |
| Sawai | Japanese | Other Asian | Wong et al. |
| Sawaki | Japanese | Other Asian | Wong et al. |
| Sawal | Filipino | Other Asian | Wong et al. |
| Sawamoto | Japanese | Other Asian | Wong et al. |
| Sawamura | Japanese | Other Asian | Wong et al. |
| Sawano | Japanese | Other Asian | Wong et al. |
| Sawant | Asian Indian | South Asian | Wong et al. |
| Sawh |  | South Asian | Lim et al. |
| Sawhny | Asian Indian | South Asian | Wong et al. |
| Sawi | Filipino | Other Asian | Wong et al. |
| Saxena | Asian Indian | South Asian | Wong et al. |
| Saya | Filipino | Other Asian | Wong et al. |
| Sayama | Japanese | Other Asian | Wong et al. |
| Sayan | Filipino | Other Asian | Wong et al. |
| Sayani | Asian Indian | South Asian | Wong et al. |
| Sayas | Filipino | Other Asian | Wong et al. |
| Sayco | Filipino | Other Asian | Wong et al. |
| Saycon | Filipino | Other Asian | Wong et al. |
| Sayed | Asian Indian | South Asian | Wong et al. |
| Sayeda |  | South Asian | Lim et al. |
| Sayeed |  | South Asian | Lim et al. |
| Sayno | Filipino | Other Asian | Wong et al. |
| Sayo | Filipino | Other Asian | Wong et al. |
| Sayoc | Filipino | Other Asian | Wong et al. |
| Sayon | Filipino | Other Asian | Wong et al. |
| Sayson | Filipino | Other Asian | Wong et al. |
| Sayyed | Asian Indian | South Asian | Wong et al. |
| Sazon | Filipino | Other Asian | Wong et al. |
| Scaria | Asian Indian | South Asian | Wong et al. |
| Seamber |  | South Asian | Lim et al. |
| Seastres | Filipino | Other Asian | Wong et al. |
| Seballos | Filipino | Other Asian | Wong et al. |
| Sebarrotin | Filipino | Other Asian | Wong et al. |
| Secretario | Filipino | Other Asian | Wong et al. |
| Sedani | Asian Indian | South Asian | Wong et al. |
| Sedano | Filipino | Other Asian | Wong et al. |
| Sedeno | Filipino | Other Asian | Wong et al. |
| Sedigo | Filipino | Other Asian | Wong et al. |
| See | Malaysian; Filipino | Other Asian | Kranick et al. |
| Seebarran |  | South Asian | Lim et al. |
| Seecharan |  | South Asian | Lim et al. |
| Seecharran | Indonesian | Other Asian | Kranick et al. |
| Seedat | Asian Indian | South Asian | Wong et al. |
| Seegobind |  | South Asian | Lim et al. |
| Seema |  | South Asian | Lim et al. |
| Seemangal |  | South Asian | Lim et al. |
| Seen | Chinese | Chinese | Wong et al. |
| Seenauth |  | South Asian | Lim et al. |
| Seepersad |  | South Asian | Lim et al. |
| Seepersaud |  | South Asian | Lim et al. |
| Seeraj |  | South Asian | Lim et al. |
| Seeram | Asian Indian | South Asian | Kranick et al. |
| Seeratan |  | South Asian | Lim et al. |
| Seetharam | Asian Indian | South Asian | Wong et al. |
| Seetharaman | Asian Indian | South Asian | Wong et al. |
| Seeto | Chinese | Chinese | Wong et al. |
| Seetoo | Chinese | Chinese | Wong et al. |
| Seetram |  | South Asian | Lim et al. |
| Segarra | Filipino | Other Asian | Wong et al. |
| Segawa | Japanese | Other Asian | Wong et al. |
| Segismundo | Filipino | Other Asian | Wong et al. |
| Segovia | Filipino | Other Asian | Wong et al. |
| Seguban | Filipino | Other Asian | Wong et al. |
| Seguchi | Japanese | Other Asian | Wong et al. |
| Seguerra | Filipino | Other Asian | Wong et al. |
| Segui | Filipino | Other Asian | Wong et al. |
| Seguisabal | Filipino | Other Asian | Wong et al. |
| Segundo | Filipino | Other Asian | Wong et al. |
| Segura | Filipino | Other Asian | Wong et al. |
| Seguritan | Filipino | Other Asian | Wong et al. |
| Sehdev | Asian Indian | South Asian | Wong et al. |
| Sehgal | Asian Indian | South Asian | Wong et al. |
| Sehwani | Filipino | Other Asian | Wong et al. |
| Sei | Chinese | Chinese | Wong et al. |
| Seid | Chinese | Chinese | Wong et al. |
| Seiji | Japanese | Other Asian | Wong et al. |
| Seike | Japanese | Other Asian | Wong et al. |
| Seiki | Japanese | Other Asian | Wong et al. |
| Seikichi | Japanese | Other Asian | Wong et al. |
| Seino | Japanese | Other Asian | Wong et al. |
| Sejuty |  | South Asian | Lim et al. |
| Sekender |  | South Asian | Lim et al. |
| Sekhar | Asian Indian | South Asian | Wong et al. |
| Sekharan | Asian Indian | South Asian | Wong et al. |
| Sekhon | Asian Indian | South Asian | Wong et al. |
| Sekhri | Asian Indian | South Asian | Wong et al. |
| Seki | Japanese | Other Asian | Wong et al. |
| Sekido | Japanese | Other Asian | Wong et al. |
| Sekiguchi | Japanese | Other Asian | Wong et al. |
| Sekihara | Japanese | Other Asian | Wong et al. |
| Sekikawa | Japanese | Other Asian | Wong et al. |
| Sekimoto | Japanese | Other Asian | Wong et al. |
| Sekine | Japanese | Other Asian | Wong et al. |
| Sekino | Japanese | Other Asian | Wong et al. |
| Sekiya | Japanese | Other Asian | Wong et al. |
| Sekizawa | Japanese | Other Asian | Wong et al. |
| Seko | Japanese | Other Asian | Wong et al. |
| Selerio | Filipino | Other Asian | Wong et al. |
| Selga | Filipino | Other Asian | Wong et al. |
| Selim | Filipino | Other Asian | Wong et al. |
| Sellona | Filipino | Other Asian | Wong et al. |
| Selorio | Filipino | Other Asian | Wong et al. |
| Seludo | Filipino | Other Asian | Wong et al. |
| Selvaraj | Asian Indian | South Asian | Wong et al. |
| Semana | Filipino | Other Asian | Wong et al. |
| Semba | Japanese | Other Asian | Wong et al. |
| Semblante | Filipino | Other Asian | Wong et al. |
| Sembrano | Filipino | Other Asian | Wong et al. |
| Semilla | Filipino | Other Asian | Wong et al. |
| Sempio | Filipino | Other Asian | Wong et al. |
| Sen | Asian Indian; Chinese; Bangladeshi | South Asian | Kranick et al. |
| Senador | Filipino | Other Asian | Wong et al. |
| Senaha | Japanese | Other Asian | Wong et al. |
| Senato | Filipino | Other Asian | Wong et al. |
| Senba | Japanese | Other Asian | Wong et al. |
| Senda | Japanese | Other Asian | Wong et al. |
| Sendaydiego | Filipino | Other Asian | Wong et al. |
| Senense | Filipino | Other Asian | Wong et al. |
| Seneres | Filipino | Other Asian | Wong et al. |
| Seneris | Filipino | Other Asian | Wong et al. |
| Senga | Japanese | Other Asian | Wong et al. |
| Sengco | Filipino | Other Asian | Wong et al. |
| Sengoku | Japanese | Other Asian | Wong et al. |
| Sengson | Filipino | Other Asian | Wong et al. |
| Sengupta |  | South Asian | Lim et al. |
| Senh | Vietnamese | Other Asian | Wong et al. |
| Senica | Filipino | Other Asian | Wong et al. |
| Seno | Filipino | Other Asian | Wong et al. |
| Senoo | Japanese | Other Asian | Wong et al. |
| Senoran | Filipino | Other Asian | Wong et al. |
| Sensano | Filipino | Other Asian | Wong et al. |
| Senson | Filipino | Other Asian | Wong et al. |
| Seo | Korean | Other Asian | Kranick et al. |
| Seok | Korean | Other Asian | Kranick et al. |
| Seolall |  | South Asian | Lim et al. |
| Seonarain |  | South Asian | Lim et al. |
| Seong | Korean | Other Asian | Kranick et al. |
| Seow | Malaysian | Other Asian | Kranick et al. |
| Separa | Filipino | Other Asian | Wong et al. |
| Sepulveda | Filipino | Other Asian | Wong et al. |
| Sequerra | Filipino | Other Asian | Wong et al. |
| Sera | Japanese | Other Asian | Wong et al. |
| Serafica | Filipino | Other Asian | Wong et al. |
| Serafico | Filipino | Other Asian | Wong et al. |
| Serame | Filipino | Other Asian | Wong et al. |
| Seranilla | Filipino | Other Asian | Wong et al. |
| Serano | Filipino | Other Asian | Wong et al. |
| Serapio | Filipino | Other Asian | Wong et al. |
| Serapion | Filipino | Other Asian | Wong et al. |
| Serdena | Filipino | Other Asian | Wong et al. |
| Serdenia | Filipino | Other Asian | Wong et al. |
| Sergodhia |  | South Asian | Lim et al. |
| Seril | Filipino | Other Asian | Wong et al. |
| Serion | Filipino | Other Asian | Wong et al. |
| Seriosa | Filipino | Other Asian | Wong et al. |
| Serizawa | Japanese | Other Asian | Wong et al. |
| Serker |  | South Asian | Lim et al. |
| Sermonia | Filipino | Other Asian | Wong et al. |
| Serna | Filipino | Other Asian | Wong et al. |
| Sernal | Filipino | Other Asian | Wong et al. |
| Serquina | Filipino | Other Asian | Wong et al. |
| Serquinia | Filipino | Other Asian | Wong et al. |
| Serran | Filipino | Other Asian | Wong et al. |
| Serrano | Filipino | Other Asian | Wong et al. |
| Serrao | Asian Indian | South Asian | Wong et al. |
| Serraon | Filipino | Other Asian | Wong et al. |
| Servando | Filipino | Other Asian | Wong et al. |
| Servanez | Filipino | Other Asian | Wong et al. |
| Servano | Filipino | Other Asian | Wong et al. |
| Servida | Filipino | Other Asian | Wong et al. |
| Servidad | Filipino | Other Asian | Wong et al. |
| Servillon | Filipino | Other Asian | Wong et al. |
| Servito | Filipino | Other Asian | Wong et al. |
| Sese | Filipino | Other Asian | Wong et al. |
| Seshadri | Asian Indian | South Asian | Wong et al. |
| Seshagiri | Asian Indian | South Asian | Kranick et al. |
| Seshan | Asian Indian | South Asian | Kranick et al. |
| Sesmundo | Filipino | Other Asian | Wong et al. |
| Sesoko | Japanese | Other Asian | Wong et al. |
| Sespene | Filipino | Other Asian | Wong et al. |
| Set | Chinese | Chinese | Wong et al. |
| Seta | Japanese | Other Asian | Wong et al. |
| Seth | Asian Indian | South Asian | Wong et al. |
| Sethi |  | South Asian | Lim et al. |
| Sethia | Asian Indian | South Asian | Wong et al. |
| Sethna | Asian Indian | South Asian | Wong et al. |
| Sethuraman | Asian Indian | South Asian | Wong et al. |
| Seto | Chinese | Chinese | Wong et al. |
| Setoguchi | Japanese | Other Asian | Wong et al. |
| Setyadharma | Indonesian | Other Asian | Kranick et al. |
| Seu | Chinese | Chinese | Wong et al. |
| Seulakhan |  | South Asian | Lim et al. |
| Seunarine |  | South Asian | Lim et al. |
| Seung | Chinese | Chinese | Wong et al. |
| Seva | Filipino | Other Asian | Wong et al. |
| Sevak | Asian Indian | South Asian | Wong et al. |
| Severo | Filipino | Other Asian | Wong et al. |
| Seveses | Filipino | Other Asian | Wong et al. |
| Sevidal | Filipino | Other Asian | Wong et al. |
| Sevilla | Filipino | Other Asian | Kranick et al. |
| Sevillano | Filipino | Other Asian | Wong et al. |
| Sevilleja | Filipino | Other Asian | Wong et al. |
| Sevillo | Filipino | Other Asian | Wong et al. |
| Sew | Chinese | Chinese | Wong et al. |
| Seya | Japanese | Other Asian | Wong et al. |
| Sezaki | Japanese | Other Asian | Wong et al. |
| Sezto | Chinese | Chinese | Wong et al. |
| Sha | Chinese | Chinese | Wong et al. |
| Shabbir |  | South Asian | Lim et al. |
| Shafi |  | South Asian | Lim et al. |
| Shafiq | Asian Indian | South Asian | Wong et al. |
| Shah | Asian Indian; Bangladeshi | South Asian | Kranick et al. |
| Shaha |  | South Asian | Lim et al. |
| Shahabuddin | Bangladeshi | South Asian | Kranick et al. |
| Shahana | Bangladeshi; Asian Indian | South Asian | Kranick et al. |
| Shahanaj |  | South Asian | Lim et al. |
| Shahbaz |  | South Asian | Lim et al. |
| Shahed |  | South Asian | Lim et al. |
| Shaheda |  | South Asian | Lim et al. |
| Shaheed |  | South Asian | Lim et al. |
| Shaheen |  | South Asian | Lim et al. |
| Shahi | Asian Indian | South Asian | Wong et al. |
| Shahid |  | South Asian | Lim et al. |
| Shahida |  | South Asian | Lim et al. |
| Shahidullah |  | South Asian | Lim et al. |
| Shahin |  | South Asian | Lim et al. |
| Shahjahan |  | South Asian | Lim et al. |
| Shahriar |  | South Asian | Lim et al. |
| Shahzadi |  | South Asian | Lim et al. |
| Shai | Chinese | Chinese | Wong et al. |
| Shaik | Asian Indian | South Asian | Wong et al. |
| Shaikh | Asian Indian; Pakistani | South Asian | Kranick et al. |
| Shaikhsiddique |  | South Asian | Lim et al. |
| Shaju |  | South Asian | Lim et al. |
| Shak | Chinese | Chinese | Wong et al. |
| Shake |  | South Asian | Lim et al. |
| Shaki |  | South Asian | Lim et al. |
| Shakir | Asian Indian | South Asian | Wong et al. |
| Shakoo |  | South Asian | Lim et al. |
| Shakoor | Asian Indian | South Asian | Wong et al. |
| Shakur | Asian Indian | South Asian | Wong et al. |
| Sham | Chinese | Chinese | Wong et al. |
| Shamanna | Asian Indian | South Asian | Wong et al. |
| Shamdany |  | South Asian | Lim et al. |
| Shami | Asian Indian | South Asian | Wong et al. |
| Shamim | Asian Indian | South Asian | Wong et al. |
| Shamji | Asian Indian | South Asian | Wong et al. |
| Shams | Asian Indian | South Asian | Wong et al. |
| Shamsheer |  | South Asian | Lim et al. |
| Shamsi | Asian Indian | South Asian | Wong et al. |
| Shamsuddin | Bangladeshi; Malaysian | South Asian | Kranick et al. |
| Shamsuddoha |  | South Asian | Lim et al. |
| Shamsul |  | South Asian | Lim et al. |
| Shamsundar |  | South Asian | Lim et al. |
| Shamsunnaharlily |  | South Asian | Lim et al. |
| Shamsuzzaman | Bangladeshi | South Asian | Kranick et al. |
| Shan | Chinese | Chinese | Wong et al. |
| Shanbhag | Asian Indian | South Asian | Wong et al. |
| Shang | Chinese | Chinese | Wong et al. |
| Shangkuan | Chinese | Chinese | Wong et al. |
| Shankar | Asian Indian | South Asian | Wong et al. |
| Shankerbhai | Asian Indian | South Asian | Wong et al. |
| Shanmugam | Asian Indian; Malaysian | South Asian | Kranick et al. |
| Shanmugasund | Asian Indian | South Asian | Wong et al. |
| Shantaram | Asian Indian | South Asian | Wong et al. |
| Shantilal | Asian Indian | South Asian | Wong et al. |
| Shanu |  | South Asian | Lim et al. |
| Shao | Chinese | Chinese | Kranick et al. |
| Shapayet |  | South Asian | Lim et al. |
| Shapna |  | South Asian | Lim et al. |
| Shar | Chinese | Chinese | Wong et al. |
| Sharaf | Asian Indian | South Asian | Wong et al. |
| Sharda | Asian Indian | South Asian | Wong et al. |
| Shareef | Asian Indian | South Asian | Wong et al. |
| Sharfuddin | Asian Indian | South Asian | Wong et al. |
| Shariar |  | South Asian | Lim et al. |
| Sharief | Asian Indian | South Asian | Wong et al. |
| Sharieff | Asian Indian | South Asian | Wong et al. |
| Sharif | Bangladeshi; Pakistani | South Asian | Kranick et al. |
| Shariff | Asian Indian | South Asian | Wong et al. |
| Sharifuddin |  | South Asian | Lim et al. |
| Sharma | Asian Indian | South Asian | Kranick et al. |
| Sharmin |  | South Asian | Lim et al. |
| Shastri | Asian Indian | South Asian | Wong et al. |
| Shastry | Asian Indian | South Asian | Wong et al. |
| Shau | Chinese | Chinese | Wong et al. |
| Shavari |  | South Asian | Lim et al. |
| Shaw |  | South Asian | Lim et al. |
| She | Chinese | Chinese | Wong et al. |
| Shee |  | South Asian | Lim et al. |
| Sheen | Chinese | Chinese | Wong et al. |
| Sheh | Chinese | Chinese | Wong et al. |
| Shei | Chinese | Chinese | Wong et al. |
| Sheih | Chinese | Chinese | Wong et al. |
| Sheikh | Asian Indian; Pakistani | South Asian | Kranick et al. |
| Shek | Asian Indian | South Asian | Kranick et al. |
| Shekhar | Asian Indian | South Asian | Kranick et al. |
| Sheladia | Asian Indian | South Asian | Wong et al. |
| Shelat | Asian Indian | South Asian | Wong et al. |
| Shelpy |  | South Asian | Lim et al. |
| Shem | Chinese | Chinese | Wong et al. |
| Shemul |  | South Asian | Lim et al. |
| Shen | Chinese | Chinese | Kranick et al. |
| Shende | Asian Indian | South Asian | Wong et al. |
| Sheng | Chinese | Chinese | Wong et al. |
| Shenoi | Asian Indian | South Asian | Wong et al. |
| Shenoy | Asian Indian | South Asian | Wong et al. |
| Sheong | Chinese | Chinese | Wong et al. |
| Shergill | Asian Indian | South Asian | Wong et al. |
| Sheriff |  | South Asian | Lim et al. |
| Sherpa |  | South Asian | Lim et al. |
| Sherring | Asian Indian | South Asian | Wong et al. |
| Sherwani | Asian Indian | South Asian | Wong et al. |
| Sheth | Asian Indian | South Asian | Kranick et al. |
| Shetty | Asian Indian | South Asian | Wong et al. |
| Shetye | Asian Indian | South Asian | Kranick et al. |
| Sheu | Chinese | Chinese | Wong et al. |
| Sheuly |  | South Asian | Lim et al. |
| Sheung | Chinese | Chinese | Wong et al. |
| Shew | Chinese | Chinese | Wong et al. |
| Shewly |  | South Asian | Lim et al. |
| Shey | Chinese | Chinese | Wong et al. |
| Shi | Chinese | Chinese | Kranick et al. |
| Shia | Chinese | Chinese | Wong et al. |
| Shiah | Chinese | Chinese | Wong et al. |
| Shian | Chinese | Chinese | Wong et al. |
| Shiang | Chinese | Chinese | Wong et al. |
| Shiao | Chinese | Chinese | Wong et al. |
| Shiau | Chinese | Chinese | Wong et al. |
| Shiba | Japanese | Other Asian | Wong et al. |
| Shibagaki | Japanese | Other Asian | Wong et al. |
| Shibakawa | Japanese | Other Asian | Wong et al. |
| Shibamoto | Japanese | Other Asian | Wong et al. |
| Shibano | Japanese | Other Asian | Wong et al. |
| Shibasaki | Japanese | Other Asian | Wong et al. |
| Shibata | Japanese | Other Asian | Wong et al. |
| Shibayama | Japanese | Other Asian | Wong et al. |
| Shibazaki | Japanese | Other Asian | Wong et al. |
| Shibue | Japanese | Other Asian | Wong et al. |
| Shibukawa | Japanese | Other Asian | Wong et al. |
| Shibusawa | Japanese | Other Asian | Wong et al. |
| Shibuta | Japanese | Other Asian | Wong et al. |
| Shibutani | Japanese | Other Asian | Wong et al. |
| Shibuya | Japanese | Other Asian | Wong et al. |
| Shic | Chinese | Chinese | Wong et al. |
| Shichi | Japanese | Other Asian | Wong et al. |
| Shichiri | Japanese | Other Asian | Wong et al. |
| Shida | Japanese | Other Asian | Wong et al. |
| Shidara | Japanese | Other Asian | Wong et al. |
| Shiddika |  | South Asian | Lim et al. |
| Shiddque |  | South Asian | Lim et al. |
| Shido | Japanese | Other Asian | Wong et al. |
| Shie | Chinese | Chinese | Wong et al. |
| Shieh | Chinese | Chinese | Wong et al. |
| Shiekh | Asian Indian | South Asian | Wong et al. |
| Shien | Chinese | Chinese | Wong et al. |
| Shiga | Japanese | Other Asian | Wong et al. |
| Shige | Japanese | Other Asian | Wong et al. |
| Shigehara | Japanese | Other Asian | Wong et al. |
| Shigehisa | Japanese | Other Asian | Wong et al. |
| Shigematsu | Japanese | Other Asian | Wong et al. |
| Shigemi | Japanese | Other Asian | Wong et al. |
| Shigemitsu | Japanese | Other Asian | Wong et al. |
| Shigemori | Japanese | Other Asian | Wong et al. |
| Shigemoto | Japanese | Other Asian | Wong et al. |
| Shigemura | Japanese | Other Asian | Wong et al. |
| Shigeno | Japanese | Other Asian | Wong et al. |
| Shigenobu | Japanese | Other Asian | Wong et al. |
| Shigeoka | Japanese | Other Asian | Wong et al. |
| Shigeru | Japanese | Other Asian | Wong et al. |
| Shigeta | Japanese | Other Asian | Wong et al. |
| Shigetomi | Japanese | Other Asian | Wong et al. |
| Shigeyama | Japanese | Other Asian | Wong et al. |
| Shigihara | Japanese | Other Asian | Wong et al. |
| Shih | Chinese | Chinese | Kranick et al. |
| Shiiba | Japanese | Other Asian | Wong et al. |
| Shiina | Japanese | Other Asian | Wong et al. |
| Shiino | Japanese | Other Asian | Wong et al. |
| Shijo | Japanese | Other Asian | Wong et al. |
| Shik | Korean | Other Asian | Wong et al. |
| Shikada | Japanese | Other Asian | Wong et al. |
| Shikama | Japanese | Other Asian | Wong et al. |
| Shikano | Japanese | Other Asian | Wong et al. |
| Shikari |  | South Asian | Lim et al. |
| Shikata | Japanese | Other Asian | Wong et al. |
| Shiki | Japanese | Other Asian | Wong et al. |
| Shikina | Japanese | Other Asian | Wong et al. |
| Shikiya | Japanese | Other Asian | Wong et al. |
| Shikrakar |  | South Asian | Lim et al. |
| Shil |  | South Asian | Lim et al. |
| Shill |  | South Asian | Lim et al. |
| Shilpy |  | South Asian | Lim et al. |
| Shim | Korean | Other Asian | Kranick et al. |
| Shima |  | South Asian | Lim et al. |
| Shimabuku | Japanese | Other Asian | Wong et al. |
| Shimabukuro | Japanese | Other Asian | Wong et al. |
| Shimada | Japanese | Other Asian | Wong et al. |
| Shimahara | Japanese | Other Asian | Wong et al. |
| Shimajiri | Japanese | Other Asian | Wong et al. |
| Shimakawa | Japanese | Other Asian | Wong et al. |
| Shimamoto | Japanese | Other Asian | Wong et al. |
| Shimamura | Japanese | Other Asian | Wong et al. |
| Shimane | Japanese | Other Asian | Wong et al. |
| Shimano | Japanese | Other Asian | Wong et al. |
| Shimaoka | Japanese | Other Asian | Wong et al. |
| Shimasaki | Japanese | Other Asian | Wong et al. |
| Shimatani | Japanese | Other Asian | Wong et al. |
| Shimauchi | Japanese | Other Asian | Wong et al. |
| Shimaya | Japanese | Other Asian | Wong et al. |
| Shimazaki | Japanese | Other Asian | Wong et al. |
| Shimazu | Japanese | Other Asian | Wong et al. |
| Shimbo | Japanese | Other Asian | Wong et al. |
| Shimizu | Japanese | Other Asian | Kranick et al. |
| Shimo | Japanese | Other Asian | Wong et al. |
| Shimoda | Japanese | Other Asian | Wong et al. |
| Shimodaira | Japanese | Other Asian | Wong et al. |
| Shimogori | Japanese | Other Asian | Wong et al. |
| Shimoi | Japanese | Other Asian | Wong et al. |
| Shimoji | Japanese | Other Asian | Wong et al. |
| Shimojima | Japanese | Other Asian | Wong et al. |
| Shimojo | Japanese | Other Asian | Wong et al. |
| Shimokawa | Japanese | Other Asian | Wong et al. |
| Shimokawara | Japanese | Other Asian | Wong et al. |
| Shimomoto | Japanese | Other Asian | Wong et al. |
| Shimomura | Japanese | Other Asian | Wong et al. |
| Shimono | Japanese | Other Asian | Wong et al. |
| Shimotsu | Japanese | Other Asian | Wong et al. |
| Shimotsuma | Japanese | Other Asian | Wong et al. |
| Shimoyama | Japanese | Other Asian | Wong et al. |
| Shimozato | Japanese | Other Asian | Wong et al. |
| Shimura | Japanese | Other Asian | Wong et al. |
| Shin | Korean | Other Asian | Kranick et al. |
| Shinada | Japanese | Other Asian | Wong et al. |
| Shinagawa | Japanese | Other Asian | Wong et al. |
| Shinde | Asian Indian | South Asian | Wong et al. |
| Shindo | Japanese | Other Asian | Wong et al. |
| Shing | Chinese | Chinese | Wong et al. |
| Shingai | Japanese | Other Asian | Wong et al. |
| Shingaki | Japanese | Other Asian | Wong et al. |
| Shingh |  | South Asian | Lim et al. |
| Shingu | Japanese | Other Asian | Wong et al. |
| Shinhira | Japanese | Other Asian | Wong et al. |
| Shinichiro | Japanese | Other Asian | Wong et al. |
| Shinji | Japanese | Other Asian | Wong et al. |
| Shinjo | Japanese | Other Asian | Wong et al. |
| Shinkai | Japanese | Other Asian | Wong et al. |
| Shinkawa | Japanese | Other Asian | Wong et al. |
| Shinmen | Japanese | Other Asian | Wong et al. |
| Shinmoto | Japanese | Other Asian | Wong et al. |
| Shino | Japanese | Other Asian | Wong et al. |
| Shinoda | Japanese | Other Asian | Wong et al. |
| Shinohara | Japanese | Other Asian | Wong et al. |
| Shinomiya | Japanese | Other Asian | Wong et al. |
| Shinozaki | Japanese | Other Asian | Wong et al. |
| Shinozuka | Japanese | Other Asian | Wong et al. |
| Shinsato | Japanese | Other Asian | Wong et al. |
| Shintaku | Japanese | Other Asian | Wong et al. |
| Shintani | Japanese | Other Asian | Wong et al. |
| Shinya | Japanese | Other Asian | Wong et al. |
| Shinzato | Japanese | Other Asian | Wong et al. |
| Shio | Chinese | Chinese | Wong et al. |
| Shiobara | Japanese | Other Asian | Wong et al. |
| Shioda | Japanese | Other Asian | Wong et al. |
| Shioiri | Japanese | Other Asian | Wong et al. |
| Shioji | Japanese | Other Asian | Wong et al. |
| Shiojima | Japanese | Other Asian | Wong et al. |
| Shiokawa | Japanese | Other Asian | Wong et al. |
| Shiomi | Japanese | Other Asian | Wong et al. |
| Shion | Chinese | Chinese | Wong et al. |
| Shiono | Japanese | Other Asian | Wong et al. |
| Shionoya | Japanese | Other Asian | Wong et al. |
| Shiosaki | Japanese | Other Asian | Wong et al. |
| Shiota | Japanese | Other Asian | Wong et al. |
| Shiotani | Japanese | Other Asian | Wong et al. |
| Shiotsuki | Japanese | Other Asian | Wong et al. |
| Shioya | Japanese | Other Asian | Wong et al. |
| Shiozaki | Japanese | Other Asian | Wong et al. |
| Shiozawa | Japanese | Other Asian | Wong et al. |
| Shirado | Japanese | Other Asian | Wong et al. |
| Shirahama | Japanese | Other Asian | Wong et al. |
| Shirahata | Japanese | Other Asian | Wong et al. |
| Shirai | Japanese | Other Asian | Wong et al. |
| Shiraishi | Japanese | Other Asian | Wong et al. |
| Shiraiwa | Japanese | Other Asian | Wong et al. |
| Shiraj |  | South Asian | Lim et al. |
| Shirakata | Japanese | Other Asian | Wong et al. |
| Shirakawa | Japanese | Other Asian | Wong et al. |
| Shiraki | Japanese | Other Asian | Wong et al. |
| Shirakura | Japanese | Other Asian | Wong et al. |
| Shirali | Asian Indian | South Asian | Wong et al. |
| Shiramizu | Japanese | Other Asian | Wong et al. |
| Shirasaka | Japanese | Other Asian | Wong et al. |
| Shirasaki | Japanese | Other Asian | Wong et al. |
| Shirasawa | Japanese | Other Asian | Wong et al. |
| Shirasu | Japanese | Other Asian | Wong et al. |
| Shirata | Japanese | Other Asian | Wong et al. |
| Shirato | Japanese | Other Asian | Wong et al. |
| Shiratori | Japanese | Other Asian | Wong et al. |
| Shirayama | Japanese | Other Asian | Wong et al. |
| Shirazi | Asian Indian | South Asian | Wong et al. |
| Shirke | Asian Indian | South Asian | Wong et al. |
| Shiro | Japanese | Other Asian | Wong et al. |
| Shirodkar | Asian Indian | South Asian | Wong et al. |
| Shirokawa | Japanese | Other Asian | Wong et al. |
| Shiroma | Japanese | Other Asian | Wong et al. |
| Shirota | Japanese | Other Asian | Wong et al. |
| Shiroyama | Japanese | Other Asian | Wong et al. |
| Shishida | Japanese | Other Asian | Wong et al. |
| Shishido | Japanese | Other Asian | Wong et al. |
| Shishikura | Japanese | Other Asian | Wong et al. |
| Shit | Chinese | Chinese | Wong et al. |
| Shitara | Japanese | Other Asian | Wong et al. |
| Shito | Japanese | Other Asian | Wong et al. |
| Shiu | Chinese | Chinese | Wong et al. |
| Shiue | Chinese | Chinese | Wong et al. |
| Shiung | Chinese | Chinese | Wong et al. |
| Shivabhai | Asian Indian | South Asian | Wong et al. |
| Shivdasani | Asian Indian | South Asian | Wong et al. |
| Shivjattan |  | South Asian | Lim et al. |
| Shivji | Asian Indian | South Asian | Wong et al. |
| Shivprasad |  | South Asian | Lim et al. |
| Shiwcharan |  | South Asian | Lim et al. |
| Shiwdat |  | South Asian | Lim et al. |
| Shiwnath |  | South Asian | Lim et al. |
| Shizuka | Japanese | Other Asian | Wong et al. |
| Sho | Chinese | Chinese | Wong et al. |
| Shoaib | Asian Indian | South Asian | Wong et al. |
| Shoda | Japanese | Other Asian | Wong et al. |
| Shodhan | Asian Indian | South Asian | Wong et al. |
| Shohag |  | South Asian | Lim et al. |
| Shohel |  | South Asian | Lim et al. |
| Shoji | Japanese | Other Asian | Kranick et al. |
| Shokar | Asian Indian | South Asian | Wong et al. |
| Shokat |  | South Asian | Lim et al. |
| Shoker | Asian Indian | South Asian | Wong et al. |
| Shome | Asian Indian | South Asian | Wong et al. |
| Shomi |  | South Asian | Lim et al. |
| Shon | Korean | Other Asian | Kranick et al. |
| Shong | Chinese | Chinese | Wong et al. |
| Shono | Japanese | Other Asian | Wong et al. |
| Shoon | Chinese | Chinese | Wong et al. |
| Shori | Asian Indian | South Asian | Wong et al. |
| Shou | Chinese | Chinese | Wong et al. |
| Shrestha | Nepali | South Asian | Kranick et al. |
| Shrikant |  | South Asian | Lim et al. |
| Shrivastava | Asian Indian | South Asian | Wong et al. |
| Shroff | Asian Indian | South Asian | Wong et al. |
| Shu | Chinese | Chinese | Wong et al. |
| Shuai | Chinese | Chinese | Wong et al. |
| Shue | Chinese | Chinese | Wong et al. |
| Shueh | Chinese | Chinese | Wong et al. |
| Shuen | Chinese | Chinese | Wong et al. |
| Shui | Chinese | Chinese | Kranick et al. |
| Shukla | Asian Indian | South Asian | Wong et al. |
| Shum | Chinese | Chinese | Kranick et al. |
| Shumi |  | South Asian | Lim et al. |
| Shumon |  | South Asian | Lim et al. |
| Shun | Chinese | Chinese | Wong et al. |
| Shung | Chinese | Chinese | Wong et al. |
| Shuto | Japanese | Other Asian | Wong et al. |
| Shyam | Asian Indian | South Asian | Wong et al. |
| Shyn | Korean | Other Asian | Wong et al. |
| Shyong | Chinese | Chinese | Wong et al. |
| Shyr | Chinese | Chinese | Wong et al. |
| Shyu | Chinese | Chinese | Wong et al. |
| Shyy | Chinese | Chinese | Wong et al. |
| Si | Chinese | Chinese | Wong et al. |
| Sia | Filipino | Other Asian | Wong et al. |
| Siacunco | Filipino | Other Asian | Wong et al. |
| Siador | Filipino | Other Asian | Wong et al. |
| Siam | Chinese | Chinese | Wong et al. |
| Sian | Filipino | Other Asian | Wong et al. |
| Sianghio | Filipino | Other Asian | Wong et al. |
| Siao | Chinese | Chinese | Wong et al. |
| Siaotong | Filipino | Other Asian | Wong et al. |
| Siapno | Filipino | Other Asian | Wong et al. |
| Siasat | Filipino | Other Asian | Wong et al. |
| Siasoco | Filipino | Other Asian | Wong et al. |
| Siason | Filipino | Other Asian | Wong et al. |
| Siat | Filipino | Other Asian | Wong et al. |
| Siazon | Filipino | Other Asian | Wong et al. |
| Sibal | Filipino | Other Asian | Wong et al. |
| Sibayan | Filipino | Other Asian | Wong et al. |
| Sibia | Asian Indian | South Asian | Wong et al. |
| Sibonga | Filipino | Other Asian | Wong et al. |
| Sibtain |  | South Asian | Lim et al. |
| Sibucao | Filipino | Other Asian | Wong et al. |
| Sibug | Filipino | Other Asian | Wong et al. |
| Sibulo | Filipino | Other Asian | Wong et al. |
| Sic | Chinese | Chinese | Wong et al. |
| Sicam | Filipino | Other Asian | Wong et al. |
| Sicangco | Filipino | Other Asian | Wong et al. |
| Sicat | Filipino | Other Asian | Wong et al. |
| Sichon | Filipino | Other Asian | Wong et al. |
| Sico | Filipino | Other Asian | Wong et al. |
| Sid | Chinese | Chinese | Wong et al. |
| Siddappa | Asian Indian | South Asian | Wong et al. |
| Siddigi | Asian Indian | South Asian | Wong et al. |
| Siddika |  | South Asian | Lim et al. |
| Siddiki |  | South Asian | Lim et al. |
| Siddioui | Asian Indian | South Asian | Wong et al. |
| Siddiq | Asian Indian | South Asian | Wong et al. |
| Siddiqa |  | South Asian | Lim et al. |
| Siddiqi | Pakistani | South Asian | Kranick et al. |
| Siddiqua |  | South Asian | Lim et al. |
| Siddique | Asian Indian | South Asian | Wong et al. |
| Siddiquee |  | South Asian | Lim et al. |
| Siddiqui | Pakistani | South Asian | Kranick et al. |
| Siddiquie |  | South Asian | Lim et al. |
| Sideco | Filipino | Other Asian | Wong et al. |
| Sidhu | Asian Indian | South Asian | Wong et al. |
| Sidhwa | Asian Indian | South Asian | Wong et al. |
| Sien | Chinese | Chinese | Wong et al. |
| Siendo | Filipino | Other Asian | Wong et al. |
| Sierra |  | South Asian | Lim et al. |
| Siew | Chinese | Chinese | Wong et al. |
| Signey | Filipino | Other Asian | Wong et al. |
| Signo | Filipino | Other Asian | Wong et al. |
| Sigua | Filipino | Other Asian | Wong et al. |
| Siguenza | Filipino | Other Asian | Wong et al. |
| Sih | Chinese | Chinese | Wong et al. |
| Sihota | Asian Indian | South Asian | Wong et al. |
| Sik | Korean | Other Asian | Wong et al. |
| Sikand | Asian Indian | South Asian | Wong et al. |
| Sikat | Filipino | Other Asian | Wong et al. |
| Sikdar | Asian Indian | South Asian | Wong et al. |
| Sikder |  | South Asian | Lim et al. |
| Sikka | Asian Indian | South Asian | Wong et al. |
| Sikri | Asian Indian | South Asian | Wong et al. |
| Sil | Korean | Other Asian | Wong et al. |
| Silagan | Filipino | Other Asian | Wong et al. |
| Silan | Filipino | Other Asian | Wong et al. |
| Silang | Filipino | Other Asian | Wong et al. |
| Silangcruz | Filipino | Other Asian | Wong et al. |
| Silao | Filipino | Other Asian | Wong et al. |
| Silapan | Filipino | Other Asian | Wong et al. |
| Silerio | Filipino | Other Asian | Wong et al. |
| Silla | Filipino | Other Asian | Wong et al. |
| Sillona | Filipino | Other Asian | Wong et al. |
| Silo | Filipino | Other Asian | Wong et al. |
| Silos | Filipino | Other Asian | Wong et al. |
| Silva | Filipino | Other Asian | Wong et al. |
| Silvanetto | Filipino | Other Asian | Wong et al. |
| Silvano | Filipino | Other Asian | Wong et al. |
| Silverio | Filipino | Other Asian | Wong et al. |
| Silvestre | Filipino | Other Asian | Wong et al. |
| Silvino | Filipino | Other Asian | Wong et al. |
| Silvosa | Filipino | Other Asian | Kranick et al. |
| Sim | Korean | Other Asian | Wong et al. |
| Simangan | Filipino | Other Asian | Wong et al. |
| Simbajon | Filipino | Other Asian | Wong et al. |
| Simbe | Filipino | Other Asian | Wong et al. |
| Simbol | Filipino | Other Asian | Wong et al. |
| Simbra | Filipino | Other Asian | Wong et al. |
| Simbre | Filipino | Other Asian | Wong et al. |
| Simbulan | Filipino | Other Asian | Wong et al. |
| Simeon |  | South Asian | Lim et al. |
| Simpao | Filipino | Other Asian | Wong et al. |
| Simpauco | Filipino | Other Asian | Wong et al. |
| Simpelo | Filipino | Other Asian | Wong et al. |
| Simpliciano | Filipino | Other Asian | Wong et al. |
| Simsiman | Filipino | Other Asian | Wong et al. |
| Simsuangco | Filipino | Other Asian | Wong et al. |
| Simuangco | Filipino | Other Asian | Wong et al. |
| Sin | Burmese; Chinese | Other Asian | Kranick et al. |
| Sinaban | Filipino | Other Asian | Wong et al. |
| Sinaguinan | Filipino | Other Asian | Wong et al. |
| Sinah | Asian Indian | South Asian | Wong et al. |
| Sinan | Asian Indian | South Asian | Wong et al. |
| Sinang | Filipino | Other Asian | Wong et al. |
| Sinay | Filipino | Other Asian | Wong et al. |
| Sinch | Asian Indian | South Asian | Wong et al. |
| Sinchongco | Filipino | Other Asian | Wong et al. |
| Sinco | Filipino | Other Asian | Wong et al. |
| Sindac | Filipino | Other Asian | Wong et al. |
| Sindayen | Filipino | Other Asian | Wong et al. |
| Sindhu | Asian Indian | South Asian | Wong et al. |
| Sindiong | Filipino | Other Asian | Wong et al. |
| Sinense | Filipino | Other Asian | Wong et al. |
| Sinfuego | Filipino | Other Asian | Wong et al. |
| Sing |  | South Asian | Lim et al. |
| Singal | Asian Indian | South Asian | Wong et al. |
| Singco | Filipino | Other Asian | Wong et al. |
| Singh | Asian Indian | South Asian | Kranick et al. |
| Singha | Asian Indian | South Asian | Wong et al. |
| Singhal | Asian Indian | South Asian | Wong et al. |
| Singhgill | Asian Indian | South Asian | Wong et al. |
| Singh-Sahi |  | South Asian | Lim et al. |
| Singhvi | Asian Indian | South Asian | Wong et al. |
| Singian | Filipino | Other Asian | Wong et al. |
| Singl | Asian Indian | South Asian | Wong et al. |
| Singm | Asian Indian | South Asian | Wong et al. |
| Singn | Asian Indian | South Asian | Wong et al. |
| Singson | Filipino | Other Asian | Wong et al. |
| Singu | Asian Indian | South Asian | Wong et al. |
| Singzon | Filipino | Other Asian | Wong et al. |
| Sinh | Vietnamese | Other Asian | Wong et al. |
| Sinha | Asian Indian | South Asian | Kranick et al. |
| Sinhkh |  | South Asian | Lim et al. |
| Sinio | Filipino | Other Asian | Wong et al. |
| Sinlao | Filipino | Other Asian | Wong et al. |
| Sinocruz | Filipino | Other Asian | Wong et al. |
| Sinsay | Filipino | Other Asian | Wong et al. |
| Sinsh | Asian Indian | South Asian | Wong et al. |
| Sinson | Filipino | Other Asian | Wong et al. |
| Sintos | Filipino | Other Asian | Wong et al. |
| Siochi | Filipino | Other Asian | Wong et al. |
| Sioco | Filipino | Other Asian | Wong et al. |
| Siojo | Filipino | Other Asian | Wong et al. |
| Sion | Filipino | Other Asian | Wong et al. |
| Siongco | Filipino | Other Asian | Wong et al. |
| Siopongco | Filipino | Other Asian | Wong et al. |
| Sioson | Filipino | Other Asian | Wong et al. |
| Siou | Chinese | Chinese | Wong et al. |
| Siow | Chinese | Chinese | Wong et al. |
| Sipin | Filipino | Other Asian | Wong et al. |
| Siplon | Filipino | Other Asian | Wong et al. |
| Siquig | Filipino | Other Asian | Wong et al. |
| Siquijor | Filipino | Other Asian | Wong et al. |
| Sira | Asian Indian | South Asian | Wong et al. |
| Siraj | Asian Indian | South Asian | Wong et al. |
| Sirajuddin | Asian Indian | South Asian | Wong et al. |
| Siraz |  | South Asian | Lim et al. |
| Sircar | Asian Indian | South Asian | Wong et al. |
| Siriban | Filipino | Other Asian | Wong et al. |
| Sirilan | Filipino | Other Asian | Wong et al. |
| Siripant | Thai | Other Asian | Kranick et al. |
| Sirohi | Asian Indian | South Asian | Wong et al. |
| Siruelo | Filipino | Other Asian | Wong et al. |
| Siruno | Filipino | Other Asian | Wong et al. |
| Sirur | Asian Indian | South Asian | Wong et al. |
| Sisante | Filipino | Other Asian | Wong et al. |
| Siscar | Filipino | Other Asian | Wong et al. |
| Sismaet | Filipino | Other Asian | Wong et al. |
| Sisodia | Asian Indian | South Asian | Wong et al. |
| Sison | Filipino | Other Asian | Kranick et al. |
| Sistoso | Filipino | Other Asian | Wong et al. |
| Sit | Chinese | Chinese | Wong et al. |
| Sitabkhan | Asian Indian | South Asian | Wong et al. |
| Sitaram | Asian Indian | South Asian | Wong et al. |
| Sitaraman | Asian Indian | South Asian | Wong et al. |
| Sitchon | Filipino | Other Asian | Wong et al. |
| Sitjar | Filipino | Other Asian | Wong et al. |
| Sito | Chinese | Chinese | Wong et al. |
| Situ | Chinese | Chinese | Wong et al. |
| Sityar | Filipino | Other Asian | Wong et al. |
| Siu | Chinese | Chinese | Kranick et al. |
| Sivaramakris | Asian Indian | South Asian | Wong et al. |
| Sivaraman | Asian Indian | South Asian | Wong et al. |
| Sivasubraman | Asian Indian | South Asian | Wong et al. |
| Sivila | Filipino | Other Asian | Wong et al. |
| Siwa | Filipino | Other Asian | Wong et al. |
| Siy | Filipino | Other Asian | Wong et al. |
| Skaria | Asian Indian | South Asian | Wong et al. |
| Skariah | Asian Indian | South Asian | Wong et al. |
| Skievaski | Filipino | Other Asian | Wong et al. |
| Slepouhoua |  | South Asian | Lim et al. |
| Snaer | Filipino | Other Asian | Wong et al. |
| Sngh | Asian Indian | South Asian | Wong et al. |
| So | Chinese; Cambodian | Chinese | Kranick et al. |
| Soans | Asian Indian | South Asian | Wong et al. |
| Sobejana | Filipino | Other Asian | Wong et al. |
| Soberano | Filipino | Other Asian | Wong et al. |
| Sobhan |  | South Asian | Lim et al. |
| Sobredo | Filipino | Other Asian | Wong et al. |
| Sobremisana | Filipino | Other Asian | Wong et al. |
| Sobremonte | Filipino | Other Asian | Wong et al. |
| Sobrepena | Filipino | Other Asian | Wong et al. |
| Sobrevilla | Filipino | Other Asian | Wong et al. |
| Sobrevinas | Filipino | Other Asian | Wong et al. |
| Sobti | Asian Indian | South Asian | Wong et al. |
| Socias | Filipino | Other Asian | Wong et al. |
| Socito | Filipino | Other Asian | Wong et al. |
| Soco | Filipino | Other Asian | Wong et al. |
| Socorro | Filipino | Other Asian | Wong et al. |
| Sodeyama | Japanese | Other Asian | Wong et al. |
| Sodha | Asian Indian | South Asian | Wong et al. |
| Sodhi | Asian Indian | South Asian | Wong et al. |
| Sodusta | Filipino | Other Asian | Wong et al. |
| Soeda | Japanese | Other Asian | Wong et al. |
| Soejima | Japanese | Other Asian | Wong et al. |
| Soetijono | Indonesian | Other Asian | Kranick et al. |
| Soga | Japanese | Other Asian | Wong et al. |
| Sogabe | Japanese | Other Asian | Wong et al. |
| Sogani | Asian Indian | South Asian | Wong et al. |
| Sogawa | Japanese | Other Asian | Wong et al. |
| Sogi | Japanese | Other Asian | Wong et al. |
| Sogo | Japanese | Other Asian | Wong et al. |
| Soh | Korean | Other Asian | Kranick et al. |
| Sohag |  | South Asian | Lim et al. |
| Sohal | Asian Indian | South Asian | Wong et al. |
| Sohan |  | South Asian | Lim et al. |
| Sohani | Asian Indian | South Asian | Wong et al. |
| Sohel |  | South Asian | Lim et al. |
| Sohi | Asian Indian | South Asian | Wong et al. |
| Sohma | Japanese | Other Asian | Wong et al. |
| Sohn | Korean | Other Asian | Kranick et al. |
| Soho | Chinese | Chinese | Wong et al. |
| Sohoni | Asian Indian | South Asian | Wong et al. |
| Soin | Asian Indian | South Asian | Wong et al. |
| Sokhi | Asian Indian | South Asian | Wong et al. |
| Solaiman |  | South Asian | Lim et al. |
| Solamo | Filipino | Other Asian | Wong et al. |
| Solancho | Filipino | Other Asian | Wong et al. |
| Solanki | Asian Indian | South Asian | Wong et al. |
| Solanky | Asian Indian | South Asian | Wong et al. |
| Solano | Filipino | Other Asian | Wong et al. |
| Solanoy | Filipino | Other Asian | Wong et al. |
| Solanzo | Filipino | Other Asian | Wong et al. |
| Solas | Filipino | Other Asian | Wong et al. |
| Solatre | Filipino | Other Asian | Wong et al. |
| Soldevilla | Filipino | Other Asian | Wong et al. |
| Soledad | Filipino | Other Asian | Wong et al. |
| Soleman |  | South Asian | Lim et al. |
| Soler | Filipino | Other Asian | Wong et al. |
| Soleta | Filipino | Other Asian | Wong et al. |
| Solidarios | Filipino | Other Asian | Wong et al. |
| Solidum | Filipino | Other Asian | Kranick et al. |
| Solima | Filipino | Other Asian | Wong et al. |
| Soliman | Filipino | Other Asian | Wong et al. |
| Solinap | Filipino | Other Asian | Wong et al. |
| Solis | Filipino | Other Asian | Wong et al. |
| Solito | Filipino | Other Asian | Wong et al. |
| Soliva | Filipino | Other Asian | Wong et al. |
| Soliven | Filipino | Other Asian | Wong et al. |
| Solivio | Filipino | Other Asian | Wong et al. |
| Sollano | Filipino | Other Asian | Wong et al. |
| Soller | Filipino | Other Asian | Wong et al. |
| Solomon |  | South Asian | Lim et al. |
| Soloria | Filipino | Other Asian | Wong et al. |
| Soma | Japanese | Other Asian | Kranick et al. |
| Somabhai | Asian Indian | South Asian | Wong et al. |
| Somaiya | Asian Indian | South Asian | Wong et al. |
| Somal | Asian Indian | South Asian | Wong et al. |
| Soman | Asian Indian | South Asian | Wong et al. |
| Somani | Asian Indian | South Asian | Wong et al. |
| Somaroo |  | South Asian | Lim et al. |
| Somayajulu | Asian Indian | South Asian | Wong et al. |
| Sombero | Filipino | Other Asian | Wong et al. |
| Sombillo | Filipino | Other Asian | Wong et al. |
| Sombilon | Filipino | Other Asian | Wong et al. |
| Some | Chinese | Chinese | Wong et al. |
| Somedat |  | South Asian | Lim et al. |
| Somekawa | Japanese | Other Asian | Wong et al. |
| Somera | Filipino | Other Asian | Wong et al. |
| Someya | Japanese | Other Asian | Wong et al. |
| Somosierra | Filipino | Other Asian | Wong et al. |
| Somwar |  | South Asian | Lim et al. |
| Somwaru |  | South Asian | Lim et al. |
| Son | Korean | Other Asian | Kranick et al. |
| Sonam |  | South Asian | Lim et al. |
| Sonawala | Asian Indian | South Asian | Wong et al. |
| Soncuya | Filipino | Other Asian | Wong et al. |
| Sondhi | Asian Indian | South Asian | Kranick et al. |
| Sone | Japanese | Other Asian | Wong et al. |
| Soneji | Asian Indian | South Asian | Wong et al. |
| Song | Korean | Other Asian | Kranick et al. |
| Songalia | Filipino | Other Asian | Wong et al. |
| Songco | Filipino | Other Asian | Wong et al. |
| Songcuan | Filipino | Other Asian | Wong et al. |
| Soni | Asian Indian | South Asian | Wong et al. |
| Sonico | Filipino | Other Asian | Wong et al. |
| Sonido | Filipino | Other Asian | Wong et al. |
| Soniega | Filipino | Other Asian | Wong et al. |
| Sonnysingh |  | South Asian | Lim et al. |
| Sono | Japanese | Other Asian | Wong et al. |
| Sonobe | Japanese | Other Asian | Wong et al. |
| Sonoda | Japanese | Other Asian | Wong et al. |
| Sonoki | Japanese | Other Asian | Wong et al. |
| Sonoyama | Japanese | Other Asian | Wong et al. |
| Sonson | Filipino | Other Asian | Wong et al. |
| Sonu | Korean | Other Asian | Wong et al. |
| Sonza | Filipino | Other Asian | Wong et al. |
| Soo | Chinese | Chinese | Wong et al. |
| Sooch | Asian Indian | South Asian | Wong et al. |
| Sood | Asian Indian | South Asian | Wong et al. |
| Soofi | Asian Indian | South Asian | Wong et al. |
| Soohoo | Chinese | Chinese | Kranick et al. |
| Sook | Korean | Other Asian | Wong et al. |
| Sookdeo |  | South Asian | Lim et al. |
| Sooklall |  | South Asian | Lim et al. |
| Sooknanan | Trinidad & Tobago | Other Asian | Kranick et al. |
| Sookram | Trinidad & Tobago | Other Asian | Kranick et al. |
| Sool | Korean | Other Asian | Wong et al. |
| Soon | Chinese | Chinese | Wong et al. |
| Soong | Chinese | Chinese | Wong et al. |
| Sopariwala | Asian Indian | South Asian | Wong et al. |
| Soquena | Filipino | Other Asian | Wong et al. |
| Sorathia | Asian Indian | South Asian | Wong et al. |
| Soratorio | Filipino | Other Asian | Wong et al. |
| Sordilla | Filipino | Other Asian | Wong et al. |
| Soria | Filipino | Other Asian | Wong et al. |
| Soriaga | Filipino | Other Asian | Wong et al. |
| Soriano | Filipino | Other Asian | Kranick et al. |
| Sorianosos | Filipino | Other Asian | Wong et al. |
| Soriben | Filipino | Other Asian | Wong et al. |
| Soriente | Filipino | Other Asian | Wong et al. |
| Sorilla | Filipino | Other Asian | Wong et al. |
| Sorimachi | Japanese | Other Asian | Wong et al. |
| Sorio | Filipino | Other Asian | Wong et al. |
| Sornito | Filipino | Other Asian | Wong et al. |
| Sorongon | Filipino | Other Asian | Wong et al. |
| Sorra | Filipino | Other Asian | Wong et al. |
| Sortijas | Filipino | Other Asian | Wong et al. |
| Sosa | Filipino | Other Asian | Wong et al. |
| Sotelo | Filipino | Other Asian | Wong et al. |
| Sotero | Filipino | Other Asian | Wong et al. |
| Sotiangco | Filipino | Other Asian | Wong et al. |
| Soto | Filipino | Other Asian | Wong et al. |
| Sotto | Filipino | Other Asian | Wong et al. |
| Soundararaja | Asian Indian | South Asian | Wong et al. |
| Sousae | Chinese | Chinese | Wong et al. |
| Sow | Chinese | Chinese | Wong et al. |
| Soyangco | Filipino | Other Asian | Wong et al. |
| Sozon |  | South Asian | Lim et al. |
| Sra | Asian Indian | South Asian | Wong et al. |
| Sran | Asian Indian | South Asian | Wong et al. |
| Sreedharan | Asian Indian | South Asian | Wong et al. |
| Sreenivasan | Asian Indian | South Asian | Kranick et al. |
| Sricharoon | Thai | Other Asian | Kranick et al. |
| Sridharan | Asian Indian | South Asian | Wong et al. |
| Srikantaiah | Asian Indian | South Asian | Wong et al. |
| Srikantan | Asian Indian | South Asian | Wong et al. |
| Srikantiah | Asian Indian | South Asian | Wong et al. |
| Srinath | Asian Indian | South Asian | Wong et al. |
| Srinivas | Asian Indian | South Asian | Wong et al. |
| Srinivasa | Asian Indian | South Asian | Wong et al. |
| Srinivasacha | Asian Indian | South Asian | Wong et al. |
| Srinivasan | Asian Indian | South Asian | Kranick et al. |
| Srivastava | Asian Indian | South Asian | Kranick et al. |
| Sriwatte |  | South Asian | Lim et al. |
| Ssi | Korean | Other Asian | Wong et al. |
| Ssngh | Asian Indian | South Asian | Wong et al. |
| Staana | Filipino | Other Asian | Wong et al. |
| Stacruz | Filipino | Other Asian | Wong et al. |
| Staelena | Filipino | Other Asian | Wong et al. |
| Stagen | Filipino | Other Asian | Wong et al. |
| Staiglesia | Filipino | Other Asian | Wong et al. |
| Stamaria | Filipino | Other Asian | Wong et al. |
| Stamatelaky | Filipino | Other Asian | Wong et al. |
| Starosa | Filipino | Other Asian | Wong et al. |
| Stateresa | Filipino | Other Asian | Wong et al. |
| Stodomingo | Filipino | Other Asian | Wong et al. |
| Stotomas | Filipino | Other Asian | Wong et al. |
| Su | Chinese | Chinese | Kranick et al. |
| Suaco | Filipino | Other Asian | Wong et al. |
| Sual | Filipino | Other Asian | Wong et al. |
| Sualog | Filipino | Other Asian | Wong et al. |
| Suan | Filipino | Other Asian | Wong et al. |
| Suarez | Filipino | Other Asian | Wong et al. |
| Suasin | Filipino | Other Asian | Wong et al. |
| Suatengco | Filipino | Other Asian | Wong et al. |
| Suayan | Filipino | Other Asian | Wong et al. |
| Suazo | Filipino | Other Asian | Wong et al. |
| Suba | Filipino | Other Asian | Wong et al. |
| Subala | Filipino | Other Asian | Wong et al. |
| Suban | Filipino | Other Asian | Wong et al. |
| Subang | Filipino | Other Asian | Wong et al. |
| Subbaiah | Asian Indian | South Asian | Wong et al. |
| Subbaraman | Asian Indian | South Asian | Wong et al. |
| Subbarao | Asian Indian | South Asian | Wong et al. |
| Subbarayan | Asian Indian | South Asian | Wong et al. |
| Subbarayudu | Asian Indian | South Asian | Wong et al. |
| Subbiah | Asian Indian | South Asian | Wong et al. |
| Subedi | Nepali | South Asian | Kranick et al. |
| Subhan |  | South Asian | Lim et al. |
| Subhani | Asian Indian | South Asian | Wong et al. |
| Subia | Filipino | Other Asian | Wong et al. |
| Subiaga | Filipino | Other Asian | Wong et al. |
| Subida | Filipino | Other Asian | Kranick et al. |
| Subido | Filipino | Other Asian | Wong et al. |
| Subijano | Filipino | Other Asian | Wong et al. |
| Subillaga | Filipino | Other Asian | Wong et al. |
| Subrahmanian | Asian Indian | South Asian | Wong et al. |
| Subrahmanyam | Asian Indian | South Asian | Wong et al. |
| Subrahmanyan | Asian Indian | South Asian | Wong et al. |
| Subramani | Asian Indian | South Asian | Wong et al. |
| Subramania | Asian Indian | South Asian | Wong et al. |
| Subramaniai | Asian Indian | South Asian | Wong et al. |
| Subramaniam | Asian Indian | South Asian | Wong et al. |
| Subramanian | Asian Indian | South Asian | Wong et al. |
| Subramanium | Asian Indian | South Asian | Wong et al. |
| Subramanya | Asian Indian | South Asian | Wong et al. |
| Subramanyam | Asian Indian | South Asian | Kranick et al. |
| Subramanyan | Asian Indian | South Asian | Wong et al. |
| Sucaldito | Filipino | Other Asian | Wong et al. |
| Sucgang | Filipino | Other Asian | Wong et al. |
| Sud | Asian Indian | South Asian | Wong et al. |
| Suda | Japanese | Other Asian | Wong et al. |
| Sudan | Asian Indian | South Asian | Wong et al. |
| Sudaria | Filipino | Other Asian | Wong et al. |
| Sudario | Filipino | Other Asian | Wong et al. |
| Sudiacal | Filipino | Other Asian | Wong et al. |
| Sudo | Japanese | Other Asian | Wong et al. |
| Sudoh | Japanese | Other Asian | Wong et al. |
| Sue | Chinese | Chinese | Wong et al. |
| Sueda | Japanese | Other Asian | Wong et al. |
| Suehiro | Japanese | Other Asian | Wong et al. |
| Suela | Filipino | Other Asian | Wong et al. |
| Suelto | Filipino | Other Asian | Kranick et al. |
| Suematsu | Japanese | Other Asian | Wong et al. |
| Suen | Chinese | Chinese | Kranick et al. |
| Suenaga | Japanese | Other Asian | Wong et al. |
| Sueno | Filipino | Other Asian | Wong et al. |
| Sueoka | Japanese | Other Asian | Wong et al. |
| Suero | Filipino | Other Asian | Wong et al. |
| Suerte | Filipino | Other Asian | Wong et al. |
| Suetake | Japanese | Other Asian | Wong et al. |
| Suetos | Filipino | Other Asian | Wong et al. |
| Suetsugu | Japanese | Other Asian | Wong et al. |
| Suey | Chinese | Chinese | Wong et al. |
| Sueyoshi | Japanese | Other Asian | Wong et al. |
| Suez | Chinese | Chinese | Wong et al. |
| Sufi | Asian Indian | South Asian | Wong et al. |
| Sufian |  | South Asian | Lim et al. |
| Suficiencia | Filipino | Other Asian | Wong et al. |
| Suga | Japanese | Other Asian | Wong et al. |
| Sugahara | Japanese | Other Asian | Wong et al. |
| Sugai | Japanese | Other Asian | Wong et al. |
| Suganami | Japanese | Other Asian | Wong et al. |
| Sugano | Japanese | Other Asian | Wong et al. |
| Suganob | Filipino | Other Asian | Wong et al. |
| Suganuma | Japanese | Other Asian | Wong et al. |
| Sugata | Japanese | Other Asian | Wong et al. |
| Sugatan | Filipino | Other Asian | Wong et al. |
| Sugawa | Japanese | Other Asian | Wong et al. |
| Sugawara | Japanese | Other Asian | Wong et al. |
| Sugay | Filipino | Other Asian | Wong et al. |
| Sugaya | Japanese | Other Asian | Wong et al. |
| Sugi | Japanese | Other Asian | Wong et al. |
| Sugie | Japanese | Other Asian | Wong et al. |
| Sugihara | Japanese | Other Asian | Wong et al. |
| Sugimori | Japanese | Other Asian | Wong et al. |
| Sugimoto | Japanese | Other Asian | Wong et al. |
| Sugimura | Japanese | Other Asian | Wong et al. |
| Sugino | Japanese | Other Asian | Wong et al. |
| Sugioka | Japanese | Other Asian | Wong et al. |
| Sugisaki | Japanese | Other Asian | Wong et al. |
| Sugita | Japanese | Other Asian | Wong et al. |
| Sugitani | Japanese | Other Asian | Wong et al. |
| Sugito | Japanese | Other Asian | Wong et al. |
| Sugiura | Japanese | Other Asian | Kranick et al. |
| Sugiyama | Japanese | Other Asian | Kranick et al. |
| Sugizaki | Japanese | Other Asian | Wong et al. |
| Sugrim |  | South Asian | Lim et al. |
| Sugui | Filipino | Other Asian | Wong et al. |
| Suguitan | Filipino | Other Asian | Wong et al. |
| Suh | Korean | Other Asian | Kranick et al. |
| Suhara | Japanese | Other Asian | Wong et al. |
| Sui | Chinese | Chinese | Wong et al. |
| Suico | Filipino | Other Asian | Wong et al. |
| Suing | Filipino | Other Asian | Wong et al. |
| Suita | Japanese | Other Asian | Wong et al. |
| Sujo | Chinese | Chinese | Wong et al. |
| Suk | Korean | Other Asian | Wong et al. |
| Sukegawa | Japanese | Other Asian | Wong et al. |
| Sukhadia | Asian Indian | South Asian | Wong et al. |
| Sukhai |  | South Asian | Lim et al. |
| Sukhan |  | South Asian | Lim et al. |
| Sukhdeo |  | South Asian | Lim et al. |
| Sukhia | Asian Indian | South Asian | Wong et al. |
| Sukhnanan |  | South Asian | Lim et al. |
| Sukhnandan |  | South Asian | Lim et al. |
| Sukhram |  | South Asian | Lim et al. |
| Sukhu |  | South Asian | Lim et al. |
| Sukigara | Japanese | Other Asian | Wong et al. |
| Sukumaran | Asian Indian | South Asian | Wong et al. |
| Sul | Korean | Other Asian | Wong et al. |
| Sulaiman | Malaysian; Indonesian | Other Asian | Kranick et al. |
| Sulayman |  | South Asian | Lim et al. |
| Suleman | Asian Indian | South Asian | Wong et al. |
| Sulit | Filipino | Other Asian | Wong et al. |
| Sullano | Filipino | Other Asian | Wong et al. |
| Sulquiano | Filipino | Other Asian | Wong et al. |
| Sulse | Filipino | Other Asian | Wong et al. |
| Sultama |  | South Asian | Lim et al. |
| Sultan |  | South Asian | Lim et al. |
| Sultana | Bangladeshi | South Asian | Kranick et al. |
| Sum | Chinese | Chinese | Wong et al. |
| Suma | Japanese | Other Asian | Wong et al. |
| Sumabat | Filipino | Other Asian | Wong et al. |
| Sumagang | Filipino | Other Asian | Wong et al. |
| Sumagaysay | Filipino | Other Asian | Wong et al. |
| Sumagit | Filipino | Other Asian | Wong et al. |
| Sumague | Filipino | Other Asian | Wong et al. |
| Sumagui | Filipino | Other Asian | Wong et al. |
| Sumalbag | Filipino | Other Asian | Wong et al. |
| Sumalde | Filipino | Other Asian | Wong et al. |
| Sumalpong | Filipino | Other Asian | Wong et al. |
| Sumampong | Filipino | Other Asian | Wong et al. |
| Suman |  | South Asian | Lim et al. |
| Sumang | Filipino | Other Asian | Wong et al. |
| Sumangil | Filipino | Other Asian | Wong et al. |
| Sumaoang | Filipino | Other Asian | Wong et al. |
| Sumaray | Filipino | Other Asian | Wong et al. |
| Sumaria | Filipino | Other Asian | Wong et al. |
| Sumasar |  | South Asian | Lim et al. |
| Sumaway | Filipino | Other Asian | Kranick et al. |
| Sumaya | Filipino | Other Asian | Wong et al. |
| Sumayao | Filipino | Other Asian | Wong et al. |
| Sumaylo | Filipino | Other Asian | Wong et al. |
| Sumayo | Filipino | Other Asian | Wong et al. |
| Sumbang | Filipino | Other Asian | Wong et al. |
| Sumbillo | Filipino | Other Asian | Wong et al. |
| Sumcad | Filipino | Other Asian | Wong et al. |
| Sume |  | South Asian | Lim et al. |
| Sumera | Filipino | Other Asian | Wong et al. |
| Sumi |  | South Asian | Lim et al. |
| Sumibcay | Filipino | Other Asian | Wong et al. |
| Sumida | Japanese | Other Asian | Wong et al. |
| Sumikawa | Japanese | Other Asian | Wong et al. |
| Sumilang | Filipino | Other Asian | Wong et al. |
| Sumino | Japanese | Other Asian | Wong et al. |
| Sumira | Indonesian | Other Asian | Kranick et al. |
| Sumit |  | South Asian | Lim et al. |
| Sumita | Japanese | Other Asian | Wong et al. |
| Sumitani | Japanese | Other Asian | Wong et al. |
| Sumitomo | Japanese | Other Asian | Wong et al. |
| Sumiya | Japanese | Other Asian | Wong et al. |
| Sumiyama | Japanese | Other Asian | Wong et al. |
| Sumiyoshi | Japanese | Other Asian | Wong et al. |
| Summan | Asian Indian | South Asian | Wong et al. |
| Sumoba | Filipino | Other Asian | Wong et al. |
| Sumon |  | South Asian | Lim et al. |
| Sumona |  | South Asian | Lim et al. |
| Sumrah |  | South Asian | Lim et al. |
| Sumugat | Filipino | Other Asian | Wong et al. |
| Sumulong | Filipino | Other Asian | Wong et al. |
| Sun | Chinese | Chinese | Kranick et al. |
| Sunada | Japanese | Other Asian | Wong et al. |
| Sunagawa | Japanese | Other Asian | Wong et al. |
| Sunahara | Japanese | Other Asian | Wong et al. |
| Sunakawa | Japanese | Other Asian | Wong et al. |
| Sunami | Japanese | Other Asian | Wong et al. |
| Sunamoto | Japanese | Other Asian | Wong et al. |
| Sundaram | Asian Indian | South Asian | Wong et al. |
| Sundararaj | Asian Indian | South Asian | Wong et al. |
| Sundararajan | Asian Indian | South Asian | Wong et al. |
| Sundararaman | Asian Indian | South Asian | Wong et al. |
| Sundaresan | Asian Indian | South Asian | Wong et al. |
| Sunderji | Asian Indian | South Asian | Wong et al. |
| Sundiam | Filipino | Other Asian | Wong et al. |
| Sundita | Filipino | Other Asian | Wong et al. |
| Sune | Filipino | Other Asian | Wong et al. |
| Sung | Chinese; Korean | Chinese | Kranick et al. |
| Sunga | Filipino | Other Asian | Wong et al. |
| Sungahid | Filipino | Other Asian | Wong et al. |
| Sunglao | Filipino | Other Asian | Wong et al. |
| Sunico | Filipino | Other Asian | Wong et al. |
| Suniega | Filipino | Other Asian | Wong et al. |
| Suniga | Filipino | Other Asian | Wong et al. |
| Sunio | Filipino | Other Asian | Wong et al. |
| Sunkara | Asian Indian | South Asian | Wong et al. |
| Sunoo | Korean | Other Asian | Wong et al. |
| Sunpongco | Filipino | Other Asian | Kranick et al. |
| Suntay | Filipino | Other Asian | Wong et al. |
| Sunu | Korean | Other Asian | Wong et al. |
| Sunwoo | Korean | Other Asian | Wong et al. |
| Suo | Korean | Other Asian | Wong et al. |
| Suobiron | Filipino | Other Asian | Wong et al. |
| Suol | Vietnamese | Other Asian | Wong et al. |
| Suoribio | Filipino | Other Asian | Kranick et al. |
| Supangan | Filipino | Other Asian | Wong et al. |
| Supapo | Filipino | Other Asian | Wong et al. |
| Supena | Filipino | Other Asian | Wong et al. |
| Superio | Filipino | Other Asian | Wong et al. |
| Suplemento | Filipino | Other Asian | Wong et al. |
| Suplido | Filipino | Other Asian | Wong et al. |
| Supnet | Filipino | Other Asian | Wong et al. |
| Supremo | Filipino | Other Asian | Wong et al. |
| Supsup | Filipino | Other Asian | Wong et al. |
| Sur |  | South Asian | Lim et al. |
| Suralta | Filipino | Other Asian | Wong et al. |
| Surana | Asian Indian | South Asian | Wong et al. |
| Surani | Asian Indian | South Asian | Wong et al. |
| Surapaneni | Asian Indian | South Asian | Wong et al. |
| Surati | Asian Indian | South Asian | Wong et al. |
| Suratos | Filipino | Other Asian | Wong et al. |
| Surban | Filipino | Other Asian | Wong et al. |
| Surell | Filipino | Other Asian | Wong et al. |
| Suresh | Asian Indian | South Asian | Wong et al. |
| Surh | Korean | Other Asian | Wong et al. |
| Suri | Asian Indian; Pakistani; Indonesian | South Asian | Kranick et al. |
| Suriaga | Filipino | Other Asian | Wong et al. |
| Surinder |  | South Asian | Lim et al. |
| Surio | Filipino | Other Asian | Wong et al. |
| Surla | Filipino | Other Asian | Wong et al. |
| Surti | Asian Indian | South Asian | Wong et al. |
| Surtida | Filipino | Other Asian | Wong et al. |
| Surujlall |  | South Asian | Lim et al. |
| Surujpaulganesh |  | South Asian | Lim et al. |
| Suruki | Japanese | Other Asian | Wong et al. |
| Surve | Asian Indian | South Asian | Wong et al. |
| Suryanarayan | Asian Indian | South Asian | Wong et al. |
| Susada | Filipino | Other Asian | Wong et al. |
| Susai | Asian Indian | South Asian | Wong et al. |
| Susaki | Japanese | Other Asian | Wong et al. |
| Susara | Filipino | Other Asian | Wong et al. |
| Susbilla | Filipino | Other Asian | Wong et al. |
| Suson | Filipino | Other Asian | Wong et al. |
| Sustal | Filipino | Other Asian | Wong et al. |
| Sustento | Filipino | Other Asian | Wong et al. |
| Susuki | Japanese | Other Asian | Wong et al. |
| Susumu | Japanese | Other Asian | Wong et al. |
| Sut | Chinese | Chinese | Wong et al. |
| Sutaria | Asian Indian | South Asian | Wong et al. |
| Sutariya | Asian Indian | South Asian | Wong et al. |
| Sutarwala | Asian Indian | South Asian | Wong et al. |
| Suthar | Asian Indian | South Asian | Wong et al. |
| Suthayalai | Thai | Other Asian | Kranick et al. |
| Suto | Japanese | Other Asian | Kranick et al. |
| Sutradhar |  | South Asian | Lim et al. |
| Suu | Vietnamese | Other Asian | Wong et al. |
| Suvarna | Asian Indian | South Asian | Wong et al. |
| Suwa | Japanese | Other Asian | Wong et al. |
| Suwabe | Japanese | Other Asian | Wong et al. |
| Suyama | Japanese | Other Asian | Kranick et al. |
| Suyao | Filipino | Other Asian | Wong et al. |
| Suyat | Filipino | Other Asian | Wong et al. |
| Suyom | Filipino | Other Asian | Wong et al. |
| Suyu | Filipino | Other Asian | Wong et al. |
| Suzaki | Japanese | Other Asian | Wong et al. |
| Suzara | Filipino | Other Asian | Wong et al. |
| Suzawa | Japanese | Other Asian | Wong et al. |
| Suzue | Japanese | Other Asian | Wong et al. |
| Suzukawa | Japanese | Other Asian | Wong et al. |
| Suzuki | Japanese | Other Asian | Wong et al. |
| Suzumura | Japanese | Other Asian | Wong et al. |
| Swaby | Japanese | Other Asian | Wong et al. |
| Swami | Asian Indian | South Asian | Wong et al. |
| Swamidoss | Asian Indian | South Asian | Wong et al. |
| Swaminarayan | Asian Indian | South Asian | Wong et al. |
| Swaminathan | Asian Indian | South Asian | Wong et al. |
| Swamy | Asian Indian | South Asian | Wong et al. |
| Swaroop | Asian Indian | South Asian | Wong et al. |
| Swarup | Asian Indian | South Asian | Wong et al. |
| Swei | Chinese | Chinese | Wong et al. |
| Swen | Chinese | Chinese | Wong et al. |
| Sy | Filipino | Other Asian | Kranick et al. |
| Sycip | Filipino | Other Asian | Wong et al. |
| Sye | Chinese | Chinese | Wong et al. |
| Syed | Pakistani; Asian Indian | South Asian | Kranick et al. |
| Syfu | Filipino | Other Asian | Wong et al. |
| Sylla |  | South Asian | Lim et al. |
| Syn | Korean | Other Asian | Wong et al. |
| Synn | Korean | Other Asian | Wong et al. |
| Syquia | Filipino | Other Asian | Wong et al. |
| Syyap | Filipino | Other Asian | Wong et al. |
| Sze | Chinese | Chinese | Wong et al. |
| Szeto | Chinese | Chinese | Wong et al. |
| Szetu | Chinese | Chinese | Wong et al. |
| Szto | Chinese | Chinese | Wong et al. |
| Szu | Chinese | Chinese | Wong et al. |
| Szutu | Chinese | Chinese | Wong et al. |
| Ta | Vietnamese | Other Asian | Kranick et al. |
| Taa | Filipino | Other Asian | Wong et al. |
| Taaca | Filipino | Other Asian | Wong et al. |
| Taal | Filipino | Other Asian | Wong et al. |
| Taam | Chinese | Chinese | Wong et al. |
| Taasin | Filipino | Other Asian | Wong et al. |
| Taay | Filipino | Other Asian | Wong et al. |
| Taba | Japanese | Other Asian | Wong et al. |
| Tababa | Filipino | Other Asian | Wong et al. |
| Tabaco | Filipino | Other Asian | Wong et al. |
| Tabada | Filipino | Other Asian | Wong et al. |
| Tabadero | Filipino | Other Asian | Wong et al. |
| Tabafunda | Filipino | Other Asian | Wong et al. |
| Tabag | Filipino | Other Asian | Wong et al. |
| Tabago | Filipino | Other Asian | Wong et al. |
| Tabajonda | Filipino | Other Asian | Wong et al. |
| Tabalanza | Filipino | Other Asian | Wong et al. |
| Tabalbag | Filipino | Other Asian | Wong et al. |
| Tabaldo | Filipino | Other Asian | Wong et al. |
| Tabalno | Filipino | Other Asian | Wong et al. |
| Tabalon | Filipino | Other Asian | Wong et al. |
| Tabamo | Filipino | Other Asian | Wong et al. |
| Tabanao | Filipino | Other Asian | Wong et al. |
| Tabanera | Filipino | Other Asian | Wong et al. |
| Tabangay | Filipino | Other Asian | Wong et al. |
| Tabangcura | Filipino | Other Asian | Wong et al. |
| Tabaniag | Filipino | Other Asian | Wong et al. |
| Tabao | Filipino | Other Asian | Wong et al. |
| Tabaosares | Filipino | Other Asian | Wong et al. |
| Tabaquero | Filipino | Other Asian | Wong et al. |
| Tabar | Filipino | Other Asian | Wong et al. |
| Tabaranza | Filipino | Other Asian | Wong et al. |
| Tabares | Filipino | Other Asian | Wong et al. |
| Tabasa | Filipino | Other Asian | Wong et al. |
| Tabassum |  | South Asian | Lim et al. |
| Tabata | Japanese | Other Asian | Wong et al. |
| Tabay | Filipino | Other Asian | Wong et al. |
| Tabayoyong | Filipino | Other Asian | Wong et al. |
| Tabbada | Filipino | Other Asian | Wong et al. |
| Tabe | Japanese | Other Asian | Wong et al. |
| Tabei | Japanese | Other Asian | Wong et al. |
| Tabelin | Filipino | Other Asian | Wong et al. |
| Tabernero | Filipino | Other Asian | Wong et al. |
| Tabernilla | Filipino | Other Asian | Wong et al. |
| Tabeta | Filipino | Other Asian | Wong et al. |
| Tabieros | Filipino | Other Asian | Wong et al. |
| Tabigne | Filipino | Other Asian | Wong et al. |
| Tabije | Filipino | Other Asian | Kranick et al. |
| Tabil | Filipino | Other Asian | Wong et al. |
| Tabilas | Filipino | Other Asian | Wong et al. |
| Tabile | Filipino | Other Asian | Wong et al. |
| Tabili | Filipino | Other Asian | Wong et al. |
| Tabilin | Filipino | Other Asian | Wong et al. |
| Tabilisma | Filipino | Other Asian | Wong et al. |
| Tabin | Filipino | Other Asian | Wong et al. |
| Tabing | Filipino | Other Asian | Wong et al. |
| Tabinga | Filipino | Other Asian | Wong et al. |
| Tabingo | Filipino | Other Asian | Wong et al. |
| Tabiolo | Filipino | Other Asian | Wong et al. |
| Tabion | Filipino | Other Asian | Wong et al. |
| Tabios | Filipino | Other Asian | Wong et al. |
| Tabisola | Filipino | Other Asian | Wong et al. |
| Tabisula | Filipino | Other Asian | Wong et al. |
| Tabita | Filipino | Other Asian | Wong et al. |
| Tablada | Filipino | Other Asian | Wong et al. |
| Tabladillo | Filipino | Other Asian | Wong et al. |
| Tablan | Filipino | Other Asian | Wong et al. |
| Tablang | Filipino | Other Asian | Kranick et al. |
| Tablante | Filipino | Other Asian | Wong et al. |
| Tabligan | Filipino | Other Asian | Wong et al. |
| Tablit | Filipino | Other Asian | Wong et al. |
| Tablizo | Filipino | Other Asian | Wong et al. |
| Tabo | Filipino | Other Asian | Wong et al. |
| Taboada | Filipino | Other Asian | Wong et al. |
| Tabobo | Filipino | Other Asian | Wong et al. |
| Tabon | Filipino | Other Asian | Wong et al. |
| Tabones | Filipino | Other Asian | Wong et al. |
| Tabora | Filipino | Other Asian | Wong et al. |
| Tabotabo | Filipino | Other Asian | Wong et al. |
| Taboy | Filipino | Other Asian | Wong et al. |
| Tabtab | Filipino | Other Asian | Wong et al. |
| Tabu | Filipino | Other Asian | Wong et al. |
| Tabucbuc | Filipino | Other Asian | Wong et al. |
| Tabuchi | Japanese | Other Asian | Wong et al. |
| Tabucol | Filipino | Other Asian | Wong et al. |
| Tabudlo | Filipino | Other Asian | Wong et al. |
| Tabudlong | Filipino | Other Asian | Wong et al. |
| Tabuena | Filipino | Other Asian | Wong et al. |
| Tabujara | Filipino | Other Asian | Wong et al. |
| Tabula | Filipino | Other Asian | Wong et al. |
| Tabunan | Filipino | Other Asian | Wong et al. |
| Tabunar | Filipino | Other Asian | Wong et al. |
| Tabuno | Filipino | Other Asian | Wong et al. |
| Tabunot | Filipino | Other Asian | Wong et al. |
| Tabura | Filipino | Other Asian | Wong et al. |
| Tabuso | Filipino | Other Asian | Wong et al. |
| Tabutol | Filipino | Other Asian | Wong et al. |
| Tabuyo | Filipino | Other Asian | Wong et al. |
| Tabuzo | Filipino | Other Asian | Wong et al. |
| Tac | Vietnamese | Other Asian | Wong et al. |
| Tacad | Filipino | Other Asian | Wong et al. |
| Tacadena | Filipino | Other Asian | Wong et al. |
| Tacadina | Filipino | Other Asian | Wong et al. |
| Tacal | Filipino | Other Asian | Wong et al. |
| Tacan | Filipino | Other Asian | Wong et al. |
| Tacang | Filipino | Other Asian | Wong et al. |
| Tacardon | Filipino | Other Asian | Wong et al. |
| Tacata | Filipino | Other Asian | Wong et al. |
| Tacazon | Filipino | Other Asian | Wong et al. |
| Tacderan | Filipino | Other Asian | Wong et al. |
| Tacderas | Filipino | Other Asian | Wong et al. |
| Tacdol | Filipino | Other Asian | Wong et al. |
| Tachi | Japanese | Other Asian | Wong et al. |
| Tachibana | Japanese | Other Asian | Wong et al. |
| Tachikawa | Japanese | Other Asian | Wong et al. |
| Tachiki | Japanese | Other Asian | Wong et al. |
| Tacla | Filipino | Other Asian | Wong et al. |
| Taclan | Filipino | Other Asian | Wong et al. |
| Taclas | Filipino | Other Asian | Wong et al. |
| Taclay | Filipino | Other Asian | Wong et al. |
| Tacmo | Filipino | Other Asian | Wong et al. |
| Tacorda | Filipino | Other Asian | Wong et al. |
| Tacotaco | Filipino | Other Asian | Wong et al. |
| Tactacan | Filipino | Other Asian | Wong et al. |
| Tactaquin | Filipino | Other Asian | Wong et al. |
| Tactay | Filipino | Other Asian | Wong et al. |
| Tacub | Filipino | Other Asian | Wong et al. |
| Tacuboy | Filipino | Other Asian | Wong et al. |
| Taculog | Filipino | Other Asian | Wong et al. |
| Tada | Japanese | Other Asian | Wong et al. |
| Tadaki | Japanese | Other Asian | Wong et al. |
| Tadano | Japanese | Other Asian | Wong et al. |
| Tadashi | Japanese | Other Asian | Wong et al. |
| Taday | Filipino | Other Asian | Wong et al. |
| Tadeja | Filipino | Other Asian | Wong et al. |
| Tadena | Filipino | Other Asian | Wong et al. |
| Tadeo | Filipino | Other Asian | Wong et al. |
| Tadiar | Filipino | Other Asian | Wong et al. |
| Tadifa | Filipino | Other Asian | Wong et al. |
| Tadina | Filipino | Other Asian | Wong et al. |
| Tadio | Filipino | Other Asian | Wong et al. |
| Tadokoro | Japanese | Other Asian | Wong et al. |
| Taduran | Filipino | Other Asian | Wong et al. |
| Tae | Korean | Other Asian | Wong et al. |
| Taek | Korean | Other Asian | Wong et al. |
| Taeza | Filipino | Other Asian | Wong et al. |
| Tafalla | Filipino | Other Asian | Wong et al. |
| Taga | Japanese | Other Asian | Wong et al. |
| Tagaban | Filipino | Other Asian | Wong et al. |
| Tagaca | Filipino | Other Asian | Wong et al. |
| Tagal | Filipino | Other Asian | Wong et al. |
| Tagala | Filipino | Other Asian | Wong et al. |
| Tagalan | Filipino | Other Asian | Wong et al. |
| Tagalicod | Filipino | Other Asian | Wong et al. |
| Tagalicud | Filipino | Other Asian | Wong et al. |
| Tagalog | Filipino | Other Asian | Wong et al. |
| Tagama | Filipino | Other Asian | Wong et al. |
| Tagami | Japanese | Other Asian | Wong et al. |
| Tagamolila | Filipino | Other Asian | Wong et al. |
| Taganap | Filipino | Other Asian | Wong et al. |
| Taganas | Filipino | Other Asian | Wong et al. |
| Tagarao | Filipino | Other Asian | Wong et al. |
| Tagare | Filipino | Other Asian | Wong et al. |
| Tagari | Filipino | Other Asian | Wong et al. |
| Tagarino | Filipino | Other Asian | Wong et al. |
| Tagashira | Japanese | Other Asian | Wong et al. |
| Tagata | Filipino | Other Asian | Wong et al. |
| Tagatac | Filipino | Other Asian | Wong et al. |
| Tagavilla | Filipino | Other Asian | Wong et al. |
| Tagawa | Japanese | Other Asian | Wong et al. |
| Tagaya | Japanese | Other Asian | Wong et al. |
| Tagayuna | Filipino | Other Asian | Wong et al. |
| Taghap | Filipino | Other Asian | Wong et al. |
| Tagle | Filipino | Other Asian | Kranick et al. |
| Tagnipez | Filipino | Other Asian | Wong et al. |
| Tago | Japanese | Other Asian | Wong et al. |
| Tagorda | Filipino | Other Asian | Wong et al. |
| Tagore | Asian Indian | South Asian | Wong et al. |
| Taguba | Filipino | Other Asian | Wong et al. |
| Taguchi | Japanese | Other Asian | Wong et al. |
| Tagudar | Filipino | Other Asian | Wong et al. |
| Tagudin | Filipino | Other Asian | Wong et al. |
| Taguiam | Filipino | Other Asian | Wong et al. |
| Taguibao | Filipino | Other Asian | Wong et al. |
| Taguinod | Filipino | Other Asian | Wong et al. |
| Tagulao | Filipino | Other Asian | Wong et al. |
| Tagulinao | Filipino | Other Asian | Wong et al. |
| Tagum | Filipino | Other Asian | Wong et al. |
| Tagupa | Filipino | Other Asian | Wong et al. |
| Tagura | Filipino | Other Asian | Wong et al. |
| Tahara | Japanese | Other Asian | Wong et al. |
| Taher | Bangladeshi | South Asian | Kranick et al. |
| Tahilan | Filipino | Other Asian | Wong et al. |
| Tahimic | Filipino | Other Asian | Wong et al. |
| Tahira | Japanese | Other Asian | Wong et al. |
| Tahk | Korean | Other Asian | Wong et al. |
| Tahmith |  | South Asian | Lim et al. |
| Tai | Chinese | Chinese | Wong et al. |
| Taik | Korean | Other Asian | Wong et al. |
| Tailor | Asian Indian | South Asian | Wong et al. |
| Taima | Japanese | Other Asian | Wong et al. |
| Tain | Chinese | Chinese | Wong et al. |
| Taino | Filipino | Other Asian | Wong et al. |
| Taira | Japanese | Other Asian | Wong et al. |
| Taja | Filipino | Other Asian | Wong et al. |
| Tajani | Asian Indian | South Asian | Wong et al. |
| Tajanlangit | Filipino | Other Asian | Wong et al. |
| Tajima | Japanese | Other Asian | Wong et al. |
| Tajiri | Japanese | Other Asian | Wong et al. |
| Tajon | Filipino | Other Asian | Wong et al. |
| Tajonera | Filipino | Other Asian | Wong et al. |
| Tak | Asian Indian; Japanese; Korean | South Asian | Kranick et al. |
| Taka | Japanese | Other Asian | Wong et al. |
| Takabatake | Japanese | Other Asian | Wong et al. |
| Takabayashi | Japanese | Other Asian | Wong et al. |
| Takabe | Japanese | Other Asian | Wong et al. |
| Takada | Japanese | Other Asian | Wong et al. |
| Takae | Japanese | Other Asian | Wong et al. |
| Takaesu | Japanese | Other Asian | Wong et al. |
| Takagaki | Japanese | Other Asian | Wong et al. |
| Takagawa | Japanese | Other Asian | Wong et al. |
| Takagi | Japanese | Other Asian | Wong et al. |
| Takagishi | Japanese | Other Asian | Wong et al. |
| Takaguchi | Japanese | Other Asian | Wong et al. |
| Takahagi | Japanese | Other Asian | Wong et al. |
| Takahama | Japanese | Other Asian | Wong et al. |
| Takahara | Japanese | Other Asian | Wong et al. |
| Takahash | Japanese | Other Asian | Wong et al. |
| Takahashi | Japanese | Other Asian | Kranick et al. |
| Takahasi | Japanese | Other Asian | Wong et al. |
| Takahata | Japanese | Other Asian | Wong et al. |
| Takahira | Japanese | Other Asian | Wong et al. |
| Takahshi | Japanese | Other Asian | Wong et al. |
| Takai | Japanese | Other Asian | Wong et al. |
| Takaishi | Japanese | Other Asian | Wong et al. |
| Takaiwa | Japanese | Other Asian | Wong et al. |
| Takaki | Japanese | Other Asian | Wong et al. |
| Takaku | Japanese | Other Asian | Wong et al. |
| Takakura | Japanese | Other Asian | Wong et al. |
| Takakuwa | Japanese | Other Asian | Wong et al. |
| Takamatsu | Japanese | Other Asian | Wong et al. |
| Takami | Japanese | Other Asian | Wong et al. |
| Takamine | Japanese | Other Asian | Wong et al. |
| Takamiya | Japanese | Other Asian | Wong et al. |
| Takamizawa | Japanese | Other Asian | Wong et al. |
| Takamori | Japanese | Other Asian | Wong et al. |
| Takamoto | Japanese | Other Asian | Wong et al. |
| Takamura | Japanese | Other Asian | Wong et al. |
| Takanabe | Japanese | Other Asian | Wong et al. |
| Takanashi | Japanese | Other Asian | Wong et al. |
| Takano | Japanese | Other Asian | Wong et al. |
| Takao | Japanese | Other Asian | Wong et al. |
| Takaoka | Japanese | Other Asian | Wong et al. |
| Takara | Japanese | Other Asian | Wong et al. |
| Takarabe | Japanese | Other Asian | Wong et al. |
| Takarada | Japanese | Other Asian | Wong et al. |
| Takasaka | Japanese | Other Asian | Wong et al. |
| Takasaki | Japanese | Other Asian | Wong et al. |
| Takasawa | Japanese | Other Asian | Wong et al. |
| Takase | Japanese | Other Asian | Wong et al. |
| Takashi | Japanese | Other Asian | Wong et al. |
| Takashima | Japanese | Other Asian | Wong et al. |
| Takashina | Japanese | Other Asian | Wong et al. |
| Takasu | Japanese | Other Asian | Wong et al. |
| Takasugi | Japanese | Other Asian | Wong et al. |
| Takata | Japanese | Other Asian | Wong et al. |
| Takato | Japanese | Other Asian | Wong et al. |
| Takatori | Japanese | Other Asian | Wong et al. |
| Takatsu | Japanese | Other Asian | Wong et al. |
| Takatsuka | Japanese | Other Asian | Wong et al. |
| Takatsuki | Japanese | Other Asian | Wong et al. |
| Takatu | Japanese | Other Asian | Kranick et al. |
| Takaura | Japanese | Other Asian | Wong et al. |
| Takaya | Japanese | Other Asian | Wong et al. |
| Takayama | Japanese | Other Asian | Wong et al. |
| Takayanagi | Japanese | Other Asian | Wong et al. |
| Takayasu | Japanese | Other Asian | Wong et al. |
| Takayoshi | Japanese | Other Asian | Wong et al. |
| Takazawa | Japanese | Other Asian | Wong et al. |
| Takebayashi | Japanese | Other Asian | Wong et al. |
| Takebe | Japanese | Other Asian | Wong et al. |
| Takechi | Japanese | Other Asian | Wong et al. |
| Takeda | Japanese | Other Asian | Wong et al. |
| Takegami | Japanese | Other Asian | Wong et al. |
| Takegawa | Japanese | Other Asian | Wong et al. |
| Takeguchi | Japanese | Other Asian | Wong et al. |
| Takehara | Japanese | Other Asian | Wong et al. |
| Takehisa | Japanese | Other Asian | Wong et al. |
| Takei | Japanese | Other Asian | Wong et al. |
| Takeichi | Japanese | Other Asian | Wong et al. |
| Takeishi | Japanese | Other Asian | Wong et al. |
| Takekawa | Japanese | Other Asian | Wong et al. |
| Takekoshi | Japanese | Other Asian | Wong et al. |
| Takel | Japanese | Other Asian | Wong et al. |
| Takemasa | Japanese | Other Asian | Wong et al. |
| Takematsu | Japanese | Other Asian | Wong et al. |
| Takemori | Japanese | Other Asian | Wong et al. |
| Takemoto | Japanese | Other Asian | Wong et al. |
| Takemura | Japanese | Other Asian | Wong et al. |
| Takenaga | Japanese | Other Asian | Wong et al. |
| Takenaka | Japanese | Other Asian | Wong et al. |
| Takeno | Japanese | Other Asian | Wong et al. |
| Takenouchi | Japanese | Other Asian | Wong et al. |
| Takeo | Japanese | Other Asian | Kranick et al. |
| Takeoka | Japanese | Other Asian | Wong et al. |
| Takesako | Japanese | Other Asian | Wong et al. |
| Takeshi | Japanese | Other Asian | Wong et al. |
| Takeshima | Japanese | Other Asian | Wong et al. |
| Takeshita | Japanese | Other Asian | Wong et al. |
| Takesue | Japanese | Other Asian | Wong et al. |
| Taketa | Japanese | Other Asian | Wong et al. |
| Taketani | Japanese | Other Asian | Wong et al. |
| Taketomi | Japanese | Other Asian | Wong et al. |
| Takeuchi | Japanese | Other Asian | Kranick et al. |
| Takeya | Japanese | Other Asian | Wong et al. |
| Takeyama | Japanese | Other Asian | Wong et al. |
| Takezawa | Japanese | Other Asian | Wong et al. |
| Takhar | Asian Indian | South Asian | Wong et al. |
| Taki | Japanese | Other Asian | Wong et al. |
| Takida | Japanese | Other Asian | Wong et al. |
| Takido | Japanese | Other Asian | Wong et al. |
| Takigawa | Japanese | Other Asian | Wong et al. |
| Takiguchi | Japanese | Other Asian | Wong et al. |
| Takii | Japanese | Other Asian | Wong et al. |
| Takikawa | Japanese | Other Asian | Wong et al. |
| Takimoto | Japanese | Other Asian | Wong et al. |
| Takino | Japanese | Other Asian | Wong et al. |
| Takishima | Japanese | Other Asian | Wong et al. |
| Takita | Japanese | Other Asian | Wong et al. |
| Takiuchi | Japanese | Other Asian | Wong et al. |
| Takiyama | Japanese | Other Asian | Wong et al. |
| Takizawa | Japanese | Other Asian | Wong et al. |
| Takubo | Japanese | Other Asian | Wong et al. |
| Takuma | Japanese | Other Asian | Wong et al. |
| Takumi | Japanese | Other Asian | Wong et al. |
| Takushi | Japanese | Other Asian | Wong et al. |
| Tala | Filipino | Other Asian | Wong et al. |
| Talabis | Filipino | Other Asian | Wong et al. |
| Talabucon | Filipino | Other Asian | Wong et al. |
| Talactac | Filipino | Other Asian | Wong et al. |
| Taladua | Filipino | Other Asian | Wong et al. |
| Talag | Filipino | Other Asian | Wong et al. |
| Talagtag | Filipino | Other Asian | Wong et al. |
| Talamayan | Filipino | Other Asian | Wong et al. |
| Talampas | Filipino | Other Asian | Wong et al. |
| Talana | Filipino | Other Asian | Wong et al. |
| Talanay | Filipino | Other Asian | Wong et al. |
| Talania | Filipino | Other Asian | Wong et al. |
| Talao | Filipino | Other Asian | Wong et al. |
| Talaro | Filipino | Other Asian | Wong et al. |
| Talastas | Filipino | Other Asian | Wong et al. |
| Talatala | Filipino | Other Asian | Wong et al. |
| Talati | Asian Indian | South Asian | Wong et al. |
| Talato | Filipino | Other Asian | Wong et al. |
| Talavera | Filipino | Other Asian | Wong et al. |
| Talay | Filipino | Other Asian | Wong et al. |
| Talbo | Filipino | Other Asian | Wong et al. |
| Taleb |  | South Asian | Lim et al. |
| Talens | Filipino | Other Asian | Wong et al. |
| Talento | Filipino | Other Asian | Wong et al. |
| Taleon | Filipino | Other Asian | Wong et al. |
| Taliman | Filipino | Other Asian | Wong et al. |
| Talla | Filipino | Other Asian | Wong et al. |
| Talledo | Filipino | Other Asian | Wong et al. |
| Tallungan | Filipino | Other Asian | Wong et al. |
| Talob | Filipino | Other Asian | Wong et al. |
| Taloma | Filipino | Other Asian | Wong et al. |
| Talon | Filipino | Other Asian | Wong et al. |
| Talorda | Filipino | Other Asian | Wong et al. |
| Talosig | Filipino | Other Asian | Wong et al. |
| Taloza | Filipino | Other Asian | Wong et al. |
| Talpade | Asian Indian | South Asian | Wong et al. |
| Talplacido | Filipino | Other Asian | Wong et al. |
| Talsania | Asian Indian | South Asian | Wong et al. |
| Taluban | Filipino | Other Asian | Wong et al. |
| Talucad | Filipino | Other Asian | Wong et al. |
| Talukdar | Asian Indian; Bangladeshi | South Asian | Kranick et al. |
| Talukder | Bangladeshi; Asian Indian | South Asian | Kranick et al. |
| Talusan | Filipino | Other Asian | Wong et al. |
| Talvo | Filipino | Other Asian | Wong et al. |
| Talwar | Asian Indian | South Asian | Kranick et al. |
| Tam | Chinese; Vietnamese | Chinese | Kranick et al. |
| Tamada | Japanese | Other Asian | Wong et al. |
| Tamagawa | Japanese | Other Asian | Wong et al. |
| Tamai | Japanese | Other Asian | Wong et al. |
| Tamaki | Japanese | Other Asian | Wong et al. |
| Tamamura | Japanese | Other Asian | Wong et al. |
| Tamanaha | Japanese | Other Asian | Wong et al. |
| Tamang |  | South Asian | Lim et al. |
| Tamano | Filipino | Other Asian | Wong et al. |
| Tamaoki | Japanese | Other Asian | Wong et al. |
| Tamaray | Filipino | Other Asian | Wong et al. |
| Tamares | Filipino | Other Asian | Wong et al. |
| Tamargo | Filipino | Other Asian | Wong et al. |
| Tamari | Japanese | Other Asian | Wong et al. |
| Tamarra | Filipino | Other Asian | Wong et al. |
| Tamaru | Japanese | Other Asian | Wong et al. |
| Tamase | Filipino | Other Asian | Wong et al. |
| Tamashiro | Japanese | Other Asian | Wong et al. |
| Tamate | Japanese | Other Asian | Wong et al. |
| Tamayao | Filipino | Other Asian | Wong et al. |
| Tamayo | Filipino | Other Asian | Wong et al. |
| Tamayose | Japanese | Other Asian | Wong et al. |
| Tamba | Filipino | Other Asian | Wong et al. |
| Tambalque | Filipino | Other Asian | Wong et al. |
| Tambanillo | Filipino | Other Asian | Wong et al. |
| Tambaoan | Filipino | Other Asian | Wong et al. |
| Tambio | Filipino | Other Asian | Wong et al. |
| Tambo | Filipino | Other Asian | Wong et al. |
| Tamboli | Asian Indian | South Asian | Wong et al. |
| Tambongco | Filipino | Other Asian | Wong et al. |
| Tamboong | Filipino | Other Asian | Wong et al. |
| Tambot | Filipino | Other Asian | Wong et al. |
| Tambunting | Filipino | Other Asian | Wong et al. |
| Tameta | Filipino | Other Asian | Wong et al. |
| Tamhane | Asian Indian | South Asian | Wong et al. |
| Tamiya | Japanese | Other Asian | Wong et al. |
| Tamonan | Filipino | Other Asian | Wong et al. |
| Tamondong | Filipino | Other Asian | Kranick et al. |
| Tamonte | Filipino | Other Asian | Wong et al. |
| Tamoria | Filipino | Other Asian | Wong et al. |
| Tamoro | Filipino | Other Asian | Wong et al. |
| Tamotsu | Japanese | Other Asian | Wong et al. |
| Tamparong | Filipino | Other Asian | Wong et al. |
| Tampoc | Filipino | Other Asian | Wong et al. |
| Tampol | Filipino | Other Asian | Wong et al. |
| Tampon | Filipino | Other Asian | Wong et al. |
| Tampos | Filipino | Other Asian | Wong et al. |
| Tampoya | Filipino | Other Asian | Wong et al. |
| Tampus | Filipino | Other Asian | Wong et al. |
| Tamula | Filipino | Other Asian | Wong et al. |
| Tamura | Japanese | Other Asian | Wong et al. |
| Tan | Chinese; Malaysian; Vietnamese; Filipino | Chinese | Kranick et al. |
| Tana | Filipino | Other Asian | Wong et al. |
| Tanabe | Japanese | Other Asian | Wong et al. |
| Tanada | Filipino | Other Asian | Wong et al. |
| Tanael | Filipino | Other Asian | Wong et al. |
| Tanafranca | Filipino | Other Asian | Wong et al. |
| Tanagon | Filipino | Other Asian | Wong et al. |
| Tanahara | Japanese | Other Asian | Wong et al. |
| Tanahashi | Japanese | Other Asian | Wong et al. |
| Tanaka | Japanese | Other Asian | Wong et al. |
| Tanala | Filipino | Other Asian | Wong et al. |
| Tanalega | Filipino | Other Asian | Wong et al. |
| Tanalgo | Filipino | Other Asian | Wong et al. |
| Tanap | Filipino | Other Asian | Wong et al. |
| Tanawan | Filipino | Other Asian | Wong et al. |
| Tanbara | Japanese | Other Asian | Wong et al. |
| Tancinco | Filipino | Other Asian | Wong et al. |
| Tancioco | Filipino | Other Asian | Wong et al. |
| Tandaguen | Filipino | Other Asian | Wong et al. |
| Tandan | Asian Indian | South Asian | Wong et al. |
| Tandel | Asian Indian | South Asian | Wong et al. |
| Tandingan | Filipino | Other Asian | Wong et al. |
| Tandoc | Filipino | Other Asian | Wong et al. |
| Tandon | Asian Indian | South Asian | Wong et al. |
| Taneda | Japanese | Other Asian | Wong et al. |
| Tanedo | Filipino | Other Asian | Wong et al. |
| Tanega | Filipino | Other Asian | Wong et al. |
| Tanemura | Japanese | Other Asian | Wong et al. |
| Taneo | Filipino | Other Asian | Wong et al. |
| Tang | Chinese; Vietnamese | Chinese | Kranick et al. |
| Tangalin | Filipino | Other Asian | Wong et al. |
| Tangatue | Filipino | Other Asian | Wong et al. |
| Tangca | Filipino | Other Asian | Wong et al. |
| Tangchakkrachai | Thai | Other Asian | Kranick et al. |
| Tangco | Filipino | Other Asian | Wong et al. |
| Tange | Japanese | Other Asian | Wong et al. |
| Tanghal | Filipino | Other Asian | Wong et al. |
| Tanglao | Filipino | Other Asian | Wong et al. |
| Tango | Filipino | Other Asian | Wong et al. |
| Tangonan | Filipino | Other Asian | Wong et al. |
| Tangri | Asian Indian | South Asian | Wong et al. |
| Tanguilig | Filipino | Other Asian | Wong et al. |
| Tangunan | Filipino | Other Asian | Wong et al. |
| Tanh | Vietnamese | Other Asian | Kranick et al. |
| Tanhueco | Filipino | Other Asian | Wong et al. |
| Tani | Japanese | Other Asian | Wong et al. |
| Tanicala | Filipino | Other Asian | Wong et al. |
| Tanida | Japanese | Other Asian | Wong et al. |
| Tanifuji | Japanese | Other Asian | Wong et al. |
| Tanigaki | Japanese | Other Asian | Wong et al. |
| Tanigawa | Japanese | Other Asian | Wong et al. |
| Taniguchi | Japanese | Other Asian | Wong et al. |
| Tanikawa | Japanese | Other Asian | Wong et al. |
| Tanimoto | Japanese | Other Asian | Wong et al. |
| Tanimura | Japanese | Other Asian | Wong et al. |
| Taninaka | Japanese | Other Asian | Wong et al. |
| Taningco | Filipino | Other Asian | Wong et al. |
| Tanino | Japanese | Other Asian | Wong et al. |
| Tanioka | Japanese | Other Asian | Wong et al. |
| Taniwaki | Japanese | Other Asian | Wong et al. |
| Taniyama | Japanese | Other Asian | Wong et al. |
| Tanizaki | Japanese | Other Asian | Wong et al. |
| Tanizawa | Japanese | Other Asian | Wong et al. |
| Tanji | Japanese | Other Asian | Wong et al. |
| Tanjuakio | Filipino | Other Asian | Wong et al. |
| Tanjuaquio | Filipino | Other Asian | Wong et al. |
| Tanjutco | Filipino | Other Asian | Wong et al. |
| Tanna | Asian Indian | South Asian | Wong et al. |
| Tanno | Japanese | Other Asian | Wong et al. |
| Tano | Filipino | Other Asian | Wong et al. |
| Tanoue | Japanese | Other Asian | Wong et al. |
| Tanouye | Japanese | Other Asian | Wong et al. |
| Tanseco | Filipino | Other Asian | Wong et al. |
| Tansioco | Filipino | Other Asian | Wong et al. |
| Tantay | Filipino | Other Asian | Wong et al. |
| Tantingco | Filipino | Other Asian | Wong et al. |
| Tantoco | Filipino | Other Asian | Wong et al. |
| Tantoy | Filipino | Other Asian | Wong et al. |
| Tantuico | Filipino | Other Asian | Wong et al. |
| Tanudra | Filipino | Other Asian | Wong et al. |
| Tanudtanud | Filipino | Other Asian | Wong et al. |
| Tanveer | Pakistani | South Asian | Kranick et al. |
| Tanwangco | Filipino | Other Asian | Wong et al. |
| Tanyag | Filipino | Other Asian | Wong et al. |
| Tanzil | Bangladeshi | South Asian | Kranick et al. |
| Tanzo | Filipino | Other Asian | Wong et al. |
| Tao | Chinese | Chinese | Wong et al. |
| Taoatao | Filipino | Other Asian | Wong et al. |
| Taoka | Japanese | Other Asian | Wong et al. |
| Taong | Filipino | Other Asian | Wong et al. |
| Tapado | Filipino | Other Asian | Wong et al. |
| Tapales | Filipino | Other Asian | Wong et al. |
| Tapalla | Filipino | Other Asian | Wong et al. |
| Tapang | Filipino | Other Asian | Wong et al. |
| Tapaoan | Filipino | Other Asian | Wong et al. |
| Tapat | Filipino | Other Asian | Wong et al. |
| Tapawan | Filipino | Other Asian | Wong et al. |
| Tapay | Filipino | Other Asian | Wong et al. |
| Tapayan | Filipino | Other Asian | Wong et al. |
| Tapec | Filipino | Other Asian | Wong et al. |
| Tapia | Filipino | Other Asian | Wong et al. |
| Tapiador | Filipino | Other Asian | Wong et al. |
| Tapnio | Filipino | Other Asian | Wong et al. |
| Tapuro | Filipino | Other Asian | Wong et al. |
| Taqi | Asian Indian | South Asian | Wong et al. |
| Taracatac | Filipino | Other Asian | Wong et al. |
| Tarachand | Asian Indian | South Asian | Wong et al. |
| Tarampi | Filipino | Other Asian | Wong et al. |
| Tarantan | Filipino | Other Asian | Wong et al. |
| Tarape | Filipino | Other Asian | Wong et al. |
| Taraya | Filipino | Other Asian | Wong et al. |
| Tare | Asian Indian | South Asian | Wong et al. |
| Tarectecan | Filipino | Other Asian | Wong et al. |
| Targa | Filipino | Other Asian | Wong et al. |
| Tariga | Filipino | Other Asian | Wong et al. |
| Tariq | Pakistani | South Asian | Kranick et al. |
| Tark | Korean | Other Asian | Wong et al. |
| Tarlit | Filipino | Other Asian | Wong et al. |
| Tarn | Chinese | Chinese | Wong et al. |
| Tarnate | Filipino | Other Asian | Wong et al. |
| Tarng | Chinese | Chinese | Wong et al. |
| Taroma | Filipino | Other Asian | Wong et al. |
| Tarongoy | Filipino | Other Asian | Wong et al. |
| Taroy | Filipino | Other Asian | Wong et al. |
| Tarrayo | Filipino | Other Asian | Wong et al. |
| Tarrosa | Filipino | Other Asian | Wong et al. |
| Tarroza | Filipino | Other Asian | Wong et al. |
| Taruc | Filipino | Other Asian | Wong et al. |
| Tarui | Japanese | Other Asian | Wong et al. |
| Tarumoto | Japanese | Other Asian | Wong et al. |
| Tarun | Filipino | Other Asian | Wong et al. |
| Tasaka | Japanese | Other Asian | Wong et al. |
| Tasaki | Japanese | Other Asian | Wong et al. |
| Tashima | Japanese | Other Asian | Wong et al. |
| Tashiro | Japanese | Other Asian | Wong et al. |
| Taslim |  | South Asian | Lim et al. |
| Tatad | Filipino | Other Asian | Wong et al. |
| Tatara | Japanese | Other Asian | Wong et al. |
| Tatari |  | South Asian | Lim et al. |
| Tatebe | Japanese | Other Asian | Wong et al. |
| Tateishi | Japanese | Other Asian | Wong et al. |
| Tateiwa | Japanese | Other Asian | Wong et al. |
| Tatematsu | Japanese | Other Asian | Wong et al. |
| Tateno | Japanese | Other Asian | Wong et al. |
| Tatewaki | Japanese | Other Asian | Wong et al. |
| Tateyama | Japanese | Other Asian | Wong et al. |
| Tatheer |  | South Asian | Lim et al. |
| Tating | Filipino | Other Asian | Wong et al. |
| Tatlonghari | Filipino | Other Asian | Wong et al. |
| Taton | Filipino | Other Asian | Wong et al. |
| Tatoy | Filipino | Other Asian | Wong et al. |
| Tatsukawa | Japanese | Other Asian | Wong et al. |
| Tatsumi | Japanese | Other Asian | Wong et al. |
| Tatsumoto | Japanese | Other Asian | Wong et al. |
| Tatsuno | Japanese | Other Asian | Wong et al. |
| Tatsuta | Japanese | Other Asian | Wong et al. |
| Tau | Chinese | Chinese | Wong et al. |
| Tauchi | Japanese | Other Asian | Wong et al. |
| Taur | Chinese | Chinese | Wong et al. |
| Taura | Japanese | Other Asian | Wong et al. |
| Tauro | Filipino | Other Asian | Wong et al. |
| Tauyan | Filipino | Other Asian | Wong et al. |
| Tavanlar | Filipino | Other Asian | Wong et al. |
| Tavares | Chinese | Chinese | Wong et al. |
| Tavera | Filipino | Other Asian | Wong et al. |
| Tavora | Filipino | Other Asian | Wong et al. |
| Taw | Chinese | Chinese | Wong et al. |
| Tawa | Japanese | Other Asian | Wong et al. |
| Tawara | Japanese | Other Asian | Wong et al. |
| Tawatao | Filipino | Other Asian | Wong et al. |
| Taya | Japanese | Other Asian | Wong et al. |
| Tayaba | Filipino | Other Asian | Wong et al. |
| Tayag | Filipino | Other Asian | Wong et al. |
| Tayal | Asian Indian | South Asian | Wong et al. |
| Tayama | Japanese | Other Asian | Wong et al. |
| Tayamen | Filipino | Other Asian | Kranick et al. |
| Tayao | Filipino | Other Asian | Wong et al. |
| Tayco | Filipino | Other Asian | Wong et al. |
| Tayengco | Filipino | Other Asian | Wong et al. |
| Tayko | Filipino | Other Asian | Wong et al. |
| Taylan | Filipino | Other Asian | Wong et al. |
| Tayo | Filipino | Other Asian | Wong et al. |
| Tayong | Filipino | Other Asian | Wong et al. |
| Tayza | Burmese | Other Asian | Kranick et al. |
| Tazaki | Japanese | Other Asian | Wong et al. |
| Tazawa | Japanese | Other Asian | Wong et al. |
| Tchan | Vietnamese | Other Asian | Wong et al. |
| Tchang | Vietnamese | Other Asian | Wong et al. |
| Tchao | Chinese | Chinese | Wong et al. |
| Tche | Vietnamese | Other Asian | Wong et al. |
| Tcheng | Vietnamese | Other Asian | Wong et al. |
| Tchong | Vietnamese | Other Asian | Wong et al. |
| Tchou | Chinese | Chinese | Wong et al. |
| Te | Chinese; Filipino; Cambodian; Burmese | Chinese | Kranick et al. |
| Teang | Chinese | Chinese | Wong et al. |
| Teano | Filipino | Other Asian | Wong et al. |
| Tecson | Filipino | Other Asian | Kranick et al. |
| Ted | Chinese | Chinese | Wong et al. |
| Tee | Chinese | Chinese | Wong et al. |
| Tegio | Filipino | Other Asian | Wong et al. |
| Teh | Chinese | Chinese | Wong et al. |
| Tei | Chinese | Chinese | Wong et al. |
| Tejada | Filipino | Other Asian | Wong et al. |
| Tejam | Filipino | Other Asian | Wong et al. |
| Tejani | Asian Indian | South Asian | Wong et al. |
| Tejano | Filipino | Other Asian | Wong et al. |
| Tejero | Filipino | Other Asian | Wong et al. |
| Tejima | Japanese | Other Asian | Wong et al. |
| Tejpal | Asian Indian | South Asian | Wong et al. |
| Tejuco | Filipino | Other Asian | Wong et al. |
| Tejuja | Asian Indian | South Asian | Wong et al. |
| Tela | Filipino | Other Asian | Wong et al. |
| Telan | Filipino | Other Asian | Wong et al. |
| Telang | Asian Indian | South Asian | Wong et al. |
| Telebrico | Filipino | Other Asian | Wong et al. |
| Tello | Filipino | Other Asian | Wong et al. |
| Telmo | Filipino | Other Asian | Wong et al. |
| Tembe | Asian Indian | South Asian | Wong et al. |
| Templado | Filipino | Other Asian | Wong et al. |
| Templo | Filipino | Other Asian | Wong et al. |
| Templonuevo | Filipino | Other Asian | Wong et al. |
| Tempongko | Filipino | Other Asian | Wong et al. |
| Temporal | Filipino | Other Asian | Wong et al. |
| Ten | Chinese | Chinese | Wong et al. |
| Tena | Filipino | Other Asian | Wong et al. |
| Tenazas | Filipino | Other Asian | Wong et al. |
| Tence | Filipino | Other Asian | Wong et al. |
| Tenchavez | Filipino | Other Asian | Wong et al. |
| Tendencia | Filipino | Other Asian | Wong et al. |
| Tendero | Filipino | Other Asian | Wong et al. |
| Tenebro | Filipino | Other Asian | Wong et al. |
| Tenedero | Filipino | Other Asian | Wong et al. |
| Tenefrancia | Filipino | Other Asian | Wong et al. |
| Teneza | Filipino | Other Asian | Wong et al. |
| Teng | Chinese | Chinese | Wong et al. |
| Tengan | Japanese | Other Asian | Wong et al. |
| Tengasantos | Filipino | Other Asian | Wong et al. |
| Tengco | Filipino | Other Asian | Wong et al. |
| Tenno | Japanese | Other Asian | Wong et al. |
| Tenorio | Filipino | Other Asian | Wong et al. |
| Tenoso | Filipino | Other Asian | Wong et al. |
| Tentado | Filipino | Other Asian | Wong et al. |
| Tenzin |  | South Asian | Lim et al. |
| Teodoro | Filipino | Other Asian | Kranick et al. |
| Teodosio | Filipino | Other Asian | Wong et al. |
| Teofilo | Filipino | Other Asian | Wong et al. |
| Teologo | Filipino | Other Asian | Wong et al. |
| Teong | Chinese | Chinese | Wong et al. |
| Teopaco | Filipino | Other Asian | Wong et al. |
| Teope | Filipino | Other Asian | Wong et al. |
| Teotico | Filipino | Other Asian | Wong et al. |
| Teppang | Filipino | Other Asian | Wong et al. |
| Terada | Japanese | Other Asian | Wong et al. |
| Terai | Japanese | Other Asian | Wong et al. |
| Teraji | Japanese | Other Asian | Wong et al. |
| Terakado | Japanese | Other Asian | Wong et al. |
| Terakawa | Japanese | Other Asian | Wong et al. |
| Teramoto | Japanese | Other Asian | Wong et al. |
| Teramura | Japanese | Other Asian | Wong et al. |
| Teranishi | Japanese | Other Asian | Wong et al. |
| Terante | Filipino | Other Asian | Wong et al. |
| Terao | Japanese | Other Asian | Wong et al. |
| Teraoka | Japanese | Other Asian | Wong et al. |
| Terasaka | Japanese | Other Asian | Wong et al. |
| Terasaki | Japanese | Other Asian | Wong et al. |
| Terasawa | Japanese | Other Asian | Kranick et al. |
| Terashi | Japanese | Other Asian | Wong et al. |
| Terashima | Japanese | Other Asian | Wong et al. |
| Teratani | Japanese | Other Asian | Wong et al. |
| Terauchi | Japanese | Other Asian | Wong et al. |
| Terayama | Japanese | Other Asian | Wong et al. |
| Terazaki | Japanese | Other Asian | Wong et al. |
| Terazawa | Japanese | Other Asian | Wong et al. |
| Terencio | Filipino | Other Asian | Wong et al. |
| Ternio | Filipino | Other Asian | Wong et al. |
| Teroy | Filipino | Other Asian | Wong et al. |
| Terrado | Filipino | Other Asian | Wong et al. |
| Terre | Filipino | Other Asian | Wong et al. |
| Terrenal | Filipino | Other Asian | Wong et al. |
| Terrible | Filipino | Other Asian | Wong et al. |
| Tersol | Filipino | Other Asian | Wong et al. |
| Teruel | Filipino | Other Asian | Wong et al. |
| Terui | Japanese | Other Asian | Wong et al. |
| Terukina | Japanese | Other Asian | Wong et al. |
| Teruya | Japanese | Other Asian | Wong et al. |
| Tesalona | Filipino | Other Asian | Wong et al. |
| Teshima | Japanese | Other Asian | Wong et al. |
| Teshirogi | Japanese | Other Asian | Wong et al. |
| Tesorero | Filipino | Other Asian | Wong et al. |
| Tesorio | Filipino | Other Asian | Wong et al. |
| Tesoro | Filipino | Other Asian | Wong et al. |
| Testado | Filipino | Other Asian | Wong et al. |
| Teu | Chinese | Chinese | Wong et al. |
| Teung | Chinese | Chinese | Wong et al. |
| Teves | Filipino | Other Asian | Wong et al. |
| Tewari | Asian Indian | South Asian | Wong et al. |
| Tezuka | Japanese | Other Asian | Wong et al. |
| Thach | Vietnamese | Other Asian | Wong et al. |
| Thachet | Asian Indian | South Asian | Wong et al. |
| Thadathil | Asian Indian | South Asian | Wong et al. |
| Thai | Vietnamese | Other Asian | Wong et al. |
| Thakali |  | South Asian | Lim et al. |
| Thakar | Asian Indian | South Asian | Wong et al. |
| Thaker | Asian Indian | South Asian | Wong et al. |
| Thakkar | Asian Indian | South Asian | Wong et al. |
| Thakker | Asian Indian | South Asian | Wong et al. |
| Thakor | Asian Indian | South Asian | Wong et al. |
| Thakore | Asian Indian | South Asian | Wong et al. |
| Thakorlal | Asian Indian | South Asian | Wong et al. |
| Thakral | Asian Indian | South Asian | Wong et al. |
| Thakrar | Asian Indian | South Asian | Wong et al. |
| Thakur | Asian Indian | South Asian | Wong et al. |
| Tham | Vietnamese; Malaysian | Other Asian | Kranick et al. |
| Thamman | Asian Indian | South Asian | Wong et al. |
| Than | Burmese | Other Asian | Kranick et al. |
| Thanawala | Asian Indian | South Asian | Wong et al. |
| Thandi | Asian Indian | South Asian | Wong et al. |
| Thang | Vietnamese | Other Asian | Wong et al. |
| Thangavelu | Asian Indian | South Asian | Wong et al. |
| Thanh | Vietnamese | Other Asian | Wong et al. |
| Thanki | Asian Indian | South Asian | Wong et al. |
| Thapa |  | South Asian | Lim et al. |
| Thapar | Asian Indian | South Asian | Wong et al. |
| Thaper | Asian Indian | South Asian | Wong et al. |
| Tharakan | Asian Indian | South Asian | Wong et al. |
| Tharani | Asian Indian | South Asian | Wong et al. |
| Tharayil | Asian Indian | South Asian | Wong et al. |
| Tharian | Asian Indian | South Asian | Wong et al. |
| That | Vietnamese | Other Asian | Wong et al. |
| Thater | Filipino | Other Asian | Wong et al. |
| Thatte | Asian Indian | South Asian | Wong et al. |
| Thau | Vietnamese | Other Asian | Wong et al. |
| Thaver | Asian Indian | South Asian | Wong et al. |
| Thayaparan | Sri Lankan | South Asian | Kranick et al. |
| Thayil | Asian Indian | South Asian | Wong et al. |
| Them | Vietnamese | Other Asian | Wong et al. |
| Therattil | Asian Indian | South Asian | Wong et al. |
| Theung | Chinese | Chinese | Wong et al. |
| Thi | Vietnamese | Other Asian | Wong et al. |
| Thiagarajan | Asian Indian | South Asian | Wong et al. |
| Thiam | Filipino | Other Asian | Wong et al. |
| Thiara | Asian Indian | South Asian | Wong et al. |
| Thich | Vietnamese | Other Asian | Wong et al. |
| Thien | Vietnamese | Other Asian | Wong et al. |
| Thiep | Vietnamese | Other Asian | Wong et al. |
| Thiet | Vietnamese | Other Asian | Wong et al. |
| Thieu | Vietnamese | Other Asian | Wong et al. |
| Thind |  | South Asian | Lim et al. |
| Thinh | Vietnamese | Other Asian | Wong et al. |
| Thiruvengada | Asian Indian | South Asian | Wong et al. |
| Thiu | Chinese | Chinese | Wong et al. |
| Tho | Vietnamese | Other Asian | Wong et al. |
| Thoa | Vietnamese | Other Asian | Wong et al. |
| Thoai | Vietnamese | Other Asian | Wong et al. |
| Thobani | Asian Indian | South Asian | Wong et al. |
| Thoi | Vietnamese | Other Asian | Wong et al. |
| Thommy | Asian Indian | South Asian | Wong et al. |
| Thompson |  | South Asian | Lim et al. |
| Thoon | Chinese | Chinese | Wong et al. |
| Thoonkuzhy | Asian Indian | South Asian | Wong et al. |
| Thoppil | Asian Indian | South Asian | Wong et al. |
| Thorlaksson | Japanese | Other Asian | Wong et al. |
| Thoy | Chinese | Chinese | Wong et al. |
| Thu | Vietnamese | Other Asian | Wong et al. |
| Thua | Vietnamese | Other Asian | Wong et al. |
| Thuan | Vietnamese | Other Asian | Wong et al. |
| Thuc | Vietnamese | Other Asian | Wong et al. |
| Thui | Chinese | Chinese | Wong et al. |
| Thukral | Asian Indian | South Asian | Wong et al. |
| Thundathil | Asian Indian | South Asian | Wong et al. |
| Thundiyil | Asian Indian | South Asian | Wong et al. |
| Thuong | Vietnamese | Other Asian | Wong et al. |
| Thuy | Vietnamese | Other Asian | Wong et al. |
| Thuyen | Vietnamese | Other Asian | Wong et al. |
| Thuyet | Vietnamese | Other Asian | Wong et al. |
| Thyagarajan | Asian Indian | South Asian | Wong et al. |
| Ti | Chinese | Chinese | Wong et al. |
| Tiam | Filipino | Other Asian | Wong et al. |
| Tiambeng | Filipino | Other Asian | Wong et al. |
| Tiamson | Filipino | Other Asian | Wong et al. |
| Tiamzon | Filipino | Other Asian | Wong et al. |
| Tian | Chinese | Chinese | Wong et al. |
| Tiang | Chinese | Chinese | Wong et al. |
| Tiangco | Filipino | Other Asian | Kranick et al. |
| Tiangha | Filipino | Other Asian | Wong et al. |
| Tiangsing | Filipino | Other Asian | Wong et al. |
| Tiangson | Filipino | Other Asian | Wong et al. |
| Tiantawach | Thai | Other Asian | Kranick et al. |
| Tiao | Chinese | Chinese | Wong et al. |
| Tibay | Filipino | Other Asian | Wong et al. |
| Tibayan | Filipino | Other Asian | Wong et al. |
| Tibon | Filipino | Other Asian | Wong et al. |
| Tiburcio | Filipino | Other Asian | Wong et al. |
| Tica | Filipino | Other Asian | Wong et al. |
| Ticar | Filipino | Other Asian | Wong et al. |
| Tich | Vietnamese | Other Asian | Wong et al. |
| Ticman | Filipino | Other Asian | Wong et al. |
| Ticsay | Filipino | Other Asian | Wong et al. |
| Ticzon | Filipino | Other Asian | Wong et al. |
| Tidalgo | Filipino | Other Asian | Wong et al. |
| Tie | Chinese | Chinese | Wong et al. |
| Tiebhowan |  | South Asian | Lim et al. |
| Tieh | Chinese | Chinese | Wong et al. |
| Tiem | Vietnamese | Other Asian | Wong et al. |
| Tiempo | Filipino | Other Asian | Wong et al. |
| Tien | Vietnamese; Chinese | Other Asian | Kranick et al. |
| Tienzo | Filipino | Other Asian | Wong et al. |
| Tiep | Vietnamese | Other Asian | Wong et al. |
| Tierra | Filipino | Other Asian | Wong et al. |
| Tiet | Vietnamese | Other Asian | Wong et al. |
| Tieu | Vietnamese | Other Asian | Wong et al. |
| Tigcal | Filipino | Other Asian | Wong et al. |
| Tiglao | Filipino | Other Asian | Wong et al. |
| Tigno | Filipino | Other Asian | Wong et al. |
| Tijam | Filipino | Other Asian | Wong et al. |
| Tikku | Asian Indian | South Asian | Wong et al. |
| Tiku | Asian Indian | South Asian | Wong et al. |
| Tila | Filipino | Other Asian | Wong et al. |
| Tilan | Filipino | Other Asian | Wong et al. |
| Tillo | Filipino | Other Asian | Wong et al. |
| Tilos | Filipino | Other Asian | Wong et al. |
| Tim | Chinese | Chinese | Wong et al. |
| Timario | Filipino | Other Asian | Wong et al. |
| Timbal | Filipino | Other Asian | Wong et al. |
| Timbang | Filipino | Other Asian | Wong et al. |
| Timbol | Filipino | Other Asian | Wong et al. |
| Timbreza | Filipino | Other Asian | Wong et al. |
| Timilsina |  | South Asian | Lim et al. |
| Timog | Filipino | Other Asian | Wong et al. |
| Timoteo | Filipino | Other Asian | Wong et al. |
| Timple | Filipino | Other Asian | Wong et al. |
| Timtim | Filipino | Other Asian | Wong et al. |
| Timtiman | Filipino | Other Asian | Wong et al. |
| Tin | Chinese | Chinese | Wong et al. |
| Tina | Filipino | Other Asian | Wong et al. |
| Tinawin | Filipino | Other Asian | Wong et al. |
| Tinaza | Filipino | Other Asian | Wong et al. |
| Ting | Chinese | Chinese | Kranick et al. |
| Tinga | Filipino | Other Asian | Wong et al. |
| Tingin | Filipino | Other Asian | Wong et al. |
| Tingson | Filipino | Other Asian | Wong et al. |
| Tingzon | Filipino | Other Asian | Wong et al. |
| Tinh | Vietnamese | Other Asian | Wong et al. |
| Tinio | Filipino | Other Asian | Wong et al. |
| Tinitigan | Filipino | Other Asian | Wong et al. |
| Tinni |  | South Asian | Lim et al. |
| Tinoso | Filipino | Other Asian | Wong et al. |
| Tinsay | Filipino | Other Asian | Wong et al. |
| Tintiangco | Filipino | Other Asian | Wong et al. |
| Tio | Filipino | Other Asian | Wong et al. |
| Tiomico | Filipino | Other Asian | Wong et al. |
| Tiong | Filipino | Other Asian | Wong et al. |
| Tiongco | Filipino | Other Asian | Wong et al. |
| Tiongko | Filipino | Other Asian | Wong et al. |
| Tiongson | Filipino | Other Asian | Wong et al. |
| Tionko | Filipino | Other Asian | Wong et al. |
| Tioseco | Filipino | Other Asian | Wong et al. |
| Tiotuico | Filipino | Other Asian | Wong et al. |
| Tiozon | Filipino | Other Asian | Wong et al. |
| Tip | Chinese | Chinese | Wong et al. |
| Tipnis | Asian Indian | South Asian | Wong et al. |
| Tipon | Filipino | Other Asian | Wong et al. |
| Tique | Filipino | Other Asian | Wong et al. |
| Tiqui | Filipino | Other Asian | Wong et al. |
| Tira | Filipino | Other Asian | Wong et al. |
| Tirado | Filipino | Other Asian | Wong et al. |
| Tirador | Filipino | Other Asian | Wong et al. |
| Tirao | Filipino | Other Asian | Wong et al. |
| Tiratira | Filipino | Other Asian | Wong et al. |
| Tirazona | Filipino | Other Asian | Wong et al. |
| Tirbeni |  | South Asian | Lim et al. |
| Tirmizi | Asian Indian | South Asian | Wong et al. |
| Tiro | Filipino | Other Asian | Wong et al. |
| Tirol | Filipino | Other Asian | Wong et al. |
| Tirona | Filipino | Other Asian | Wong et al. |
| Tirso | Filipino | Other Asian | Wong et al. |
| Tirumala | Asian Indian | South Asian | Wong et al. |
| Tisado | Filipino | Other Asian | Wong et al. |
| Tisbe | Filipino | Other Asian | Wong et al. |
| Tismo | Filipino | Other Asian | Wong et al. |
| Titong | Filipino | Other Asian | Wong et al. |
| Titu |  | South Asian | Lim et al. |
| Tiu | Filipino | Other Asian | Kranick et al. |
| Tividad | Filipino | Other Asian | Wong et al. |
| Tiwana | Asian Indian | South Asian | Wong et al. |
| Tiwari | Asian Indian | South Asian | Wong et al. |
| Tizon | Filipino | Other Asian | Wong et al. |
| Tjo | Korean | Other Asian | Wong et al. |
| Tjon | Chinese | Chinese | Wong et al. |
| Tng | Chinese | Chinese | Wong et al. |
| To | Chinese; Vietnamese | Chinese | Kranick et al. |
| Toa | Chinese | Chinese | Wong et al. |
| Toan | Vietnamese | Other Asian | Wong et al. |
| Toba | Japanese | Other Asian | Wong et al. |
| Tobari | Japanese | Other Asian | Wong et al. |
| Tobaru | Japanese | Other Asian | Wong et al. |
| Tobita | Japanese | Other Asian | Wong et al. |
| Tobo | Japanese | Other Asian | Wong et al. |
| Tochio | Japanese | Other Asian | Wong et al. |
| Toda | Japanese | Other Asian | Wong et al. |
| Todo | Japanese | Other Asian | Wong et al. |
| Todoroki | Japanese | Other Asian | Wong et al. |
| Togami | Japanese | Other Asian | Wong et al. |
| Togashi | Japanese | Other Asian | Wong et al. |
| Togawa | Japanese | Other Asian | Wong et al. |
| Togi | Japanese | Other Asian | Wong et al. |
| Togle | Filipino | Other Asian | Wong et al. |
| Togo | Japanese | Other Asian | Wong et al. |
| Togonon | Filipino | Other Asian | Wong et al. |
| Toguchi | Japanese | Other Asian | Wong et al. |
| Tohyama | Japanese | Other Asian | Wong et al. |
| Toida | Japanese | Other Asian | Wong et al. |
| Toji | Japanese | Other Asian | Wong et al. |
| Tojima | Japanese | Other Asian | Wong et al. |
| Tojino | Filipino | Other Asian | Wong et al. |
| Tojo | Japanese | Other Asian | Wong et al. |
| Tojong | Filipino | Other Asian | Wong et al. |
| Tok | Korean | Other Asian | Wong et al. |
| Tokashiki | Japanese | Other Asian | Wong et al. |
| Tokeshi | Japanese | Other Asian | Wong et al. |
| Toki | Japanese | Other Asian | Wong et al. |
| Tokita | Japanese | Other Asian | Wong et al. |
| Tokiwa | Japanese | Other Asian | Wong et al. |
| Tokko | Korean | Other Asian | Wong et al. |
| Tokoro | Japanese | Other Asian | Wong et al. |
| Toku | Japanese | Other Asian | Wong et al. |
| Tokuda | Japanese | Other Asian | Wong et al. |
| Tokugawa | Japanese | Other Asian | Wong et al. |
| Tokuhiro | Japanese | Other Asian | Wong et al. |
| Tokuhisa | Japanese | Other Asian | Wong et al. |
| Tokui | Japanese | Other Asian | Wong et al. |
| Tokumaru | Japanese | Other Asian | Wong et al. |
| Tokumasu | Japanese | Other Asian | Wong et al. |
| Tokumitsu | Japanese | Other Asian | Wong et al. |
| Tokumoto | Japanese | Other Asian | Wong et al. |
| Tokunaga | Japanese | Other Asian | Wong et al. |
| Tokuno | Japanese | Other Asian | Wong et al. |
| Tokuoka | Japanese | Other Asian | Wong et al. |
| Tokura | Japanese | Other Asian | Wong et al. |
| Tokushige | Japanese | Other Asian | Wong et al. |
| Tokushima | Japanese | Other Asian | Wong et al. |
| Tokuyama | Japanese | Other Asian | Wong et al. |
| Tokuyasu | Japanese | Other Asian | Wong et al. |
| Tol | Korean | Other Asian | Wong et al. |
| Tolat | Asian Indian | South Asian | Wong et al. |
| Toledano | Filipino | Other Asian | Wong et al. |
| Toledo | Filipino | Other Asian | Wong et al. |
| Tolenada | Filipino | Other Asian | Wong et al. |
| Tolentino | Filipino | Other Asian | Wong et al. |
| Tolero | Filipino | Other Asian | Wong et al. |
| Tolete | Filipino | Other Asian | Wong et al. |
| Tolia | Asian Indian | South Asian | Wong et al. |
| Toliao | Filipino | Other Asian | Wong et al. |
| Tolibas | Filipino | Other Asian | Wong et al. |
| Tolintino | Filipino | Other Asian | Wong et al. |
| Tolo | Filipino | Other Asian | Wong et al. |
| Tolores | Filipino | Other Asian | Wong et al. |
| Tolosa | Filipino | Other Asian | Wong et al. |
| Tom | Chinese | Chinese | Kranick et al. |
| Toma | Japanese | Other Asian | Wong et al. |
| Tomabechi | Japanese | Other Asian | Wong et al. |
| Tomacder | Filipino | Other Asian | Wong et al. |
| Tomacruz | Filipino | Other Asian | Wong et al. |
| Tomada | Filipino | Other Asian | Wong et al. |
| Tomal |  | South Asian | Lim et al. |
| Tomaneng | Filipino | Other Asian | Wong et al. |
| Tomar | Asian Indian | South Asian | Wong et al. |
| Tomari | Japanese | Other Asian | Wong et al. |
| Tomas | Filipino | Other Asian | Wong et al. |
| Tomatani | Japanese | Other Asian | Wong et al. |
| Tombo | Filipino | Other Asian | Wong et al. |
| Tomboc | Filipino | Other Asian | Wong et al. |
| Tome | Japanese | Other Asian | Wong et al. |
| Tomeldan | Filipino | Other Asian | Wong et al. |
| Tomelden | Filipino | Other Asian | Wong et al. |
| Tomi | Japanese | Other Asian | Wong et al. |
| Tomihara | Japanese | Other Asian | Wong et al. |
| Tomii | Japanese | Other Asian | Wong et al. |
| Tomikawa | Japanese | Other Asian | Wong et al. |
| Tomilloso | Filipino | Other Asian | Wong et al. |
| Tomimatsu | Japanese | Other Asian | Wong et al. |
| Tomimbang | Filipino | Other Asian | Wong et al. |
| Tomimoto | Japanese | Other Asian | Wong et al. |
| Tomimura | Japanese | Other Asian | Wong et al. |
| Tominaga | Japanese | Other Asian | Wong et al. |
| Tomines | Filipino | Other Asian | Wong et al. |
| Tomioka | Japanese | Other Asian | Wong et al. |
| Tomishima | Japanese | Other Asian | Wong et al. |
| Tomita | Japanese | Other Asian | Kranick et al. |
| Tomiyama | Japanese | Other Asian | Wong et al. |
| Tomiyasu | Japanese | Other Asian | Wong et al. |
| Tomizawa | Japanese | Other Asian | Wong et al. |
| Tomizuka | Japanese | Other Asian | Wong et al. |
| Tomoda | Japanese | Other Asian | Wong et al. |
| Tomoeda | Japanese | Other Asian | Wong et al. |
| Tomonaga | Japanese | Other Asian | Wong et al. |
| Tomonari | Japanese | Other Asian | Wong et al. |
| Tomono | Japanese | Other Asian | Wong et al. |
| Tomooka | Japanese | Other Asian | Wong et al. |
| Tomori | Japanese | Other Asian | Wong et al. |
| Tomoyoshi | Japanese | Other Asian | Wong et al. |
| Tomura | Japanese | Other Asian | Wong et al. |
| Ton | Vietnamese | Other Asian | Wong et al. |
| Tonai | Japanese | Other Asian | Wong et al. |
| Tondo | Filipino | Other Asian | Wong et al. |
| Tonegawa | Japanese | Other Asian | Wong et al. |
| Tonel | Filipino | Other Asian | Wong et al. |
| Tong | Chinese | Chinese | Kranick et al. |
| Tongco | Filipino | Other Asian | Wong et al. |
| Tongohan | Filipino | Other Asian | Wong et al. |
| Tongol | Filipino | Other Asian | Wong et al. |
| Tongson | Filipino | Other Asian | Wong et al. |
| Tongu | Japanese | Other Asian | Wong et al. |
| Tonogbanua | Filipino | Other Asian | Wong et al. |
| Tonomura | Japanese | Other Asian | Wong et al. |
| Tonooka | Japanese | Other Asian | Wong et al. |
| Tonthat | Vietnamese | Other Asian | Wong et al. |
| Too | Chinese | Chinese | Wong et al. |
| Toong | Chinese | Chinese | Wong et al. |
| Toor | Asian Indian | South Asian | Wong et al. |
| Topacio | Filipino | Other Asian | Wong et al. |
| Topenio | Filipino | Other Asian | Wong et al. |
| Topinio | Filipino | Other Asian | Wong et al. |
| Topiwala | Asian Indian | South Asian | Wong et al. |
| Toquero | Filipino | Other Asian | Wong et al. |
| Tora | Japanese | Other Asian | Wong et al. |
| Toral | Filipino | Other Asian | Wong et al. |
| Toralba | Filipino | Other Asian | Wong et al. |
| Torao | Japanese | Other Asian | Wong et al. |
| Torcuato | Filipino | Other Asian | Wong et al. |
| Torculas | Filipino | Other Asian | Wong et al. |
| Torda | Filipino | Other Asian | Wong et al. |
| Tordecilla | Filipino | Other Asian | Wong et al. |
| Tordesillas | Filipino | Other Asian | Wong et al. |
| Tordil | Filipino | Other Asian | Wong et al. |
| Torga | Filipino | Other Asian | Wong et al. |
| Toribio | Filipino | Other Asian | Wong et al. |
| Torigoe | Japanese | Other Asian | Wong et al. |
| Torii | Japanese | Other Asian | Wong et al. |
| Torikai | Japanese | Other Asian | Wong et al. |
| Torino | Filipino | Other Asian | Wong et al. |
| Torio | Filipino | Other Asian | Wong et al. |
| Toriumi | Japanese | Other Asian | Wong et al. |
| Toriyabe | Japanese | Other Asian | Wong et al. |
| Toriyama | Japanese | Other Asian | Wong et al. |
| Tormon | Filipino | Other Asian | Wong et al. |
| Torne | Filipino | Other Asian | Wong et al. |
| Torng | Chinese | Chinese | Wong et al. |
| Torno | Filipino | Other Asian | Wong et al. |
| Toro | Chinese | Chinese | Wong et al. |
| Torofder |  | South Asian | Lim et al. |
| Torqueza | Filipino | Other Asian | Wong et al. |
| Torrado | Filipino | Other Asian | Wong et al. |
| Torralba | Filipino | Other Asian | Wong et al. |
| Torrecampo | Filipino | Other Asian | Wong et al. |
| Torrecer | Filipino | Other Asian | Wong et al. |
| Torrefiel | Filipino | Other Asian | Wong et al. |
| Torrefranca | Filipino | Other Asian | Wong et al. |
| Torrejos | Filipino | Other Asian | Wong et al. |
| Torreliza | Filipino | Other Asian | Wong et al. |
| Torrella | Filipino | Other Asian | Wong et al. |
| Torrente | Filipino | Other Asian | Wong et al. |
| Torreon | Filipino | Other Asian | Wong et al. |
| Torres | Filipino | Other Asian | Wong et al. |
| Torrevillas | Filipino | Other Asian | Wong et al. |
| Torrez | Filipino | Other Asian | Wong et al. |
| Torricer | Filipino | Other Asian | Wong et al. |
| Torrijos | Filipino | Other Asian | Wong et al. |
| Tortal | Filipino | Other Asian | Wong et al. |
| Tortona | Filipino | Other Asian | Wong et al. |
| Tosaka | Japanese | Other Asian | Wong et al. |
| Toshima | Japanese | Other Asian | Wong et al. |
| Toshio | Japanese | Other Asian | Wong et al. |
| Tot | Vietnamese | Other Asian | Wong et al. |
| Totaan | Filipino | Other Asian | Wong et al. |
| Totanes | Filipino | Other Asian | Wong et al. |
| Totani | Japanese | Other Asian | Wong et al. |
| Totaram |  | South Asian | Lim et al. |
| Totengco | Filipino | Other Asian | Wong et al. |
| Totsuka | Japanese | Other Asian | Wong et al. |
| Tou | Chinese | Chinese | Wong et al. |
| Toung | Chinese | Chinese | Wong et al. |
| Tovera | Filipino | Other Asian | Wong et al. |
| Tow | Chinese | Chinese | Kranick et al. |
| Toy | Chinese | Chinese | Wong et al. |
| Toya | Japanese | Other Asian | Wong et al. |
| Toyama | Japanese | Other Asian | Wong et al. |
| Toyoda | Japanese | Other Asian | Wong et al. |
| Toyofuku | Japanese | Other Asian | Wong et al. |
| Toyohara | Japanese | Other Asian | Wong et al. |
| Toyokawa | Japanese | Other Asian | Wong et al. |
| Toyonaga | Japanese | Other Asian | Wong et al. |
| Toyooka | Japanese | Other Asian | Wong et al. |
| Toyoshima | Japanese | Other Asian | Wong et al. |
| Toyota | Japanese | Other Asian | Wong et al. |
| Tozaki | Japanese | Other Asian | Wong et al. |
| Tozawa | Japanese | Other Asian | Wong et al. |
| Tra | Vietnamese | Other Asian | Wong et al. |
| Trac | Vietnamese | Other Asian | Wong et al. |
| Trach | Vietnamese | Other Asian | Wong et al. |
| Trajano | Filipino | Other Asian | Wong et al. |
| Traje | Filipino | Other Asian | Wong et al. |
| Tram | Vietnamese | Other Asian | Wong et al. |
| Tran | Vietnamese | Other Asian | Kranick et al. |
| Trance | Filipino | Other Asian | Wong et al. |
| Trang | Vietnamese | Other Asian | Wong et al. |
| Tranh | Vietnamese | Other Asian | Wong et al. |
| Tranquilino | Filipino | Other Asian | Wong et al. |
| Transfigurac | Filipino | Other Asian | Wong et al. |
| Tranvan | Vietnamese | Other Asian | Wong et al. |
| Trapsi | Filipino | Other Asian | Wong et al. |
| Trasmonte | Filipino | Other Asian | Wong et al. |
| Trat | Vietnamese | Other Asian | Wong et al. |
| Trau | Vietnamese | Other Asian | Wong et al. |
| Travilla | Filipino | Other Asian | Wong et al. |
| Traya | Filipino | Other Asian | Wong et al. |
| Trazo | Filipino | Other Asian | Wong et al. |
| Trehan | Asian Indian | South Asian | Wong et al. |
| Trenh | Vietnamese | Other Asian | Wong et al. |
| Trevias | Filipino | Other Asian | Wong et al. |
| Treyes | Filipino | Other Asian | Wong et al. |
| Tri | Vietnamese | Other Asian | Wong et al. |
| Tria | Filipino | Other Asian | Wong et al. |
| Trias | Filipino | Other Asian | Wong et al. |
| Trien | Vietnamese | Other Asian | Wong et al. |
| Trieu | Vietnamese | Other Asian | Kranick et al. |
| Trillana | Filipino | Other Asian | Wong et al. |
| Trillanes | Filipino | Other Asian | Wong et al. |
| Trilles | Filipino | Other Asian | Wong et al. |
| Trillo | Filipino | Other Asian | Wong et al. |
| Triloki |  | South Asian | Lim et al. |
| Trimor | Filipino | Other Asian | Wong et al. |
| Trinanes | Filipino | Other Asian | Wong et al. |
| Trinh | Vietnamese | Other Asian | Wong et al. |
| Trinidad | Filipino | Other Asian | Wong et al. |
| Trinos | Filipino | Other Asian | Kranick et al. |
| Trio | Filipino | Other Asian | Wong et al. |
| Tripathi | Asian Indian | South Asian | Wong et al. |
| Tripathy | Asian Indian | South Asian | Wong et al. |
| Triunfo | Filipino | Other Asian | Wong et al. |
| Trivedi | Asian Indian | South Asian | Wong et al. |
| Trivino | Filipino | Other Asian | Wong et al. |
| Trocino | Filipino | Other Asian | Wong et al. |
| Trocio | Filipino | Other Asian | Wong et al. |
| Trompeta | Filipino | Other Asian | Wong et al. |
| Tron | Vietnamese | Other Asian | Wong et al. |
| Tronco | Filipino | Other Asian | Wong et al. |
| Troncoso | Filipino | Other Asian | Wong et al. |
| Trong | Vietnamese | Other Asian | Wong et al. |
| Trono | Filipino | Other Asian | Wong et al. |
| Troung | Vietnamese | Other Asian | Wong et al. |
| Trovela | Filipino | Other Asian | Wong et al. |
| Tru | Vietnamese | Other Asian | Wong et al. |
| Truc | Vietnamese | Other Asian | Wong et al. |
| Trujillo | Filipino | Other Asian | Wong et al. |
| Trumata | Filipino | Other Asian | Wong et al. |
| Trung | Vietnamese | Other Asian | Wong et al. |
| Truong | Vietnamese | Other Asian | Kranick et al. |
| Truyen | Vietnamese | Other Asian | Wong et al. |
| Trvong | Vietnamese | Other Asian | Wong et al. |
| Tsa | Chinese | Chinese | Kranick et al. |
| Tsai | Chinese | Chinese | Kranick et al. |
| Tsan | Chinese | Chinese | Wong et al. |
| Tsang | Chinese | Chinese | Kranick et al. |
| Tsao | Chinese | Chinese | Wong et al. |
| Tsau | Chinese | Chinese | Wong et al. |
| Tsaur | Chinese | Chinese | Wong et al. |
| Tsay | Chinese | Chinese | Wong et al. |
| Tse | Chinese | Chinese | Kranick et al. |
| Tsen | Chinese | Chinese | Wong et al. |
| Tseng | Chinese | Chinese | Wong et al. |
| Tsering |  | South Asian | Lim et al. |
| Tserng | Chinese | Chinese | Wong et al. |
| Tseu | Chinese | Chinese | Wong et al. |
| Tseung | Chinese | Chinese | Kranick et al. |
| Tsi | Chinese | Chinese | Wong et al. |
| Tsia | Chinese | Chinese | Wong et al. |
| Tsiang | Chinese | Chinese | Wong et al. |
| Tsiao | Chinese | Chinese | Wong et al. |
| Tsien | Chinese | Chinese | Wong et al. |
| Tsim | Chinese | Chinese | Wong et al. |
| Tsin | Chinese | Chinese | Wong et al. |
| Tsing | Chinese | Chinese | Wong et al. |
| Tsiu | Chinese | Chinese | Wong et al. |
| Tso | Chinese | Chinese | Kranick et al. |
| Tsoi | Chinese | Chinese | Wong et al. |
| Tson | Chinese | Chinese | Wong et al. |
| Tsong | Chinese | Chinese | Wong et al. |
| Tsou | Chinese | Chinese | Wong et al. |
| Tsow | Chinese | Chinese | Wong et al. |
| Tsu | Chinese | Chinese | Wong et al. |
| Tsuan | Chinese | Chinese | Wong et al. |
| Tsuang | Chinese | Chinese | Wong et al. |
| Tsubaki | Japanese | Other Asian | Wong et al. |
| Tsuboi | Japanese | Other Asian | Wong et al. |
| Tsubokawa | Japanese | Other Asian | Wong et al. |
| Tsubone | Japanese | Other Asian | Wong et al. |
| Tsubota | Japanese | Other Asian | Wong et al. |
| Tsubouchi | Japanese | Other Asian | Wong et al. |
| Tsuchida | Japanese | Other Asian | Wong et al. |
| Tsuchihashi | Japanese | Other Asian | Wong et al. |
| Tsuchikawa | Japanese | Other Asian | Wong et al. |
| Tsuchimoto | Japanese | Other Asian | Wong et al. |
| Tsuchiya | Japanese | Other Asian | Wong et al. |
| Tsuchiyama | Japanese | Other Asian | Wong et al. |
| Tsuda | Japanese | Other Asian | Wong et al. |
| Tsue | Chinese | Chinese | Wong et al. |
| Tsuei | Chinese | Chinese | Wong et al. |
| Tsugami | Japanese | Other Asian | Wong et al. |
| Tsugawa | Japanese | Other Asian | Wong et al. |
| Tsuge | Japanese | Other Asian | Wong et al. |
| Tsuha | Japanese | Other Asian | Wong et al. |
| Tsui | Chinese | Chinese | Kranick et al. |
| Tsuiki | Japanese | Other Asian | Wong et al. |
| Tsuji | Japanese | Other Asian | Wong et al. |
| Tsujii | Japanese | Other Asian | Wong et al. |
| Tsujimoto | Japanese | Other Asian | Wong et al. |
| Tsujimura | Japanese | Other Asian | Wong et al. |
| Tsujita | Japanese | Other Asian | Wong et al. |
| Tsujiuchi | Japanese | Other Asian | Wong et al. |
| Tsukada | Japanese | Other Asian | Wong et al. |
| Tsukagoshi | Japanese | Other Asian | Wong et al. |
| Tsukahara | Japanese | Other Asian | Wong et al. |
| Tsukamoto | Japanese | Other Asian | Wong et al. |
| Tsukasa | Japanese | Other Asian | Wong et al. |
| Tsukayama | Japanese | Other Asian | Wong et al. |
| Tsukiji | Japanese | Other Asian | Wong et al. |
| Tsukioka | Japanese | Other Asian | Wong et al. |
| Tsukiyama | Japanese | Other Asian | Wong et al. |
| Tsukuda | Japanese | Other Asian | Wong et al. |
| Tsukui | Japanese | Other Asian | Wong et al. |
| Tsumura | Japanese | Other Asian | Wong et al. |
| Tsun | Chinese | Chinese | Wong et al. |
| Tsunashima | Japanese | Other Asian | Wong et al. |
| Tsunekawa | Japanese | Other Asian | Wong et al. |
| Tsunematsu | Japanese | Other Asian | Wong et al. |
| Tsuneyoshi | Japanese | Other Asian | Wong et al. |
| Tsung | Chinese | Chinese | Wong et al. |
| Tsuno | Japanese | Other Asian | Wong et al. |
| Tsunoda | Japanese | Other Asian | Wong et al. |
| Tsunoi | Japanese | Other Asian | Wong et al. |
| Tsuru | Japanese | Other Asian | Wong et al. |
| Tsuruda | Japanese | Other Asian | Wong et al. |
| Tsurudome | Japanese | Other Asian | Wong et al. |
| Tsuruga | Japanese | Other Asian | Wong et al. |
| Tsuruhara | Japanese | Other Asian | Wong et al. |
| Tsurumaki | Japanese | Other Asian | Wong et al. |
| Tsurumi | Japanese | Other Asian | Wong et al. |
| Tsuruoka | Japanese | Other Asian | Wong et al. |
| Tsuruta | Japanese | Other Asian | Wong et al. |
| Tsushima | Japanese | Other Asian | Wong et al. |
| Tsutsui | Japanese | Other Asian | Wong et al. |
| Tsutsumi | Japanese | Other Asian | Wong et al. |
| Tsuya | Japanese | Other Asian | Wong et al. |
| Tsuyama | Japanese | Other Asian | Wong et al. |
| Tsuyuki | Japanese | Other Asian | Wong et al. |
| Tsuzuki | Japanese | Other Asian | Wong et al. |
| Tsz | Chinese | Chinese | Wong et al. |
| Tu | Chinese; Vietnamese | Chinese | Kranick et al. |
| Tua | Filipino | Other Asian | Wong et al. |
| Tualla | Filipino | Other Asian | Wong et al. |
| Tuan | Chinese | Chinese | Wong et al. |
| Tuangco | Filipino | Other Asian | Wong et al. |
| Tuanmu | Chinese | Chinese | Wong et al. |
| Tuano | Filipino | Other Asian | Wong et al. |
| Tuanquin | Filipino | Other Asian | Wong et al. |
| Tuason | Filipino | Other Asian | Wong et al. |
| Tuat | Vietnamese | Other Asian | Wong et al. |
| Tuazon | Filipino | Other Asian | Wong et al. |
| Tubal | Filipino | Other Asian | Wong et al. |
| Tubao | Filipino | Other Asian | Wong et al. |
| Tubera | Filipino | Other Asian | Wong et al. |
| Tubianosa | Filipino | Other Asian | Wong et al. |
| Tubiera | Filipino | Other Asian | Wong et al. |
| Tubig | Filipino | Other Asian | Wong et al. |
| Tuble | Filipino | Other Asian | Wong et al. |
| Tubo | Filipino | Other Asian | Wong et al. |
| Tubon | Filipino | Other Asian | Wong et al. |
| Tubongbanua | Filipino | Other Asian | Wong et al. |
| Tuc | Vietnamese | Other Asian | Wong et al. |
| Tucay | Filipino | Other Asian | Wong et al. |
| Tud | Filipino | Other Asian | Wong et al. |
| Tudtud | Filipino | Other Asian | Wong et al. |
| Tue | Chinese | Chinese | Wong et al. |
| Tuen | Chinese | Chinese | Wong et al. |
| Tuey | Chinese | Chinese | Wong et al. |
| Tufail | Pakistani | South Asian | Kranick et al. |
| Tugab | Filipino | Other Asian | Wong et al. |
| Tugade | Filipino | Other Asian | Wong et al. |
| Tugadi | Filipino | Other Asian | Wong et al. |
| Tugado | Filipino | Other Asian | Wong et al. |
| Tugaoen | Filipino | Other Asian | Wong et al. |
| Tugas | Filipino | Other Asian | Wong et al. |
| Tugbang | Filipino | Other Asian | Wong et al. |
| Tuhin |  | South Asian | Lim et al. |
| Tui | Chinese | Chinese | Wong et al. |
| Tuico | Filipino | Other Asian | Wong et al. |
| Tukdeo | Asian Indian | South Asian | Kranick et al. |
| Tulabot | Filipino | Other Asian | Wong et al. |
| Tulabut | Filipino | Other Asian | Wong et al. |
| Tulagan | Filipino | Other Asian | Wong et al. |
| Tulao | Filipino | Other Asian | Wong et al. |
| Tulfo | Filipino | Other Asian | Wong et al. |
| Tuli | Asian Indian | South Asian | Wong et al. |
| Tuliao | Filipino | Other Asian | Wong et al. |
| Tulio | Filipino | Other Asian | Wong et al. |
| Tullao | Filipino | Other Asian | Wong et al. |
| Tulod | Filipino | Other Asian | Wong et al. |
| Tulshi |  | South Asian | Lim et al. |
| Tulud | Filipino | Other Asian | Wong et al. |
| Tum | Chinese | Chinese | Wong et al. |
| Tumacder | Filipino | Other Asian | Wong et al. |
| Tumale | Filipino | Other Asian | Wong et al. |
| Tumaliuan | Filipino | Other Asian | Wong et al. |
| Tumamak | Filipino | Other Asian | Wong et al. |
| Tumamao | Filipino | Other Asian | Wong et al. |
| Tumambing | Filipino | Other Asian | Wong et al. |
| Tumanda | Filipino | Other Asian | Wong et al. |
| Tumaneng | Filipino | Other Asian | Wong et al. |
| Tumang | Filipino | Other Asian | Wong et al. |
| Tumanon | Filipino | Other Asian | Wong et al. |
| Tumbaga | Filipino | Other Asian | Wong et al. |
| Tumber | Asian Indian | South Asian | Wong et al. |
| Tumbocon | Filipino | Other Asian | Wong et al. |
| Tumbokon | Filipino | Other Asian | Wong et al. |
| Tummala | Asian Indian | South Asian | Wong et al. |
| Tumulak | Filipino | Other Asian | Wong et al. |
| Tun | Burmese | Other Asian | Kranick et al. |
| Tunac | Filipino | Other Asian | Wong et al. |
| Tunacao | Filipino | Other Asian | Wong et al. |
| Tunay | Filipino | Other Asian | Wong et al. |
| Tung | Chinese; Vietnamese | Chinese | Kranick et al. |
| Tungala | Filipino | Other Asian | Wong et al. |
| Tungka | Indonesian | Other Asian | Kranick et al. |
| Tungol | Filipino | Other Asian | Wong et al. |
| Tungpalan | Filipino | Other Asian | Wong et al. |
| Tungul | Filipino | Other Asian | Wong et al. |
| Tunque | Filipino | Other Asian | Wong et al. |
| Tuoc | Vietnamese | Other Asian | Wong et al. |
| Tuoi | Vietnamese | Other Asian | Wong et al. |
| Tuong | Vietnamese | Other Asian | Wong et al. |
| Tupas | Filipino | Other Asian | Wong et al. |
| Tupaz | Filipino | Other Asian | Wong et al. |
| Tupinio | Filipino | Other Asian | Wong et al. |
| Tuquero | Filipino | Other Asian | Wong et al. |
| Tura | Filipino | Other Asian | Wong et al. |
| Turakhia | Asian Indian | South Asian | Wong et al. |
| Turgano | Filipino | Other Asian | Wong et al. |
| Turiano | Filipino | Other Asian | Wong et al. |
| Turingan | Filipino | Other Asian | Wong et al. |
| Turla | Filipino | Other Asian | Wong et al. |
| Turqueza | Filipino | Other Asian | Wong et al. |
| Tuscano | Filipino | Other Asian | Wong et al. |
| Tut | Asian Indian | South Asian | Wong et al. |
| Tutaan | Filipino | Other Asian | Wong et al. |
| Tutanes | Filipino | Other Asian | Wong et al. |
| Tuvera | Filipino | Other Asian | Wong et al. |
| Tuyen | Vietnamese | Other Asian | Wong et al. |
| Tuyet | Vietnamese | Other Asian | Wong et al. |
| Tuyor | Filipino | Other Asian | Wong et al. |
| Tuzon | Filipino | Other Asian | Wong et al. |
| Twan | Chinese | Chinese | Wong et al. |
| Twu | Chinese | Chinese | Wong et al. |
| Tyagi | Asian Indian | South Asian | Wong et al. |
| Tyan | Chinese | Chinese | Wong et al. |
| Tze | Chinese | Chinese | Wong et al. |
| Tzen | Chinese | Chinese | Wong et al. |
| Tzeng | Chinese | Chinese | Wong et al. |
| Tzo | Chinese | Chinese | Wong et al. |
| Tzong | Chinese | Chinese | Wong et al. |
| Tzou | Chinese | Chinese | Wong et al. |
| Tzu | Chinese | Chinese | Wong et al. |
| Uang | Chinese | Chinese | Wong et al. |
| Ubalde | Filipino | Other Asian | Wong et al. |
| Ubaldo | Filipino | Other Asian | Wong et al. |
| Ubando | Filipino | Other Asian | Wong et al. |
| Ubanos | Filipino | Other Asian | Kranick et al. |
| Ubas | Filipino | Other Asian | Wong et al. |
| Ubay | Filipino | Other Asian | Wong et al. |
| Uberita | Filipino | Other Asian | Wong et al. |
| Uberoi | Asian Indian | South Asian | Wong et al. |
| Ubhi | Asian Indian | South Asian | Wong et al. |
| Ubongen | Filipino | Other Asian | Wong et al. |
| Ubukata | Japanese | Other Asian | Wong et al. |
| Ubungen | Filipino | Other Asian | Wong et al. |
| Uchi | Filipino | Other Asian | Wong et al. |
| Uchibori | Japanese | Other Asian | Wong et al. |
| Uchida | Japanese | Other Asian | Wong et al. |
| Uchikawa | Japanese | Other Asian | Wong et al. |
| Uchikoshi | Japanese | Other Asian | Wong et al. |
| Uchima | Japanese | Other Asian | Wong et al. |
| Uchimura | Japanese | Other Asian | Wong et al. |
| Uchino | Japanese | Other Asian | Wong et al. |
| Uchio | Japanese | Other Asian | Wong et al. |
| Uchiumi | Japanese | Other Asian | Wong et al. |
| Uchiyama | Japanese | Other Asian | Wong et al. |
| Uchizono | Japanese | Other Asian | Wong et al. |
| Uclaray | Filipino | Other Asian | Wong et al. |
| Ucol | Filipino | Other Asian | Wong et al. |
| Uda | Japanese | Other Asian | Wong et al. |
| Udagawa | Japanese | Other Asian | Wong et al. |
| Udai |  | South Asian | Lim et al. |
| Udan | Filipino | Other Asian | Wong et al. |
| Udani | Asian Indian | South Asian | Wong et al. |
| Udarbe | Filipino | Other Asian | Wong et al. |
| Udasco | Filipino | Other Asian | Wong et al. |
| Udaundo | Filipino | Other Asian | Wong et al. |
| Uddeen |  | South Asian | Lim et al. |
| Uddin | Bangladeshi | South Asian | Kranick et al. |
| Udeshi | Asian Indian | South Asian | Kranick et al. |
| Udit |  | South Asian | Lim et al. |
| Udo | Japanese | Other Asian | Wong et al. |
| Udvadia | Asian Indian | South Asian | Wong et al. |
| Ueba | Japanese | Other Asian | Wong et al. |
| Uechi | Japanese | Other Asian | Wong et al. |
| Ueda | Japanese | Other Asian | Wong et al. |
| Uehara | Japanese | Other Asian | Wong et al. |
| Ueki | Japanese | Other Asian | Wong et al. |
| Uema | Japanese | Other Asian | Wong et al. |
| Uematsu | Japanese | Other Asian | Wong et al. |
| Uemoto | Japanese | Other Asian | Wong et al. |
| Uemura | Japanese | Other Asian | Wong et al. |
| Ueng | Chinese | Chinese | Wong et al. |
| Uenishi | Japanese | Other Asian | Wong et al. |
| Ueno | Japanese | Other Asian | Wong et al. |
| Uenoyama | Japanese | Other Asian | Wong et al. |
| Ueoka | Japanese | Other Asian | Wong et al. |
| Ueshima | Japanese | Other Asian | Wong et al. |
| Uesugi | Japanese | Other Asian | Wong et al. |
| Ueta | Japanese | Other Asian | Wong et al. |
| Uetake | Japanese | Other Asian | Wong et al. |
| Ueyama | Japanese | Other Asian | Wong et al. |
| Uezato | Japanese | Other Asian | Wong et al. |
| Uezu | Japanese | Other Asian | Wong et al. |
| Uga | Japanese | Other Asian | Wong et al. |
| Ugaban | Filipino | Other Asian | Wong et al. |
| Ugai | Japanese | Other Asian | Wong et al. |
| Ugajin | Japanese | Other Asian | Wong et al. |
| Ugalde | Filipino | Other Asian | Wong et al. |
| Ugale | Filipino | Other Asian | Wong et al. |
| Ugalino | Filipino | Other Asian | Wong et al. |
| Ugarte | Filipino | Other Asian | Wong et al. |
| Ugay | Filipino | Other Asian | Wong et al. |
| Ugerio | Filipino | Other Asian | Wong et al. |
| Ugot | Filipino | Other Asian | Wong et al. |
| Ugto | Filipino | Other Asian | Wong et al. |
| Uh | Korean | Other Asian | Wong et al. |
| Uhm | Korean | Other Asian | Wong et al. |
| Uichanco | Filipino | Other Asian | Wong et al. |
| Ujano | Filipino | Other Asian | Wong et al. |
| Uji | Japanese | Other Asian | Wong et al. |
| Ujihara | Japanese | Other Asian | Wong et al. |
| Ujiie | Japanese | Other Asian | Wong et al. |
| Ujita | Japanese | Other Asian | Wong et al. |
| Ujjol |  | South Asian | Lim et al. |
| Ukai | Japanese | Other Asian | Wong et al. |
| Ukani | Asian Indian | South Asian | Wong et al. |
| Ukawa | Japanese | Other Asian | Wong et al. |
| Ukita | Japanese | Other Asian | Wong et al. |
| Ulahannan | Asian Indian | South Asian | Wong et al. |
| Ulanday | Filipino | Other Asian | Wong et al. |
| Ulang | Filipino | Other Asian | Wong et al. |
| Ulangca | Filipino | Other Asian | Wong et al. |
| Ulat | Filipino | Other Asian | Wong et al. |
| Ulatan | Filipino | Other Asian | Wong et al. |
| Ulep | Filipino | Other Asian | Wong et al. |
| Ulgado | Filipino | Other Asian | Wong et al. |
| Ulhaq | Asian Indian | South Asian | Wong et al. |
| Ulhasan | Asian Indian | South Asian | Wong et al. |
| Ulibas | Filipino | Other Asian | Wong et al. |
| Uligan | Filipino | Other Asian | Kranick et al. |
| Ulit | Filipino | Other Asian | Wong et al. |
| Ulla |  | South Asian | Lim et al. |
| Ullah | Pakistani; Bangladeshi | South Asian | Kranick et al. |
| Ullal | Asian Indian | South Asian | Wong et al. |
| Ulnagan | Filipino | Other Asian | Wong et al. |
| Ulpindo | Filipino | Other Asian | Wong et al. |
| Um | Korean | Other Asian | Kranick et al. |
| Umadhay | Filipino | Other Asian | Wong et al. |
| Umagat | Filipino | Other Asian | Wong et al. |
| Umaguing | Filipino | Other Asian | Wong et al. |
| Umale | Filipino | Other Asian | Wong et al. |
| Umali | Filipino | Other Asian | Wong et al. |
| Umandal | Filipino | Other Asian | Wong et al. |
| Umandap | Filipino | Other Asian | Wong et al. |
| Umano | Japanese | Other Asian | Wong et al. |
| Umanos | Filipino | Other Asian | Wong et al. |
| Umar | Asian Indian | South Asian | Wong et al. |
| Umayam | Filipino | Other Asian | Wong et al. |
| Umbay | Filipino | Other Asian | Wong et al. |
| Umblas | Filipino | Other Asian | Wong et al. |
| Umeda | Japanese | Other Asian | Wong et al. |
| Umedbhai | Asian Indian | South Asian | Wong et al. |
| Umehara | Japanese | Other Asian | Wong et al. |
| Umekawa | Japanese | Other Asian | Wong et al. |
| Umeki | Japanese | Other Asian | Wong et al. |
| Umel | Filipino | Other Asian | Wong et al. |
| Umemoto | Japanese | Other Asian | Wong et al. |
| Umemura | Japanese | Other Asian | Wong et al. |
| Umengan | Filipino | Other Asian | Wong et al. |
| Umeno | Japanese | Other Asian | Wong et al. |
| Umer | Asian Indian | South Asian | Wong et al. |
| Umetani | Japanese | Other Asian | Wong et al. |
| Umetsu | Japanese | Other Asian | Wong et al. |
| Umeyama | Japanese | Other Asian | Wong et al. |
| Umezaki | Japanese | Other Asian | Wong et al. |
| Umezawa | Japanese | Other Asian | Wong et al. |
| Umezu | Japanese | Other Asian | Wong et al. |
| Umil | Filipino | Other Asian | Wong et al. |
| Umila | Filipino | Other Asian | Wong et al. |
| Uminga | Filipino | Other Asian | Wong et al. |
| Umino | Japanese | Other Asian | Wong et al. |
| Umipig | Filipino | Other Asian | Wong et al. |
| Umran |  | South Asian | Lim et al. |
| Umrigar | Asian Indian | South Asian | Wong et al. |
| Unabia | Filipino | Other Asian | Wong et al. |
| Unarce | Filipino | Other Asian | Wong et al. |
| Unay | Filipino | Other Asian | Wong et al. |
| Unciano | Filipino | Other Asian | Wong et al. |
| Undag | Filipino | Other Asian | Wong et al. |
| Une | Japanese | Other Asian | Wong et al. |
| Ungab | Filipino | Other Asian | Wong et al. |
| Ungco | Filipino | Other Asian | Wong et al. |
| Ungos | Filipino | Other Asian | Wong et al. |
| Ungria | Filipino | Other Asian | Wong et al. |
| Ungson | Filipino | Other Asian | Wong et al. |
| Uniambrol |  | South Asian | Lim et al. |
| Unico | Filipino | Other Asian | Wong et al. |
| Unite | Filipino | Other Asian | Wong et al. |
| Unknown |  | South Asian | Lim et al. |
| Unkwn | Chinese | Chinese | Wong et al. |
| Unni | Asian Indian | South Asian | Wong et al. |
| Unnithan | Asian Indian | South Asian | Wong et al. |
| Unno | Japanese | Other Asian | Wong et al. |
| Uno | Japanese | Other Asian | Wong et al. |
| Unoki | Japanese | Other Asian | Wong et al. |
| Unson | Filipino | Other Asian | Kranick et al. |
| Untal | Filipino | Other Asian | Wong et al. |
| Untalan | Filipino | Other Asian | Wong et al. |
| Unwala | Asian Indian | South Asian | Wong et al. |
| Unwalla | Asian Indian | South Asian | Wong et al. |
| Uong | Vietnamese | Other Asian | Wong et al. |
| Uozumi | Japanese | Other Asian | Wong et al. |
| Up | Korean | Other Asian | Wong et al. |
| Upadhyay | Asian Indian | South Asian | Wong et al. |
| Upadhyaya | Asian Indian; Nepali | South Asian | Kranick et al. |
| Uppal | Asian Indian | South Asian | Wong et al. |
| Ura | Japanese | Other Asian | Wong et al. |
| Urabe | Japanese | Other Asian | Wong et al. |
| Uraizee | Asian Indian | South Asian | Wong et al. |
| Urakami | Japanese | Other Asian | Wong et al. |
| Urakawa | Japanese | Other Asian | Wong et al. |
| Urano | Japanese | Other Asian | Wong et al. |
| Uranza | Filipino | Other Asian | Wong et al. |
| Urasaki | Japanese | Other Asian | Wong et al. |
| Urata | Japanese | Other Asian | Wong et al. |
| Uratani | Japanese | Other Asian | Wong et al. |
| Urayama | Japanese | Other Asian | Wong et al. |
| Urayanza | Filipino | Other Asian | Wong et al. |
| Urbanes | Filipino | Other Asian | Wong et al. |
| Urbano | Filipino | Other Asian | Wong et al. |
| Urbi | Filipino | Other Asian | Wong et al. |
| Urbien | Filipino | Other Asian | Wong et al. |
| Urbina | Filipino | Other Asian | Wong et al. |
| Urbino | Filipino | Other Asian | Wong et al. |
| Urbiztondo | Filipino | Other Asian | Wong et al. |
| Urcia | Filipino | Other Asian | Wong et al. |
| Urdaneta | Filipino | Other Asian | Wong et al. |
| Ureta | Filipino | Other Asian | Wong et al. |
| Urgel | Filipino | Other Asian | Wong et al. |
| Urgello | Filipino | Other Asian | Wong et al. |
| Urgena | Filipino | Other Asian | Wong et al. |
| Urian | Filipino | Other Asian | Wong et al. |
| Uriarte | Filipino | Other Asian | Wong et al. |
| Urmaza | Filipino | Other Asian | Wong et al. |
| Urmeneta | Filipino | Other Asian | Wong et al. |
| Urmi |  | South Asian | Lim et al. |
| Urquico | Filipino | Other Asian | Wong et al. |
| Urquiola | Filipino | Other Asian | Wong et al. |
| Urrutia | Filipino | Other Asian | Wong et al. |
| Urs | Asian Indian | South Asian | Wong et al. |
| Ursabia | Filipino | Other Asian | Wong et al. |
| Ursal | Filipino | Other Asian | Wong et al. |
| Ursua | Filipino | Other Asian | Wong et al. |
| Urubio | Filipino | Other Asian | Wong et al. |
| Urushibara | Japanese | Other Asian | Wong et al. |
| Urushihara | Japanese | Other Asian | Wong et al. |
| Uryu | Japanese | Other Asian | Wong et al. |
| Usami | Japanese | Other Asian | Wong et al. |
| Usha |  | South Asian | Lim et al. |
| Ushida | Japanese | Other Asian | Wong et al. |
| Ushijima | Japanese | Other Asian | Kranick et al. |
| Ushiki | Japanese | Other Asian | Wong et al. |
| Ushikubo | Japanese | Other Asian | Wong et al. |
| Ushio | Japanese | Other Asian | Wong et al. |
| Ushioda | Japanese | Other Asian | Wong et al. |
| Ushiyama | Japanese | Other Asian | Wong et al. |
| Usi | Filipino | Other Asian | Wong et al. |
| Usita | Filipino | Other Asian | Wong et al. |
| Usman |  | South Asian | Lim et al. |
| Usmani | Pakistani | South Asian | Kranick et al. |
| Uson | Filipino | Other Asian | Wong et al. |
| Ustaris | Filipino | Other Asian | Wong et al. |
| Usuda | Japanese | Other Asian | Wong et al. |
| Usui | Japanese | Other Asian | Wong et al. |
| Usuki | Japanese | Other Asian | Wong et al. |
| Ut | Vietnamese | Other Asian | Wong et al. |
| Utaka | Japanese | Other Asian | Wong et al. |
| Uthup | Asian Indian | South Asian | Wong et al. |
| Uthuppan | Asian Indian | South Asian | Wong et al. |
| Uto | Japanese | Other Asian | Wong et al. |
| Utrera | Filipino | Other Asian | Wong et al. |
| Utsugi | Japanese | Other Asian | Wong et al. |
| Utsumi | Japanese | Other Asian | Wong et al. |
| Utsunomiya | Japanese | Other Asian | Wong et al. |
| Utzurrum | Filipino | Other Asian | Wong et al. |
| Uvas | Filipino | Other Asian | Wong et al. |
| Uvero | Filipino | Other Asian | Wong et al. |
| Uy | Filipino | Other Asian | Wong et al. |
| Uyama | Japanese | Other Asian | Wong et al. |
| Uyan | Filipino | Other Asian | Wong et al. |
| Uyas | Asian Indian | South Asian | Wong et al. |
| Uyeda | Japanese | Other Asian | Wong et al. |
| Uyehara | Japanese | Other Asian | Wong et al. |
| Uyeki | Japanese | Other Asian | Wong et al. |
| Uyematsu | Japanese | Other Asian | Wong et al. |
| Uyemoto | Japanese | Other Asian | Wong et al. |
| Uyemura | Japanese | Other Asian | Wong et al. |
| Uyen | Vietnamese | Other Asian | Wong et al. |
| Uyeno | Japanese | Other Asian | Wong et al. |
| Uyeyama | Japanese | Other Asian | Wong et al. |
| Uytengsu | Filipino | Other Asian | Wong et al. |
| Uytingco | Filipino | Other Asian | Wong et al. |
| Uzarraga | Filipino | Other Asian | Wong et al. |
| Uzawa | Japanese | Other Asian | Wong et al. |
| Uzzal |  | South Asian | Lim et al. |
| V | Asian Indian | South Asian | Wong et al. |
| Vaca | Filipino | Other Asian | Wong et al. |
| Vachhani | Asian Indian | South Asian | Wong et al. |
| Vadlamani | Asian Indian | South Asian | Wong et al. |
| Vadlamudi | Asian Indian | South Asian | Wong et al. |
| Vagay | Filipino | Other Asian | Wong et al. |
| Vaghani | Asian Indian | South Asian | Wong et al. |
| Vaghela | Asian Indian | South Asian | Wong et al. |
| Vahora | Asian Indian | South Asian | Wong et al. |
| Vaid | Asian Indian | South Asian | Wong et al. |
| Vaidhyanathan | Asian Indian | South Asian | Kranick et al. |
| Vaidya | Asian Indian | South Asian | Kranick et al. |
| Vaidyanathan | Asian Indian | South Asian | Wong et al. |
| Vailoces | Filipino | Other Asian | Wong et al. |
| Vaish | Asian Indian | South Asian | Wong et al. |
| Vaishampayan | Asian Indian | South Asian | Wong et al. |
| Vaishnav | Asian Indian | South Asian | Wong et al. |
| Vakani | Asian Indian | South Asian | Wong et al. |
| Vakharia | Asian Indian | South Asian | Wong et al. |
| Vakil | Asian Indian | South Asian | Wong et al. |
| Valand | Asian Indian | South Asian | Wong et al. |
| Valbuena | Filipino | Other Asian | Wong et al. |
| Valcarcel | Filipino | Other Asian | Wong et al. |
| Valdecanas | Filipino | Other Asian | Wong et al. |
| Valdecantos | Filipino | Other Asian | Wong et al. |
| Valdefiera | Filipino | Other Asian | Wong et al. |
| Valdehuesa | Filipino | Other Asian | Wong et al. |
| Valdehueza | Filipino | Other Asian | Wong et al. |
| Valdellon | Filipino | Other Asian | Wong et al. |
| Valdemoro | Filipino | Other Asian | Wong et al. |
| Valderama | Filipino | Other Asian | Wong et al. |
| Valderrama | Filipino | Other Asian | Wong et al. |
| Valdes | Filipino | Other Asian | Wong et al. |
| Valdevieso | Filipino | Other Asian | Wong et al. |
| Valdez | Filipino | Other Asian | Wong et al. |
| Valdivia | Filipino | Other Asian | Wong et al. |
| Valdomar | Filipino | Other Asian | Wong et al. |
| Valdoz | Filipino | Other Asian | Wong et al. |
| Valdriz | Filipino | Other Asian | Wong et al. |
| Valecruz | Filipino | Other Asian | Wong et al. |
| Valena | Filipino | Other Asian | Wong et al. |
| Valencerina | Filipino | Other Asian | Wong et al. |
| Valencia | Filipino | Other Asian | Wong et al. |
| Valenciano | Filipino | Other Asian | Wong et al. |
| Valentin | Filipino | Other Asian | Wong et al. |
| Valenton | Filipino | Other Asian | Wong et al. |
| Valentos | Filipino | Other Asian | Wong et al. |
| Valenzona | Filipino | Other Asian | Wong et al. |
| Valenzuela | Filipino | Other Asian | Wong et al. |
| Valera | Filipino | Other Asian | Wong et al. |
| Valeriano | Filipino | Other Asian | Wong et al. |
| Valerio | Filipino | Other Asian | Wong et al. |
| Valero | Filipino | Other Asian | Wong et al. |
| Valeros | Filipino | Other Asian | Wong et al. |
| Valeroso | Filipino | Other Asian | Wong et al. |
| Vales | Filipino | Other Asian | Wong et al. |
| Valete | Filipino | Other Asian | Wong et al. |
| Valeza | Filipino | Other Asian | Wong et al. |
| Vali | Asian Indian | South Asian | Wong et al. |
| Valido | Filipino | Other Asian | Wong et al. |
| Valiente | Filipino | Other Asian | Wong et al. |
| Valin | Filipino | Other Asian | Wong et al. |
| Valino | Filipino | Other Asian | Wong et al. |
| Valisno | Filipino | Other Asian | Wong et al. |
| Valite | Filipino | Other Asian | Wong et al. |
| Valiti | Filipino | Other Asian | Wong et al. |
| Valji | Asian Indian | South Asian | Wong et al. |
| Vallabh | Asian Indian | South Asian | Wong et al. |
| Vallabhbhai | Asian Indian | South Asian | Wong et al. |
| Vallada | Filipino | Other Asian | Wong et al. |
| Valladares | Asian Indian | South Asian | Wong et al. |
| Valladolid | Filipino | Other Asian | Wong et al. |
| Vallar | Filipino | Other Asian | Wong et al. |
| Vallarta | Filipino | Other Asian | Kranick et al. |
| Valle | Filipino | Other Asian | Wong et al. |
| Valledor | Filipino | Other Asian | Wong et al. |
| Vallejera | Filipino | Other Asian | Wong et al. |
| Vallejo | Filipino | Other Asian | Wong et al. |
| Vallejos | Filipino | Other Asian | Wong et al. |
| Vallente | Filipino | Other Asian | Wong et al. |
| Valles | Filipino | Other Asian | Kranick et al. |
| Valleser | Filipino | Other Asian | Wong et al. |
| Vallespin | Filipino | Other Asian | Wong et al. |
| Vallestero | Filipino | Other Asian | Wong et al. |
| Vallesterol | Filipino | Other Asian | Wong et al. |
| Vallesteros | Filipino | Other Asian | Wong et al. |
| Vallido | Filipino | Other Asian | Wong et al. |
| Vallikolil | Asian Indian | South Asian | Kranick et al. |
| Vallo | Filipino | Other Asian | Wong et al. |
| Valluri | Asian Indian | South Asian | Wong et al. |
| Valmoja | Filipino | Other Asian | Wong et al. |
| Valmonte | Filipino | Other Asian | Wong et al. |
| Valmores | Filipino | Other Asian | Wong et al. |
| Valte | Filipino | Other Asian | Wong et al. |
| Van | Vietnamese | Other Asian | Wong et al. |
| Vanderlipe | Filipino | Other Asian | Wong et al. |
| Vangala | Asian Indian | South Asian | Wong et al. |
| Vanguardia | Filipino | Other Asian | Wong et al. |
| Vanhaeften | Asian Indian | South Asian | Wong et al. |
| Vannguyen | Vietnamese | Other Asian | Wong et al. |
| Vanta | Filipino | Other Asian | Wong et al. |
| Vanu |  | South Asian | Lim et al. |
| Vanzuela | Filipino | Other Asian | Wong et al. |
| Vapor | Filipino | Other Asian | Wong et al. |
| Vaquilar | Filipino | Other Asian | Wong et al. |
| Varadachari | Asian Indian | South Asian | Wong et al. |
| Varadarajan | Asian Indian | South Asian | Wong et al. |
| Varanasi | Asian Indian | South Asian | Wong et al. |
| Varde | Asian Indian | South Asian | Wong et al. |
| Vardeleon | Filipino | Other Asian | Wong et al. |
| Vareed | Asian Indian | South Asian | Wong et al. |
| Varela | Filipino | Other Asian | Wong et al. |
| Vargas | Filipino | Other Asian | Wong et al. |
| Varghese | Asian Indian | South Asian | Kranick et al. |
| Varias | Filipino | Other Asian | Wong et al. |
| Varilla | Filipino | Other Asian | Wong et al. |
| Varkey | Asian Indian | South Asian | Wong et al. |
| Varma | Asian Indian | South Asian | Wong et al. |
| Varona | Filipino | Other Asian | Wong et al. |
| Varquez | Filipino | Other Asian | Wong et al. |
| Varshney | Asian Indian | South Asian | Wong et al. |
| Varughese | Asian Indian | South Asian | Kranick et al. |
| Vasa | Asian Indian | South Asian | Wong et al. |
| Vasallo | Filipino | Other Asian | Wong et al. |
| Vasan | Asian Indian | South Asian | Kranick et al. |
| Vasani | Asian Indian | South Asian | Wong et al. |
| Vasavada | Asian Indian | South Asian | Wong et al. |
| Vasco | Filipino | Other Asian | Wong et al. |
| Vashi | Asian Indian | South Asian | Wong et al. |
| Vashisht | Asian Indian | South Asian | Wong et al. |
| Vasques | Filipino | Other Asian | Wong et al. |
| Vasquez | Filipino | Other Asian | Wong et al. |
| Vasudev | Asian Indian | South Asian | Wong et al. |
| Vasudeva | Asian Indian | South Asian | Wong et al. |
| Vasudevan | Asian Indian | South Asian | Wong et al. |
| Vaswani | Pakistani; Asian Indian | South Asian | Kranick et al. |
| Vats | Asian Indian | South Asian | Wong et al. |
| Vay | Vietnamese | Other Asian | Wong et al. |
| Vaz | Asian Indian | South Asian | Wong et al. |
| Vaze | Asian Indian | South Asian | Wong et al. |
| Vazir | Asian Indian | South Asian | Wong et al. |
| Vazquez | Filipino | Other Asian | Wong et al. |
| Ve | Vietnamese | Other Asian | Wong et al. |
| Vea | Filipino | Other Asian | Wong et al. |
| Vechi | Japanese | Other Asian | Wong et al. |
| Vecta | Filipino | Other Asian | Wong et al. |
| Ved | Asian Indian | South Asian | Wong et al. |
| Vedantham | Asian Indian | South Asian | Wong et al. |
| Vedar | Filipino | Other Asian | Wong et al. |
| Vedula | Asian Indian | South Asian | Wong et al. |
| Vee | Chinese | Chinese | Wong et al. |
| Veerappan | Asian Indian | South Asian | Wong et al. |
| Veeraraghava | Asian Indian | South Asian | Wong et al. |
| Veerjee | Asian Indian | South Asian | Wong et al. |
| Vega | Filipino | Other Asian | Wong et al. |
| Vehara | Japanese | Other Asian | Wong et al. |
| Vekaria | Asian Indian | South Asian | Wong et al. |
| Veki | Japanese | Other Asian | Wong et al. |
| Vela | Filipino | Other Asian | Wong et al. |
| Velada | Filipino | Other Asian | Wong et al. |
| Velandria | Filipino | Other Asian | Wong et al. |
| Velani | Asian Indian | South Asian | Wong et al. |
| Velano | Filipino | Other Asian | Wong et al. |
| Velante | Filipino | Other Asian | Wong et al. |
| Velarde | Filipino | Other Asian | Kranick et al. |
| Velasco | Filipino | Other Asian | Wong et al. |
| Velasquez | Filipino | Other Asian | Wong et al. |
| Velayo | Filipino | Other Asian | Wong et al. |
| Velayudhan | Asian Indian | South Asian | Wong et al. |
| Velazco | Filipino | Other Asian | Wong et al. |
| Velbis | Filipino | Other Asian | Wong et al. |
| Velchez | Filipino | Other Asian | Wong et al. |
| Velena | Filipino | Other Asian | Wong et al. |
| Velez | Filipino | Other Asian | Wong et al. |
| Velicaria | Filipino | Other Asian | Wong et al. |
| Velilla | Filipino | Other Asian | Wong et al. |
| Velji | Asian Indian | South Asian | Wong et al. |
| Vellanki | Asian Indian | South Asian | Wong et al. |
| Velo | Filipino | Other Asian | Wong et al. |
| Veloira | Filipino | Other Asian | Wong et al. |
| Velonza | Filipino | Other Asian | Wong et al. |
| Veloria | Filipino | Other Asian | Wong et al. |
| Veloro | Filipino | Other Asian | Wong et al. |
| Veloso | Filipino | Other Asian | Wong et al. |
| Velovic |  | South Asian | Lim et al. |
| Veloz | Filipino | Other Asian | Wong et al. |
| Veluya | Filipino | Other Asian | Wong et al. |
| Veluz | Filipino | Other Asian | Wong et al. |
| Vematsu | Japanese | Other Asian | Wong et al. |
| Vemulapalli | Asian Indian | South Asian | Wong et al. |
| Vemura | Japanese | Other Asian | Wong et al. |
| Vemuri | Asian Indian | South Asian | Wong et al. |
| Vencer | Filipino | Other Asian | Wong et al. |
| Vendiola | Filipino | Other Asian | Wong et al. |
| Venegas | Filipino | Other Asian | Wong et al. |
| Venenciano | Filipino | Other Asian | Wong et al. |
| Veneracion | Filipino | Other Asian | Wong et al. |
| Venezuela | Filipino | Other Asian | Wong et al. |
| Vengco | Filipino | Other Asian | Wong et al. |
| Venida | Filipino | Other Asian | Wong et al. |
| Veniegas | Filipino | Other Asian | Wong et al. |
| Venkata | Asian Indian | South Asian | Wong et al. |
| Venkatachala | Asian Indian | South Asian | Wong et al. |
| Venkatachar | Asian Indian | South Asian | Wong et al. |
| Venkatachari | Asian Indian | South Asian | Wong et al. |
| Venkatakrish | Asian Indian | South Asian | Wong et al. |
| Venkatappa | Asian Indian | South Asian | Wong et al. |
| Venkataramai | Asian Indian | South Asian | Wong et al. |
| Venkataraman | Asian Indian | South Asian | Wong et al. |
| Venkatasubbi | Asian Indian | South Asian | Wong et al. |
| Venkatasubra | Asian Indian | South Asian | Wong et al. |
| Venkatesan | Asian Indian | South Asian | Wong et al. |
| Venkatesh | Asian Indian | South Asian | Wong et al. |
| Venkateswara | Asian Indian | South Asian | Wong et al. |
| Venkatraman | Asian Indian | South Asian | Wong et al. |
| Ventanilla | Filipino | Other Asian | Wong et al. |
| Ventayen | Filipino | Other Asian | Wong et al. |
| Ventenilla | Filipino | Other Asian | Wong et al. |
| Ventigan | Filipino | Other Asian | Wong et al. |
| Ventinilla | Filipino | Other Asian | Wong et al. |
| Ventosa | Filipino | Other Asian | Wong et al. |
| Ventura | Filipino | Other Asian | Wong et al. |
| Venturanza | Filipino | Other Asian | Wong et al. |
| Venturillo | Filipino | Other Asian | Wong et al. |
| Venturina | Filipino | Other Asian | Kranick et al. |
| Venugopalan | Asian Indian | South Asian | Wong et al. |
| Venzon | Filipino | Other Asian | Wong et al. |
| Venzuela | Filipino | Other Asian | Wong et al. |
| Ver | Filipino | Other Asian | Wong et al. |
| Vera | Filipino | Other Asian | Wong et al. |
| Veracruz | Filipino | Other Asian | Wong et al. |
| Verador | Filipino | Other Asian | Wong et al. |
| Veran | Filipino | Other Asian | Wong et al. |
| Veranga | Filipino | Other Asian | Wong et al. |
| Verano | Filipino | Other Asian | Wong et al. |
| Veras | Filipino | Other Asian | Wong et al. |
| Verayo | Filipino | Other Asian | Wong et al. |
| Verbo | Filipino | Other Asian | Wong et al. |
| Verceles | Filipino | Other Asian | Wong et al. |
| Verdadero | Filipino | Other Asian | Wong et al. |
| Verde | Filipino | Other Asian | Wong et al. |
| Verdeflor | Filipino | Other Asian | Wong et al. |
| Verdejo | Filipino | Other Asian | Wong et al. |
| Verder | Filipino | Other Asian | Wong et al. |
| Verendia | Filipino | Other Asian | Wong et al. |
| Vergara | Filipino | Other Asian | Wong et al. |
| Vergel | Filipino | Other Asian | Wong et al. |
| Vergeldedi | Filipino | Other Asian | Wong et al. |
| Vergeldedio | Filipino | Other Asian | Wong et al. |
| Verghese | Asian Indian | South Asian | Wong et al. |
| Veridiano | Filipino | Other Asian | Wong et al. |
| Verma | Asian Indian | South Asian | Wong et al. |
| Versola | Filipino | Other Asian | Wong et al. |
| Versoza | Filipino | Other Asian | Wong et al. |
| Vertido | Filipino | Other Asian | Wong et al. |
| Vertucio | Filipino | Other Asian | Wong et al. |
| Verzola | Filipino | Other Asian | Wong et al. |
| Verzosa | Filipino | Other Asian | Wong et al. |
| Vesagas | Filipino | Other Asian | Wong et al. |
| Vestil | Filipino | Other Asian | Wong et al. |
| Vesugi | Japanese | Other Asian | Wong et al. |
| Vi | Vietnamese | Other Asian | Wong et al. |
| Viacrucis | Filipino | Other Asian | Wong et al. |
| Viacrusis | Filipino | Other Asian | Wong et al. |
| Viado | Filipino | Other Asian | Wong et al. |
| Viajar | Filipino | Other Asian | Wong et al. |
| Viaje | Filipino | Other Asian | Wong et al. |
| Viana | Filipino | Other Asian | Wong et al. |
| Vianzon | Filipino | Other Asian | Wong et al. |
| Viardo | Filipino | Other Asian | Wong et al. |
| Vibal | Filipino | Other Asian | Wong et al. |
| Vibangco | Filipino | Other Asian | Wong et al. |
| Vibar | Filipino | Other Asian | Wong et al. |
| Vibat | Filipino | Other Asian | Wong et al. |
| Vibora | Filipino | Other Asian | Wong et al. |
| Vicedo | Filipino | Other Asian | Wong et al. |
| Vicencio | Filipino | Other Asian | Wong et al. |
| Vicente | Filipino | Other Asian | Wong et al. |
| Vicera | Filipino | Other Asian | Wong et al. |
| Viceral | Filipino | Other Asian | Wong et al. |
| Vicerra | Filipino | Other Asian | Wong et al. |
| Victa | Filipino | Other Asian | Wong et al. |
| Victo | Filipino | Other Asian | Wong et al. |
| Victoria | Filipino | Other Asian | Wong et al. |
| Victoriano | Filipino | Other Asian | Kranick et al. |
| Victorino | Filipino | Other Asian | Kranick et al. |
| Victorio | Filipino | Other Asian | Wong et al. |
| Vicuna | Filipino | Other Asian | Wong et al. |
| Vidad | Filipino | Other Asian | Kranick et al. |
| Vidal | Filipino | Other Asian | Wong et al. |
| Vidallo | Filipino | Other Asian | Wong et al. |
| Vidallon | Filipino | Other Asian | Wong et al. |
| Vidamo | Filipino | Other Asian | Wong et al. |
| Vidanes | Filipino | Other Asian | Wong et al. |
| Videna | Filipino | Other Asian | Wong et al. |
| Vidola | Filipino | Other Asian | Wong et al. |
| Viduya | Filipino | Other Asian | Wong et al. |
| Vidyarthi | Asian Indian | South Asian | Wong et al. |
| Vieiraribei | Chinese | Chinese | Wong et al. |
| Viejo | Filipino | Other Asian | Wong et al. |
| Vien | Vietnamese | Other Asian | Wong et al. |
| Viernes | Filipino | Other Asian | Wong et al. |
| Viet | Vietnamese | Other Asian | Wong et al. |
| Vigilia | Filipino | Other Asian | Wong et al. |
| Vigilla | Filipino | Other Asian | Wong et al. |
| Vigo | Filipino | Other Asian | Wong et al. |
| Vij | Asian Indian | South Asian | Wong et al. |
| Vijay | Asian Indian | South Asian | Wong et al. |
| Vijayaraghav | Asian Indian | South Asian | Wong et al. |
| Vijaykumar |  | South Asian | Lim et al. |
| Vijh | Asian Indian | South Asian | Wong et al. |
| Vila | Filipino | Other Asian | Wong et al. |
| Vilar | Filipino | Other Asian | Wong et al. |
| Vilbar | Filipino | Other Asian | Wong et al. |
| Vilches | Filipino | Other Asian | Wong et al. |
| Vilda | Filipino | Other Asian | Wong et al. |
| Villa | Filipino | Other Asian | Wong et al. |
| Villaber | Filipino | Other Asian | Wong et al. |
| Villablanca | Filipino | Other Asian | Wong et al. |
| Villabroza | Filipino | Other Asian | Wong et al. |
| Villacampa | Filipino | Other Asian | Wong et al. |
| Villacarlos | Filipino | Other Asian | Wong et al. |
| Villaceran | Filipino | Other Asian | Wong et al. |
| Villacorta | Filipino | Other Asian | Wong et al. |
| Villacorte | Filipino | Other Asian | Wong et al. |
| Villacrusis | Filipino | Other Asian | Wong et al. |
| Villacruz | Filipino | Other Asian | Wong et al. |
| Villadelgado | Filipino | Other Asian | Wong et al. |
| Villadolid | Filipino | Other Asian | Wong et al. |
| Villafania | Filipino | Other Asian | Wong et al. |
| Villaflor | Filipino | Other Asian | Wong et al. |
| Villaflores | Filipino | Other Asian | Wong et al. |
| Villafranca | Filipino | Other Asian | Wong et al. |
| Villafuerte | Filipino | Other Asian | Wong et al. |
| Villagonzalo | Filipino | Other Asian | Wong et al. |
| Villagracia | Filipino | Other Asian | Wong et al. |
| Villahermosa | Filipino | Other Asian | Wong et al. |
| Villajin | Filipino | Other Asian | Kranick et al. |
| Villajuan | Filipino | Other Asian | Wong et al. |
| Villalba | Filipino | Other Asian | Wong et al. |
| Villalobos | Filipino | Other Asian | Wong et al. |
| Villalon | Filipino | Other Asian | Wong et al. |
| Villaluna | Filipino | Other Asian | Wong et al. |
| Villaluz | Filipino | Other Asian | Wong et al. |
| Villalva | Filipino | Other Asian | Wong et al. |
| Villamar | Filipino | Other Asian | Wong et al. |
| Villamaria | Filipino | Other Asian | Wong et al. |
| Villamarin | Filipino | Other Asian | Wong et al. |
| Villamater | Filipino | Other Asian | Wong et al. |
| Villamayor | Filipino | Other Asian | Wong et al. |
| Villamil | Filipino | Other Asian | Wong et al. |
| Villamin | Filipino | Other Asian | Wong et al. |
| Villamonte | Filipino | Other Asian | Wong et al. |
| Villamor | Filipino | Other Asian | Wong et al. |
| Villamora | Filipino | Other Asian | Wong et al. |
| Villan | Filipino | Other Asian | Wong et al. |
| Villaneuva | Filipino | Other Asian | Wong et al. |
| Villania | Filipino | Other Asian | Wong et al. |
| Villanos | Filipino | Other Asian | Wong et al. |
| Villanosa | Filipino | Other Asian | Wong et al. |
| Villanueva | Filipino | Other Asian | Kranick et al. |
| Villanveva | Filipino | Other Asian | Wong et al. |
| Villao | Filipino | Other Asian | Wong et al. |
| Villapa | Filipino | Other Asian | Wong et al. |
| Villapana | Filipino | Other Asian | Wong et al. |
| Villapando | Filipino | Other Asian | Wong et al. |
| Villar | Filipino | Other Asian | Wong et al. |
| Villarama | Filipino | Other Asian | Wong et al. |
| Villaran | Filipino | Other Asian | Wong et al. |
| Villaranda | Filipino | Other Asian | Wong et al. |
| Villarante | Filipino | Other Asian | Wong et al. |
| Villaraza | Filipino | Other Asian | Wong et al. |
| Villarba | Filipino | Other Asian | Wong et al. |
| Villareal | Filipino | Other Asian | Wong et al. |
| Villarente | Filipino | Other Asian | Wong et al. |
| Villarete | Filipino | Other Asian | Wong et al. |
| Villarias | Filipino | Other Asian | Wong et al. |
| Villarica | Filipino | Other Asian | Wong et al. |
| Villarico | Filipino | Other Asian | Wong et al. |
| Villarin | Filipino | Other Asian | Wong et al. |
| Villarino | Filipino | Other Asian | Wong et al. |
| Villarivera | Filipino | Other Asian | Wong et al. |
| Villarma | Filipino | Other Asian | Wong et al. |
| Villarmia | Filipino | Other Asian | Wong et al. |
| Villarmino | Filipino | Other Asian | Wong et al. |
| Villaro | Filipino | Other Asian | Wong et al. |
| Villaroman | Filipino | Other Asian | Wong et al. |
| Villaros | Filipino | Other Asian | Wong et al. |
| Villarosa | Filipino | Other Asian | Wong et al. |
| Villaroz | Filipino | Other Asian | Wong et al. |
| Villarreal | Filipino | Other Asian | Wong et al. |
| Villarroel | Filipino | Other Asian | Wong et al. |
| Villarruz | Filipino | Other Asian | Wong et al. |
| Villarta | Filipino | Other Asian | Wong et al. |
| Villaruel | Filipino | Other Asian | Wong et al. |
| Villaruz | Filipino | Other Asian | Wong et al. |
| Villas | Filipino | Other Asian | Wong et al. |
| Villasan | Filipino | Other Asian | Wong et al. |
| Villasenor | Filipino | Other Asian | Wong et al. |
| Villasin | Filipino | Other Asian | Wong et al. |
| Villasis | Filipino | Other Asian | Wong et al. |
| Villasista | Filipino | Other Asian | Wong et al. |
| Villasor | Filipino | Other Asian | Wong et al. |
| Villastiqui | Filipino | Other Asian | Wong et al. |
| Villate | Filipino | Other Asian | Wong et al. |
| Villatuya | Filipino | Other Asian | Wong et al. |
| Villaueva | Filipino | Other Asian | Wong et al. |
| Villaver | Filipino | Other Asian | Wong et al. |
| Villaverde | Filipino | Other Asian | Wong et al. |
| Villavert | Filipino | Other Asian | Wong et al. |
| Villavicenci | Filipino | Other Asian | Wong et al. |
| Villazor | Filipino | Other Asian | Wong et al. |
| Villegas | Filipino | Other Asian | Wong et al. |
| Villejo | Filipino | Other Asian | Wong et al. |
| Villena | Filipino | Other Asian | Wong et al. |
| Villero | Filipino | Other Asian | Wong et al. |
| Villeza | Filipino | Other Asian | Wong et al. |
| Villezar | Filipino | Other Asian | Wong et al. |
| Villo | Filipino | Other Asian | Wong et al. |
| Villon | Filipino | Other Asian | Wong et al. |
| Villones | Filipino | Other Asian | Wong et al. |
| Villongco | Filipino | Other Asian | Wong et al. |
| Villorente | Filipino | Other Asian | Wong et al. |
| Villoria | Filipino | Other Asian | Wong et al. |
| Villoso | Filipino | Other Asian | Wong et al. |
| Villota | Filipino | Other Asian | Wong et al. |
| Viloria | Filipino | Other Asian | Kranick et al. |
| Viluan | Filipino | Other Asian | Wong et al. |
| Vimawala | Asian Indian | South Asian | Wong et al. |
| Vina | Filipino | Other Asian | Wong et al. |
| Vinalon | Filipino | Other Asian | Wong et al. |
| Vinarao | Filipino | Other Asian | Wong et al. |
| Vinas | Filipino | Other Asian | Wong et al. |
| Vincecruz | Filipino | Other Asian | Wong et al. |
| Vincente | Filipino | Other Asian | Wong et al. |
| Vincoy | Filipino | Other Asian | Wong et al. |
| Vindua | Filipino | Other Asian | Wong et al. |
| Vinegas | Filipino | Other Asian | Wong et al. |
| Ving | Chinese | Chinese | Wong et al. |
| Vinh | Vietnamese | Other Asian | Wong et al. |
| Vinjamuri | Asian Indian | South Asian | Wong et al. |
| Vinluan | Filipino | Other Asian | Wong et al. |
| Vino | Filipino | Other Asian | Wong et al. |
| Vinoray | Filipino | Other Asian | Wong et al. |
| Vinoya | Filipino | Other Asian | Wong et al. |
| Vinteres | Filipino | Other Asian | Wong et al. |
| Vinuya | Filipino | Other Asian | Wong et al. |
| Vinzon | Filipino | Other Asian | Wong et al. |
| Viojan | Filipino | Other Asian | Wong et al. |
| Violago | Filipino | Other Asian | Wong et al. |
| Violan | Filipino | Other Asian | Wong et al. |
| Violanda | Filipino | Other Asian | Wong et al. |
| Violango | Filipino | Other Asian | Wong et al. |
| Violeta | Filipino | Other Asian | Wong et al. |
| Virani | Asian Indian | South Asian | Wong et al. |
| Virata | Filipino | Other Asian | Wong et al. |
| Viray | Filipino | Other Asian | Wong et al. |
| Virdi | Asian Indian | South Asian | Kranick et al. |
| Viri | Filipino | Other Asian | Wong et al. |
| Viril | Filipino | Other Asian | Wong et al. |
| Virini | Filipino | Other Asian | Kranick et al. |
| Virjee | Asian Indian | South Asian | Wong et al. |
| Virji | Asian Indian | South Asian | Wong et al. |
| Virkar | Asian Indian | South Asian | Wong et al. |
| Virola | Filipino | Other Asian | Wong et al. |
| Virrey | Filipino | Other Asian | Wong et al. |
| Virtucio | Filipino | Other Asian | Wong et al. |
| Virtudazo | Filipino | Other Asian | Wong et al. |
| Virtusio | Filipino | Other Asian | Wong et al. |
| Visaya | Filipino | Other Asian | Wong et al. |
| Visda | Filipino | Other Asian | Wong et al. |
| Vishram | Asian Indian | South Asian | Wong et al. |
| Visitacion | Filipino | Other Asian | Wong et al. |
| Vismanos | Filipino | Other Asian | Wong et al. |
| Vismonte | Filipino | Other Asian | Wong et al. |
| Vison | Filipino | Other Asian | Wong et al. |
| Visoria | Filipino | Other Asian | Wong et al. |
| Visperas | Filipino | Other Asian | Wong et al. |
| Vispo | Filipino | Other Asian | Wong et al. |
| Vista | Filipino | Other Asian | Wong et al. |
| Vistan | Filipino | Other Asian | Wong et al. |
| Viste | Filipino | Other Asian | Wong et al. |
| Vistro | Filipino | Other Asian | Wong et al. |
| Visvanathan | Asian Indian | South Asian | Wong et al. |
| Viswanath | Asian Indian | South Asian | Kranick et al. |
| Viswanatha | Asian Indian | South Asian | Wong et al. |
| Viswanathan | Asian Indian | South Asian | Wong et al. |
| Vitales | Filipino | Other Asian | Wong et al. |
| Vitalis | Filipino | Other Asian | Wong et al. |
| Vitan | Filipino | Other Asian | Wong et al. |
| Vitangcol | Filipino | Other Asian | Wong et al. |
| Vite | Filipino | Other Asian | Wong et al. |
| Vitente | Filipino | Other Asian | Wong et al. |
| Viterbo | Filipino | Other Asian | Wong et al. |
| Vithalbhai | Asian Indian | South Asian | Wong et al. |
| Vithayathil | Asian Indian | South Asian | Wong et al. |
| Vitin | Filipino | Other Asian | Wong et al. |
| Vito | Filipino | Other Asian | Wong et al. |
| Vitocruz | Filipino | Other Asian | Wong et al. |
| Vitor | Filipino | Other Asian | Wong et al. |
| Vittal | Asian Indian | South Asian | Wong et al. |
| Vitto | Filipino | Other Asian | Wong et al. |
| Vitug | Filipino | Other Asian | Wong et al. |
| Viudez | Filipino | Other Asian | Wong et al. |
| Vivar | Filipino | Other Asian | Wong et al. |
| Vivas | Filipino | Other Asian | Wong et al. |
| Vivero | Filipino | Other Asian | Wong et al. |
| Vives | Filipino | Other Asian | Wong et al. |
| Vivit | Filipino | Other Asian | Wong et al. |
| Vivo | Filipino | Other Asian | Wong et al. |
| Vizcarra | Filipino | Other Asian | Wong et al. |
| Vizconde | Filipino | Other Asian | Wong et al. |
| Vizmanos | Filipino | Other Asian | Wong et al. |
| Vizon | Filipino | Other Asian | Wong et al. |
| Vo | Vietnamese | Other Asian | Kranick et al. |
| Vocal | Filipino | Other Asian | Wong et al. |
| Vohra | Pakistani | South Asian | Kranick et al. |
| Voluntad | Filipino | Other Asian | Wong et al. |
| Vong | Vietnamese | Other Asian | Wong et al. |
| Vongiese | Filipino | Other Asian | Wong et al. |
| Voo | Chinese | Chinese | Wong et al. |
| Voong | Vietnamese | Other Asian | Wong et al. |
| Vora | Asian Indian | South Asian | Kranick et al. |
| Voung | Vietnamese | Other Asian | Wong et al. |
| Vu | Vietnamese | Other Asian | Wong et al. |
| Vung | Chinese | Chinese | Wong et al. |
| Vuong | Vietnamese | Other Asian | Wong et al. |
| Vuu | Vietnamese | Other Asian | Wong et al. |
| Vy | Vietnamese | Other Asian | Wong et al. |
| Vyas | Asian Indian | South Asian | Wong et al. |
| W | Chinese | Chinese | Wong et al. |
| Wa | Chinese | Chinese | Wong et al. |
| Wabe | Filipino | Other Asian | Wong et al. |
| Wachi | Japanese | Other Asian | Wong et al. |
| Wada | Japanese | Other Asian | Kranick et al. |
| Wadhera | Asian Indian | South Asian | Wong et al. |
| Wadia | Asian Indian | South Asian | Wong et al. |
| Wadiwala | Asian Indian | South Asian | Wong et al. |
| Wadiwalla | Asian Indian | South Asian | Wong et al. |
| Wagan | Filipino | Other Asian | Wong et al. |
| Wagas | Filipino | Other Asian | Wong et al. |
| Wagatsuma | Japanese | Other Asian | Wong et al. |
| Wagh | Asian Indian | South Asian | Wong et al. |
| Waghela | Asian Indian | South Asian | Wong et al. |
| Wah | Chinese | Chinese | Wong et al. |
| Wahab | Asian Indian | South Asian | Wong et al. |
| Wahed |  | South Asian | Lim et al. |
| Waheed | Asian Indian | South Asian | Wong et al. |
| Waheed | Pakistani | South Asian | Kranick et al. |
| Waheeduddin | Asian Indian | South Asian | Wong et al. |
| Wahi | Asian Indian | South Asian | Wong et al. |
| Wahid |  | South Asian | Lim et al. |
| Wai | Chinese | Chinese | Wong et al. |
| Waje | Filipino | Other Asian | Wong et al. |
| Wajid | Asian Indian | South Asian | Wong et al. |
| Wajima | Japanese | Other Asian | Wong et al. |
| Wakabayashi | Japanese | Other Asian | Wong et al. |
| Wakahara | Japanese | Other Asian | Wong et al. |
| Wakai | Japanese | Other Asian | Wong et al. |
| Wakamatsu | Japanese | Other Asian | Wong et al. |
| Wakamiya | Japanese | Other Asian | Wong et al. |
| Wakamoto | Japanese | Other Asian | Wong et al. |
| Wakamura | Japanese | Other Asian | Wong et al. |
| Wakao | Japanese | Other Asian | Wong et al. |
| Wakasa | Japanese | Other Asian | Wong et al. |
| Wakasugi | Japanese | Other Asian | Wong et al. |
| Wakatsuki | Japanese | Other Asian | Wong et al. |
| Wakayama | Japanese | Other Asian | Wong et al. |
| Waki | Japanese | Other Asian | Wong et al. |
| Wakida | Japanese | Other Asian | Wong et al. |
| Wakimoto | Japanese | Other Asian | Wong et al. |
| Wakisaka | Japanese | Other Asian | Wong et al. |
| Wakita | Japanese | Other Asian | Wong et al. |
| Wakiya | Japanese | Other Asian | Wong et al. |
| Wakiyama | Japanese | Other Asian | Wong et al. |
| Wako | Japanese | Other Asian | Wong et al. |
| Wakui | Japanese | Other Asian | Wong et al. |
| Walavalkar | Asian Indian | South Asian | Wong et al. |
| Wali | Asian Indian | South Asian | Wong et al. |
| Walia | Asian Indian | South Asian | Wong et al. |
| Waliullaha |  | South Asian | Lim et al. |
| Walji | Asian Indian | South Asian | Wong et al. |
| Wan | Chinese | Chinese | Kranick et al. |
| Wanchoo | Asian Indian | South Asian | Wong et al. |
| Wane | Chinese | Chinese | Wong et al. |
| Wang | Chinese | Chinese | Kranick et al. |
| Wani | Asian Indian | South Asian | Wong et al. |
| War | Chinese | Chinese | Wong et al. |
| Warrier | Asian Indian | South Asian | Wong et al. |
| Warsi | Asian Indian | South Asian | Wong et al. |
| Wasan | Filipino | Other Asian | Wong et al. |
| Waseem |  | South Asian | Lim et al. |
| Waseque |  | South Asian | Lim et al. |
| Washino | Japanese | Other Asian | Wong et al. |
| Washio | Japanese | Other Asian | Wong et al. |
| Washizu | Japanese | Other Asian | Wong et al. |
| Wat | Chinese | Chinese | Wong et al. |
| Watabe | Japanese | Other Asian | Wong et al. |
| Watai | Japanese | Other Asian | Wong et al. |
| Watanaba | Japanese | Other Asian | Wong et al. |
| Watanabe | Japanese | Other Asian | Kranick et al. |
| Watanbe | Japanese | Other Asian | Wong et al. |
| Watanuki | Japanese | Other Asian | Wong et al. |
| Watarai | Japanese | Other Asian | Wong et al. |
| Watari | Japanese | Other Asian | Wong et al. |
| Watase | Japanese | Other Asian | Wong et al. |
| Wataya | Japanese | Other Asian | Wong et al. |
| Waung | Chinese | Chinese | Wong et al. |
| Wazed |  | South Asian | Lim et al. |
| Wazir | Asian Indian | South Asian | Wong et al. |
| We | Chinese | Chinese | Wong et al. |
| Wee | Chinese | Chinese | Wong et al. |
| Weerawardena |  | South Asian | Lim et al. |
| Wei | Chinese | Chinese | Kranick et al. |
| Wen | Chinese | Chinese | Wong et al. |
| Wenceslao | Filipino | Other Asian | Wong et al. |
| Weng | Chinese | Chinese | Kranick et al. |
| Wha | Korean | Other Asian | Wong et al. |
| Whang | Korean | Other Asian | Wong et al. |
| Who | Chinese | Chinese | Wong et al. |
| Whon | Korean | Other Asian | Wong et al. |
| Why | Chinese | Chinese | Wong et al. |
| Wi | Korean | Other Asian | Wong et al. |
| Wico | Filipino | Other Asian | Wong et al. |
| Wijangco | Filipino | Other Asian | Wong et al. |
| Wijesekera | Sri Lankan | South Asian | Kranick et al. |
| Wijesinghe | Sri Lankan | South Asian | Kranick et al. |
| Wijesooriya |  | South Asian | Lim et al. |
| wijeyesinghe | Sri Lankan | South Asian | Kranick et al. |
| Wilwayco | Filipino | Other Asian | Wong et al. |
| Win | Burmese | Other Asian | Kranick et al. |
| Wing | Chinese | Chinese | Wong et al. |
| Wisco | Filipino | Other Asian | Wong et al. |
| Wn | Chinese | Chinese | Wong et al. |
| Wng | Chinese | Chinese | Wong et al. |
| Wo | Chinese | Chinese | Wong et al. |
| Woey | Chinese | Chinese | Wong et al. |
| Woh | Chinese | Chinese | Wong et al. |
| Won | Korean; Chinese | Other Asian | Kranick et al. |
| Wone | Chinese | Chinese | Wong et al. |
| Wong | Chinese | Chinese | Kranick et al. |
| Woo | Chinese | Chinese | Kranick et al. |
| Wooh | Korean | Other Asian | Wong et al. |
| Woon | Chinese | Chinese | Wong et al. |
| Wor | Chinese | Chinese | Wong et al. |
| Wot | Chinese | Chinese | Wong et al. |
| Wou | Chinese | Chinese | Wong et al. |
| Wu | Chinese | Chinese | Kranick et al. |
| Wue | Chinese | Chinese | Wong et al. |
| Wui | Chinese | Chinese | Wong et al. |
| Wun | Chinese | Chinese | Wong et al. |
| Wung | Chinese | Chinese | Wong et al. |
| Wuu | Chinese | Chinese | Wong et al. |
| Wy | Chinese | Chinese | Wong et al. |
| Wycoco | Filipino | Other Asian | Kranick et al. |
| Xam | Vietnamese | Other Asian | Wong et al. |
| Xavier | Chinese | Chinese | Wong et al. |
| Xe | Vietnamese | Other Asian | Wong et al. |
| Xi | Chinese | Chinese | Wong et al. |
| Xia | Chinese | Chinese | Wong et al. |
| Xian | Chinese | Chinese | Wong et al. |
| Xiang | Chinese | Chinese | Wong et al. |
| Xiao | Chinese | Chinese | Wong et al. |
| Xie | Chinese | Chinese | Kranick et al. |
| Xieu | Vietnamese | Other Asian | Wong et al. |
| Xin | Chinese | Chinese | Wong et al. |
| Xing | Chinese | Chinese | Wong et al. |
| Xinh | Vietnamese | Other Asian | Wong et al. |
| Xiu | Chinese | Chinese | Wong et al. |
| Xu | Chinese | Chinese | Kranick et al. |
| Xua | Vietnamese | Other Asian | Wong et al. |
| Xuan | Vietnamese | Other Asian | Wong et al. |
| Xue | Chinese | Chinese | Kranick et al. |
| Xun | Chinese | Chinese | Wong et al. |
| Xung | Vietnamese | Other Asian | Wong et al. |
| Xuong | Vietnamese | Other Asian | Wong et al. |
| Xuyen | Vietnamese | Other Asian | Wong et al. |
| Y | Vietnamese | Other Asian | Wong et al. |
| Yaba | Filipino | Other Asian | Wong et al. |
| Yabe | Japanese | Other Asian | Wong et al. |
| Yabes | Filipino | Other Asian | Wong et al. |
| Yabiku | Japanese | Other Asian | Wong et al. |
| Yabot | Filipino | Other Asian | Wong et al. |
| Yabu | Japanese | Other Asian | Wong et al. |
| Yabuki | Japanese | Other Asian | Wong et al. |
| Yabumoto | Japanese | Other Asian | Wong et al. |
| Yabuno | Japanese | Other Asian | Wong et al. |
| Yabusaki | Japanese | Other Asian | Wong et al. |
| Yabut | Filipino | Other Asian | Wong et al. |
| Yabuta | Japanese | Other Asian | Wong et al. |
| Yabuuchi | Japanese | Other Asian | Wong et al. |
| Yacap | Filipino | Other Asian | Wong et al. |
| Yacapin | Filipino | Other Asian | Wong et al. |
| Yacas | Filipino | Other Asian | Wong et al. |
| Yachi | Japanese | Other Asian | Wong et al. |
| Yacoob | Asian Indian | South Asian | Wong et al. |
| Yada | Japanese | Other Asian | Wong et al. |
| Yadao | Filipino | Other Asian | Wong et al. |
| Yadav | Asian Indian | South Asian | Wong et al. |
| Yadava | Asian Indian | South Asian | Wong et al. |
| Yae | Korean | Other Asian | Wong et al. |
| Yaegashi | Japanese | Other Asian | Wong et al. |
| Yafuso | Japanese | Other Asian | Wong et al. |
| Yagasaki | Japanese | Other Asian | Wong et al. |
| Yagi | Japanese | Other Asian | Wong et al. |
| Yagihashi | Japanese | Other Asian | Wong et al. |
| Yagin | Filipino | Other Asian | Wong et al. |
| Yaginuma | Japanese | Other Asian | Wong et al. |
| Yagishita | Japanese | Other Asian | Wong et al. |
| Yagnik | Asian Indian | South Asian | Wong et al. |
| Yago | Filipino | Other Asian | Wong et al. |
| Yaguchi | Japanese | Other Asian | Wong et al. |
| Yague | Filipino | Other Asian | Wong et al. |
| Yagyagan | Filipino | Other Asian | Wong et al. |
| Yagyu | Japanese | Other Asian | Wong et al. |
| Yah | Chinese | Chinese | Wong et al. |
| Yahagi | Japanese | Other Asian | Wong et al. |
| Yahata | Japanese | Other Asian | Wong et al. |
| Yahiro | Japanese | Other Asian | Wong et al. |
| Yahya | Indonesian; Pakistani | Other Asian | Kranick et al. |
| Yai | Chinese | Chinese | Wong et al. |
| Yajima | Japanese | Other Asian | Wong et al. |
| Yajnik | Asian Indian | South Asian | Wong et al. |
| Yakoob | Asian Indian | South Asian | Wong et al. |
| Yakura | Japanese | Other Asian | Wong et al. |
| Yakuwa | Japanese | Other Asian | Wong et al. |
| Yalamanchili | Asian Indian | South Asian | Wong et al. |
| Yalong | Filipino | Other Asian | Wong et al. |
| Yalung | Filipino | Other Asian | Wong et al. |
| Yam | Asian Indian; Chinese | South Asian | Kranick et al. |
| Yama | Japanese | Other Asian | Wong et al. |
| Yamabe | Japanese | Other Asian | Wong et al. |
| Yamachi | Japanese | Other Asian | Wong et al. |
| Yamada | Japanese | Other Asian | Wong et al. |
| Yamaga | Japanese | Other Asian | Wong et al. |
| Yamagami | Japanese | Other Asian | Wong et al. |
| Yamagata | Japanese | Other Asian | Wong et al. |
| Yamagishi | Japanese | Other Asian | Wong et al. |
| Yamagiwa | Japanese | Other Asian | Wong et al. |
| Yamaguchi | Japanese | Other Asian | Wong et al. |
| Yamahata | Japanese | Other Asian | Wong et al. |
| Yamaji | Japanese | Other Asian | Wong et al. |
| Yamakawa | Japanese | Other Asian | Wong et al. |
| Yamaki | Japanese | Other Asian | Wong et al. |
| Yamakita | Japanese | Other Asian | Wong et al. |
| Yamakoshi | Japanese | Other Asian | Wong et al. |
| Yamamichi | Japanese | Other Asian | Wong et al. |
| Yamamori | Japanese | Other Asian | Wong et al. |
| Yamamoto | Japanese | Other Asian | Wong et al. |
| Yamamura | Japanese | Other Asian | Wong et al. |
| Yamamuro | Japanese | Other Asian | Wong et al. |
| Yamana | Japanese | Other Asian | Wong et al. |
| Yamanaka | Japanese | Other Asian | Wong et al. |
| Yamanashi | Japanese | Other Asian | Wong et al. |
| Yamane | Japanese | Other Asian | Wong et al. |
| Yamanishi | Japanese | Other Asian | Wong et al. |
| Yamano | Japanese | Other Asian | Wong et al. |
| Yamanobe | Japanese | Other Asian | Wong et al. |
| Yamanoha | Japanese | Other Asian | Wong et al. |
| Yamanoi | Japanese | Other Asian | Wong et al. |
| Yamanouchi | Japanese | Other Asian | Wong et al. |
| Yamao | Japanese | Other Asian | Wong et al. |
| Yamaoka | Japanese | Other Asian | Wong et al. |
| Yamasaki | Japanese | Other Asian | Wong et al. |
| Yamashige | Japanese | Other Asian | Wong et al. |
| Yamashina | Japanese | Other Asian | Wong et al. |
| Yamashiro | Japanese | Other Asian | Wong et al. |
| Yamashita | Japanese | Other Asian | Wong et al. |
| Yamat | Filipino | Other Asian | Wong et al. |
| Yamatani | Japanese | Other Asian | Wong et al. |
| Yamate | Japanese | Other Asian | Wong et al. |
| Yamato | Japanese | Other Asian | Wong et al. |
| Yamatsu | Japanese | Other Asian | Wong et al. |
| Yamauchi | Japanese | Other Asian | Wong et al. |
| Yamaura | Japanese | Other Asian | Wong et al. |
| Yamawaki | Japanese | Other Asian | Wong et al. |
| Yamaya | Japanese | Other Asian | Wong et al. |
| Yamazaki | Japanese | Other Asian | Wong et al. |
| Yamazato | Japanese | Other Asian | Wong et al. |
| Yamazoe | Japanese | Other Asian | Wong et al. |
| Yamba | Filipino | Other Asian | Wong et al. |
| Yambao | Filipino | Other Asian | Wong et al. |
| Yambot | Filipino | Other Asian | Wong et al. |
| Yamori | Japanese | Other Asian | Wong et al. |
| Yamson | Filipino | Other Asian | Wong et al. |
| Yamsuan | Filipino | Other Asian | Wong et al. |
| Yamzon | Filipino | Other Asian | Wong et al. |
| Yan | Chinese | Chinese | Kranick et al. |
| Yana | Indonesian | Other Asian | Kranick et al. |
| Yanaga | Japanese | Other Asian | Wong et al. |
| Yanagawa | Japanese | Other Asian | Wong et al. |
| Yanagi | Japanese | Other Asian | Wong et al. |
| Yanagibashi | Japanese | Other Asian | Wong et al. |
| Yanagida | Japanese | Other Asian | Wong et al. |
| Yanagihara | Japanese | Other Asian | Wong et al. |
| Yanagimachi | Japanese | Other Asian | Wong et al. |
| Yanagimoto | Japanese | Other Asian | Wong et al. |
| Yanagisawa | Japanese | Other Asian | Wong et al. |
| Yanagita | Japanese | Other Asian | Wong et al. |
| Yanagiya | Japanese | Other Asian | Wong et al. |
| Yanagizawa | Japanese | Other Asian | Wong et al. |
| Yanai | Japanese | Other Asian | Wong et al. |
| Yanase | Japanese | Other Asian | Wong et al. |
| Yance | Filipino | Other Asian | Wong et al. |
| Yandoc | Filipino | Other Asian | Wong et al. |
| Yanes | Filipino | Other Asian | Wong et al. |
| Yanez | Filipino | Other Asian | Wong et al. |
| Yaneza | Filipino | Other Asian | Wong et al. |
| Yang | Chinese; Korean | Chinese | Kranick et al. |
| Yanga | Filipino | Other Asian | Wong et al. |
| Yangat | Filipino | Other Asian | Wong et al. |
| Yangco | Filipino | Other Asian | Wong et al. |
| Yango | Filipino | Other Asian | Wong et al. |
| Yanit | Filipino | Other Asian | Wong et al. |
| Yankarran |  | South Asian | Lim et al. |
| Yano | Japanese | Other Asian | Wong et al. |
| Yanos | Filipino | Other Asian | Wong et al. |
| Yanson | Filipino | Other Asian | Wong et al. |
| Yanto | Filipino | Other Asian | Wong et al. |
| Yanuaria | Filipino | Other Asian | Wong et al. |
| Yao | Chinese | Chinese | Kranick et al. |
| Yap | Malaysian; Filipino; Chinese | Other Asian | Kranick et al. |
| Yapching | Filipino | Other Asian | Wong et al. |
| Yape | Filipino | Other Asian | Wong et al. |
| Yapit | Filipino | Other Asian | Wong et al. |
| Yapo | Filipino | Other Asian | Wong et al. |
| Yapp | Chinese | Chinese | Wong et al. |
| Yaptangco | Filipino | Other Asian | Wong et al. |
| Yaptinchay | Filipino | Other Asian | Wong et al. |
| Yaqoob |  | South Asian | Lim et al. |
| Yara | Japanese | Other Asian | Wong et al. |
| Yaranon | Filipino | Other Asian | Wong et al. |
| Yarimizu | Japanese | Other Asian | Wong et al. |
| Yarlagadda | Asian Indian | South Asian | Wong et al. |
| Yarte | Filipino | Other Asian | Wong et al. |
| Yarza | Filipino | Other Asian | Wong et al. |
| Yasaka | Japanese | Other Asian | Wong et al. |
| Yasana | Filipino | Other Asian | Wong et al. |
| Yasay | Filipino | Other Asian | Wong et al. |
| Yaseen |  | South Asian | Lim et al. |
| Yashiki | Japanese | Other Asian | Wong et al. |
| Yashima | Japanese | Other Asian | Wong et al. |
| Yashiro | Japanese | Other Asian | Wong et al. |
| Yasis | Filipino | Other Asian | Wong et al. |
| Yasmin |  | South Asian | Lim et al. |
| Yassim |  | South Asian | Lim et al. |
| Yasuda | Japanese | Other Asian | Wong et al. |
| Yasue | Japanese | Other Asian | Wong et al. |
| Yasuhara | Japanese | Other Asian | Wong et al. |
| Yasui | Japanese | Other Asian | Wong et al. |
| Yasukawa | Japanese | Other Asian | Wong et al. |
| Yasukochi | Japanese | Other Asian | Wong et al. |
| Yasuma | Japanese | Other Asian | Wong et al. |
| Yasumatsu | Japanese | Other Asian | Wong et al. |
| Yasumi | Japanese | Other Asian | Wong et al. |
| Yasumoto | Japanese | Other Asian | Wong et al. |
| Yasumura | Japanese | Other Asian | Wong et al. |
| Yasunaga | Japanese | Other Asian | Wong et al. |
| Yasuno | Japanese | Other Asian | Wong et al. |
| Yasuo | Japanese | Other Asian | Wong et al. |
| Yasuoka | Japanese | Other Asian | Wong et al. |
| Yasutake | Japanese | Other Asian | Wong et al. |
| Yasutomi | Japanese | Other Asian | Wong et al. |
| Yat | Chinese | Chinese | Wong et al. |
| Yata | Japanese | Other Asian | Wong et al. |
| Yatabe | Japanese | Other Asian | Wong et al. |
| Yatar | Filipino | Other Asian | Wong et al. |
| Yatco | Filipino | Other Asian | Wong et al. |
| Yato | Japanese | Other Asian | Wong et al. |
| Yatsu | Japanese | Other Asian | Wong et al. |
| Yau | Chinese | Chinese | Wong et al. |
| Yawata | Japanese | Other Asian | Wong et al. |
| Yazaki | Japanese | Other Asian | Wong et al. |
| Yazawa | Japanese | Other Asian | Wong et al. |
| Yazdani | Asian Indian | South Asian | Wong et al. |
| Yazon | Filipino | Other Asian | Wong et al. |
| Yballe | Filipino | Other Asian | Wong et al. |
| Ybanez | Filipino | Other Asian | Wong et al. |
| Ybardolaza | Filipino | Other Asian | Wong et al. |
| Ybay | Filipino | Other Asian | Wong et al. |
| Ybiernas | Filipino | Other Asian | Wong et al. |
| Ycasas | Filipino | Other Asian | Wong et al. |
| Ycasiano | Filipino | Other Asian | Wong et al. |
| Yco | Filipino | Other Asian | Wong et al. |
| Ycoy | Filipino | Other Asian | Wong et al. |
| Ye | Chinese | Chinese | Wong et al. |
| Yea | Chinese | Chinese | Wong et al. |
| Yean | Chinese | Chinese | Wong et al. |
| Yeasmin |  | South Asian | Lim et al. |
| Yeban | Filipino | Other Asian | Wong et al. |
| Yee | Chinese; Burmese; Malaysian | Chinese | Kranick et al. |
| Yeen | Chinese | Chinese | Wong et al. |
| Yeh | Chinese | Chinese | Kranick et al. |
| Yei | Chinese | Chinese | Wong et al. |
| Yein | Chinese | Chinese | Wong et al. |
| Yek | Chinese | Chinese | Wong et al. |
| Yel | Chinese | Chinese | Wong et al. |
| Yeldandi | Asian Indian | South Asian | Wong et al. |
| Yem | Cambodian | Other Asian | Kranick et al. |
| Yen | Chinese | Chinese | Wong et al. |
| Yenchitra | Thai | Other Asian | Kranick et al. |
| Yeng | Chinese | Chinese | Wong et al. |
| Yeo | Korean | Other Asian | Wong et al. |
| Yeom | Korean | Other Asian | Wong et al. |
| Yeon | Korean | Other Asian | Wong et al. |
| Yeong | Chinese | Chinese | Wong et al. |
| Yep | Chinese | Chinese | Wong et al. |
| Yepes | Filipino | Other Asian | Wong et al. |
| Yere | Filipino | Other Asian | Wong et al. |
| Yerro | Filipino | Other Asian | Wong et al. |
| Yesmin | Bangladeshi | South Asian | Kranick et al. |
| Yesudas | Asian Indian | South Asian | Wong et al. |
| Yet | Chinese | Chinese | Wong et al. |
| Yeu | Chinese | Chinese | Wong et al. |
| Yeum | Korean | Other Asian | Wong et al. |
| Yeun | Chinese | Chinese | Wong et al. |
| Yeung | Chinese | Chinese | Kranick et al. |
| Yew | Chinese | Chinese | Wong et al. |
| Ygnacio | Filipino | Other Asian | Wong et al. |
| Yi | Chinese; Korean | Chinese | Kranick et al. |
| Yian | Chinese | Chinese | Wong et al. |
| Yiang | Chinese | Chinese | Wong et al. |
| Yick | Chinese | Chinese | Wong et al. |
| Yie | Chinese | Chinese | Wong et al. |
| Yieh | Chinese | Chinese | Wong et al. |
| Yien | Chinese | Chinese | Wong et al. |
| Yih | Taiwanese; Malaysian | Other Asian | Kranick et al. |
| Yii | Chinese | Chinese | Wong et al. |
| Yik | Chinese | Chinese | Wong et al. |
| Yim | Chinese; Korean | Chinese | Kranick et al. |
| Yin | Chinese | Chinese | Kranick et al. |
| Ying | Chinese | Chinese | Wong et al. |
| Yip | Chinese | Chinese | Kranick et al. |
| Yisuf |  | South Asian | Lim et al. |
| Yit | Chinese | Chinese | Wong et al. |
| Yiu | Chinese | Chinese | Kranick et al. |
| Ylagan | Filipino | Other Asian | Wong et al. |
| Ylanan | Filipino | Other Asian | Wong et al. |
| Ylarde | Filipino | Other Asian | Wong et al. |
| Ylaya | Filipino | Other Asian | Wong et al. |
| Ylo | Filipino | Other Asian | Wong et al. |
| Ymson | Filipino | Other Asian | Wong et al. |
| Yn | Chinese | Chinese | Wong et al. |
| Yniguez | Filipino | Other Asian | Wong et al. |
| Yo | Korean | Other Asian | Wong et al. |
| Yock | Chinese | Chinese | Wong et al. |
| Yoda | Japanese | Other Asian | Wong et al. |
| Yodh | Asian Indian | South Asian | Wong et al. |
| Yoen | Chinese | Chinese | Wong et al. |
| Yogi | Japanese | Other Asian | Wong et al. |
| Yogo | Japanese | Other Asian | Wong et al. |
| Yoh | Chinese | Chinese | Wong et al. |
| Yohannan | Asian Indian | South Asian | Wong et al. |
| Yoingco | Filipino | Other Asian | Wong et al. |
| Yok | Chinese | Chinese | Wong et al. |
| Yoke | Chinese | Chinese | Wong et al. |
| Yoko | Japanese | Other Asian | Wong et al. |
| Yokobori | Japanese | Other Asian | Wong et al. |
| Yokoda | Japanese | Other Asian | Wong et al. |
| Yokoe | Japanese | Other Asian | Wong et al. |
| Yokogawa | Japanese | Other Asian | Wong et al. |
| Yokohama | Japanese | Other Asian | Wong et al. |
| Yokoi | Japanese | Other Asian | Wong et al. |
| Yokokawa | Japanese | Other Asian | Wong et al. |
| Yokol | Japanese | Other Asian | Wong et al. |
| Yokomizo | Japanese | Other Asian | Wong et al. |
| Yokoo | Japanese | Other Asian | Wong et al. |
| Yokoro | Japanese | Other Asian | Wong et al. |
| Yokosawa | Japanese | Other Asian | Wong et al. |
| Yokose | Japanese | Other Asian | Wong et al. |
| Yokota | Japanese | Other Asian | Wong et al. |
| Yokote | Japanese | Other Asian | Wong et al. |
| Yokotsuka | Japanese | Other Asian | Wong et al. |
| Yokouchi | Japanese | Other Asian | Wong et al. |
| Yokoyama | Japanese | Other Asian | Wong et al. |
| Yokozawa | Japanese | Other Asian | Wong et al. |
| Yol | Korean | Other Asian | Wong et al. |
| Yolangco | Filipino | Other Asian | Wong et al. |
| Yolo | Filipino | Other Asian | Wong et al. |
| Yom | Korean | Other Asian | Wong et al. |
| Yonaha | Japanese | Other Asian | Wong et al. |
| Yonamine | Japanese | Other Asian | Wong et al. |
| Yonashiro | Japanese | Other Asian | Wong et al. |
| Yoneda | Japanese | Other Asian | Wong et al. |
| Yonehara | Japanese | Other Asian | Wong et al. |
| Yonei | Japanese | Other Asian | Wong et al. |
| Yonekawa | Japanese | Other Asian | Wong et al. |
| Yonekura | Japanese | Other Asian | Wong et al. |
| Yonemitsu | Japanese | Other Asian | Wong et al. |
| Yonemoto | Japanese | Other Asian | Wong et al. |
| Yonemura | Japanese | Other Asian | Wong et al. |
| Yonenaga | Japanese | Other Asian | Wong et al. |
| Yoneoka | Japanese | Other Asian | Wong et al. |
| Yoneya | Japanese | Other Asian | Wong et al. |
| Yoneyama | Japanese | Other Asian | Wong et al. |
| Yonezawa | Japanese | Other Asian | Wong et al. |
| Yonzon | Filipino | Other Asian | Wong et al. |
| Yoo | Korean | Other Asian | Kranick et al. |
| Yook | Chinese | Chinese | Wong et al. |
| Yoon | Korean | Other Asian | Kranick et al. |
| Yop | Korean | Other Asian | Wong et al. |
| Yor | Chinese | Chinese | Wong et al. |
| Yorac | Filipino | Other Asian | Wong et al. |
| Yoro | Filipino | Other Asian | Wong et al. |
| Yorozu | Japanese | Other Asian | Wong et al. |
| Yorro | Filipino | Other Asian | Wong et al. |
| Yoshi | Japanese | Other Asian | Wong et al. |
| Yoshida | Japanese | Other Asian | Kranick et al. |
| Yoshidome | Japanese | Other Asian | Wong et al. |
| Yoshie | Japanese | Other Asian | Wong et al. |
| Yoshihara | Japanese | Other Asian | Wong et al. |
| Yoshihashi | Japanese | Other Asian | Wong et al. |
| Yoshihiro | Japanese | Other Asian | Wong et al. |
| Yoshii | Japanese | Other Asian | Wong et al. |
| Yoshiike | Japanese | Other Asian | Wong et al. |
| Yoshikawa | Japanese | Other Asian | Kranick et al. |
| Yoshiki | Japanese | Other Asian | Wong et al. |
| Yoshikura | Japanese | Other Asian | Wong et al. |
| Yoshimaru | Japanese | Other Asian | Wong et al. |
| Yoshimatsu | Japanese | Other Asian | Wong et al. |
| Yoshimi | Japanese | Other Asian | Wong et al. |
| Yoshimine | Japanese | Other Asian | Wong et al. |
| Yoshimitsu | Japanese | Other Asian | Wong et al. |
| Yoshimizu | Japanese | Other Asian | Wong et al. |
| Yoshimori | Japanese | Other Asian | Wong et al. |
| Yoshimoto | Japanese | Other Asian | Wong et al. |
| Yoshimura | Japanese | Other Asian | Wong et al. |
| Yoshinaga | Japanese | Other Asian | Wong et al. |
| Yoshinari | Japanese | Other Asian | Wong et al. |
| Yoshino | Japanese | Other Asian | Wong et al. |
| Yoshio | Japanese | Other Asian | Wong et al. |
| Yoshioka | Japanese | Other Asian | Wong et al. |
| Yoshitaka | Japanese | Other Asian | Wong et al. |
| Yoshitake | Japanese | Other Asian | Wong et al. |
| Yoshitaro | Japanese | Other Asian | Wong et al. |
| Yoshitomi | Japanese | Other Asian | Wong et al. |
| Yoshiya | Japanese | Other Asian | Wong et al. |
| Yoshiyama | Japanese | Other Asian | Wong et al. |
| Yoshizaki | Japanese | Other Asian | Wong et al. |
| Yoshizawa | Japanese | Other Asian | Wong et al. |
| Yoshizumi | Japanese | Other Asian | Wong et al. |
| Yosida | Japanese | Other Asian | Wong et al. |
| Yosores | Filipino | Other Asian | Wong et al. |
| Yosuico | Filipino | Other Asian | Wong et al. |
| Yot | Chinese | Chinese | Wong et al. |
| Yota | Japanese | Other Asian | Wong et al. |
| Yotsumoto | Japanese | Other Asian | Wong et al. |
| Yotsuya | Japanese | Other Asian | Wong et al. |
| You | Korean; Chinese | Other Asian | Kranick et al. |
| Youh | Korean | Other Asian | Wong et al. |
| Youm | Korean | Other Asian | Wong et al. |
| Youn | Korean | Other Asian | Kranick et al. |
| Younis | Pakistani | South Asian | Kranick et al. |
| Younus |  | South Asian | Lim et al. |
| Yousef | Pakistani | South Asian | Kranick et al. |
| Yousif | Asian Indian | South Asian | Wong et al. |
| Youssuff |  | South Asian | Lim et al. |
| Yousuf |  | South Asian | Lim et al. |
| Yousuff | Asian Indian | South Asian | Wong et al. |
| Yousufuddin | Asian Indian | South Asian | Wong et al. |
| Youtso |  | South Asian | Lim et al. |
| Yow | Chinese | Chinese | Wong et al. |
| Yoza | Japanese | Other Asian | Wong et al. |
| Ypil | Filipino | Other Asian | Wong et al. |
| Yra | Filipino | Other Asian | Wong et al. |
| Yraola | Filipino | Other Asian | Wong et al. |
| Yrastorza | Filipino | Other Asian | Wong et al. |
| Yray | Filipino | Other Asian | Wong et al. |
| Yrigan | Filipino | Other Asian | Wong et al. |
| Ysip | Filipino | Other Asian | Wong et al. |
| Ysit | Filipino | Other Asian | Wong et al. |
| Ysla | Filipino | Other Asian | Wong et al. |
| Ysmael | Filipino | Other Asian | Wong et al. |
| Yson | Filipino | Other Asian | Wong et al. |
| Ytem | Filipino | Other Asian | Wong et al. |
| Yturralde | Filipino | Other Asian | Wong et al. |
| Yu | Chinese | Chinese | Kranick et al. |
| Yuam | Chinese | Chinese | Wong et al. |
| Yuan | Chinese | Chinese | Kranick et al. |
| Yuang | Chinese | Chinese | Wong et al. |
| Yuasa | Japanese | Other Asian | Wong et al. |
| Yuck | Chinese | Chinese | Wong et al. |
| Yuda | Japanese | Other Asian | Wong et al. |
| Yue | Chinese | Chinese | Wong et al. |
| Yueh | Chinese | Chinese | Wong et al. |
| Yuen | Chinese | Chinese | Kranick et al. |
| Yueng | Chinese | Chinese | Wong et al. |
| Yuet | Chinese | Chinese | Wong et al. |
| Yuey | Chinese | Chinese | Wong et al. |
| Yuge | Japanese | Other Asian | Wong et al. |
| Yuh | Korean | Other Asian | Wong et al. |
| Yuhara | Japanese | Other Asian | Wong et al. |
| Yui | Japanese | Other Asian | Kranick et al. |
| Yuin | Chinese | Chinese | Wong et al. |
| Yuji | Japanese | Other Asian | Wong et al. |
| Yujuico | Filipino | Other Asian | Wong et al. |
| Yuk | Chinese | Chinese | Wong et al. |
| Yukawa | Japanese | Other Asian | Wong et al. |
| Yuki | Japanese | Other Asian | Wong et al. |
| Yul | Korean | Other Asian | Wong et al. |
| Yulo | Filipino | Other Asian | Wong et al. |
| Yum | Korean | Other Asian | Wong et al. |
| Yumang | Filipino | Other Asian | Wong et al. |
| Yumol | Filipino | Other Asian | Wong et al. |
| Yumoto | Japanese | Other Asian | Wong et al. |
| Yumul | Filipino | Other Asian | Wong et al. |
| Yumura | Japanese | Other Asian | Wong et al. |
| Yun | Chinese; Korean | Chinese | Kranick et al. |
| Yune | Korean | Other Asian | Wong et al. |
| Yung | Chinese | Chinese | Kranick et al. |
| Yuno | Japanese | Other Asian | Wong et al. |
| Yunoki | Japanese | Other Asian | Wong et al. |
| Yuo | Chinese | Chinese | Wong et al. |
| Yup | Chinese | Chinese | Wong et al. |
| Yuri | Japanese | Other Asian | Wong et al. |
| Yusa | Japanese | Other Asian | Wong et al. |
| Yusay | Filipino | Other Asian | Wong et al. |
| Yusi | Filipino | Other Asian | Wong et al. |
| Yuson | Filipino | Other Asian | Wong et al. |
| Yusuf |  | South Asian | Lim et al. |
| Yut | Chinese | Chinese | Wong et al. |
| Yutani | Japanese | Other Asian | Wong et al. |
| Yutuc | Filipino | Other Asian | Wong et al. |
| Yuu | Chinese | Chinese | Wong et al. |
| Yuvienco | Filipino | Other Asian | Wong et al. |
| Yuyama | Japanese | Other Asian | Wong et al. |
| Yuzawa | Japanese | Other Asian | Wong et al. |
| Yuzon | Filipino | Other Asian | Wong et al. |
| Zabala | Filipino | Other Asian | Wong et al. |
| Zaballa | Filipino | Other Asian | Wong et al. |
| Zaballero | Filipino | Other Asian | Wong et al. |
| Zabat | Filipino | Other Asian | Wong et al. |
| Zabate | Filipino | Other Asian | Wong et al. |
| Zablan | Filipino | Other Asian | Wong et al. |
| Zacarias | Filipino | Other Asian | Wong et al. |
| Zacharia | Asian Indian | South Asian | Wong et al. |
| Zachariah | Asian Indian | South Asian | Wong et al. |
| Zafar | Bangladeshi | South Asian | Kranick et al. |
| Zafra | Filipino | Other Asian | Wong et al. |
| Zafranco | Filipino | Other Asian | Wong et al. |
| Zagala | Filipino | Other Asian | Kranick et al. |
| Zaguirre | Filipino | Other Asian | Wong et al. |
| Zahed |  | South Asian | Lim et al. |
| Zaheer | Asian Indian | South Asian | Wong et al. |
| Zahid |  | South Asian | Lim et al. |
| Zahir |  | South Asian | Lim et al. |
| Zahiruddin |  | South Asian | Lim et al. |
| Zahoor | Asian Indian | South Asian | Wong et al. |
| Zai | Chinese | Chinese | Wong et al. |
| Zaide | Filipino | Other Asian | Wong et al. |
| Zaidi | Asian Indian | South Asian | Wong et al. |
| Zaima | Japanese | Other Asian | Wong et al. |
| Zainuddin | Asian Indian | South Asian | Wong et al. |
| Zaka |  | South Asian | Lim et al. |
| Zakaria |  | South Asian | Lim et al. |
| Zaki | Asian Indian | South Asian | Wong et al. |
| Zakir |  | South Asian | Lim et al. |
| Zalamea | Filipino | Other Asian | Wong et al. |
| Zalameda | Filipino | Other Asian | Wong et al. |
| Zalavadia | Asian Indian | South Asian | Wong et al. |
| Zaldarriaga | Filipino | Other Asian | Wong et al. |
| Zaldivar | Filipino | Other Asian | Wong et al. |
| Zaldua | Filipino | Other Asian | Wong et al. |
| Zamader |  | South Asian | Lim et al. |
| Zamal |  | South Asian | Lim et al. |
| Zamaluddin |  | South Asian | Lim et al. |
| Zaman |  | South Asian | Lim et al. |
| Zamboanga | Filipino | Other Asian | Wong et al. |
| Zambrano | Filipino | Other Asian | Wong et al. |
| Zameer |  | South Asian | Lim et al. |
| Zamir |  | South Asian | Lim et al. |
| Zamora | Filipino | Other Asian | Wong et al. |
| Zamoranos | Filipino | Other Asian | Wong et al. |
| Zampaga | Filipino | Other Asian | Wong et al. |
| Zamuco | Filipino | Other Asian | Wong et al. |
| Zamudio | Filipino | Other Asian | Wong et al. |
| Zandueta | Filipino | Other Asian | Wong et al. |
| Zane | Chinese | Chinese | Wong et al. |
| Zannat |  | South Asian | Lim et al. |
| Zano | Filipino | Other Asian | Wong et al. |
| Zanoria | Filipino | Other Asian | Wong et al. |
| Zantua | Filipino | Other Asian | Wong et al. |
| Zao | Chinese | Chinese | Wong et al. |
| Zapanta | Filipino | Other Asian | Wong et al. |
| Zapata | Filipino | Other Asian | Wong et al. |
| Zapatos | Filipino | Other Asian | Wong et al. |
| Zara | Filipino | Other Asian | Wong et al. |
| Zaragosa | Filipino | Other Asian | Wong et al. |
| Zaragoza | Filipino | Other Asian | Wong et al. |
| Zarandin | Filipino | Other Asian | Wong et al. |
| Zaraspe | Filipino | Other Asian | Wong et al. |
| Zarate | Filipino | Other Asian | Wong et al. |
| Zarco | Filipino | Other Asian | Wong et al. |
| Zareno | Filipino | Other Asian | Wong et al. |
| Zarraga | Filipino | Other Asian | Wong et al. |
| Zarsadias | Filipino | Other Asian | Wong et al. |
| Zarza | Filipino | Other Asian | Wong et al. |
| Zarzoso | Filipino | Other Asian | Wong et al. |
| Zarzuela | Filipino | Other Asian | Wong et al. |
| Zaspa | Filipino | Other Asian | Wong et al. |
| Zau | Chinese | Chinese | Wong et al. |
| Zaver | Asian Indian | South Asian | Wong et al. |
| Zaverbhai | Asian Indian | South Asian | Wong et al. |
| Zaveri | Asian Indian | South Asian | Wong et al. |
| Zavosh | Asian Indian | South Asian | Wong et al. |
| Zayas | Filipino | Other Asian | Wong et al. |
| Zayco | Filipino | Other Asian | Wong et al. |
| Ze | Chinese | Chinese | Wong et al. |
| Zee | Pakistani | South Asian | Kranick et al. |
| Zehra |  | South Asian | Lim et al. |
| Zen | Chinese | Chinese | Wong et al. |
| Zenarosa | Filipino | Other Asian | Wong et al. |
| Zeng | Chinese | Chinese | Kranick et al. |
| Zepeda | Filipino | Other Asian | Wong et al. |
| Zerda | Filipino | Other Asian | Wong et al. |
| Zerna | Filipino | Other Asian | Wong et al. |
| Zerrudo | Filipino | Other Asian | Wong et al. |
| Zervoulakos | Filipino | Other Asian | Wong et al. |
| Zeta | Filipino | Other Asian | Wong et al. |
| Zha | Chinese | Chinese | Wong et al. |
| Zhad | Chinese | Chinese | Wong et al. |
| Zhai | Chinese | Chinese | Wong et al. |
| Zhan | Chinese | Chinese | Kranick et al. |
| Zhang | Chinese | Chinese | Kranick et al. |
| Zhao | Chinese | Chinese | Kranick et al. |
| Zhen | Chinese | Chinese | Wong et al. |
| Zheng | Chinese | Chinese | Kranick et al. |
| Zhi | Chinese | Chinese | Wong et al. |
| Zhon | Chinese | Chinese | Wong et al. |
| Zhong | Chinese | Chinese | Wong et al. |
| Zhou | Chinese | Chinese | Kranick et al. |
| Zhu | Chinese | Chinese | Kranick et al. |
| Zhuang | Chinese | Chinese | Kranick et al. |
| Zhuo | Chinese | Chinese | Wong et al. |
| Zi | Chinese | Chinese | Wong et al. |
| Zia |  | South Asian | Lim et al. |
| Zialcita | Filipino | Other Asian | Wong et al. |
| Ziang | Chinese | Chinese | Wong et al. |
| Ziauddin | Asian Indian | South Asian | Wong et al. |
| Ziaullah |  | South Asian | Lim et al. |
| Zien | Chinese | Chinese | Wong et al. |
| Zing | Chinese | Chinese | Wong et al. |
| Zinnath |  | South Asian | Lim et al. |
| Zipagan | Filipino | Other Asian | Wong et al. |
| Zita | Filipino | Other Asian | Wong et al. |
| Zohra | Bangladeshi; Pakistani | South Asian | Kranick et al. |
| Zoleta | Filipino | Other Asian | Wong et al. |
| Zong | Chinese | Chinese | Wong et al. |
| Zonio | Filipino | Other Asian | Wong et al. |
| Zoo | Chinese | Chinese | Wong et al. |
| Zorilla | Filipino | Other Asian | Wong et al. |
| Zorrilla | Filipino | Other Asian | Wong et al. |
| Zosa | Filipino | Other Asian | Wong et al. |
| Zou | Chinese | Chinese | Kranick et al. |
| Zozobrado | Filipino | Other Asian | Wong et al. |
| Zu | Chinese | Chinese | Wong et al. |
| Zubair | Pakistani | South Asian | Kranick et al. |
| Zubairi | Asian Indian | South Asian | Wong et al. |
| Zuberi | Asian Indian | South Asian | Wong et al. |
| Zubiri | Filipino | Other Asian | Wong et al. |
| Zue | Chinese | Chinese | Wong et al. |
| Zulaybar | Filipino | Other Asian | Wong et al. |
| Zulfiker |  | South Asian | Lim et al. |
| Zulfiqar |  | South Asian | Lim et al. |
| Zulfiquar |  | South Asian | Lim et al. |
| Zuluaga | Filipino | Other Asian | Wong et al. |
| Zulueta | Filipino | Other Asian | Wong et al. |
| Zumel | Filipino | Other Asian | Wong et al. |
| Zun | Chinese | Chinese | Wong et al. |
| Zung | Chinese | Chinese | Wong et al. |
| Zuniga | Filipino | Other Asian | Wong et al. |
| Zuno | Filipino | Other Asian | Wong et al. |
| Zuo | Chinese | Chinese | Wong et al. |
| Zuraek | Filipino | Other Asian | Wong et al. |
| Zurbano | Filipino | Other Asian | Wong et al. |
| Zurbito | Filipino | Other Asian | Wong et al. |
| Zurita | Filipino | Other Asian | Wong et al. |
| Zutshi | Asian Indian | South Asian | Wong et al. |
